# Identification of 64 new risk loci for major depression, refinement of the genetic architecture and risk prediction of recurrence and comorbidities

**DOI:** 10.1101/2022.08.24.22279149

**Authors:** Thomas D. Als, Mitja Kurki, Jakob Grove, Georgios Voloudakis, Karen Therrien, Elisa Tasanko, Trine Tollerup Nielsen, Joonas Naamanka, Kumar Veerapen, Daniel Levey, Jaroslav Bendl, Jonas Bybjerg-Grauholm, Biao Zheng, Ditte Demontis, Anders Rosengren, Georgios Athanasiadis, Marie Bækved-Hansen, Per Qvist, Bragi Walters, Thorgeir Thorgeirsson, Hreinn Stefánsson, Katherine L Musliner, Veera Manikandan, Leila Farajzadeh, Janne Thirstrup, Bjarni J. Vilhjálmsson, John J. McGrath, Manuel Mattheisen, Sandra Meier, iPSYCH-Broad Consortium, Esben Agerbo, Kári Stefánsson, Merete Nordentoft, Thomas Werge, David M. Hougaard, Preben B. Mortensen, Murray Stein, Joel Gelernter, Iiris Hovatta, Panos Roussos, Mark J. Daly, Ole Mors, Aarno Palotie, Anders D. Børglum

**Affiliations:** Department of Biomedicine, Aarhus University, Aarhus, Denmark; The Lundbeck Foundation Initiative for Integrative Psychiatric Research, iPSYCH, Denmark; Center for Genomics and Personalized Medicine, Aarhus, Denmark; Institute for Molecular Medicine Finland, University of Helsinki, Helsinki, Finland; Bioinformatics Research Centre, Aarhus University, 8000 Aarhus C, Denmark; Center for Disease Neurogenomics, Icahn School of Medicine at Mount Sinai, NY, USA; Department of Psychiatry, Icahn School of Medicine at Mount Sinai, NY, USA; Department of Genetics and Genomic Sciences, Icahn School of Medicine at Mount Sinai, New York, NY, USA; Icahn Institute for Data Science and Genomic Technology, Icahn School of Medicine at Mount Sinai, NY, USA; Friedman Brain Institute, Icahn School of Medicine at Mount Sinai, New York, NY, USA; Mental Illness Research, Education, and Clinical Center (VISN 2 South), James J. Peters VA Medical Center, Bronx, NY, USA; Nash Family Department of Neuroscience, Icahn School of Medicine at Mount Sinai, New York, NY, USA; Department of Psychology and Logopedics, SleepWell Research Program, University of Helsinki, Finland; Analytic and Translational Genetics Unit, Department of Medicine, Massachusetts General Hospital and Harvard Medical School, Boston, MA, USA; Stanley Center for Psychiatric Research, Broad Institute of MIT and Harvard, Cambridge, MA, USA; Department of Medicine, Harvard Medical School, Boston, MA, USA; Division of Human Genetics, Department of Psychiatry, Yale University School of Medicine, New Haven, CT, USA; Department of Psychiatry, Veterans Affairs Connecticut Healthcare Center, West Haven, CT, USA; Center for Neonatal Screening, Department for Congenital Disorders, Statens Serum Institut, Copenhagen, Denmark; Mental Health Centre Sct. Hans, Capital Region of Denmark, Institute of Biological Psychiatry, Copenhagen University Hospital, Copenhagen, Denmark; deCODE genetics / Amgen, Reykjavik, Iceland; NCRR - National Centre for Register-Based Research, Business and Social Sciences, Aarhus University, Aarhus, Denmark; Centre for Integrated Register-based Research, CIRRAU, Aarhus University, Aarhus, Denmark; Department of Affective Disorders, Aarhus University Hospital-Psychiatry, Aarhus Denmark; National Centre for Register-Based Research, Aarhus University, 8210 Aarhus V, Denmark; Queensland Centre for Mental Health Research, The Park Centre for Mental Health, Brisbane, QLD 4076, Australia; Queensland Brain Institute, University of Queensland, Brisbane, QLD 4072, Australia; Mental Health Centre Copenhagen, Capital Region of Denmark, Copenhagen University Hospital, Copenhagen, Denmark; Psychiatry Service, VA San Diego Healthcare System, San Diego, CA, USA; Departments of Psychiatry and Herbert Wertheim School of Public Health, University of California, San Diego, La Jolla, CA, USA; Center for Dementia Research, Nathan Kline Institute for Psychiatric Research, Orangeburg, NY 10962, USA; Psychosis Research Unit, Aarhus University Hospital-Psychiatry, Denmark

## Abstract

Major depression (MD) is a common mental disorder and a leading cause of disability worldwide. We conducted a GWAS meta-analysis of more than 1.3 million individuals, including 371,184 with MD, identifying 243 risk loci. Sixty-four loci are novel, including glutamate and GABA receptors that are targets for antidepressant drugs. Several biological pathways and components were enriched for genetic MD risk, implicating neuronal development and function. Intersection with functional genomics data prioritized likely causal genes and revealed novel enrichment of prenatal GABAergic neurons, astrocytes and oligodendrocyte lineages.

We found MD to be highly polygenic, with around 11,700 variants explaining 90% of the SNP heritability. Bivariate Gaussian mixture modeling estimated that > 97% of risk variants for other psychiatric disorders (anxiety, schizophrenia, bipolar disorder and ADHD) are influencing MD risk when both concordant and discordant variants are considered, and nearly all MD risk variants influence educational attainment. Additionally, we demonstrated that MD genetic risk is associated with impaired complex cognition, including verbal reasoning, attention, abstraction and mental flexibility.

Analyzing Danish nation-wide longitudinal data, we dissected the genetic and clinical heterogeneity, revealing distinct polygenic architectures across case subgroups of MD recurrency and psychiatric comorbidity and demonstrating two-to six-fold increases in absolute risks for developing comorbid psychiatric disorders among MD cases with the highest versus the lowest polygenic burden.

The results deepen the understanding of the biology underlying MD and its progression and inform precision medicine approaches in MD.

## Introduction

Major depression (MD) is a genetic and phenotypic complex disorder with a lifetime prevalence of 15-20%^1-3^. It is often recurrent and accompanied by considerable morbidity and co-morbidity, excess mortality, increased risk of suicide and substantial costs worldwide^4-8^. Individuals diagnosed with MD have an increased risk of developing practically all other types of mental disorders, particularly anxiety (ANX), bipolar disorder (BD), schizophrenia (SZ) and substance use disorder (SUD)^9,10^.

Heritability estimates based on twin studies (*h*^2^ = 0.37) have indicated that familial aggregation of MD is influenced by additive genetic effects^11^, and several recent studies have documented a considerable genetic overlap between MD and multiple psychiatric as well as somatic disorders and traits^12-17^. Despite the substantial heritability of MD and other mental disorders, the potential for translating genetic insights into precision psychiatry has yet to be fulfilled, including demonstrating clinical utility of polygenic risk scores (PRS)^18-20^.

Major advancement in understanding the genetic architecture of MD has only recently been achieved, primarily via genome-wide association studies (GWAS) led by the Psychiatric Genomics Consortium (PGC). Most recently, GWAS results from the PGC^12^, UK Biobank (UKB)^14,21^, FinnGen and 23andMe, Inc.^22^ were combined with data from the Million Veteran Program (MVP) in a large meta-analysis^23^, identifying 178 risk loci. The identified loci explain a small fraction of the overall heritability of MD^12,21,23^ and even larger GWASs are needed to further elucidate genetic factors contributing to the risk of developing MD and advance genetically informed patient stratification and outcome prediction towards clinical utility.

Here, we present the largest MD GWAS to date, including expanded iPSYCH^24^ and FinnGen cohorts, meta-analyzed with PGC, UKB, 23andMe and MVP data^23^, revealing numerous novel risk loci and characterizing functional implications of associated variants by intersecting with functional genomics data. We refine the genetic architecture of MD and case subgroups, and demonstrate the impact of MD genetic risk on domains of cognitive performance. Leveraging nation-wide longitudinal health data on the Danish iPSYCH cohort, we dissect the genetic architecture of single-episode and recurrent MD as well as cases who have developed ANX, BD, SZ and SUD. Furthermore, to inform precision psychiatry approaches, we calculate time-dependent absolute risks and hazard rate ratios for developing recurrent depression, ANX, BP, SZ and SUD depending on different polygenic burdens of MD cases.

## Results

### Genome-wide association

We analyzed data from the large population-based case-cohort of iPSYCH^24,25^, which include genotypes from all individuals born in Denmark between 1981 and 2008 who have received treatment for MD in hospitals and outpatient clinics (ICD-10 codes F32-F33 in the Danish Psychiatric Central Research Register^26^). Compared to the latest GWAS of MD^23^, which included samples from the initial iPSYCH2012 cohort^12,25^, we added 11,710 cases and 18,410 controls from the expanded iPSYCH2015 cohort^24,25^, summarizing to a total of 30,618 cases and 38,200 controls after relatedness pruning and removal of ancestry outliers. In addition, we included an updated dataset consisting of 28,098 cases and 228,817 controls from the FinnGen study. When combining these with data from previously published samples from the PGC, UKB, 23andMe and MVP, the number of samples added up to a total of 371,184 MD cases and 978,703 controls (Supplementary Table S1).

We performed a variance weighted fixed effects meta-analysis using METAL^27^, testing the effects of 6,037,120 SNPs common across all the data sets. This revealed a total of 303 genome-wide significant LD-independent (*r*^2^ < 0.1) lead variants located in 243 distinct loci. A conditional association analysis using GCTA-COJO^28,29^ retained 251 independent SNPs in the 243 loci. Manhattan plots are shown in Figure 1, regional plots are provided in Supplementary Figure S2 and details on lead variants are provided in Supplementary Table S2A, while GCTA-COJO independent variants are listed in Supplementary Table S2B. No statistically significant heterogeneity was observed between the datasets (Supplementary Figure S3). Sixty-four (64) of the 243 loci are novel, i.e., not overlapping with the two most recent MD meta-analyses^14,23^ (Supplementary Table S2A). The three most significant loci were located near *NEGR1*, in *SORCS3* and in the *HIST1* histone cluster, respectively. Among the novel loci, the three strongest associations were in *BPTF, LINGO1* and *GRIA1* (Table 1). All three genes have been associated with monogenic forms of neurodevelopmental disorders^30-32^ and *GRIA1*, encoding glutamate ionotropic receptor AMPA type subunit 1 (GluA1), is the first genome-wide significant locus implicating an AMPA receptor subunit in MD. We also note that the seventh-strongest novel locus is located in *GABRA1*, which is the first time a GABA receptor locus has been identified in GWAS of MD. Both AMPA and GABA receptors are targets for antidepressants^33,34^.

**Table 1:** Top 5 index SNPs for previous and top 15 index SNPs for novel MD GWAS loci. Results for the top 5 among previously identified loci and top 15 novel genome-wide significant index variants identified in the GWAS meta-analysis of 371,184 cases with Major Depression and 978,703 controls. The location (chromosome (Chr)) base position (BP)), alleles (A1/A2), odds ratio (OR) of the effect with respect to A1, standard error (SE) and association P-values (P) from inverse-variance weighted fixed effects model of the index variants are given. “Novel” indicates if the locus is a novel MD risk locus i.e. not identified previously. Genes located within 50 kb from index variants are listed (See supplementary table 2 for details on all 303 index SNPs)

| Loc | Chr | indexSNP | BP | A1/A2 | cases | ctrls | OR | SE | P | Nearby genes | Novel |
| --- | --- | --- | --- | --- | --- | --- | --- | --- | --- | --- | --- |
| 8 | 1 | rs7531118 | 72837239 | T/C | 0.473 | 0.461 | 0.97 | 0.0029 | 1.5e-27 | *NEGR1* | no |
| 9 | 1 | rs10890020 | 73668836 | A/G | 0.508 | 0.510 | 0.97 | 0.0029 | 1.7e-21 | *LINC01360* | no |
| 38 | 2 | rs1320138 | 144158287 | T/C | 0.428 | 0.435 | 1.02 | 0.0028 | 4.8e-09 | *ARHGAP15* | yes |
| 45 | 2 | rs6715105 | 198445601 | T/C | 0.326 | 0.341 | 1.02 | 0.0030 | 1.3e-09 | *ANKRD44, ANKRD44-IT1, SF3B1, COQ10B, HSPD1, SNORA105A, SNORA105B, HSPE1, HSPE1-MOB4, MOB4, RFTN2, MARS2, BOLL, PLCL1* | yes |
| 53 | 3 | rs56029819 | 43478295 | T/C | 0.853 | 0.849 | 0.98 | 0.0040 | 2.5e-09 | *SNRK, SNRK-AS1, ANO10* | yes |
| 80 | 5 | rs4262121 | 31078958 | G/C | 0.528 | 0.546 | 0.98 | 0.0029 | 4.3e-09 | - | yes |
| 89 | 5 | rs62379847 | 120109119 | A/C | 0.655 | 0.652 | 0.98 | 0.0030 | 1.8e-09 | *PRR16* | yes |
| 93 | 5 | rs2964003 | 153216733 | A/G | 0.829 | 0.820 | 1.02 | 0.0038 | 8.1e-10 | *GRIA1, LINC01861* | yes |
| 94 | 5 | rs4596421 | 161269895 | C/T | 0.708 | 0.693 | 0.98 | 0.0031 | 2.4e-09 | *GABRA1* | yes |
| 98 | 6 | rs35883476 | 28368508 | G/C | 0.910 | 0.918 | 1.05 | 0.0052 | 1.3e-21 | *HIST1 histone cluster, BTN3A2, BTN2A2, BTN3A1, BTN2A3P, BTN3A3, BTN2A1, LOC285819, BTN1A1, HCG11, HMGN4, LOC105374988, ABT1, ZNF322, GUSBP2, LINC00240, LOC100270746, MIR3143, PRSS16, POM121L2, VN1R10P, ZNF204P, ZNF391, ZNF184, LINC01012, LOC100131289, OR2B2, OR2B6, ZNF165, ZSCAN12P1, ZSCAN16-AS1, ZSCAN16, ZKSCAN8, ZNF192P1, TOB2P1, ZSCAN9, ZKSCAN4, NKAPL, ZSCAN26, PGBD1, ZSCAN31, ZKSCAN3, ZSCAN12, ZSCAN23* | no |
| 99 | 6 | rs10947690 | 37631768 | A/G | 0.751 | 0.756 | 0.98 | 0.0033 | 6.3e-09 | *MDGA1* | yes |
| 114 | 7 | rs957360 | 3660918 | C/G | 0.714 | 0.702 | 1.02 | 0.0031 | 1.3e-09 | *SDK1* | yes |
| 124 | 7 | rs1986692 | 133743393 | A/G | 0.612 | 0.610 | 1.02 | 0.0029 | 2.8e-09 | *EXOC4* | yes |
| 156 | 10 | rs1909696 | 77582203 | G/T | 0.337 | 0.328 | 0.98 | 0.0030 | 3.3e-09 | *LRMDA, LOC105378367* | yes |
| 158 | 10 | rs1021363 | 106610839 | A/G | 0.349 | 0.335 | 1.03 | 0.0029 | 5.3e-25 | *SORCS3* | no |
| 191 | 13 | rs9561331 | 94017476 | G/A | 0.869 | 0.873 | 0.98 | 0.0042 | 3.7e-09 | *GPC6* | yes |
| 199 | 14 | rs7141014 | 98667928 | T/C | 0.796 | 0.806 | 0.98 | 0.0035 | 3.3e-09 | - | yes |
| 210 | 15 | rs4886915 | 78075023 | A/G | 0.421 | 0.423 | 0.98 | 0.0028 | 6.1e-10 | *LINGO1* | yes |
| 225 | 17 | rs60856912 | 65892343 | G/T | 0.825 | 0.817 | 0.97 | 0.0038 | 1.8e-11 | *BPTF, C17orf58, KPNA2* | yes |
| 233 | 18 | rs12967143 | 53099012 | G/C | 0.301 | 0.292 | 1.03 | 0.0031 | 2.0e-21 | *TCF4, TCF4-AS1, MIR4529* | no |

**Figure 1:**
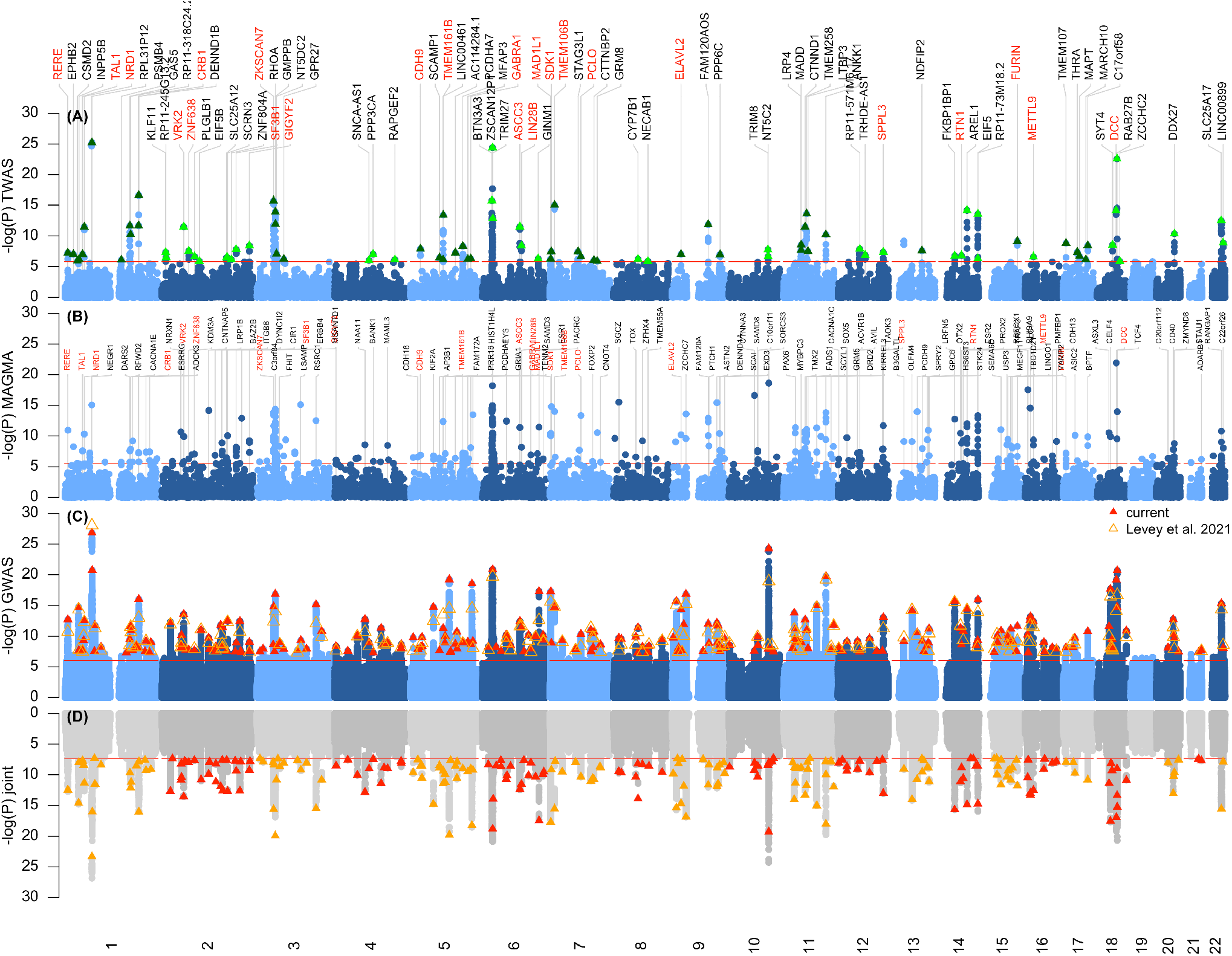
Miami plots… **(A)** TWAS, genes are represented by both gene expression and isoform expression (= features, represented by the dots). A red line indicates Bonferroni corrected genome-wide significance within analyses; p < 1.44×10-6; corresponding to Bonferroni correction of all the 34,646 features. The top transcript is labeled for each independent linkage disequilibrium block as previously described (PMID: 26395773). **(B)** Gene-based MAGMA analysis: A total of 268 significant genes were located in 93 of the 238 genomic risk loci identified using FUMA, while the remaining 143 significant genes were located outside these loci (table S7). The most significant gene within each of the 93 genomic risk loci are labeled in this plot. Gene names are labelled red if they overlap with the top gene within each independent locus of the TWAS in (A). **(C)** Manhattanplot of the meta-analysis GWAS of major depression (broad definition). Neighbouring index SNPs were considered independent when *r*^2^ *<* 0.1 in a sliding 3 Mb window. For each index SNP we defined the associated LD-region by recording the left and rightmost variant with *r*^2^ *<* 0.1. To define GWAS loci, a 50kb window was added on each side of the LD-region and overlapping LD-regions were combined into a single locus. Only a single SNP was kept from within the MHC region, due to extended linkage disequilibrium and a strong association signal of the MHC region (chr 6:25-35 Mb). **(D)** Joint p-values as implemented in GCTA-COJO ingreen, while original p-values are in shades of grey.

**Figure 2:**
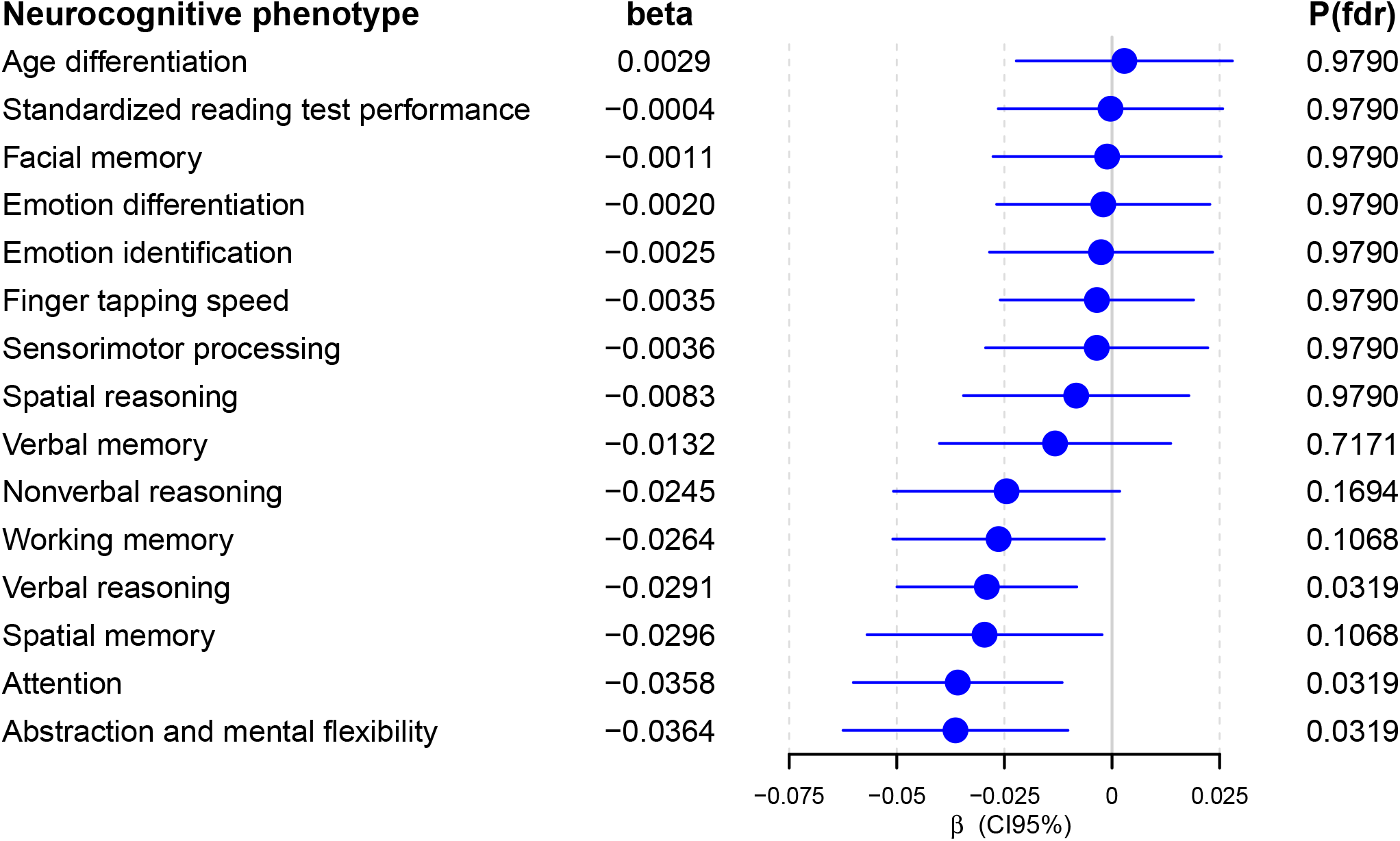
Association of MD-PGS with measures of cognitive abilities in the PNC cohort (N=4,973). Beta values (and 95% confidence intervals indicated as horizontal bars) from linear regression testing for the association of MD-PGS with the 15 neurocognitive measures listed on the y-axis. *β* values and FDR adjusted p-values are indicated.

### Genetic correlations and heritability

Analyzing the GWAS meta-analysis by LD score regression (LDsc)^35^ produced a genomic inflation factor (*λGC*) estimate of 1.89 with an intercept of 1.06 (SE = 0.01) and an attenuation ratio of 0.047 (SE = 0.012), indicating that 95% of the observed inflation of the test statistics (Supplementary Figure S1) is due to polygenic signal rather than population structure. The individual GWASs (iPSYCH2015, Howard et al. 2019^14^, Wray et al. 2018^12^, Levey et al. 2020^23^ and FinnGen) showed significant pairwise genetic correlations, ranging from *rG* = 0.77 to *rG* = 0.95 (Supplementary Table S4 and Figure S6), thus corroborating that the GWAS results are combined in a meta-analysis. We note that the SNP-heritability estimate^35^ for the iPSYCH cohort (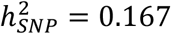, SE = 0.014, prevalence = 0.2) was significantly higher (around double) compared to the other cohorts (Supplementary Table S5 and Figure S7). This may reflect that the iPSYCH sample is more homogeneous and includes relatively severe cases who have been treated in hospitals.

We investigated genetic correlations with other phenotypes available at LD Hub^36^ and in-house, including published GWASs (N = 258) and GWAS results for UKB traits (N = 597). MD was significantly correlated (P < 2 × 10^−4^) with 364 phenotypes representing several domains and overall confirming previous observations^12-15,37-42^ (Supplementary Table S6A and Figure S8A). Among psychiatric disorders, MD showed significant correlation with e.g., ADHD (*r*_*G*_ = 0.56, SE = 0.022, P = 1 × 10^−135^), autism spectrum disorder (ASD) (*r*_*G*_ = 0.35, SE = 0.033, P = 6.5 × 10^−24^), BP (*r*_*G*_ = 0.31, SE = 0.033, P = 3 × 10^−18^), SZ (*r*_*G*_ = 0.33, SE = 0.021, P = 4 × 10^−53^), ANX (*r*_*G*_ = 0.79, SE = 0.017, P = 3.2 × 10^−193^), alcohol dependence (*r*_*G*_ = 0.65, SE = 0.097, P = 4.3 × 10^−9^) and cannabis use disorder (*r*_*G*_ = 0.44, SE = 0.036, P = 2.2 × 10^−31^). In UKB data, the three strongest genetic correlations were seen for “Seen doctor (GP) for nerves_ anxiety_ tension or depression”, “Mood swings” and “Miserableness” (*r*_*G*_ = 0.96, 0.71, 0.71, respectively; Supplementary Table S6B and Figure S8B).

To dissect the observed genetic overlap further, we used uni- and bivariate gaussian mixture modeling, as implemented in MiXeR^43^, to quantify the actual number of variants that (i) explain 90% of the SNP heritability of MD, and (ii) overlap between MD and MD-correlated phenotypes. Approximately 11,700 (SE = 310) common variants were estimated to confer liability to MD, suggesting that MD is the most polygenic of the major psychiatric disorders evaluated, showing number of risk variants ranging between 6-10,000^43,44^ (Figure 3, Supplementary Table S7). MiXeR considers all variants irrespective of the direction of correlation (i.e. both variants with same and opposite direction of effects, hereafter collectively referred to as “influencing” variants). Strikingly, the vast majority of variants conferring risk to the other psychiatric disorders investigated were found to influence MD (range 85-98%; Figure 3; Supplementary Table S7), most pronounced for ANX, SZ, BP and ADHD with 97-99%^44^ of their risk variants also influencing MD. The other investigated traits showed also substantial overlap with MD (range 85-98%). Notably, the fraction of concordant variants within the shared part varied considerably; lowest for educational attainment (42%) and highest for SUD (88%) and ANX (90%).

**Figure 3:**
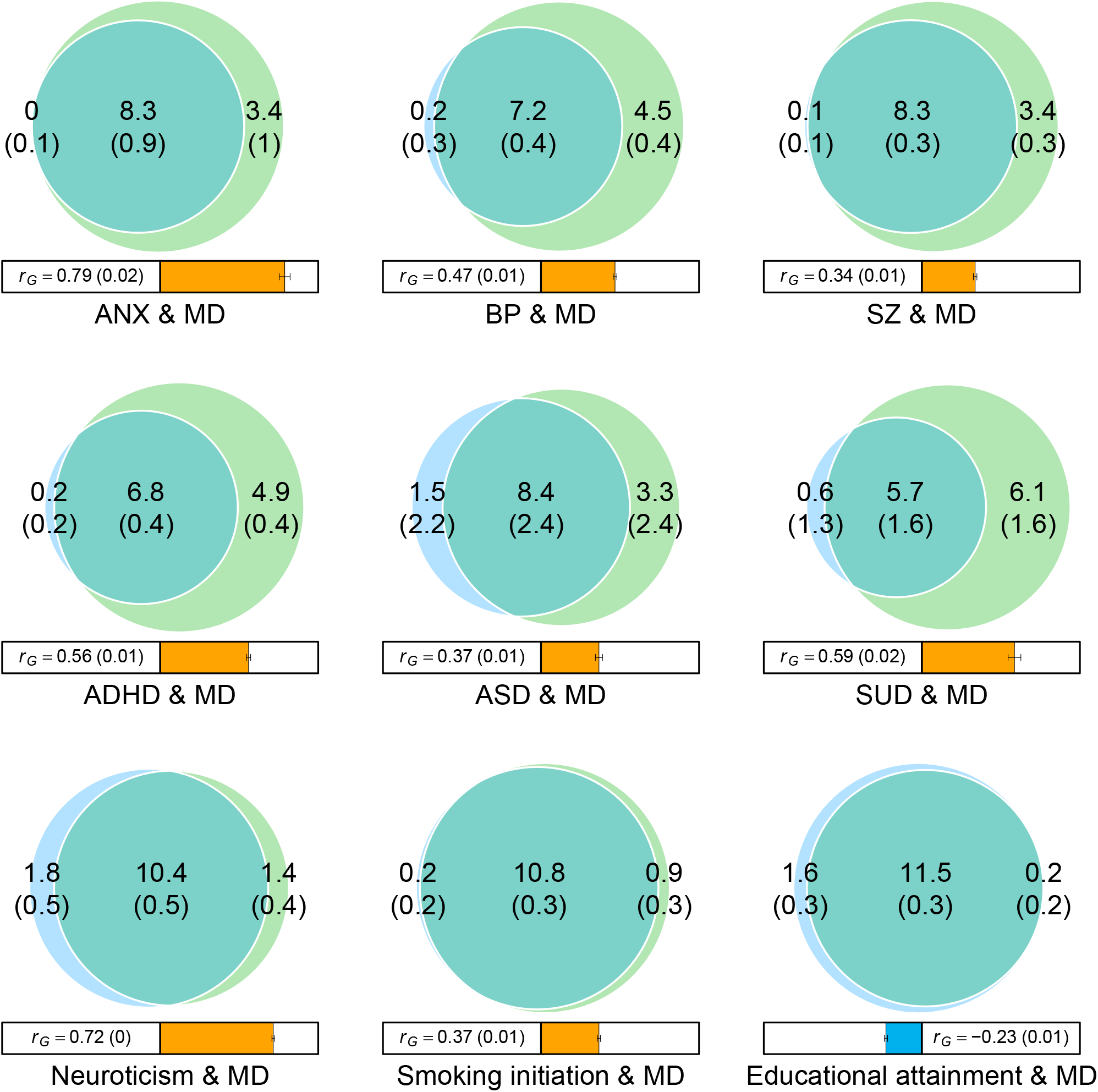
MiXeR bivariate analysis. Venn diagrams showing MiXeR results of the estimated number of variants shared between MD and psychiatric disorders (with significant genetic correlations with MD) and phenotypes representing other domains with high genetic correlation with MD. Circles represent loci unique to MD (green) and unique to the phenotype of interest (blue) and corresponding overlapping (shared) loci. The number of shared variants (and standard errors) are shown in thousands. The size of the circles reflects the polygenicity of each phenotype, with larger circles corresponding to greater polygenicity. The estimated genetic correlation (*r*_*g*_ and 95% confidence intervals) between MD and each phenotype from LDSC is shown below the corresponding Venn diagram, with an accompanying scale (−1 to +1) with blue and orange representing negative and positive genetic correlations, respectively. Bivariate results for the following comparisons are shown: BIPxMDD, SCZxMDD, ADHDxMDD, ASDxMDD, ALDCUDxMDD, neuroxMDD, SmoInixMDD, EAxMDD, (see also Supplemenatry Table XXX).

### Association with cognitive performance

Educational attainment was among the most prominent negatively correlated traits. To further evaluate the impact of MD genetic risk on cognition, we analyzed the association of MD polygenic scores (MD-PRS) with 15 cognitive measures in the Philadelphia Neurodevelopmental Cohort (PNC, N = 4,973)^45,46^. Cognitive performance was measured by the Computerized Neurocognitive Battery^47^, including 14 tests in 5 domains: executive-control, episodic memory, complex cognitive processing, social cognition, and sensorimotor speed. In addition, the Wide Range Achievement Test (WRAT-4)^48^ was used as a proxy measure for overall IQ^46^. MD-PRS was negatively associated with attention (β = −0.036, SE = 0.012, P = 0.032) and abstraction and mental flexibility (β = −0.036, SE = 0.013, P = 0.032) in the executive-control domain, and verbal reasoning (β = −0.029, SE = 0.011, P = 0.032) in the reasoning domain, while e.g. the WRAT-4 test did not show association (Figure 2, Supplementary Table S8). The results demonstrate, for the first time, that genetic MD risk is associated with attenuated functioning in specific cognitive domains. This is consistent with observations that individuals suffering from MD show lower performance in cognitive domains such as executive function, memory, language and attention^49-51^.

### Gene-wise and pathway analysis

A genome-wide gene-based association study conducted in Multi-Marker Analysis of GenoMic Annotation (MAGMA)^52^, mapped the GWAS SNPs to 17,840 protein coding genes and revealed 411 genes significantly associated with MD after Bonferroni correction for the number of genes tested (P < 2.8 × 10^−6^). A total of 314 significant genes were located in 141 of the 242 identified GWAS loci, while the remaining 97 significant genes were located outside these loci (Supplementary Table S9D). The most significant gene within each of the 141 genomic risk loci are labeled in Figure 1B.

To investigate enrichment of biological pathways we analyzed 8,664 gene-sets derived from GO Biological Process (N = 7,658) and GO Cellular Components (N = 1,006) ontology in the MSigDB database, identifying 479 significant gene-sets after correction for multiple testing (FDR < 0.05, Supplementary Table S10B), including several gene-sets that have not previously shown significant enrichment^14,23^. The majority of the top-ranking sets related to neuronal development and function including the five most significant gene-sets (P_adj_ < 10^−14^): GO_SYNAPSE, GO_NEURON_PART, GO_SYNAPSE_ORGANIZATION, GO_SYNAPSE_ASSEMBLY and GO_NEURON_DIFFERENTIATION.

### Transcriptome-wide association and GWAS-eQTL prioritisation

To identify and prioritize putative causal genes, we performed a transcriptome-wide association study (TWAS), imputing the genetically regulated gene expression using EpiXcan^53^ and models trained on expression data from the PsychENCODE Consortium^54,55^ for genes and isoforms detected in the dorsolateral prefrontal cortex (DLPFC). Among 34,646 transcripts (genes and isoforms) tested, we identified 2,541 transcripts at FDR < 0.05 and, after Bonferroni correction of all transcripts tested, 324 transcripts from 201 genes showing significant differential imputed gene expression in DLPFC between MD cases and controls (Supplementary Table S11). The Bonferroni significant transcripts were located in 88 independent^56^ regions. The top gene/isoform is labeled in Figure 1A and regional TWAS/GWAS Miami-plots for each of the 88 regions are shown in Figure S9. In 38 of the 88 regions, the top transcript was > 100 times more significant than the second-most associated gene/isoform in the region (Supplementary Table S11B), appointing those as plausible causal candidates.

To further prioritize likely causal genes and variants, we performed co-localization analyses integrating fine-mapped GWAS results, using the CAUSALdb pipeline (Methods), and eQTL data from a meta-analysis of three brain datasets^57^ applying a fixed-effect model. First, we adopted the Coloc method^58,59^, which revealed 13 genes with strong evidence for both GWAS-association, eQTL-association and co-localization (i.e., with a posterior probability of PPH4 > 0.8; Supplementary Table S12). The three top-ranked genes were: *FURIN, NEGR1* and *CKS2*. Secondly, we conducted the eQTL and GWAS CAusal Variants Identification in Associated Regions (eCAVIAR) approach^60^, in which both eQTL and GWAS were fine-mapped and the product of posterior probability (CLPP) was calculated, prioritizing variants with at least a single variant with CLPP >= 0.01. The eCAVIAR approach revealed five prioritized variants located in three genes: *FURIN, GPR27* and *TCTA* (Supplementary Table S13 and Figure S10).

### Tissue and cell type enrichment

We next tested whether the GWAS results were enriched with respect to the transcriptomic profiles of human tissues. At the specific tissue level, we found significant enrichment exclusively in brain tissues, including all the brain tissues analyzed (Supplementary Figure S11). Cell-type enrichment analyses revealed experiment-wide significant association (across all 13 datasets tested) of primarily neuronal cell-types, including dopaminergic and GABAergic neurons (Supplementary Figure S12 and S13). Interestingly, both GABAergic neurons and oligodendrocyte progenitor cells of the human prefrontal cortex were enriched already at prenatal stages.

To further evaluate cell-type enrichment, we intersected the GWAS results with two recent epigenomic maps of cell-specific open chromatin^61,62^ using an LD score partitioned heritability approach^63^. Again, we observed a clear contrast between the enrichments in the brain and non-brain tissue (Supplementary Figure S14). Consistent with our FUMA-based results and prior reports^12,14^, the strongest associations were measured for neuronal cell types, phenotypically manifested by severe synaptic loss and deficits in functional connectivity^64,65^. Conversely, to our knowledge, the reported association of MD with astrocytes and oligodendrocyte lineages have not yet been described by genetic data, albeit having support in behavioral and postmortem studies^66-68^.

### Polygenic architecture and co-morbidity rates for single-episode and recurrent depression

To dissect the polygenic architecture of single-episode and recurrent depression, we used dates of diagnosis in the Danish Psychiatric Central Research Register to group the 30,618 iPSYCH MD cases into 24,101 cases with single-episode and 6,517 with recurrent depression (see Methods and Supplementary Table S14 for details). We conducted three GWASs: 1) single-episode vs controls, 2) recurrent depression vs. controls and 3) recurrent vs. single-episode depression (excluding all controls). SNP-heritability estimates were similar for recurrent and single-episode depression and, for the case-only analysis, not significantly different from zero (Supplementary Table S5 and Figure S7). Likewise, the genetic correlations with other phenotypes showed similar patterns for single-episode and recurrent depression (Supplementary Figure S15 and Table S15).

We next investigated the polygenic load for MD in the recurrent and single-episode case groups, using a multivariate polygenic risk score (mvPRS) approach^15^ and the MD GWAS meta-analysis^14,23^ without iPSYCH samples for training. This analysis showed significant association of single-episode (*β* = 0.36, SE = 0.0081, P < 10^−7^) and recurrent (*β* = 0.43, SE = 0.014, P < 10^−7^) subgroups with PRS for MD (MD-PRS), but with a significantly larger effect size for recurrent than for single-episode depression (P = 4.8 × 10^−6^, Supplementary Table S16A and Figure S16A). This increased polygenic load among recurrent cases reinforces previous observations^11,12,69,70^.

To further dissect the genetic architecture of the two subgroups we investigated the PRS load for psychiatric disorders and traits showing strong genetic correlation with MD (Figure S16B-I, Table S16A) including ANX, BD, SZ, ASD, ADHD, SUD, substance use (SU) and neuroticism. The results showed an overall pattern of increased PRS among recurrent cases compared to single-episode cases (P = 0.00075), primarily driven by significantly different burdens of MD, ANX, BP and neuroticism PRS (Figure S16 and Table S16A).

To complement these analyses, we compared the rates of comorbid psychiatric disorders between single-episode and recurrent cases, using data from the Danish Psychiatric Central Research Register, and found highly significant increase in comorbidity among recurrent cases (Supplementary Table S17). This is consistent with well-established correlations between MD symptom severity, recurrence, chronicity and comorbidities (e.g.^10,71^) and, to our knowledge, it is the first quantification of differences in psychiatric comorbidity rates between single-episode and recurrent case groups. The most compelling differences were observed for ANX, BP and SZ.

Summarizing, compared to single-episode cases, recurrent cases showed significantly increased polygenic load of particularly MD-, BP- and neuroticism-PRS as well as increased rates of comorbid psychiatric disorders.

### Hazard rate ratio and absolute risk of recurrent depression

To investigate whether PRS can prospectively predict recurrence among individuals with first-onset depression, we performed Cox regression analyses estimating hazard rate ratio (HRR) and absolute risk of developing a second episode of depression over time among individuals having a first diagnosis of MD. This was done for groups of increasing polygenic load, in PRS deciles. We found that the HRR of developing a second-episode generally increased with increasing MD-PRS, most significantly for the 10^th^ MD-PRS decile compared to the first decile (*HRR*10 = 1.33, SE = 0.06, P = 2.5 × 10^−6^; Supplementary Figure S18A and Table S18). Similarly, the absolute risk of a second diagnosis of depression increased with time since first diagnosis for all MD-PRS deciles, with the trajectories for the 10^th^ decile reaching an absolute risk of 0.35 compared to 0.24 for the lowest decile (Supplementary Figure S18A).

Prompted by the results of the mvPRS analysis, we performed the same analysis for PRSs from other psychiatric disorders, but without yielding significant results (Figure S18, Table S18). The HRR for developing single-episode and recurrent MD in the general population can be found in Supplementary Figure S17.

### Dissecting the polygenic architecture of psychiatric comorbidity among MD cases

Individuals diagnosed with MD are at an increased risk of developing other mental disorders as documented in Supplementary Table S17 and a large body of studies (e.g.^9,10,72^), but little is known about the genetic constitution of those that develop these conditions. We examined the polygenic architecture of MD cases who have developed ANX, BP, SZ and SUD, using mvPRS analysis and PRSs from MD and eight psychiatric phenotypes genetically correlated with MD (ANX, BD, SZ, ASD, ADHD, SUD, SU and neuroticism; Supplementary Table S6A and Figure S8). This analysis showed that the subgroup of MD cases who had transitioned to BP, had a significantly increased PRS for three out of the eight psychiatric phenotypes tested compared to MD cases without a later BP diagnosis, including BP-PRS (P = 5.2 × 10^−17^), SZ-PRS (P = 3.2 × 10^−12^) and MD-PRS (P = 8.3 × 10^−5^) (Figure S19 (B), Table S19B). MD cases with a co-diagnosis of SZ showed increased PRS load for all eight psychiatric phenotypes except ASD (Figure S19 (C), Table S19C), most significantly for SZ-PRS (P = 2.9 × 10^−14^), BP-PRS (P = 5.5 × 10^−8^), substance use (SU) PRS (P = 2.4 × 10^−5^) and MD-PRS (P = 2.5 × 10^−5^). The most compelling results were revealed for SUD comorbidity, showing highly increased PRS loads for most of the psychiatric phenotypes (Figure S19 (D), Table S19D), most significantly for SU-PRS (P = 2.3 × 10^−84^), ADHD-PRS (P = 1.2 × 10^−31^) and SUD-PRS (P = 2.4 × 10^−30^).

Overall, we demonstrated that all three comorbid subgroups (MD-BP, MD-SZ, MD-SUD) have increased polygenic burdens of common risk variants for several psychiatric phenotypes compared to the non-comorbid MD (MD-only) case groups (overall mvPRS p-values: P_MD-ANX_ = 1.2 × 10^−21^, P_MD-BP_ = 3.7 × 10^−16^, P_MD-SZ_ = 3.2 × 10^−14^, P_MD-SUD_ = 1 × 10^−93^), revealing an overall PRS pattern that distinguishes the comorbid case groups from their non-comorbid counterparts. We also note that all three comorbid subgroups showed an increased load of MD-PRS compared to the MD-only case groups.

### Hazard rate ratio and absolute risk of comorbidity among MD cases

To assess the use of PRS in prediction of developing comorbid disorders, we performed Cox regression analysis on MD cases with and without additional diagnosis of ANX, BP, SZ and SUD, using the same eight PRSs as in the above-mentioned mvPRS analysis. In addition, we included four aggregate scores combining all PRSs into a single score weighted by their association with ANX, BP, SZ and SUD respectively (see Methods).

We found that the HRR for developing BP increased with increasing polygenic loads (Supplementary Figure S21 and Table S20B), most significantly for BP-PRS (HRR_10_ = 1.84, SE = 0.12, P = 9.3 × 10^−7^) and the aggregate score (BP-SUM-PRS; HRR_10_ = 2.32, SE = 0.14, P = 7.5 × 10^−10^) with the absolute risk trajectory reaching 0.11 for the 10^th^ BP-SUM-PRS decile compared to 0.04 for the 1^st^ decile as shown in Figure 4.

**Figure 4:**
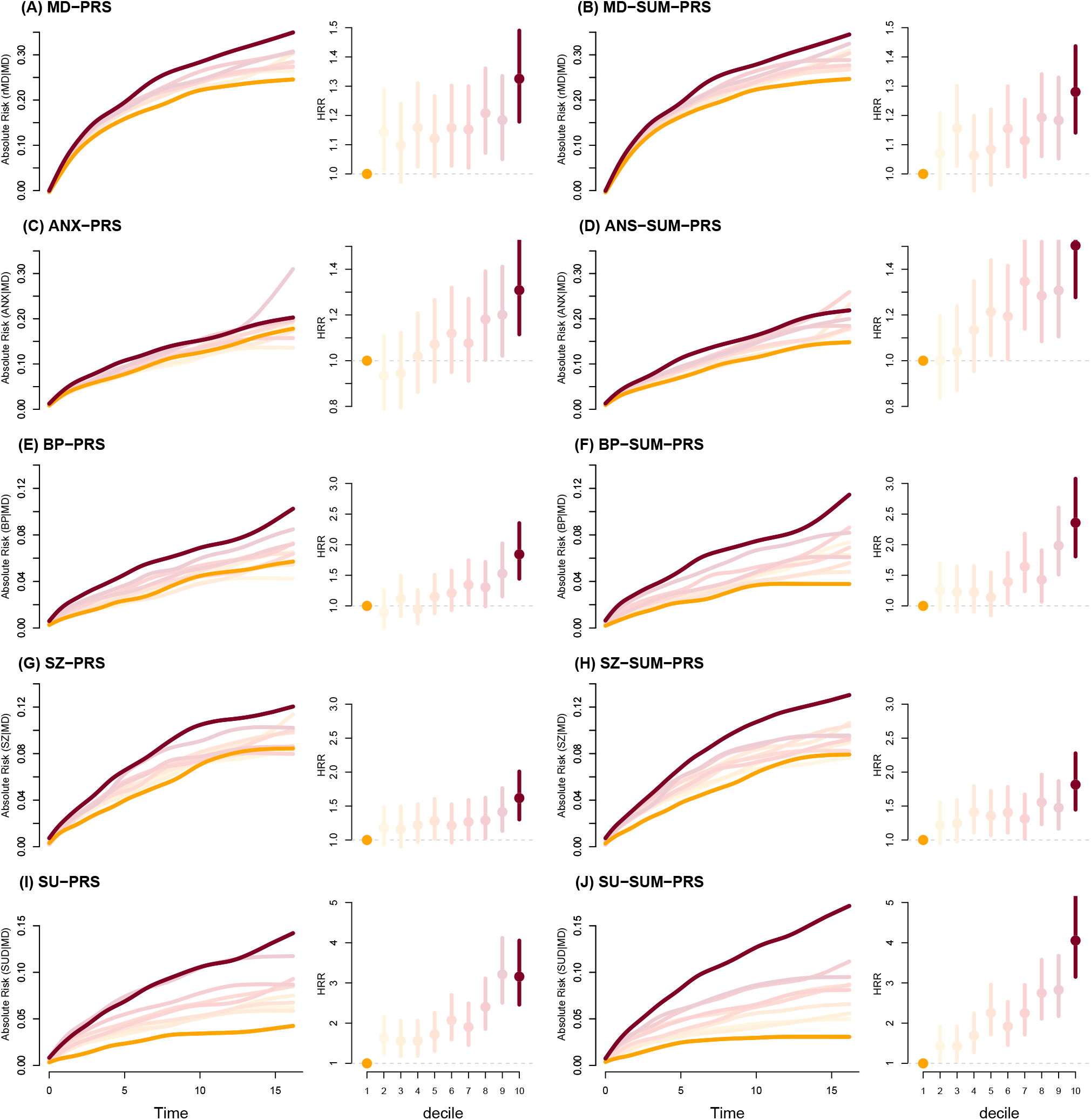
Absolute risk and HRR of developing a second episode of depression stratified by MD-PRS deciles (A) and MD-SUM-PRS (B). Absolute risk and HRR of transitioning into bipolar disorder stratified by BP-PRS deciles (C) and BP-SUM-PRS (D). Absolute risk and HRR of transitioning into schizophrenia stratified by SZ-PRS deciles (C) and SZ-SUM-PRS (D). Absolute risk and HRR of transitioning into substance-use-disorder stratified by SU-PRS deciles (C) and SUD-SUM-PRS (D). MD-SUM-PRS, BP-SUM-PRS, SZ-SUM-PRS and SUD-SUM-PRS were calculated by adding PRSs for multiple phenotypes weighted by log(*OR*) with the aim of optimising prediction (see methods for details). See supplementary figures S15, S17, S18 and S19 for results based on all PRSs analysed in the current study.

A similar pattern was observed for MD cases developing SZ (Supplementary Figure S22, Table S20C) where the highest HRR was seen for the aggregate SZ-SUM-PRS (HRR_10_ = 1.82, SE = 0.11, P = 1 × 10^−7^), with the absolute risk curve for the 10^th^ decile reaching 0.14, well separated from the 1^st^ decile trajectory approaching 0.067 (Figure 4).

The most striking results were obtained for SUD showing substantial differences in HHR and absolute risk estimates across PRS deciles for several phenotypes (Supplementary Figure S23, Table S20D). Again, the most significant prediction was obtained for the aggregate SUD-SUM-PRS (HRR_10_= 3.92, SE = 0.13, P = 3.6 × 10^−27^) with the absolute risk trajectory reaching 0.17 at 15 years after first episode for the 10^th^ decile in contrast to the first decile reaching a maximum risk of 0.03 (Figure 4), indicating that the absolute risk for developing SUD is almost 6 times higher for the most genetically burdened (top decile) MD case group compared to the least burdened group in the lowest decile at 15 years after first episode.

## Discussion

We performed an MD GWAS meta-analysis of more than 1.3 million individuals, identifying 251 independent risk variants in 243 genomic loci, of which 64 are novel. Among the novel loci, we highlight *GRIA1* and *GABRA1*, encoding a glutamate AMPA receptor subunit (GluA1) and a GABA receptor subunit (α1), respectively. Neither of these two loci have previously been identified in GWAS of MD or other mental disorders (Supplementary Table S2). The two genes showed also significant imputed differential expression in the DLPFC (at Bonferroni and FDR<0.05 significance, respectively; Supplementary Table S11) in our TWAS. We note that another GABA receptor subunit gene, *GABRB1*, was also significant (at FDR < 0.05) in the TWAS (Supplementary Table S11). *GABRB1*, although not significantly associated with MD in the current GWAS, has previously been implicated in schizoaffective disorder^73^ and SZ^74,75^. Our pathway analysis reinforced previous reports^12,14,23^ and extended the enrichment of glutamatergic and GABAergic synapses and functions, further indicating that glutamatergic and GABAergic dysfunctions are key etiologic components in MD. Along with the observed enrichment of GABAergic cell-types present already prenatally, this supports the accumulating multidisciplinary evidence that implicate excitatory/inhibitory (E/I) imbalance with MD^76^ as well as other psychiatric disorders^77-80^.

Our TWAS found that *GRIA1* expression in DLPFC was decreased in cases compared to controls. This is consistent with findings in preclinical pharmacological studies of mood disorders showing promising results of AMPA receptor potentiators^34^, as well as the convergent rapid and sustained increase in GluA1 and other synaptic proteins associated with most fast-acting antidepressants^33^. Thus, our results provide human genetic evidence that emphasizes positive modulation of the AMPA receptor as an interesting pharmacological approach.

In contrast to a reported decrease in *GABRA1* expression in the DLFPC of depressed suicide cases^81,82^, our TWAS pointed to an increased expression in MD cases. GABA_A_ receptors provide critical inhibitory control of the firing of glutamatergic excitatory neurons and they are the binding partner of several drugs in mood disorders and potential drug targets for other mental disorders^83-85^. Inhibitory synapse formation depends on the expression and selective postsynaptic clustering of GABA_A_ receptor subunits, which is a combinatorial process in which pentamers are assembled from nineteen different subunits (α1-6, β1-3, γ1-3, δ, ε, π, and θ). The subunit composition varies according to brain region and neuronal subtype, and confers unique physiological and pharmacological properties. Cortical expression of *GABRA1* increases drastically in the first few years of postnatal human development^86^, and in the adult brain, α1 contributes to the majority of both synaptic and extra-synaptic GABA_A_ receptors complexes. Antidepressant effects have been associated with pharmacological modulation of both synaptic^87^ and, more promising, extra-synaptic GABA_A_ receptors^88^. Although a recent study indicated that reversing the E/I imbalance alone only induces short-term antidepressant effects, but not long-term clinical response^89^, our findings support the use of pharmacological modulators of the α1βxδ subset of extra-synaptic GABA_A_ complexes.

The MiXeR bivariate Gaussian mixture modeling revealed compelling genetic overlap between MD and other traits when considering both concordant and discordant variants, suggesting that psychiatric disorders and correlated traits are substantially more intertwined than indicated by their genetic correlations. Perhaps the most surprising results were seen for educational attainment (EA), showing that 98% of the MD risk variants are also influencing EA and, vice-versa, 88% of EA variants are influencing MD. This almost complete overlap in influencing variants, with concordant (42% of the shared variants) and discordant effects, is consistent with the traits’ overall negative genetic correlation (r_G_ = −0.23) and refines the understanding of their polygenic architecture appreciably. Although generally less pronounced, a similar picture was observed for the other mental and behavioural traits examined. Furthermore, the notable overlap suggests that future fine-mapping efforts to pinpoint the causal variants could increase power by combining data from traits with overlapping influencing variants when accounting for the directional effects.

In the Danish iPSYCH cohort, we assessed PRS-based predictions for developing recurrent depression and psychiatric comorbidity over time and found significant differences in hazard rates and absolute risks between MD cases in the highest and lowest PRS deciles. The iPSYCH cohort includes relatively young individuals (mean age 23.37 years, SE = 6.9, at follow-up December 31^st^ 2016) and thus, a substantial number of the MD cases will develop recurrent MD and/or comorbid disorders later in life. This indicates that the observed risks, and likely also the differences across PRS deciles, will increase further over time. Moreover, the prediction could probably be improved by including other risk factors such as e.g. family history^90^ and clinical/phenotypic variables, as it has been shown in other complex disorders^91^. In clinical settings, a targeted effort could be envisaged that offer intensified monitoring for development of e.g. BP, SZ or ANX among MD cases with the highest PRS burden (or combined genetic and clinical risk) to obtain early diagnosis and initiate early treatment, which may have beneficial effects^92,93^. Similarly, identifying high risk for developing SUD could potentially be an informative point of attention for both the physician and the patient^94^. However, assessment of the clinical utility at the individual level of such potential applications is warranted^95^.

## Methods

### Samples

The iPSYCH2015 sample is a nation-wide population sample extracted from a baseline cohort consisting of all children born in Denmark between May 1, 1981, and December 31, 2008, who were alive and resided in Denmark on their one-year birthday, and who have a known mother^24,25^. Individuals diagnosed with one (or more) of six major psychiatric disorders (ADHD, ASD, MDD, BP, SZ, Anorexia) were identified via the Danish Psychiatric Central Research Register^26^, which includes data on all individuals treated in Denmark at psychiatric hospitals (from 1969 onward) as well as at outpatient psychiatric clinics (from 1995 onward). A random sample from the same birth cohort were chosen as control group. The iPSYCH2015 cohort consists of the initial case-cohort sample iPSYCH2012^25^ and the recent extension iPSYCH2015i^24^.

Individuals with a ICD10 F32-F33 diagnosis in 2016 or earlier were considered as MD cases and the random population-based sample excluding individuals with an MD diagnosis were used as controls. All individuals diagnosed with BP were excluded, except for the analyses involving co-occurrence of MD and BP diagnosis. MD cases were divided into two groups; 1) individuals diagnosed with a single episode of depression, and 2) those fulfilling the criteria for recurrent depression. The date of the first depression episode for each individual was defined as the start date of the first contact in the Danish Psychiatric Central Research Register with an ICD-10 code of F32-F33 diagnoses at an age of 10 years or older. When defining later depression episodes only contacts with the ICD-10 codes: F32, F33.0-F33.3, F33.8-F33.9 were considered. Individuals with the ICD-10 code F33.4 “Major depressive disorder, recurrent, in remission” were omitted because these contacts are unlikely to indicate a new episode of depression. Only ICD-10: F32, F33.0-F33.3, F33.8-F33.9 diagnosis at dates later than 60 days after the end date for all previous ICD10: F32-F33 diagnosis were considered as new episodes of depression, thus categorizing that particular individual as having recurrent depression.

In addition to depression phenotypes, individuals with the following diagnoses were recorded in the iPSYCH sample: BP (F30-F31), SZ (F20), SUD (F10-F19, excluding F1X.0 acute intoxication), ANX (F40-F43), ASD (F84.0, F84.1, F84.5, F84.8 or F84), ADHD (F90.0). In the iPSYCH2015 cohort^24^, we have added 11,710 MD cases and 18,410 controls to the iPSYCH2012 sample^12,25^, summarizing to a total of 34,095 cases and 45,393 controls prior to relatedness pruning and removal of ancestry outliers (Supplementary Table S14). A total of 30,618 cases and 38,200 controls were retained after relatedness pruning and removal of ancestry outliers (Supplementary Table S14). Of these, 6,517 had recurrent episodes of depression (see definition in Methods), while 24,101 were classified as single-episode depression cases (Supplementary Table S14).

The number of cases with an additional psychiatric diagnosis (BP, SZ, ANX, ASD, ADHD, SUD) are shown in Supplementary Table S17. Individuals with a diagnosis of BP were excluded from all subsequent analyses except for analyses involving BP.

Whether recurrent depression was associated with increased comorbidity compared to single-episode depression was tested using logistic regression for each additional diagnosis while adjusting for age. Individuals were grouped in five-year age bins in order to construct dummy variables for the age adjustment (Supplementary Table S17).

All analyses of the iPSYCH sample were performed at the secured national GenomeDK high-performance computing cluster in Denmark. The study was approved by the Scientific Ethics Committee in the Central Denmark Region and the Danish Data Protection Agency. The study was approved by the appropriate local scientific ethics committees and IRBs.

### Genotyping, QC and imputation

iPSYCH2015 samples were linked via the unique national personal identification number to the Danish Neonatal Screening Biobank (DNSB) at Statens Serum Institute (SSI), where DNA was extracted from Guthrie cards, and whole-genome amplification was performed in triplicate, as described previously^96,97^.

Genotyping in iPSYCH2012 was performed using the PsychArray V1.0 (Illumina, San Diego, California), while genotyping of iPSYCH2015i was done using the Global Screening Array v2 (Illumina, San Diego, California). Since the two samples were genotyped on different platforms, they were QCed and imputed separately.

Genotyping of the iPSYCH2012 sample was performed at the Broad Institute of Harvard and MIT (Cambridge, MA, USA) with PsychChip arrays from Illumina according to the manufacturer’s instructions. Genotype calling of markers with MAF ≥ 0.01 was performed by merging call sets from GenCall^98^ and Birdseed^99^, and less frequent variants (MAF < 0.01) were called with zCall^100^. Genotyping and data processing were carried out in 23 waves. Genotyping of the iPSYCH2015i sample was performed at Statens Serum Institut (SSI, Copenhagen, Denmark) using the Global Screening Array v2 with a Multi disease drop in (Illumina, San Diego, California) according to the manufacturer’s instructions. Genotype calling of markers was performed using GenTrain V3. Preimputation quality control was performed using the Ricopili^101^ pipeline with the specified parameters in the following order. Initially SNPs with a call rate < 0.95 were removed, and subsequently all individuals with a call rate in cases or controls of < 0.95 or an autosomal heterozygosity deviation *F*_*HET*_ outside the interval [-0.2;0.2] were removed. Individuals where stated sex was not consistent with sex derived from genotypes were flagged. Subsequently QC was conducted at the marker level, keeping markers with call rate ≥ 0.98, missing difference ≤ 0.02 between cases and controls, with MAF ≥ 0.01, Hardy-Weinberg equilibrium (HWE) in controls p value ≥ 1 × 10^−06^ and Hardy-Weinberg equilibrium (HWE) in cases p value ≥ 1 × 10^−10^ (See https://sites.google.com/a/broadinstitute.org/ricopili/preimputation-qc for further details). The effects of three batch variables on marker genotypes were tested in iPSYCH2012 (ArrayPlate.ID, PreProc.Plate and wave) and iPSYCH2015i separately (Array.Batch, ArrayPlate.ID and PreProc.Plate). This was done using relatedness-pruned dataset with ancestry outliers removed in order to avoid removal of markers where batch effects were caused by population structure or cryptic relatedness rather than genotype artefacts.

Pairwise relatedness coefficients 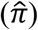 were estimated with plink using a LD pruned and MAF filtered set of SNPs (snps-only, window size = 5,000, step size = 300, *r*^2^ < 0.05, MAF > 0.05). Principal Component Analysis was conducted using the same set of LD pruned and MAF filtered SNPs, with random removal of one member of each pair with a relatedness coefficient 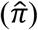 higher than 0.2. Eigenvectors were inferred using EIGENSOFT version 6.1.4 on the relatedness-pruned set of individuals, and subsequently projecting all individuals onto those eigenvectors based on their genotypes. Individuals with all four grandparents born in Denmark were used as a reference for constructing a 3-dimensional ellipsoid using principal components 1, 2 and 3 with a radius of 5 standard deviations from the mean. Individuals located outside this ellipsoid were removed prior to the testing for batch effects. Each genotyped marker was tested for association with each batch versus the remaining batches pooled. This was done for the following batch-variables: ArrayPlate.ID, PreProc.Plate and wave for iPSYCH2012 and Array.Batch, ArrayPlate.ID and PreProc.Plate for iPSYCH2015i. Batch-association testing was conducted using plink v1.90b4.

The exclusion of SNPs strongly associated with any of the batch-variables were based on their minimum P-value across all associations per variable. The cut-off for the wave and Array.Batch was min(p) < 2 × 10^−10^ and for PreProc.Plate and ArrayPlate.ID min(p) < 2 × 10^−12^, based on a Bonferroni correction for the number of markers tested and the number of associations done per batch-variable trying to incorporate the degree of nestedness of these. After removing SNPs falling for any of the above cut-off the remaining distribution was evaluated using QQ-plots. The expected minimum p-distribution was calculated using the inverse cumulative distribution of N independent distributions as suggested in supplementary of Schork et al.^102^, N being the number batch-variable values. Reviewing these QQ-plots it is evident that despite filtering by p-value some signal from the batch-variable remains in the dataset.

iPSYCH2012 and iPSYCH2015i were imputed using the ricopili pipeline^101^. Prephasing was done using Eagle v2.3.5^103^ and the subsequent imputation was conducted using Minimac3^104^, using the downloadable version of the Haplotype Reference Consortium (HRC) (accession number: EGAD00001002729)^105^ as reference.

### Relatedness pruning and removal of ancestry outliers

Best guest genotypes from iPSYCH2012 and iPSYCH2015i were merged, filtered and LD-pruned down to a set of roughly 30K markers, with imputation INFO score > 0.8, *r*^2^ < 0.075, located outside regions of long-range LD as defined by Price et al.^106^, minor allele frequency > 0.05 and no deviation from Hardy-Weinberg proportions (P > 1 × 10^−4^). Relatedness coefficients, based on “identity-by-state”, were estimated using plink v1.9, in order to identify related (and duplicated samples), with 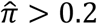 and one individual of each such pair was subsequently excluded at random. PCA was carried out using the same set of filtered and LD-pruned SNPs as implemented in the ricopili pipeline^101^. A subsample of European ancestry was selected as an ellipsoid in the space of PC1-3 centred and scaled using the mean and 8 standard deviation of the subsample whose grandparents were all known to be born in Denmark.

### FinnGen

The FinnGen (https://www.finngen.fi/en) study combines genotype data with longitudinal health register data of Finland, including the causes of death, inpatient, outpatient, and drug reimbursement registers.

A GWAS for depression was conducted in FinnGen release 6 samples containing 28098 cases (ICD-10/9: F32 or F33) and 228817 controls without manic episodes (ICD10 F30), bipolar affective disorder (ICD-10: F31, ICD-9: 296[2-7], ICD8: 296), persistent mood disorders (ICD10: F34) or other or unspecified mood disorder (ICD10: F38,F39, ICD-8: 29699). Variants with an imputation info score < 0.6 were excluded. GWAS was run with SAIGE logistic mixed model (version 0.39.1) using age, sex, 10 PCs and genotyping batch as covariates.

A GWAS for anxiety disorders was conducted in the FinnGen release 6 sample consisting of 7,671 cases with a generalized anxiety disorder (ICD-10: F41.1, ICD-9: 3000C, ICD-8: 300,00), panic disorder (ICD-10: F41.0; ICD-9: 3000B, 3002B) or phobic anxiety (ICD-10: F40, ICD-9: 3002C, 3002D, 3002X) diagnoses and 161,438 controls without a history of any psychiatric diagnoses (ICD-10: F00-F99, ICD-9: 290-319, ICD-8: 290-315). With ICD-9 codes the first three digits signify numerical value (e.g., the code **300**2B is included in the code range 290-319). Cases with psychotic disorders (ICD-10: F2; ICD-9: 295, 297, 298; ICD-8: 295, 297, 298), autism spectrum disorders (ICD-10: F84.0, F84.1, F84.5; ICD-9: 2990; ICD-8: 299,99) or intellectual disability (ICD-10: F7, ICD-9: 317-319; ICD-8: 311-315) were excluded. Additionally, the age range of controls was adjusted to match to that of cases.

Subjects were genotyped with Illumina and Affymetrix arrays (Illumina Inc., San Diego, and Thermo Fisher Scientific, Santa Clara, CA, USA) as described (https://www.finngen.fi/en/researchers/genotyping). Genotyping and imputation with the Finnish population-specific SISu v3 reference panel were conducted, as described (https://www.protocols.io/view/genotype-imputation-workflow-v3-0-xbgfijw). SNPs were pruned for minor allele frequency (MAF) ≥ 0.01 and imputation info score > 0.8. GWAS was performed using the Scalable and Accurate Implementation of GEneralized mixed model (SAIGE) v0.20^107^ with a kinship matrix as a random effect and age, sex, the first 10 principal components (PCs), and genotyping batch as fixed effects.

### GWAS analyses

Genome-wide-association analyses within iPSYCH were conducted using the RICOPILI pipline^101^, applying an additive logistic regression model using dosages of the imputed genotypes. Analyses were adjusted for PC 1-10 from a PCA using the remaining subsample after removal of ancestry outliers and pruning for relatedness. In addition, MD cases with a diagnosis of bipolar disorder were excluded.

Summary statistics from iPSYCH2015 and the following external samples were included in a fixed-effects variance-weighted meta-analysis in METAL^27^: 1) 230,118 broadly defined MD cases and 545,339 controls from Howard et al. 2019^14^, including self-reported MD of 23andMe^22^ and the broadly defined MD phenotype of UKB; 2) 35,077 narrowly defined MD cases and 95,406 controls from Wray et al. 2018^12^, excluding 23andMe^22^; 3) depression phenotypes from the Million Veteran Program (MVP)^23^ based on ICD codes derived from electronic health records (83,810 cases and 166,405 controls); 4) 28,098 cases with ICD-10 F32 and/or F33 diagnoses and 228,817 controls from FinnGen.

For comparison, we also conducted a GWAS and several downstream analyses using a narrower definition of MD that excluded self-reported MD of 23andMe^12,22^ and the broadly defined MD phenotype of UKB^14,21^ (Supplementary Figure S5, Table S3). The results were very similar to the primary analyses without noteworthy differences.

### SNP-heritability, genetic correlations and overlap with other phenotypes

We estimated SNP-heritability 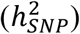 for iPSYCH2015 cases with MD, single episode and recurrent depression and for the external GWAS summary statistics outlined above using LD score regression^35^. Genetic correlations within iPSYCH and between iPSYCH and external GWAS summary statistics was estimated using LD score regression^35^.

In addition, the genetic correlations of MD meta-analysis, single-episode and recurrent depression with other phenotypes were, evaluated using LD score regression^108^ at the LD Hub^36^ website. We used all available phenotypes on LD Hub, but we performed analyses for the UKB traits (N = 597) and the remaining individual phenotypes (N = 258) separately. Levels of experiment-wide significance (Bonferroni correction for number of tests applied) were also established separately within the two groups, i.e. in the UKB traits (P < 8.38 × 10-5) and the remaining individual phenotypes (p < 0.00019), respectively. LD Hub traits were supplemented with LD score regression^35^ analyses performed locally using updated or in-house GWAS summary statistics for traits not available at LD-hub. These include ADHD, ASD^15^, cannabis use disorder^37^, cannabis use^109^, alcohol dependence^110^, drinks per week, smoking ever^111^, age of initiation as a regular smoker, current versus a former smoker (smoking cessation), number of cigarettes per day, smoking initiation^112^

We applied MiXeR^43^ on our MD GWAS summary statistics and a selection of additional traits (Supplementary Table S7 and Figure 3) to estimate (i) the number of variants explaining 90% of the SNP heritability of each trait and (ii) the genetic overlap between MD and each trait. MiXeR analysis were conducted with default settings (https://github.com/precimed/mixer) in a two-step process: 1) a univariate model for each trait to produce estimates of the proportion of variants with non-zero additive genetic effect on the trait (i.e. “polygenicity”) and the variance of effect sizes of these non-zero variants (i.e. “discoverability’”). 2) the variance estimates obtained in the univariate analysis were applied in the bivariate model (i.e. MD vs. each of the additional traits) to obtain four estimates representing (i) zero-effect SNPs in both traits; (ii) SNPs with a specific non-zero effect on one of the two traits; and (iii) SNPs with a non-zero effect on both traits. Estimates of polygenic overlap and genetic correlation between pairs of traits were obtained by combining these four parameters.

### Conditional analysis and finemapping

We identified potential independent genome-wide significant lead variants for each of the broadly defined genome-wide-significant locus identified pipeline from our GWAS meta-analysis results^101^: Neighboring index SNPs were considered independent when *r*^2^ < 0.1 in a sliding 3 Mb window. For each index SNP we defined the associated LD-region by recording the left and rightmost variant with *r*^2^ < 0.1. To define GWAS loci, a 50kb window was added on each side of the LD-region and overlapping LD-regions were combined into a single locus. Only a single SNP was kept from within the MHC region, due to extended linkage disequilibrium and a strong association signal of the MHC region (chr 6:25-35 Mb).

In order to identify additional independent index variants, we performed a stepwise model selection procedure to select independently associated SNPs also implemented in GCTA-COJO^28,29^.

We assigned posterior probabilities (PP) of being causal to SNPs and constructed credible sets of SNPs that cumulatively capture 95% of the regional posterior probability^113^ using PAINTOR^114-116^, CAVIARBF and FINEMAP^117^, using the CUASALdb pipeline (https://github.com/mulinlab/CAUSALdb-finemapping-pip). We applied a conservative approach and assumed one causal variant for each locus.

Co-localization analyses were performed in order to evaluate the extent of overlap between eQTL and GWAS signatures in MD and to identify putative causal genes from GWAS associations. Considering only the 95% credible SNPs from the fine-mapped MD-meta-analysis, we integrated GWAS results and eQTL data from a previous meta-analysis integrating signals among three brain datasets^57^ applying a fixed-effect model. The eQTL data originates from eQTL meta-analysis on RNA-sequenced gene expression data from the dorsolateral prefrontal cortex from PsychENCODE^54^ and ROSMAP^118^, and from 13 brain regions from GTEx^119^.

Using the Coloc method^58,59^, we extraced eQTL signals of genes within 200 kb distance to significant GWAS variants (P < 5 × 10^−8^) using effect sizes (*β*-values) and standard errors from eQTL and GWAS as input. Four hierarchical hypotheses^59^ were tested: H0, no association; H1, GWAS association only; H2, eQTL association only; H3, both GWAS and eQTL association but no co-localization; H4, both GWAS and eQTL association and co-localization, considering PPH4 > 0.8 as strong evidence from both GWAS, eQTL and co-localization. In addition, we conducted the eQTL and GWAS CAusal Variants Identification in Associated Regions (eCAVIAR) approach^60^, in which, both eQTL and GWAS were fine-mapped, and the product of posterior probability (CLPP) was calculated, prioritizing genes with at least a single variant with CLPP >= 0.01.

### Gene-wise and pathway analysis

We used a number of different approaches and data including those available via the FUMA v1.3.6a^52^ website (http://fuma.ctglab.nl) for downstream annotation and functional characterization of significant loci.

We performed a genome-wide gene-based association study using the Multi-Marker Analysis of GenoMic Annotation (MAGMA) tool, as implemented in FUMA version 1.3.6a^52^. Using this we mapped SNPs from the MD GWAS meta-analysis to 17,840 protein coding genes, and performed Bonferroni correction for the total number of protein-coding genes (P < 2.8 × 10^−6^, Supplementary Table S9D).

Protein coding genes were mapped if they were located with a distance of 10Kb up- or downstream index variants or if a credible variant was annotated to the gene based on eQTL data or chromatin interaction data from human brain (data sets used in the mapping can be found in the Supplementary Note). No additional variant filtering by functional annotation was applied in the eQTL and chromatin interaction mapping. This analysis identified 411 MD risk genes, which were used in a gene-set enrichment analysis within the GENE2FUNC module of FUMA, where we analyzed 8,664 gene-sets derived from GO Biological Process (N = 7,658; 393 gene-sets) and GO Cellular Components (N = 1,006; 86 gene-sets) ontology in the MSigDB database.

### Tissue and cell type enrichment

We also used FUMA to perform tissue expression analyses on data available through their website, by tested whether the identified genetic associations for both the primary and narrow MD phenotype definitions were enriched regarding transcriptome profiles of human tissues using summary statistics based on all SNPs. Finally, we used FUMA to perform cell-type enrichment analyses^52^ based the MD GWAS summary statistics. We use MAGMA gene-property analysis to test cell type specificity of phenotype with GWAS summary statistics. MAGMA gene-property analysis with scRNA-seq: The gene-property analysis aims to test relationships between cell specific gene expression profiles and disease-gene associations. In all above-mentioned analyses implemented in FUMA default settings were applied. To further evaluate whether the genomic loci implicated in MD are enriched in any particular cell type, we intersected common MD risk variants with two recent epigenomic maps of cell-specific open chromatin^61,62^ using LD score partitioned heritability approach^63^(Supplementary Figure S14).

### Transcriptome-wide association study

In addition, we performed a transcriptome-wide association study (TWAS), imputing the genetically regulated gene expression using EpiXcan^53^ and using models trained on PsychENCODE Consortium (PEC)^54,55^ expression data for genes and transcripts detected in the dorsolateral prefrontal cortex (DLPFC), with the aim of identifying and prioritizing putative causal loci for the broad MD phenotype definition. A total of 34,646 genes/transcripts were tested (transcripts with prediction performance R^2^ > 0.01 and prediction performance q value < 0.05 with the Benjamini-Hochberg method were retained), and applying a significance threshold of P < 1.44 × 10^−6^ (corresponding to Bonferroni correction of all genes and isoforms tested; Figure 1A, S9 and Table S11), using information on approximately independent linkage disequilibrium blocks in human populations^56^ to identify independent genomic regions with genes/transcripts showing significant differential gene expression between MD cases and controls.

### Polygenic risk scores

Using available summary statistics form published GWAS as training datasets, we calculated polygenic risk scores (PRS) for individuals in the iPSYCH2015 sample using LDpred2^120^. Summary statistics were filtered for an imputation info score INFO ≥ 0.9 if available. When using external summary stats not processed using RICOPILI or imputed using different imputation references, we excluded all ambiguous markers to avoid potential strand conflicts. To improve performance of the scores and avoid including artefacts from batch effects, we restricted the summary stats to include only SNPs known to be present in both the iPSYCH2012 and iPSYCH2015i data at a reasonable quality (info score 0.6 and MAF 0.01). This step also checked for allele flips.

To derive a PRS for MD within the iPSYCH2015 sample we used the MD meta-analysis^12,14,22,23^ excluding all iPSYCH samples as training. We also calculated PRS based on GWAS summary statistics from BP^121^, SZ^122^, ADHD^13^ and ASD^15^. The PRSs for ADHD and ASD were based on a combination of external GWAS sumstats and internal training. A meta-analysis of two GWAS for anxiety, one based on self-report of physician diagnosis of ANX in the MVP^123^ and the other being core anxiety in the FinnGen cohort (see FinnGen description above), served as weights for generating an ANX-PRS summarizing genetic risk of anxiety. To derive a PRS for neuroticism we used GWAS summary stats of a weighted neuroticism sum-score, constructed by adding up ten individual item responses (Fed-up, Guilt, Irr, Miss, Mood, Tense, Nerv feel, Suf Nerv, worry emb and worry) by Nagel et al.^124^. In addition, we calculated PRSs using the following GWAS summary stats for traits related to substance-use and substance-use-disorder as training datasets. These include *cannabis use disorder*^37^(excluding iPSYCH samples, i.e., training only based on data from PGC and data from decode genetics), *cannabis use*^109^, *alcohol dependence*^110^, *drinks per week, smoking ever*^111^, *age of initiation* as a regular smoker, current versus a former smoker (*smoking cessation*), *number of cigarettes per day, smoking initiation*^112^. All summary statistics were combined in a pseudo-meta-analysis, subsequently used for generating a SU-PRS summarizing genetic liability for substance use while summary stats for *alcohol dependence*^110^ and *cannabis use disorder*^37^ were pseudo-meta-analyzed to generate a SUD-PRS summarizing genetic liability of substance use disorder.

With the aim at improving the PRS prediction we attempted to exploit the genetic overlap of MD with other phenotypes. We therefore combined all of the above PRSs into a single score as a weighted sum^15^. We chose here to use the log(OR) for the logistic regression of sub-phenotype of interest on each score as a continuous factor as a measure of ‘importance’ in context of the sub-phenotype of interest (either recurrent vs. single-episode or MD with additional diagnosis of BP, SZ or SUD) and adjusting for PCs. We added a PRS weighted by its log(OR) and the standardized one PRS at the time, starting with the phenotype with highest abs(log(OR)) and ending with the lowest. This way we ended up with a sequence of scores starting with *S*_*0*_ and continuing with:

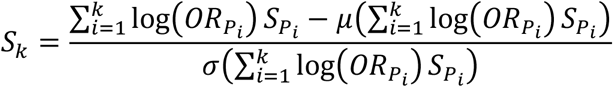

Where 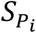 is the score for phenotype *P*_*i*_, *OR*_*Pi*_ the odds ratio from the logistic regression of sub-phenotype of interest on *S*_*Pi*_ as a continuous factor adjusting for PCs, and *μ* is the mean and *σ* the standard deviation.

To examine potential polygenic heterogeneity across MD sub-phenotypes, we investigated how PRS trained on the different phenotypes described above were distributed across MD sub-phenotypes in iPSYCH using a multivariate PRS approach. The method is described in detail in Grove et al.^15^, and is a regression of multiple outcome variables, and in principle a linear regression for each PRS on the MD sub-phenotypes allowing for comparisons on the average PRS across sub-phenotypes for PRS from a number of phenotypes while accounting for the inherent correlation between scores and adjusting for necessary covariates. MD-BP cases were excluded from the analyses not involving BP. In addition, we applied a Cox proportional hazard model to estimate hazard rate ratios and absolute risk of developing 1) a second episode of MD, 2) BP, 3) SZ and 4) SUD, among individuals already being diagnosed with their first episode of MD. Analyses were stratified by PRS deciles of the phenotypes described above and using functions from the R-packages survival (https://CRAN.R-project.org/package=survival)^125,126^ and survminer (https://CRAN.R-project.org/package=survminer), while correcting for batch (iPSYCH2012 and iPSYCH2015i) and PCs using an in-house pipeline.

### MD-PRS in the Philadelphia Neurodevelopmental Cohort (PNC)

MD-PRS were calculated for 4,973 individuals of European ancestry from the Philadelphia Neurodevelopmental Cohort (PNC)^45,46^ using the primary MD GWAS meta-analysis of the current study as training. Genotypes from the first PNC release (dbGaP phs000607.v1.p1) and neurocognitive phenotypes from the third release (dbGaP phs000607.v3.p2) were utilized. Individuals whose genotypically inferred and phenotypically reported sex did not match, those who did not meet the identity by descent (IBD) filter 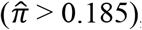, those who did not meet the individual-level missingness filter of 0.05, and those with heterozygosity rates +/- three standard deviations from the mean were removed. SNPs that failed to meet the Hardy-Weinberg proportions (HWP), minor allele frequency (MAF), and SNP-level missingness filters of 0.00001, 0.01, and 0.05, respectively, were also removed. Genotype imputation was performed using the reference panel HRC r1.1 2016 on the Michigan Imputation Server (https://imputationserver.sph.umich.edu/index.html#!), selecting the “Mixed population” option. Imputed SNPs with an imputation R^2^ of < 0.03 and those who failed to meet the missingness, MAF, and HWP thresholds stated above were removed. The first two Multi-Dimensional Scaling (MDS) dimensions were plotted to identify and remove outlier genotypes. The GemTools package in R (http://www.compgen.pitt.edu/GemTools/GEM%20Documentation.pdf) and Ward’s hierarchical clustering methods were used to identify individuals of European ancestry.

PRS-CS^127^ was used to assign per-allele posterior SNP effect sizes to MD GWAS summary statistics. A European LD reference panel provided by the developers of PRS-CS was utilized (https://github.com/getian107/PRScs), which draws from the 1000 Genomes Project data. The following PRS-CS default settings were used: parameter a in the γ-γ prior = 1, parameter b in the γ-γ prior = 0.5, MCMC iterations = 1,000, number of burn-in iterations = 500, and thinning of the Markov chain factor = 5. The global shrinkage parameter phi was set using a fully Bayesian determination method. Individual-level MD-PRS were calculated using Plink v2.0^128^. The associations between scaled (mean = 0, SD = 1) MD-PRS and 15 scaled neurocognitive phenotypes in the PNC were assessed using linear regression. Covariates included in the analysis were age at neurocognitive testing, age squared, the first 10 MDS dimensions, sex, and genotyping batch. Adjusted R^2^ was used to report the total variance explained by MD-PRS and model covariates for the 15 tested neurocognitive phenotypes. Additionally, a variance partitioning tool (https://github.com/GabrielHoffman/misc_vp/blob/master/calcVarPart.R) was used to determine the variance explained by MD-PRS and each covariate individually. FDR-adjusted p-values were reported.

## Data Availability

Summary statistics from this publication are available at http://ipsych.au.dk/downloads/. All relevant iPSYCH data are available from the authors after approval by the iPSYCH Data Access Committee and can only be accessed on the secured Danish server (GenomeDK, https://genome.au.dk) as the data are protected by Danish legislation. For data access please contact: Anders D. Børglum. Data used for brain transcriptome model generation are available from PsychENCODE (overview of available data sets at http://resource.psychencode.org/); genotypes are controlled data and access instructions are provided at https://www.synapse.org/#!Synapse:syn4921369/wiki/477467. Note that some datasets have been indirectly accessed at the respective analytical websites (through the FUMA website). Please refer to these websites (e.g., for FUMA https://fuma.ctglab.nl/links and https://fuma.ctglab.nl/tutorial#datasets) for availability of datasets used in the respective follow-up analyses / lookups.

## Code availability

Please refer to individual sections of the methods above for published code (e.g., Ricopili, LDpred2, MiXeR). Because the in-house scripts used for data processing and analysis of the iPSYCH data on the GenomeDK HPC infrastructure is highly dependent on that context, they can only be obtained from the authors upon request. This way we can ensure the proper context is explained in dialogue with the interested parties.

## Inclusion and ethics

The research has included local researchers throughout the research process, including study design, study implementation, data ownership, intellectual property and authorship of publications. Roles and responsibilities were agreed amongst collaborators ahead of the research.

This research does not result in stigmatization, incrimination, discrimination or any personal risk of the participants. Local and regional research relevant to the study have been considered in the listed citations.

## Acknowledgements

The iPSYCH team was supported by grants from the Lundbeck Foundation (R102-A9118, R155-2014-1724, and R248-2017-2003), the EU H2020 Program (Grant No. 667302, “CoCA”), NIH/NIMH (1U01MH109514-01 and 1R01MH124851-01 to ADB) and the Universities and University Hospitals of Aarhus and Copenhagen. The Danish National Biobank resource was supported by the Novo Nordisk Foundation. High-performance computer capacity for handling and statistical analysis of iPSYCH data on the GenomeDK HPC facility was provided by the Center for Genomics and Personalized Medicine and the Centre for Integrative Sequencing, iSEQ, Aarhus University, Denmark (grant to ADB).

We would like to thank the research participants and employees of 23andMe, Inc. for making this work possible.

The team at the Center for Disease Neurogenomics at Icahn School of Medicine at Mount Sinai was supported by the NIH (K08MH122911 to GV; T32MH087004 to KT; R01MH125246, R01AG067025, U01MH116442 and R01MH109677 to PR).

## Author contributions

### Analysis

TDA, MK, JG, GV, KT, ET, TTN, JN, KV, DL, JB, and BZ

### Sample and/or data provider and processing

J.B.Grauholm, D.D., A.R., G.A., M.B.Hansen, P.Q., B.W., T.T., H.S., K.L.M., V.M., L.F., J.T., B.J.V., J.J.M., M.M, S.M., iPSYCH-Broad Consortium, E.A., K.S., M.M., T.W., D.M.H., P.B.M., M.S., J.G., I.H., P.R., M.J.D., O.M., A.P., and A.D.B.

### Supervision

P.Q., K.L.M., B.J.V., J.J.M, I.H., P.R., M.J.D., O.M., A.P., and A.D.B.

### Writing

T.D.A. and A.D.B.

### Study design and direction

A.D.B.

All authors contributed with critical revision of the manuscript.

