## Supplementary tables and figures for "Identification of 64 new risk loci for major depression, refinement of the genetic architecture and risk prediction of recurrence and comorbidities"

### 1 Supplemental tables

Table S1: Overvies of number of cases and controls by study and MD-subphenotype. Overview is grouped in iPSYCH-only, Meta-analysis of iPSYCH and external summary statistics and external summary statistics.

| Study | Cases | Controls | Total |
|---|---|---|---|
| **iPSYCH-only** | | | |
| iPSYCH2015 MD | 29158 | 38142 | 67300 |
| iPSYCH2015 recurrent | 6018 | 38142 | 44160 |
| iPSYCH2015 recurrent vs single-episode | 6018 | 23140 | 29158 |
| iPSYCH2015 single-episode | 23140 | 38142 | 61282 |
| **Meta** | | | |
| MD-meta primary | 371184 | 978703 | 1349887 |
| MD-meta narrowly defined | 176143 | 528770 | 704913 |
| **External** | | | |
| PGC2 (Wray et al. 2018) | 35077 | 95406 | 130483 |
| Howard et al. 2019 | 230118 | 545339 | 775457 |
| FinnGen | 28098 | 228817 | 256915 |
| MVP (Levey et al. 2021) | 83810 | 166405 | 250215 |

Table S2: Independent GWAS index SNPs (N = 303) of the primary MD-meta (See TableS02.xlsx): A total of 1349887 samples, 371184 cases and 978703 controls were analyzed using RICOPILI. (A) Columns are locus number (rplocus) as indentified using RICOPILI pipeline; indication of whether the locus is consider novel or not (Novel); chromosome (CHR); start position of locus (start); end position of locus (end); marker name / rs-numer for the lead SNP within each locus (LeadSNP); two-sided p-value of lead SNP (P.leadSNP); marker name / rs-numer for the independent SNP within each locus (index.SNP); lead SNP (if any) from Howard et al. within the specific locus (Howard.LeadSNP); lead SNP (if any) from Levey et al. within the specific locus (Levey.LeadSNP); base-pair position of index SNP (BP.index.SNP); two-sided p-value of index SNP (P.index.SNP); odds-ratio of effect allele of index SNP (OR.index.SNP); standard error of odds-ratio (SE.index.SNP); allele names (A1A2.index.SNP), where A1 is the effect allele with regards to OR); A1 allele frequency among 371,184 MD cases (FRQ_A_371184.index.SNP); A1 allele frequency among 978,703 controls (FRQ_U_978703.index.SNP); imputation info score of index SNP (INFO.index.SNP); list of all variants with LD-r2 > 0.1 to index SNP (LD-friends(0.1).p0.001.index.SNP), in brackets LD-$r^2$ and distance in kb sorted by LD-$r^2$; left margin of region defined by LD friends (range.left.index.SNP); right margin of region defined by LD friends (range.right.index.SNP); right margin - left margin in kb (span(kb).index.SNP); list of all variants with LD-r2 > 0.6 to index SNP (LD-friends(0.6).p0.001.index.SNP), in brackets LD-$r^2$ and distance in kb sorted by LD-$r^2$; left margin of region defined by LD friends (range.left.6.index.SNP); right margin of region defined by LD friends (range.right.6.index.SNP); right margin - left margin in kb (span.6(kb).index.SNP); list of entries in NHGRI GWAS catalogue among entries in column friends(.6) (gwas_catalog_span.6.index.SNP); list of genes within the region of friends.6 (±50 kb), in brackets distance to index SNP in kb (genes.6.50kb(dist2index).index.SNP); number of genes within the region of friends.6 (±50 kb), (N.genes.6.50kb.index.SNP). Neighboring index SNPs were considered independent when $r^2 < 0.1$ in a sliding 3 Mb window. For each index SNP we defined the associated LD-region by recording the left and rightmost variant with $r^2 < 0.1$. To define GWAS loci, a 50kb window was added on each side of the LD-region and overlapping LD-regions were combined into a single locus. (B) Results from the stepwise selection procedure selecting independently associated SNPs implemented in GCTA-COJO. Columns are chromosome (Chr); marker ID (SNP); physical position (bp); frequency of the effect allele in the original data (freqA); the effect allele (refA); effect size (b = $\beta$), standard error (se) and p-value (p) from the original GWAS or meta-analysis; estimated effective sample size (n); frequency of the effect allele in the reference sample (freq_geno); effect size (bJ), standard error (bJ_se) and p-value (pJ) from a joint analysis of all the selected SNPs; LD correlation (LD_r) between the SNP i and SNP i + 1 for the SNPs on the list, locus number from ricopili output (rplocus) in table S2A, index SNPs of GWAS locus from ricopili output (rpclump) in table S2A.

| excel sheet | content |
|---|---|
| TableS2A | Index SNPs (303) from RICOPILI output |
| TableS2B | GCTA-COJO (251) independent variants |

Table S3: Independent index SNPs (N = 76) of the narrow MD GWAS meta-analysis (See TableS3.xlsx): A total of 704913 samples, 176143 cases and 528770 controls were analyzed using RICOPILI. Index SNP (SNP); chromosome (CHR); position (BP); two-sided p-value of index SNP (P); odds-ratio of effect allele of index SNP (OR); standard error of odds-ratio (SE); allele names (A1A2), where A1 is the effect allele with regards to OR); A1 allele frequency among 176,143 MD cases (FRQ_A_176143); A1 allele frequency among 528,770 controls (FRQ_U_528770); imputation info score of index SNP (INFO); list of all variants with LD-r2 > 0.1 to index SNP (LD-friends(0.1).p0.001), in brackets LD-$r^2$ and distance in kb sorted by LD-$r^2$; left margin of region defined by LD friends (range.left); right margin of region defined by LD friends (range.right); right margin - left margin in kb (span(kb)); list of all variants with LD-$r^2 > 0.6$ to index SNP (LD-friends(0.6).p0.001), in brackets LD-$r^2$ and distance in kb sorted by LD-$r^2$; left margin of region defined by LD friends (range.left.6); right margin of region defined by LD friends (range.right.6); right margin - left margin in kb (span.6(kb)); list of entries in NHGRI GWAS catalogue among entries in column friends(.6) (gwas_catalog_span.6); list of genes within the region of friends.6 (±50 kb), in brackets distance to index SNP in kb (genes.6.50kb(dist2index)); number of genes within the region of friends.6 (±50 kb), (N.genes.6.50kb). Neighboring index SNPs were considered independent when $r^2 < 0.1$ in a sliding 3 Mb window. For each index SNP we defined the associated LD-region by recording the left and rightmost variant with $r^2 < 0.1$. To define GWAS loci, a 50kb window was added on each side of the LD-region and overlapping LD-regions were combined into a single locus.

| excel sheet | table content |
|---|---|
| TableS3 | Index SNPs from RICOPILI output |

Table S4: Genetic correlations $r_G$ among MD studies/subtypes using LD score regression. Genetic correlations ($r_G$) and its standard error (s.e.) between different MD-GWAS summary statistics and MD-subtypes (MD-primary, MD-narrow,single-episode, recurrent depression) used in the current study. Columns are: MD-subtype/study one (Dataset 1); MD-subtype/study two (Dataset 2); estimate of genetic correlation ($r_G$) using LD-score regression; Standar error of $r_G$ (s.e.); test statistics (z); two-sided nominal p-value (P) of tests for $r_G = 0$ (see figure S6). Sample sizes are: iPSYCH2015 MD $N_{cases} = 29158$, $N_{ctrls} = 38142$, $N_{total} = 67300$; iPSYCH2015 recurrent $N_{cases} = 6018$, $N_{ctrls} = 38142$, $N_{total} = 44160$; iPSYCH2015 recurrent vs single-episode $N_{cases} = 6018$, $N_{ctrls} = 23140$, $N_{total} = 29158$; iPSYCH2015 single-episode $N_{cases} = 23140$, $N_{ctrls} = 38142$, $N_{total} = 61282$; MD-meta primary $N_{cases} = 371184$, $N_{ctrls} = 978703$, $N_{total} = 1349887$; MD-meta narrowly defined $N_{cases} = 176143$, $N_{ctrls} = 528770$, $N_{total} = 704913$; PGC2 (Wray et al. 2018) $N_{cases} = 35077$, $N_{ctrls} = 95406$, $N_{total} = 130483$; Howard et al. 2019 $N_{cases} = 230118$, $N_{ctrls} = 545339$, $N_{total} = 775457$; FinnGen $N_{cases} = 28098$, $N_{ctrls} = 228817$, $N_{total} = 256915$; MVP (Levey et al. 2021) $N_{cases} = 83810$, $N_{ctrls} = 166405$, $N_{total} = 250215$ (see table S1)

| Dataset 1 | Dataset 2 | $r_G$ | s.e | z | p |
|---|---|---|---|---|---|
| Howard et al. 2019 MD | FinnGen MD | 0.92 | 0.05 | 19.25 | 1.5273e-82 |
| Howard et al. 2019 MD | MVP MD | 0.83 | 0.03 | 29.61 | 1.2781e-192 |
| Howard et al. 2019 MD | Wray et al. 2018 (narrow) MD | 0.92 | 0.03 | 36.77 | 5.9824e-296 |
| iPSYCH2015 MD | FinnGen MD | 0.95 | 0.07 | 13.60 | 4.0231e-42 |
| iPSYCH2015 MD | Howard et al. 2019 MD | 0.79 | 0.04 | 21.15 | 2.8694e-99 |
| iPSYCH2015 MD | MVP MD | 0.77 | 0.05 | 14.47 | 1.8517e-47 |
| iPSYCH2015 MD | Wray et al. 2018 (narrow) MD | 0.80 | 0.06 | 13.48 | 2.0766e-41 |
| iPSYCH2015 MD | iPSYCH2015 recurrent | 1.07 | 0.05 | 20.12 | 4.5988e-90 |
| iPSYCH2015 MD | iPSYCH2015 single-episode | 0.99 | 0.01 | 84.93 | 0 |
| MVP MD | FinnGen MD | 0.88 | 0.06 | 13.82 | 1.8591e-43 |
| Wray et al. 2018 (narrow) MD | FinnGen MD | 0.92 | 0.07 | 14.00 | 1.6604e-44 |
| Wray et al. 2018 (narrow) MD | MVP MD | 0.81 | 0.05 | 17.98 | 2.9128e-72 |
| iPSYCH2015 recurrent | FinnGen MD | 0.94 | 0.11 | 8.77 | 1.8397e-18 |
| iPSYCH2015 recurrent | Howard et al. 2019 MD | 0.74 | 0.06 | 11.68 | 1.5293e-31 |
| iPSYCH2015 recurrent | MVP MD | 0.76 | 0.08 | 9.51 | 1.9291e-21 |
| iPSYCH2015 recurrent | Wray et al. 2018 (narrow) MD | 0.75 | 0.08 | 8.87 | 7.1974e-19 |
| iPSYCH2015 single-episode | FinnGen MD | 0.97 | 0.08 | 12.50 | 7.0898e-36 |
| iPSYCH2015 single-episode | Howard et al. 2019 MD | 0.82 | 0.05 | 17.47 | 2.3191e-68 |
| iPSYCH2015 single-episode | MVP MD | 0.80 | 0.06 | 13.00 | 1.1956e-38 |
| iPSYCH2015 single-episode | Wray et al. 2018 (narrow) MD | 0.83 | 0.07 | 12.29 | 9.8752e-35 |
| iPSYCH2015 single-episode | iPSYCH2015 recurrent | 1.10 | 0.09 | 11.73 | 9.2443e-32 |
| iPSYCH2012 MD | FinnGen MD | 0.93 | 0.07 | 12.51 | 6.7681e-36 |
| iPSYCH2012 MD | Howard et al. 2019 MD | 0.78 | 0.05 | 16.52 | 2.6136e-61 |
| iPSYCH2012 MD | iPSYCH2015 MD | 0.99 | 0.02 | 65.43 | 0.0 |
| iPSYCH2012 MD | iPSYCH2015i MD | 0.86 | 0.11 | 7.55 | 4.4411e-14 |
| iPSYCH2012 MD | MVP MD | 0.76 | 0.06 | 12.65 | 1.1732e-36 |
| iPSYCH2012 MD | Wray et al. 2018 (narrow) MD | 0.77 | 0.07 | 11.23 | 3.0487e-29 |
| iPSYCH2012 MD | iPSYCH2015 recurrent | 1.02 | 0.05 | 19.48 | 1.7903e-84 |
| iPSYCH2012 MD | iPSYCH2015 single-episode | 0.98 | 0.03 | 33.31 | 2.8529e-243 |
| iPSYCH2015i MD | FinnGen MD | 0.96 | 0.13 | 7.35 | 2.0155e-13 |
| iPSYCH2015i MD | Howard et al. 2019 MD | 0.74 | 0.07 | 9.95 | 2.5962e-23 |
| iPSYCH2015i MD | iPSYCH2015 MD | 0.89 | 0.05 | 19.23 | 2.0443e-82 |
| iPSYCH2015i MD | MVP MD | 0.70 | 0.09 | 7.73 | 1.0630e-14 |
| iPSYCH2015i MD | Wray et al. 2018 (narrow) MD | 0.76 | 0.09 | 8.10 | 5.3550e-16 |
| iPSYCH2015i MD | iPSYCH2015 recurrent | 1.05 | 0.11 | 9.37 | 7.3875e-21 |
| iPSYCH2015i MD | iPSYCH2015 single-episode | 0.88 | 0.05 | 17.01 | 7.3004e-65 |

Table S5: Heritability estimates of MD studies/subtypes using LD score regression. SNP-based heritability ($h^2_{SNP}$) and its standard error (s.e.) of different MD-GWAS summary statistics and MD-subtypes (MD-primary, MD-narrow,single-episode, recurrent depression) used in the current study. Columns are: MD-subtype/study (Dataset); estimate of SNP-heritability ($h^2_G$) using LD-score regression; Standar error of $h^2_{SNP}$ (s.e.); test statistics (z-score); two-sided nominal p-value (P) of tests for $h^2_{SNP} = 0$; genomic inflation factor $\lambda_{GC}$ (lambda); mean $\chi^2$ ($\chi^2$); intercept (Intercept) and corresponding standard error (s.e); attenuation ratio (Ratio) and corresponding standard error (s.e.) (see figure SX). Sample sizes are: iPSYCH2015 MD $N_{cases} = 29158$, $N_{ctrls} = 38142$, $N_{total} = 67300$; iPSYCH2015 recurrent $N_{cases} = 6018$, $N_{ctrls} = 38142$, $N_{total} = 44160$; iPSYCH2015 recurrent vs single-episode $N_{cases} = 6018$, $N_{ctrls} = 23140$, $N_{total} = 29158$; iPSYCH2015 single-episode $N_{cases} = 23140$, $N_{ctrls} = 38142$, $N_{total} = 61282$; MD-meta primary $N_{cases} = 371184$, $N_{ctrls} = 978703$, $N_{total} = 1349887$; MD-meta narrowly defined $N_{cases} = 176143$, $N_{ctrls} = 528770$, $N_{total} = 704913$; PGC2 (Wray et al. 2018) $N_{cases} = 35077$, $N_{ctrls} = 95406$, $N_{total} = 130483$; Howard et al. 2019 $N_{cases} = 230118$, $N_{ctrls} = 545339$, $N_{total} = 775457$; FinnGen $N_{cases} = 28098$, $N_{ctrls} = 228817$, $N_{total} = 256915$; MVP (Levey et al. 2021) $N_{cases} = 83810$, $N_{ctrls} = 166405$, $N_{total} = 250215$ (see table S1)

| Dataset | $h^2_{SNP}$ | s.e | z | p | $\lambda_{GC}$ | $\chi^2$ | Intercept | s.e. | Ratio | s.e |
|---|---|---|---|---|---|---|---|---|---|---|
| **Meta analyses** | | | | | | | | | | |
| MD | 0.070 | 0.002 | 31.591 | 7.25e-191 | 1.893 | 2.250 | 1.059 | 0.015 | 0.0474 | 0.0118 |
| MD narrow | 0.072 | 0.003 | 23.967 | 5.78e-112 | 1.471 | 1.608 | 1.034 | 0.010 | 0.056 | 0.0165 |
| **iPSYCH2015** | | | | | | | | | | |
| MD | 0.167 | 0.014 | 11.583 | 1.42e-28 | 1.178 | 1.203 | 1.028 | 0.009 | 0.136 | 0.0462 |
| recurrent | 0.123 | 0.019 | 6.484 | 2.42e-10 | 1.074 | 1.083 | 1.011 | 0.009 | 0.1266 | 0.1037 |
| single-episode | 0.134 | 0.014 | 9.278 | 3.63e-19 | 1.140 | 1.162 | 1.031 | 0.009 | 0.1893 | 0.0546 |
| recurrent vs. single-episode | -0.014 | 0.019 | 0.743 | 4.66e-01 | 0.999 | 1.001 | 1.008 | 0.007 | 6.5708 | 5.6742 |
| **External** | | | | | | | | | | |
| Howard et al. 2019 MD | 0.080 | 0.003 | 27.655 | 1.63e-147 | 1.637 | 1.839 | 1.022 | 0.013 | 0.0268 | 0.0159 |
| Wray et al. 2018 MD-narrow | 0.110 | 0.008 | 13.762 | 3.13e-39 | 1.165 | 1.173 | 0.997 | 0.007 | $< 0$ | - |
| FinnGen MD | 0.069 | 0.010 | 7.124 | 4.11e-12 | 1.162 | 1.173 | 1.066 | 0.008 | 0.3792 | 0.0487 |
| MVP MD | 0.072 | 0.005 | 14.896 | 1.88e-45 | 1.220 | 1.261 | 1.021 | 0.009 | 0.0805 | 0.0337 |

Table S6: Genetic correlations of MD primary meta-analysis and other traits (see TableS6.xlsx): Genetic correlation ($r_G$) of the primary (N=1349887) and narrow (N=704913) MD meta-analysis and MD-narrow meta-analysis (see table S1) verus published GWAS summary statistics available at LDhub or locally grouped in non-UKBB (Table S6A) and UKBB traits (Table S6B). (see figure S8). Columns are as follows: Trait of the external summary statistics (Trait); Category of trait (Category); Ethnicity of trait (ethnicity), Pubmed ID of external study (PMID); Genetic correlation with the primary MD meta-analysis (rg.primary); standard error (se.primary); 95% confidence limits(CIL.primary, CIU.primary); z-score (z.primary); two-sided nominal p-value (p.primary); bonferroni adjusted p-value (padj.primary); SNP-heritability on the observed scale (h2_obs.primary), standard error (h2_obs_se.primary) LD score regression intercept (h2_int.primary) and standard error (h2_int_se.primary) for the external trait in analysis with the primary trait; Cross-trait LD score regression intercept (gcov_int.primary) and standard error (gcov_int_se.primary) of external trait and the MD-primary GWAS; The remaining columns are from the comparison of the external trait with the meta-analysis of the narrow definition of MD: Genetic correlation (rg.narrow), standar error (se.narrow) and 95% CI limits (CIL.narrow, CIU.narrow); z-score (z.narrow); nominal p-value (p.narrow); bonferroni adjusted p-value (padj.narrow); SNP-heritability on the observed scale (h2_obs.narrow) and standard error (h2_obs_se.narrow); single-trait LD score regression intercept (h2_int.narrow) and standard error (h2_int_se.narrow) of the external trait from the analysis with MD-narrow; Cross-trait LD score regression intercept (gcov_int.narrow) and standard error (gcov_int_se.narrow) of external trait and the MD-narrow meta-analysis (see figure S8B).

| excel sheet | content |
|---|---|
| TableS6A | Significant genetic correlations of primary/narrow MD meta-analysis with non-UKB traits |
| TableS6B | Significant genetic correlations of primary/narrow MD meta-analysis with UKB traits |

Table S7: MiXeR uni- and bivariate analyses: $n$ is the number of variants from the univariate analysis, while $n_1$, $n_2$ and $n_{12}$ are the number of unique variants for each trait and the overlap between MD and each trait, respectively, from the bivariate analysis. Fraction shared $f_{12}$ = fraction shared out of all trait 1 influencing variants, while $f_{21}$ = fraction shared out of all MD influencing variants; $\rho_{12}$, correlation of genetic effects for the overlapping variants; $r_g$, genetic correlation; concordant fraction (fraction_concordant_within_shared) indicates the fraction of influencing variants within the shared component that have concordant effect directions; best_vs_min_overlap AIC and BIC estimates indicate if MiXeR can accurately distinguish the reported overlap from the minimum possible overlap allowed; best vs max overlap AIC and BIC estimates indicate if MiXeR can accurately distinguish the reported overlap from the maximum possible overlap allowed. Traits are MD = major depression (N = 1349887), ANX = anxiety (N = 361365), BP = bipolar disorder (N = 405771), SZ = schizophrenia (N = 153808), ADHD = Attention deficit hyperactivity disorder (N = 225534), ASD = autism spectrum disorder (N = 46350), SUD = substance use disorder (N = 46568), Neuroticism = neuroticism (N = 380506), Smoking = smoking initiation (N = 632803), EA = educational attainment (N = XXXX). See main figure 3.

| | | | | | | | | | Fraction shared | | | | | | Concordant | | Best vs min | | Best vs max | |
|---|---|---|---|---|---|---|---|---|---|---|---|---|---|---|---|---|---|---|---|---|
| Trait | $n$ | $sd(n)$ | $n_1$ | $sd(n_1)$ | $n_2$ | $sd(n_2)$ | $n_{12}$ | $sd(n_{12})$ | $f_{12}$ | $f_{21}$ | $\rho_0$ | $sd(\rho_0)$ | $r_G$ | sd($r_G$) | fraction | sd | AIC | BIC | AIC | BIC |
| MD | 11750.27 | 310.02 | - | - | - | - | - | - | - | - | - | - | - | - | - | - | - | - | - | - |
| ANX | 8361.81 | 999.78 | 48.59 | 90.66 | 3437.06 | 1019.91 | 8313.21 | 948.94 | 0.99 | 0.71 | 0.2 | 0.004 | 0.79 | 0.017 | 0.91 | 0.057 | 0.47 | -8.66 | -1.07 | -10.2 |
| BP | 7438.87 | 264.09 | 198.68 | 259.57 | 4510.08 | 396.53 | 7240.19 | 393.84 | 0.97 | 0.62 | 0.11 | 0.002 | 0.47 | 0.006 | 0.71 | 0.011 | 6.87 | -2.28 | -0.6 | -9.75 |
| SZ | 8459.31 | 242.14 | 129.49 | 82.79 | 3420.44 | 270.57 | 8329.83 | 269.52 | 0.98 | 0.71 | 0.09 | 0.003 | 0.34 | 0.006 | 0.63 | 0.003 | 21.2 | 12.08 | -0.49 | -9.61 |
| ADHD | 6989.62 | 285.85 | 173.56 | 169.35 | 4934.21 | 422.61 | 6816.06 | 356.96 | 0.98 | 0.58 | 0.15 | 0.003 | 0.56 | 0.007 | 0.77 | 0.015 | 5.11 | -4.02 | -0.59 | -9.71 |
| ASD | 9936.47 | 1050.76 | 1501.32 | 2181.86 | 3315.12 | 2388.17 | 8435.15 | 2437.09 | 0.85 | 0.72 | 0.09 | 0.002 | 0.37 | 0.012 | 0.7 | 0.119 | -0.26 | -9.33 | -0.84 | -9.91 |
| SUD | 6289.39 | 2418.43 | 604.45 | 1335.94 | 6065.33 | 1630.71 | 5684.94 | 1555.94 | 0.9 | 0.48 | 0.05 | 0.003 | 0.59 | 0.021 | 0.88 | 0.083 | -0.41 | -9.57 | -0.83 | -9.99 |
| Neuroticism | 12225.76 | 336.22 | 1846.97 | 478.19 | 1371.49 | 425.65 | 10378.78 | 466.39 | 0.85 | 0.88 | 0.26 | 0.002 | 0.72 | 0.004 | 0.81 | 0.019 | 10.53 | 1.37 | 1.24 | -7.92 |
| Smoking | 11058.76 | 325.62 | 230.11 | 201.4 | 921.63 | 306.54 | 10828.64 | 298.83 | 0.98 | 0.92 | 0.08 | 0.003 | 0.37 | 0.005 | 0.63 | 0.003 | 24.58 | 15.43 | -0.17 | -9.32 |
| EA | 13083.90 | 231.28 | 1554.95 | 335.22 | 221.32 | 163.5 | 11528.95 | 300.68 | 0.88 | 0.98 | -0.05 | 0.003 | -0.23 | 0.005 | 0.42 | 0.002 | 206.65 | 197.49 | 0.48 | -8.68 |

Table S8: Association of MD-PRS with neurocognitive phenotypes in the Philadelphia Neurodevelopmental cohhort (N=4,973). Neurocognitive phenotype, broad neurocognitive domain, meteric used as response varianle, sample size (N), beta ($\beta$) from linear regression, lower (CIL) and upper (CIU) 95% confidence limits, standard error of the beta (S.E.), (t), nominal p-value ($P$), FDR adjusted p-value ($P_{FDR}$). The total variance explained by MD-PRS and model covariates for each neurocognitive phenotype was reported using $R^2$, adjusted ($R^2_{adj}$) and variance explaied by the MD-PRS.

| Neurocog. phenotype | Broad Neurocog. domain | Metric used as response variable | N | $\beta$ | $CI_L$ | $CI_U$ | S.E. | t | $P$ | $P_{FDR}$ | $R^2$ | $R^2_{adj}$ | Variance explained by MD-PRS |
|---|---|---|---|---|---|---|---|---|---|---|---|---|---|
| Age differentiation | Social cognition | Penn Age Differentiation Test – PADT PC (Percent Correct Responses for All Test Trials) | 4740 | 0.003 | -0.022 | 0.028 | 0.013 | 0.22 | 0.823 | 0.979 | 0.21 | 0.20 | 1.4e-06 |
| Standardized reading test performance | Other | Wide Range Achievement Test (reading component of test, used as IQ estimate) – WRAT CR STD (Wide Range Assessment Test Total Standard Score, age-adjusted) | 4854 | 0.000 | -0.026 | 0.026 | 0.013 | -0.03 | 0.979 | 0.979 | 0.04 | 0.04 | 5.8e-08 |
| Facial memory | Episodic memory | Penn Face Memory Test – PFMT IFAC TOT (Total Correct Responses for All Test Trials) | 4744 | -0.001 | -0.028 | 0.025 | 0.014 | -0.08 | 0.933 | 0.979 | 0.13 | 0.13 | 4.9e-07 |
| Emotion differentiation | Social cognition | Penn Emotion Differentiation Test – PEDT PC (Percent of Correct Responses for All Test Trials) | 4845 | -0.002 | -0.027 | 0.023 | 0.013 | -0.16 | 0.873 | 0.979 | 0.18 | 0.17 | 7.0e-07 |
| Emotion identification | Social cognition | Penn Emotion Identification Test – PEIT CR (Total Correct Responses for All Test Trials) | 4868 | -0.003 | -0.028 | 0.023 | 0.013 | -0.19 | 0.848 | 0.979 | 0.12 | 0.12 | 3.2e-06 |
| Finger tapping speed | Motor/sensorimotor speed | Finger Tapping Test – TAP TOT (Sum of Mean of Tap Responses for Dominant/Non-Dominant Hand Trials) | 4656 | -0.004 | -0.026 | 0.019 | 0.011 | -0.31 | 0.759 | 0.979 | 0.41 | 0.41 | 1.8e-06 |
| Sensorimotor processing | Motor/sensorimotor speed | Motor Praxis – MP MP2RTCR (Median Response Time for Correct Mouse Click Responses, then reverse-scored so that higher = better performance) | 4929 | -0.004 | -0.029 | 0.022 | 0.013 | -0.27 | 0.787 | 0.979 | 0.09 | 0.09 | 3.1e-06 |
| Spatial reasoning | Reasoning | Penn Line Orientation Test – PLOT PC (Percent Correct Responses for All Test Trials) | 4553 | -0.008 | -0.034 | 0.018 | 0.013 | -0.62 | 0.533 | 0.979 | 0.17 | 0.17 | 1.3e-05 |
| Verbal memory | Episodic memory | Penn Word Memory Test – PWMT KIWRD TOT (Total Correct Responses for All Test Trials) | 4809 | -0.013 | -0.040 | 0.014 | 0.014 | -0.96 | 0.335 | 0.717 | 0.03 | 0.03 | 1.5e-04 |
| Nonverbal reasoning | Reasoning | Penn Matrix Reasoning Test – PMAT CR (Total Correct Responses for All Test Trials) | 4841 | -0.024 | -0.051 | 0.002 | 0.013 | -1.83 | 0.068 | 0.169 | 0.13 | 0.13 | 2.7e-04 |
| Working memory | Executive control | Letter N-Back Test – LNB TP (Total Correct Responses to 0-Back, 1-Back, and 2-Back Trials) | 4526 | -0.026 | -0.051 | -0.002 | 0.013 | -2.10 | 0.036 | 0.107 | 0.15 | 0.15 | 2.5e-04 |
| Verbal reasoning | Reasoning | Penn Verbal Reasoning Test – PVRT CR (Total Correct Responses for All Test Trials) | 4848 | -0.029 | -0.050 | -0.008 | 0.011 | -2.73 | 0.006 | 0.032 | 0.34 | 0.34 | 1.3e-04 |
| Spatial memory | Episodic memory | Visual Object Learning Test – VOLT SVT (Total Correct Responses for All Test Trials) | 4786 | -0.030 | -0.057 | -0.002 | 0.014 | -2.13 | 0.034 | 0.107 | 0.02 | 0.01 | 4.9e-04 |
| Abstraction and mental flexibility | Executive control | Penn Conditional Exclusion Test – PCET ACC2 (Calculated Accuracy Measure) | 4821 | -0.036 | -0.062 | -0.010 | 0.013 | -2.73 | 0.006 | 0.032 | 0.06 | 0.05 | 8.0e-04 |
| Attention | Executive control | Penn Continuous Performance Test – PCPT T TP (Total of Correct Responses to Number/Letter Trials) | 4527 | -0.036 | -0.060 | -0.012 | 0.012 | -2.90 | 0.004 | 0.032 | 0.21 | 0.21 | 5.3e-04 |

Table S9: Summary of the FUMA SNP2GENE analyses: see TableS9.xlsx Significant genes from the gene-based MAGMA analysis of the primary MD GWAS (N = 1349887). GRL collumn indicate whether a gene was within a region identified as a genomic risk locus (+) or not (−). NA

| excel sheet | content |
|---|---|
| TableS9A | FUMA datasets |
| TableS9B | FUMA mapped genes |
| TableS9C | FUMA snps |
| TableS9D | FUMA gene-based MAGMA |

Table S10: FUMA GENE2FUNC analysis of 411 significant genes (See TableS10.xlsx). A total of 8,664 gene-sets derived from GO Biological Process (n=7,658) and GO Cellular Components (n=1,006) ontology in the MSigDB database were analysed. **Supplementary table 10A:** list of parameters in the FUMA GENE2FUNC analysis. **Supplementary table 10B:** The gene-to-function analysis identifyed 479 significant gene-sets after correction for multiple testing (FDR<0.05). Columns are: One of the category from MsigDB (Category); Name of gene set as provided by MsigDB (GeneSet); Number of genes in a gene set (N__genes); Number of input genes overlapping with the gene set (N__overlap); Hypergeometric test (upper tail) P-value (p); FDR adjusted p-value per category (adjP); FDR corrected p-value across categories (adjPtotalBH); Genes overlapping with the gene set (genes) and Link to the MsigDB page if available (link). Hypergeometric tests are performed to test if genes of interest are overrepresented in any of the pre-defined gene sets.

| excel sheet | table content |
|---|---|
| TableS10A | FUMA GENE2FUNC parameters |
| TableS10B | FUMA GENE2FUNC analysis of MAGMA genes |

Table S11: TWAS results: see TableS11.xlsx. **Supplementary table 11A:** TWAS summary. Number of imputed genes and transcripts (n.imputed); Number of significant genes/transcripts after bonferroni correction (p.sig) and after FDR (fdr.sig). **Supplementary table 11B:** List of bonferroni significant genes and transcripts by LD-block. The ENSEMBL ID of the lead gene or transcript within an LD-block (Lead), lead genes or transcripts that have a p-value a 100 fold lower than the gene/transcript with the second lowest p-value are highlighted in red and bold; The gene's ENSEMBL ID (gene); Gene or transcript name as listed by the transcriptome model, typically HUGO (feature_name); Indication of level of gene or transcript (level); A gene's or transcript's ENSEMBL ID as listed in the tissue transcriptome model (feature); (R); chromosome (CHR); position (BP); Description of the imputation model used (tissue); S-EpiXcan's association z-score for the feature (zscore); S-EpiXcan's association p-value for the feature (pvalue); S-EpiXcan's association FDR-adjusted p-value for the feature, i.e. study-wide multiple testing correction (FDR); S-EpiXcan's association Bonferroni-adjusted p-value for the feature study-wide multiple testing (Bonferroni); Gene or transcript name as listed by the transcriptome model, typically HUGO. It is the same as feature.name in the case of genes (gene_name); S-EpiXcan's association effect size for the gene. Can only be computed when beta from the GWAS is used (effect_size); Variance of the gene expression, calculated as W' * G * W (where W is the vector of SNP weights in a gene's model, W' is its transpose, and G is the covariance matrix) (var_g); Cross-validated R2 of tissue model's correlation to gene's measured transcriptome prediction performance (pred_perf_r2); p-value of tissue model's correlation to gene's measured transcriptome prediction performance (pred_perf_pval); q-value of tissue model's correlation to gene's measured transcriptome prediction performance (pred_perf_qval); Number of SNPs from GWAS that were used in S-EpiXcan analysis (n_snps_used); Number of SNPs in the covariance matrix (n_snps_in_cov); Number of SNPs in the imputation model (n_snps_in_model); feature type (type); internal imputation model tracking ID; it contains inofrmation about the tissue of origin, cell specificity, assayed molecular species and model training method (model_ID); S-EpiXcan's association FDR-adjusted p-value for the feature, multiple testing correction only for this phenotype-imputation model combination (fdr_tissue_trait); Disregard; this is the S-EpiXcan's association FDR-adjusted p-value for the feature, multiple testing correction for all possible imputation models for the specific trait (fdr_gwas); GRCh37 is based on ENSEMBL 87 release on the GRCh37 assembly (REFERENCE); Number of credible SNPs from finemapping results used in the imputation model (NcrSNPs); SNP names of SNPs in the imputation model (SNPs); LD-block number (LDblock); starting position of LD-block (LDblock.start); ending poistion of LD-block (LDblock.stop); GW-significant index SNPs located within the LD-block if any (RPiSNP). **Supplementary table 11C:** List of FDR significant genes. Titles of columns overlap with supplementary table 11B. **Supplementary table 11D:** List of FDR significant genes. Titles of columns overlap with supplementary table 11B.

| excel sheet | table content |
|---|---|
| TableS11A | TWAS summary |
| TableS11B | Bonferroni significant genes and transcripts by LD block |
| TableS11C | FDR significant genes |
| TableS11D | FDR significant transcripts |

Table S12: eQTL colocalization results. Gene name / symbol (symbol); Gene ID (Gene ID); Chromosome (chr); starting position of gene (start); ending position of gene (end); Posterior probability (PPH4) for both GWAS and eQTL association and co-localization (hypothesis 4), considering PPH4 > 0.8 as strong evidence from both GWAS, eQTL and co-localization (PPH4), Number of credible SNPs within gene (NcrSNPs)

| Gene symbol | Gene ID | chr | start | end | PPH4 | NcrSNPs |
|---|---|---|---|---|---|---|
| NEGR1 | ENSG00000172260 | 1 | 71361623 | 71861623 | 0.97 | 15 |
| PRMT6 | ENSG00000198890 | 1 | 107099267 | 108000000 | 0.91 | 0 |
| GPR27 | ENSG00000170837 | 3 | 71803201 | 71805647 | 0.92 | 17 |
| ARAP3 | ENSG00000120318 | 5 | 140532968 | 141000000 | 0.83 | 0 |
| ZNHIT1 | ENSG00000106400 | 7 | 100360949 | 101000000 | 0.90 | 0 |
| ASPH | ENSG00000198363 | 8 | 61913116 | 62413116 | 0.85 | 0 |
| CKS2 | ENSG00000123975 | 9 | 91426113 | 91926113 | 0.95 | 0 |
| HOGA1 | ENSG00000241935 | 10 | 98844080 | 99344080 | 0.94 | 0 |
| KLC1 | ENSG00000126214 | 14 | 103528233 | 104000000 | 0.82 | 2 |
| FURIN | ENSG00000140564 | 15 | 90911822 | 91411822 | 1.00 | 1 |
| MT1X | ENSG00000187193 | 16 | 56216336 | 56716336 | 0.86 | 0 |
| ALDH16A1 | ENSG00000161618 | 19 | 49456426 | 49956426 | 0.87 | 0 |
| MACROD2 | ENSG00000172264 | 20 | 13476015 | 13976015 | 0.83 | 0 |

Table S13: eCaviar eQTL colocalization results. Gene name / symbol (Gene symbol); Gene ID (Gene ID); Chromosome (CHR); Starting position of gene (start); Ending position of gene (end); index SNP (iSNP); Posterior probability (PP).

| Gene symbol | Gene ID | CHR | start | end | iSNP | CLPP |
|---|---|---|---|---|---|---|
| FURIN | ENSG00000140564 | 15 | 91411822 | 91426688 | rs4702 | 0.962 |
| FURIN | ENSG00000140564 | 15 | 91411822 | 91426688 | rs4702 | 0.962 |
| TCTA | ENSG00000145022 | 3 | 49449639 | 49453908 | rs4955414 | 0.010 |
| GPR27 | ENSG00000170837 | 3 | 71803201 | 71805647 | rs73090632 | 0.023 |
| GPR27 | ENSG00000170837 | 3 | 71803201 | 71805647 | rs73090632 | 0.023 |

Table S14: Number of samples in iPSYCH2015Number of MD, signle-episode and recurrent cases and controls in iPSYCH2015 by dataset (iPSYCH2012 and iPSYCH2015i) and by sex (Females and Males) before and after relatedness pruning / removal of ancestry outliers. Mean age and standard error by subgroup after relatedness pruning / removal of ancestry outliers are reported.

| | iPSYCH2012 | | | iPSYCH2015i | | | iPSYCH2015 | Age | |
|---|---|---|---|---|---|---|---|---|---|
| Phenotype | Females | Males | total | Femals | Males | total | total | mean | sd |
| MD | 15219 | 7166 | 22385 | 7777 | 3933 | 11710 | 34095 | - | - |
| | 13695 | 6463 | 20158 | 6962 | 3498 | 10460 | 30618 | 25.9 | 5.6 |
| single | 11130 | 5644 | 16774 | 6634 | 3399 | 10033 | 26807 | - | - |
| | 10030 | 5096 | 15126 | 5947 | 3028 | 8975 | 24101 | 25.7 | 5.7 |
| recurrent | 4089 | 1522 | 5611 | 1143 | 534 | 1677 | 7288 | - | - |
| | 3665 | 1367 | 5032 | 1015 | 470 | 1485 | 6517 | 26.8 | 5.1 |
| ctrl | 13005 | 13978 | 26983 | 9103 | 9307 | 18410 | 45393 | - | - |
| | 10950 | 11800 | 22750 | 7641 | 7809 | 15450 | 38200 | 21.2 | 7.3 |

Table S15: Genetic correlations of other traits with MD-subtypes within iPSYCH2015 (see TableS15.xlsx). Genetic correlation ($r_G$) of within iPSYCH2015 MD, single-episode and recurrent depression (see table S1) verus published GWAS summary statistics available at LDhub or locally grouped in non-UKBB (Table S15A, see figure S15A) and UKBB traits (Table S15B, see figure S15B). Columns are as follows: Trait of the external summary statistics (Trait); Category of trait (Category); Ethnicity of trait (ethnicity), Pubmed ID of external study (PMID); Genetic correlation with the iPSYCH2015 MD-subtype (rg.); standard error (se.); 95% confidence limits(CIL., CIU.); z-score (z.); two-sided nominal p-value (p.); bonferroni adjusted p-value (padj.); SNP-heritability on the observed scale (h2_obs.), standard error (h2_obs_se.) LD score regression intercept (h2_int.) and standard error (h2_int_se.) for the external trait in analysis with the primary trait; Cross-trait LD score regression intercept (gcov_int.) and standard error (gcov_int_se.) of external trait and the MD-subtype GWAS; The remaining columns are from the comparison of the external trait with iPSYCH2015 single-episode and recurrent depression.

| excel sheet | table content |
|---|---|
| TableS15A | Significant genetic correlations of iPSYCH MD with non-UKB traits |
| TableS15B | Significant genetic correlations of iPSYCH MD with UKB traits |

Table S16A: Multivariate PRS analyses of single-episode vs. recurrent depression . Left-most collumn indicate which trait the training of the PRS is based upon and the sub-phenotype-group within the target sample (iPSYCH2015) the summary statistics is based upon. N = sample size by sub-phenotype in target, $\beta$ = slope of the linear regresssion, $SE$ = stand error, $Preg$ are p-values directly from the regression, ie. first entry is for $H_0$ : $intercept = 0$, the remaing are for $H_0$ : no difference to reference group ctrl, $CI_l$ and $C_u$ is lower and upper 95% confidence limits, $P_0$ corresponds to $H_0 : \beta + interscept = 0$, except for the control group columns where it is $H_0$ : $intercept = 0$ as in $P_{reg}$. The remaining $P$-columns holds the p-values for the Wald test of equal group effect: $P_{ctrl}$ is p-value for the control group, while $P_{sMD}$ and $P_{rMD}$ are p-values for the Wald test for single-episode and recurrent depression respectively. Note that all individuals with BP were excluded. . Overall p-value = 0.00075

| PRS based on: | $N$ | $\beta$ | $SE$ | $P_{reg}$ | $CI_l$ | $CI_u$ | $P_0$ | $P_{ctrl}$ | $P_{sMD}$ | $P_{rMD}$ |
|---|---|---|---|---|---|---|---|---|---|---|
| **Major Depression** | | | | | | | | | | |
| ctrl | 38142 | -0.25 | 0.0059 | < 1e-300 | -0.260 | -0.240 | < 1e-300 | - | <1e-300 | 4.4e-218 |
| single episode | 23140 | 0.36 | 0.0081 | < 1e-300 | 0.350 | 0.380 | 1.1e-60 | <1e-300 | - | 4.8e-06 |
| recurrent | 6018 | 0.43 | 0.0140 | 4.4e-218 | 0.400 | 0.450 | 7.9e-46 | 4.4e-218 | 4.8e-06 | - |
| **Anxiety** | | | | | | | | | | |
| ctrl | 38142 | -0.11 | 0.0060 | 2.2e-74 | -0.120 | -0.099 | 2.2e-74 | - | 2.5e-81 | 1.1e-46 |
| single episode | 23140 | 0.16 | 0.0083 | 2.5e-81 | 0.140 | 0.180 | 1.3e-11 | 2.5e-81 | - | 5.0e-03 |
| recurrent | 6018 | 0.20 | 0.0140 | 1.1e-46 | 0.170 | 0.230 | 6.0e-12 | 1.1e-46 | 5.0e-03 | - |
| **Bipolar Disorder** | | | | | | | | | | |
| ctrl | 38142 | -0.19 | 0.0055 | 3.0e-263 | -0.200 | -0.180 | 3.0e-263 | - | 8.5e-68 | 8.1e-41 |
| single episode | 23140 | 0.13 | 0.0076 | 8.5e-68 | 0.120 | 0.150 | 1.0e-19 | 8.5e-68 | - | 4.3e-03 |
| recurrent | 6018 | 0.17 | 0.0130 | 8.1e-41 | 0.140 | 0.190 | 6.3e-02 | 8.1e-41 | 4.3e-03 | - |
| **Schizophrenia** | | | | | | | | | | |
| ctrl | 38142 | -0.19 | 0.0056 | 9.2e-243 | -0.200 | -0.170 | 9.2e-243 | - | 5.2e-59 | 6.0e-28 |
| single episode | 23140 | 0.12 | 0.0077 | 5.2e-59 | 0.110 | 0.140 | 1.4e-20 | 5.2e-59 | - | 2.2e-01 |
| recurrent | 6018 | 0.14 | 0.0130 | 6.0e-28 | 0.120 | 0.170 | 1.5e-04 | 6.0e-28 | 2.2e-01 | - |
| **ADHD** | | | | | | | | | | |
| ctrl | 38142 | -0.24 | 0.0059 | < 1e-300 | -0.250 | -0.230 | < 1e-300 | - | 4.5e-156 | 2.9e-64 |
| single episode | 23140 | 0.22 | 0.0082 | 4.5e-156 | 0.200 | 0.230 | 3.2e-03 | 4.5e-156 | - | 3.4e-01 |
| recurrent | 6018 | 0.23 | 0.0140 | 2.9e-64 | 0.200 | 0.260 | 5.6e-01 | 2.9e-64 | 3.4e-01 | - |
| **Autism** | | | | | | | | | | |
| ctrl | 38142 | -0.12 | 0.0060 | 1.2e-84 | -0.130 | -0.100 | 1.2e-84 | - | 2.4e-167 | 1.5e-85 |
| single episode | 23140 | 0.23 | 0.0082 | 2.4e-167 | 0.210 | 0.240 | 1.5e-54 | 2.4e-167 | - | 3.0e-03 |
| recurrent | 6018 | 0.27 | 0.0140 | 1.5e-85 | 0.240 | 0.300 | 7.9e-33 | 1.5e-85 | 3.0e-03 | - |
| **Neurotisism** | | | | | | | | | | |
| ctrl | 38142 | -0.21 | 0.0057 | 2.2e-280 | -0.220 | -0.200 | 2.2e-280 | - | 1.8e-119 | 8.7e-46 |
| single episode | 23140 | 0.18 | 0.0079 | 1.8e-119 | 0.170 | 0.200 | 1.1e-03 | 1.8e-119 | - | 7.8e-01 |
| recurrent | 6018 | 0.19 | 0.0130 | 8.7e-46 | 0.160 | 0.210 | 1.3e-01 | 8.7e-46 | 7.8e-01 | - |
| **Substance Use** | | | | | | | | | | |
| ctrl | 38142 | -0.21 | 0.0050 | < 1e-300 | -0.220 | -0.200 | < 1e-300 | - | 8.3e-51 | 8.1e-24 |
| single episode | 23140 | 0.10 | 0.0069 | 8.3e-51 | 0.089 | 0.120 | 5.6e-72 | 8.3e-51 | - | 2.9e-01 |
| recurrent | 6018 | 0.12 | 0.0110 | 8.1e-24 | 0.093 | 0.140 | 1.5e-18 | 8.1e-24 | 2.9e-01 | - |

Table S16B: Comparison of effect size ($\beta$) of mvPRS analysis of single-episode and recurrent depression from the current study with estimates reported in Wray et al. 2018. Please note that although estimates reported by Wray et al. are based on iPSYCH2012, and thus overlap with the current study, a more strict definition of recurrent depression has been used in the current study, which is also indicated by a modest difference in sample size between iPSYCH2012 from Wray et al. 2018 and the current study that includes iPSYCH2012 + iPSYCH2015i.

| | Wray et al. | | | Current study | | | Test | |
|---|---|---|---|---|---|---|---|---|
| | $\beta$ | s.e. | n | $\beta$ | s.e. | n | t | p |
| single-episode | 0.257 | 0.012 | 12968 | 0.36 | 0.008 | 23140 | -891.116 | < 2.2e-16 |
| recurrent | 0.306 | 0.016 | 5574 | 0.43 | 0.014 | 6018 | -443.82 | < 2.2e-16 |

Table S16C: mvPRS analyses of single-episode vs. recurrent without co-diagnoses. Left-most collumn indicate which trait the training of the PRS is based upon and the sub-phenotype-group within the target sample (iPSYCH2015) the summary statistics is based upon. Subset indicates which co-diagnosis is excluded, which is either all or one of the following ANX, ADHD, ASD, SZ, SUD. Thus woANX, woADHD, woASD, woSZ and woSUD is analyses excluding anxiety, ADHD, ASD, SZ and SUD respectively. Only analysis excluding the signle co-diagnosis for the given trait of the PRS is included. N = sample size by sub-phenotype in target, $\beta$ = slope of the linear regresssion, $SE$ = stand error, $Preg$ are p-values directly from the regression, ie. first entry is for $H_0$: $intercept = 0$, the remaing are for $H_0$: no difference to reference group ctrl, $CI_l$ and $C_u$ is lower and upper 95% confidence limits, $P_0$ corresponds to $H_0$: $\beta + interscept = 0$, except for the control group columns where it is $H_0$: $intercept = 0$ as in $P_{reg}$. The remaining $P$-columns holds the p-values for the Wald test of equal group effect: $P_{ctrl}$ is p-value for the control group, while $P_{sMD}$ and $P_{rMD}$ are p-values for the Wald test for single-episode and recurrent depression respectively. Note that all individuals with BP were excluded.

| PRS based on: | Subset | $N$ | $\beta$ | $SE$ | $P_{reg}$ | $CI_l$ | $CI_u$ | $P_0$ | $P_{ctrl}$ | $P_{sMD}$ | $P_{rMD}$ |
|---|---|---|---|---|---|---|---|---|---|---|---|
| **Anxiety** | | | | | | | | | | | |
| ctrl | woAll | 53592 | -0.040 | 0.005 | 1.7e-17 | -0.060 | -0.030 | 1.7e-17 | - | 5.3e-10 | 1.3e-06 |
| single episode MD | woAll | 12765 | 0.060 | 0.010 | 5.3e-10 | 0.040 | 0.080 | 8.1e-02 | 5.3e-10 | - | 7.9e-02 |
| recurrent episodes MD | woAll | 2461 | 0.100 | 0.020 | 1.3e-06 | 0.060 | 0.100 | 6.6e-03 | 1.3e-06 | 7.9e-02 | - |
| ctrl | woANX | 46704 | -0.060 | 0.006 | 2.1e-29 | -0.070 | -0.050 | 2.1e-29 | - | 2.3e-24 | 8.5e-17 |
| single episode MD | woANX | 18036 | 0.090 | 0.009 | 2.3e-24 | 0.070 | 0.100 | 8.5e-04 | 2.3e-24 | - | 6.8e-03 |
| recurrent episodes MD | woANX | 4078 | 0.100 | 0.020 | 8.5e-17 | 0.100 | 0.200 | 3.0e-06 | 8.5e-17 | 6.8e-03 | - |
| **Schizophrenia** | | | | | | | | | | | |
| ctrl | woAll | 53592 | -0.100 | 0.005 | 7.7e-155 | -0.100 | -0.100 | 7.7e-155 | - | 7.9e-04 | 1.5e-02 |
| single episode MD | woAll | 12765 | 0.030 | 0.009 | 7.9e-04 | 0.010 | 0.050 | 1.5e-30 | 7.9e-04 | - | 4.4e-01 |
| recurrent episodes MD | woAll | 2461 | 0.050 | 0.020 | 1.5e-02 | 0.009 | 0.080 | 8.6e-06 | 1.5e-02 | 4.4e-01 | - |
| ctrl | woSZ | 43565 | -0.200 | 0.005 | 1.2e-182 | -0.200 | -0.100 | 1.2e-182 | - | 2.1e-23 | 4.2e-10 |
| single episode MD | woSZ | 20367 | 0.080 | 0.008 | 2.1e-23 | 0.060 | 0.090 | 1.0e-26 | 2.1e-23 | - | 5.3e-01 |
| recurrent episodes MD | woSZ | 4886 | 0.090 | 0.010 | 4.2e-10 | 0.060 | 0.100 | 7.7e-07 | 4.2e-10 | 5.3e-01 | - |
| **ADHD** | | | | | | | | | | | |
| ctrl | woAll | 53592 | -0.200 | 0.005 | 4.0e-199 | -0.200 | -0.100 | 4.0e-199 | - | 8.5e-23 | 7.6e-08 |
| single episode MD | woAll | 12765 | 0.100 | 0.010 | 8.5e-23 | 0.080 | 0.100 | 2.0e-11 | 8.5e-23 | - | 5.2e-01 |
| recurrent episodes MD | woAll | 2461 | 0.100 | 0.020 | 7.6e-08 | 0.070 | 0.100 | 1.7e-02 | 7.6e-08 | 5.2e-01 | - |
| ctrl | woADHD | 42231 | -0.200 | 0.006 | 1.5e-255 | -0.200 | -0.200 | 1.5e-255 | - | 3.3e-72 | 1.7e-26 |
| single episode MD | woADHD | 21207 | 0.100 | 0.008 | 3.3e-72 | 0.100 | 0.200 | 6.6e-10 | 3.3e-72 | - | 8.3e-01 |
| recurrent episodes MD | woADHD | 5380 | 0.200 | 0.010 | 1.7e-26 | 0.100 | 0.200 | 1.8e-03 | 1.7e-26 | 8.3e-01 | - |
| **Autism** | | | | | | | | | | | |
| ctrl | woAll | 53592 | -0.005 | 0.005 | 3.5e-01 | -0.020 | 0.005 | 3.5e-01 | - | 1.7e-01 | 5.6e-01 |
| single episode MD | woAll | 12765 | 0.010 | 0.010 | 1.7e-01 | -0.006 | 0.030 | 3.6e-01 | 1.7e-01 | - | 9.4e-01 |
| recurrent episodes MD | woAll | 2461 | 0.010 | 0.020 | 5.6e-01 | -0.030 | 0.050 | 7.3e-01 | 5.6e-01 | 9.4e-01 | - |
| **Substance Use** | | | | | | | | | | | |
| ctrl | woAll | 53592 | -0.100 | 0.005 | 1.1e-159 | -0.100 | -0.100 | 1.1e-159 | - | 6.9e-16 | 8.3e-04 |
| single episode MD | woAll | 12765 | 0.080 | 0.009 | 6.9e-16 | 0.060 | 0.090 | 1.5e-11 | 6.9e-16 | - | 6.3e-01 |
| recurrent episodes MD | woAll | 2461 | 0.070 | 0.020 | 8.3e-04 | 0.030 | 0.100 | 2.9e-04 | 8.3e-04 | 6.3e-01 | - |
| ctrl | woSUD | 43198 | -0.200 | 0.005 | 1.6e-179 | -0.200 | -0.100 | 1.6e-179 | - | 3.5e-36 | 1.6e-08 |
| single episode MD | woSUD | 20595 | 0.100 | 0.008 | 3.5e-36 | 0.090 | 0.100 | 3.9e-14 | 3.5e-36 | - | 1.6e-01 |
| recurrent episodes MD | woSUD | 5025 | 0.080 | 0.010 | 1.6e-08 | 0.050 | 0.100 | 2.4e-08 | 1.6e-08 | 1.6e-01 | - |
| **Substance Use Disorder** | | | | | | | | | | | |
| ctrl | woAll | 53592 | -0.200 | 0.004 | 1.0e-300 | -0.200 | -0.200 | 1.0e-300 | - | 3.8e-06 | 1.2e-01 |
| single episode MD | woAll | 12765 | 0.040 | 0.008 | 3.8e-06 | 0.020 | 0.050 | 6.9e-64 | 3.8e-06 | - | 5.4e-01 |
| recurrent episodes MD | woAll | 2461 | 0.030 | 0.020 | 1.2e-01 | -0.007 | 0.060 | 3.2e-17 | 1.2e-01 | 5.4e-01 | - |
| ctrl | woSUD | 43198 | -0.200 | 0.005 | 1.0e-300 | -0.200 | -0.200 | 1.0e-300 | - | 8.5e-18 | 1.3e-06 |
| single episode MD | woSUD | 20595 | 0.060 | 0.007 | 8.5e-18 | 0.050 | 0.070 | 4.2e-85 | 8.5e-18 | - | 9.8e-01 |
| recurrent episodes MD | woSUD | 5025 | 0.060 | 0.010 | 1.3e-06 | 0.040 | 0.080 | 1.2e-25 | 1.3e-06 | 9.8e-01 | - |

Table S17: Numbers are after relatedness pruning / removal of ancestry outliers. Number in brackets are fraction of MD cases being comorbid with BP, SZ, ANX, ASD, ADHD, SUD or having MD as the only diagnosis or being the fraction of MD cases that do not have BP. Except for population-sample where numbers in brackets indicate phenotype fractions in the population-based iPSYCH2015 sample. All numbers are without BP except for the BP collumn. Whether recurrent depression was associated with increased comorbidity compared to single-episode depression was tested using logistic regression for each additional diagnosis while adjusting for age. Individuals were grouped in age bins in five year intervals in order to construct dummy variables for the age adjustment. Odds-Ratios ($OR = \log(\beta)$) and p-values are shown.

| Phenotype | BP | SZ | ANX | ASD | ADHD | SUD | MD only | MD wo BP | MD total |
|---|---|---|---|---|---|---|---|---|---|
| **MD** | | | | | | | | | |
| No. samples | 1460 | 3905 | 7044 | 2255 | 2571 | 3538 | 15226 | 29158 | 30618 |
| $F(subtype\|MD)$ | (0.0501) | (0.134) | (0.242) | (0.0773) | (0.0882) | (0.121) | (0.522) | (0.95) | - |
| **Single-episode** | | | | | | | | | |
| No. samples | 961 | 2773 | 5104 | 1758 | 1933 | 2545 | 12765 | 23140 | 24101 |
| $F(subtype\|single)$ | (0.0415) | (0.12) | (0.221) | (0.076) | (0.0835) | (0.11) | (0.552) | (0.96) | - |
| **Recurrent** | | | | | | | | | |
| No. samples | 499 | 1132 | 1940 | 497 | 638 | 993 | 2461 | 6018 | 6517 |
| $F(subtype\|recurrent)$ | (0.0829) | (0.188) | (0.322) | (0.0826) | (0.106) | (0.165) | (0.409) | (0.92) | - |
| **Population-sample** | | | | | | | | | |
| No. samples | 106 | 401 | 888 | 636 | 774 | 502 | 556 | 1019 | 1067 |
| $F(subtype\|pop)$ | (0.0027) | (0.01) | (0.023) | (0.016) | (0.02) | (0.013) | (0.014) | (0.026) | (0.027) |
| **Test for difference in comorbidity between singel-episode and recurrent** | | | | | | | | | |
| $OR$ | 1.86 | 1.76 | 1.64 | 1.48 | 1.39 | 1.64 | 0.53 | - | - |
| $p-value$ | 3.1e-27 | 9.6e-48 | 2e-53 | 4.2e-12 | 1.3e-11 | 3.7e-32 | 5.8e-102 | - | - |

Table S18: Hazard rate ratios (HRR) of developing second-episode of depression of PRS decile 2-10 compared to the first decile. Left most column is PRS/subtype (PRS/Phenotype) here a second diagnosis of recurrent depression among cases with depression; PRS decile 2-10 (Category); Harzard Rate Ratio (HRR) by decile 2-10 compared to the first PRS decile estimated by cox-regression; lower 95% confidence limit (CIl) and upper 95% confidence limit (CIu) of HRR; standard error of HRR (SE); two-sided p-value of test for HRR being different from zero.

| PRS/Phenotype | Category | HRR | CIl | CIu | SE | P |
|---|---|---|---|---|---|---|
| **MD** | | | | | | |
| rMD\|MD | 2 | 1.1422 | 1.0127 | 1.2882 | 0.0614 | 3.0e-02 |
| rMD\|MD | 3 | 1.0981 | 0.9723 | 1.2402 | 0.0621 | 1.3e-01 |
| rMD\|MD | 4 | 1.1587 | 1.0246 | 1.3104 | 0.0628 | 1.9e-02 |
| rMD\|MD | 5 | 1.1212 | 0.9928 | 1.2662 | 0.0620 | 6.5e-02 |
| rMD\|MD | 6 | 1.1572 | 1.0282 | 1.3024 | 0.0603 | 1.5e-02 |
| rMD\|MD | 7 | 1.1521 | 1.0212 | 1.2997 | 0.0615 | 2.1e-02 |
| rMD\|MD | 8 | 1.2077 | 1.0718 | 1.3609 | 0.0609 | 1.9e-03 |
| rMD\|MD | 9 | 1.1841 | 1.0504 | 1.3349 | 0.0611 | 5.7e-03 |
| rMD\|MD | 10 | 1.3253 | 1.1788 | 1.4901 | 0.0598 | 2.5e-06 |
| **ANX** | | | | | | |
| rMD\|MD | 2 | 1.0121 | 0.8991 | 1.1393 | 0.0604 | 8.4e-01 |
| rMD\|MD | 3 | 1.0504 | 0.9332 | 1.1822 | 0.0603 | 4.2e-01 |
| rMD\|MD | 4 | 1.0835 | 0.9644 | 1.2173 | 0.0594 | 1.8e-01 |
| rMD\|MD | 5 | 0.9705 | 0.8611 | 1.0938 | 0.0610 | 6.2e-01 |
| rMD\|MD | 6 | 1.1377 | 1.0134 | 1.2772 | 0.0590 | 2.9e-02 |
| rMD\|MD | 7 | 1.1680 | 1.0397 | 1.3121 | 0.0594 | 8.9e-03 |
| rMD\|MD | 8 | 1.0678 | 0.9495 | 1.2008 | 0.0599 | 2.7e-01 |
| rMD\|MD | 9 | 1.1202 | 0.9979 | 1.2575 | 0.0590 | 5.4e-02 |
| rMD\|MD | 10 | 1.1528 | 1.0271 | 1.2939 | 0.0589 | 1.6e-02 |
| **BP** | | | | | | |
| rMD\|MD | 2 | 0.8974 | 0.7850 | 1.0258 | 0.0683 | 1.1e-01 |
| rMD\|MD | 3 | 0.9992 | 0.8859 | 1.1271 | 0.0614 | 9.9e-01 |
| rMD\|MD | 4 | 1.0118 | 0.9041 | 1.1323 | 0.0574 | 8.4e-01 |
| rMD\|MD | 5 | 0.9837 | 0.8797 | 1.0999 | 0.0570 | 7.7e-01 |
| rMD\|MD | 6 | 1.0092 | 0.9040 | 1.1267 | 0.0562 | 8.7e-01 |
| rMD\|MD | 7 | 1.0789 | 0.9661 | 1.2049 | 0.0564 | 1.8e-01 |
| rMD\|MD | 8 | 1.0080 | 0.8970 | 1.1327 | 0.0595 | 8.9e-01 |
| rMD\|MD | 9 | 0.9708 | 0.8550 | 1.1024 | 0.0648 | 6.5e-01 |
| rMD\|MD | 10 | 1.1068 | 0.9931 | 1.2336 | 0.0553 | 6.6e-02 |
| **SZ** | | | | | | |
| rMD\|MD | 2 | 0.9881 | 0.8838 | 1.1048 | 0.0569 | 8.3e-01 |
| rMD\|MD | 3 | 0.9836 | 0.8683 | 1.1143 | 0.0636 | 8.0e-01 |
| rMD\|MD | 4 | 0.9635 | 0.8610 | 1.0782 | 0.0574 | 5.2e-01 |
| rMD\|MD | 5 | 1.0651 | 0.9525 | 1.1909 | 0.0570 | 2.7e-01 |
| rMD\|MD | 6 | 1.0511 | 0.9395 | 1.1761 | 0.0573 | 3.8e-01 |
| rMD\|MD | 7 | 0.9663 | 0.8622 | 1.0830 | 0.0582 | 5.6e-01 |
| rMD\|MD | 8 | 1.0234 | 0.9124 | 1.1480 | 0.0586 | 6.9e-01 |
| rMD\|MD | 9 | 1.0238 | 0.9152 | 1.1453 | 0.0572 | 6.8e-01 |
| rMD\|MD | 10 | 1.0266 | 0.9163 | 1.1501 | 0.0580 | 6.5e-01 |
| **ADHD** | | | | | | |
| rMD\|MD | 2 | 0.9693 | 0.8608 | 1.0916 | 0.0606 | 6.1e-01 |
| rMD\|MD | 3 | 0.9941 | 0.8837 | 1.1183 | 0.0601 | 9.2e-01 |
| rMD\|MD | 4 | 1.0227 | 0.9091 | 1.1505 | 0.0601 | 7.1e-01 |
| rMD\|MD | 5 | 1.0114 | 0.8993 | 1.1374 | 0.0599 | 8.5e-01 |
| rMD\|MD | 6 | 1.0129 | 0.9005 | 1.1392 | 0.0600 | 8.3e-01 |
| rMD\|MD | 7 | 1.0235 | 0.9105 | 1.1505 | 0.0597 | 7.0e-01 |
| rMD\|MD | 8 | 1.0194 | 0.9065 | 1.1463 | 0.0599 | 7.5e-01 |
| rMD\|MD | 9 | 1.0379 | 0.9228 | 1.1674 | 0.0600 | 5.3e-01 |
| rMD\|MD | 10 | 1.0333 | 0.9190 | 1.1617 | 0.0598 | 5.8e-01 |
| **ASD** | | | | | | |

Table S18: Hazard rate ratios (HRR) of developing second-episode of depression of PRS decile 2-10 compared to the first decile. Left most column is PRS/subtype (PRS/Phenotype) here a second diagnosis of recurrent depression among cases with depression; PRS decile 2-10 (Category); Harzard Rate Ratio (HRR) by decile 2-10 compared to the first PRS decile estimated by cox-regression; lower 95% confidence limit (CIl) and upper 95% confidence limit (CIu) of HRR; standard error of HRR (SE); two-sided p-value of test for HRR being different from zero. *(continued)*

| PRS/Phenotype | Category | HRR | CIl | CIu | SE | P |
|---|---|---|---|---|---|---|
| rMD\|MD | 2 | 1.0336 | 0.9210 | 1.1601 | 0.0589 | 5.7e-01 |
| rMD\|MD | 3 | 1.0250 | 0.9139 | 1.1496 | 0.0585 | 6.7e-01 |
| rMD\|MD | 4 | 1.0557 | 0.9414 | 1.1838 | 0.0585 | 3.5e-01 |
| rMD\|MD | 5 | 0.9797 | 0.8717 | 1.1010 | 0.0596 | 7.3e-01 |
| rMD\|MD | 6 | 0.9450 | 0.8398 | 1.0634 | 0.0602 | 3.5e-01 |
| rMD\|MD | 7 | 0.9459 | 0.8419 | 1.0627 | 0.0594 | 3.5e-01 |
| rMD\|MD | 8 | 0.9962 | 0.8871 | 1.1186 | 0.0591 | 9.5e-01 |
| rMD\|MD | 9 | 0.9881 | 0.8792 | 1.1105 | 0.0596 | 8.4e-01 |
| rMD\|MD | 10 | 1.0977 | 0.9797 | 1.2299 | 0.0580 | 1.1e-01 |
| **Neuroticism** | | | | | | |
| rMD\|MD | 2 | 0.9819 | 0.8722 | 1.1054 | 0.0605 | 7.6e-01 |
| rMD\|MD | 3 | 1.0716 | 0.9536 | 1.2042 | 0.0595 | 2.5e-01 |
| rMD\|MD | 4 | 1.0882 | 0.9683 | 1.2231 | 0.0596 | 1.6e-01 |
| rMD\|MD | 5 | 0.9776 | 0.8681 | 1.1008 | 0.0606 | 7.1e-01 |
| rMD\|MD | 6 | 1.0822 | 0.9629 | 1.2164 | 0.0596 | 1.8e-01 |
| rMD\|MD | 7 | 1.0673 | 0.9495 | 1.1996 | 0.0596 | 2.7e-01 |
| rMD\|MD | 8 | 1.1345 | 1.0105 | 1.2736 | 0.0590 | 3.3e-02 |
| rMD\|MD | 9 | 1.0892 | 0.9697 | 1.2235 | 0.0593 | 1.5e-01 |
| rMD\|MD | 10 | 1.1178 | 0.9956 | 1.2551 | 0.0591 | 5.9e-02 |
| **Substance Use** | | | | | | |
| rMD\|MD | 2 | 0.9352 | 0.8335 | 1.0492 | 0.0587 | 2.5e-01 |
| rMD\|MD | 3 | 0.9603 | 0.8564 | 1.0768 | 0.0584 | 4.9e-01 |
| rMD\|MD | 4 | 1.0081 | 0.9004 | 1.1286 | 0.0576 | 8.9e-01 |
| rMD\|MD | 5 | 0.9269 | 0.8259 | 1.0404 | 0.0589 | 2.0e-01 |
| rMD\|MD | 6 | 0.9110 | 0.8115 | 1.0226 | 0.0590 | 1.1e-01 |
| rMD\|MD | 7 | 0.8929 | 0.7948 | 1.0030 | 0.0593 | 5.6e-02 |
| rMD\|MD | 8 | 0.9882 | 0.8822 | 1.1069 | 0.0579 | 8.4e-01 |
| rMD\|MD | 9 | 0.9759 | 0.8710 | 1.0935 | 0.0580 | 6.7e-01 |
| rMD\|MD | 10 | 0.9889 | 0.8830 | 1.1076 | 0.0578 | 8.5e-01 |
| **Substance Use Disorder** | | | | | | |
| rMD\|MD | 2 | 1.0885 | 0.9710 | 1.2202 | 0.0583 | 1.5e-01 |
| rMD\|MD | 3 | 0.9358 | 0.8322 | 1.0522 | 0.0598 | 2.7e-01 |
| rMD\|MD | 4 | 0.9880 | 0.8791 | 1.1104 | 0.0596 | 8.4e-01 |
| rMD\|MD | 5 | 0.9389 | 0.8348 | 1.0561 | 0.0600 | 2.9e-01 |
| rMD\|MD | 6 | 1.0165 | 0.9053 | 1.1415 | 0.0591 | 7.8e-01 |
| rMD\|MD | 7 | 1.1008 | 0.9821 | 1.2339 | 0.0582 | 9.9e-02 |
| rMD\|MD | 8 | 0.9634 | 0.8568 | 1.0832 | 0.0598 | 5.3e-01 |
| rMD\|MD | 9 | 1.0117 | 0.9008 | 1.1362 | 0.0592 | 8.4e-01 |
| rMD\|MD | 10 | 1.0544 | 0.9400 | 1.1827 | 0.0586 | 3.7e-01 |
| **rMDsMD wsumPRS** | | | | | | |
| rMD\|MD | 2 | 1.0713 | 0.9501 | 1.2078 | 0.0612 | 2.6e-01 |
| rMD\|MD | 3 | 1.1563 | 1.0276 | 1.3011 | 0.0602 | 1.6e-02 |
| rMD\|MD | 4 | 1.0636 | 0.9433 | 1.1992 | 0.0612 | 3.1e-01 |
| rMD\|MD | 5 | 1.0836 | 0.9621 | 1.2204 | 0.0607 | 1.9e-01 |
| rMD\|MD | 6 | 1.1549 | 1.0261 | 1.2998 | 0.0603 | 1.7e-02 |
| rMD\|MD | 7 | 1.1141 | 0.9897 | 1.2542 | 0.0604 | 7.4e-02 |
| rMD\|MD | 8 | 1.1928 | 1.0605 | 1.3415 | 0.0599 | 3.3e-03 |
| rMD\|MD | 9 | 1.1831 | 1.0527 | 1.3297 | 0.0596 | 4.8e-03 |
| rMD\|MD | 10 | 1.2804 | 1.1410 | 1.4369 | 0.0588 | 2.7e-05 |

Table S19A: Multivariate PRS analyses of MD vs. ANX. Left-most collumn indicate which trait the training of the PRS is based upon and the sub-phenotype-group within the target sample (iPSYCH2015) the summary statistics is based upon. N = sample size by sub-phenotype in target, $\beta$ = slope of the linear regresssion, $SE$ = stand error, $Preg$ are p-values directly from the regression, ie. first entry is for $H_0 : intercept = 0$, the remaing are for $H_0$ : no difference to reference group ctrl, $CI_l$ and $C_u$ is lower and upper 95% confidence limits, $P_0$ corresponds to $H_0 : \beta + interscept = 0$, except for the control group columns where it is $H_0 : intercept = 0$ as in $P_{reg}$. The remaining $P$-columns holds the p-values for the Wald test of equal group effect: $P_{ctrl}$ is p-value for the control group, while $P_{MDwoANX}$ and $P_{MDwANX}$ are p-values for the Wald test for MD cases without and with an diagnosis of anxiety. Note that all individuals with BP were excluded. Overall p-value =1.2e-21.

| PRS based on: | $N$ | $\beta$ | $SE$ | $P_{reg}$ | $CI_l$ | $CI_u$ | $P_0$ | $P_{ctrl}$ | $P_{MDwoANX}$ | $P_{MDwANX}$ |
|---|---|---|---|---|---|---|---|---|---|---|
| **Major Depression** | | | | | | | | | | |
| ctrl | 38142 | -0.250 | 0.0059 | < 1e-300 | -0.2600 | -0.2400 | < 1e-300 | - | <1e-300 | 2.6e-264 |
| MDwoANX | 22114 | 0.360 | 0.0082 | < 1e-300 | 0.3400 | 0.3700 | 2.8e-54 | <1e-300 | - | 4.9e-10 |
| MDANX | 7044 | 0.440 | 0.0130 | 2.6e-264 | 0.4100 | 0.4600 | 5.8e-59 | 2.6e-264 | 4.9e-10 | - |
| **Anxiety** | | | | | | | | | | |
| ctrl | 38142 | -0.110 | 0.0060 | 9.4e-74 | -0.1200 | -0.0980 | 9.4e-74 | - | 2.0e-63 | 8.4e-81 |
| MDwoANX | 22114 | 0.140 | 0.0085 | 2.0e-63 | 0.1300 | 0.1600 | 7.8e-06 | 2.0e-63 | - | 1.8e-14 |
| MDANX | 7044 | 0.250 | 0.0130 | 8.4e-81 | 0.2200 | 0.2700 | 2.5e-29 | 8.4e-81 | 1.8e-14 | - |
| **Bipolar Disorder** | | | | | | | | | | |
| ctrl | 38142 | -0.190 | 0.0055 | 5.9e-262 | -0.2000 | -0.1800 | 5.9e-262 | - | 5.9e-68 | 2.9e-40 |
| MDwoANX | 22114 | 0.130 | 0.0077 | 5.9e-68 | 0.1200 | 0.1500 | 8.4e-18 | 5.9e-68 | - | 6.9e-02 |
| MDANX | 7044 | 0.160 | 0.0120 | 2.9e-40 | 0.1300 | 0.1800 | 2.1e-03 | 2.9e-40 | 6.9e-02 | - |
| **Schizophrenia** | | | | | | | | | | |
| ctrl | 38142 | -0.190 | 0.0056 | 1.3e-242 | -0.2000 | -0.1700 | 1.3e-242 | - | 8.9e-50 | 1.2e-43 |
| MDwoANX | 22114 | 0.120 | 0.0078 | 8.9e-50 | 0.1000 | 0.1300 | 7.9e-26 | 8.9e-50 | - | 7.4e-05 |
| MDANX | 7044 | 0.170 | 0.0120 | 1.2e-43 | 0.1400 | 0.1900 | 7.0e-02 | 1.2e-43 | 7.4e-05 | - |
| **ADHD** | | | | | | | | | | |
| ctrl | 38142 | -0.240 | 0.0059 | < 1e-300 | -0.2500 | -0.2300 | < 1e-300 | - | 5.6e-155 | 1.3e-67 |
| MDwoANX | 22114 | 0.220 | 0.0083 | 5.6e-155 | 0.2000 | 0.2400 | 1.1e-02 | 5.6e-155 | - | 9.6e-01 |
| MDANX | 7044 | 0.220 | 0.0130 | 1.3e-67 | 0.2000 | 0.2500 | 1.5e-01 | 1.3e-67 | 9.6e-01 | - |
| **Autism** | | | | | | | | | | |
| ctrl | 38142 | -0.016 | 0.0060 | 6.6e-03 | -0.0280 | -0.0046 | 6.6e-03 | - | 5.9e-03 | 2.4e-05 |
| MDwoANX | 22114 | 0.023 | 0.0084 | 5.9e-03 | 0.0067 | 0.0400 | 3.4e-01 | 5.9e-03 | - | 2.1e-02 |
| MDANX | 7044 | 0.055 | 0.0130 | 2.4e-05 | 0.0290 | 0.0800 | 1.7e-03 | 2.4e-05 | 2.1e-02 | - |
| **Neuroticism** | | | | | | | | | | |
| ctrl | 38142 | -0.120 | 0.0060 | 6.4e-84 | -0.1300 | -0.1000 | 6.4e-84 | - | 1.5e-141 | 1.8e-130 |
| MDwoANX | 22114 | 0.210 | 0.0083 | 1.5e-141 | 0.2000 | 0.2300 | 6.0e-41 | 1.5e-141 | - | 1.5e-13 |
| MDANX | 7044 | 0.310 | 0.0130 | 1.8e-130 | 0.2900 | 0.3400 | 2.7e-59 | 1.8e-130 | 1.5e-13 | - |
| **Substance Use** | | | | | | | | | | |
| ctrl | 38142 | -0.210 | 0.0057 | 1.2e-280 | -0.2200 | -0.2000 | 1.2e-280 | - | 1.7e-121 | 6.1e-45 |
| MDwoANX | 22114 | 0.190 | 0.0080 | 1.7e-121 | 0.1700 | 0.2000 | 1.0e-02 | 1.7e-121 | - | 2.4e-01 |
| MDANX | 7044 | 0.170 | 0.0120 | 6.1e-45 | 0.1500 | 0.2000 | 4.3e-03 | 6.1e-45 | 2.4e-01 | - |
| **Substance Use Disorder** | | | | | | | | | | |
| ctrl | 38142 | -0.210 | 0.0050 | < 1e-300 | -0.2200 | -0.2000 | < 1e-300 | - | 5.5e-48 | 2.1e-28 |
| MDwoANX | 22114 | 0.100 | 0.0070 | 5.5e-48 | 0.0880 | 0.1200 | 1.0e-72 | 5.5e-48 | - | 1.4e-01 |
| MDANX | 7044 | 0.120 | 0.0110 | 2.1e-28 | 0.0970 | 0.1400 | 1.0e-19 | 2.1e-28 | 1.4e-01 | - |

Table S19B: Multivariate PRS analyses of MD vs. BP. Left-most collumn indicate which trait the training of the PRS is based upon and the sub-phenotype-group within the target sample (iPSYCH2015) the summary statistics is based upon. N = sample size by sub-phenotype in target, $\beta$ = slope of the linear regresssion, $SE$ = stand error, $Preg$ are p-values directly from the regression, ie. first entry is for $H_0 : intercept = 0$, the remaing are for $H_0$ : no difference to reference group ctrl, $CI_l$ and $C_u$ is lower and upper 95% confidence limits, $P_0$ corresponds to $H_0 : \beta + interscept = 0$, except for the control group columns where it is $H_0 : intercept = 0$ as in $P_{reg}$. The remaining $P$-columns holds the p-values for the Wald test of equal group effect: $P_{ctrl}$ is p-value for the control group, while $P_{MDwoBP}$ and $P_{MDwBP}$ are p-values for the Wald test for MD cases without and with an diagnosis of BP. Overall p-value =1.5e-15.

| PRS based on: | $N$ | $\beta$ | $SE$ | $P_{reg}$ | $CI_l$ | $CI_u$ | $P_0$ | $P_{ctrl}$ | $P_{MDwoBP}$ | $P_{MDwBP}$ |
|---|---|---|---|---|---|---|---|---|---|---|
| **Major Depression** | | | | | | | | | | |
| ctrl | 38200 | -0.250 | 0.0058 | < 1e-300 | -0.2600 | -0.2400 | < 1e-300 | - | <1e-300 | 3.8e-76 |
| MDwoBP | 29158 | 0.380 | 0.0076 | < 1e-300 | 0.3600 | 0.3900 | 4.6e-95 | <1e-300 | - | 8.6e-05 |
| MDBP | 1460 | 0.480 | 0.0260 | 3.8e-76 | 0.4300 | 0.5300 | 9.9e-20 | 3.8e-76 | 8.6e-05 | - |
| **Anxiety** | | | | | | | | | | |
| ctrl | 38200 | -0.110 | 0.0060 | 2.4e-72 | -0.1200 | -0.0970 | 2.4e-72 | - | 5.0e-102 | 6.9e-14 |
| MDwoBP | 29158 | 0.170 | 0.0078 | 5.0e-102 | 0.1500 | 0.1800 | 7.4e-20 | 5.0e-102 | - | 2.3e-01 |
| MDBP | 1460 | 0.200 | 0.0270 | 6.9e-14 | 0.1500 | 0.2500 | 5.2e-04 | 6.9e-14 | 2.3e-01 | - |
| **Bipolar Disorder** | | | | | | | | | | |
| ctrl | 38200 | -0.190 | 0.0055 | 5.3e-262 | -0.2000 | -0.1800 | 5.3e-262 | - | 2.1e-85 | 2.4e-45 |
| MDwoBP | 29158 | 0.140 | 0.0071 | 2.1e-85 | 0.1300 | 0.1500 | 2.0e-18 | 2.1e-85 | - | 6.1e-17 |
| MDBP | 1460 | 0.340 | 0.0240 | 2.4e-45 | 0.3000 | 0.3900 | 1.7e-10 | 2.4e-45 | 6.1e-17 | - |
| **Schizophrenia** | | | | | | | | | | |
| ctrl | 38200 | -0.190 | 0.0055 | 2.6e-243 | -0.2000 | -0.1700 | 2.6e-243 | - | 1.6e-70 | 4.3e-34 |
| MDwoBP | 29158 | 0.130 | 0.0072 | 1.6e-70 | 0.1100 | 0.1400 | 1.4e-22 | 1.6e-70 | - | 3.7e-12 |
| MDBP | 1460 | 0.300 | 0.0240 | 4.3e-34 | 0.2500 | 0.3500 | 2.8e-06 | 4.3e-34 | 3.7e-12 | - |
| **ADHD** | | | | | | | | | | |
| ctrl | 38200 | -0.240 | 0.0059 | < 1e-300 | -0.2500 | -0.2300 | < 1e-300 | - | 3.9e-182 | 4.2e-16 |
| MDwoBP | 29158 | 0.220 | 0.0076 | 3.9e-182 | 0.2100 | 0.2400 | 6.4e-03 | 3.9e-182 | - | 7.6e-01 |
| MDBP | 1460 | 0.210 | 0.0260 | 4.2e-16 | 0.1600 | 0.2600 | 3.3e-01 | 4.2e-16 | 7.6e-01 | - |
| **Autism** | | | | | | | | | | |
| ctrl | 38200 | -0.017 | 0.0060 | 5.0e-03 | -0.0290 | -0.0051 | 5.0e-03 | - | 6.6e-05 | 6.7e-02 |
| MDwoBP | 29158 | 0.031 | 0.0078 | 6.6e-05 | 0.0160 | 0.0460 | 2.8e-02 | 6.6e-05 | - | 5.1e-01 |
| MDBP | 1460 | 0.049 | 0.0270 | 6.7e-02 | -0.0035 | 0.1000 | 2.3e-01 | 6.7e-02 | 5.1e-01 | - |
| **Neuroticism** | | | | | | | | | | |
| ctrl | 38200 | -0.120 | 0.0059 | 1.9e-84 | -0.1300 | -0.1000 | 1.9e-84 | - | 5.3e-206 | 1.2e-24 |
| MDwoBP | 29158 | 0.240 | 0.0077 | 5.3e-206 | 0.2200 | 0.2500 | 1.6e-79 | 5.3e-206 | - | 2.1e-01 |
| MDBP | 1460 | 0.270 | 0.0260 | 1.2e-24 | 0.2200 | 0.3200 | 3.0e-09 | 1.2e-24 | 2.1e-01 | - |
| **Substance Use** | | | | | | | | | | |
| ctrl | 38200 | -0.210 | 0.0057 | 2.6e-282 | -0.2200 | -0.2000 | 2.6e-282 | - | 2.9e-137 | 1.7e-18 |
| MDwoBP | 29158 | 0.180 | 0.0074 | 2.9e-137 | 0.1700 | 0.2000 | 4.4e-04 | 2.9e-137 | - | 1.4e-01 |
| MDBP | 1460 | 0.220 | 0.0250 | 1.7e-18 | 0.1700 | 0.2700 | 5.3e-01 | 1.7e-18 | 1.4e-01 | - |
| **Substance Use Disorder** | | | | | | | | | | |
| ctrl | 38200 | -0.210 | 0.0050 | < 1e-300 | -0.2200 | -0.2000 | < 1e-300 | - | 1.3e-60 | 8.4e-08 |
| MDwoBP | 29158 | 0.110 | 0.0064 | 1.3e-60 | 0.0930 | 0.1200 | 1.6e-84 | 1.3e-60 | - | 5.8e-01 |
| MDBP | 1460 | 0.120 | 0.0220 | 8.4e-08 | 0.0750 | 0.1600 | 2.4e-05 | 8.4e-08 | 5.8e-01 | - |

Table S19C: Multivariate PRS analyses of MD vs. SZ. Left-most collumn indicate which trait the training of the PRS is based upon and the sub-phenotype-group within the target sample (iPSYCH2015) the summary statistics is based upon. N = sample size by sub-phenotype in target, $\beta$ = slope of the linear regresssion, $SE$ = stand error, $Preg$ are p-values directly from the regression, ie. first entry is for $H_0 : intercept = 0$, the remaing are for $H_0$ : no difference to reference group ctrl, $CI_l$ and $C_u$ is lower and upper 95% confidence limits, $P_0$ corresponds to $H_0 : \beta + interscept = 0$, except for the control group columns where it is $H_0 : intercept = 0$ as in $P_{reg}$. The remaining $P$-columns holds the p-values for the Wald test of equal group effect: $P_{ctrl}$ is p-value for the control group, while $P_{MDwoSZ}$ and $P_{MDwSZ}$ are p-values for the Wald test for MD cases without and with an diagnosis of SZ. Note that all individuals with BP were excluded. Overall p-value =1.7e-15.

| PRS based on: | $N$ | $\beta$ | $SE$ | $P_{reg}$ | $CI_l$ | $CI_u$ | $P_0$ | $P_{ctrl}$ | $P_{MDwoSZ}$ | $P_{MDwSZ}$ |
|---|---|---|---|---|---|---|---|---|---|---|
| **Major Depression** | | | | | | | | | | |
| ctrl | 38142 | -0.250 | 0.0059 | < 1e-300 | -0.2600 | -0.2400 | < 1e-300 | - | <1e-300 | 2.0e-158 |
| MDwoSZ | 25253 | 0.370 | 0.0079 | < 1e-300 | 0.3500 | 0.3800 | 4.7e-73 | <1e-300 | - | 1.9e-05 |
| MDSZ | 3905 | 0.440 | 0.0160 | 2.0e-158 | 0.4100 | 0.4700 | 9.3e-34 | 2.0e-158 | 1.9e-05 | - |
| **Anxiety** | | | | | | | | | | |
| ctrl | 38142 | -0.110 | 0.0060 | 1.8e-73 | -0.1200 | -0.0980 | 1.8e-73 | - | 2.1e-91 | 4.2e-28 |
| MDwoSZ | 25253 | 0.160 | 0.0081 | 2.1e-91 | 0.1500 | 0.1800 | 6.3e-16 | 2.1e-91 | - | 2.5e-01 |
| MDSZ | 3905 | 0.180 | 0.0170 | 4.2e-28 | 0.1500 | 0.2200 | 4.2e-06 | 4.2e-28 | 2.5e-01 | - |
| **Bipolar Disorder** | | | | | | | | | | |
| ctrl | 38142 | -0.190 | 0.0055 | 4.9e-261 | -0.2000 | -0.1800 | 4.9e-261 | - | 4.4e-67 | 7.9e-45 |
| MDwoSZ | 25253 | 0.130 | 0.0074 | 4.4e-67 | 0.1100 | 0.1400 | 8.0e-24 | 4.4e-67 | - | 3.0e-08 |
| MDSZ | 3905 | 0.210 | 0.0150 | 7.9e-45 | 0.1800 | 0.2400 | 1.0e-01 | 7.9e-45 | 3.0e-08 | - |
| **Schizophrenia** | | | | | | | | | | |
| ctrl | 38142 | -0.180 | 0.0056 | 2.3e-241 | -0.2000 | -0.1700 | 2.3e-241 | - | 5.7e-50 | 1.6e-51 |
| MDwoSZ | 25253 | 0.110 | 0.0075 | 5.7e-50 | 0.0960 | 0.1300 | 5.3e-32 | 5.7e-50 | - | 1.1e-14 |
| MDSZ | 3905 | 0.230 | 0.0150 | 1.6e-51 | 0.2000 | 0.2600 | 1.3e-03 | 1.6e-51 | 1.1e-14 | - |
| **ADHD** | | | | | | | | | | |
| ctrl | 38142 | -0.240 | 0.0059 | < 1e-300 | -0.2500 | -0.2300 | < 1e-300 | - | 1.5e-157 | 7.3e-59 |
| MDwoSZ | 25253 | 0.210 | 0.0080 | 1.5e-157 | 0.2000 | 0.2300 | 2.2e-04 | 1.5e-157 | - | 1.7e-03 |
| MDSZ | 3905 | 0.270 | 0.0160 | 7.3e-59 | 0.2300 | 0.3000 | 7.7e-02 | 7.3e-59 | 1.7e-03 | - |
| **Autism** | | | | | | | | | | |
| ctrl | 38142 | -0.016 | 0.0060 | 7.3e-03 | -0.0280 | -0.0044 | 7.3e-03 | - | 2.2e-03 | 3.4e-05 |
| MDwoSZ | 25253 | 0.025 | 0.0081 | 2.2e-03 | 0.0089 | 0.0410 | 2.0e-01 | 2.2e-03 | - | 9.2e-03 |
| MDSZ | 3905 | 0.069 | 0.0170 | 3.4e-05 | 0.0370 | 0.1000 | 1.0e-03 | 3.4e-05 | 9.2e-03 | - |
| **Neuroticism** | | | | | | | | | | |
| ctrl | 38142 | -0.120 | 0.0060 | 1.7e-83 | -0.1300 | -0.1000 | 1.7e-83 | - | 6.1e-181 | 1.4e-59 |
| MDwoSZ | 25253 | 0.230 | 0.0080 | 6.1e-181 | 0.2100 | 0.2500 | 4.2e-65 | 6.1e-181 | - | 2.1e-02 |
| MDSZ | 3905 | 0.270 | 0.0170 | 1.4e-59 | 0.2400 | 0.3000 | 7.8e-22 | 1.4e-59 | 2.1e-02 | - |
| **Substance Use** | | | | | | | | | | |
| ctrl | 38142 | -0.210 | 0.0057 | 5.6e-280 | -0.2200 | -0.2000 | 5.6e-280 | - | 3.2e-114 | 1.8e-53 |
| MDwoSZ | 25253 | 0.180 | 0.0077 | 3.2e-114 | 0.1600 | 0.1900 | 2.4e-06 | 3.2e-114 | - | 1.8e-05 |
| MDSZ | 3905 | 0.250 | 0.0160 | 1.8e-53 | 0.2100 | 0.2800 | 1.1e-02 | 1.8e-53 | 1.8e-05 | - |
| **Substance Use Disorder** | | | | | | | | | | |
| ctrl | 38142 | -0.210 | 0.0050 | < 1e-300 | -0.2200 | -0.2000 | < 1e-300 | - | 9.6e-50 | 7.5e-26 |
| MDwoSZ | 25253 | 0.099 | 0.0067 | 9.6e-50 | 0.0860 | 0.1100 | 1.1e-84 | 9.6e-50 | - | 1.1e-03 |
| MDSZ | 3905 | 0.150 | 0.0140 | 7.5e-26 | 0.1200 | 0.1700 | 1.9e-06 | 7.5e-26 | 1.1e-03 | - |

Table S19D: Multivariate PRS analyses of MD vs. SUD. Left-most collumn indicate which trait the training of the PRS is based upon and the sub-phenotype-group within the target sample (iPSYCH2015) the summary statistics is based upon. N = sample size by sub-phenotype in target, $\beta$ = slope of the linear regresssion, $SE$ = stand error, $Preg$ are p-values directly from the regression, ie. first entry is for $H_0 : intercept = 0$, the remaing are for $H_0$ : no difference to reference group ctrl, $CI_l$ and $C_u$ is lower and upper 95% confidence limits, $P_0$ corresponds to $H_0 : \beta + interscept = 0$, except for the control group columns where it is $H_0 : intercept = 0$ as in $P_{reg}$. The remaining $P$-columns holds the p-values for the Wald test of equal group effect: $P_{ctrl}$ is p-value for the control group, while $P_{MDwoSUD}$ and $P_{MDwSUD}$ are p-values for the Wald test for MD cases without and with an diagnosis of SUD. Note that all individuals with BP were excluded. Overall p-value =6.4e-97.

| PRS based on: | $N$ | $\beta$ | $SE$ | $P_{reg}$ | $CI_l$ | $CI_u$ | $P_0$ | $P_{ctrl}$ | $P_{MDwoSUD}$ | $P_{MDwSUD}$ |
|---|---|---|---|---|---|---|---|---|---|---|
| **Major Depression** | | | | | | | | | | |
| ctrl | 38142 | -0.2500 | 0.0059 | $<$ 1e-300 | -0.260 | -0.2400 | $<$ 1e-300 | - | <1e-300 | 2.1e-182 |
| MDwoSUD | 25620 | 0.3600 | 0.0079 | $<$ 1e-300 | 0.350 | 0.3800 | 1.5e-64 | <1e-300 | - | 2.7e-14 |
| MDSUD | 3538 | 0.4900 | 0.0170 | 2.1e-182 | 0.460 | 0.5300 | 3.5e-50 | 2.1e-182 | 2.7e-14 | - |
| **Anxiety** | | | | | | | | | | |
| ctrl | 38142 | -0.1100 | 0.0060 | 2.1e-74 | -0.120 | -0.0990 | 2.1e-74 | - | 1.6e-80 | 1.2e-52 |
| MDwoSUD | 25620 | 0.1500 | 0.0081 | 1.6e-80 | 0.140 | 0.1700 | 2.3e-10 | 1.6e-80 | - | 1.4e-10 |
| MDSUD | 3538 | 0.2700 | 0.0180 | 1.2e-52 | 0.230 | 0.3000 | 9.6e-21 | 1.2e-52 | 1.4e-10 | - |
| **Bipolar Disorder** | | | | | | | | | | |
| ctrl | 38142 | -0.1900 | 0.0055 | 5.0e-263 | -0.200 | -0.1800 | 5.0e-263 | - | 2.5e-70 | 4.2e-38 |
| MDwoSUD | 25620 | 0.1300 | 0.0074 | 2.5e-70 | 0.120 | 0.1400 | 1.4e-22 | 2.5e-70 | - | 3.1e-06 |
| MDSUD | 3538 | 0.2100 | 0.0160 | 4.2e-38 | 0.180 | 0.2400 | 3.2e-01 | 4.2e-38 | 3.1e-06 | - |
| **Schizophrenia** | | | | | | | | | | |
| ctrl | 38142 | -0.1900 | 0.0056 | 1.6e-244 | -0.200 | -0.1800 | 1.6e-244 | - | 1.6e-50 | 1.2e-52 |
| MDwoSUD | 25620 | 0.1100 | 0.0074 | 1.6e-50 | 0.097 | 0.1300 | 6.3e-33 | 1.6e-50 | - | 1.7e-16 |
| MDSUD | 3538 | 0.2500 | 0.0160 | 1.2e-52 | 0.220 | 0.2800 | 9.9e-05 | 1.2e-52 | 1.7e-16 | - |
| **ADHD** | | | | | | | | | | |
| ctrl | 38142 | -0.2400 | 0.0059 | $<$ 1e-300 | -0.250 | -0.2300 | $<$ 1e-300 | - | 2.4e-134 | 1.3e-120 |
| MDwoSUD | 25620 | 0.2000 | 0.0079 | 2.4e-134 | 0.180 | 0.2100 | 5.3e-11 | 2.4e-134 | - | 3.9e-32 |
| MDSUD | 3538 | 0.4000 | 0.0170 | 1.3e-120 | 0.370 | 0.4400 | 7.9e-23 | 1.3e-120 | 3.9e-32 | - |
| **Autism** | | | | | | | | | | |
| ctrl | 38142 | -0.0160 | 0.0060 | 7.3e-03 | -0.028 | -0.0044 | 7.3e-03 | - | 1.6e-05 | 9.0e-01 |
| MDwoSUD | 25620 | 0.0350 | 0.0081 | 1.6e-05 | 0.019 | 0.0510 | 6.5e-03 | 1.6e-05 | - | 6.9e-02 |
| MDSUD | 3538 | 0.0022 | 0.0180 | 9.0e-01 | -0.032 | 0.0370 | 4.1e-01 | 9.0e-01 | 6.9e-02 | - |
| **Neuroticism** | | | | | | | | | | |
| ctrl | 38142 | -0.1200 | 0.0060 | 5.1e-84 | -0.130 | -0.1000 | 5.1e-84 | - | 1.1e-183 | 1.5e-54 |
| MDwoSUD | 25620 | 0.2300 | 0.0080 | 1.1e-183 | 0.220 | 0.2500 | 3.0e-65 | 1.1e-183 | - | 2.8e-02 |
| MDSUD | 3538 | 0.2700 | 0.0170 | 1.5e-54 | 0.240 | 0.3000 | 3.1e-20 | 1.5e-54 | 2.8e-02 | - |
| **Substance Use** | | | | | | | | | | |
| ctrl | 38142 | -0.2100 | 0.0057 | 4.1e-287 | -0.220 | -0.2000 | 4.1e-287 | - | 1.4e-79 | 1.5e-180 |
| MDwoSUD | 25620 | 0.1400 | 0.0077 | 1.4e-79 | 0.130 | 0.1600 | 1.1e-22 | 1.4e-79 | - | 7.5e-86 |
| MDSUD | 3538 | 0.4800 | 0.0170 | 1.5e-180 | 0.450 | 0.5100 | 1.6e-63 | 1.5e-180 | 7.5e-86 | - |
| **Substance Use Disorder** | | | | | | | | | | |
| ctrl | 38142 | -0.2100 | 0.0050 | $<$ 1e-300 | -0.220 | -0.2000 | $<$ 1e-300 | - | 1.9e-37 | 4.8e-69 |
| MDwoSUD | 25620 | 0.0850 | 0.0066 | 1.9e-37 | 0.072 | 0.0980 | 2.8e-109 | 1.9e-37 | - | 1.8e-30 |
| MDSUD | 3538 | 0.2500 | 0.0140 | 4.8e-69 | 0.230 | 0.2800 | 1.6e-03 | 4.8e-69 | 1.8e-30 | - |

Table S20A: Hazard rate ratios (HRR) of developing anxiety among MD cases of PRS decile 2-10 compared to the first decile. Left most column is PRS/subtype (PRS/Phenotype) here a second diagnosis of recurrent depression among cases with depression; PRS decile 2-10 (Category); Harzard Rate Ratio (HRR) by decile 2-10 compared to the first PRS decile estimated by cox-regression; lower 95% confidence limit (CIl) and upper 95% confidence limit (CIu) of HRR; standard error of HRR (SE); two-sided p-value of test for HRR being different from zero.

| PRS/Phenotype | Category | HRR | CIl | CIu | SE | P |
|---|---|---|---|---|---|---|
| **MD** | | | | | | |
| ANX\|MD | 2 | 1.0745 | 0.9044 | 1.2764 | 0.0879 | 4.1e-01 |
| ANX\|MD | 3 | 1.0518 | 0.8840 | 1.2513 | 0.0886 | 5.7e-01 |
| ANX\|MD | 4 | 1.0853 | 0.9088 | 1.2962 | 0.0906 | 3.7e-01 |
| ANX\|MD | 5 | 1.1179 | 0.9419 | 1.3267 | 0.0874 | 2.0e-01 |
| ANX\|MD | 6 | 1.1727 | 0.9915 | 1.3871 | 0.0856 | 6.3e-02 |
| ANX\|MD | 7 | 1.3104 | 1.1092 | 1.5480 | 0.0850 | 1.5e-03 |
| ANX\|MD | 8 | 1.1530 | 0.9725 | 1.3671 | 0.0869 | 1.0e-01 |
| ANX\|MD | 9 | 1.3087 | 1.1078 | 1.5461 | 0.0850 | 1.6e-03 |
| ANX\|MD | 10 | 1.2890 | 1.0921 | 1.5214 | 0.0846 | 2.7e-03 |
| **ANX** | | | | | | |
| ANX\|MD | 2 | 0.9351 | 0.7875 | 1.1102 | 0.0876 | 4.4e-01 |
| ANX\|MD | 3 | 0.9451 | 0.7959 | 1.1222 | 0.0876 | 5.2e-01 |
| ANX\|MD | 4 | 1.0200 | 0.8621 | 1.2067 | 0.0858 | 8.2e-01 |
| ANX\|MD | 5 | 1.0718 | 0.9072 | 1.2664 | 0.0851 | 4.1e-01 |
| ANX\|MD | 6 | 1.1194 | 0.9498 | 1.3192 | 0.0838 | 1.8e-01 |
| ANX\|MD | 7 | 1.0766 | 0.9116 | 1.2714 | 0.0849 | 3.8e-01 |
| ANX\|MD | 8 | 1.1812 | 1.0030 | 1.3910 | 0.0834 | 4.6e-02 |
| ANX\|MD | 9 | 1.2006 | 1.0212 | 1.4115 | 0.0826 | 2.7e-02 |
| ANX\|MD | 10 | 1.3083 | 1.1151 | 1.5349 | 0.0815 | 9.8e-04 |
| **BP** | | | | | | |
| ANX\|MD | 2 | 1.0284 | 0.8539 | 1.2386 | 0.0949 | 7.7e-01 |
| ANX\|MD | 3 | 1.1525 | 0.9740 | 1.3636 | 0.0858 | 9.8e-02 |
| ANX\|MD | 4 | 1.1102 | 0.9465 | 1.3021 | 0.0814 | 2.0e-01 |
| ANX\|MD | 5 | 1.0812 | 0.9224 | 1.2672 | 0.0810 | 3.4e-01 |
| ANX\|MD | 6 | 1.0111 | 0.8624 | 1.1855 | 0.0812 | 8.9e-01 |
| ANX\|MD | 7 | 1.0203 | 0.8686 | 1.1984 | 0.0821 | 8.1e-01 |
| ANX\|MD | 8 | 1.0656 | 0.9014 | 1.2597 | 0.0854 | 4.6e-01 |
| ANX\|MD | 9 | 1.0973 | 0.9192 | 1.3099 | 0.0904 | 3.0e-01 |
| ANX\|MD | 10 | 1.1449 | 0.9808 | 1.3363 | 0.0789 | 8.6e-02 |
| **SZ** | | | | | | |
| ANX\|MD | 2 | 1.0105 | 0.8611 | 1.1858 | 0.0816 | 9.0e-01 |
| ANX\|MD | 3 | 1.1028 | 0.9263 | 1.3128 | 0.0890 | 2.7e-01 |
| ANX\|MD | 4 | 0.9418 | 0.8003 | 1.1083 | 0.0831 | 4.7e-01 |
| ANX\|MD | 5 | 1.0357 | 0.8806 | 1.2180 | 0.0827 | 6.7e-01 |
| ANX\|MD | 6 | 1.1517 | 0.9823 | 1.3503 | 0.0811 | 8.2e-02 |
| ANX\|MD | 7 | 0.9998 | 0.8465 | 1.1808 | 0.0849 | 1.0e+00 |
| ANX\|MD | 8 | 1.0092 | 0.8578 | 1.1873 | 0.0829 | 9.1e-01 |
| ANX\|MD | 9 | 1.0724 | 0.9137 | 1.2586 | 0.0817 | 3.9e-01 |
| ANX\|MD | 10 | 1.1934 | 1.0191 | 1.3975 | 0.0805 | 2.8e-02 |
| **ADHD** | | | | | | |
| ANX\|MD | 2 | 0.9529 | 0.8099 | 1.1212 | 0.0830 | 5.6e-01 |
| ANX\|MD | 3 | 0.9038 | 0.7670 | 1.0649 | 0.0837 | 2.3e-01 |
| ANX\|MD | 4 | 0.9299 | 0.7899 | 1.0948 | 0.0833 | 3.8e-01 |
| ANX\|MD | 5 | 0.9516 | 0.8094 | 1.1189 | 0.0826 | 5.5e-01 |
| ANX\|MD | 6 | 1.0263 | 0.8744 | 1.2045 | 0.0817 | 7.5e-01 |
| ANX\|MD | 7 | 0.9539 | 0.8114 | 1.1214 | 0.0825 | 5.7e-01 |
| ANX\|MD | 8 | 0.9690 | 0.8243 | 1.1391 | 0.0825 | 7.0e-01 |
| ANX\|MD | 9 | 0.9592 | 0.8158 | 1.1279 | 0.0826 | 6.1e-01 |
| ANX\|MD | 10 | 1.0211 | 0.8705 | 1.1978 | 0.0814 | 8.0e-01 |
| **ASD** | | | | | | |
| ANX\|MD | 2 | 0.9798 | 0.8291 | 1.1577 | 0.0852 | 8.1e-01 |

Table S20A: Hazard rate ratios (HRR) of developing anxiety among MD cases of PRS decile 2-10 compared to the first decile. Left most column is PRS/subtype (PRS/Phenotype) here a second diagnosis of recurrent depression among cases with depression; PRS decile 2-10 (Category); Harzard Rate Ratio (HRR) by decile 2-10 compared to the first PRS decile estimated by cox-regression; lower 95% confidence limit (CIl) and upper 95% confidence limit (CIu) of HRR; standard error of HRR (SE); two-sided p-value of test for HRR being different from zero. *(continued)*

| PRS/Phenotype | Category | HRR | CIl | CIu | SE | P |
|---|---|---|---|---|---|---|
| ANX\|MD | 3 | 1.0260 | 0.8696 | 1.2105 | 0.0844 | 7.6e-01 |
| ANX\|MD | 4 | 0.9872 | 0.8367 | 1.1648 | 0.0844 | 8.8e-01 |
| ANX\|MD | 5 | 1.0987 | 0.9353 | 1.2907 | 0.0822 | 2.5e-01 |
| ANX\|MD | 6 | 1.0013 | 0.8481 | 1.1822 | 0.0847 | 9.9e-01 |
| ANX\|MD | 7 | 0.9663 | 0.8177 | 1.1418 | 0.0852 | 6.9e-01 |
| ANX\|MD | 8 | 0.9510 | 0.8042 | 1.1247 | 0.0856 | 5.6e-01 |
| ANX\|MD | 9 | 1.1431 | 0.9734 | 1.3425 | 0.0820 | 1.0e-01 |
| ANX\|MD | 10 | 1.1276 | 0.9595 | 1.3252 | 0.0824 | 1.4e-01 |
| **Neuroticism** | | | | | | |
| ANX\|MD | 2 | 0.9939 | 0.8337 | 1.1848 | 0.0897 | 9.5e-01 |
| ANX\|MD | 3 | 1.1206 | 0.9441 | 1.3300 | 0.0874 | 1.9e-01 |
| ANX\|MD | 4 | 1.0997 | 0.9253 | 1.3069 | 0.0881 | 2.8e-01 |
| ANX\|MD | 5 | 1.0663 | 0.8969 | 1.2677 | 0.0883 | 4.7e-01 |
| ANX\|MD | 6 | 1.2485 | 1.0546 | 1.4781 | 0.0861 | 1.0e-02 |
| ANX\|MD | 7 | 1.1971 | 1.0111 | 1.4172 | 0.0861 | 3.7e-02 |
| ANX\|MD | 8 | 1.2519 | 1.0586 | 1.4805 | 0.0856 | 8.7e-03 |
| ANX\|MD | 9 | 1.3494 | 1.1441 | 1.5915 | 0.0842 | 3.7e-04 |
| ANX\|MD | 10 | 1.3875 | 1.1772 | 1.6353 | 0.0839 | 9.4e-05 |
| **Substance Use** | | | | | | |
| ANX\|MD | 2 | 0.8897 | 0.7566 | 1.0461 | 0.0827 | 1.6e-01 |
| ANX\|MD | 3 | 0.9403 | 0.8011 | 1.1037 | 0.0818 | 4.5e-01 |
| ANX\|MD | 4 | 0.8540 | 0.7255 | 1.0051 | 0.0831 | 5.8e-02 |
| ANX\|MD | 5 | 0.8833 | 0.7515 | 1.0382 | 0.0824 | 1.3e-01 |
| ANX\|MD | 6 | 0.9242 | 0.7874 | 1.0848 | 0.0817 | 3.3e-01 |
| ANX\|MD | 7 | 0.9296 | 0.7918 | 1.0913 | 0.0818 | 3.7e-01 |
| ANX\|MD | 8 | 0.8914 | 0.7586 | 1.0475 | 0.0823 | 1.6e-01 |
| ANX\|MD | 9 | 0.9309 | 0.7934 | 1.0922 | 0.0815 | 3.8e-01 |
| ANX\|MD | 10 | 1.0203 | 0.8723 | 1.1933 | 0.0799 | 8.0e-01 |
| **Substance Use Disorder** | | | | | | |
| ANX\|MD | 2 | 1.1379 | 0.9632 | 1.3443 | 0.0850 | 1.3e-01 |
| ANX\|MD | 3 | 1.0688 | 0.9026 | 1.2657 | 0.0863 | 4.4e-01 |
| ANX\|MD | 4 | 1.0177 | 0.8576 | 1.2078 | 0.0874 | 8.4e-01 |
| ANX\|MD | 5 | 1.1123 | 0.9410 | 1.3149 | 0.0853 | 2.1e-01 |
| ANX\|MD | 6 | 1.1420 | 0.9673 | 1.3481 | 0.0847 | 1.2e-01 |
| ANX\|MD | 7 | 1.1862 | 1.0057 | 1.3991 | 0.0842 | 4.3e-02 |
| ANX\|MD | 8 | 1.0560 | 0.8917 | 1.2507 | 0.0863 | 5.3e-01 |
| ANX\|MD | 9 | 1.1536 | 0.9768 | 1.3624 | 0.0849 | 9.2e-02 |
| ANX\|MD | 10 | 1.1922 | 1.0112 | 1.4057 | 0.0840 | 3.6e-02 |
| **ANX wsumPRS** | | | | | | |
| ANX\|MD | 2 | 1.0013 | 0.8383 | 1.1960 | 0.0906 | 9.9e-01 |
| ANX\|MD | 3 | 1.0388 | 0.8706 | 1.2396 | 0.0902 | 6.7e-01 |
| ANX\|MD | 4 | 1.1344 | 0.9538 | 1.3492 | 0.0885 | 1.5e-01 |
| ANX\|MD | 5 | 1.2140 | 1.0230 | 1.4406 | 0.0873 | 2.6e-02 |
| ANX\|MD | 6 | 1.1939 | 1.0065 | 1.4161 | 0.0871 | 4.2e-02 |
| ANX\|MD | 7 | 1.3465 | 1.1395 | 1.5910 | 0.0851 | 4.8e-04 |
| ANX\|MD | 8 | 1.2841 | 1.0845 | 1.5205 | 0.0862 | 3.7e-03 |
| ANX\|MD | 9 | 1.3073 | 1.1054 | 1.5461 | 0.0856 | 1.7e-03 |
| ANX\|MD | 10 | 1.5037 | 1.2773 | 1.7702 | 0.0832 | 9.6e-07 |

Table S20B: Hazard rate ratios (HRR) of transition into bipolar disorder among MD cases of PRS decile 2-10 compared to the first decile. Left most column is PRS/subtype (PRS/Phenotype) here a second diagnosis of recurrent depression among cases with depression; PRS decile 2-10 (Category); Harzard Rate Ratio (HRR) by decile 2-10 compared to the first PRS decile estimated by cox-regression; lower 95% confidence limit (CIl) and upper 95% confidence limit (CIu) of HRR; standard error of HRR (SE); two-sided p-value of test for HRR being different from zero. see Musliner, K. L. et al. Polygenic Risk and Progression to Bipolar or Psychotic Disorders Among Individuals Diagnosed With Unipolar Depression in Early Life. Am J Psychiatry 177, 936-943, doi:10.1176/appi.ajp.2020.19111195 (2020).

| PRS/Phenotype | Category | HRR | CIl | CIu | SE | P |
|---|---|---|---|---|---|---|
| **MD** | | | | | | |
| BP\|MD | 2 | 1.1419 | 0.8493 | 1.5352 | 0.1510 | 3.8e-01 |
| BP\|MD | 3 | 1.3234 | 0.9932 | 1.7633 | 0.1464 | 5.6e-02 |
| BP\|MD | 4 | 1.4107 | 1.0565 | 1.8836 | 0.1475 | 2.0e-02 |
| BP\|MD | 5 | 1.4008 | 1.0537 | 1.8623 | 0.1453 | 2.0e-02 |
| BP\|MD | 6 | 1.3717 | 1.0375 | 1.8135 | 0.1425 | 2.7e-02 |
| BP\|MD | 7 | 1.4546 | 1.0961 | 1.9302 | 0.1443 | 9.4e-03 |
| BP\|MD | 8 | 1.4199 | 1.0693 | 1.8854 | 0.1447 | 1.5e-02 |
| BP\|MD | 9 | 1.5258 | 1.1537 | 2.0180 | 0.1426 | 3.1e-03 |
| BP\|MD | 10 | 1.4562 | 1.0988 | 1.9297 | 0.1437 | 8.9e-03 |
| **ANX** | | | | | | |
| BP\|MD | 2 | 0.9613 | 0.7335 | 1.2599 | 0.1380 | 7.8e-01 |
| BP\|MD | 3 | 0.8973 | 0.6806 | 1.1831 | 0.1410 | 4.4e-01 |
| BP\|MD | 4 | 1.2458 | 0.9660 | 1.6066 | 0.1298 | 9.0e-02 |
| BP\|MD | 5 | 1.1183 | 0.8608 | 1.4528 | 0.1335 | 4.0e-01 |
| BP\|MD | 6 | 1.2012 | 0.9298 | 1.5518 | 0.1307 | 1.6e-01 |
| BP\|MD | 7 | 1.0290 | 0.7866 | 1.3461 | 0.1371 | 8.3e-01 |
| BP\|MD | 8 | 1.0753 | 0.8266 | 1.3988 | 0.1342 | 5.9e-01 |
| BP\|MD | 9 | 1.0875 | 0.8368 | 1.4131 | 0.1337 | 5.3e-01 |
| BP\|MD | 10 | 1.0322 | 0.7907 | 1.3474 | 0.1360 | 8.2e-01 |
| **BP** | | | | | | |
| BP\|MD | 2 | 0.8965 | 0.6398 | 1.2562 | 0.1721 | 5.3e-01 |
| BP\|MD | 3 | 1.1148 | 0.8326 | 1.4927 | 0.1489 | 4.7e-01 |
| BP\|MD | 4 | 0.9447 | 0.7094 | 1.2580 | 0.1461 | 7.0e-01 |
| BP\|MD | 5 | 1.1513 | 0.8793 | 1.5074 | 0.1375 | 3.1e-01 |
| BP\|MD | 6 | 1.2108 | 0.9292 | 1.5776 | 0.1350 | 1.6e-01 |
| BP\|MD | 7 | 1.3442 | 1.0347 | 1.7464 | 0.1335 | 2.7e-02 |
| BP\|MD | 8 | 1.3063 | 0.9922 | 1.7197 | 0.1403 | 5.7e-02 |
| BP\|MD | 9 | 1.5282 | 1.1552 | 2.0219 | 0.1428 | 3.0e-03 |
| BP\|MD | 10 | 1.8428 | 1.4436 | 2.3524 | 0.1246 | 9.3e-07 |
| **SZ** | | | | | | |
| BP\|MD | 2 | 1.1505 | 0.8593 | 1.5405 | 0.1489 | 3.5e-01 |
| BP\|MD | 3 | 1.3084 | 0.9591 | 1.7849 | 0.1584 | 9.0e-02 |
| BP\|MD | 4 | 1.4498 | 1.1024 | 1.9068 | 0.1398 | 7.9e-03 |
| BP\|MD | 5 | 1.2439 | 0.9241 | 1.6744 | 0.1516 | 1.5e-01 |
| BP\|MD | 6 | 1.4535 | 1.0960 | 1.9276 | 0.1440 | 9.4e-03 |
| BP\|MD | 7 | 1.4924 | 1.1286 | 1.9735 | 0.1426 | 5.0e-03 |
| BP\|MD | 8 | 1.5142 | 1.1451 | 2.0024 | 0.1426 | 3.6e-03 |
| BP\|MD | 9 | 1.9581 | 1.4970 | 2.5613 | 0.1370 | 9.3e-07 |
| BP\|MD | 10 | 1.9335 | 1.4805 | 2.5252 | 0.1362 | 1.3e-06 |
| **ADHD** | | | | | | |
| BP\|MD | 2 | 1.2048 | 0.9303 | 1.5603 | 0.1319 | 1.6e-01 |
| BP\|MD | 3 | 1.0314 | 0.7885 | 1.3489 | 0.1370 | 8.2e-01 |
| BP\|MD | 4 | 1.0153 | 0.7745 | 1.3310 | 0.1381 | 9.1e-01 |
| BP\|MD | 5 | 1.1799 | 0.9099 | 1.5301 | 0.1326 | 2.1e-01 |
| BP\|MD | 6 | 1.1008 | 0.8439 | 1.4360 | 0.1356 | 4.8e-01 |
| BP\|MD | 7 | 1.0267 | 0.7846 | 1.3436 | 0.1372 | 8.5e-01 |
| BP\|MD | 8 | 0.9838 | 0.7493 | 1.2917 | 0.1389 | 9.1e-01 |
| BP\|MD | 9 | 0.8946 | 0.6763 | 1.1834 | 0.1427 | 4.4e-01 |

Table S20B: Hazard rate ratios (HRR) of transition into bipolar disorder among MD cases of PRS decile 2-10 compared to the first decile. Left most column is PRS/subtype (PRS/Phenotype) here a second diagnosis of recurrent depression among cases with depression; PRS decile 2-10 (Category); Harzard Rate Ratio (HRR) by decile 2-10 compared to the first PRS decile estimated by cox-regression; lower 95% confidence limit (CIl) and upper 95% confidence limit (CIu) of HRR; standard error of HRR (SE); two-sided p-value of test for HRR being different from zero. see Musliner, K. L. et al. Polygenic Risk and Progression to Bipolar or Psychotic Disorders Among Individuals Diagnosed With Unipolar Depression in Early Life. Am J Psychiatry 177, 936-943, doi:10.1176/appi.ajp.2020.19111195 (2020). *(continued)*

| PRS/Phenotype | Category | HRR | CIl | CIu | SE | P |
|---|---|---|---|---|---|---|
| BP\|MD | 10 | 1.1488 | 0.8832 | 1.4943 | 0.1341 | 3.0e-01 |
| **ASD** | | | | | | |
| BP\|MD | 2 | 1.0901 | 0.8433 | 1.4092 | 0.1310 | 5.1e-01 |
| BP\|MD | 3 | 1.0827 | 0.8365 | 1.4014 | 0.1316 | 5.5e-01 |
| BP\|MD | 4 | 0.8329 | 0.6322 | 1.0975 | 0.1407 | 1.9e-01 |
| BP\|MD | 5 | 0.9548 | 0.7335 | 1.2429 | 0.1345 | 7.3e-01 |
| BP\|MD | 6 | 1.0134 | 0.7813 | 1.3144 | 0.1327 | 9.2e-01 |
| BP\|MD | 7 | 1.0165 | 0.7810 | 1.3231 | 0.1345 | 9.0e-01 |
| BP\|MD | 8 | 1.0612 | 0.8187 | 1.3755 | 0.1323 | 6.5e-01 |
| BP\|MD | 9 | 1.0015 | 0.7707 | 1.3014 | 0.1336 | 9.9e-01 |
| BP\|MD | 10 | 1.1046 | 0.8549 | 1.4273 | 0.1307 | 4.5e-01 |
| **Neuroticism** | | | | | | |
| BP\|MD | 2 | 1.0347 | 0.7919 | 1.3519 | 0.1365 | 8.0e-01 |
| BP\|MD | 3 | 1.0896 | 0.8366 | 1.4190 | 0.1348 | 5.2e-01 |
| BP\|MD | 4 | 1.1194 | 0.8600 | 1.4570 | 0.1345 | 4.0e-01 |
| BP\|MD | 5 | 1.0535 | 0.8079 | 1.3737 | 0.1354 | 7.0e-01 |
| BP\|MD | 6 | 1.2581 | 0.9730 | 1.6268 | 0.1311 | 8.0e-02 |
| BP\|MD | 7 | 0.9646 | 0.7345 | 1.2667 | 0.1390 | 8.0e-01 |
| BP\|MD | 8 | 1.1028 | 0.8477 | 1.4347 | 0.1342 | 4.7e-01 |
| BP\|MD | 9 | 0.9714 | 0.7406 | 1.2741 | 0.1384 | 8.3e-01 |
| BP\|MD | 10 | 1.1169 | 0.8580 | 1.4539 | 0.1346 | 4.1e-01 |
| BP\|MD | 2 | 1.0347 | 0.7919 | 1.3519 | 0.1365 | 8.0e-01 |
| BP\|MD | 3 | 1.0896 | 0.8366 | 1.4190 | 0.1348 | 5.2e-01 |
| BP\|MD | 4 | 1.1194 | 0.8600 | 1.4570 | 0.1345 | 4.0e-01 |
| BP\|MD | 5 | 1.0535 | 0.8079 | 1.3737 | 0.1354 | 7.0e-01 |
| BP\|MD | 6 | 1.2581 | 0.9730 | 1.6268 | 0.1311 | 8.0e-02 |
| BP\|MD | 7 | 0.9646 | 0.7345 | 1.2667 | 0.1390 | 8.0e-01 |
| BP\|MD | 8 | 1.1028 | 0.8477 | 1.4347 | 0.1342 | 4.7e-01 |
| BP\|MD | 9 | 0.9714 | 0.7406 | 1.2741 | 0.1384 | 8.3e-01 |
| BP\|MD | 10 | 1.1169 | 0.8580 | 1.4539 | 0.1346 | 4.1e-01 |
| **Substance Use** | | | | | | |
| BP\|MD | 2 | 0.9548 | 0.7306 | 1.2477 | 0.1365 | 7.3e-01 |
| BP\|MD | 3 | 1.0125 | 0.7779 | 1.3180 | 0.1345 | 9.3e-01 |
| BP\|MD | 4 | 1.0023 | 0.7687 | 1.3069 | 0.1354 | 9.9e-01 |
| BP\|MD | 5 | 1.0508 | 0.8089 | 1.3651 | 0.1335 | 7.1e-01 |
| BP\|MD | 6 | 1.1024 | 0.8509 | 1.4282 | 0.1321 | 4.6e-01 |
| BP\|MD | 7 | 1.0150 | 0.7790 | 1.3224 | 0.1350 | 9.1e-01 |
| BP\|MD | 8 | 1.1792 | 0.9136 | 1.5219 | 0.1302 | 2.1e-01 |
| BP\|MD | 9 | 1.0752 | 0.8296 | 1.3934 | 0.1323 | 5.8e-01 |
| BP\|MD | 10 | 0.9677 | 0.7402 | 1.2650 | 0.1367 | 8.1e-01 |
| **Substance Use Disorder** | | | | | | |
| BP\|MD | 2 | 1.0266 | 0.7885 | 1.3367 | 0.1346 | 8.5e-01 |
| BP\|MD | 3 | 1.0018 | 0.7678 | 1.3072 | 0.1357 | 9.9e-01 |
| BP\|MD | 4 | 1.1509 | 0.8895 | 1.4891 | 0.1314 | 2.9e-01 |
| BP\|MD | 5 | 0.9650 | 0.7375 | 1.2625 | 0.1371 | 7.9e-01 |
| BP\|MD | 6 | 1.1998 | 0.9298 | 1.5481 | 0.1301 | 1.6e-01 |
| BP\|MD | 7 | 1.1683 | 0.9036 | 1.5105 | 0.1311 | 2.4e-01 |
| BP\|MD | 8 | 1.0047 | 0.7703 | 1.3104 | 0.1355 | 9.7e-01 |
| BP\|MD | 9 | 0.9706 | 0.7421 | 1.2693 | 0.1369 | 8.3e-01 |
| BP\|MD | 10 | 0.9293 | 0.7090 | 1.2181 | 0.1381 | 6.0e-01 |

Table S20B: Hazard rate ratios (HRR) of transition into bipolar disorder among MD cases of PRS decile 2-10 compared to the first decile. Left most column is PRS/subtype (PRS/Phenotype) here a second diagnosis of recurrent depression among cases with depression; PRS decile 2-10 (Category); Harzard Rate Ratio (HRR) by decile 2-10 compared to the first PRS decile estimated by cox-regression; lower 95% confidence limit (CIl) and upper 95% confidence limit (CIu) of HRR; standard error of HRR (SE); two-sided p-value of test for HRR being different from zero. see Musliner, K. L. et al. Polygenic Risk and Progression to Bipolar or Psychotic Disorders Among Individuals Diagnosed With Unipolar Depression in Early Life. Am J Psychiatry 177, 936-943, doi:10.1176/appi.ajp.2020.19111195 (2020). *(continued)*

| PRS/Phenotype | Category | HRR | CIl | CIu | SE | P |
|---|---|---|---|---|---|---|
| **BP wsumPRS** | | | | | | |
| BP\|MD | 2 | 1.2555 | 0.9314 | 1.6925 | 0.1524 | 1.4e-01 |
| BP\|MD | 3 | 1.2240 | 0.9077 | 1.6506 | 0.1525 | 1.9e-01 |
| BP\|MD | 4 | 1.2193 | 0.9030 | 1.6464 | 0.1532 | 2.0e-01 |
| BP\|MD | 5 | 1.1422 | 0.8435 | 1.5466 | 0.1547 | 3.9e-01 |
| BP\|MD | 6 | 1.3940 | 1.0412 | 1.8664 | 0.1489 | 2.6e-02 |
| BP\|MD | 7 | 1.6429 | 1.2382 | 2.1800 | 0.1443 | 5.8e-04 |
| BP\|MD | 8 | 1.4275 | 1.0680 | 1.9081 | 0.1481 | 1.6e-02 |
| BP\|MD | 9 | 1.9852 | 1.5115 | 2.6073 | 0.1391 | 8.2e-07 |
| BP\|MD | 10 | 2.3583 | 1.8072 | 3.0774 | 0.1358 | 2.6e-10 |

Table S20C: Hazard rate ratios (HRR) of transition into schizophrenia among MD cases of PRS decile 2-10 compared to the first decile. Left most column is PRS/subtype (PRS/Phenotype) here a second diagnosis of recurrent depression among cases with depression; PRS decile 2-10 (Category); Harzard Rate Ratio (HRR) by decile 2-10 compared to the first PRS decile estimated by cox-regression; lower 95% confidence limit (CIl) and upper 95% confidence limit (CIu) of HRR; standard error of HRR (SE); two-sided p-value of test for HRR being different from zero.

| PRS/Phenotype | Category | HRR | CIl | CIu | SE | P |
|---|---|---|---|---|---|---|
| **MD** | | | | | | |
| SZ\|MD | 2 | 1.0841 | 0.8539 | 1.3763 | 0.1218 | 5.1e-01 |
| SZ\|MD | 3 | 1.2447 | 0.9861 | 1.5710 | 0.1188 | 6.5e-02 |
| SZ\|MD | 4 | 1.2716 | 1.0030 | 1.6123 | 0.1211 | 4.7e-02 |
| SZ\|MD | 5 | 1.1342 | 0.8935 | 1.4398 | 0.1217 | 3.0e-01 |
| SZ\|MD | 6 | 1.1895 | 0.9437 | 1.4994 | 0.1181 | 1.4e-01 |
| SZ\|MD | 7 | 1.2616 | 0.9991 | 1.5929 | 0.1190 | 5.1e-02 |
| SZ\|MD | 8 | 1.3456 | 1.0694 | 1.6931 | 0.1172 | 1.1e-02 |
| SZ\|MD | 9 | 1.3616 | 1.0826 | 1.7126 | 0.1170 | 8.3e-03 |
| SZ\|MD | 10 | 1.3570 | 1.0781 | 1.7081 | 0.1174 | 9.3e-03 |
| **ANX** | | | | | | |
| SZ\|MD | 2 | 0.9496 | 0.7640 | 1.1803 | 0.1110 | 6.4e-01 |
| SZ\|MD | 3 | 0.9061 | 0.7269 | 1.1294 | 0.1124 | 3.8e-01 |
| SZ\|MD | 4 | 0.9578 | 0.7708 | 1.1901 | 0.1108 | 7.0e-01 |
| SZ\|MD | 5 | 0.9389 | 0.7540 | 1.1692 | 0.1119 | 5.7e-01 |
| SZ\|MD | 6 | 0.8963 | 0.7187 | 1.1177 | 0.1126 | 3.3e-01 |
| SZ\|MD | 7 | 1.0749 | 0.8699 | 1.3283 | 0.1080 | 5.0e-01 |
| SZ\|MD | 8 | 0.9051 | 0.7260 | 1.1284 | 0.1125 | 3.8e-01 |
| SZ\|MD | 9 | 0.9873 | 0.7949 | 1.2263 | 0.1106 | 9.1e-01 |
| SZ\|MD | 10 | 0.9745 | 0.7846 | 1.2104 | 0.1106 | 8.2e-01 |
| **BP** | | | | | | |
| SZ\|MD | 2 | 0.8995 | 0.6900 | 1.1725 | 0.1353 | 4.3e-01 |
| SZ\|MD | 3 | 0.9352 | 0.7344 | 1.1910 | 0.1233 | 5.9e-01 |
| SZ\|MD | 4 | 1.0573 | 0.8480 | 1.3181 | 0.1125 | 6.2e-01 |
| SZ\|MD | 5 | 1.1694 | 0.9453 | 1.4467 | 0.1086 | 1.5e-01 |
| SZ\|MD | 6 | 1.0302 | 0.8294 | 1.2795 | 0.1106 | 7.9e-01 |
| SZ\|MD | 7 | 1.0784 | 0.8678 | 1.3401 | 0.1108 | 5.0e-01 |
| SZ\|MD | 8 | 1.0591 | 0.8428 | 1.3309 | 0.1166 | 6.2e-01 |
| SZ\|MD | 9 | 1.2270 | 0.9696 | 1.5526 | 0.1201 | 8.9e-02 |
| SZ\|MD | 10 | 1.4385 | 1.1745 | 1.7618 | 0.1035 | 4.4e-04 |
| **SZ** | | | | | | |
| SZ\|MD | 2 | 1.1769 | 0.9369 | 1.4782 | 0.1163 | 1.6e-01 |
| SZ\|MD | 3 | 1.1605 | 0.9018 | 1.4933 | 0.1287 | 2.5e-01 |
| SZ\|MD | 4 | 1.2202 | 0.9738 | 1.5288 | 0.1150 | 8.4e-02 |
| SZ\|MD | 5 | 1.2792 | 1.0193 | 1.6052 | 0.1158 | 3.4e-02 |
| SZ\|MD | 6 | 1.2119 | 0.9626 | 1.5259 | 0.1175 | 1.0e-01 |
| SZ\|MD | 7 | 1.2666 | 1.0094 | 1.5893 | 0.1158 | 4.1e-02 |
| SZ\|MD | 8 | 1.2896 | 1.0251 | 1.6223 | 0.1171 | 3.0e-02 |
| SZ\|MD | 9 | 1.4130 | 1.1329 | 1.7623 | 0.1127 | 2.2e-03 |
| SZ\|MD | 10 | 1.6171 | 1.3020 | 2.0085 | 0.1106 | 1.4e-05 |
| **ADHD** | | | | | | |
| SZ\|MD | 2 | 1.0101 | 0.7846 | 1.3005 | 0.1289 | 9.4e-01 |
| SZ\|MD | 3 | 0.8573 | 0.6597 | 1.1139 | 0.1336 | 2.5e-01 |
| SZ\|MD | 4 | 0.9857 | 0.7649 | 1.2702 | 0.1294 | 9.1e-01 |
| SZ\|MD | 5 | 1.2029 | 0.9451 | 1.5309 | 0.1231 | 1.3e-01 |
| SZ\|MD | 6 | 0.9298 | 0.7193 | 1.2019 | 0.1310 | 5.8e-01 |
| SZ\|MD | 7 | 0.9793 | 0.7599 | 1.2621 | 0.1294 | 8.7e-01 |
| SZ\|MD | 8 | 1.0248 | 0.7979 | 1.3161 | 0.1277 | 8.5e-01 |
| SZ\|MD | 9 | 1.0924 | 0.8532 | 1.3987 | 0.1261 | 4.8e-01 |
| SZ\|MD | 10 | 1.3147 | 1.0368 | 1.6672 | 0.1212 | 2.4e-02 |
| **ASD** | | | | | | |

Table S20C: Hazard rate ratios (HRR) of transition into schizophrenia among MD cases of PRS decile 2-10 compared to the first decile. Left most column is PRS/subtype (PRS/Phenotype) here a second diagnosis of recurrent depression among cases with depression; PRS decile 2-10 (Category); Harzard Rate Ratio (HRR) by decile 2-10 compared to the first PRS decile estimated by cox-regression; lower 95% confidence limit (CIl) and upper 95% confidence limit (CIu) of HRR; standard error of HRR (SE); two-sided p-value of test for HRR being different from zero. *(continued)*

| PRS/Phenotype | Category | HRR | CIl | CIu | SE | P |
|---|---|---|---|---|---|---|
| SZ\|MD | 2 | 0.9496 | 0.7640 | 1.1803 | 0.1110 | 6.4e-01 |
| SZ\|MD | 3 | 0.9061 | 0.7269 | 1.1294 | 0.1124 | 3.8e-01 |
| SZ\|MD | 4 | 0.9578 | 0.7708 | 1.1901 | 0.1108 | 7.0e-01 |
| SZ\|MD | 5 | 0.9389 | 0.7540 | 1.1692 | 0.1119 | 5.7e-01 |
| SZ\|MD | 6 | 0.8963 | 0.7187 | 1.1177 | 0.1126 | 3.3e-01 |
| SZ\|MD | 7 | 1.0749 | 0.8699 | 1.3283 | 0.1080 | 5.0e-01 |
| SZ\|MD | 8 | 0.9051 | 0.7260 | 1.1284 | 0.1125 | 3.8e-01 |
| SZ\|MD | 9 | 0.9873 | 0.7949 | 1.2263 | 0.1106 | 9.1e-01 |
| SZ\|MD | 10 | 0.9745 | 0.7846 | 1.2104 | 0.1106 | 8.2e-01 |
| **Neuroticism** | | | | | | |
| SZ\|MD | 2 | 1.0247 | 0.8114 | 1.2941 | 0.1191 | 8.4e-01 |
| SZ\|MD | 3 | 1.1090 | 0.8821 | 1.3942 | 0.1168 | 3.8e-01 |
| SZ\|MD | 4 | 1.1389 | 0.9072 | 1.4298 | 0.1160 | 2.6e-01 |
| SZ\|MD | 5 | 1.2784 | 1.0238 | 1.5962 | 0.1133 | 3.0e-02 |
| SZ\|MD | 6 | 1.1505 | 0.9166 | 1.4441 | 0.1160 | 2.3e-01 |
| SZ\|MD | 7 | 1.1045 | 0.8777 | 1.3898 | 0.1172 | 4.0e-01 |
| SZ\|MD | 8 | 1.1060 | 0.8799 | 1.3903 | 0.1167 | 3.9e-01 |
| SZ\|MD | 9 | 1.1480 | 0.9151 | 1.4402 | 0.1157 | 2.3e-01 |
| SZ\|MD | 10 | 1.3309 | 1.0673 | 1.6596 | 0.1126 | 1.1e-02 |
| **Substance Use** | | | | | | |
| SZ\|MD | 2 | 0.9803 | 0.7754 | 1.2393 | 0.1196 | 8.7e-01 |
| SZ\|MD | 3 | 1.2029 | 0.9623 | 1.5036 | 0.1139 | 1.0e-01 |
| SZ\|MD | 4 | 0.9565 | 0.7554 | 1.2110 | 0.1204 | 7.1e-01 |
| SZ\|MD | 5 | 1.0949 | 0.8713 | 1.3758 | 0.1165 | 4.4e-01 |
| SZ\|MD | 6 | 1.3092 | 1.0512 | 1.6304 | 0.1120 | 1.6e-02 |
| SZ\|MD | 7 | 1.2723 | 1.0197 | 1.5875 | 0.1129 | 3.3e-02 |
| SZ\|MD | 8 | 1.2098 | 0.9678 | 1.5122 | 0.1138 | 9.4e-02 |
| SZ\|MD | 9 | 1.1264 | 0.8987 | 1.4118 | 0.1152 | 3.0e-01 |
| SZ\|MD | 10 | 1.2741 | 1.0212 | 1.5897 | 0.1129 | 3.2e-02 |
| **Substance Use Disorder** | | | | | | |
| SZ\|MD | 2 | 1.0226 | 0.8207 | 1.2741 | 0.1122 | 8.4e-01 |
| SZ\|MD | 3 | 0.8862 | 0.7054 | 1.1133 | 0.1164 | 3.0e-01 |
| SZ\|MD | 4 | 1.0161 | 0.8154 | 1.2664 | 0.1123 | 8.9e-01 |
| SZ\|MD | 5 | 1.0577 | 0.8503 | 1.3157 | 0.1114 | 6.1e-01 |
| SZ\|MD | 6 | 0.9810 | 0.7861 | 1.2242 | 0.1130 | 8.7e-01 |
| SZ\|MD | 7 | 1.0106 | 0.8113 | 1.2591 | 0.1121 | 9.2e-01 |
| SZ\|MD | 8 | 0.9780 | 0.7831 | 1.2213 | 0.1134 | 8.4e-01 |
| SZ\|MD | 9 | 1.0933 | 0.8806 | 1.3573 | 0.1104 | 4.2e-01 |
| SZ\|MD | 10 | 1.1409 | 0.9210 | 1.4133 | 0.1092 | 2.3e-01 |
| **BP wsumPRS** | | | | | | |
| SZ\|MD | 2 | 1.2189 | 0.9537 | 1.5580 | 0.1252 | 1.1e-01 |
| SZ\|MD | 3 | 1.2453 | 0.9754 | 1.5898 | 0.1246 | 7.8e-02 |
| SZ\|MD | 4 | 1.4112 | 1.1117 | 1.7913 | 0.1217 | 4.6e-03 |
| SZ\|MD | 5 | 1.3572 | 1.0682 | 1.7244 | 0.1222 | 1.2e-02 |
| SZ\|MD | 6 | 1.4051 | 1.1072 | 1.7832 | 0.1216 | 5.2e-03 |
| SZ\|MD | 7 | 1.3108 | 1.0286 | 1.6704 | 0.1237 | 2.9e-02 |
| SZ\|MD | 8 | 1.5551 | 1.2312 | 1.9642 | 0.1192 | 2.1e-04 |
| SZ\|MD | 9 | 1.4757 | 1.1663 | 1.8671 | 0.1201 | 1.2e-03 |
| SZ\|MD | 10 | 1.8152 | 1.4471 | 2.2770 | 0.1156 | 2.5e-07 |

Table S20D: Hazard rate ratios (HRR) of developing substance-use-disorder among MD cases of PRS decile 2-10 compared to the first decile. Left most column is PRS/subtype (PRS/Phenotype) here a second diagnosis of recurrent depression among cases with depression; PRS decile 2-10 (Category); Harzard Rate Ratio (HRR) by decile 2-10 compared to the first PRS decile estimated by cox-regression; lower 95% confidence limit (CIl) and upper 95% confidence limit (CIu) of HRR; standard error of HRR (SE); two-sided p-value of test for HRR being different from zero.

| PRS/Phenotype | Category | HRR | CIl | CIu | SE | P |
|---|---|---|---|---|---|---|
| **MD** | | | | | | |
| SUD\|MD | 2 | 1.2787 | 1.0059 | 1.6253 | 0.1224 | 4.5e-02 |
| SUD\|MD | 3 | 1.2327 | 0.9678 | 1.5701 | 0.1234 | 9.0e-02 |
| SUD\|MD | 4 | 1.2329 | 0.9617 | 1.5805 | 0.1267 | 9.9e-02 |
| SUD\|MD | 5 | 1.1936 | 0.9342 | 1.5249 | 0.1250 | 1.6e-01 |
| SUD\|MD | 6 | 1.3052 | 1.0290 | 1.6555 | 0.1213 | 2.8e-02 |
| SUD\|MD | 7 | 1.3945 | 1.1003 | 1.7673 | 0.1209 | 6.0e-03 |
| SUD\|MD | 8 | 1.5238 | 1.2068 | 1.9241 | 0.1190 | 4.0e-04 |
| SUD\|MD | 9 | 1.4598 | 1.1536 | 1.8472 | 0.1201 | 1.6e-03 |
| SUD\|MD | 10 | 1.7197 | 1.3698 | 2.1588 | 0.1160 | 3.0e-06 |
| **ANX** | | | | | | |
| BP\|MD | 2 | 1.0118 | 0.7997 | 1.2802 | 0.1200 | 9.2e-01 |
| BP\|MD | 3 | 1.1115 | 0.8826 | 1.3996 | 0.1176 | 3.7e-01 |
| BP\|MD | 4 | 1.1288 | 0.8985 | 1.4180 | 0.1164 | 3.0e-01 |
| BP\|MD | 5 | 1.1421 | 0.9087 | 1.4354 | 0.1166 | 2.5e-01 |
| BP\|MD | 6 | 1.1279 | 0.8979 | 1.4169 | 0.1164 | 3.0e-01 |
| BP\|MD | 7 | 1.3209 | 1.0575 | 1.6499 | 0.1135 | 1.4e-02 |
| BP\|MD | 8 | 1.1300 | 0.9000 | 1.4188 | 0.1161 | 2.9e-01 |
| BP\|MD | 9 | 1.2763 | 1.0214 | 1.5948 | 0.1137 | 3.2e-02 |
| BP\|MD | 10 | 1.4676 | 1.1818 | 1.8225 | 0.1105 | 5.2e-04 |
| **BP** | | | | | | |
| SUD\|MD | 2 | 0.8481 | 0.6547 | 1.0985 | 0.1320 | 2.1e-01 |
| SUD\|MD | 3 | 0.9677 | 0.7693 | 1.2173 | 0.1171 | 7.8e-01 |
| SUD\|MD | 4 | 0.9319 | 0.7497 | 1.1583 | 0.1110 | 5.2e-01 |
| SUD\|MD | 5 | 1.0179 | 0.8245 | 1.2567 | 0.1075 | 8.7e-01 |
| SUD\|MD | 6 | 0.9924 | 0.8053 | 1.2230 | 0.1066 | 9.4e-01 |
| SUD\|MD | 7 | 0.9633 | 0.7784 | 1.1922 | 0.1088 | 7.3e-01 |
| SUD\|MD | 8 | 1.0660 | 0.8568 | 1.3262 | 0.1114 | 5.7e-01 |
| SUD\|MD | 9 | 1.1410 | 0.9068 | 1.4357 | 0.1172 | 2.6e-01 |
| SUD\|MD | 10 | 1.1651 | 0.9513 | 1.4269 | 0.1034 | 1.4e-01 |
| **SZ** | | | | | | |
| SUD\|MD | 2 | 1.4262 | 1.1337 | 1.7941 | 0.1171 | 2.4e-03 |
| SUD\|MD | 3 | 1.4254 | 1.1091 | 1.8318 | 0.1280 | 5.6e-03 |
| SUD\|MD | 4 | 1.2732 | 1.0057 | 1.6119 | 0.1203 | 4.5e-02 |
| SUD\|MD | 5 | 1.5054 | 1.1943 | 1.8976 | 0.1181 | 5.3e-04 |
| SUD\|MD | 6 | 1.3947 | 1.1029 | 1.7636 | 0.1197 | 5.5e-03 |
| SUD\|MD | 7 | 1.3575 | 1.0686 | 1.7244 | 0.1221 | 1.2e-02 |
| SUD\|MD | 8 | 1.4422 | 1.1435 | 1.8189 | 0.1184 | 2.0e-03 |
| SUD\|MD | 9 | 1.6083 | 1.2781 | 2.0238 | 0.1172 | 5.1e-05 |
| SUD\|MD | 10 | 1.7371 | 1.3877 | 2.1744 | 0.1146 | 1.4e-06 |
| **ADHD** | | | | | | |
| SUD\|MD | 2 | 1.0450 | 0.8106 | 1.3472 | 0.1296 | 7.3e-01 |
| SUD\|MD | 3 | 1.1492 | 0.8966 | 1.4730 | 0.1266 | 2.7e-01 |
| SUD\|MD | 4 | 1.3039 | 1.0232 | 1.6616 | 0.1237 | 3.2e-02 |
| SUD\|MD | 5 | 1.1273 | 0.8788 | 1.4461 | 0.1271 | 3.5e-01 |
| SUD\|MD | 6 | 1.3699 | 1.0783 | 1.7405 | 0.1221 | 1.0e-02 |
| SUD\|MD | 7 | 1.2655 | 0.9928 | 1.6131 | 0.1238 | 5.7e-02 |
| SUD\|MD | 8 | 1.4781 | 1.1677 | 1.8711 | 0.1203 | 1.2e-03 |
| SUD\|MD | 9 | 1.6182 | 1.2833 | 2.0405 | 0.1183 | 4.7e-05 |
| SUD\|MD | 10 | 1.8191 | 1.4482 | 2.2850 | 0.1163 | 2.7e-07 |
| **ASD** | | | | | | |

Table S20D: Hazard rate ratios (HRR) of developing substance-use-disorder among MD cases of PRS decile 2-10 compared to the first decile. Left most column is PRS/subtype (PRS/Phenotype) here a second diagnosis of recurrent depression among cases with depression; PRS decile 2-10 (Category); Harzard Rate Ratio (HRR) by decile 2-10 compared to the first PRS decile estimated by cox-regression; lower 95% confidence limit (CIl) and upper 95% confidence limit (CIu) of HRR; standard error of HRR (SE); two-sided p-value of test for HRR being different from zero. *(continued)*

| PRS/Phenotype | Category | HRR | CIl | CIu | SE | P |
|---|---|---|---|---|---|---|
| BP\|MD | 2 | 0.9111 | 0.7309 | 1.1357 | 0.1124 | 4.1e-01 |
| BP\|MD | 3 | 1.1909 | 0.9673 | 1.4663 | 0.1061 | 1.0e-01 |
| BP\|MD | 4 | 0.9494 | 0.7627 | 1.1818 | 0.1117 | 6.4e-01 |
| BP\|MD | 5 | 0.9729 | 0.7841 | 1.2073 | 0.1101 | 8.0e-01 |
| BP\|MD | 6 | 0.9019 | 0.7237 | 1.1240 | 0.1123 | 3.6e-01 |
| BP\|MD | 7 | 0.9286 | 0.7446 | 1.1580 | 0.1127 | 5.1e-01 |
| BP\|MD | 8 | 1.0439 | 0.8444 | 1.2906 | 0.1082 | 6.9e-01 |
| BP\|MD | 9 | 0.8600 | 0.6871 | 1.0764 | 0.1145 | 1.9e-01 |
| BP\|MD | 10 | 0.8652 | 0.6923 | 1.0814 | 0.1138 | 2.0e-01 |
| **Neuroticism** | | | | | | |
| SUD\|MD | 2 | 1.0523 | 0.8348 | 1.3265 | 0.1181 | 6.7e-01 |
| SUD\|MD | 3 | 1.3067 | 1.0479 | 1.6294 | 0.1126 | 1.8e-02 |
| SUD\|MD | 4 | 1.0007 | 0.7914 | 1.2655 | 0.1198 | 1.0e+00 |
| SUD\|MD | 5 | 1.3249 | 1.0632 | 1.6510 | 0.1123 | 1.2e-02 |
| SUD\|MD | 6 | 1.3167 | 1.0556 | 1.6424 | 0.1128 | 1.5e-02 |
| SUD\|MD | 7 | 1.1306 | 0.9002 | 1.4199 | 0.1163 | 2.9e-01 |
| SUD\|MD | 8 | 1.0987 | 0.8740 | 1.3812 | 0.1168 | 4.2e-01 |
| SUD\|MD | 9 | 1.0976 | 0.8728 | 1.3804 | 0.1169 | 4.3e-01 |
| SUD\|MD | 10 | 1.1217 | 0.8935 | 1.4082 | 0.1161 | 3.2e-01 |
| **Substance Use** | | | | | | |
| SUD\|MD | 2 | 1.6368 | 1.2422 | 2.1569 | 0.1408 | 4.6e-04 |
| SUD\|MD | 3 | 1.5567 | 1.1786 | 2.0561 | 0.1420 | 1.8e-03 |
| SUD\|MD | 4 | 1.5626 | 1.1840 | 2.0621 | 0.1415 | 1.6e-03 |
| SUD\|MD | 5 | 1.7199 | 1.3092 | 2.2595 | 0.1392 | 9.8e-05 |
| SUD\|MD | 6 | 2.0721 | 1.5905 | 2.6995 | 0.1350 | 6.7e-08 |
| SUD\|MD | 7 | 1.9049 | 1.4569 | 2.4906 | 0.1368 | 2.5e-06 |
| SUD\|MD | 8 | 2.4026 | 1.8551 | 3.1115 | 0.1319 | 3.1e-11 |
| SUD\|MD | 9 | 3.2124 | 2.5052 | 4.1193 | 0.1269 | 3.6e-20 |
| SUD\|MD | 10 | 3.1582 | 2.4607 | 4.0534 | 0.1273 | 1.7e-19 |
| **Substance Use Disorder** | | | | | | |
| SUD\|MD | 2 | 1.1070 | 0.8603 | 1.4243 | 0.1286 | 4.3e-01 |
| SUD\|MD | 3 | 1.1813 | 0.9221 | 1.5135 | 0.1264 | 1.9e-01 |
| SUD\|MD | 4 | 1.2973 | 1.0166 | 1.6555 | 0.1244 | 3.6e-02 |
| SUD\|MD | 5 | 1.4394 | 1.1342 | 1.8266 | 0.1216 | 2.7e-03 |
| SUD\|MD | 6 | 1.7491 | 1.3906 | 2.2002 | 0.1170 | 1.8e-06 |
| SUD\|MD | 7 | 1.2869 | 1.0095 | 1.6405 | 0.1239 | 4.2e-02 |
| SUD\|MD | 8 | 1.3999 | 1.1018 | 1.7785 | 0.1221 | 5.9e-03 |
| SUD\|MD | 9 | 1.6716 | 1.3262 | 2.1070 | 0.1181 | 1.4e-05 |
| SUD\|MD | 10 | 1.8450 | 1.4696 | 2.3162 | 0.1161 | 1.3e-07 |
| **BP wsumPRS** | | | | | | |
| SUD\|MD | 2 | 1.4324 | 1.0690 | 1.9193 | 0.1493 | 1.6e-02 |
| SUD\|MD | 3 | 1.4268 | 1.0635 | 1.9142 | 0.1499 | 1.8e-02 |
| SUD\|MD | 4 | 1.6909 | 1.2735 | 2.2452 | 0.1447 | 2.8e-04 |
| SUD\|MD | 5 | 2.2563 | 1.7214 | 2.9574 | 0.1381 | 3.8e-09 |
| SUD\|MD | 6 | 1.9192 | 1.4535 | 2.5340 | 0.1418 | 4.3e-06 |
| SUD\|MD | 7 | 2.2502 | 1.7167 | 2.9494 | 0.1381 | 4.3e-09 |
| SUD\|MD | 8 | 2.7480 | 2.1116 | 3.5760 | 0.1344 | 5.4e-14 |
| SUD\|MD | 9 | 2.8270 | 2.1748 | 3.6749 | 0.1338 | 8.1e-15 |
| SUD\|MD | 10 | 4.0545 | 3.1511 | 5.2168 | 0.1286 | 1.4e-27 |

#### 2 Supplemental figures

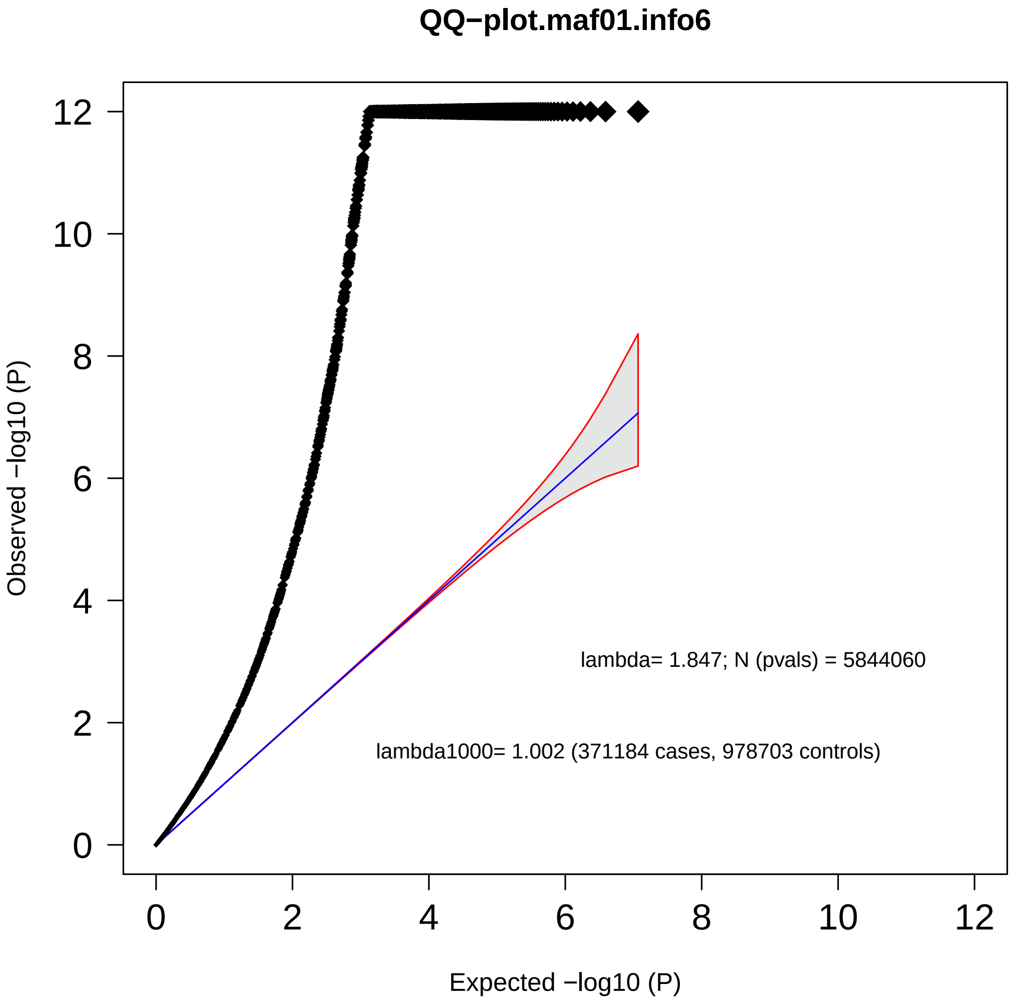

Figure S1: QQ-plot for the primary MD meta-analysis Observed p-value is plotted on the y axis and expected p-value is plotted on the x axis. The red line represents a result with no inflation. LDscore regression showed that % of observed inflation is due to the high polygenicity of depression, with no evidence for inflation due to stratification or confounds indicated by the intercept () or attenuation ratio ().

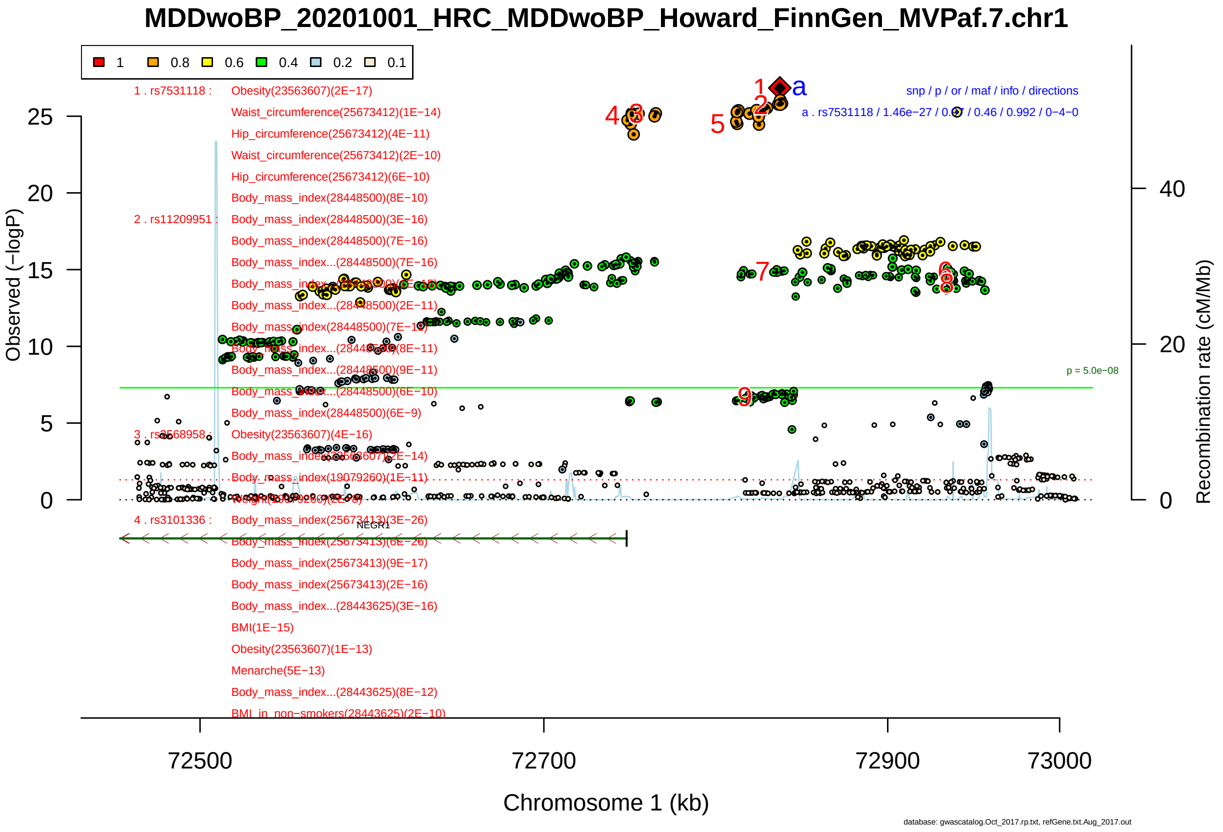

Figure S2-1: Regional plot of GWAS locus No. 1 , with rs7531118 as lead SNP. Neighbouring index SNPs were considered independent when $r^2 < 0.1$ in a sliding 3 Mb window. For each index SNP we defined the associated LD-region by recording the left and rightmost variant with $r^2 < 0.1$. To define GWAS loci, a 50kb window was added on each side of the LD-region and overlapping LD-regions were combined into a single locus.

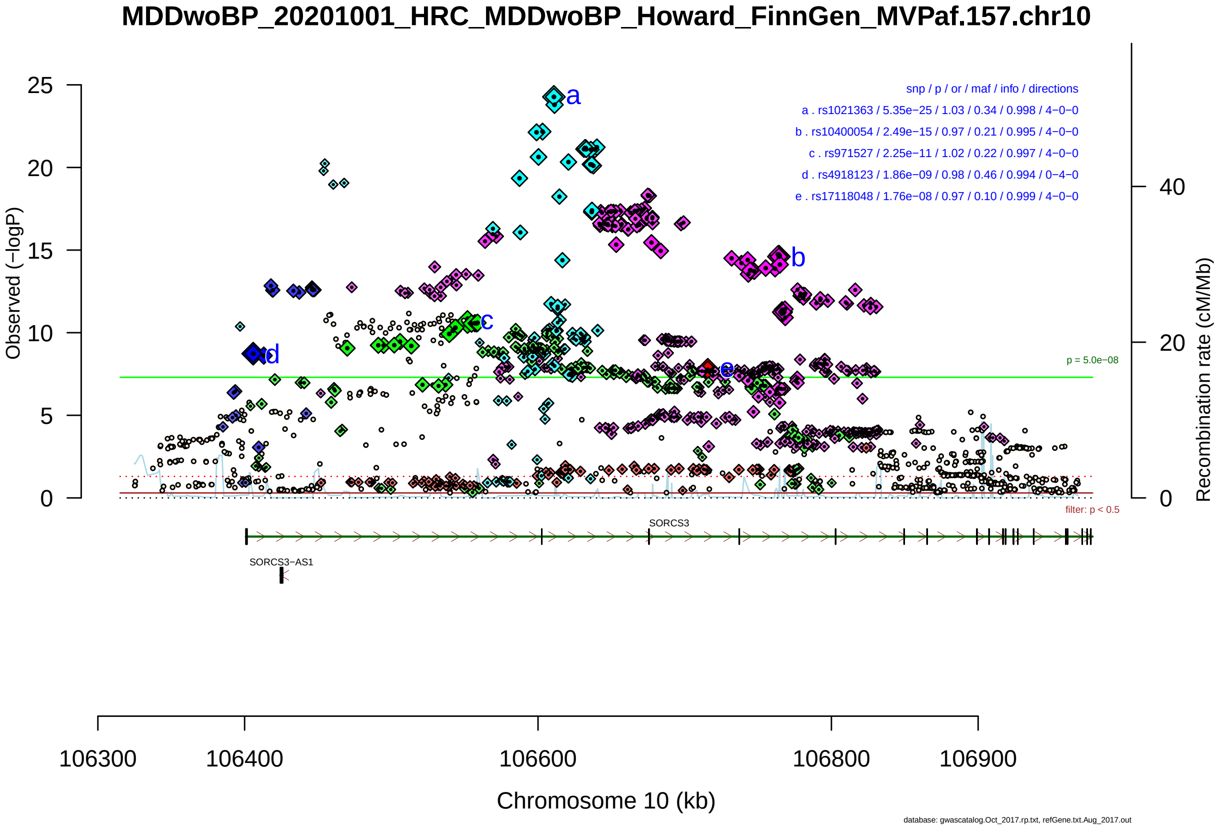

Figure S2-2: Regional plot of GWAS locus No. 2 , with rs1021363 as lead SNP.

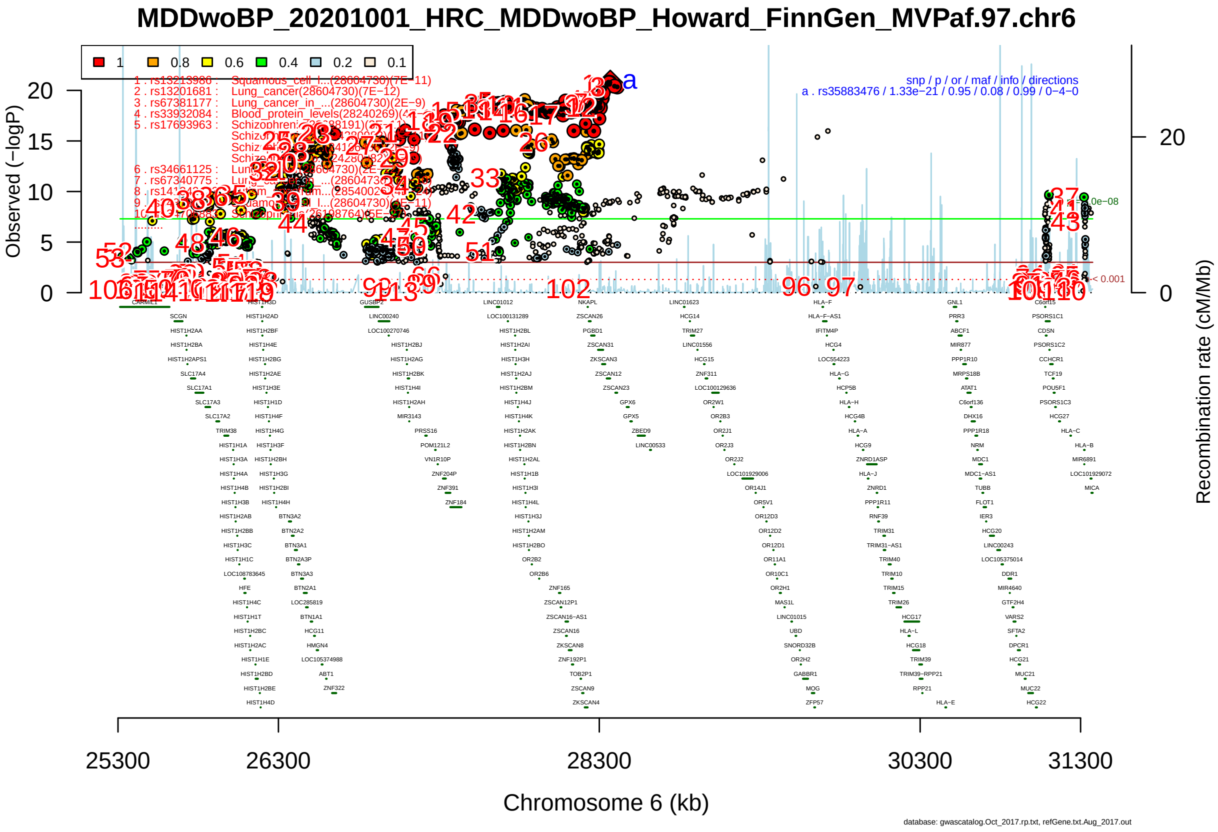

Figure S2-3: Regional plot of GWAS locus No. 3 , with rs35883476 as lead SNP.

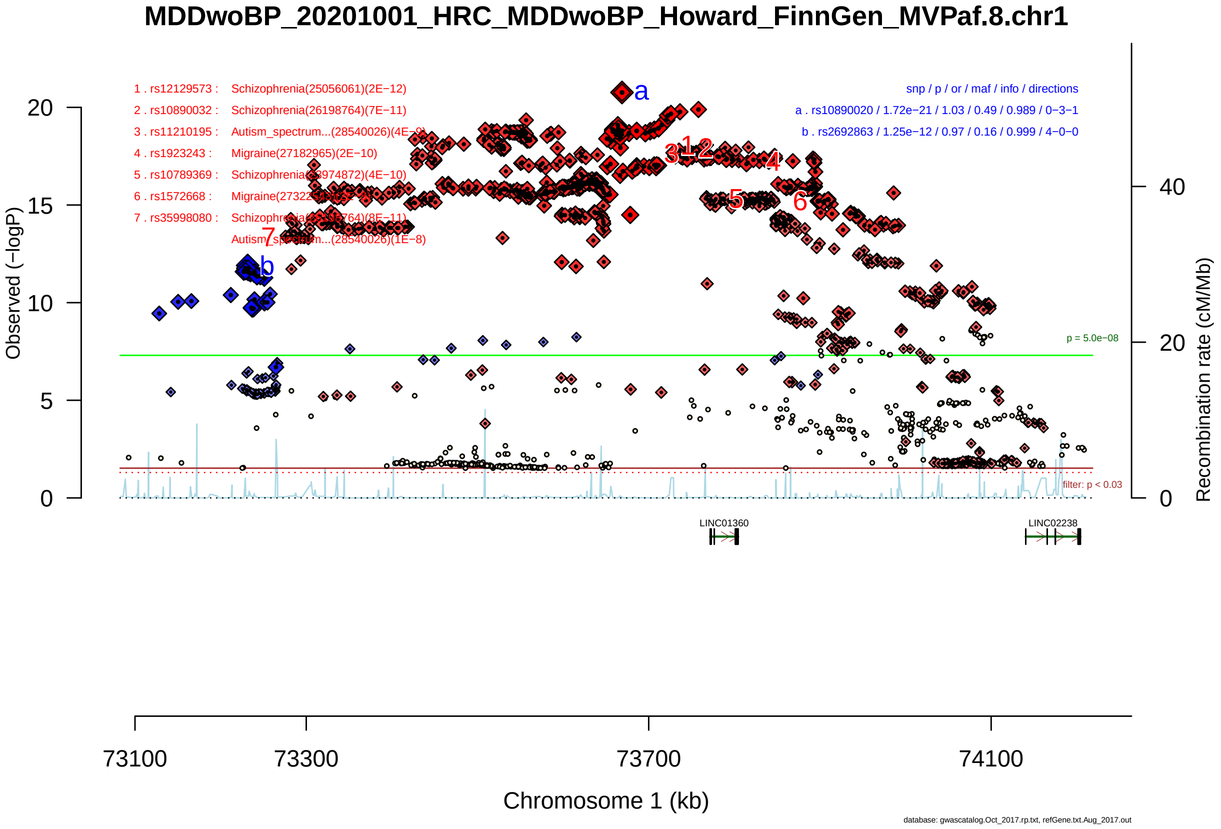

Figure S2-4: Regional plot of GWAS locus No. 4 , with rs10890020 as lead SNP.

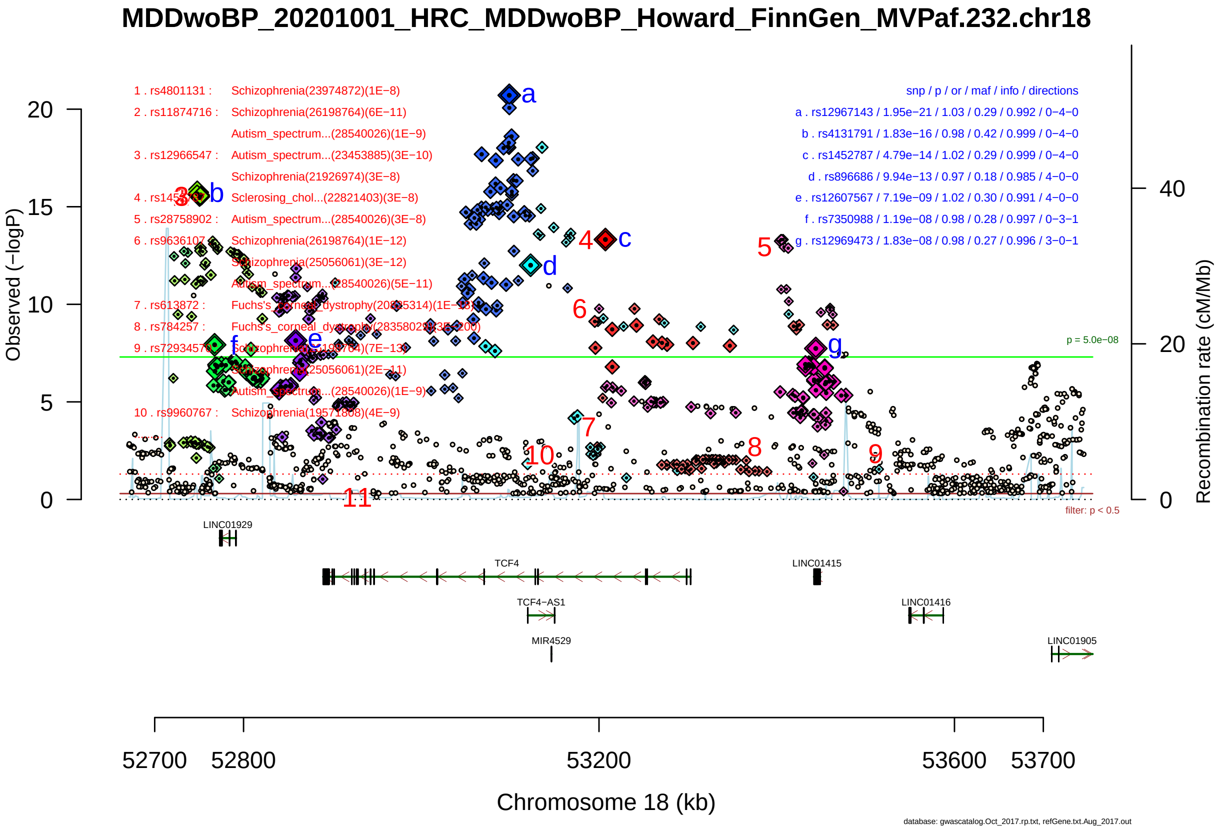

Figure S2-5: Regional plot of GWAS locus No. 5 , with rs12967143 as lead SNP.

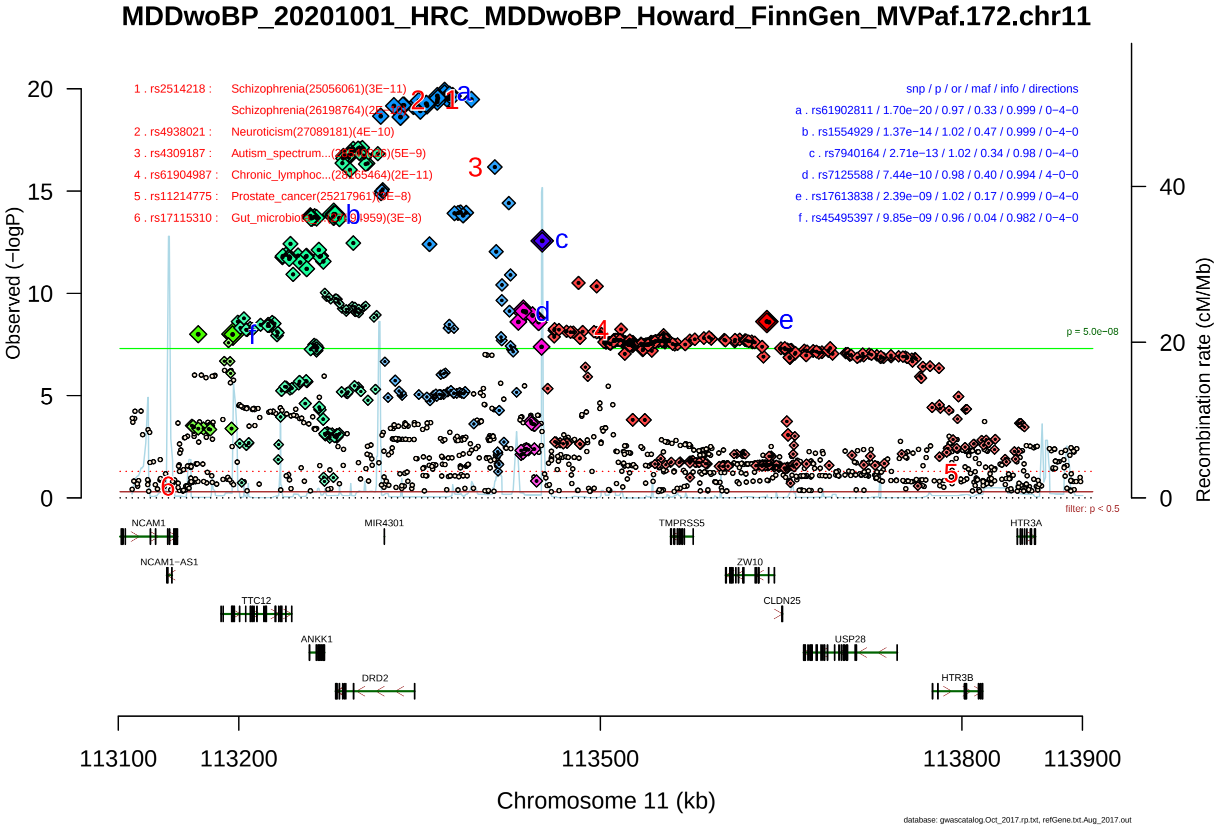

Figure S2-6: Regional plot of GWAS locus No. 6 , with rs61902811 as lead SNP.

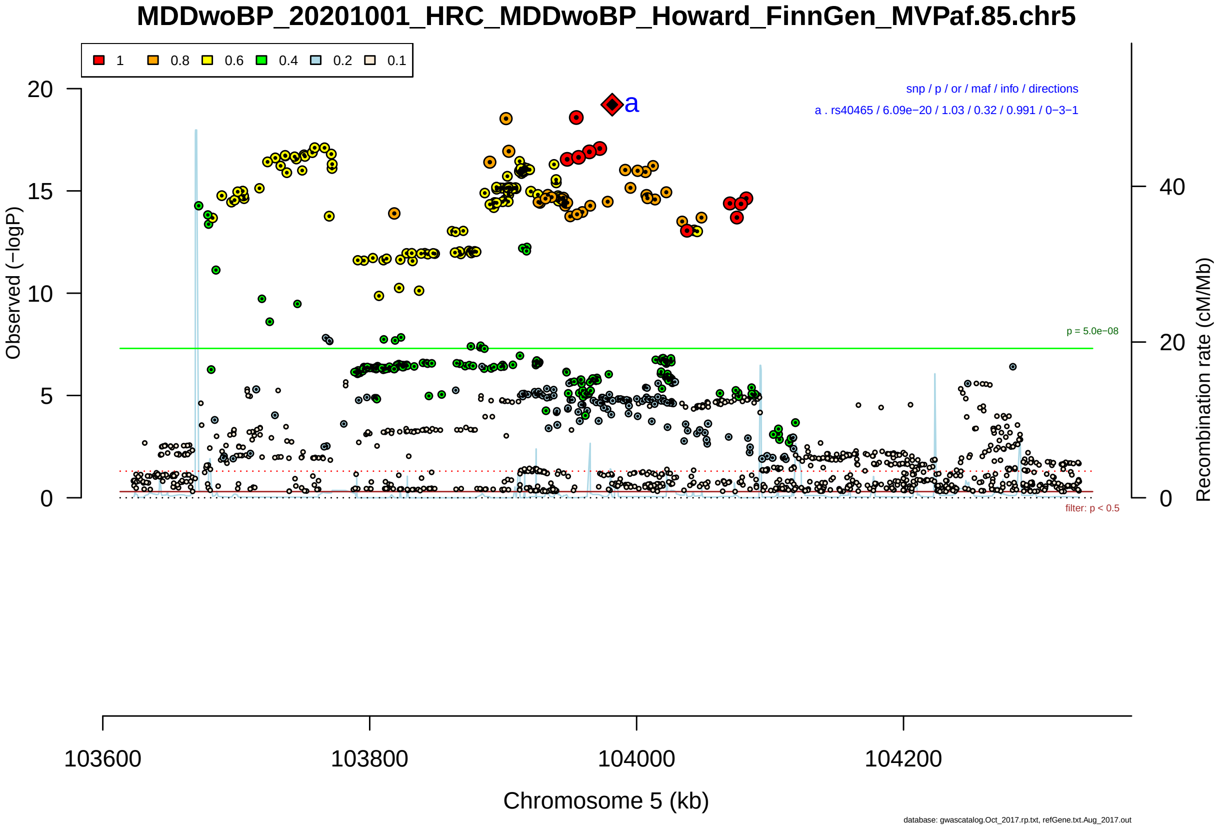

Figure S2-7: Regional plot of GWAS locus No. 7 , with rs40465 as lead SNP.

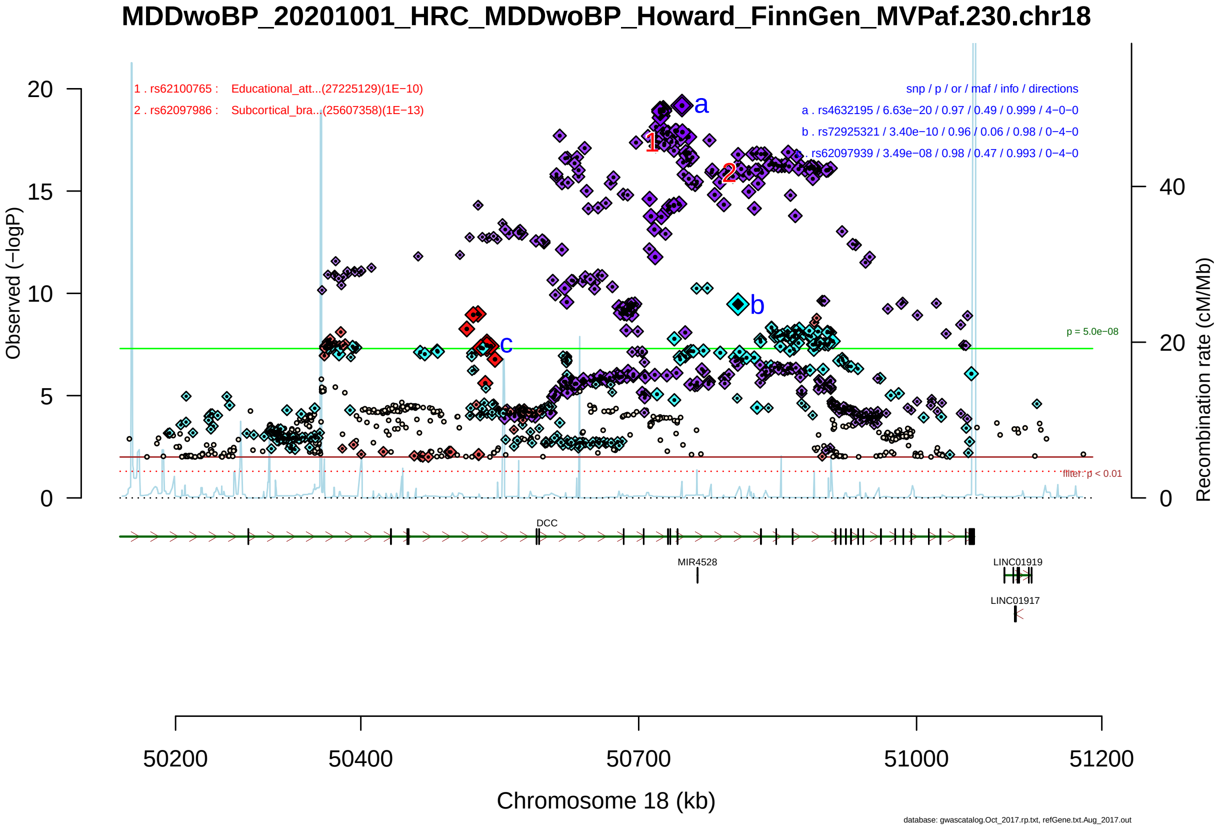

Figure S2-8: Regional plot of GWAS locus No. 8 , with rs4632195 as lead SNP.

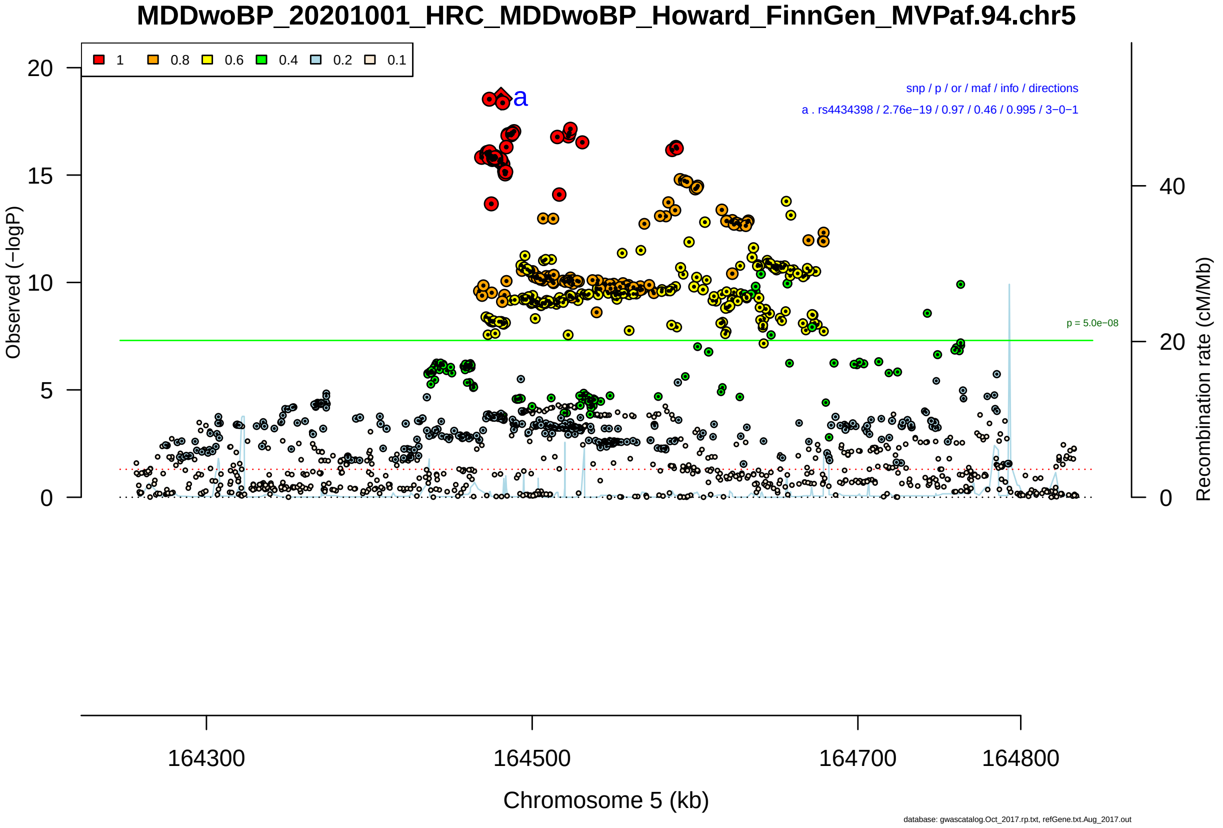

Figure S2-9: Regional plot of GWAS locus No. 9 , with rs4434398 as lead SNP.

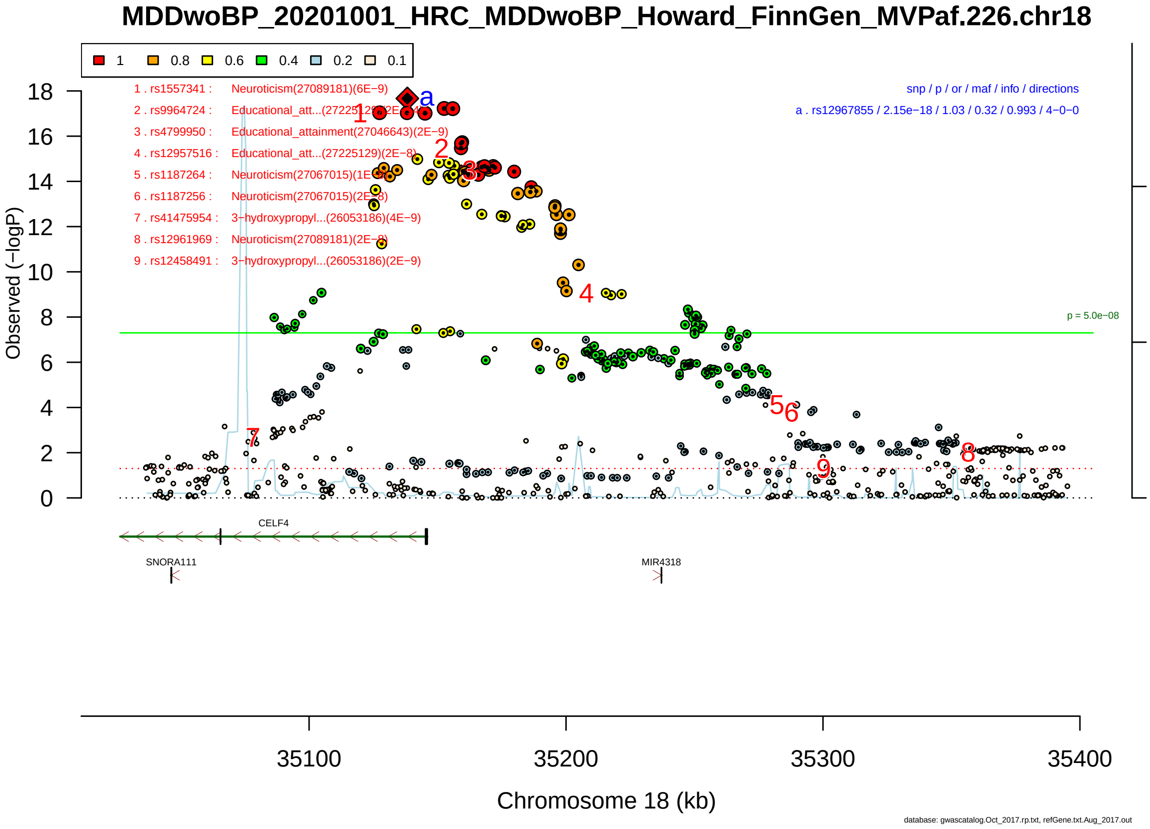

Figure S2-10: Regional plot of GWAS locus No. 10 , with rs12967855 as lead SNP.

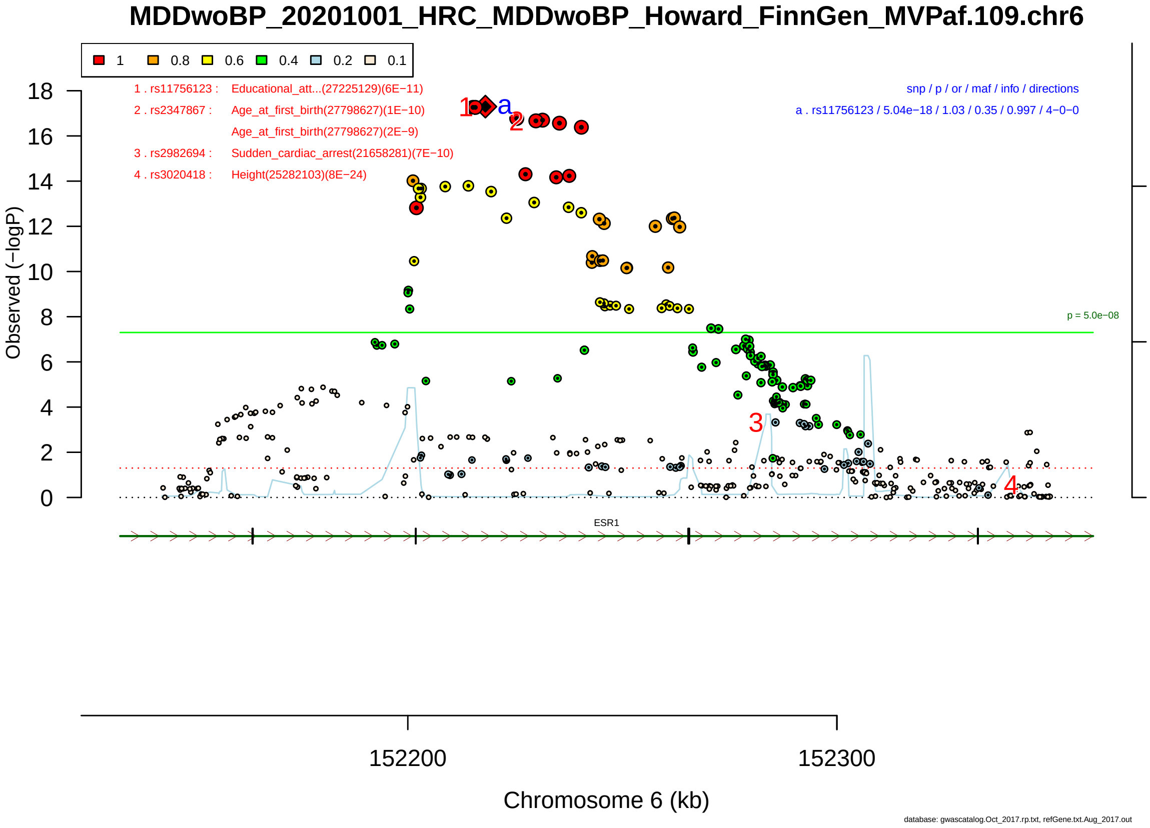

Figure S2-11: Regional plot of GWAS locus No. 11 , with rs11756123 as lead SNP.

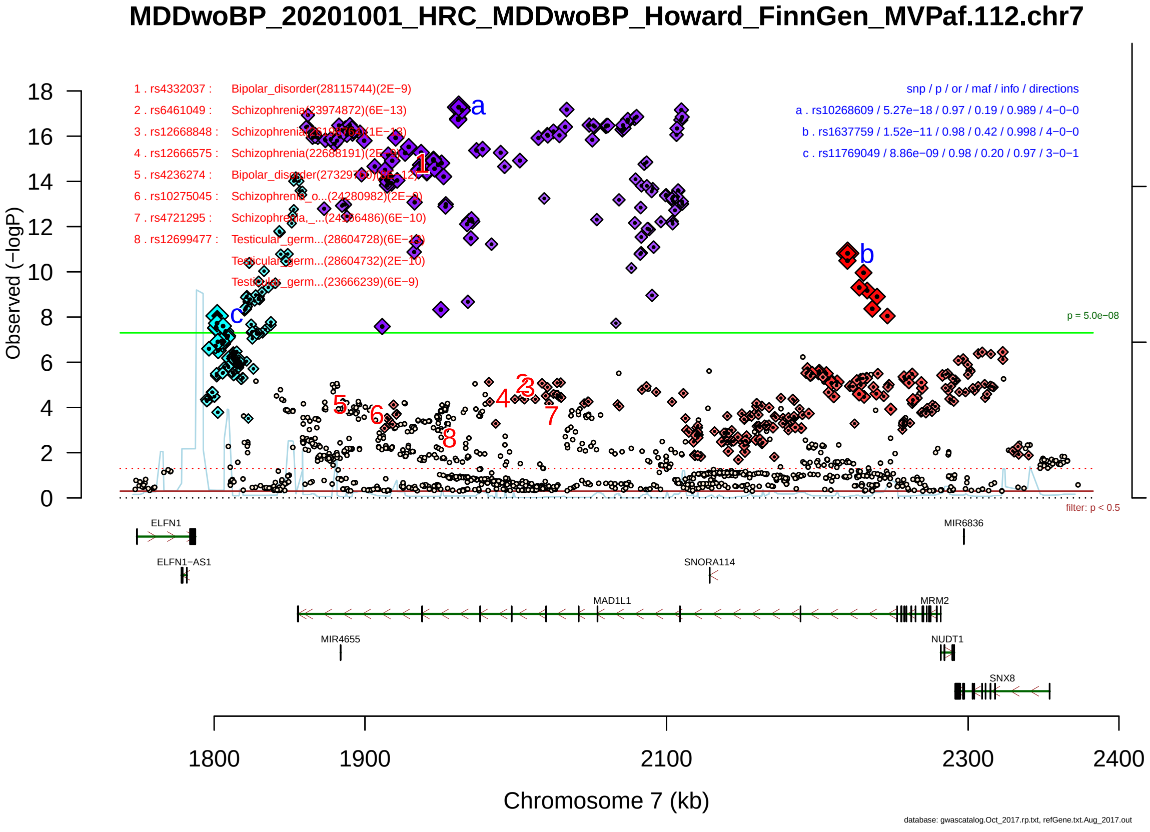

Figure S2-12: Regional plot of GWAS locus No. 12 , with rs10268609 as lead SNP.

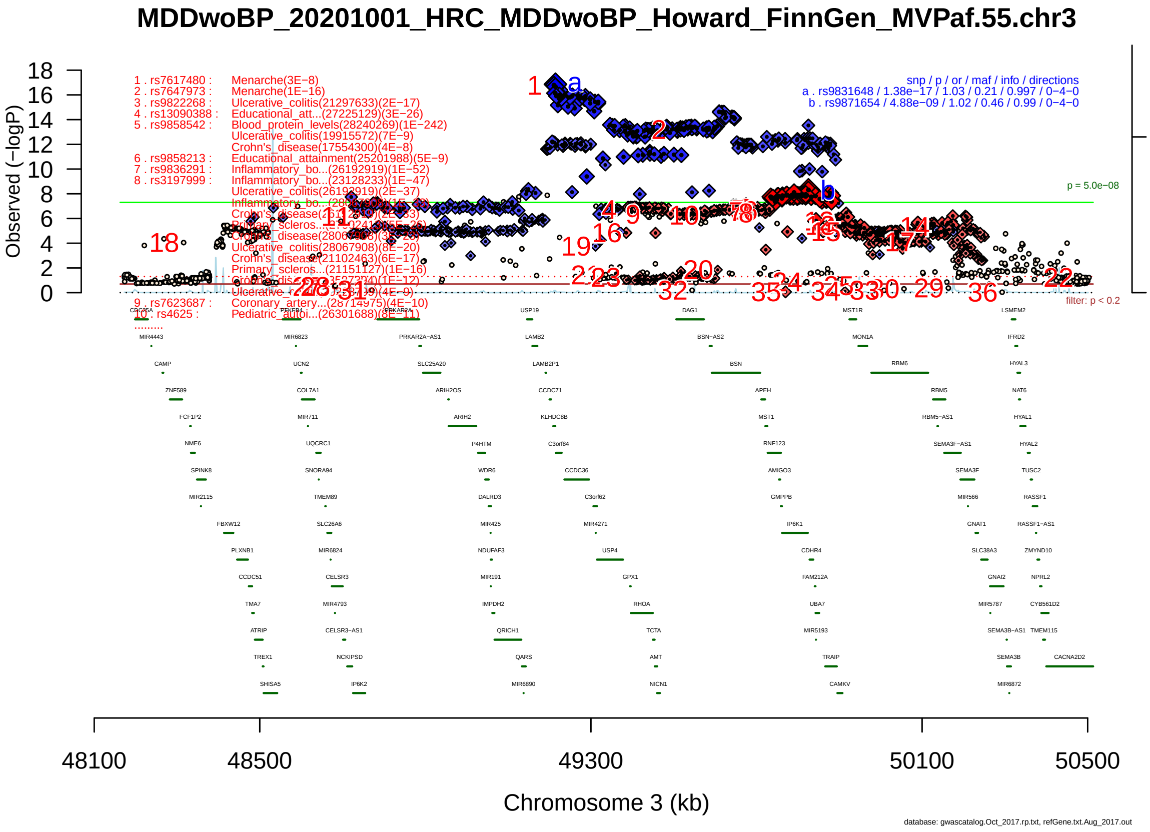

Figure S2-13: Regional plot of GWAS locus No. 13 , with rs9831648 as lead SNP.

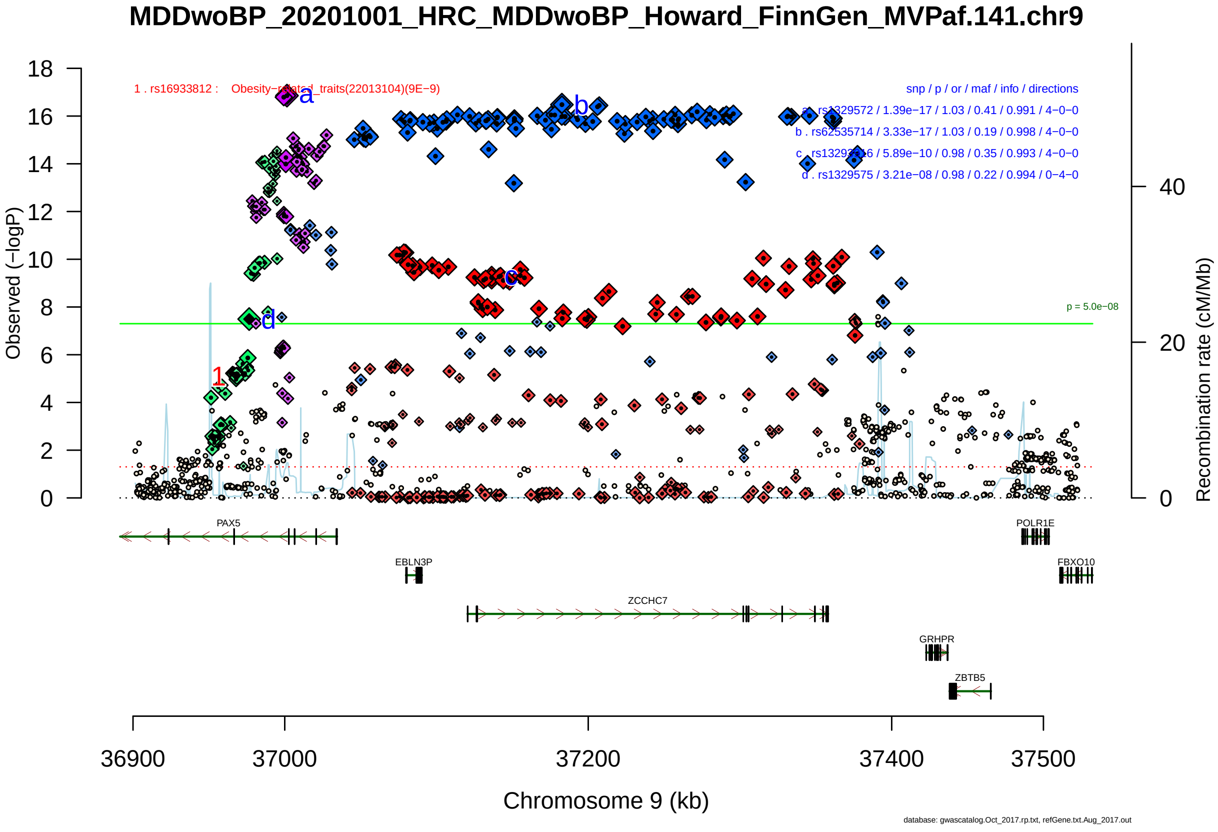

Figure S2-14: Regional plot of GWAS locus No. 14 , with rs1329572 as lead SNP.

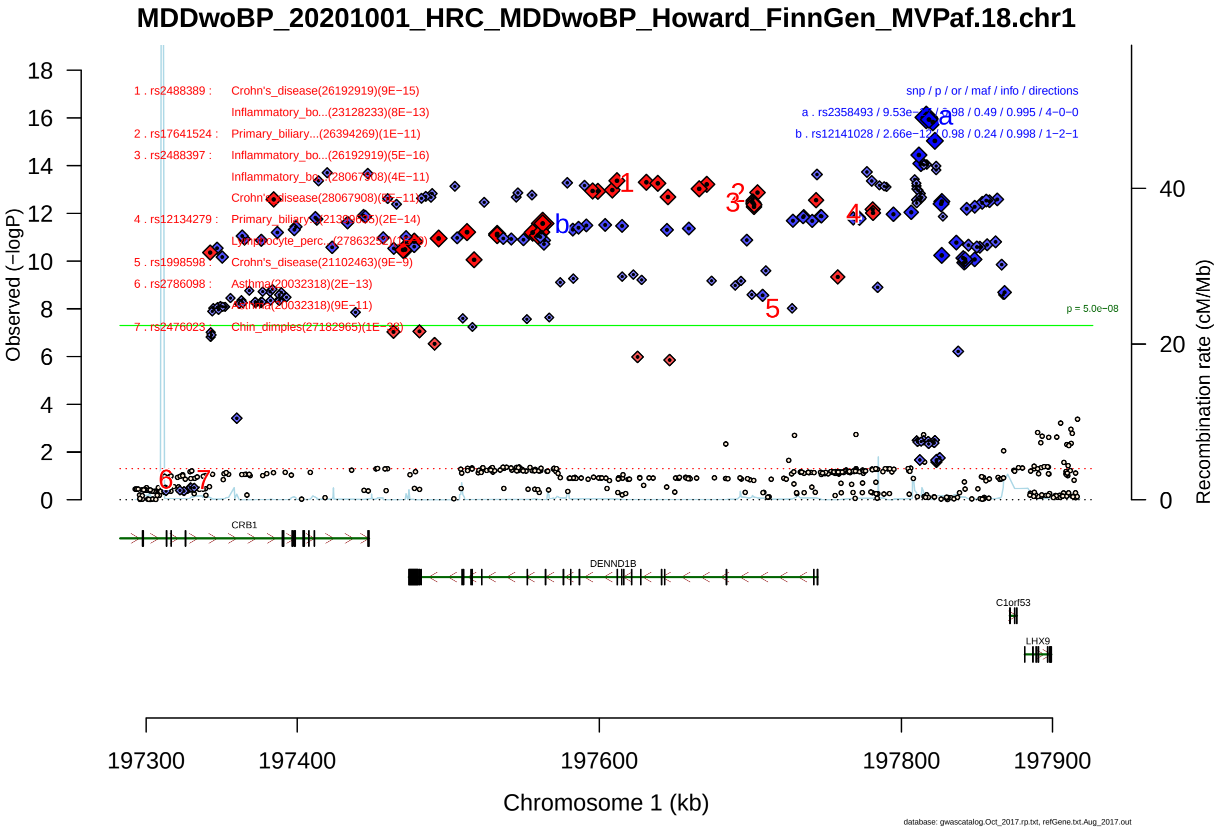

Figure S2-15: Regional plot of GWAS locus No. 15 , with rs2358493 as lead SNP.

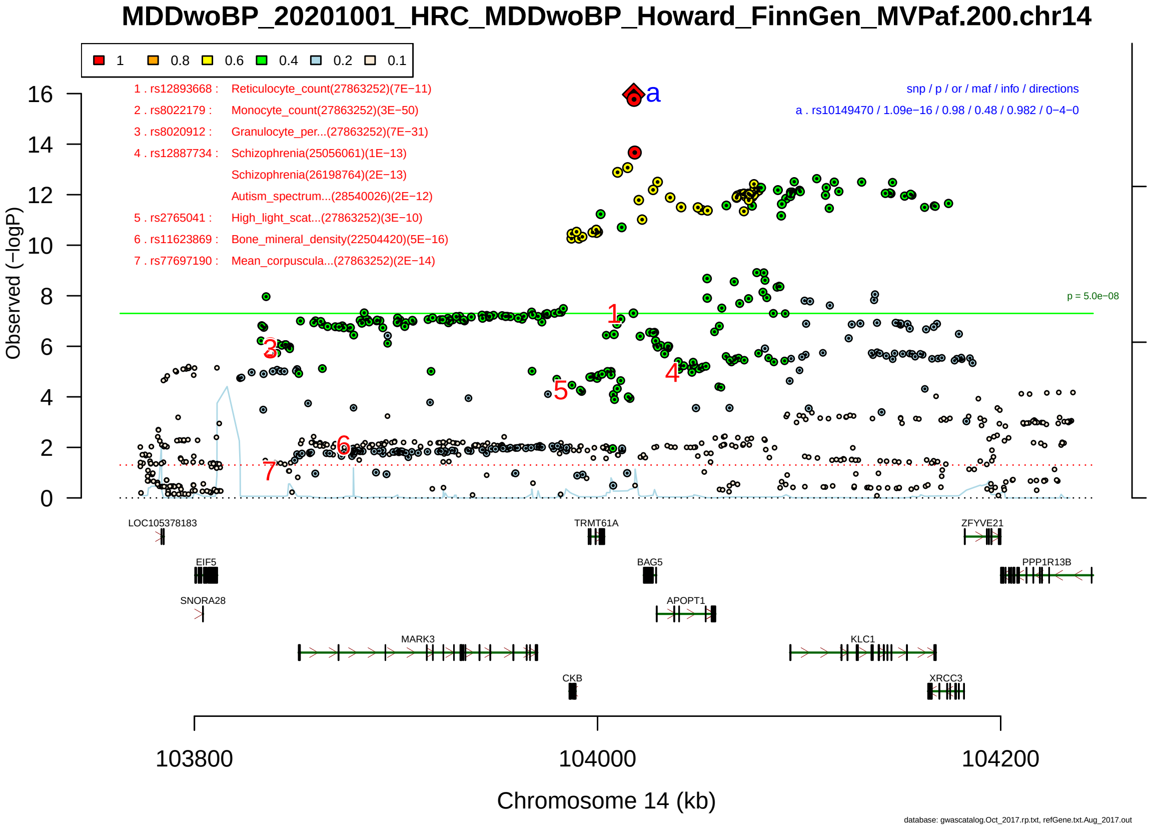

Figure S2-16: Regional plot of GWAS locus No. 16 , with rs10149470 as lead SNP.

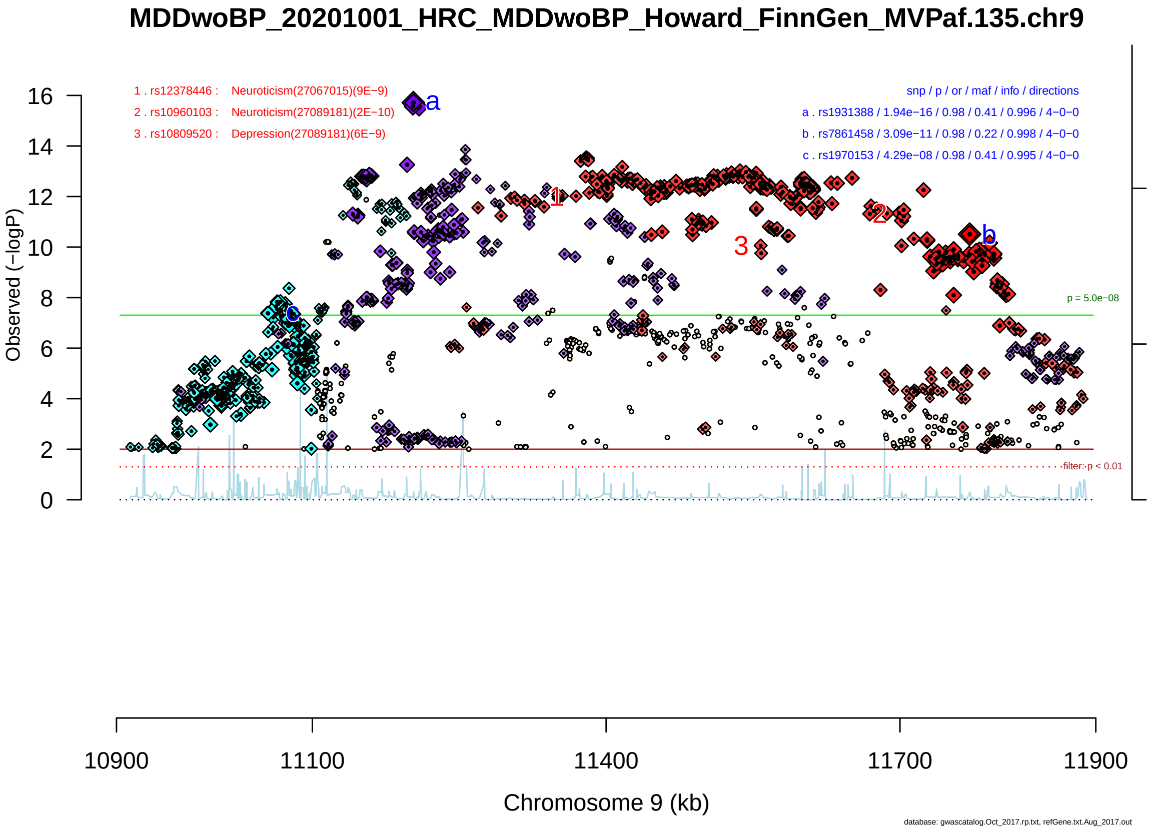

Figure S2-17: Regional plot of GWAS locus No. 17 , with rs1931388 as lead SNP.

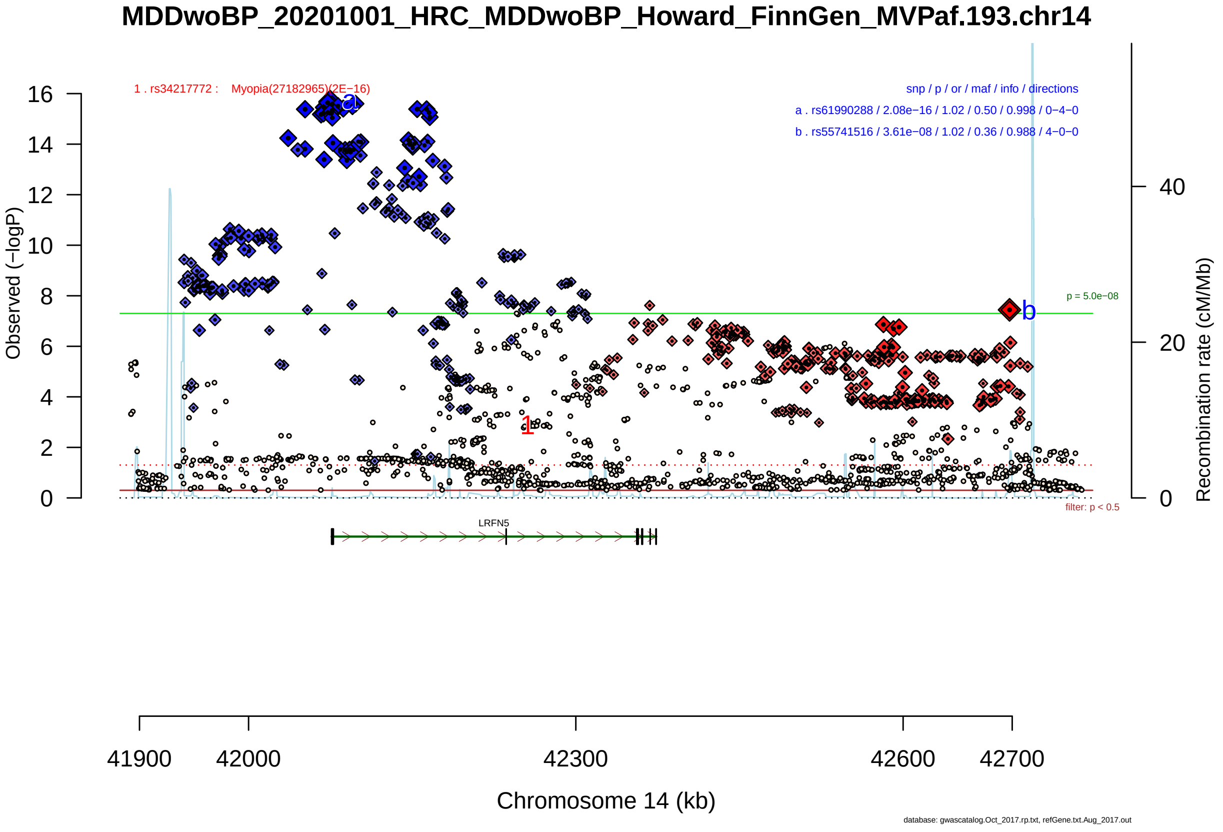

Figure S2-18: Regional plot of GWAS locus No. 18 , with rs61990288 as lead SNP.

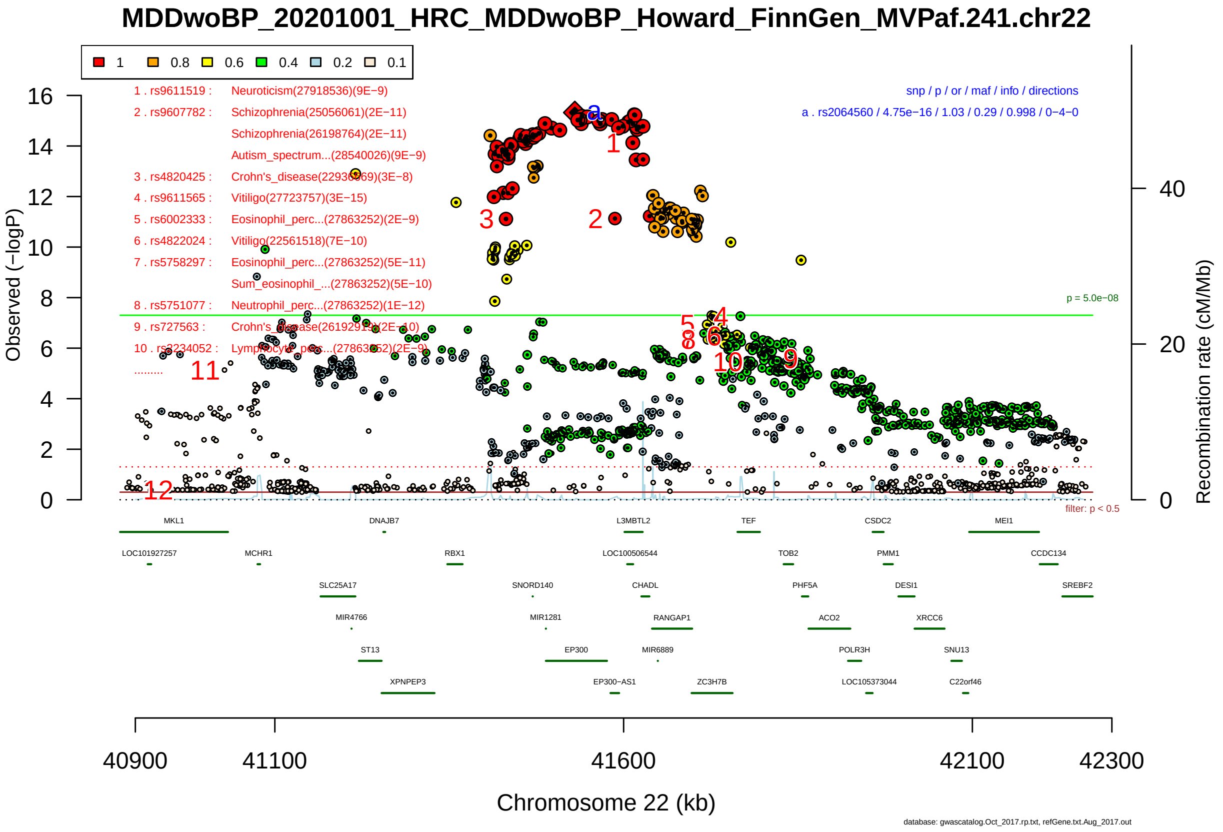

Figure S2-19: Regional plot of GWAS locus No. 19 , with rs2064560 as lead SNP.

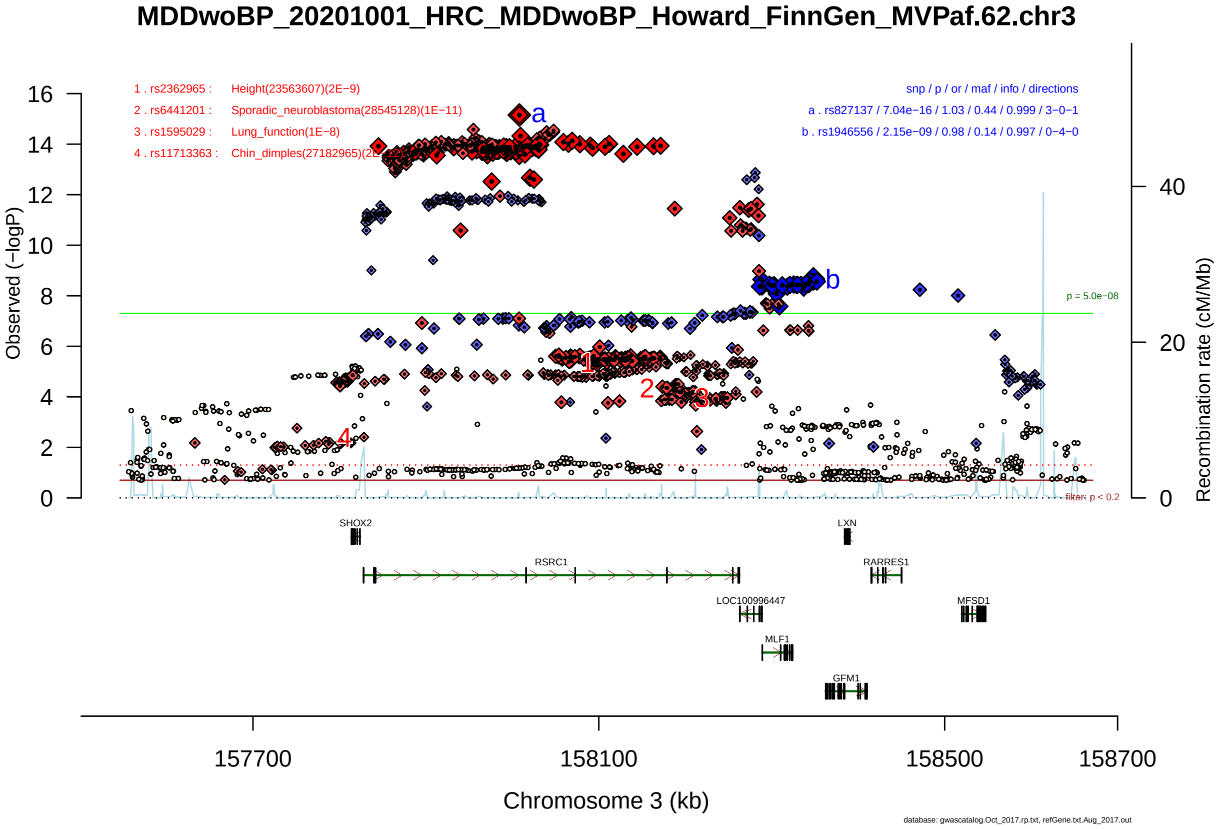

Figure S2-20: Regional plot of GWAS locus No. 20 , with rs827137 as lead SNP.

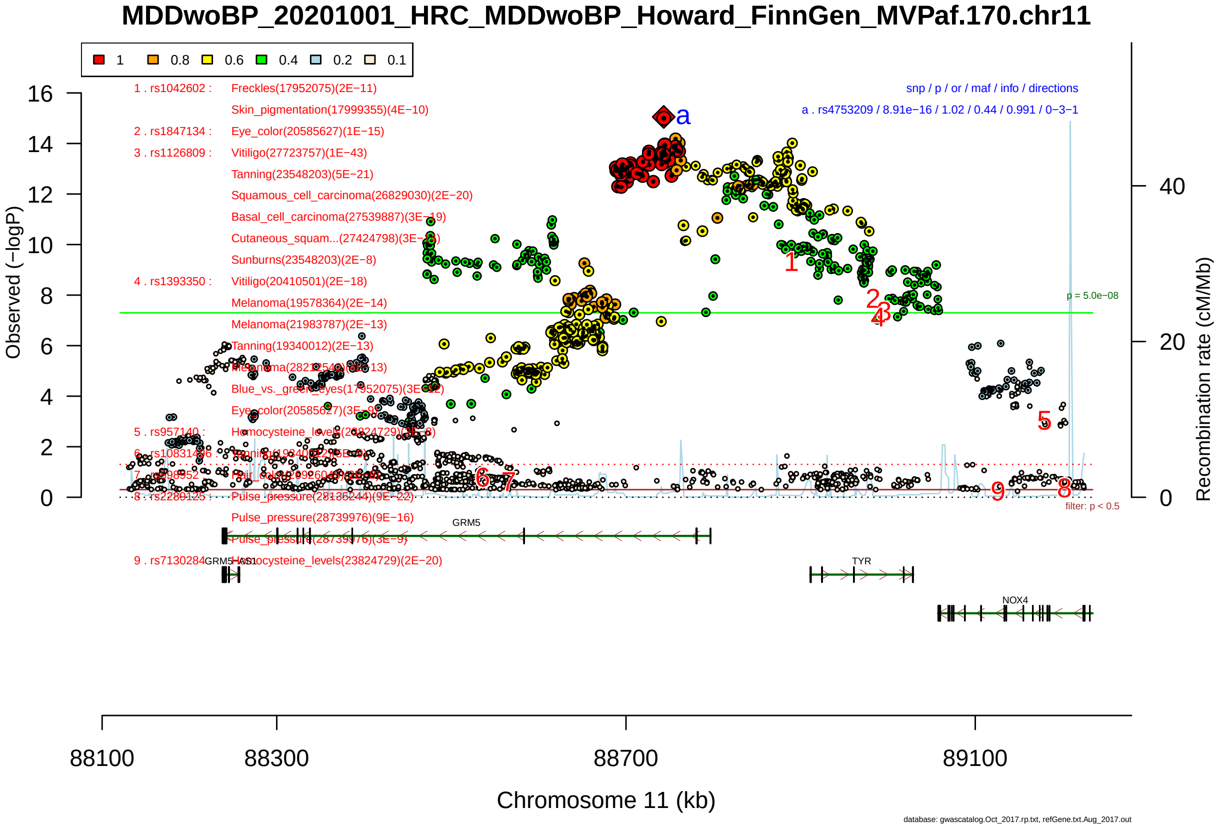

Figure S2-21: Regional plot of GWAS locus No. 21 , with rs4753209 as lead SNP.

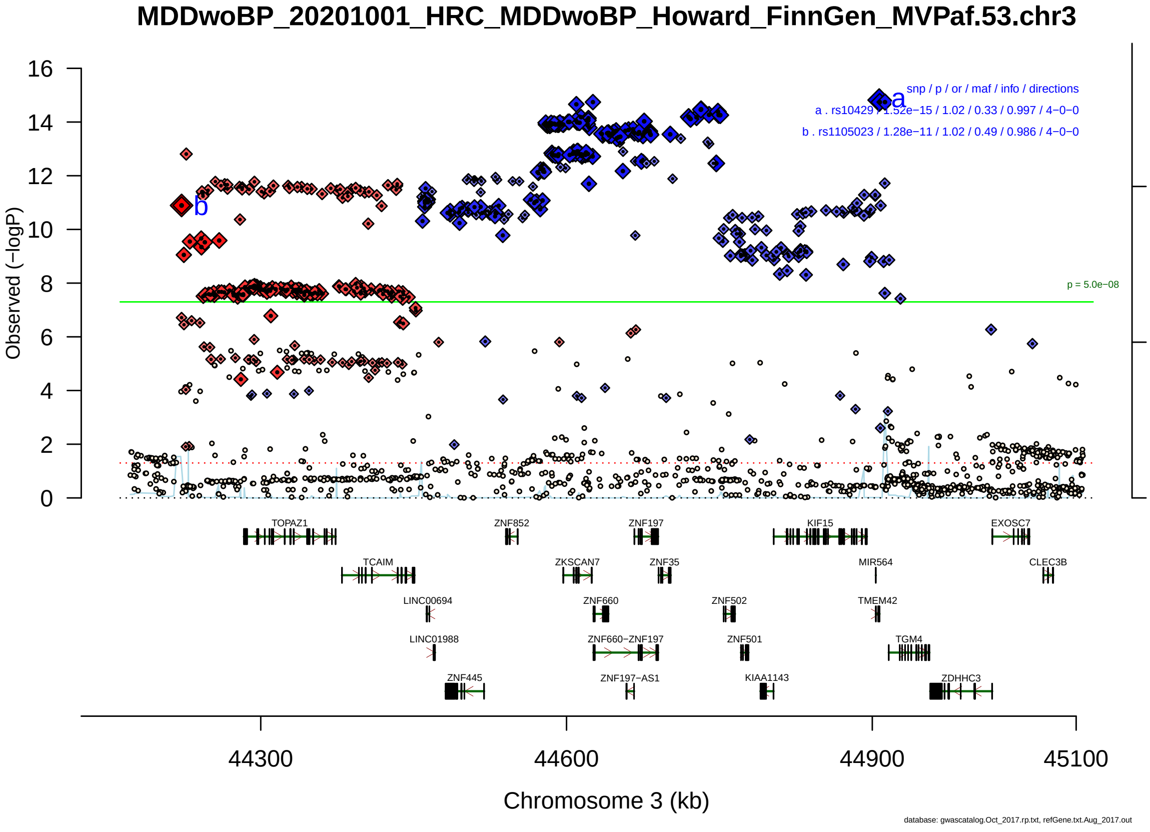

Figure S2-22: Regional plot of GWAS locus No. 22 , with rs10429 as lead SNP.

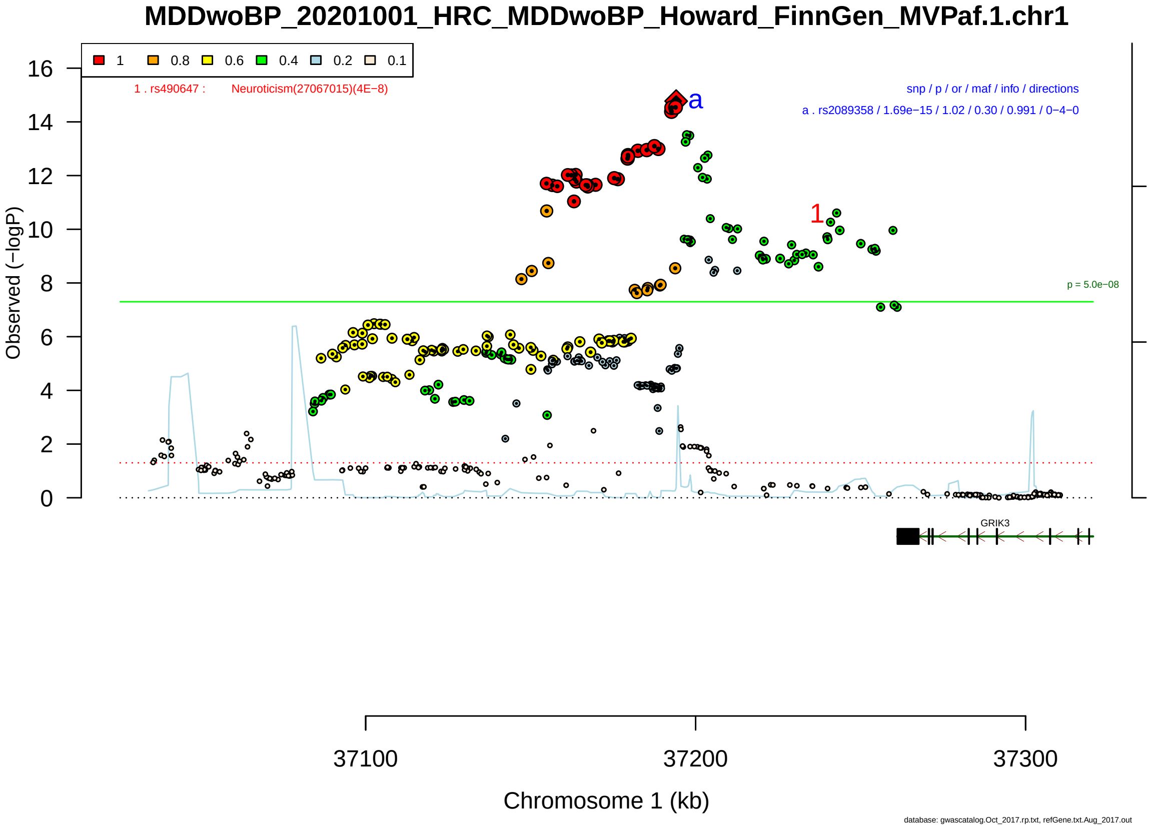

Figure S2-23: Regional plot of GWAS locus No. 23 , with rs2089358 as lead SNP.

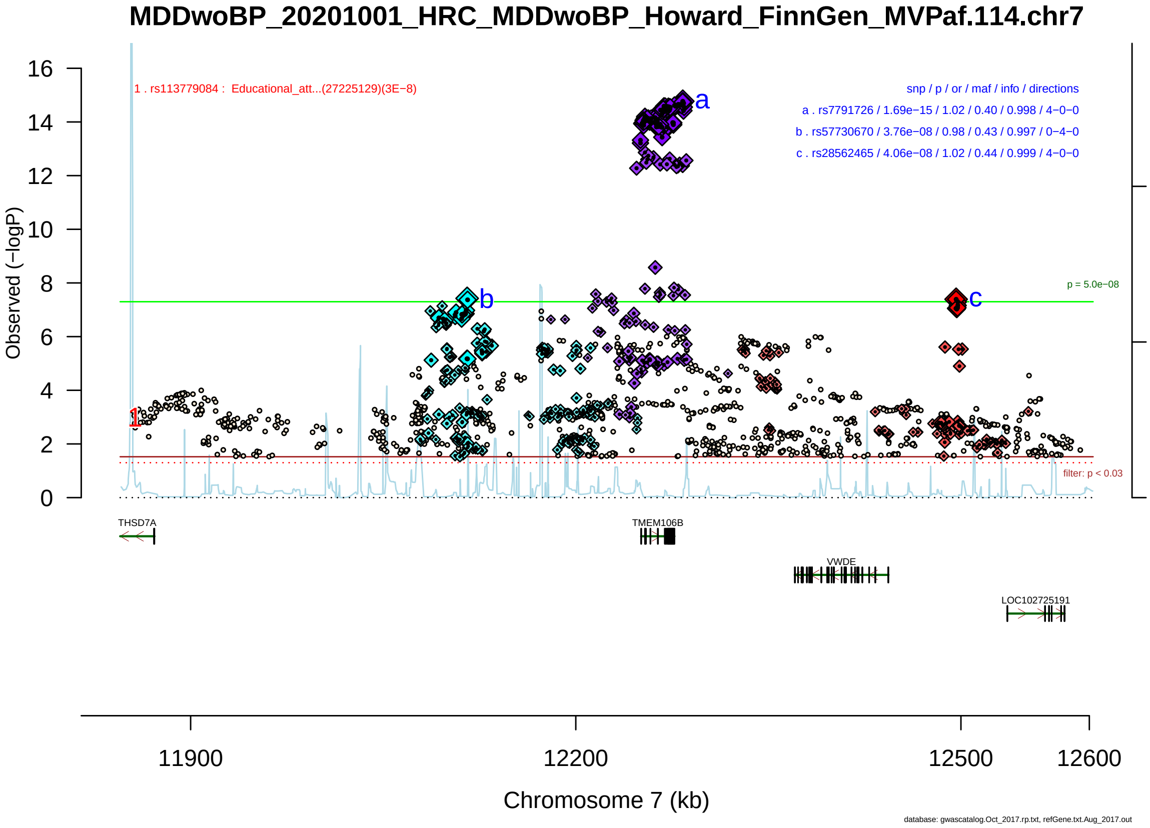

Figure S2-24: Regional plot of GWAS locus No. 24 , with rs7791726 as lead SNP.

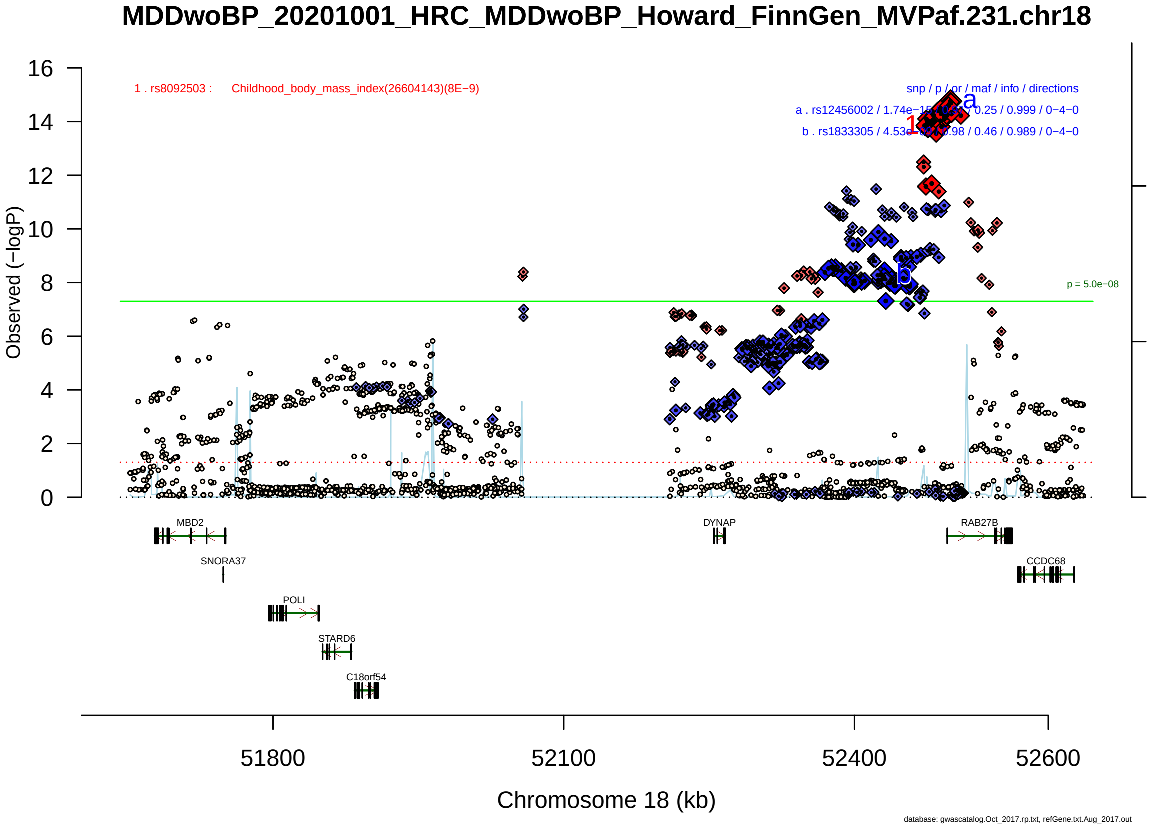

Figure S2-25: Regional plot of GWAS locus No. 25 , with rs12456002 as lead SNP.

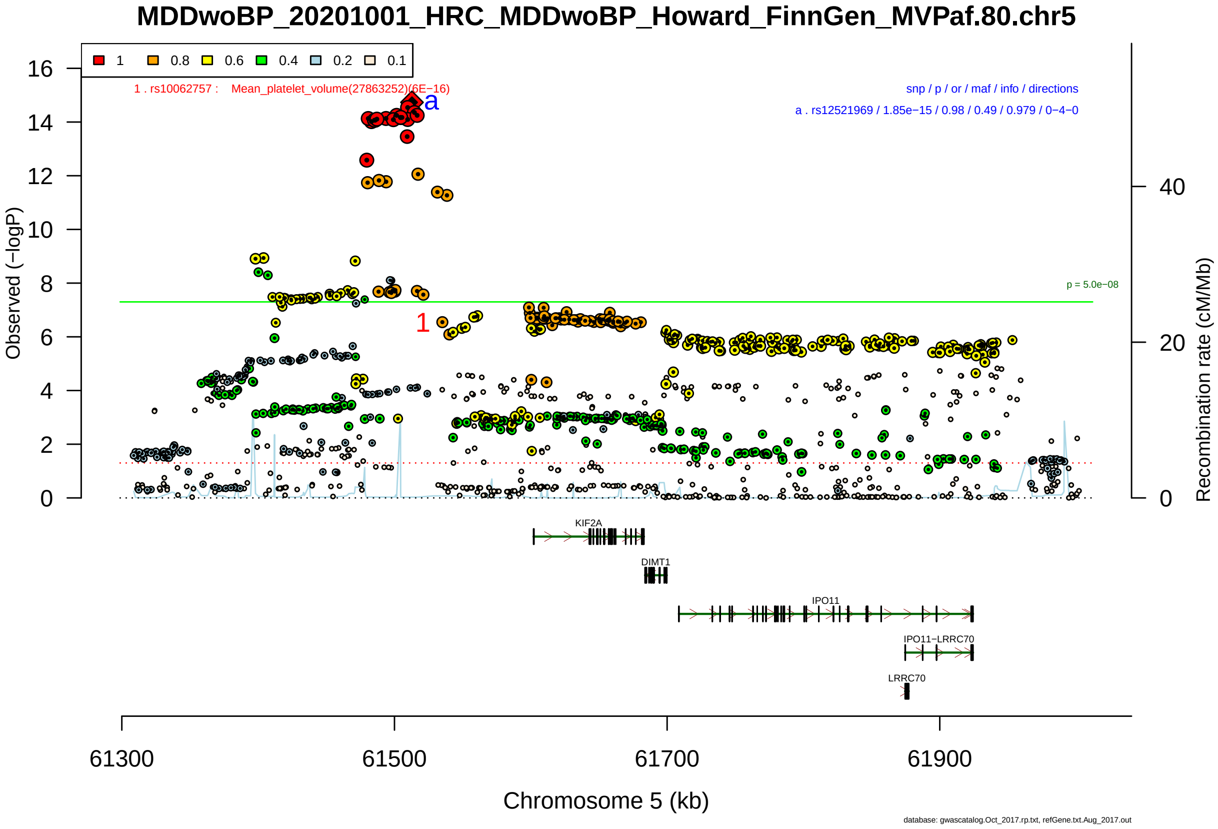

Figure S2-26: Regional plot of GWAS locus No. 26 , with rs12521969 as lead SNP.

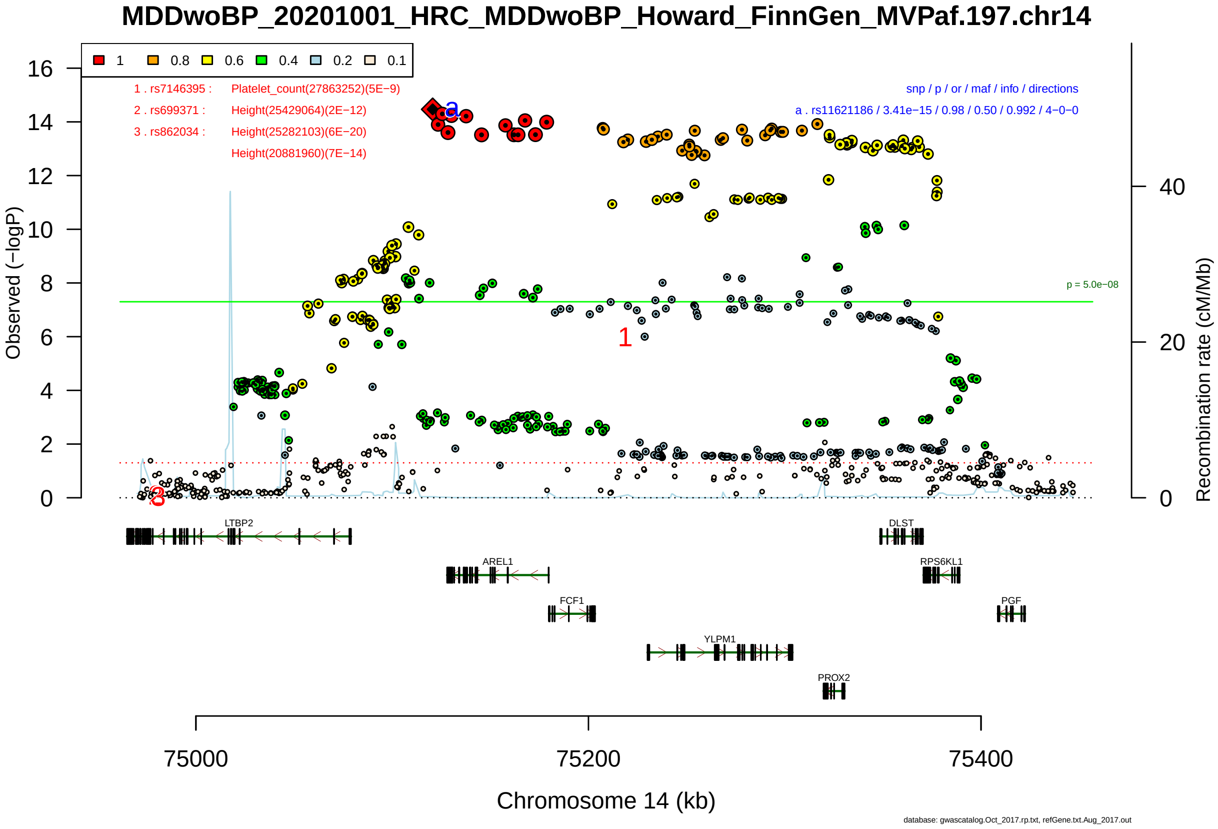

Figure S2-27: Regional plot of GWAS locus No. 27 , with rs11621186 as lead SNP.

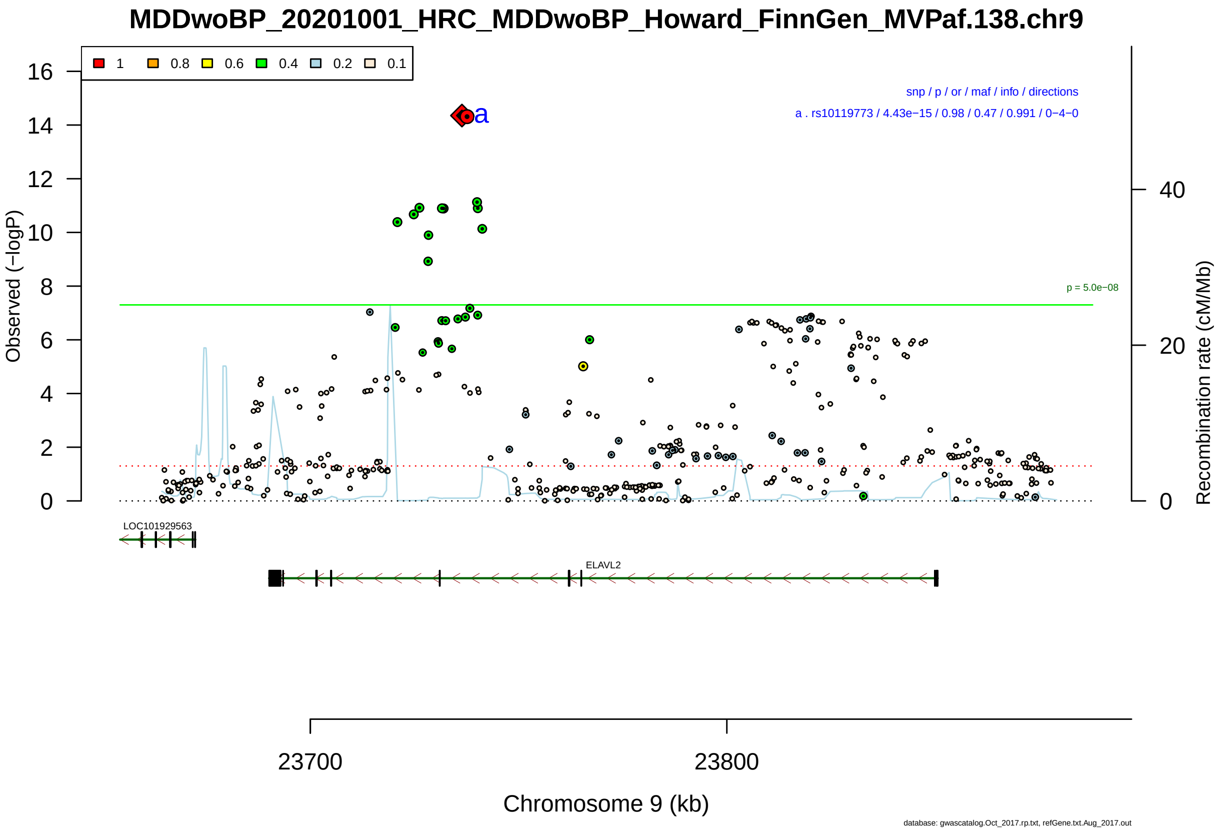

Figure S2-28: Regional plot of GWAS locus No. 28 , with rs10119773 as lead SNP.

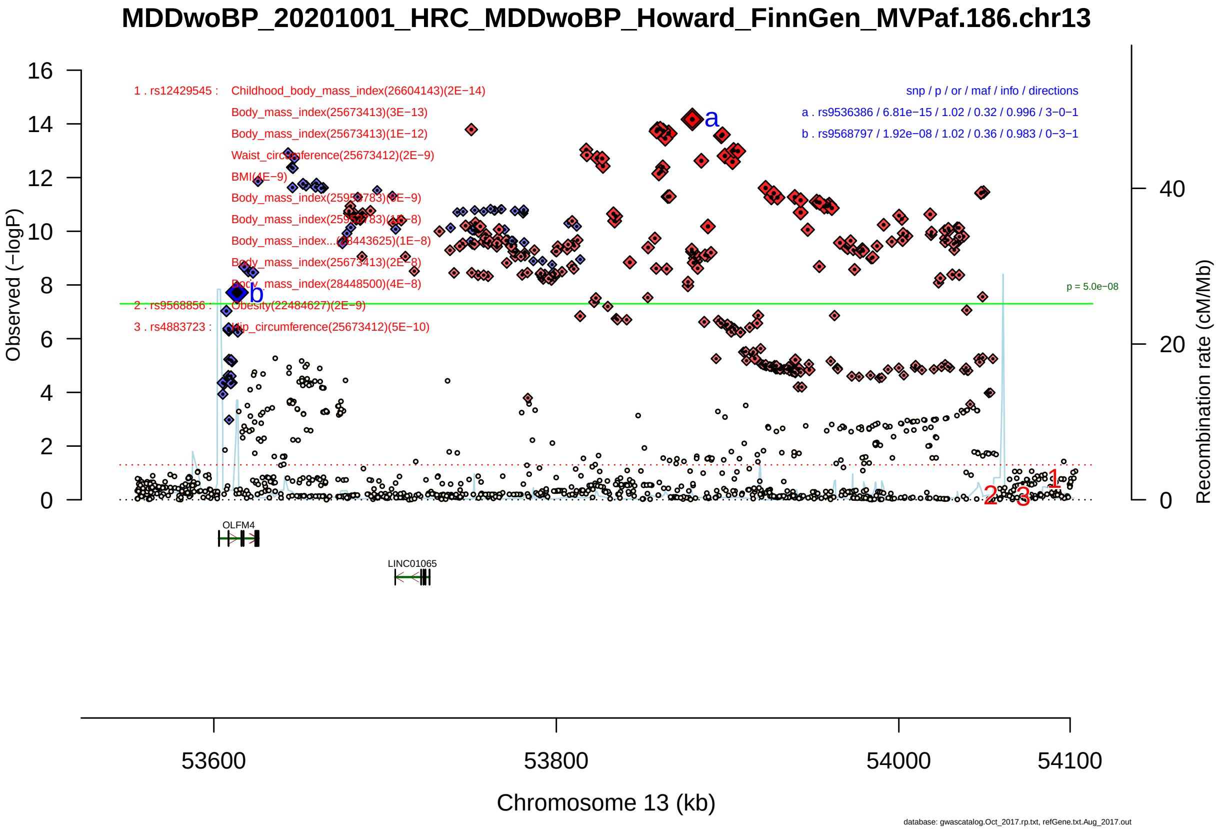

Figure S2-29: Regional plot of GWAS locus No. 29 , with rs9536386 as lead SNP.

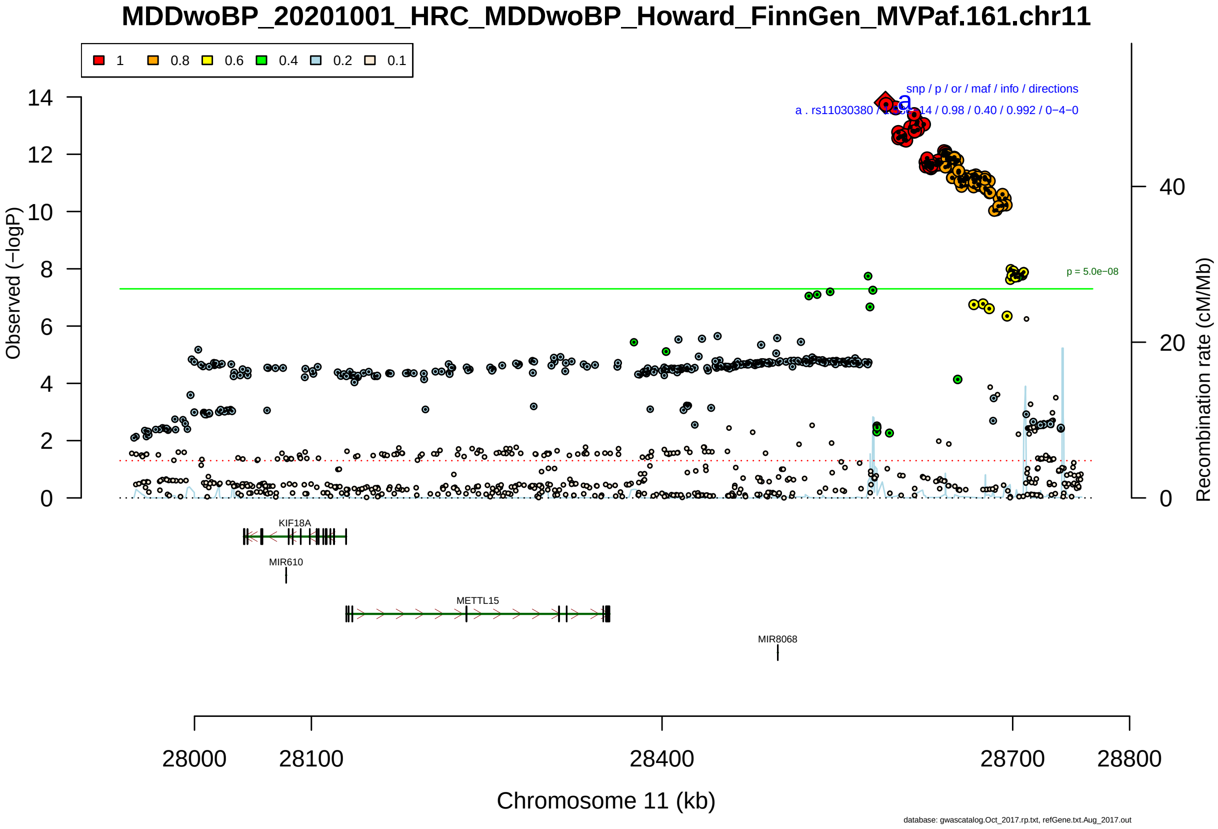

Figure S2-30: Regional plot of GWAS locus No. 30 , with rs11030380 as lead SNP.

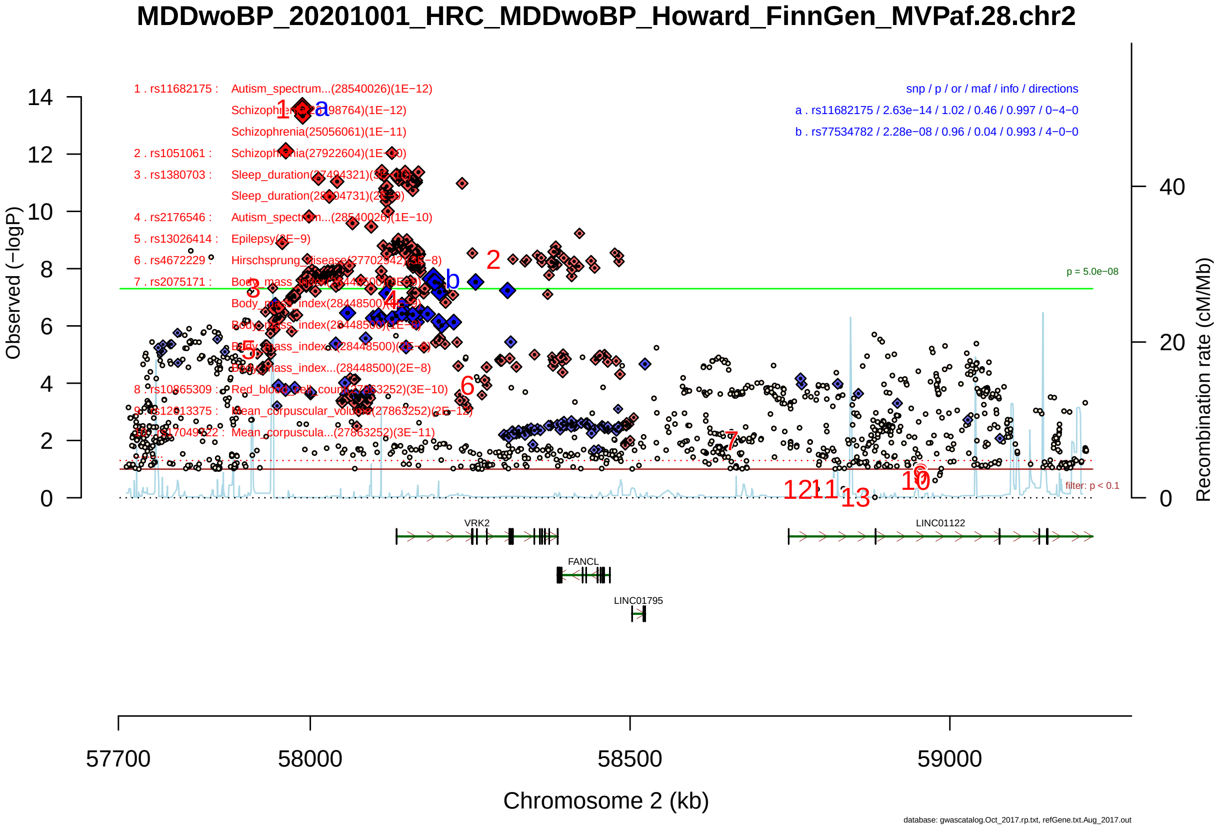

Figure S2-31: Regional plot of GWAS locus No. 31 , with rs11682175 as lead SNP.

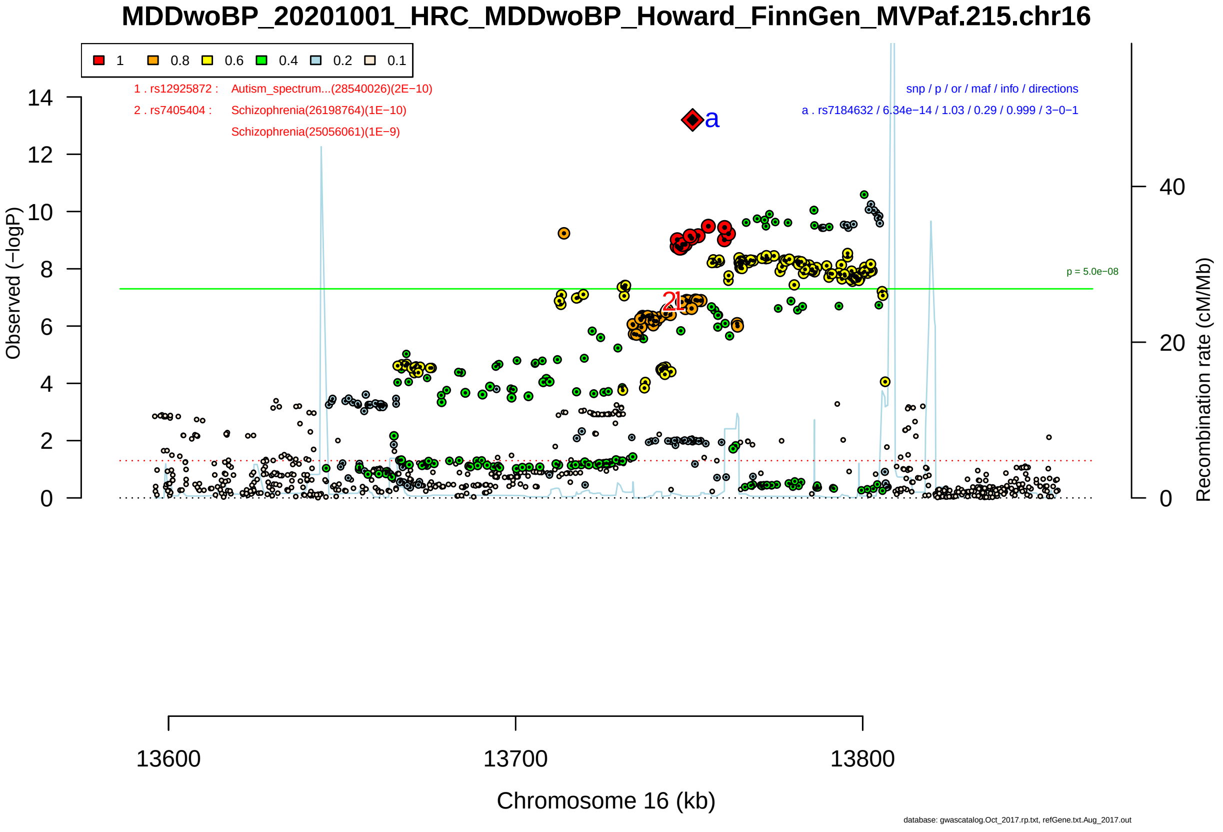

Figure S2-32: Regional plot of GWAS locus No. 32 , with rs7184632 as lead SNP.

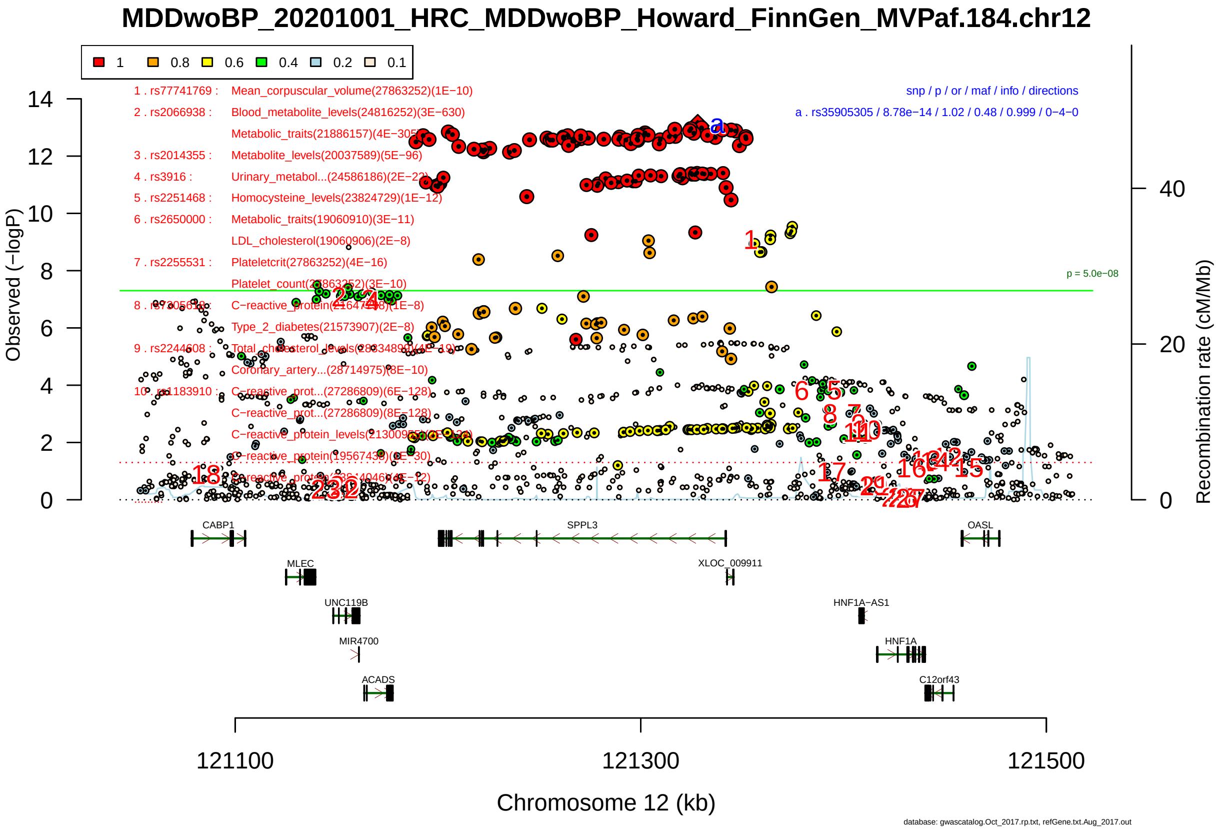

Figure S2-33: Regional plot of GWAS locus No. 33 , with rs35905305 as lead SNP.

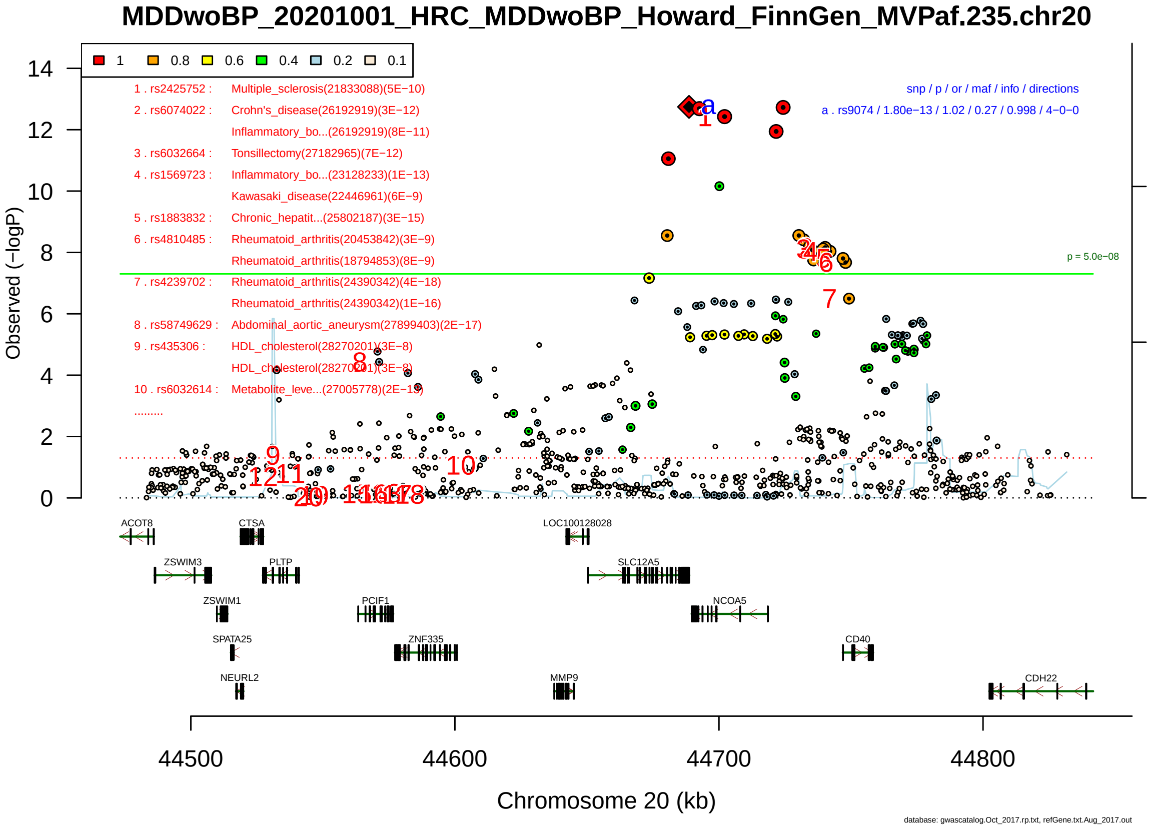

Figure S2-34: Regional plot of GWAS locus No. 34 , with rs9074 as lead SNP.

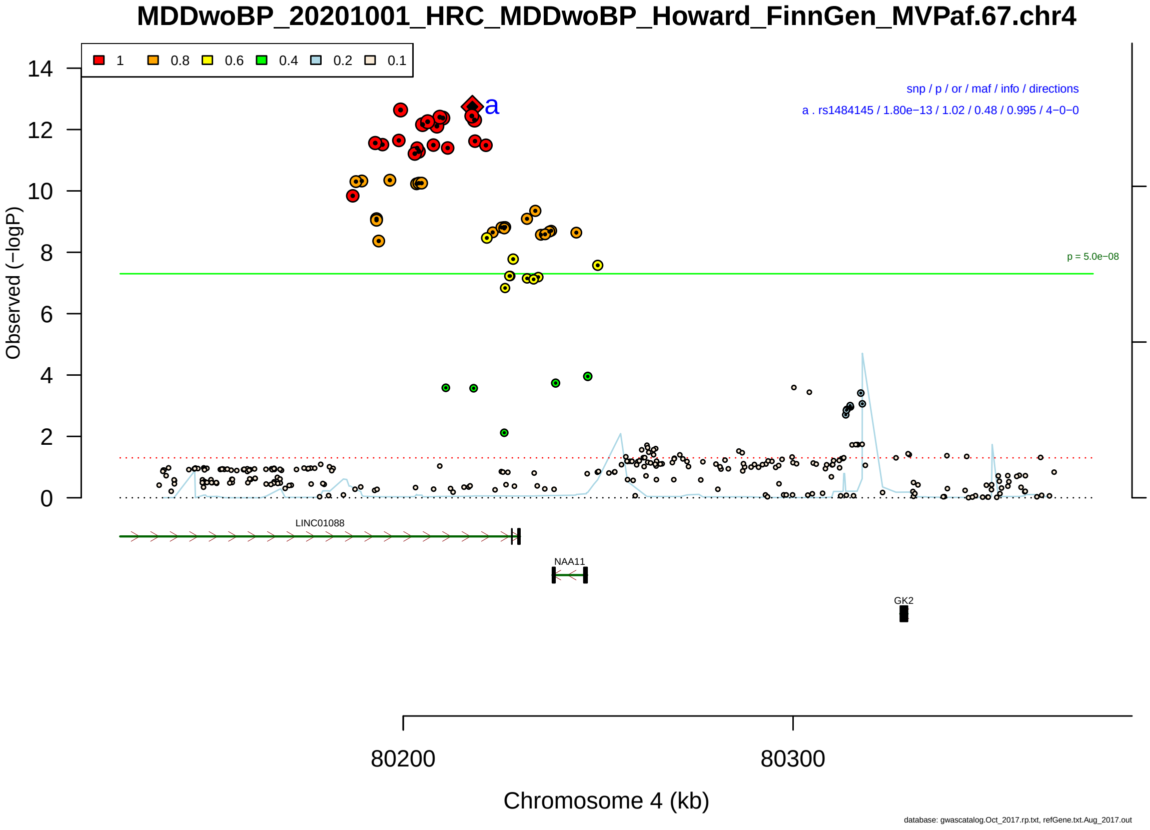

Figure S2-35: Regional plot of GWAS locus No. 35 , with rs1484145 as lead SNP.

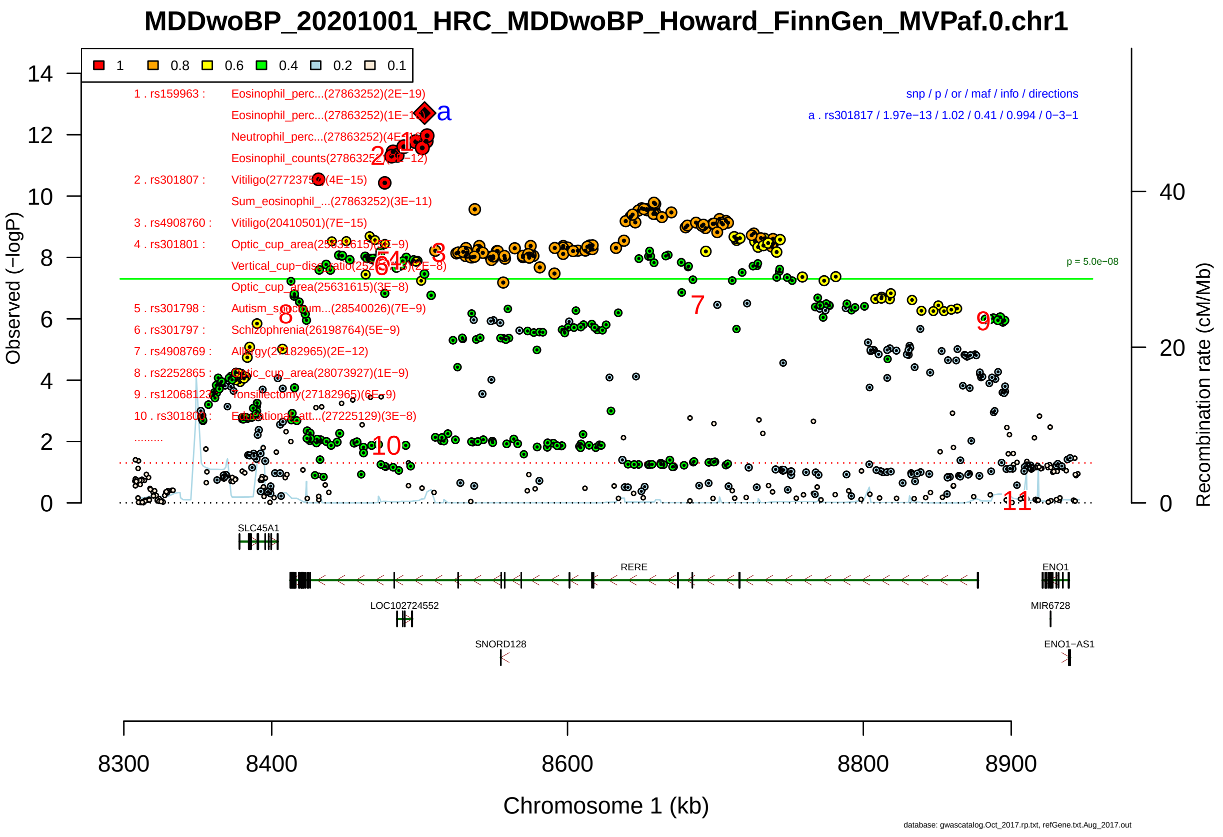

Figure S2-36: Regional plot of GWAS locus No. 36 , with rs301817 as lead SNP.

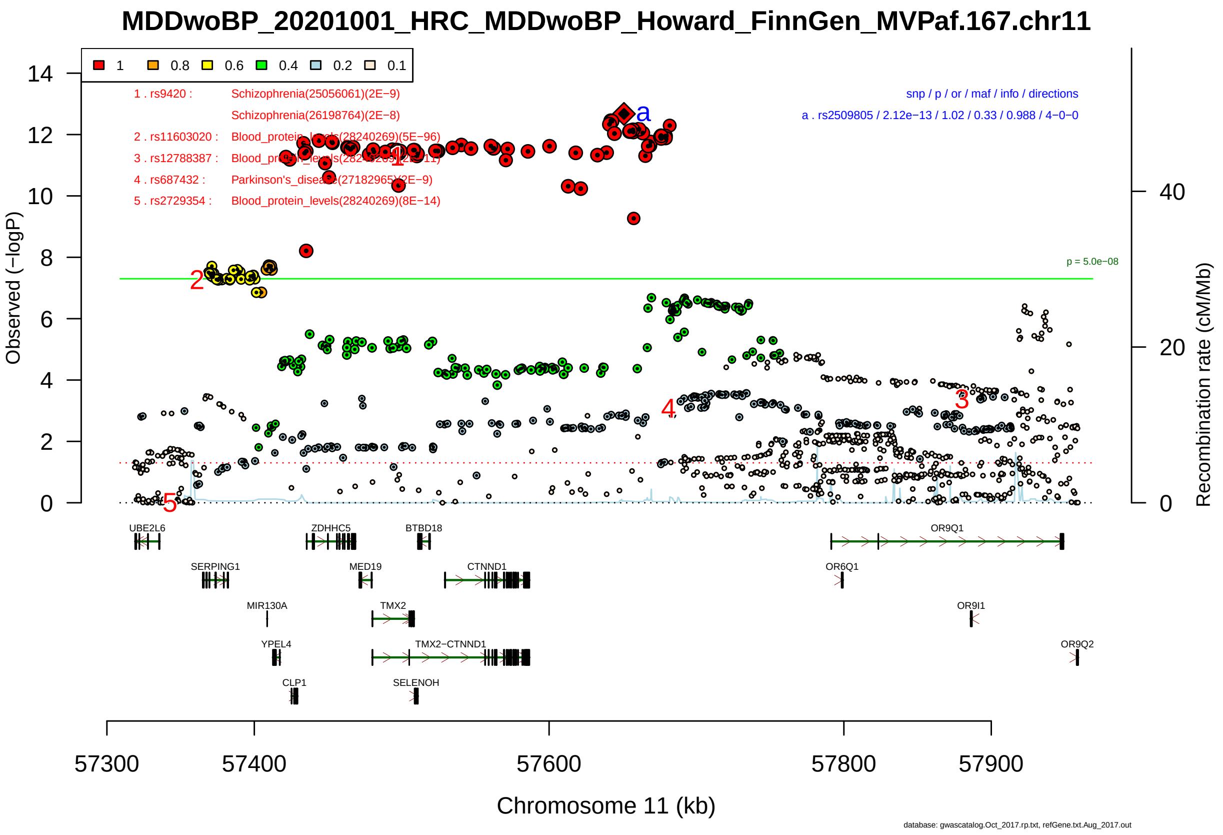

Figure S2-37: Regional plot of GWAS locus No. 37 , with rs2509805 as lead SNP.

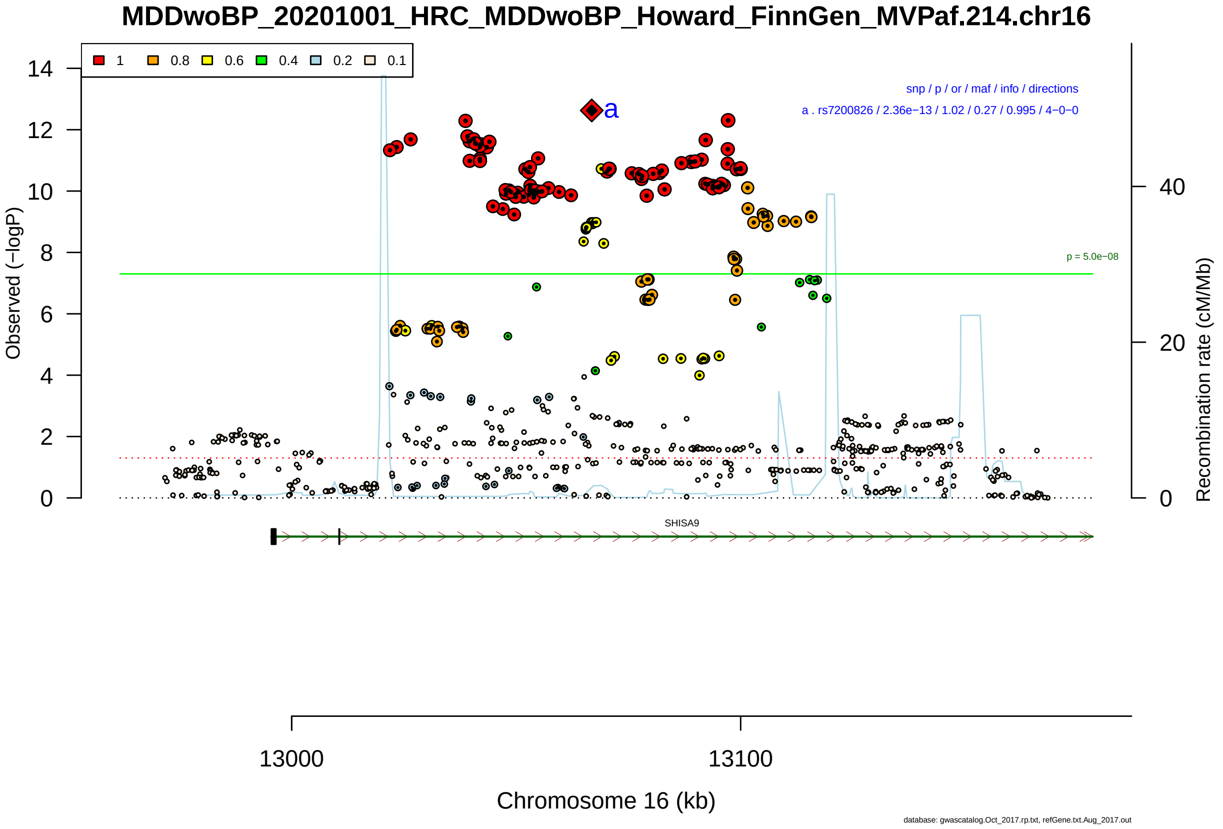

Figure S2-38: Regional plot of GWAS locus No. 38 , with rs7200826 as lead SNP.

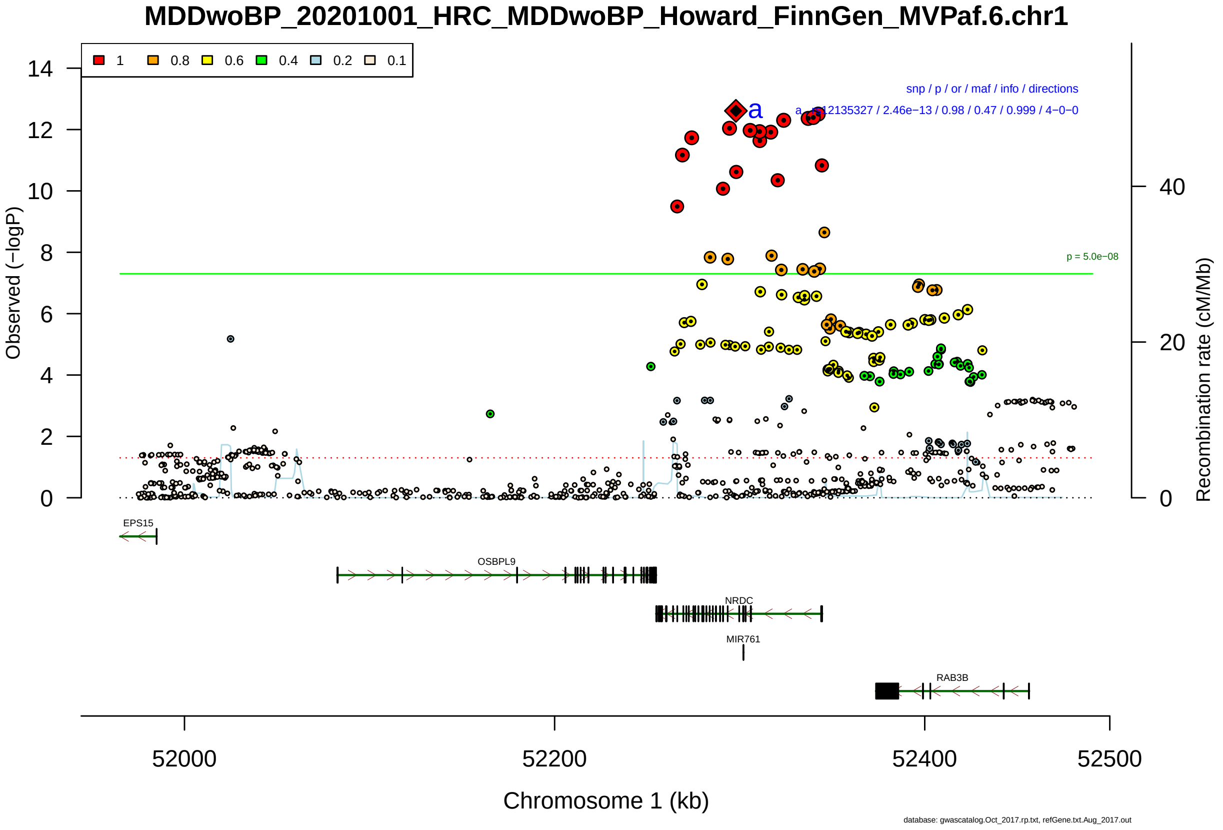

Figure S2-39: Regional plot of GWAS locus No. 39 , with rs12135327 as lead SNP.

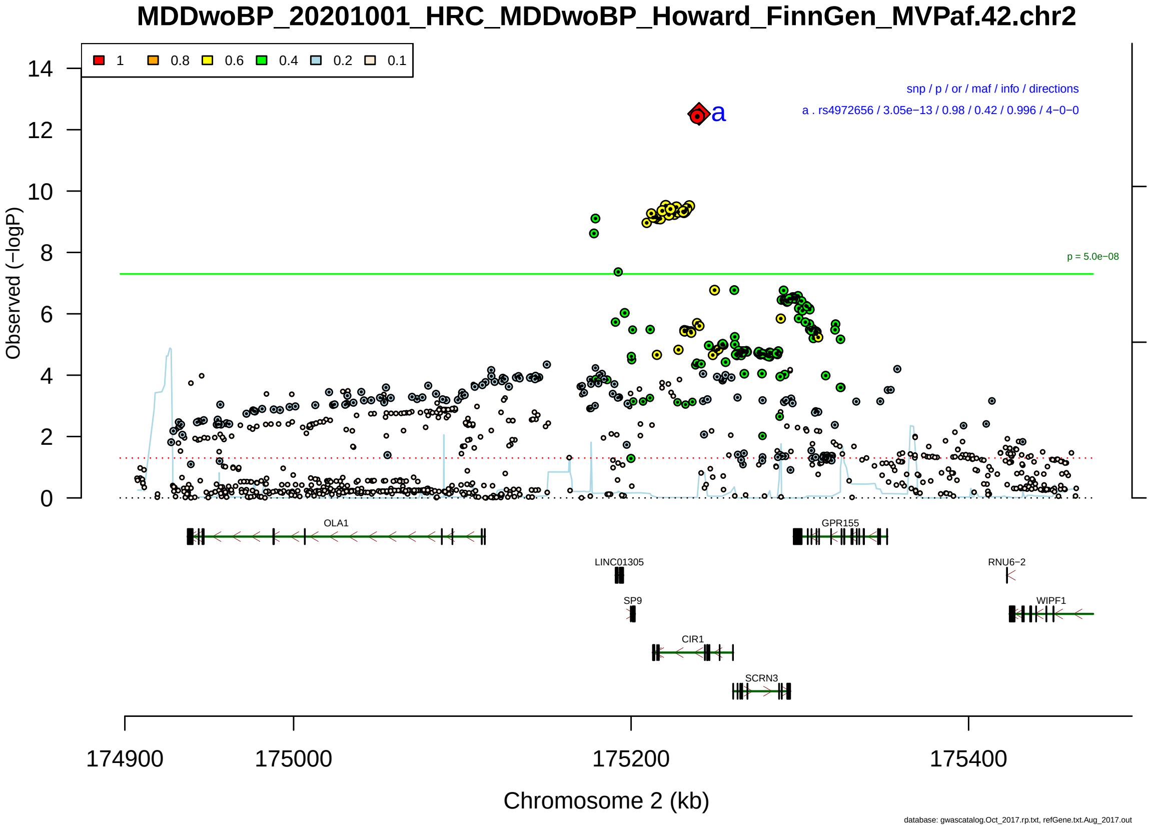

Figure S2-40: Regional plot of GWAS locus No. 40 , with rs4972656 as lead SNP.

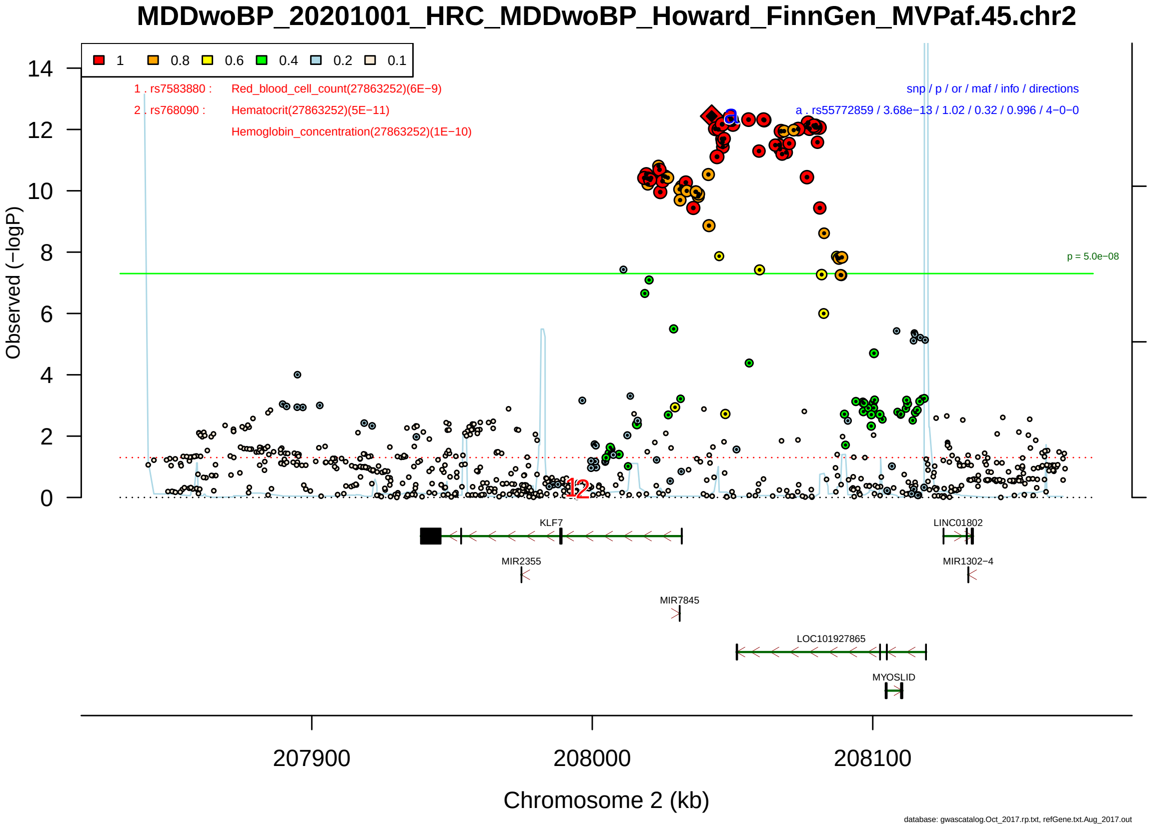

Figure S2-41: Regional plot of GWAS locus No. 41 , with rs55772859 as lead SNP.

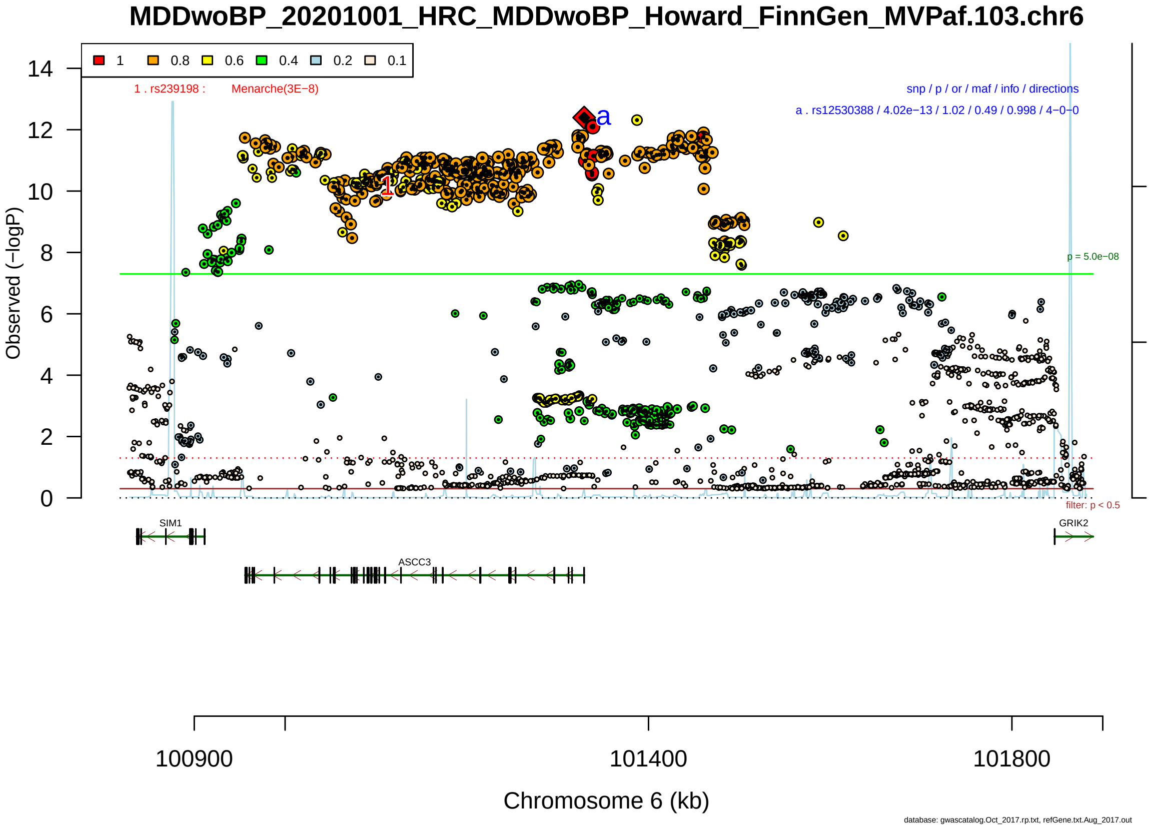

Figure S2-42: Regional plot of GWAS locus No. 42 , with rs12530388 as lead SNP.

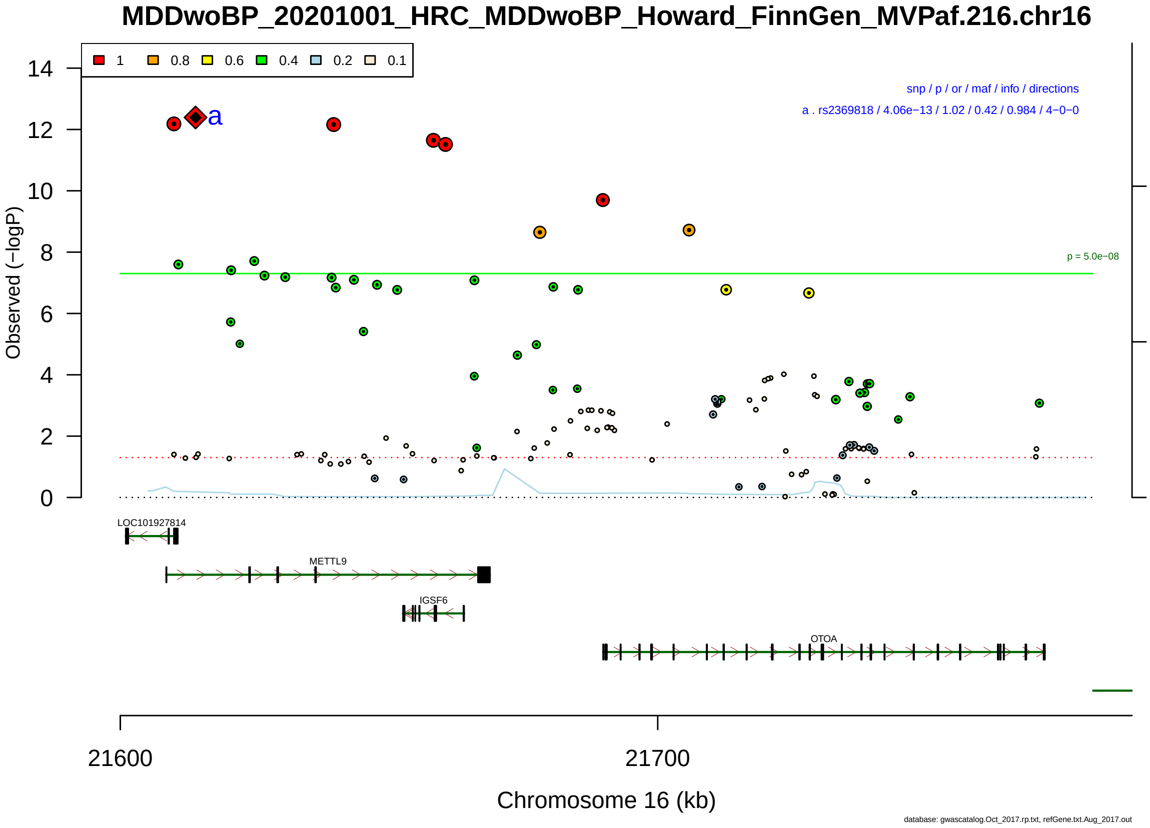

Figure S2-43: Regional plot of GWAS locus No. 43 , with rs2369818 as lead SNP.

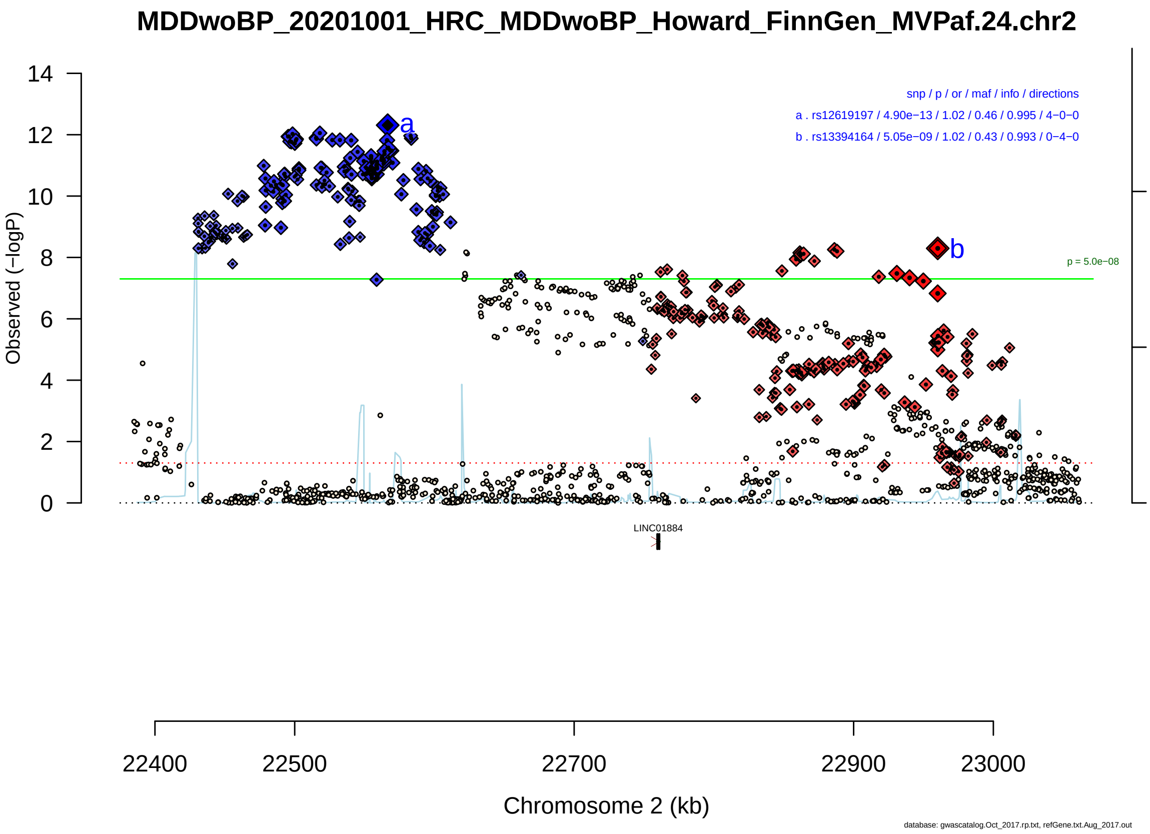

Figure S2-44: Regional plot of GWAS locus No. 44 , with rs12619197 as lead SNP.

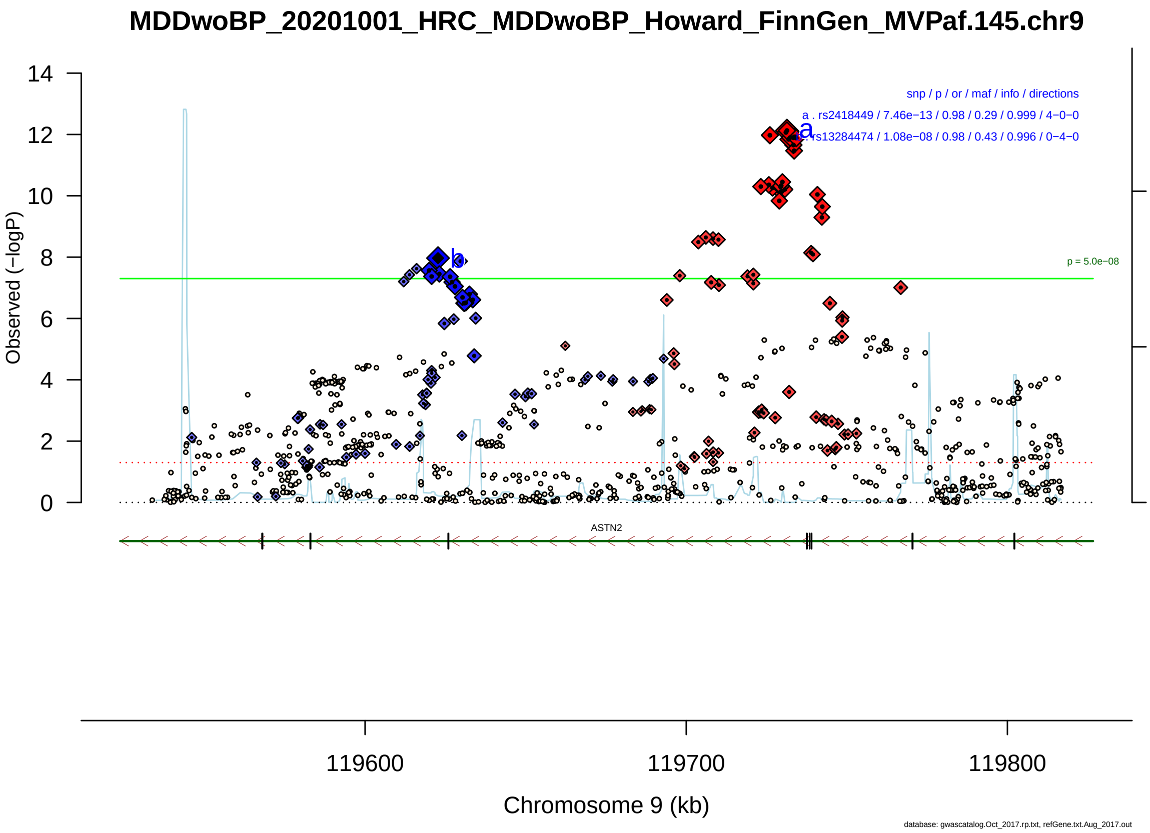

Figure S2-45: Regional plot of GWAS locus No. 45 , with rs2418449 as lead SNP.

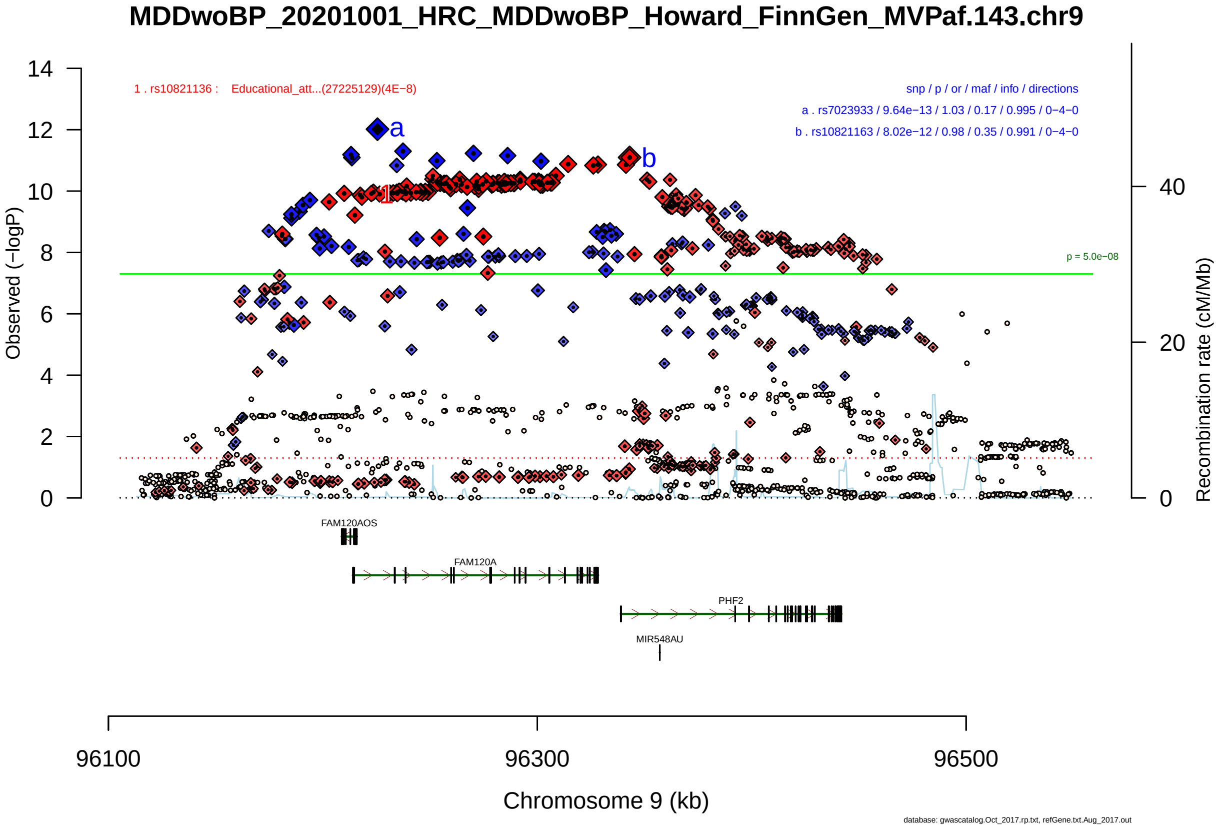

Figure S2-46: Regional plot of GWAS locus No. 46 , with rs7023933 as lead SNP.

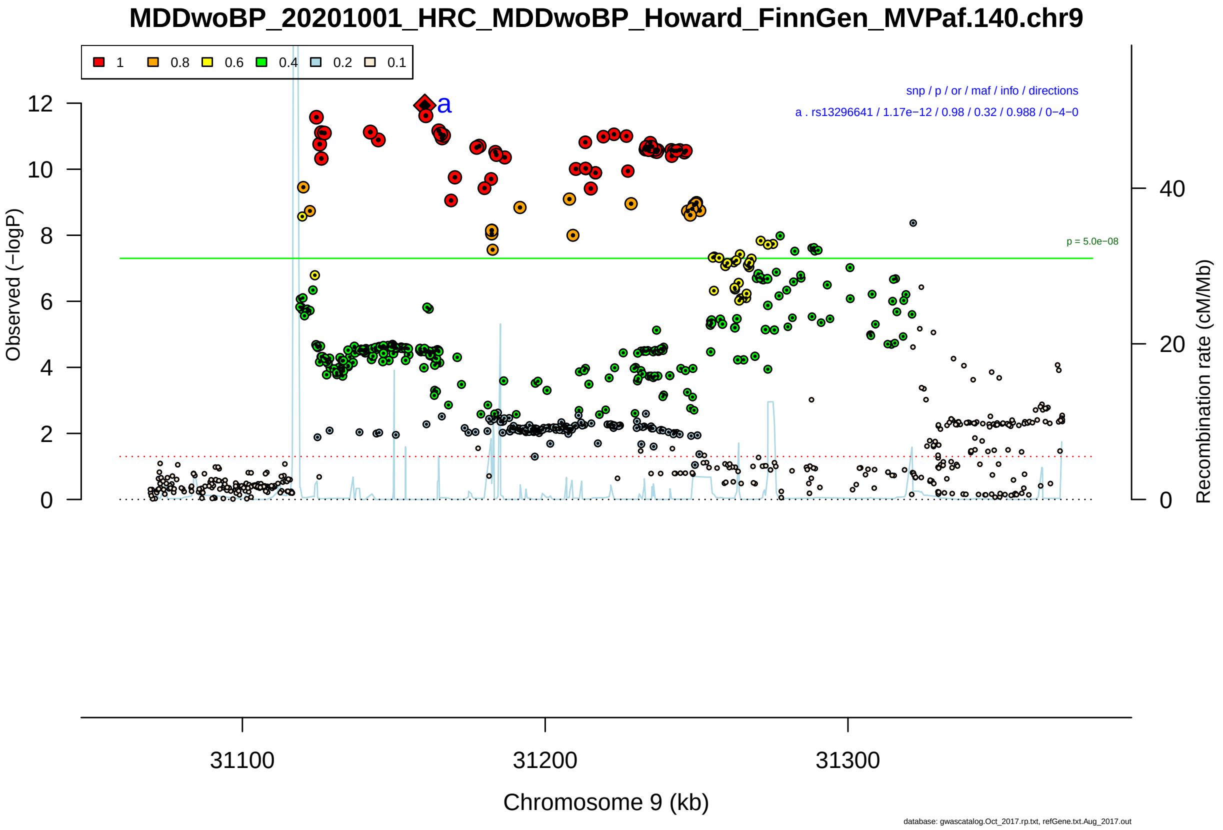

Figure S2-47: Regional plot of GWAS locus No. 47 , with rs13296641 as lead SNP.

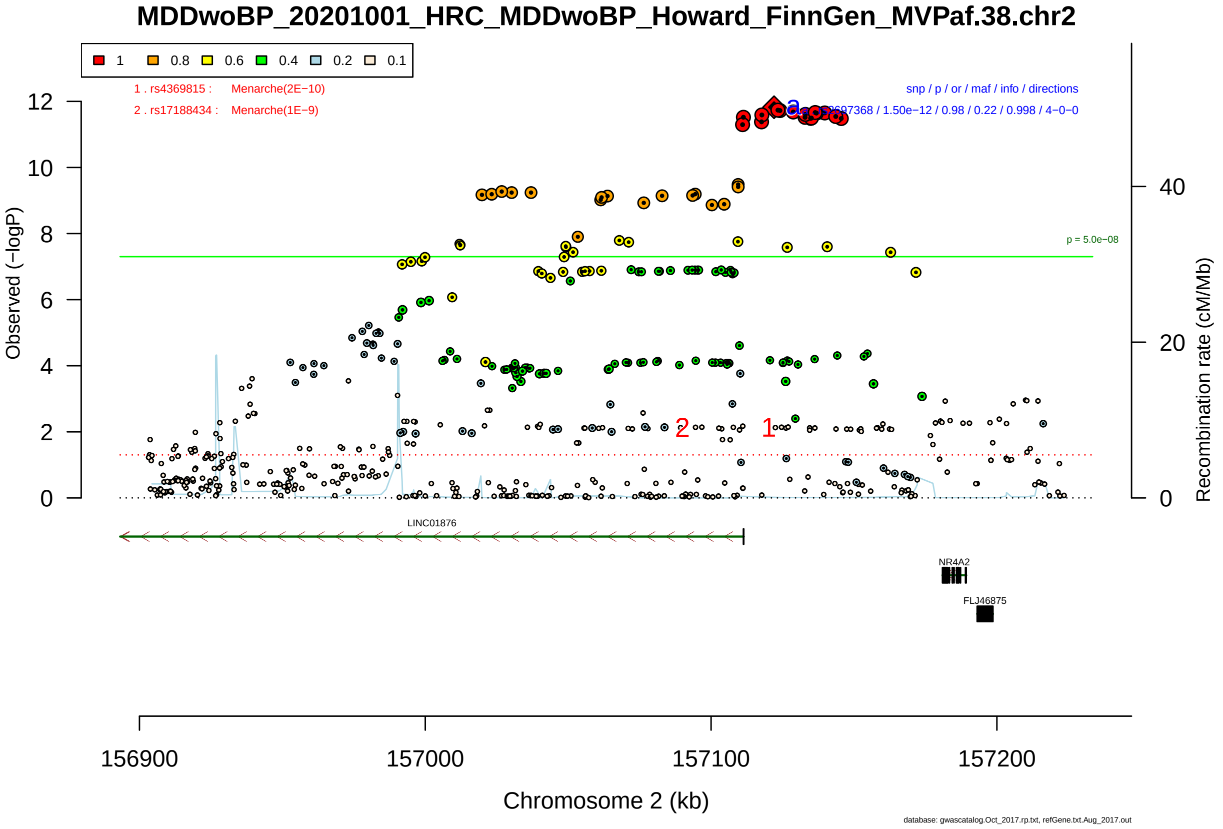

Figure S2-48: Regional plot of GWAS locus No. 48 , with rs2697368 as lead SNP.

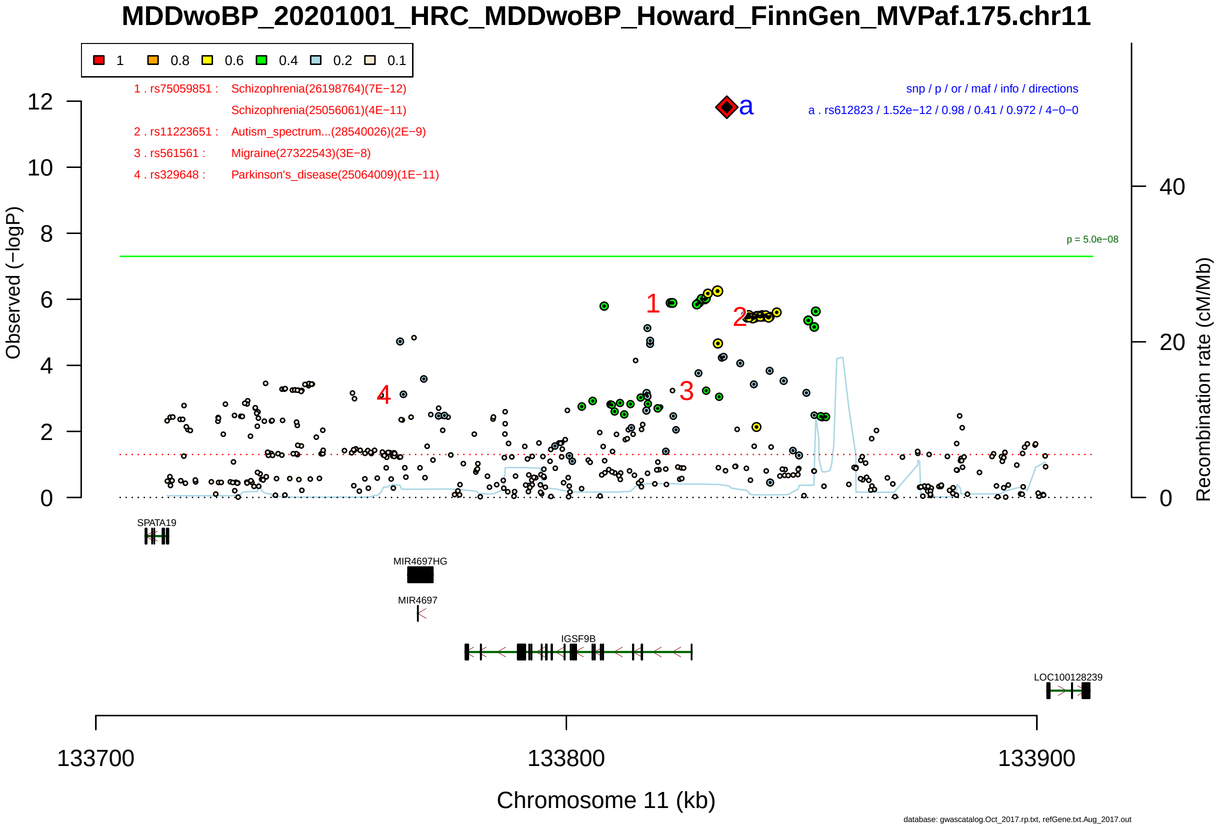

Figure S2-49: Regional plot of GWAS locus No. 49 , with rs612823 as lead SNP.

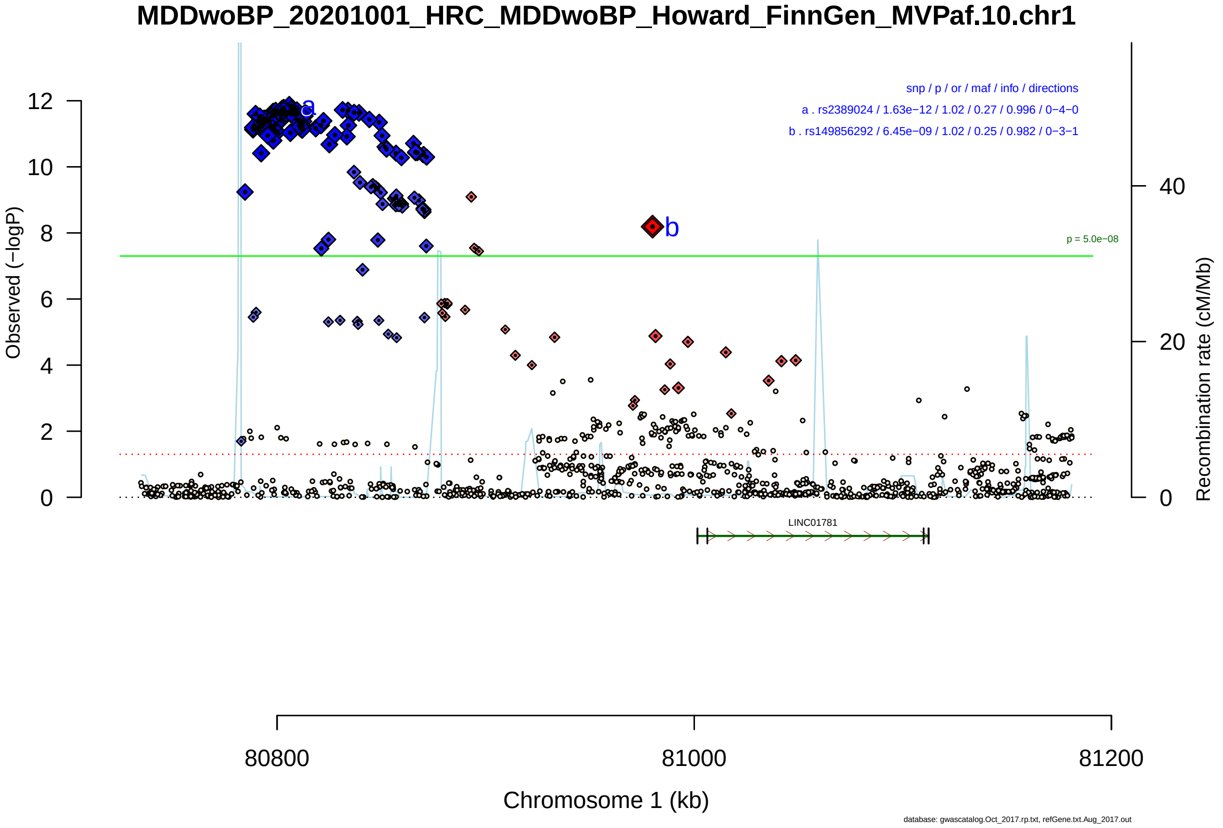

Figure S2-50: Regional plot of GWAS locus No. 50 , with rs2389024 as lead SNP.

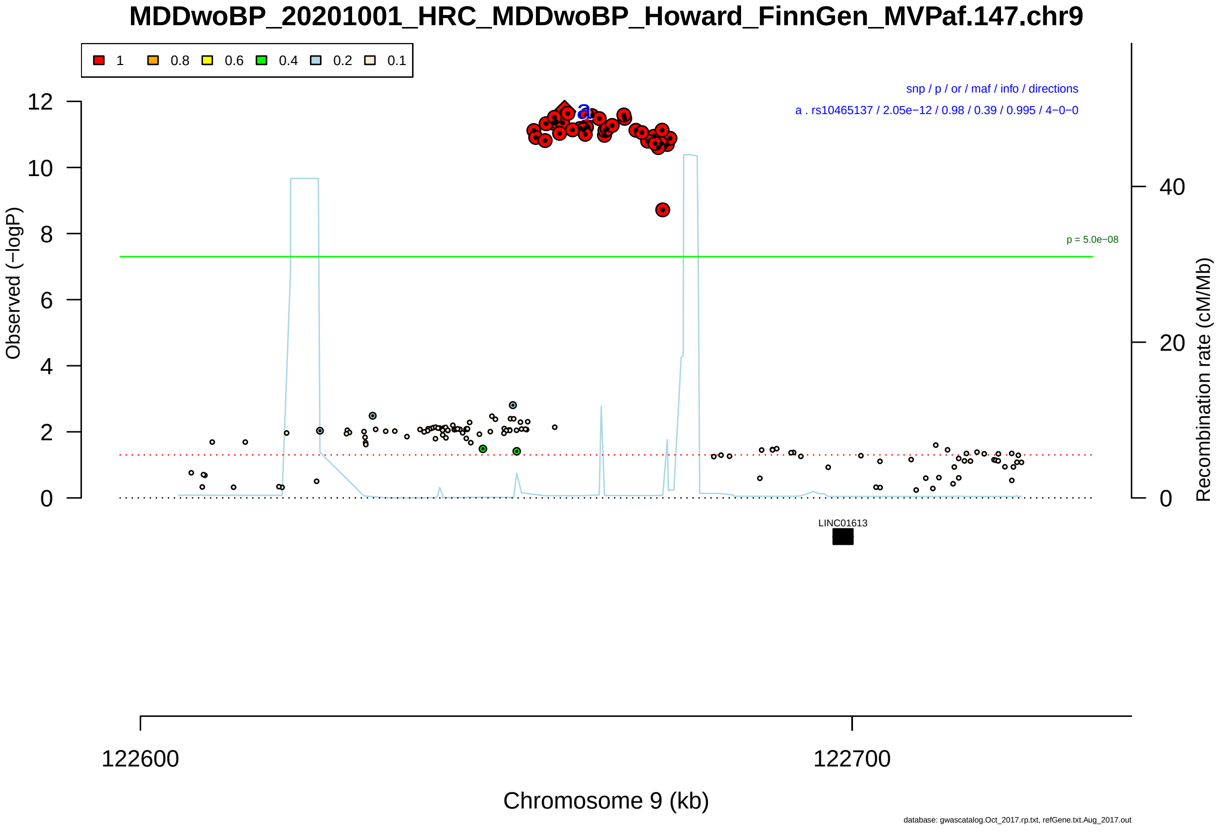

Figure S2-51: Regional plot of GWAS locus No. 51 , with rs10465137 as lead SNP.

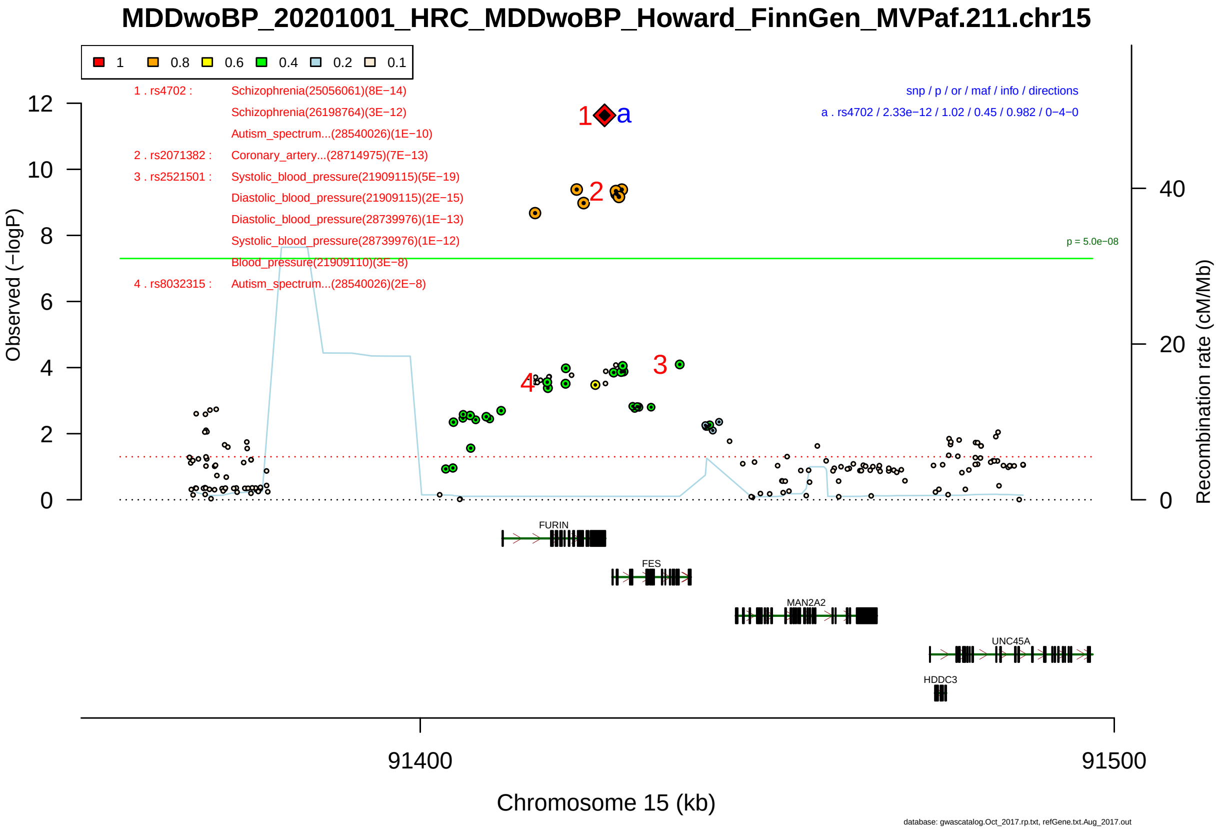

Figure S2-52: Regional plot of GWAS locus No. 52 , with rs4702 as lead SNP.

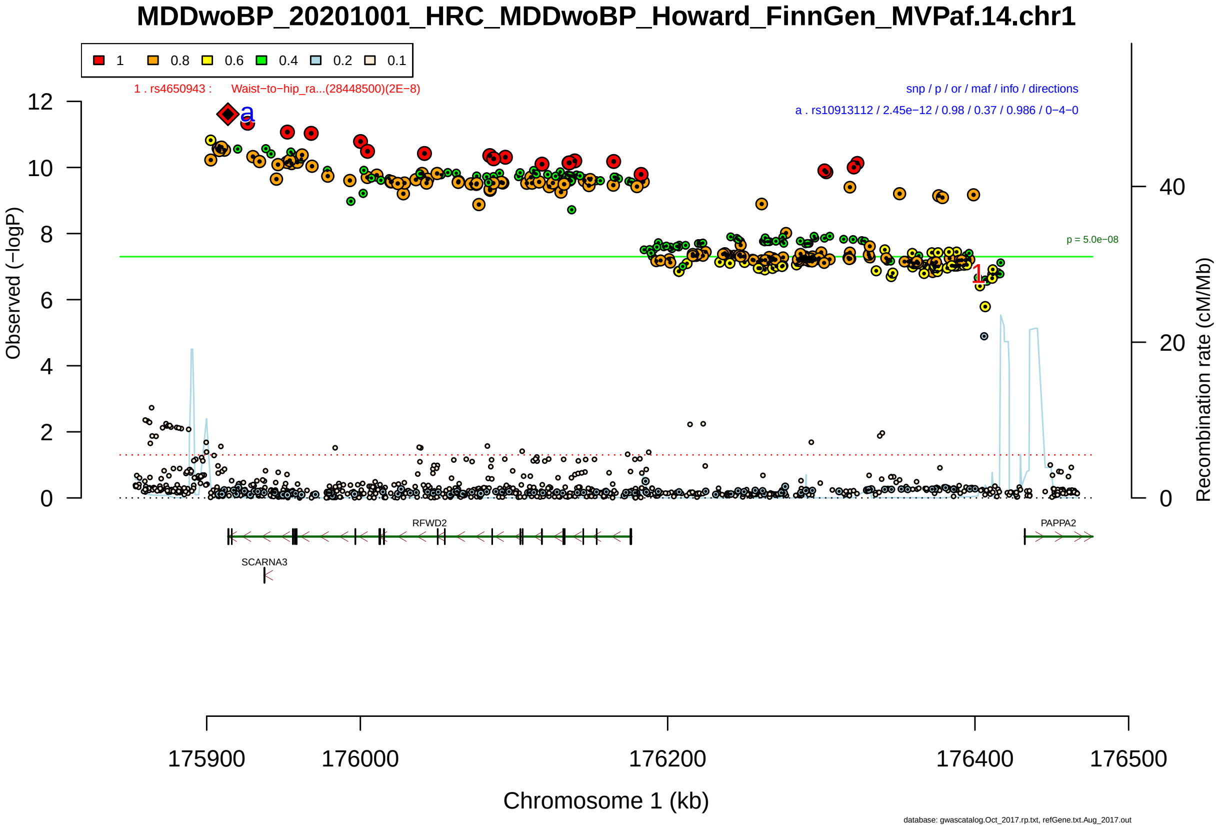

Figure S2-53: Regional plot of GWAS locus No. 53 , with rs10913112 as lead SNP.

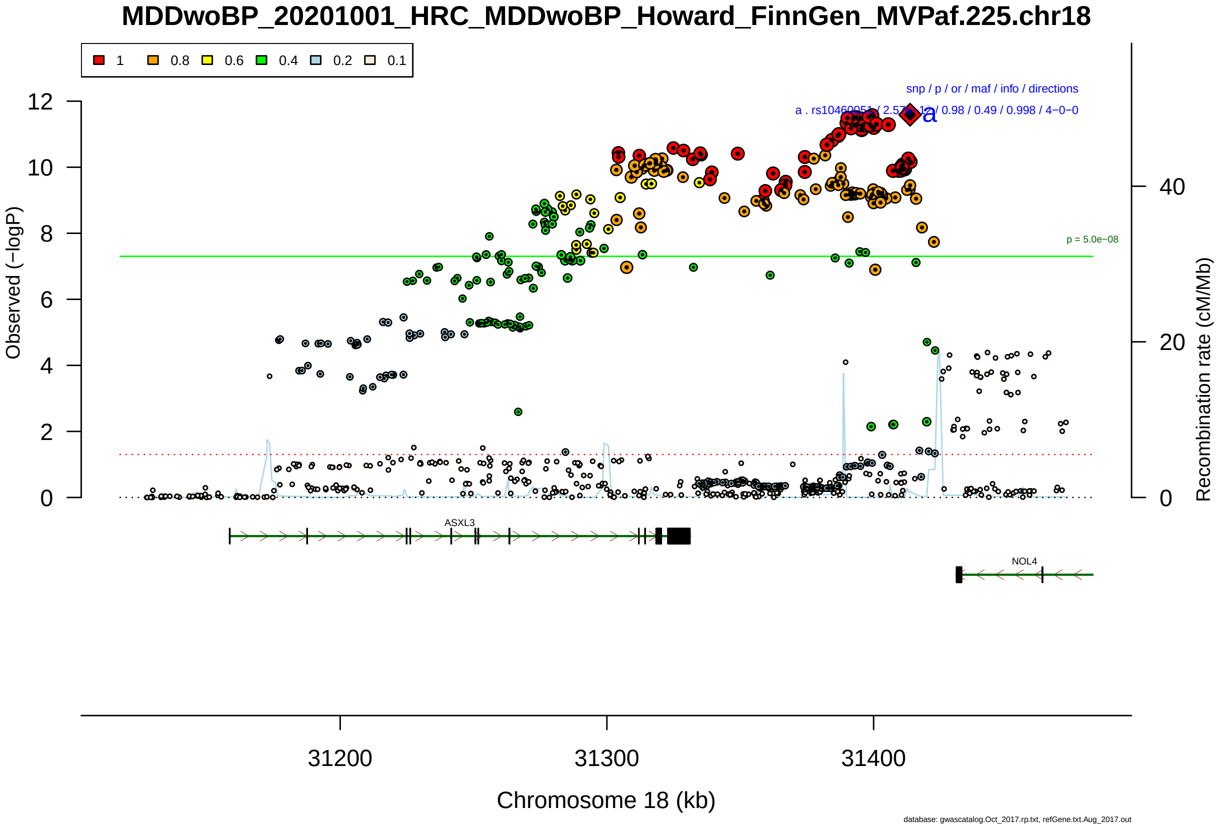

Figure S2-54: Regional plot of GWAS locus No. 54 , with rs10460051 as lead SNP.

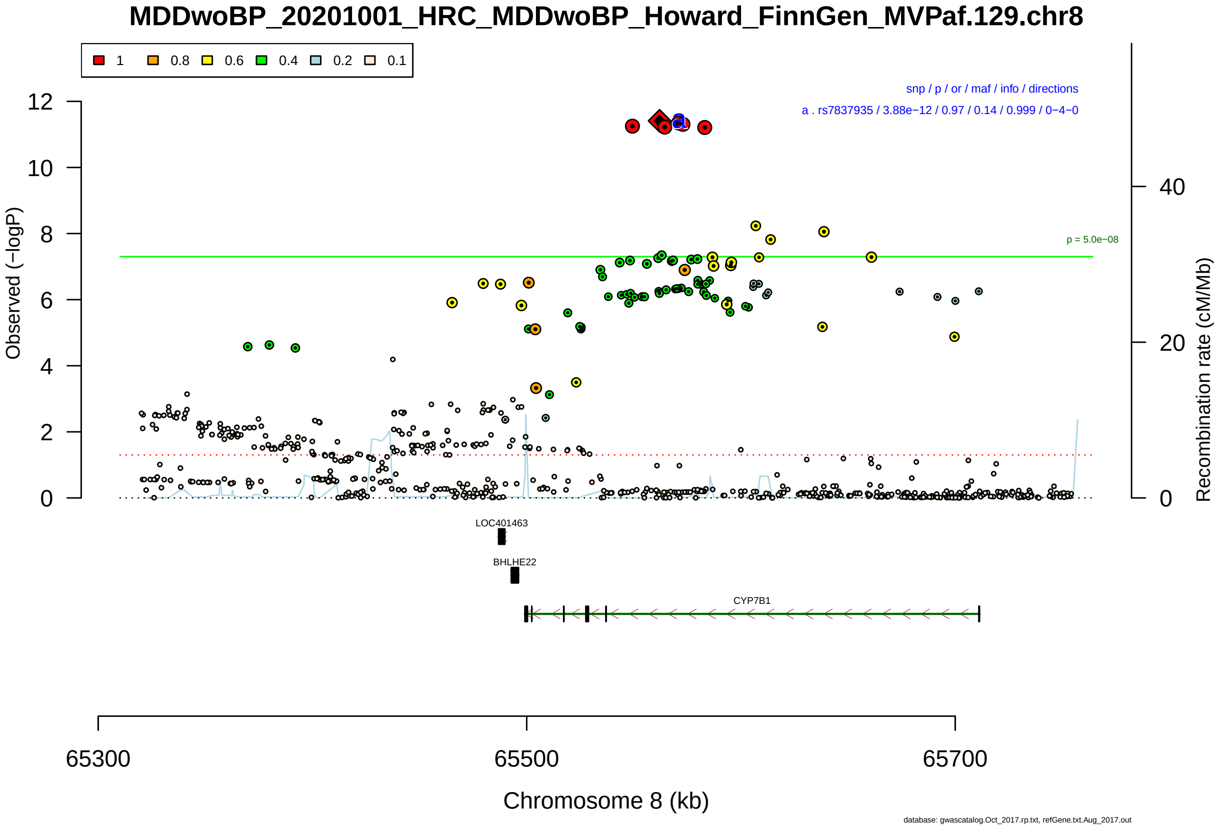

Figure S2-55: Regional plot of GWAS locus No. 55 , with rs7837935 as lead SNP.

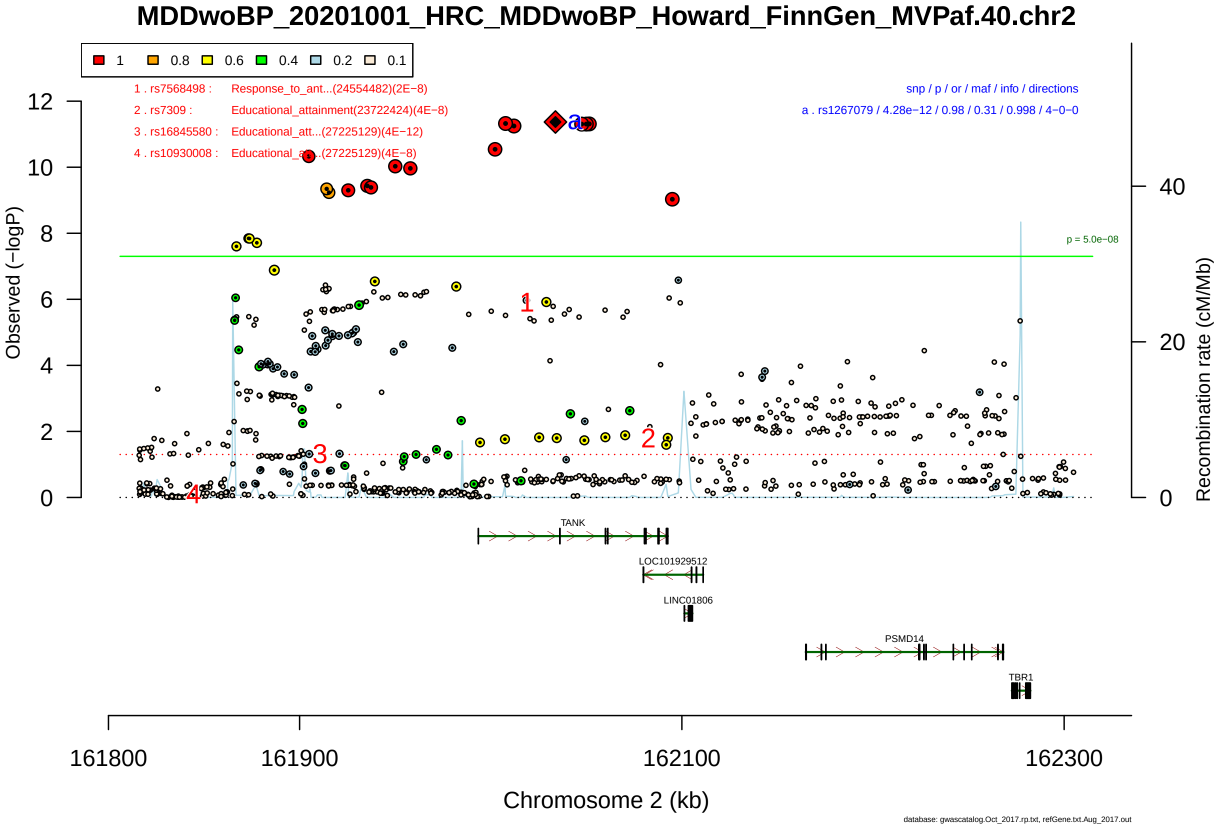

Figure S2-56: Regional plot of GWAS locus No. 56 , with rs1267079 as lead SNP.

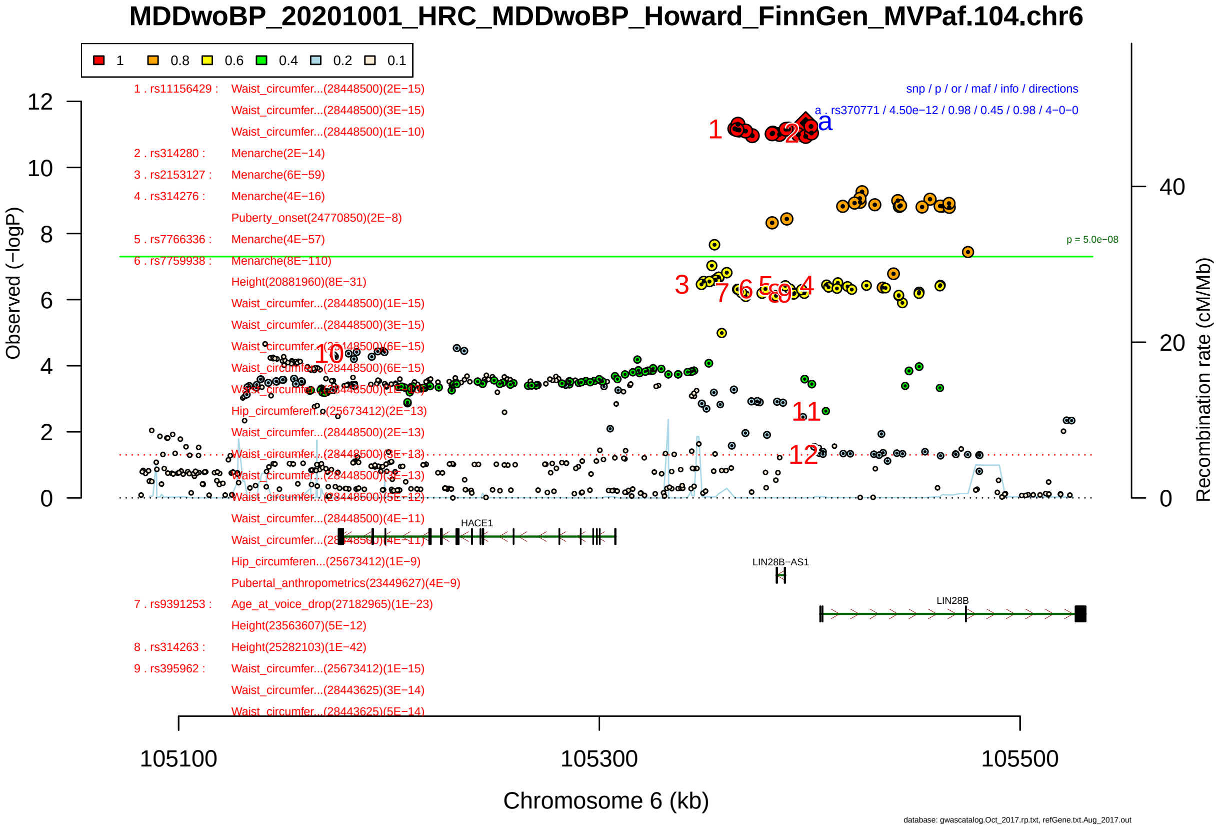

Figure S2-57: Regional plot of GWAS locus No. 57 , with rs370771 as lead SNP.

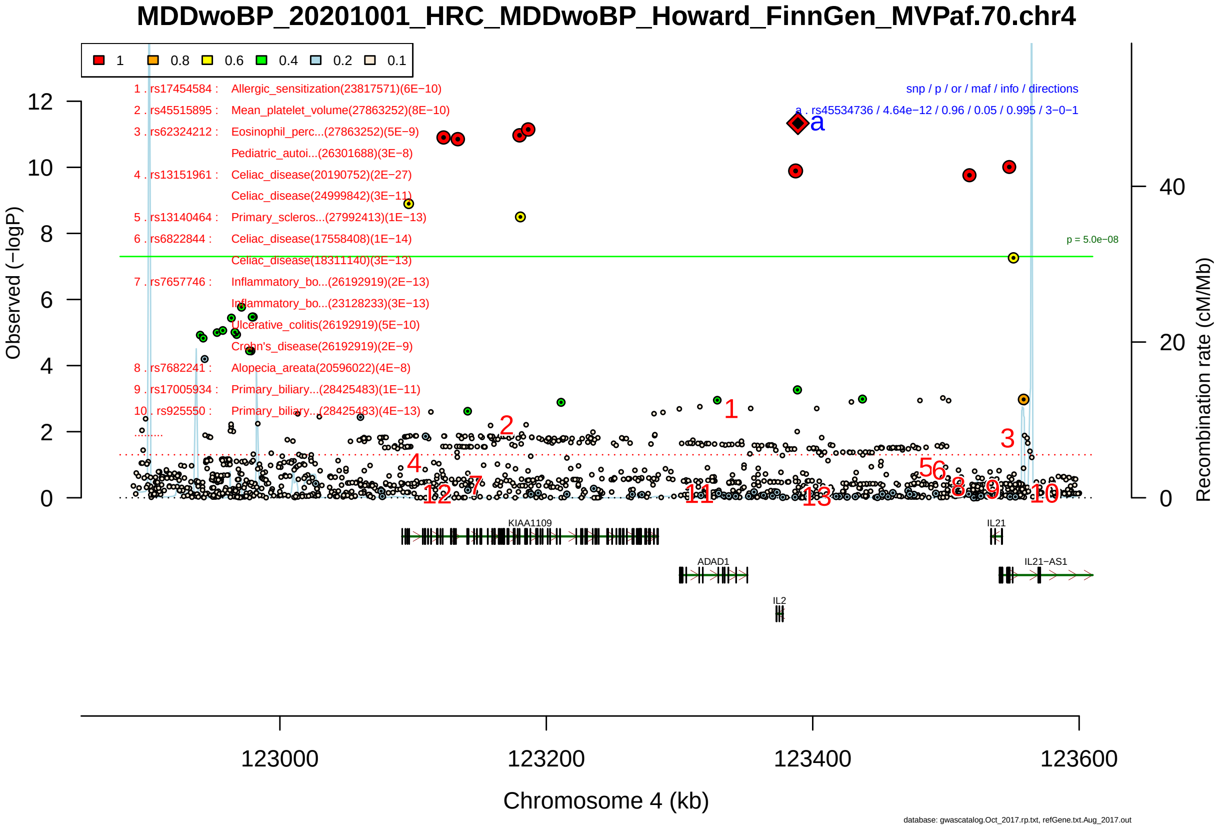

Figure S2-58: Regional plot of GWAS locus No. 58 , with rs45534736 as lead SNP.

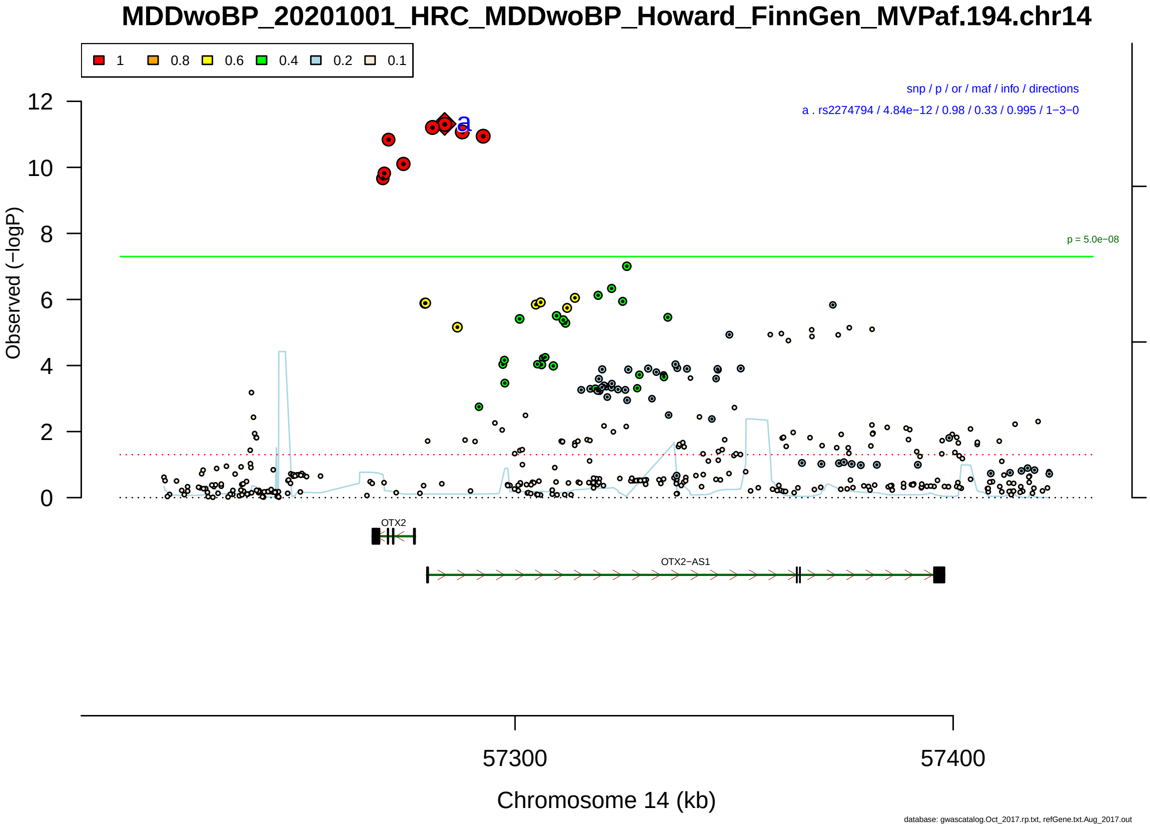

Figure S2-59: Regional plot of GWAS locus No. 59 , with rs2274794 as lead SNP.

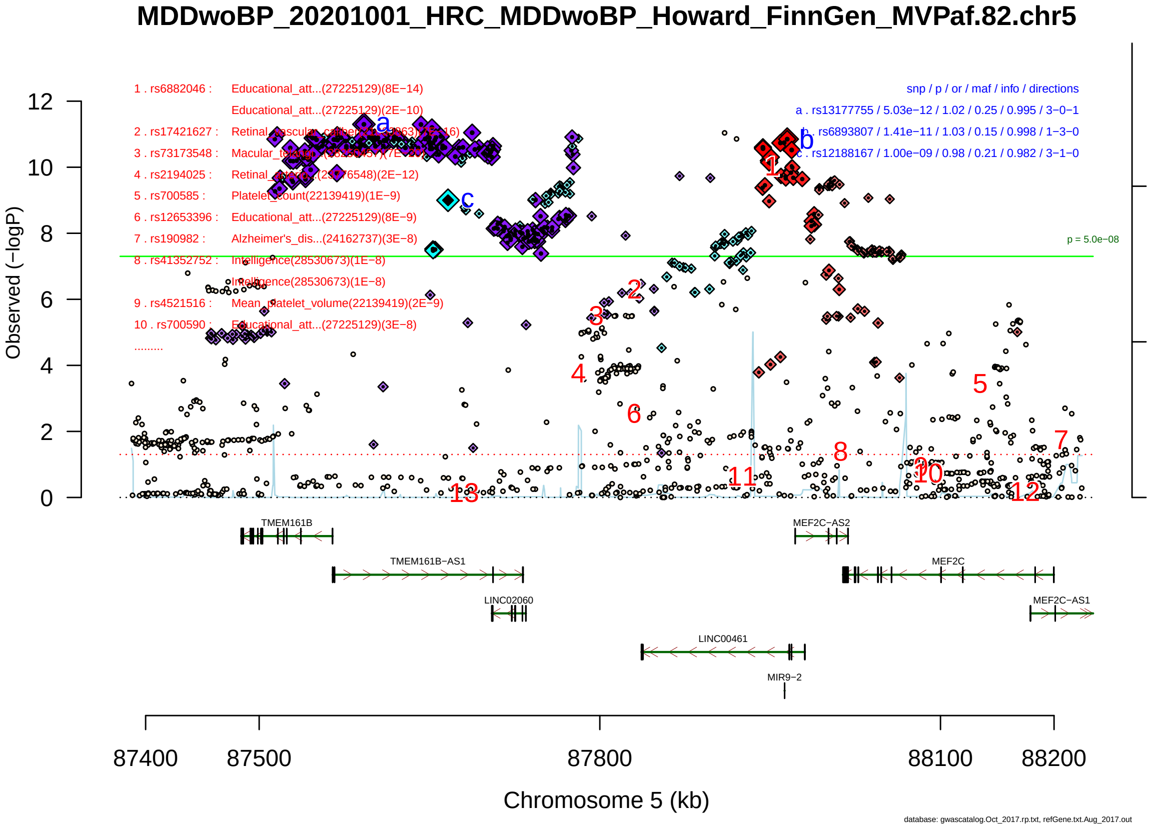

Figure S2-60: Regional plot of GWAS locus No. 60 , with rs13177755 as lead SNP.

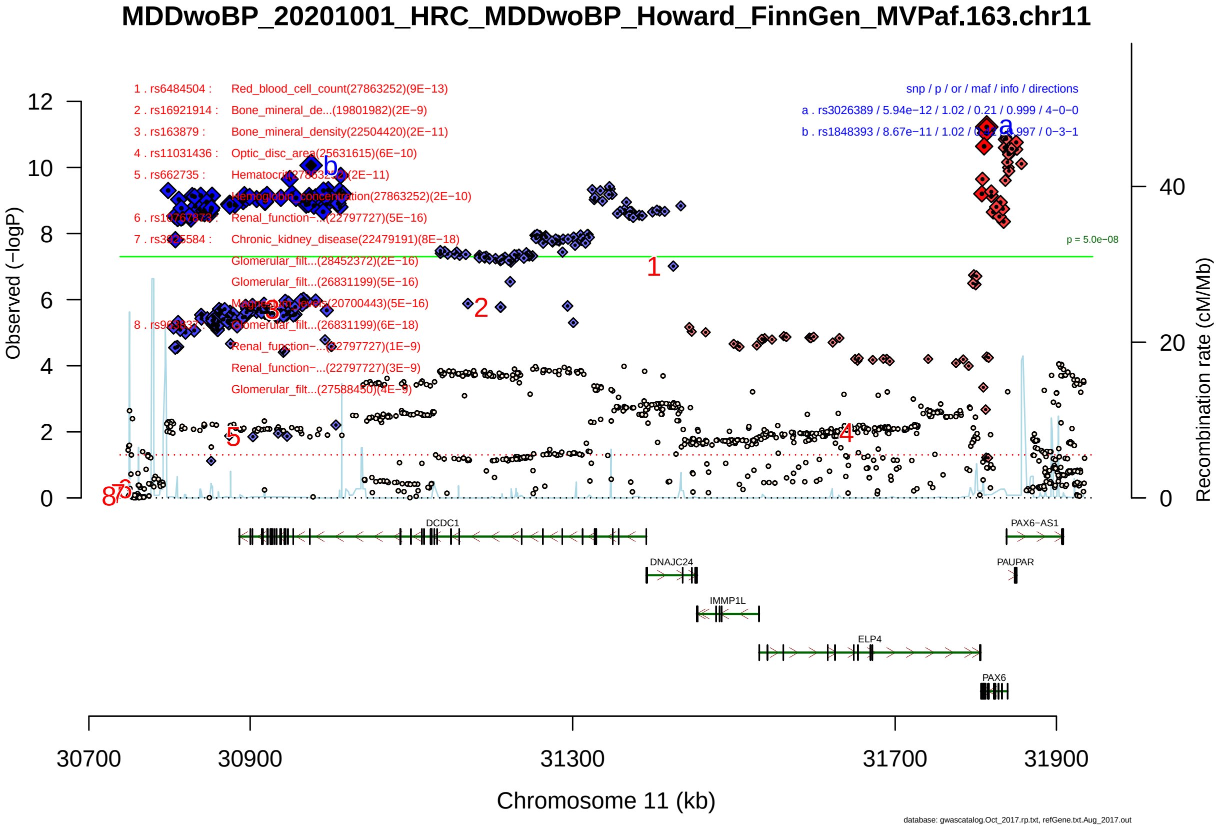

Figure S2-61: Regional plot of GWAS locus No. 61 , with rs3026389 as lead SNP.

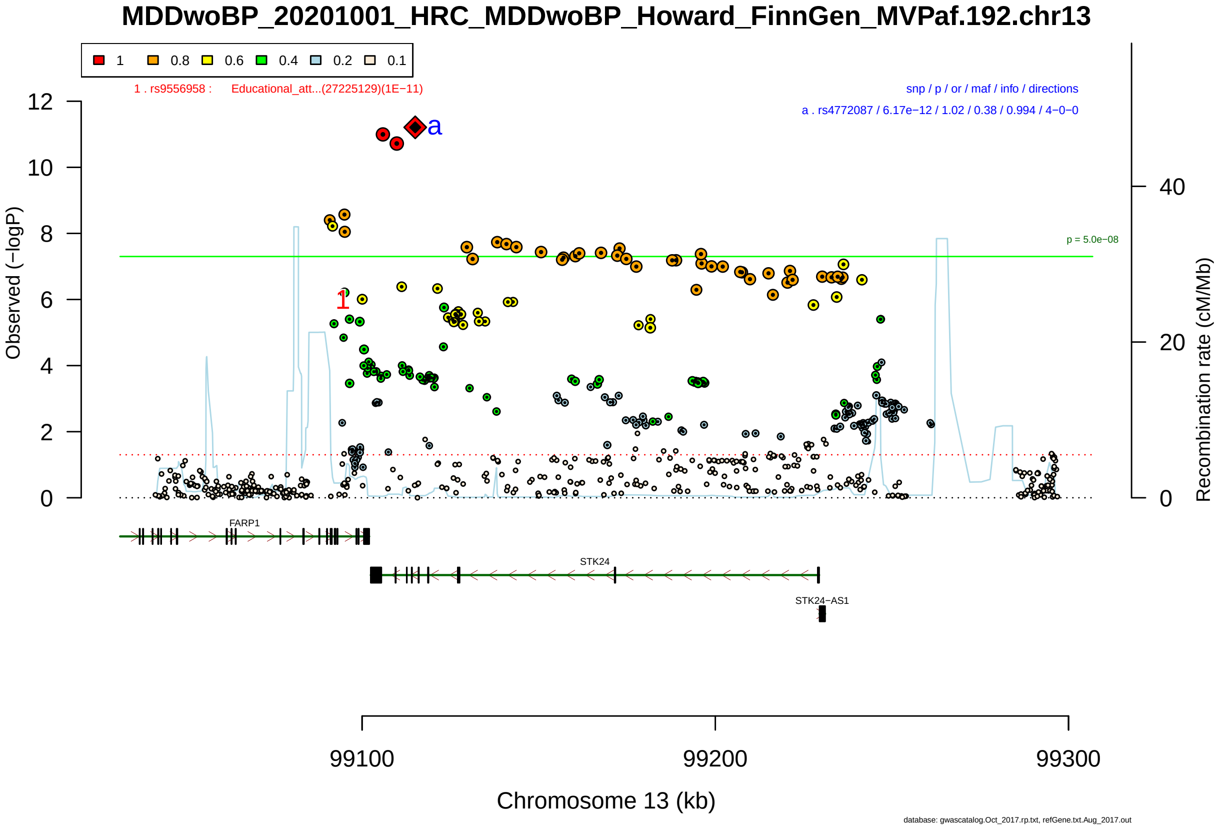

Figure S2-62: Regional plot of GWAS locus No. 62 , with rs4772087 as lead SNP.

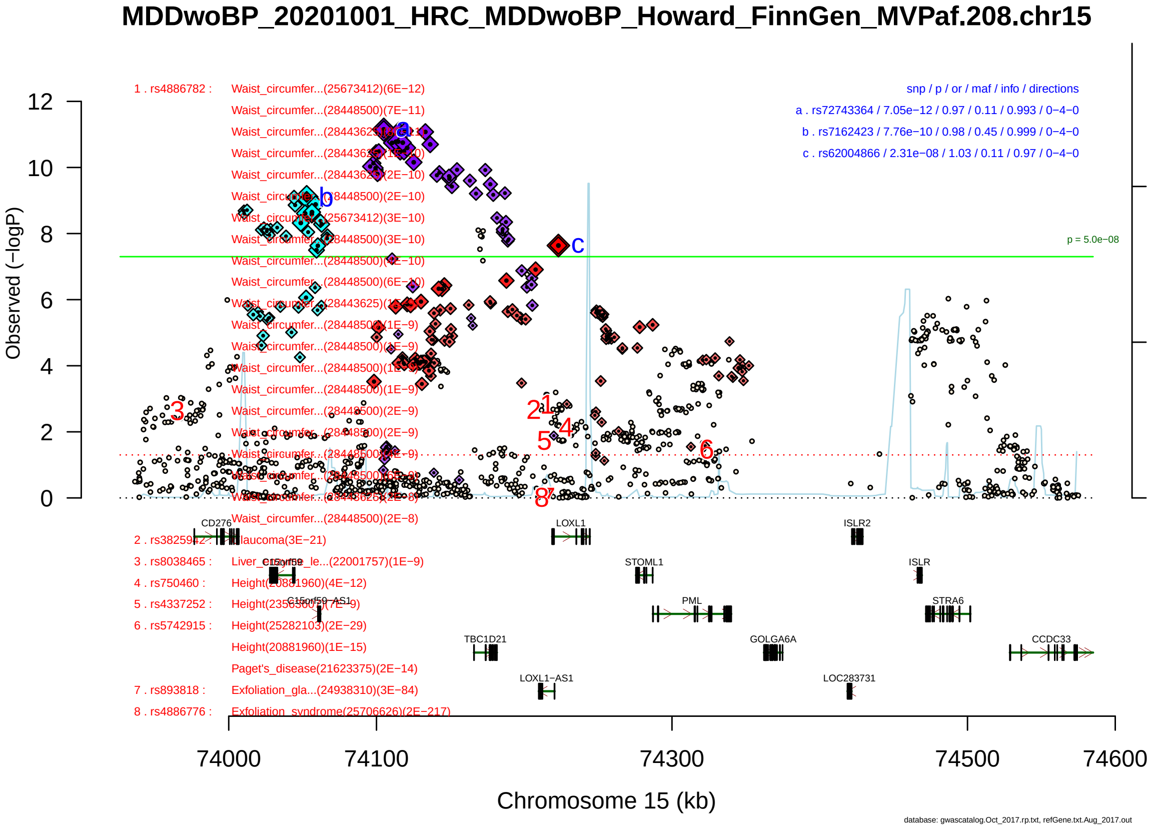

Figure S2-63: Regional plot of GWAS locus No. 63 , with rs72743364 as lead SNP.

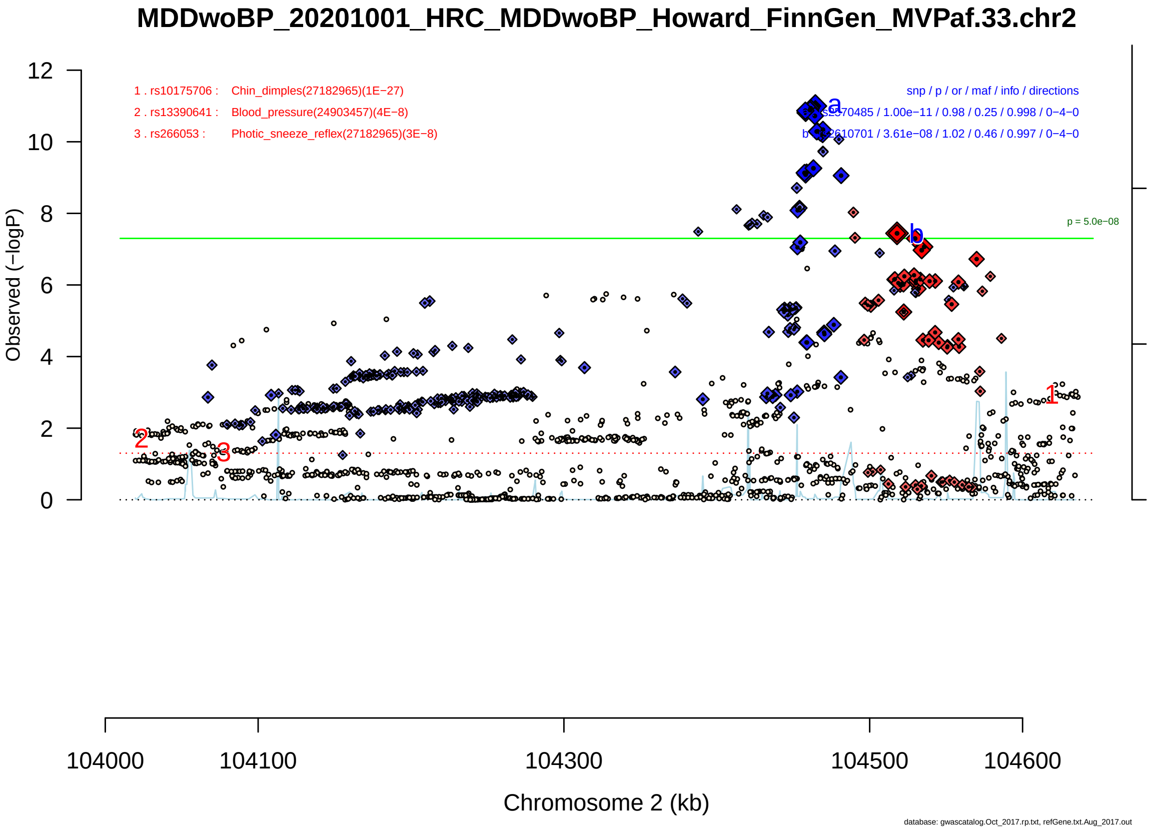

Figure S2-64: Regional plot of GWAS locus No. 64 , with rs2570485 as lead SNP.

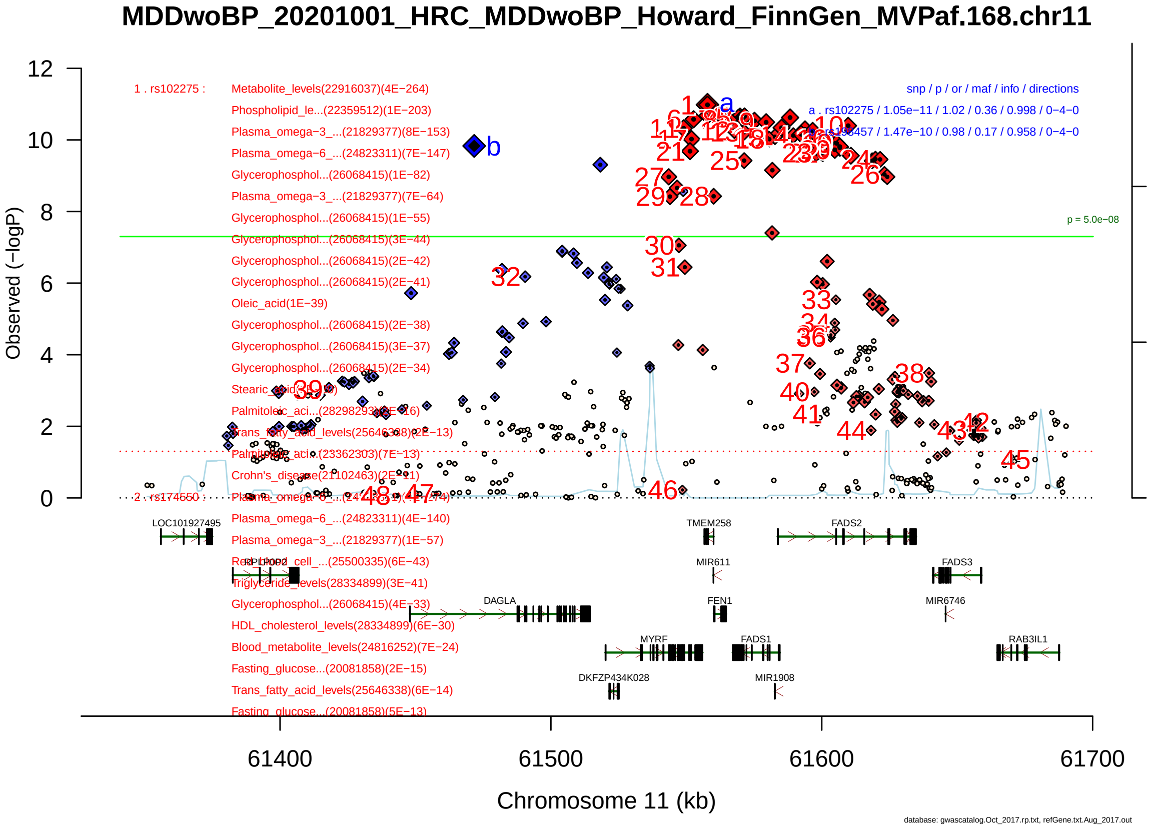

Figure S2-65: Regional plot of GWAS locus No. 65 , with rs102275 as lead SNP.

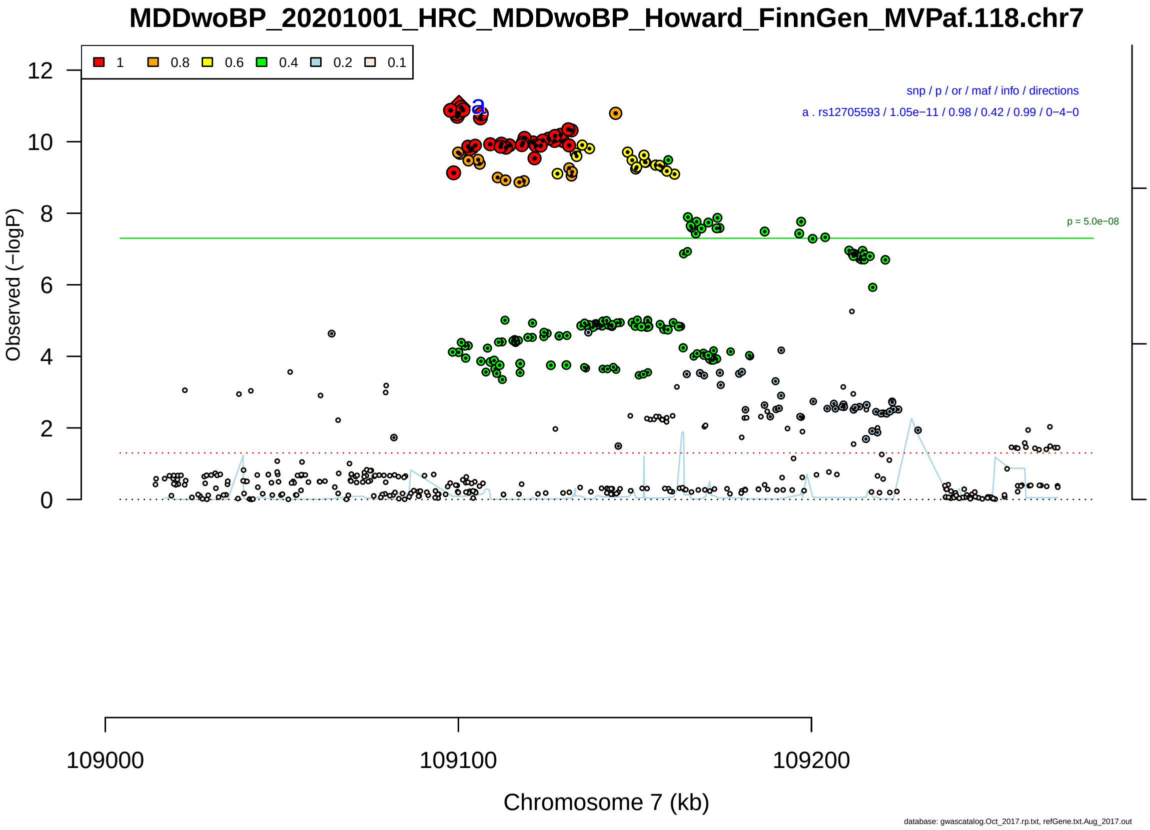

Figure S2-66: Regional plot of GWAS locus No. 66 , with rs12705593 as lead SNP.

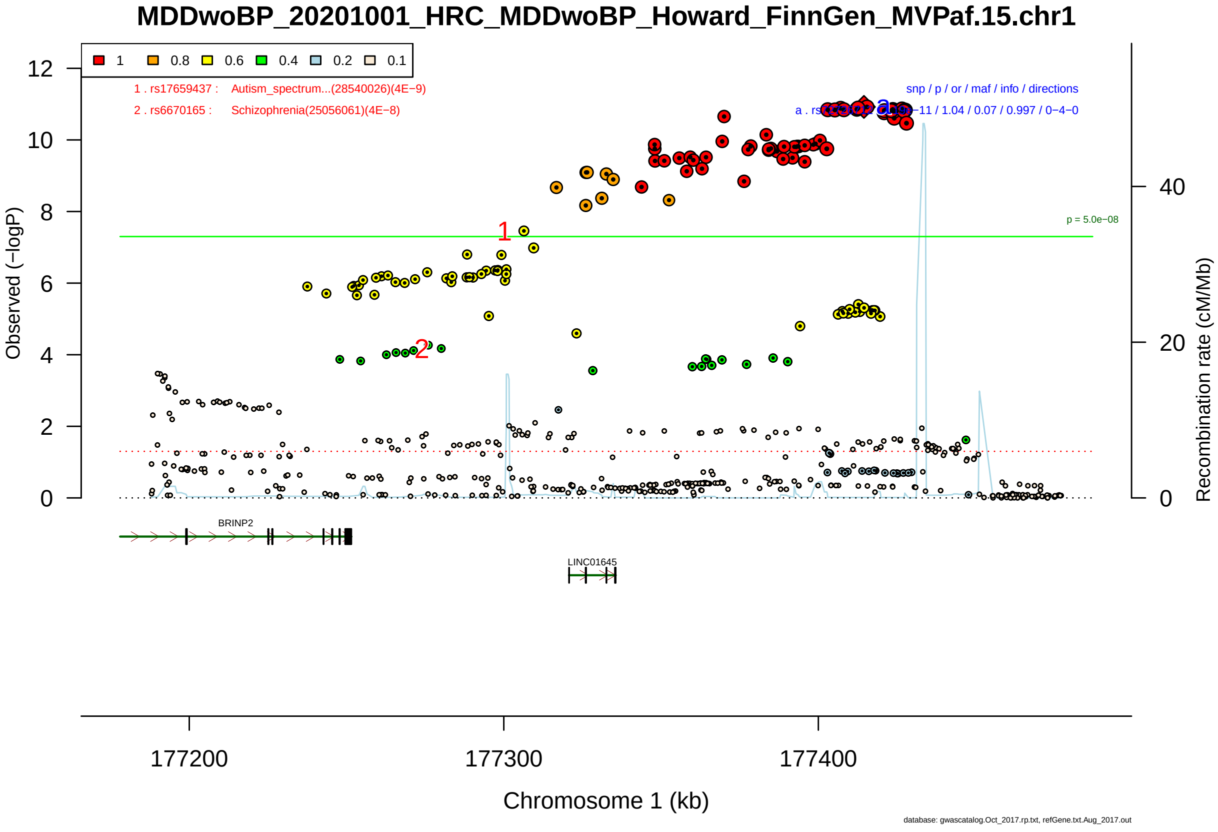

Figure S2-67: Regional plot of GWAS locus No. 67 , with rs72726712 as lead SNP.

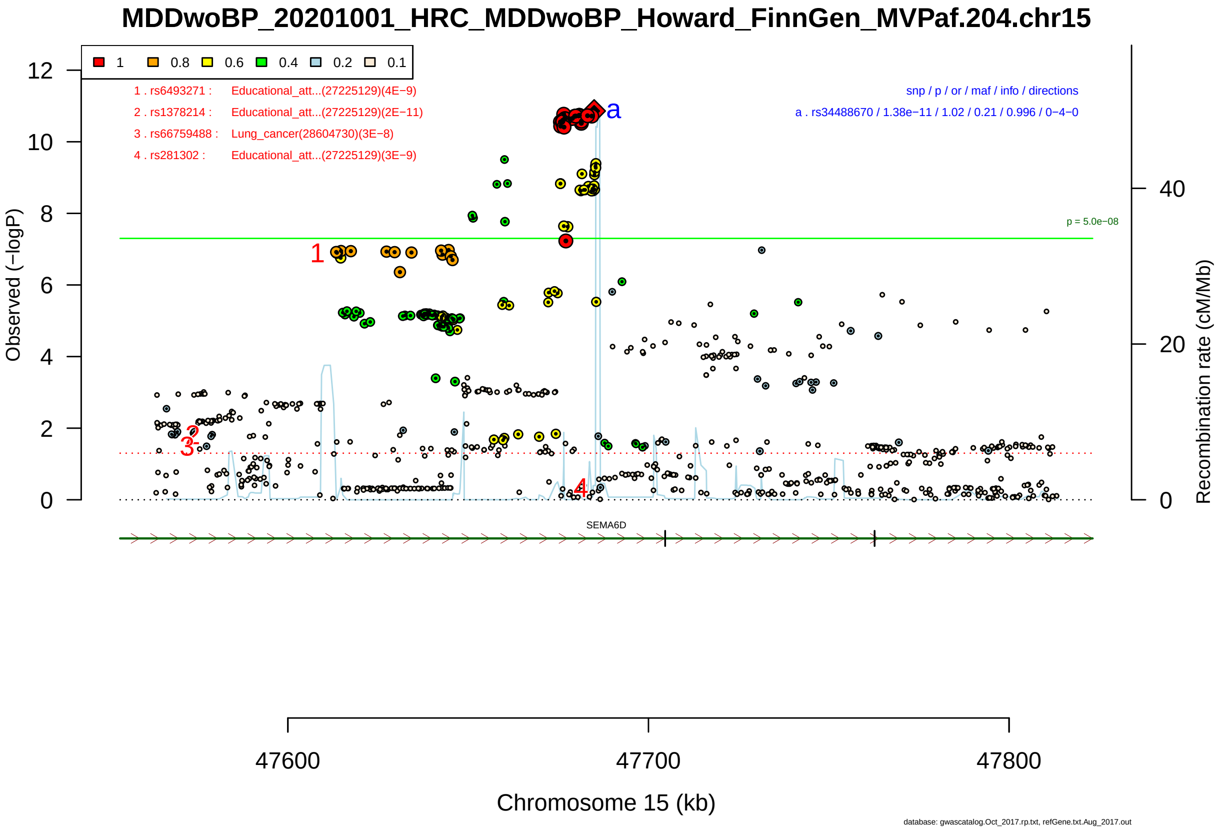

Figure S2-68: Regional plot of GWAS locus No. 68 , with rs34488670 as lead SNP.

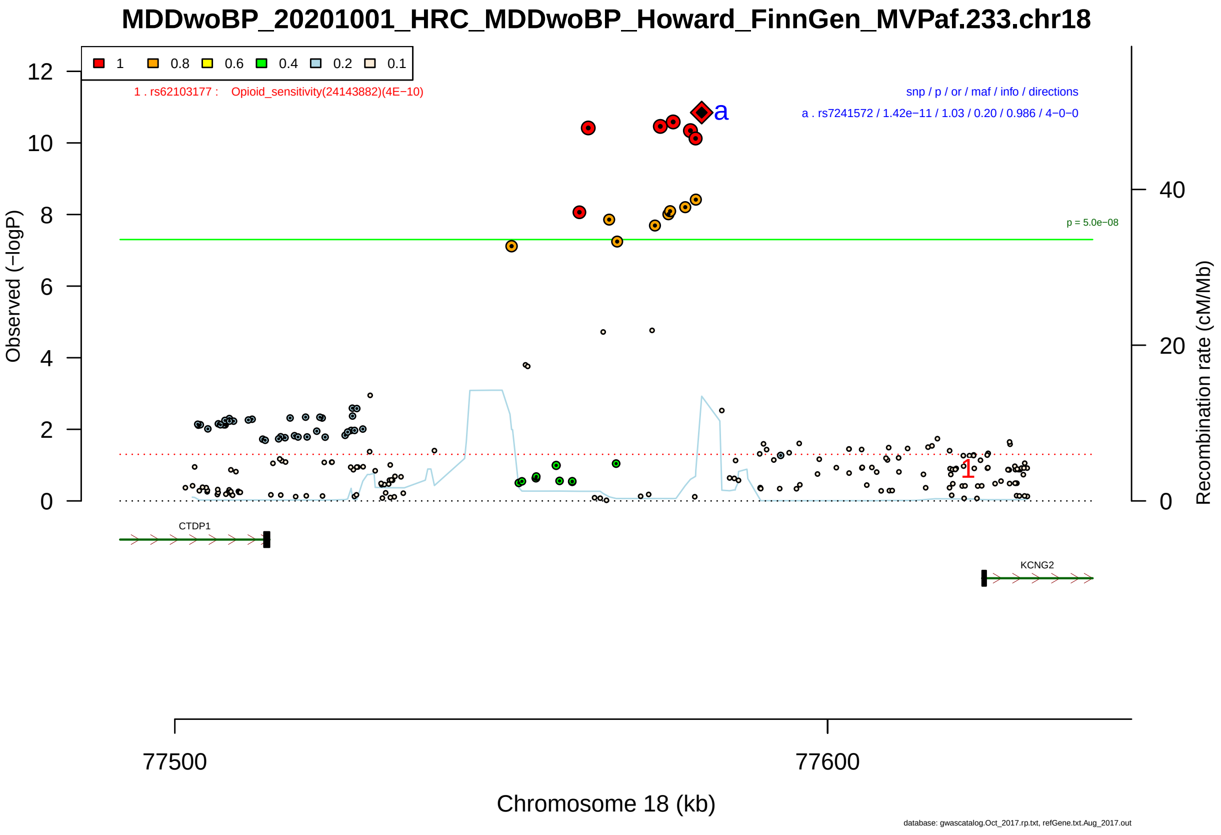

Figure S2-69: Regional plot of GWAS locus No. 69 , with rs7241572 as lead SNP.

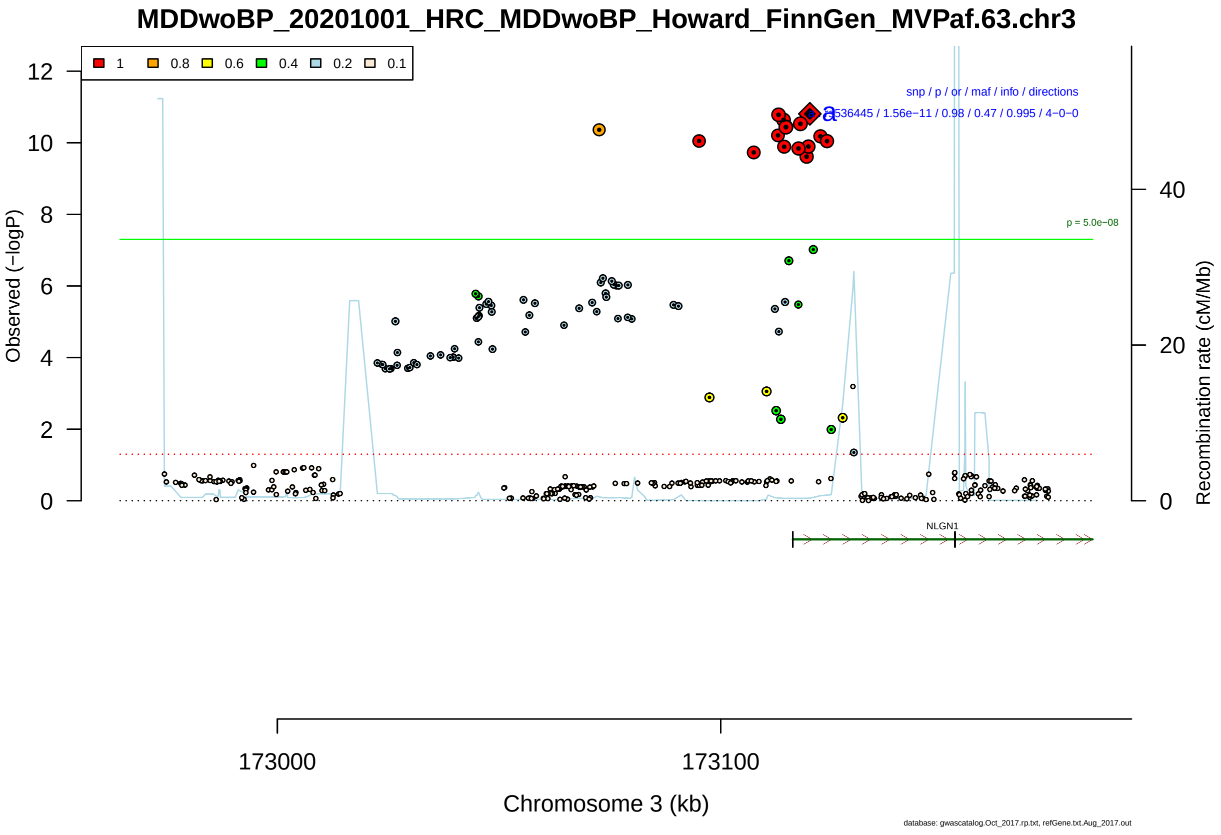

Figure S2-70: Regional plot of GWAS locus No. 70 , with rs536445 as lead SNP.

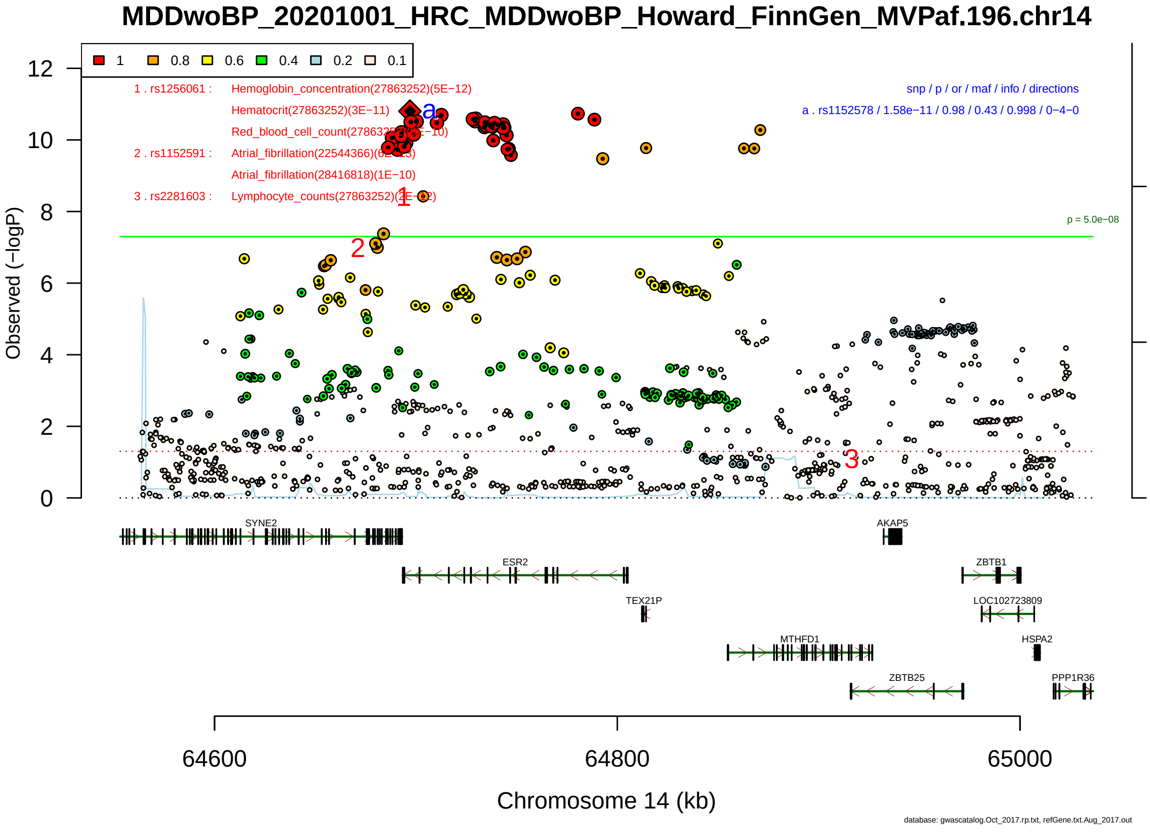

Figure S2-71: Regional plot of GWAS locus No. 71 , with rs1152578 as lead SNP.

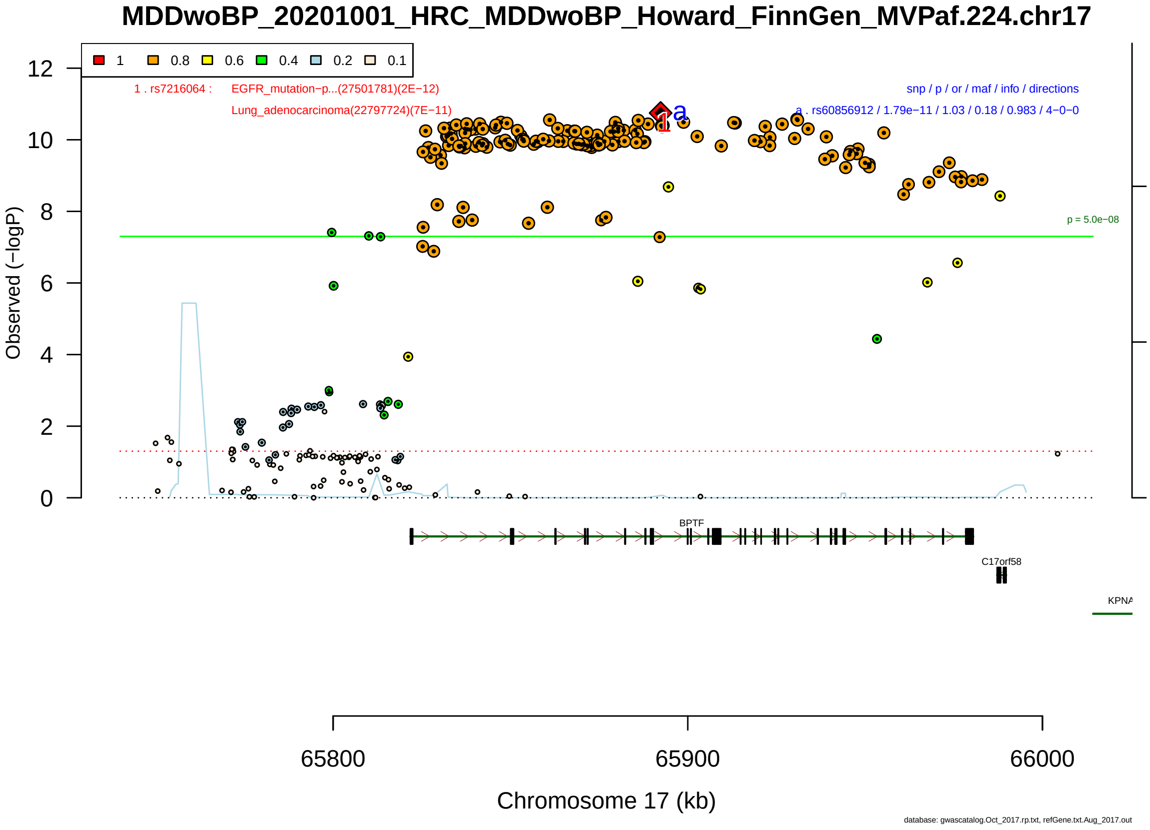

Figure S2-72: Regional plot of GWAS locus No. 72 , with rs60856912 as lead SNP.

**MDDwoBP_20201001_HRC_MDDwoBP_Howard_FinnGen_MVPaf.122.chr7**

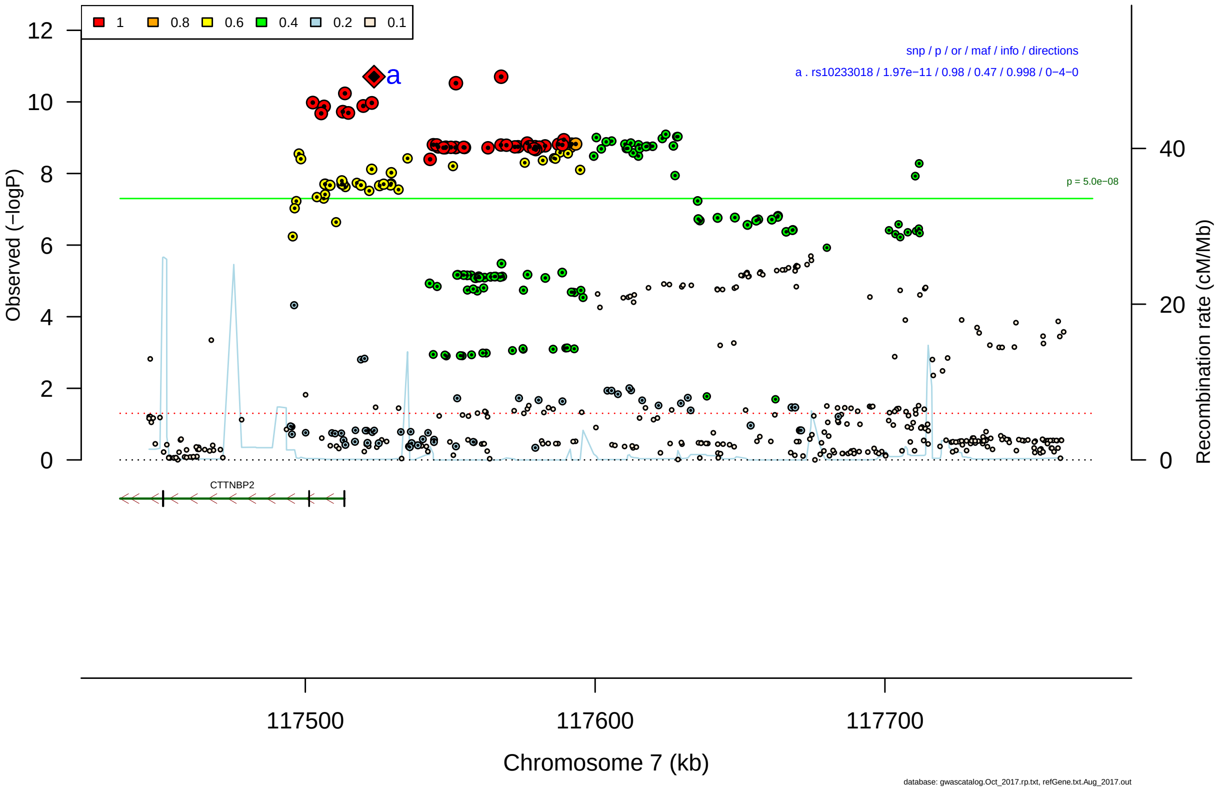

Figure S2-73: Regional plot of GWAS locus No. 73 , with rs10233018 as lead SNP.

**MDDwoBP_20201001_HRC_MDDwoBP_Howard_FinnGen_MVPaf.89.chr5**

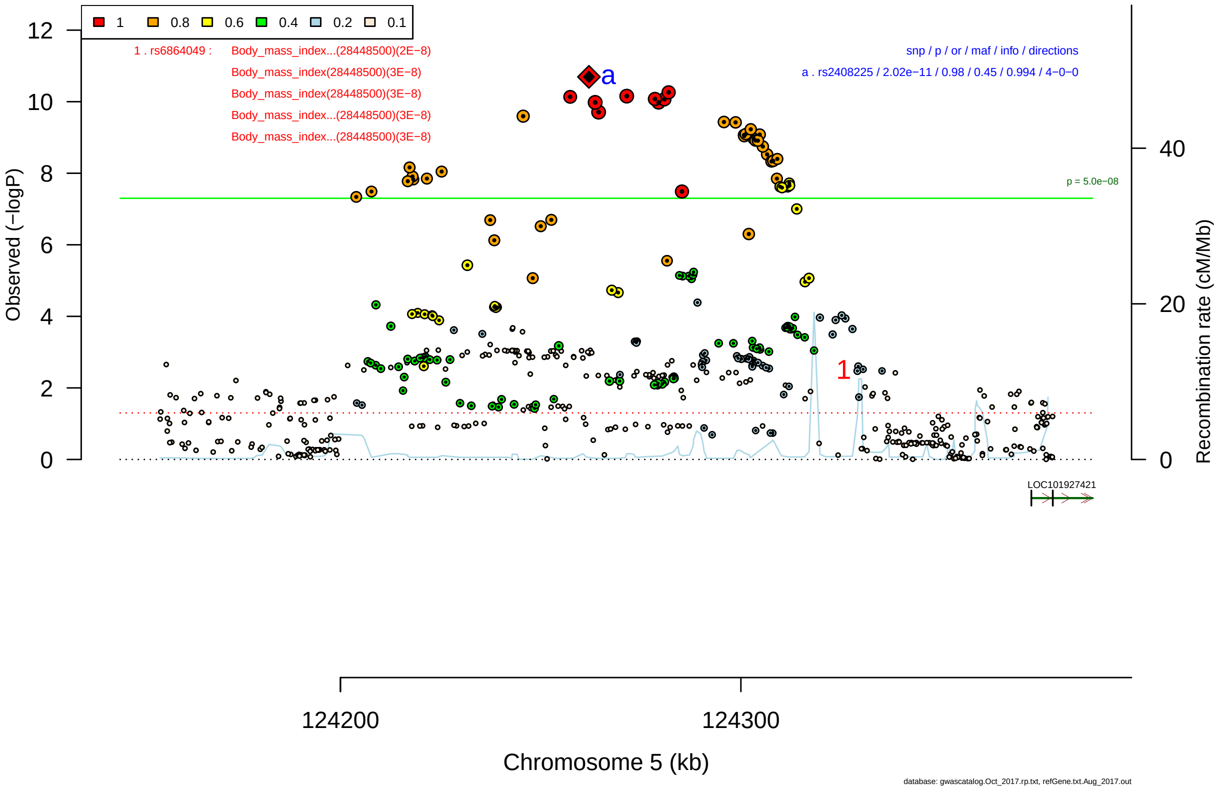

Figure S2-74: Regional plot of GWAS locus No. 74 , with rs2408225 as lead SNP.

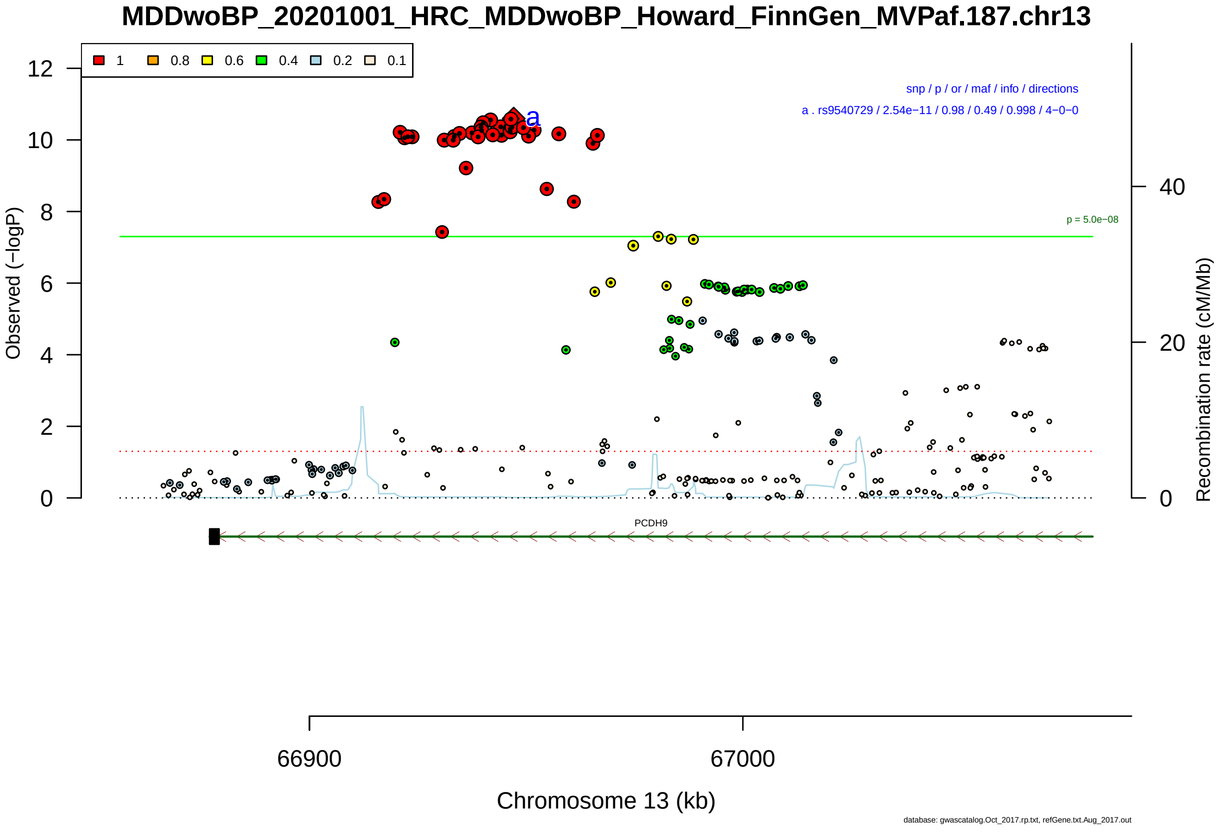

Figure S2-75: Regional plot of GWAS locus No. 75 , with rs9540729 as lead SNP.

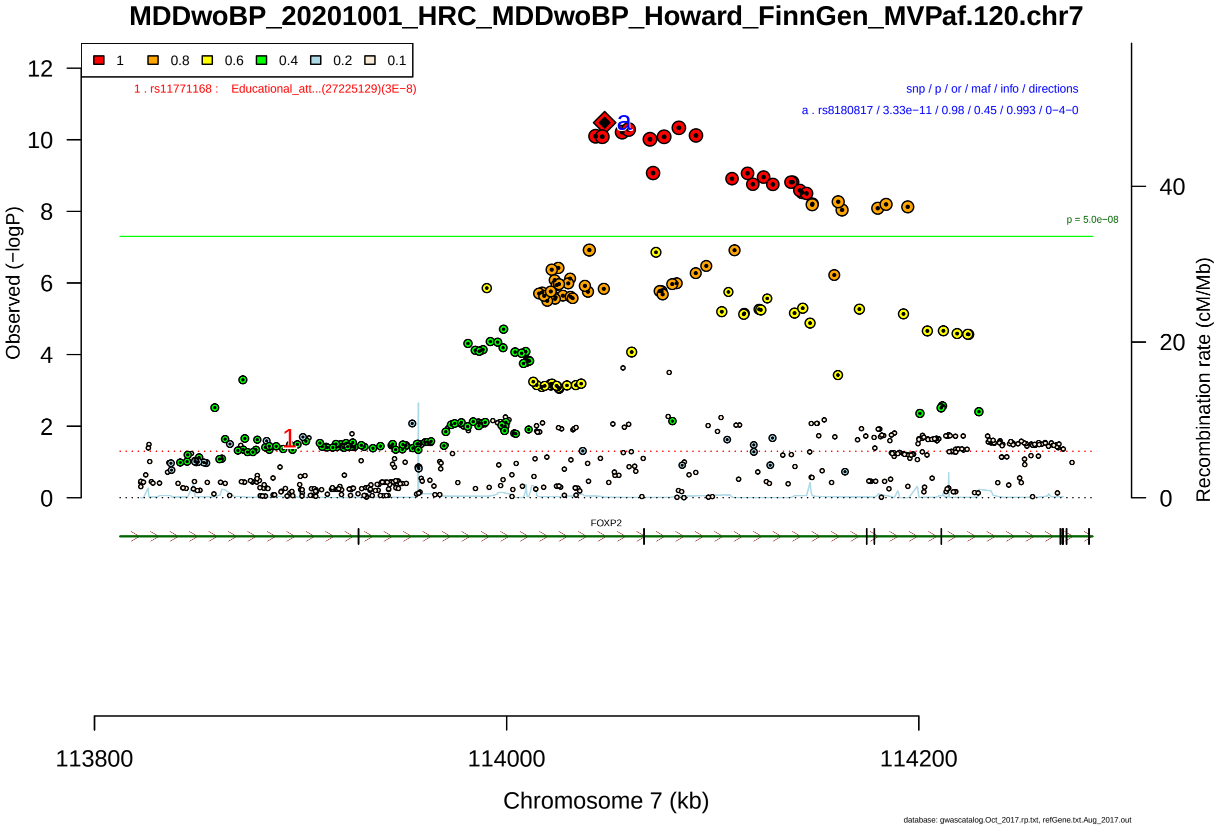

Figure S2-76: Regional plot of GWAS locus No. 76 , with rs8180817 as lead SNP.

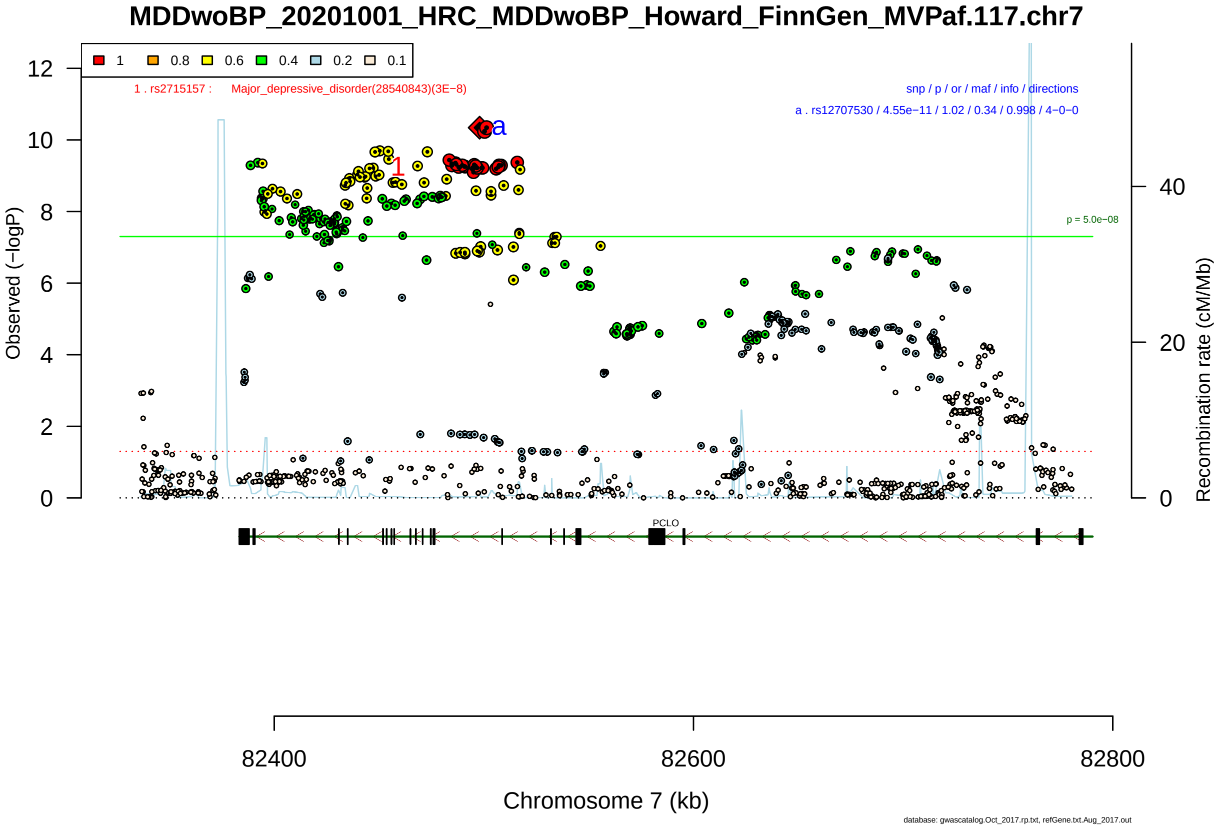

Figure S2-77: Regional plot of GWAS locus No. 77 , with rs12707530 as lead SNP.

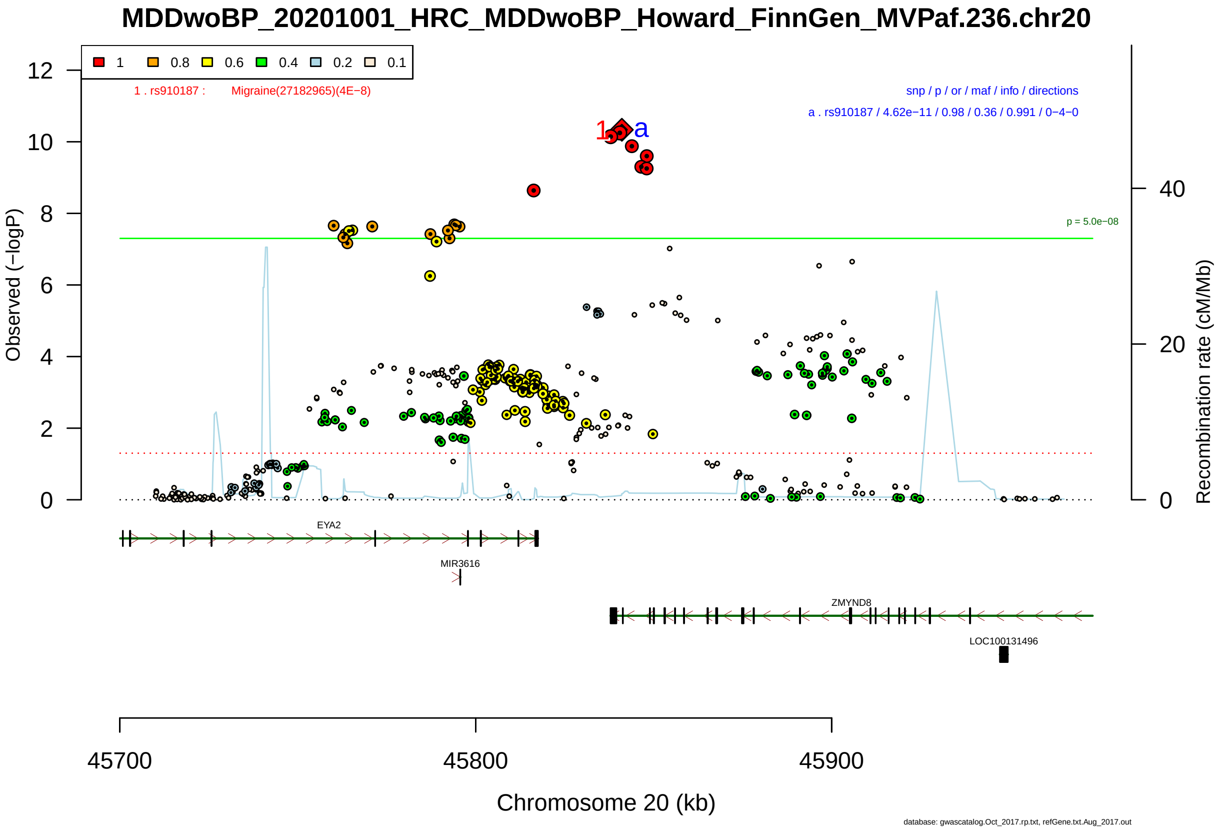

Figure S2-78: Regional plot of GWAS locus No. 78 , with rs910187 as lead SNP.

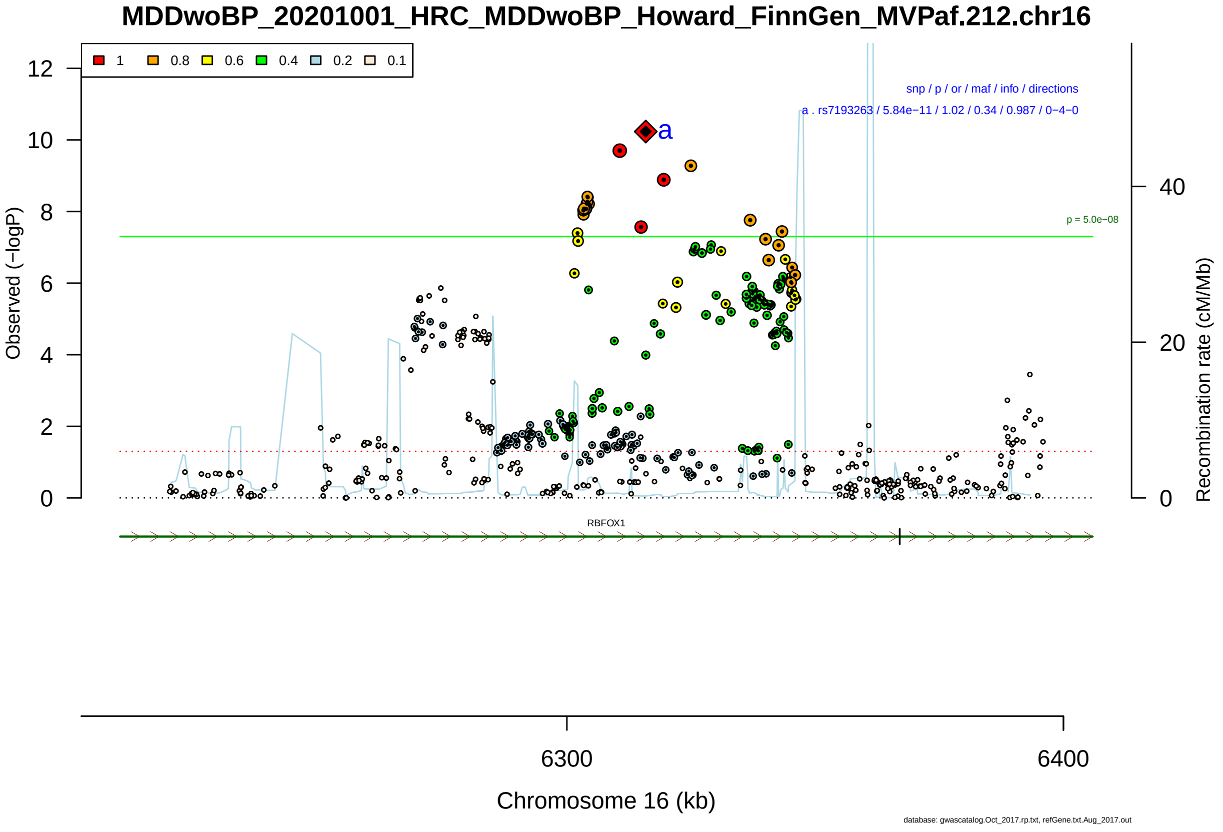

Figure S2-79: Regional plot of GWAS locus No. 79 , with rs7193263 as lead SNP.

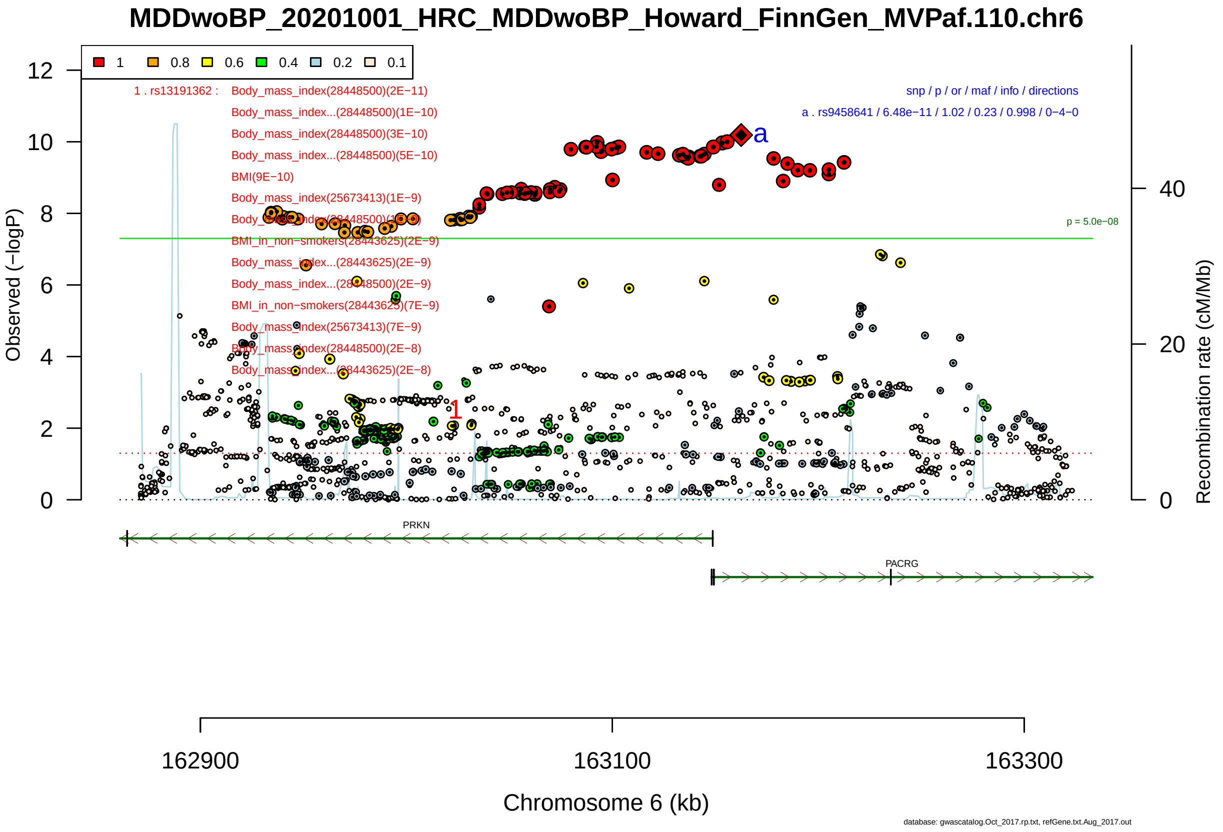

Figure S2-80: Regional plot of GWAS locus No. 80 , with rs9458641 as lead SNP.

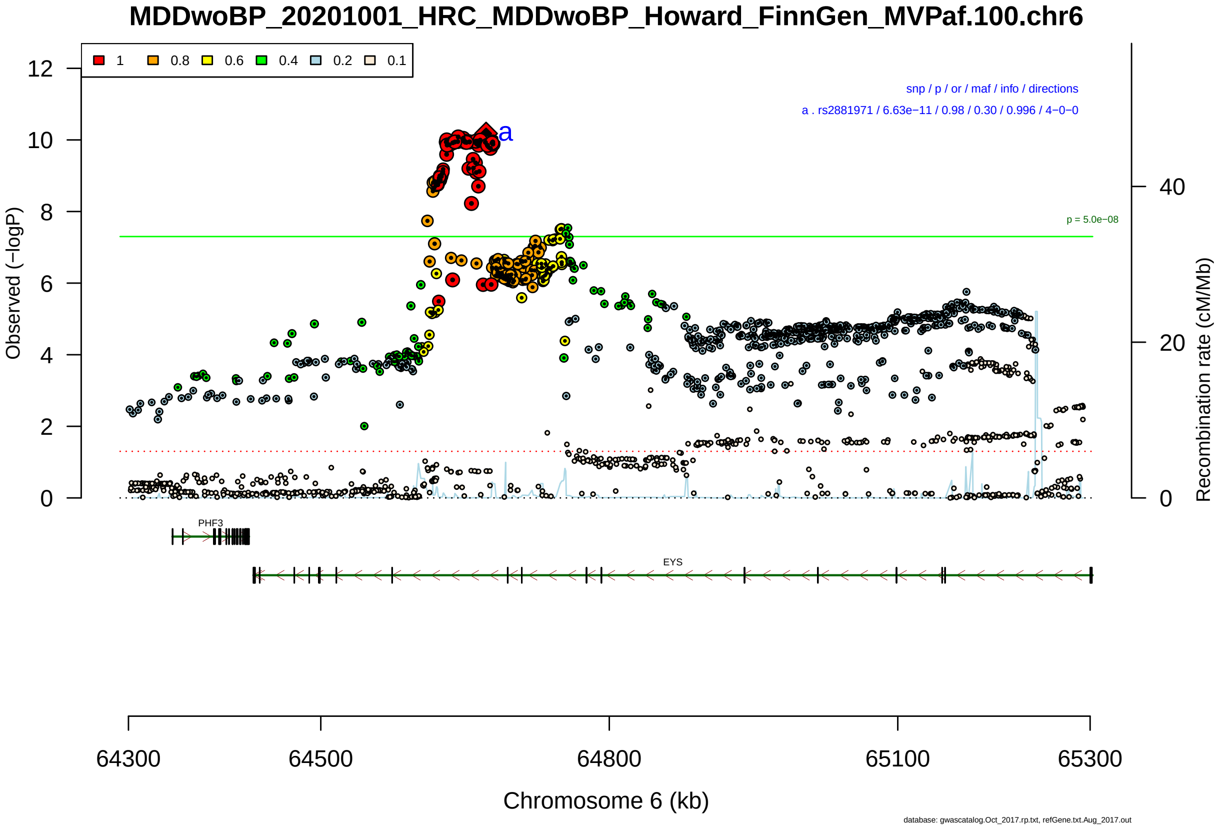

Figure S2-81: Regional plot of GWAS locus No. 81 , with rs2881971 as lead SNP.

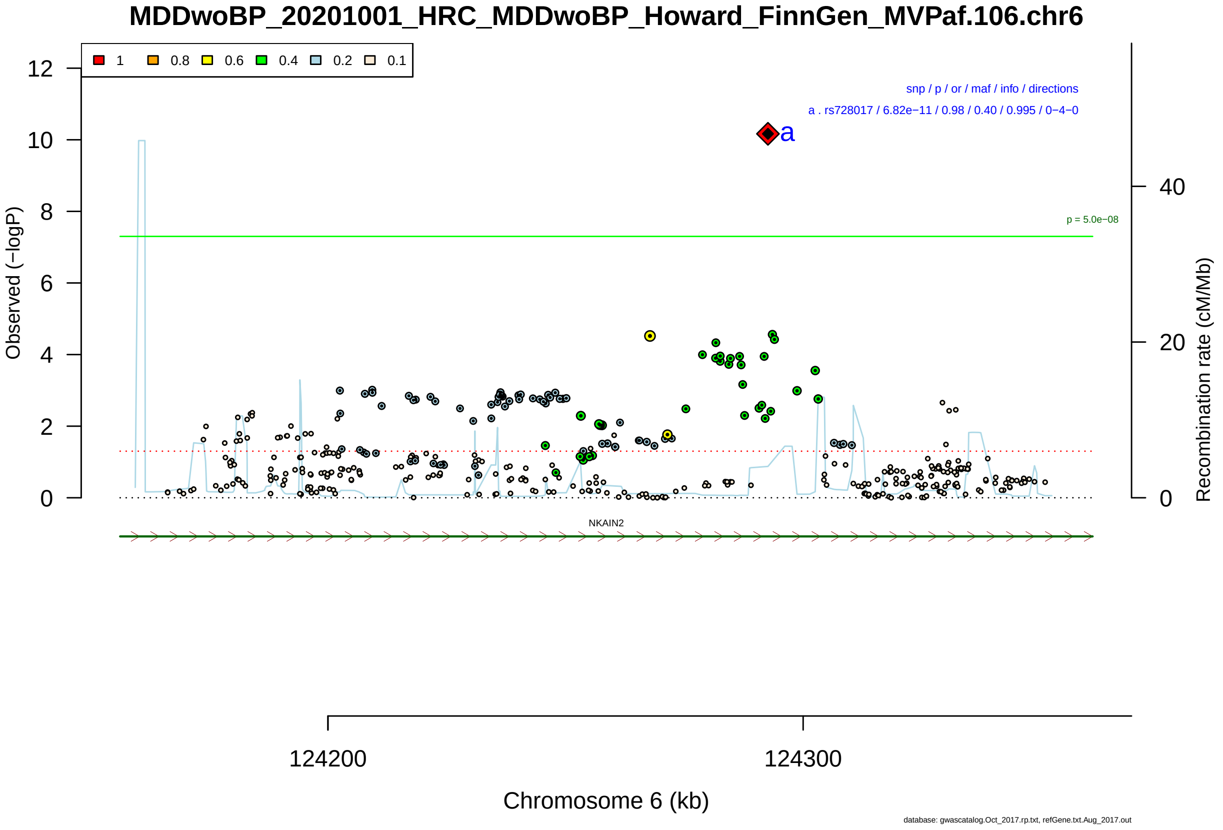

Figure S2-82: Regional plot of GWAS locus No. 82 , with rs728017 as lead SNP.

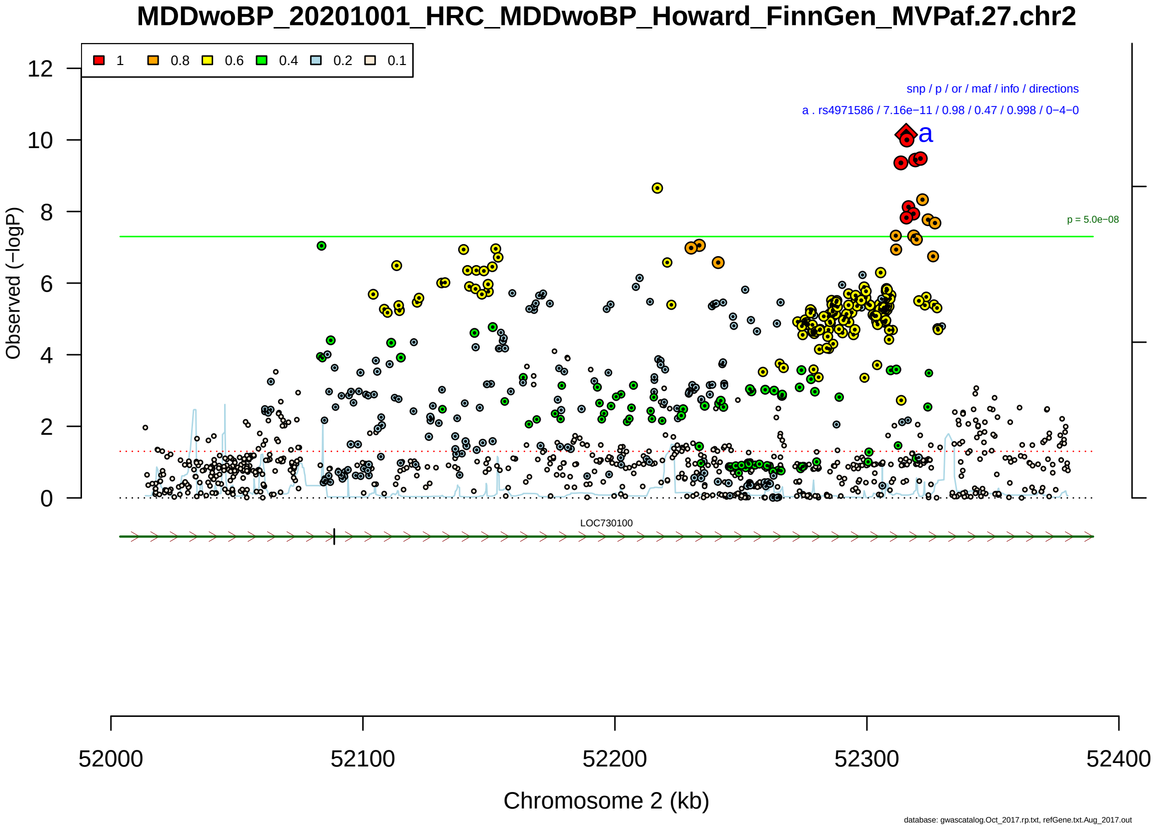

Figure S2-83: Regional plot of GWAS locus No. 83 , with rs4971586 as lead SNP.

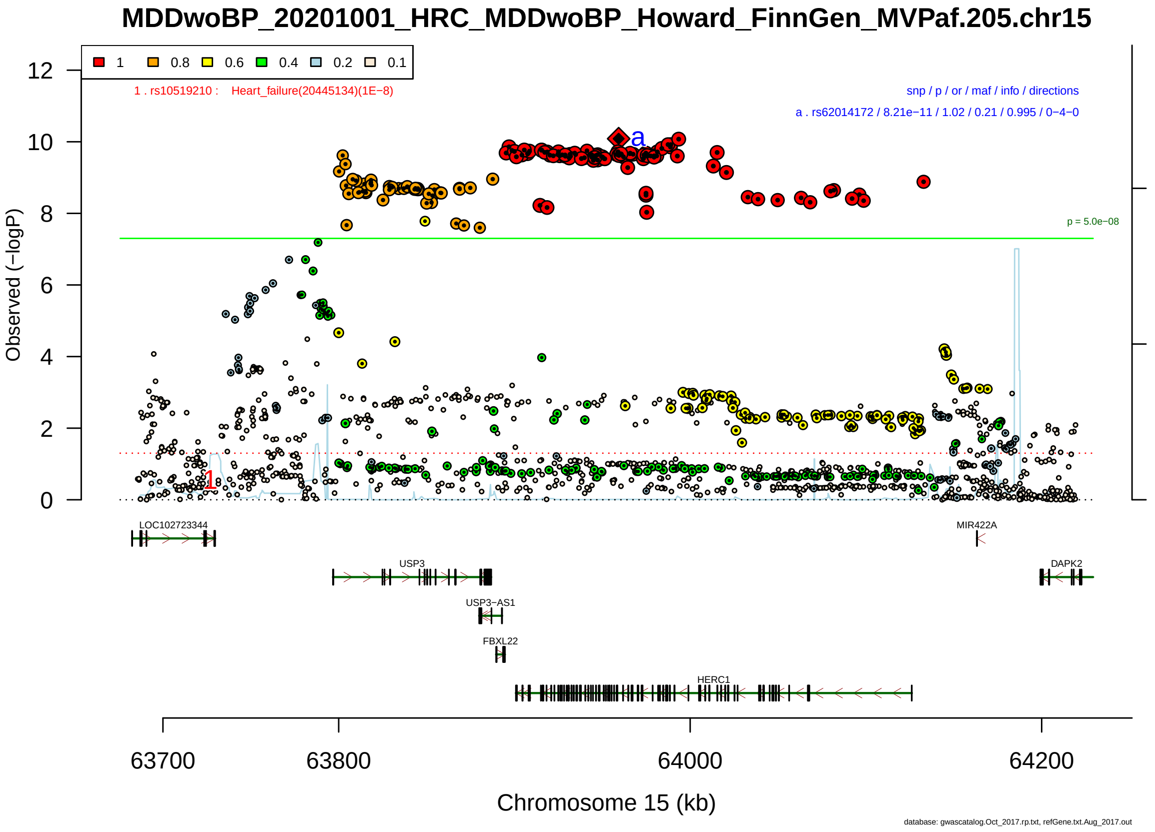

Figure S2-84: Regional plot of GWAS locus No. 84 , with rs62014172 as lead SNP.

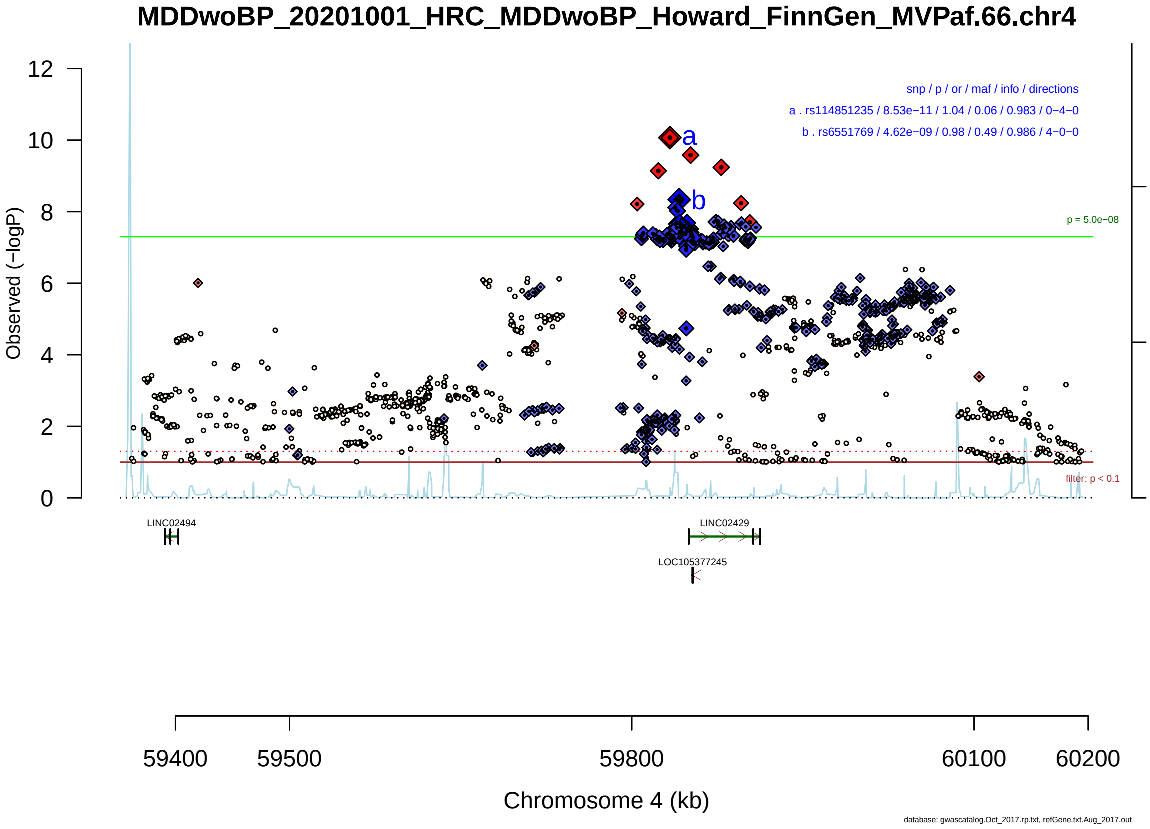

Figure S2-85: Regional plot of GWAS locus No. 85 , with rs114851235 as lead SNP.

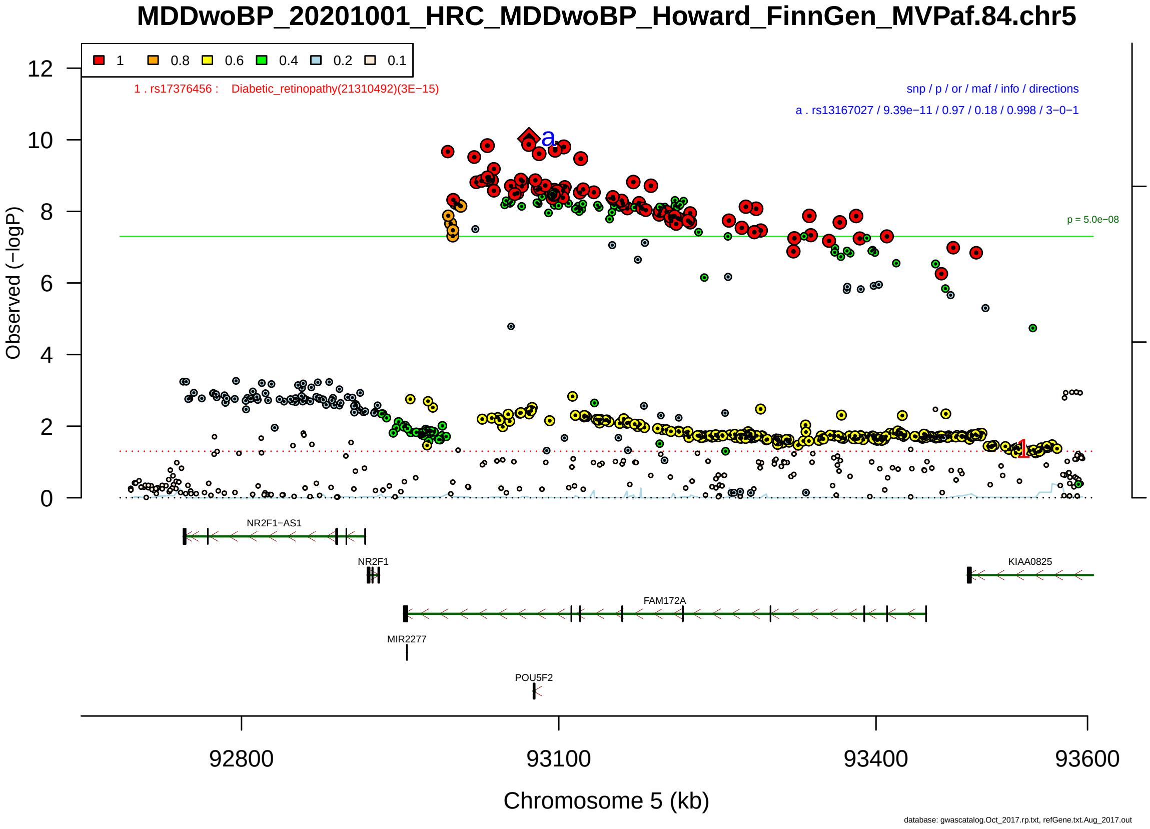

Figure S2-86: Regional plot of GWAS locus No. 86 , with rs13167027 as lead SNP.

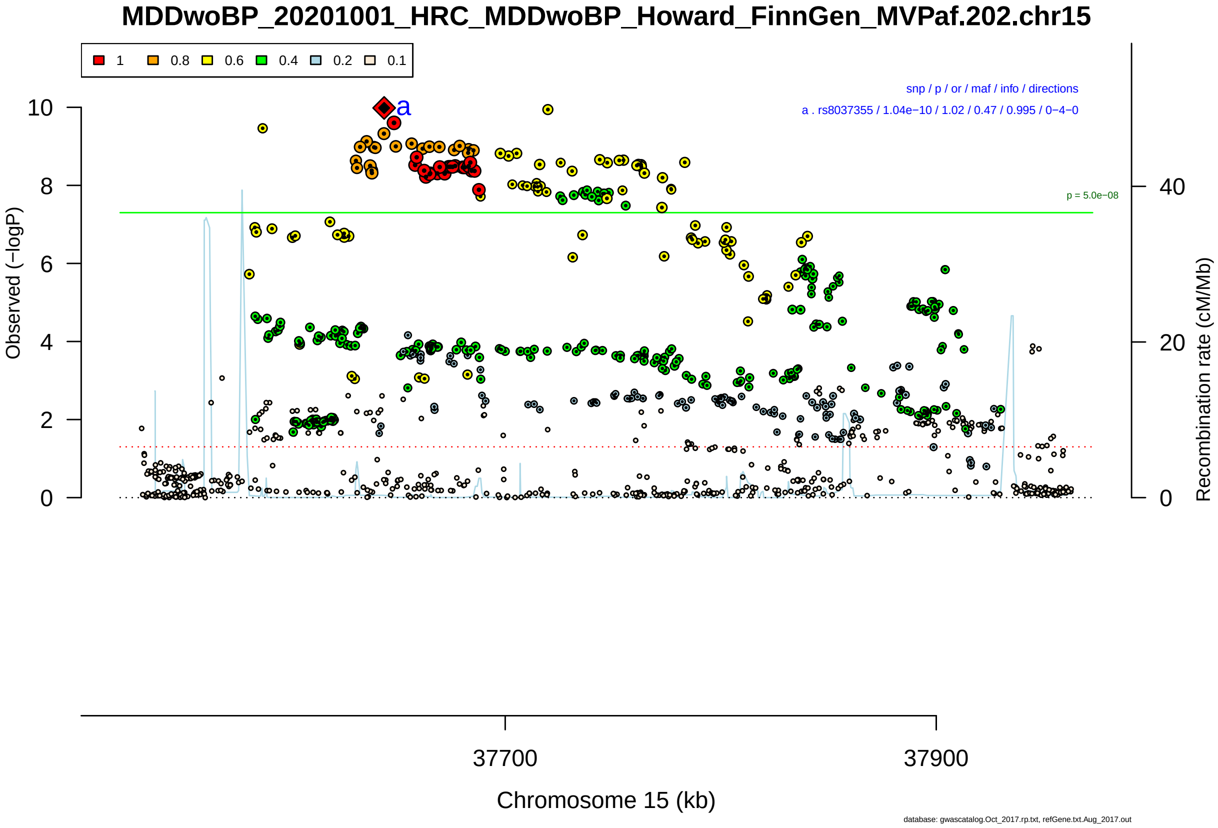

Figure S2-87: Regional plot of GWAS locus No. 87 , with rs8037355 as lead SNP.

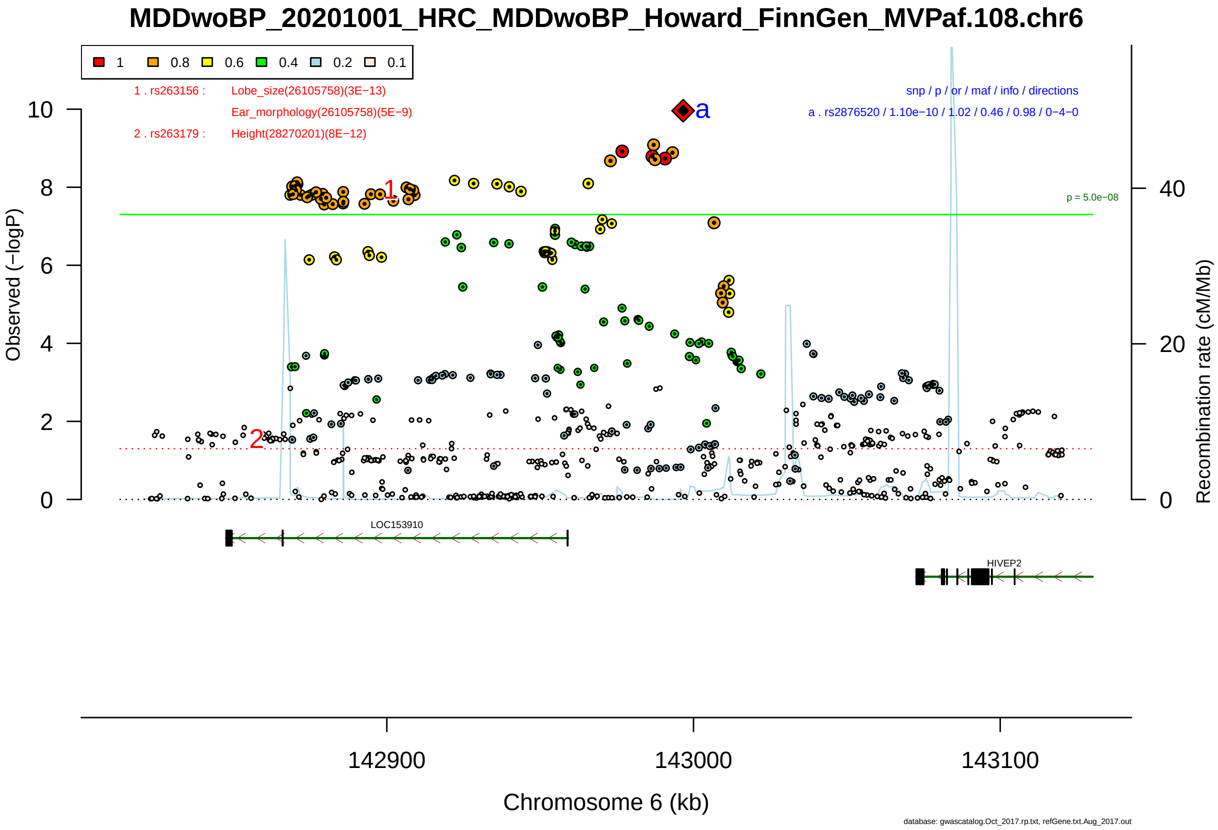

Figure S2-88: Regional plot of GWAS locus No. 88 , with rs2876520 as lead SNP.

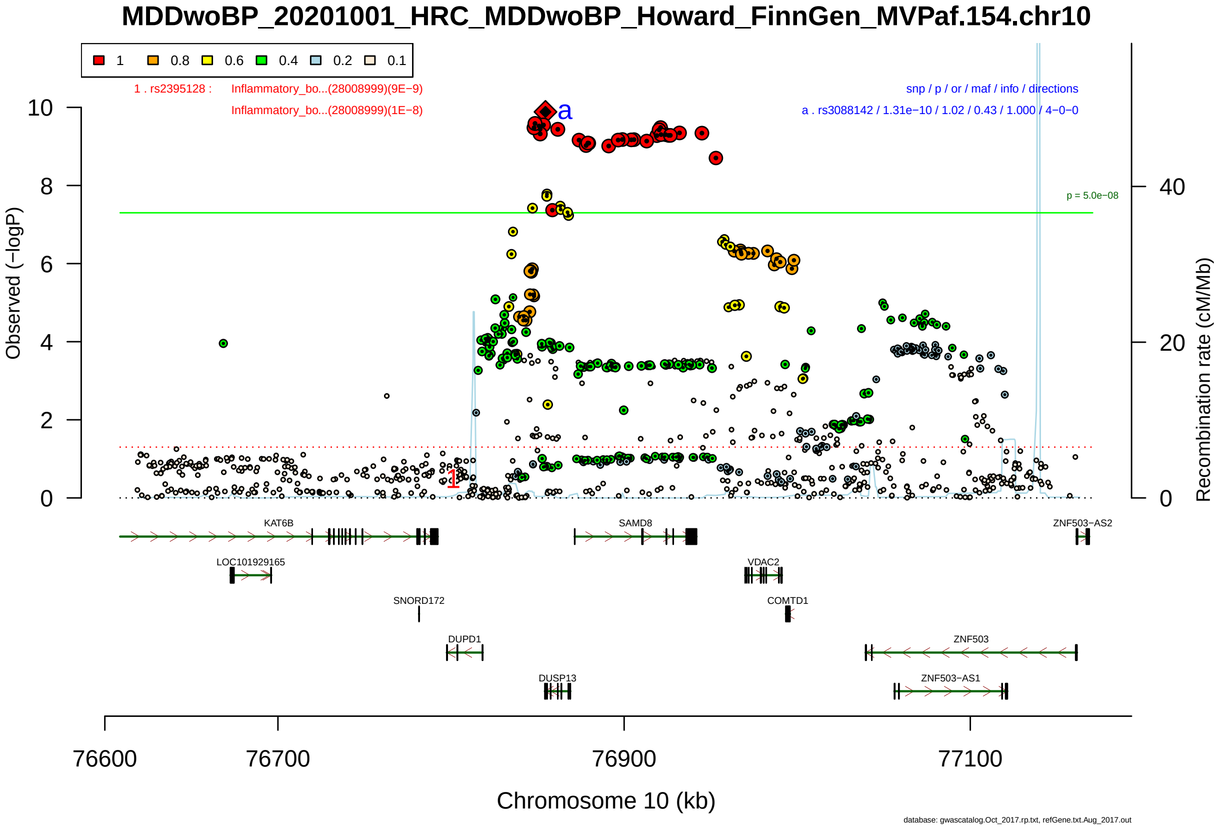

Figure S2-89: Regional plot of GWAS locus No. 89 , with rs3088142 as lead SNP.

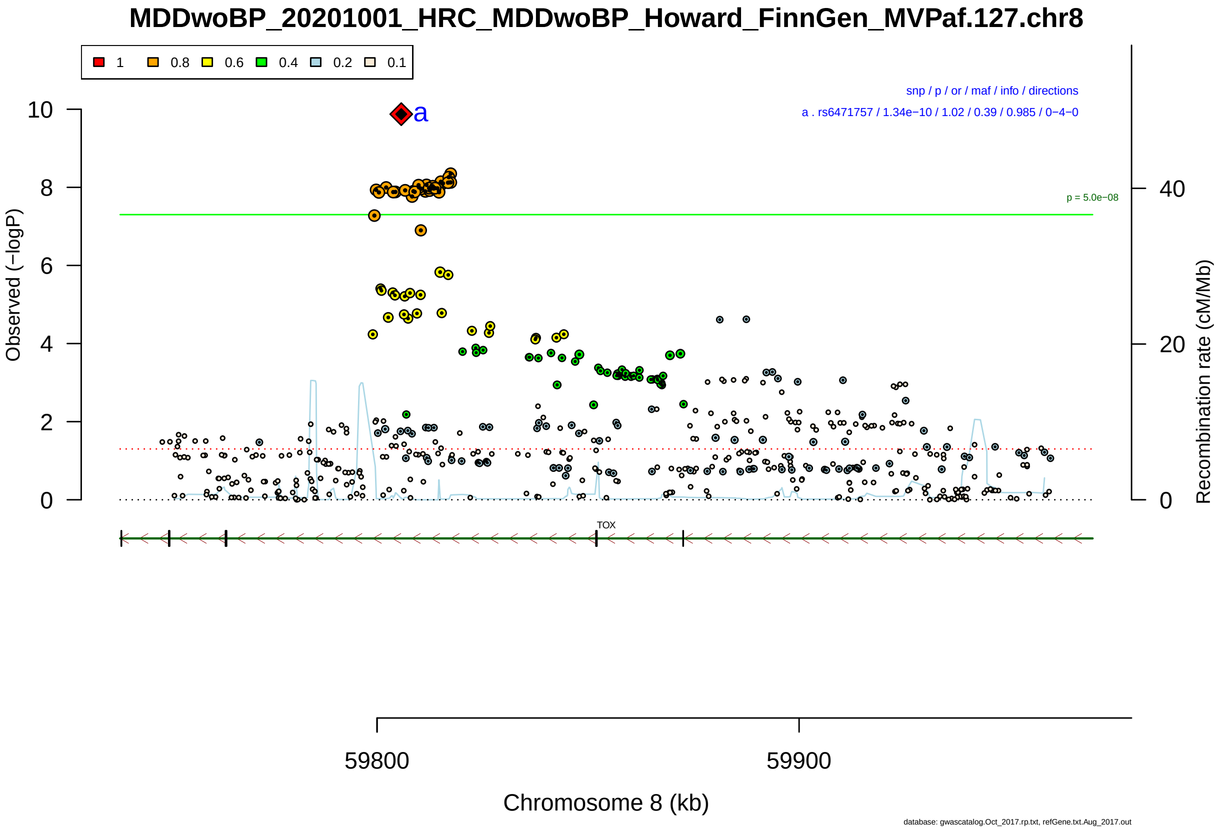

Figure S2-90: Regional plot of GWAS locus No. 90 , with rs6471757 as lead SNP.

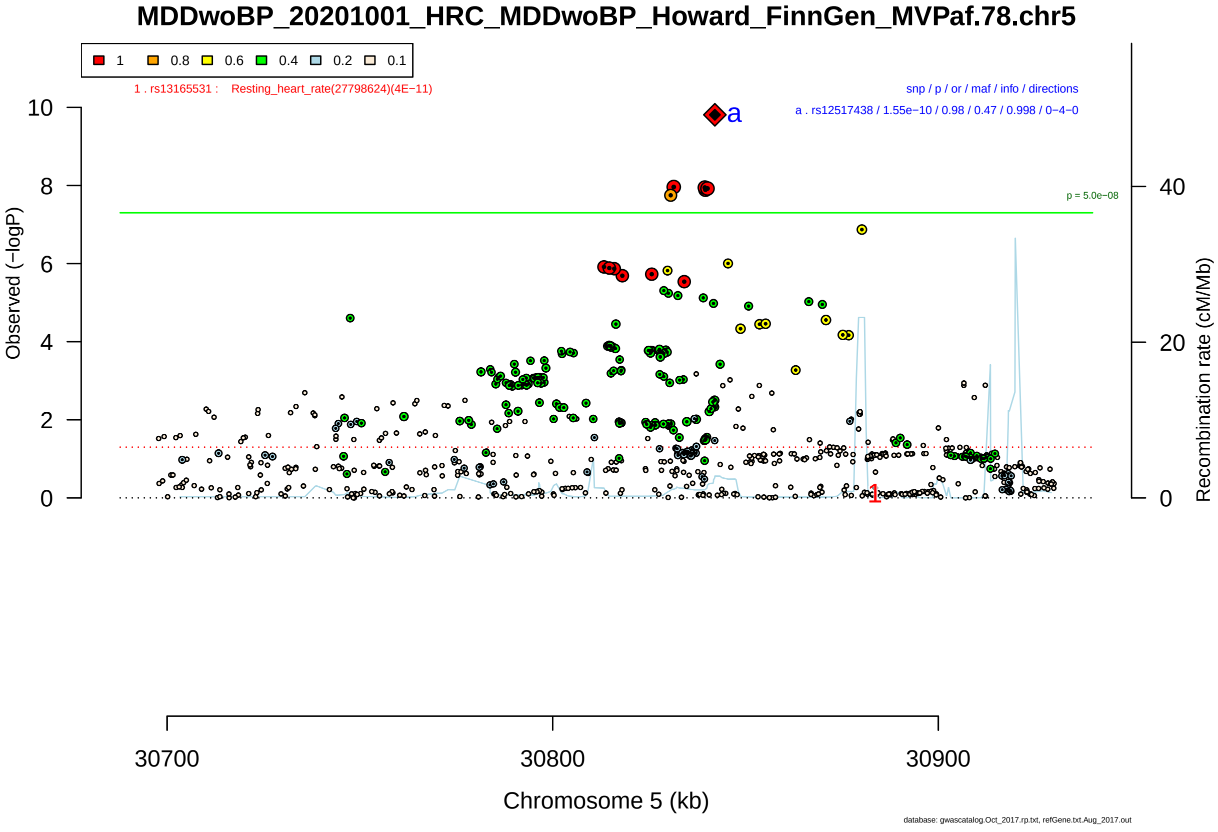

Figure S2-91: Regional plot of GWAS locus No. 91 , with rs12517438 as lead SNP.

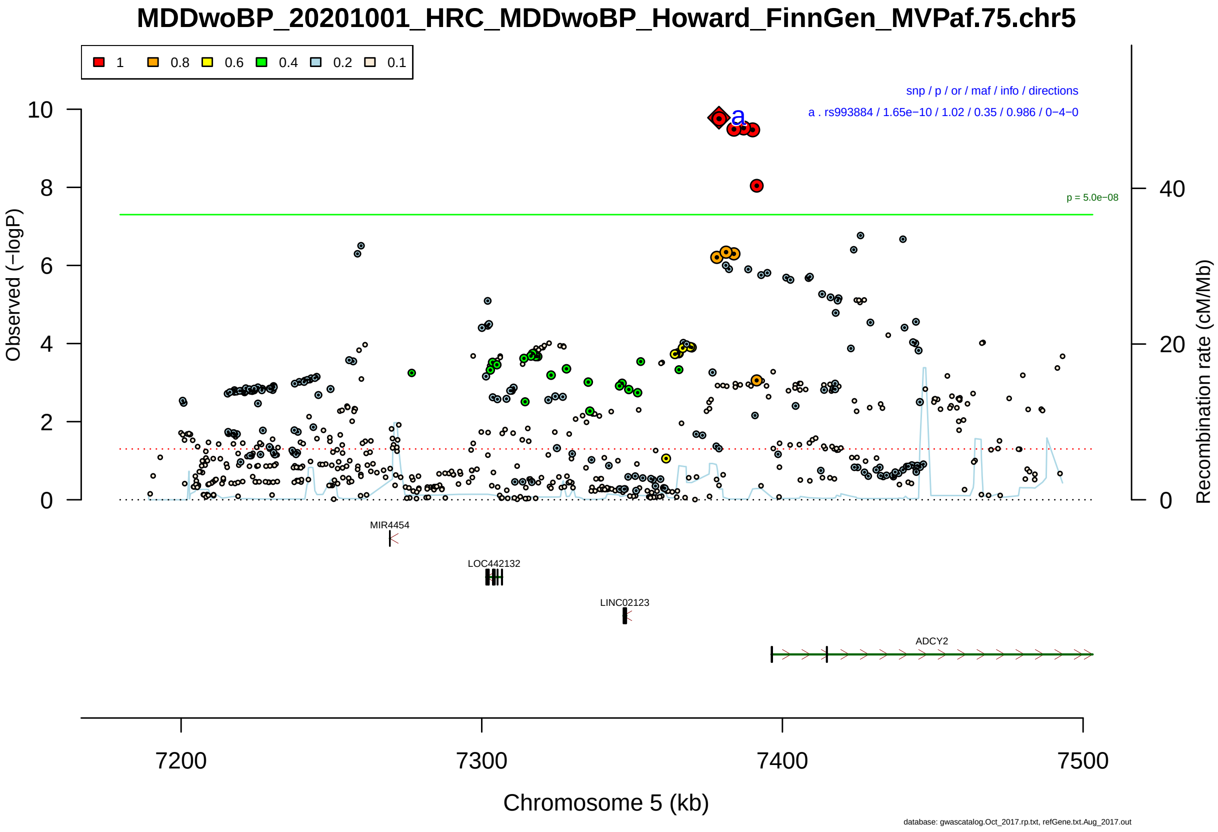

Figure S2-92: Regional plot of GWAS locus No. 92 , with rs993884 as lead SNP.

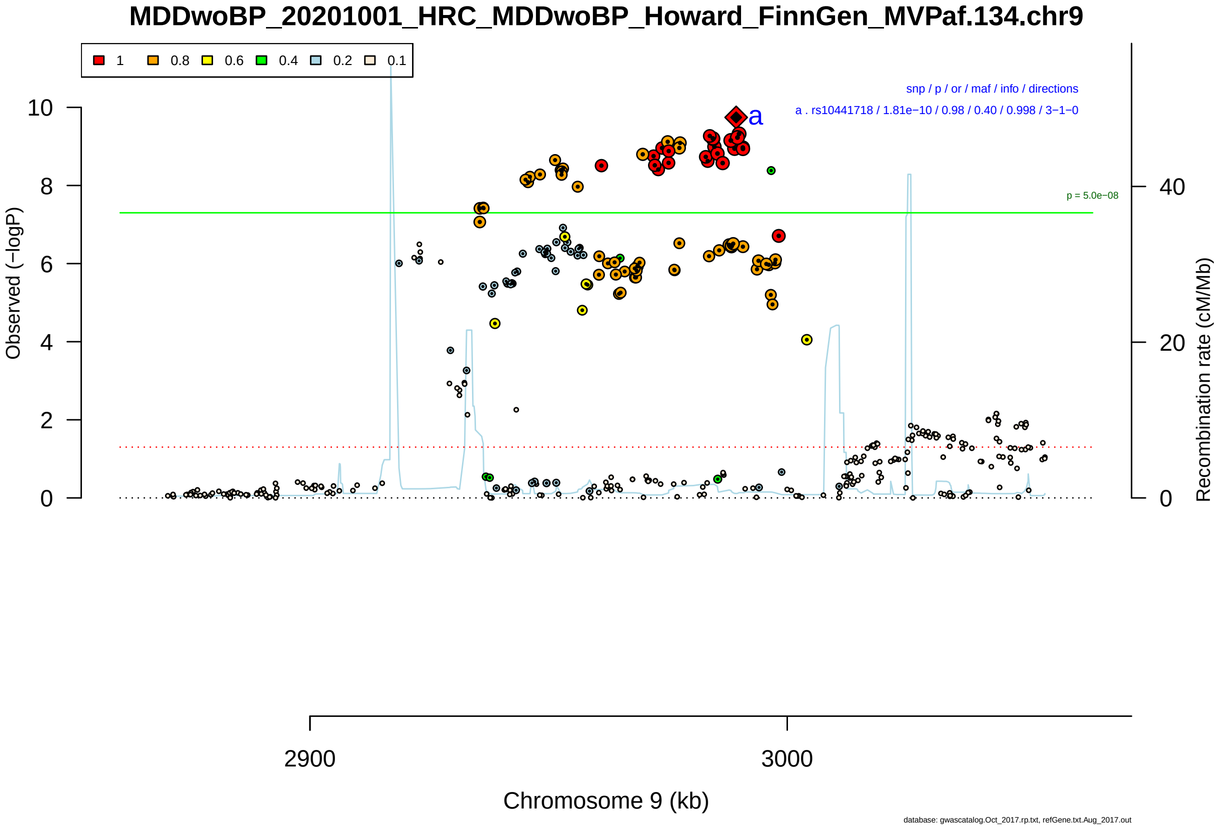

Figure S2-93: Regional plot of GWAS locus No. 93 , with rs10441718 as lead SNP.

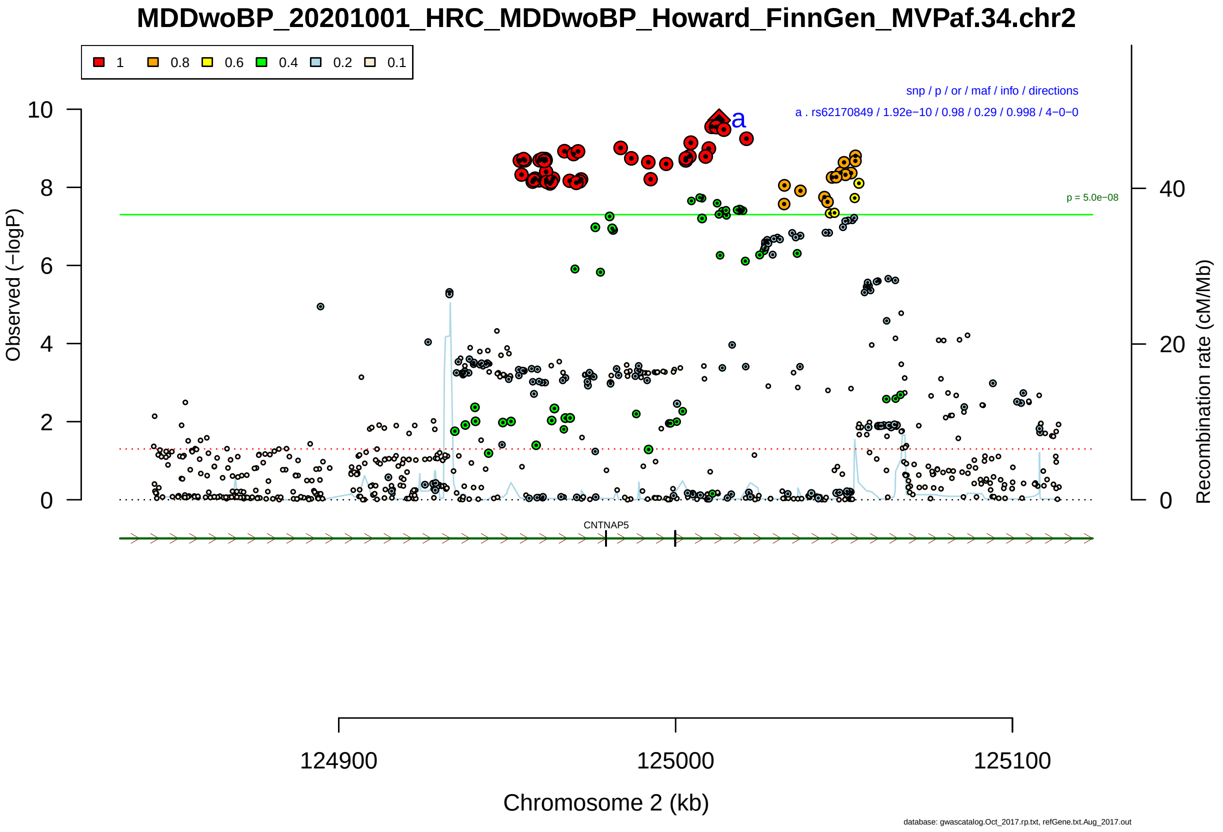

Figure S2-94: Regional plot of GWAS locus No. 94 , with rs62170849 as lead SNP.

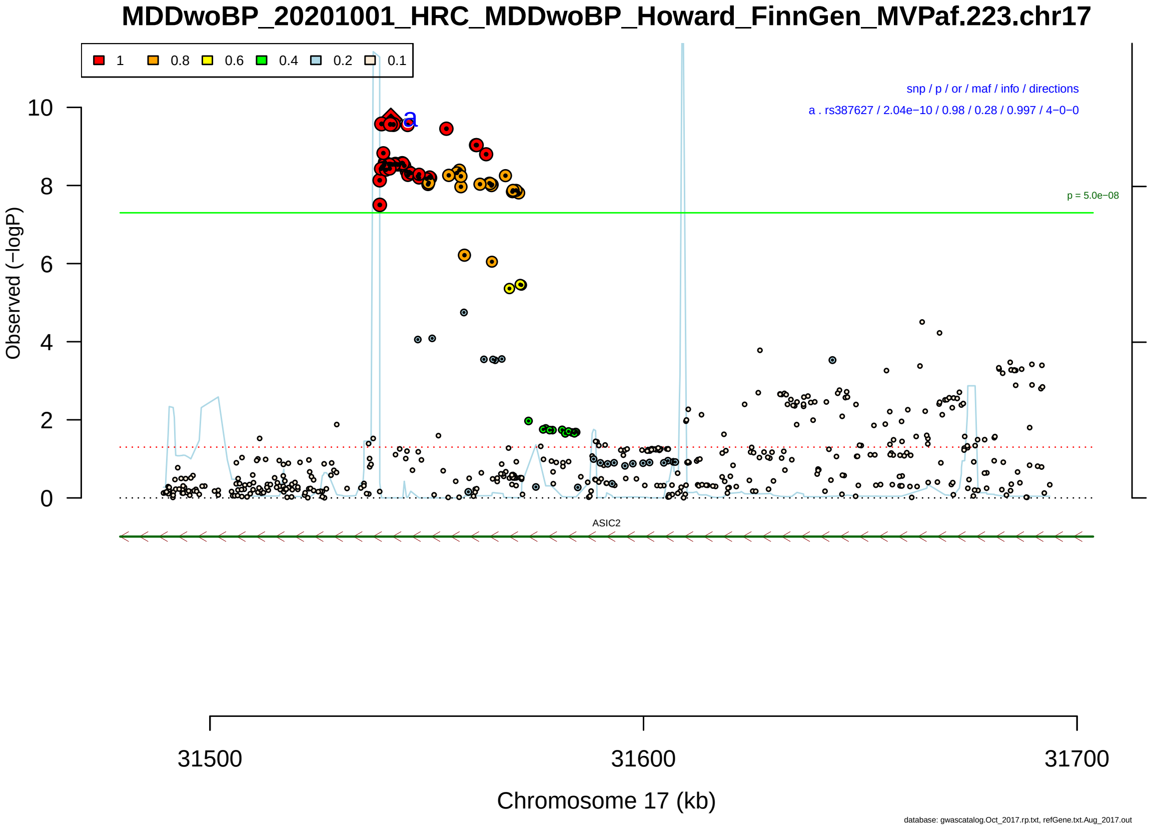

Figure S2-95: Regional plot of GWAS locus No. 95 , with rs387627 as lead SNP.

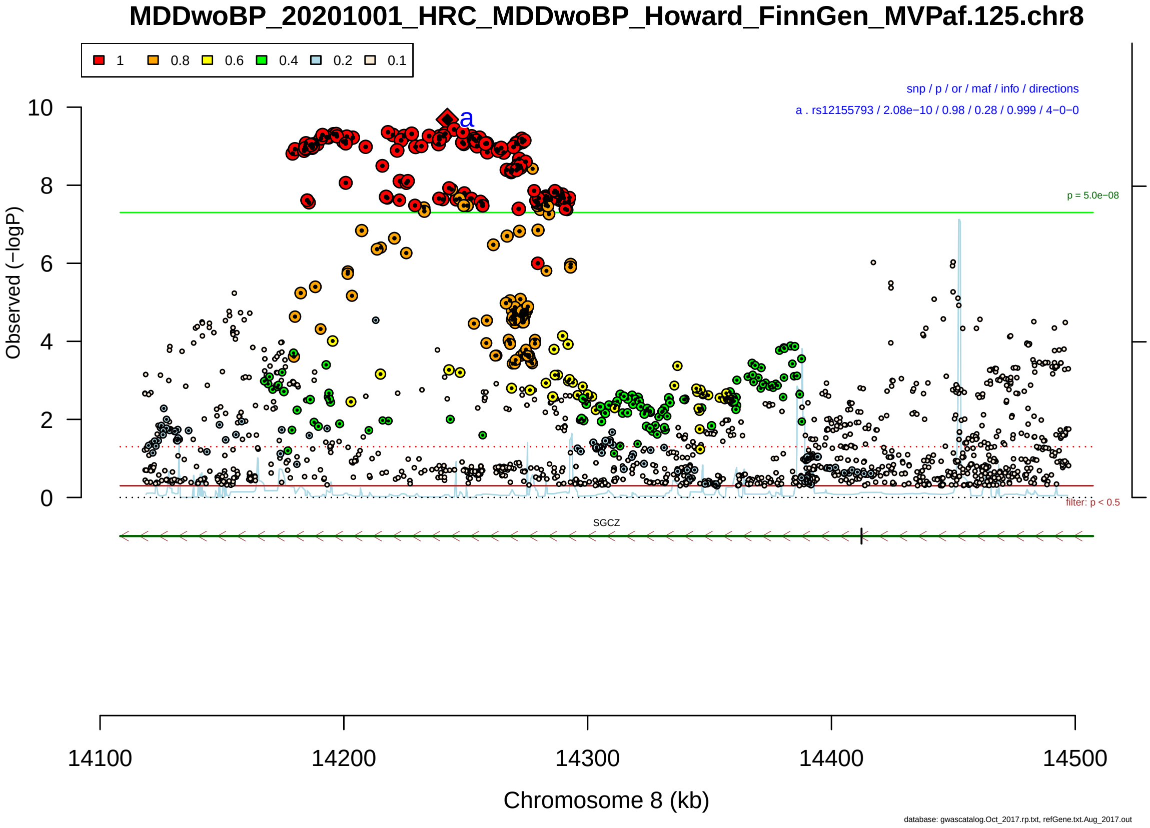

Figure S2-96: Regional plot of GWAS locus No. 96 , with rs12155793 as lead SNP.

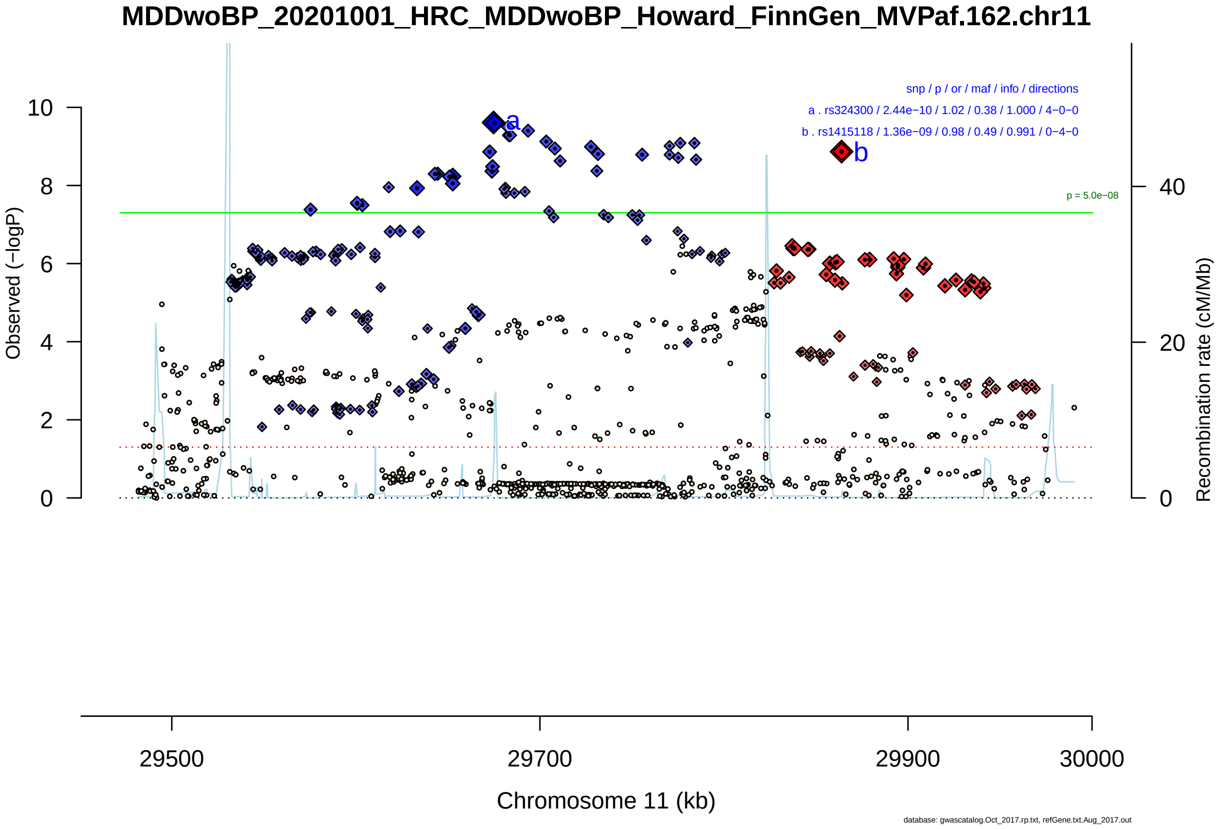

Figure S2-97: Regional plot of GWAS locus No. 97 , with rs324300 as lead SNP.

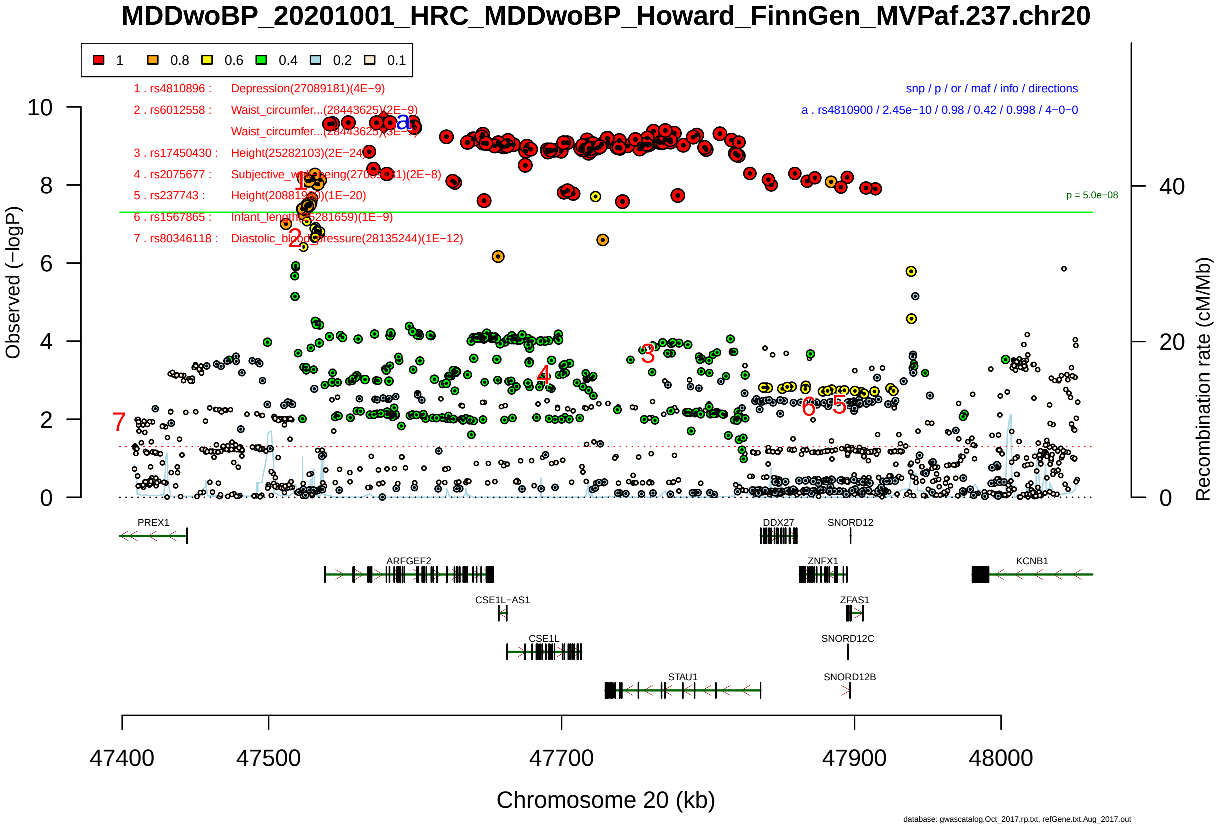

Figure S2-98: Regional plot of GWAS locus No. 98 , with rs4810900 as lead SNP.

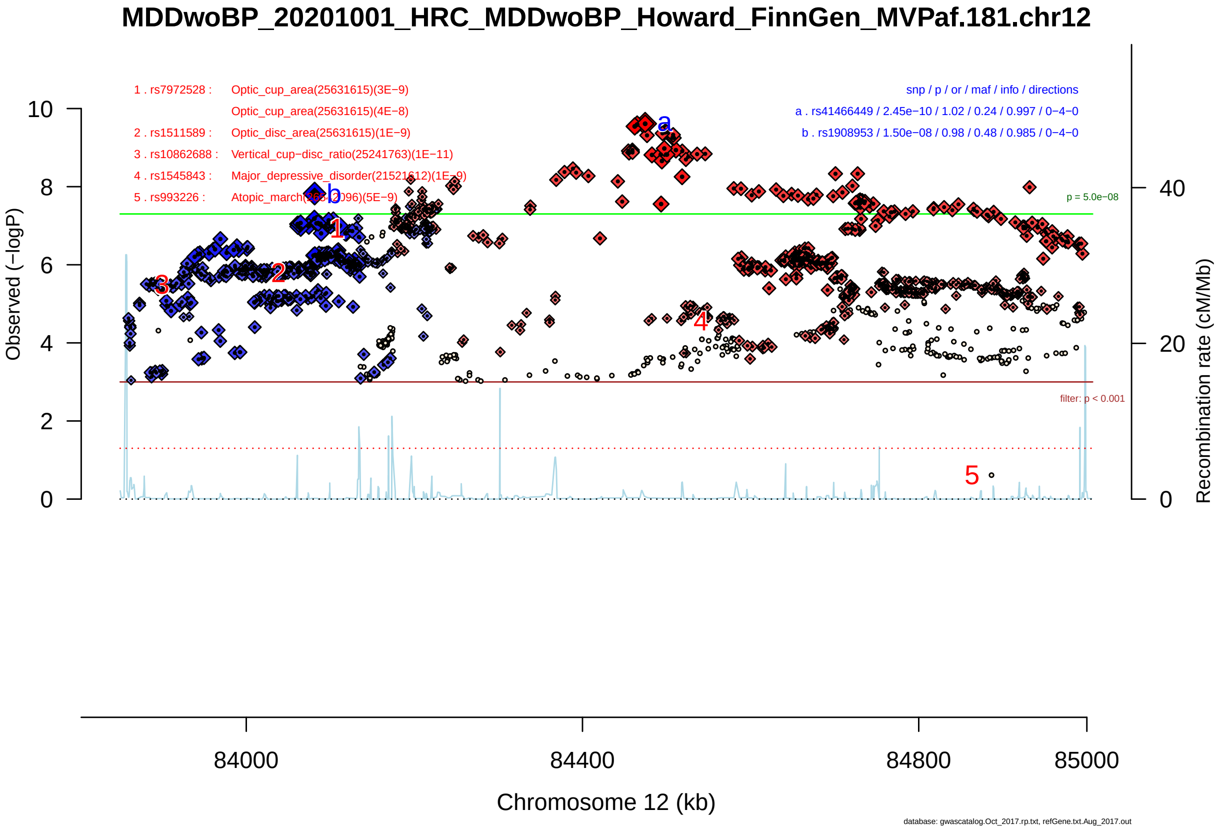

Figure S2-99: Regional plot of GWAS locus No. 99 , with rs41466449 as lead SNP.

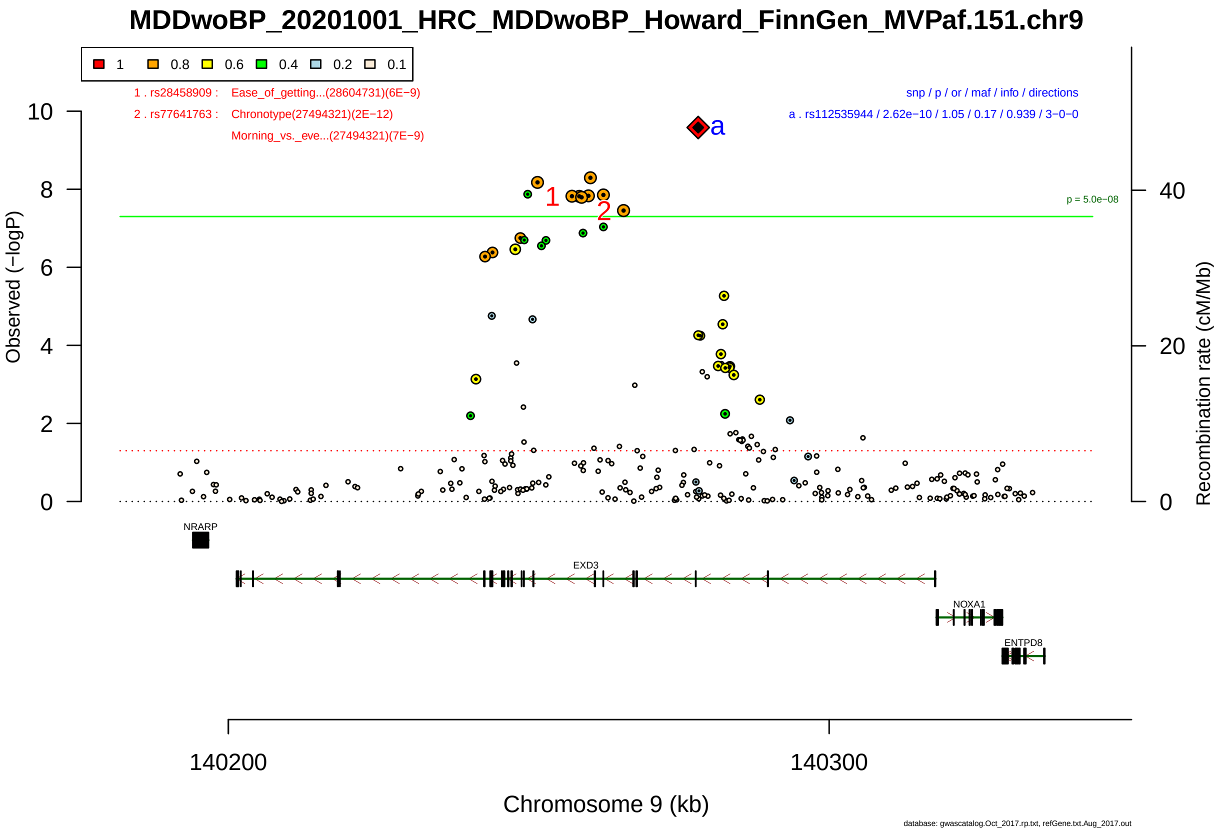

Figure S2-100: Regional plot of GWAS locus No. 100 , with rs112535944 as lead SNP.

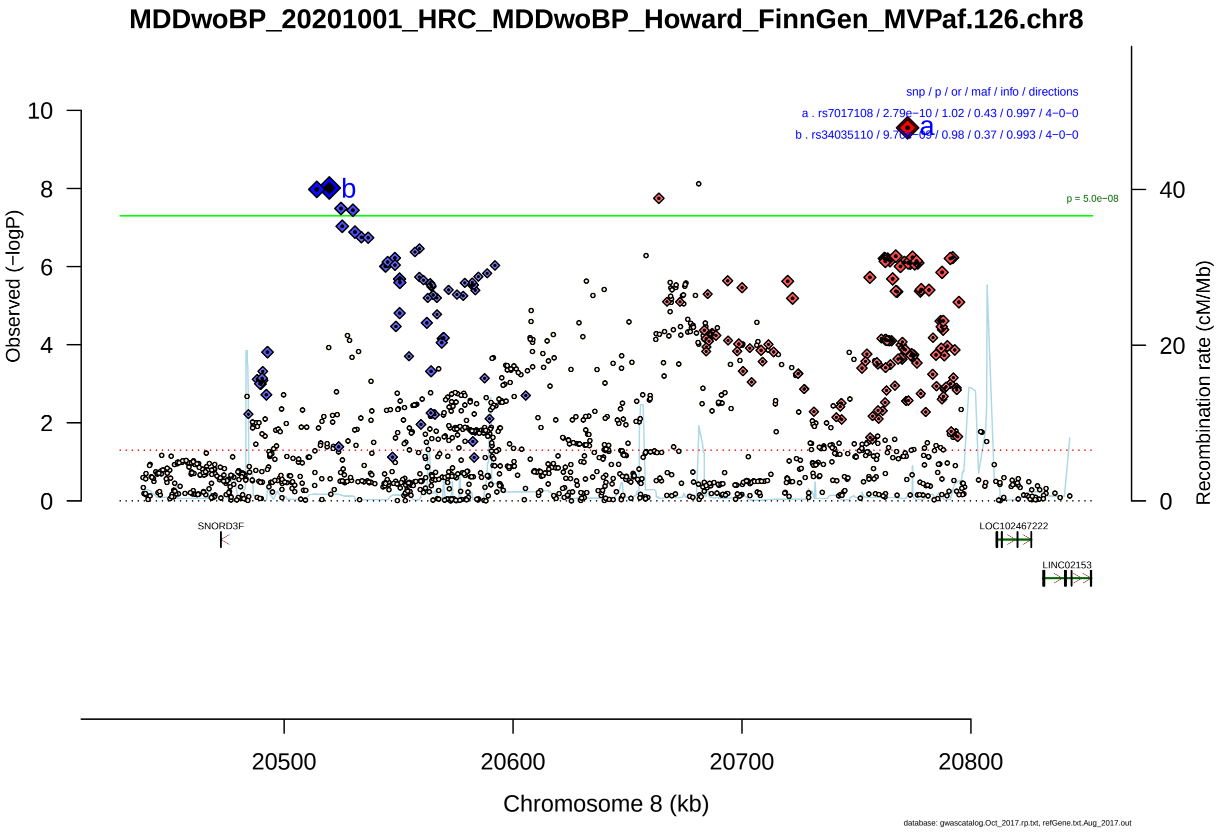

Figure S2-101: Regional plot of GWAS locus No. 101 , with rs7017108 as lead SNP.

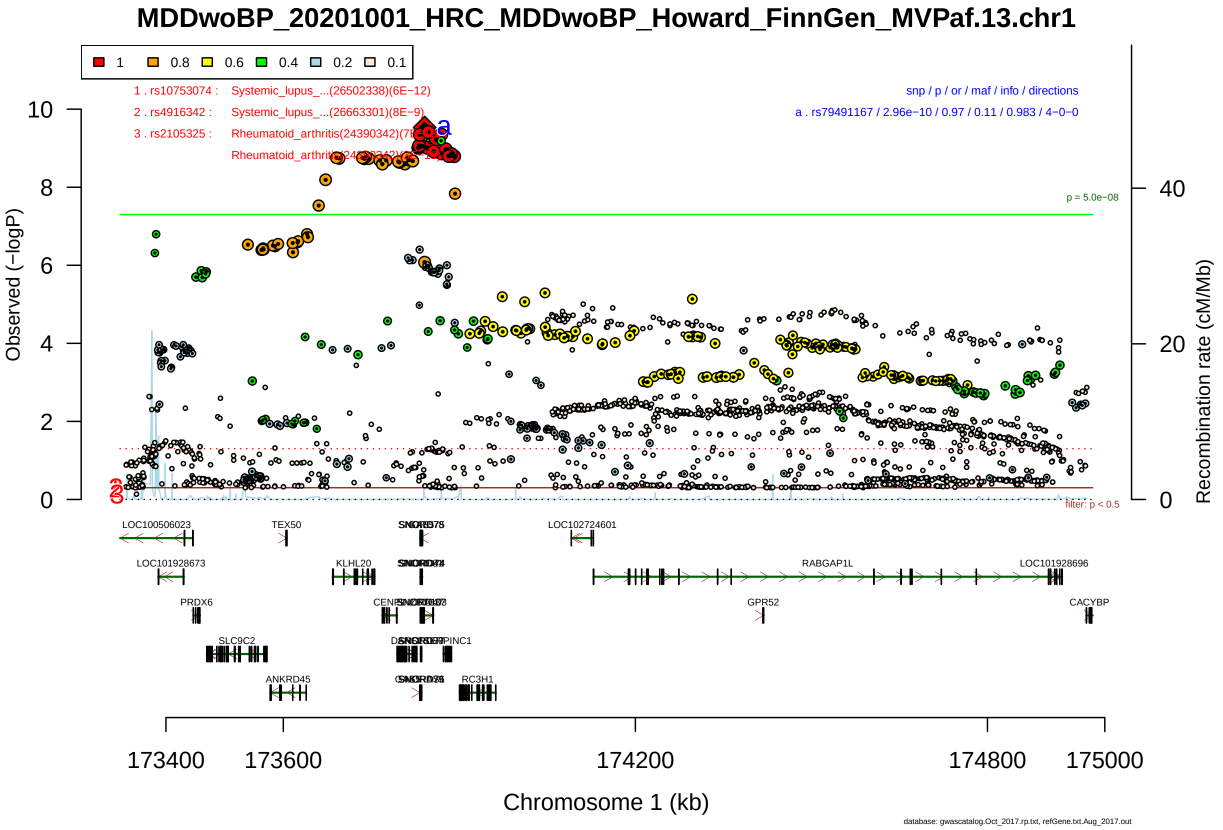

Figure S2-102: Regional plot of GWAS locus No. 102 , with rs79491167 as lead SNP.

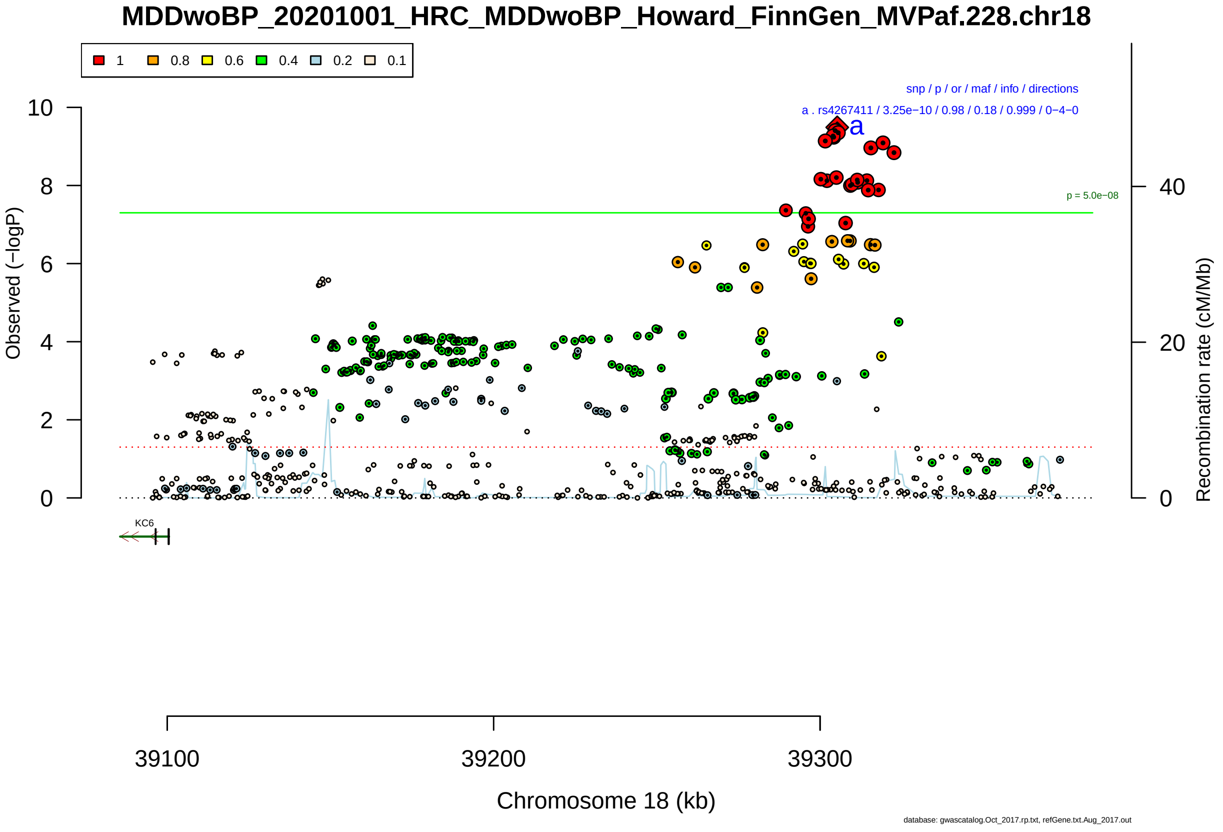

Figure S2-103: Regional plot of GWAS locus No. 103 , with rs4267411 as lead SNP.

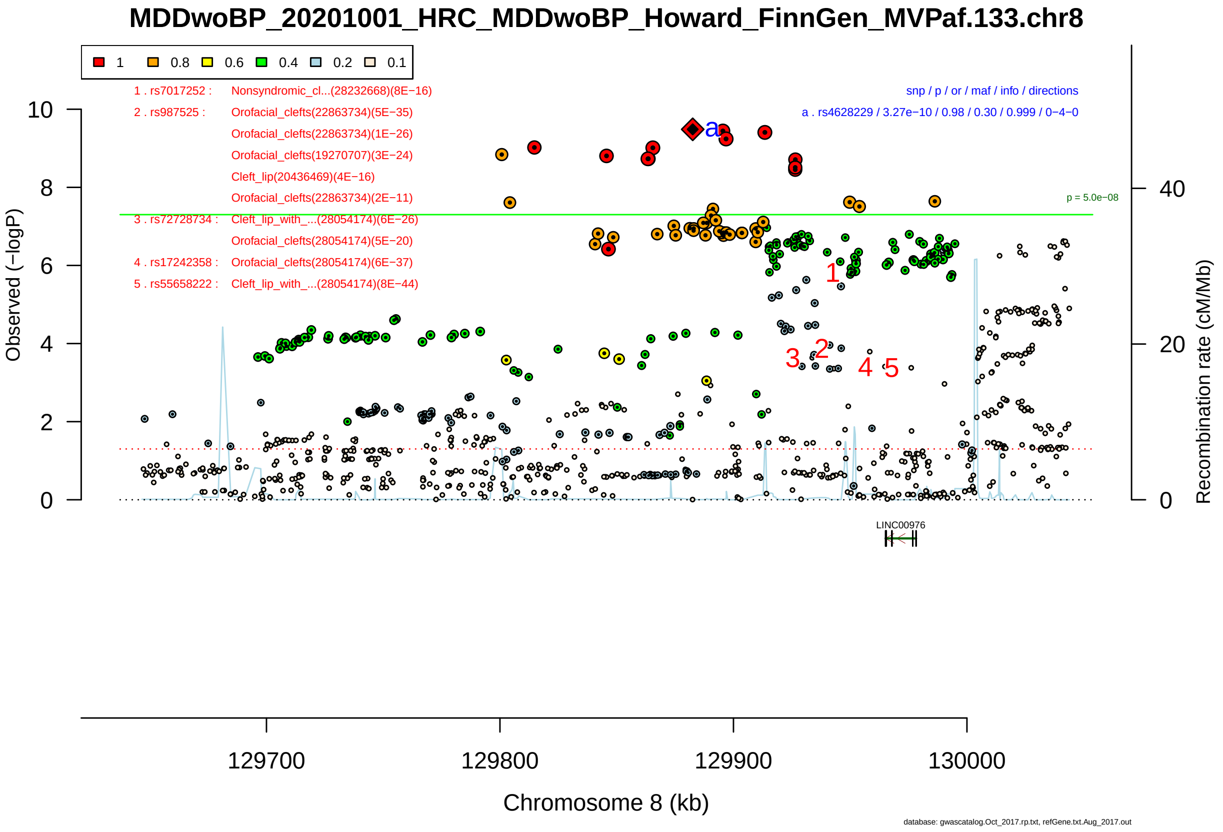

Figure S2-104: Regional plot of GWAS locus No. 104 , with rs4628229 as lead SNP.

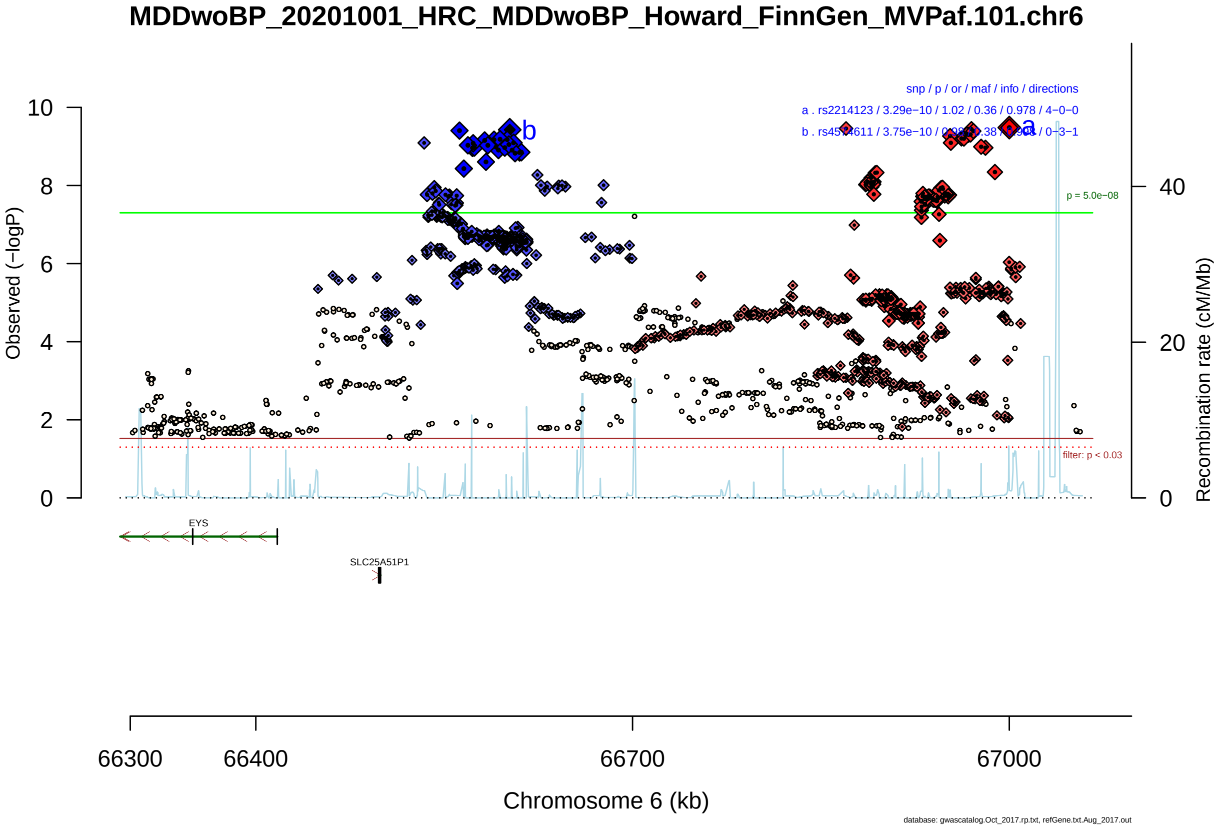

Figure S2-105: Regional plot of GWAS locus No. 105 , with rs2214123 as lead SNP.

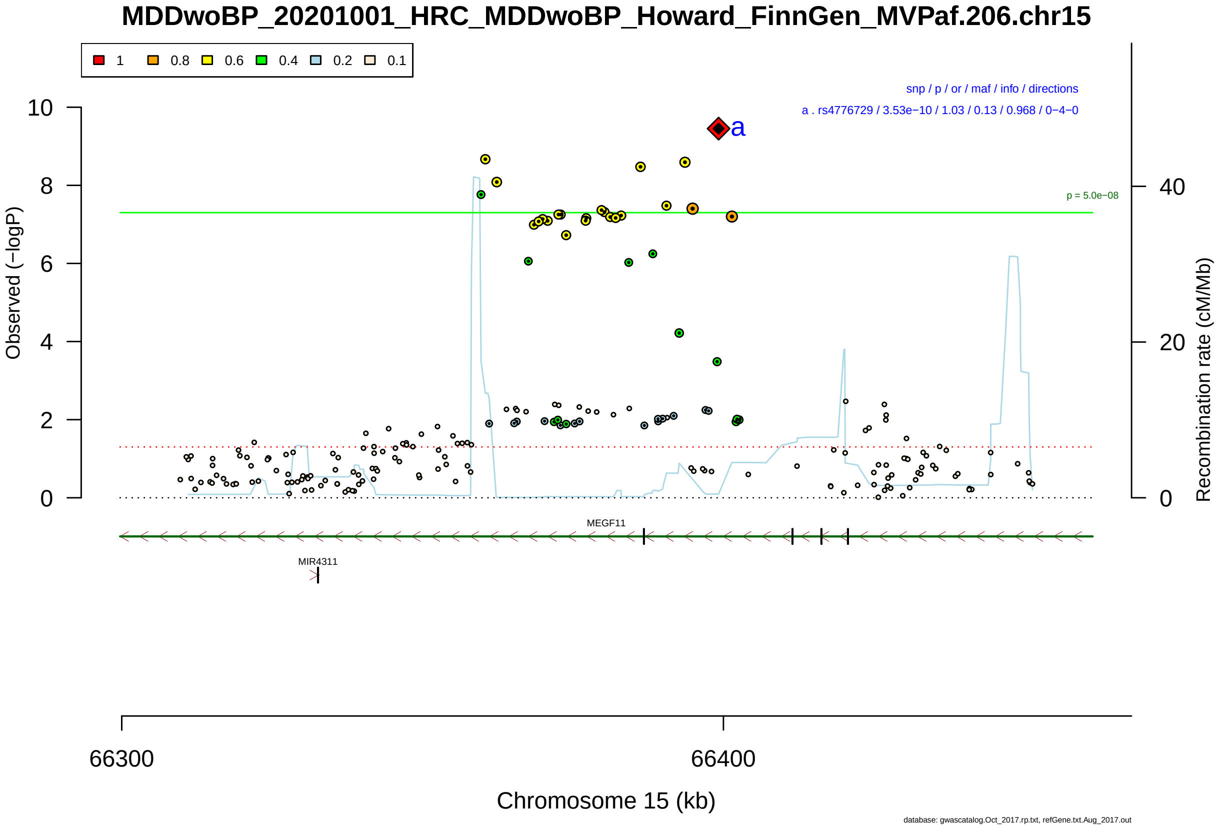

Figure S2-106: Regional plot of GWAS locus No. 106 , with rs4776729 as lead SNP.

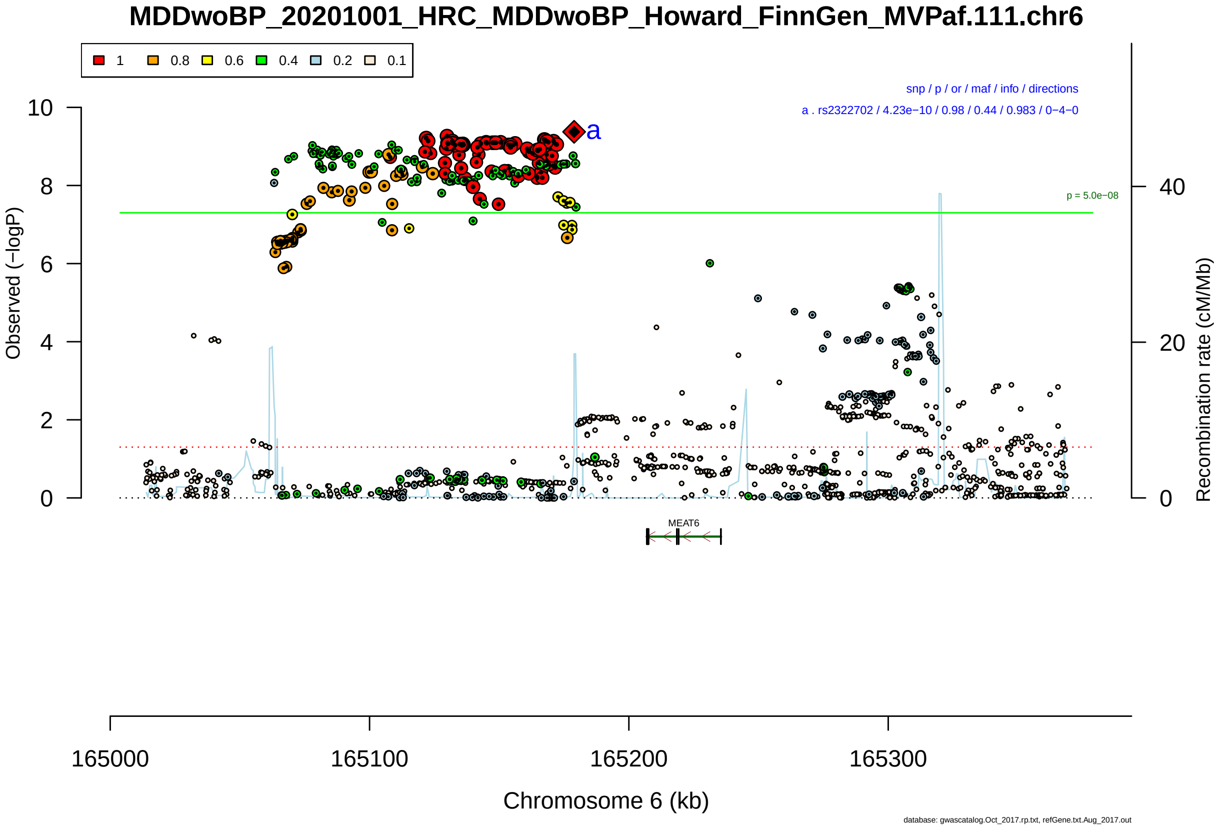

Figure S2-107: Regional plot of GWAS locus No. 107 , with rs2322702 as lead SNP.

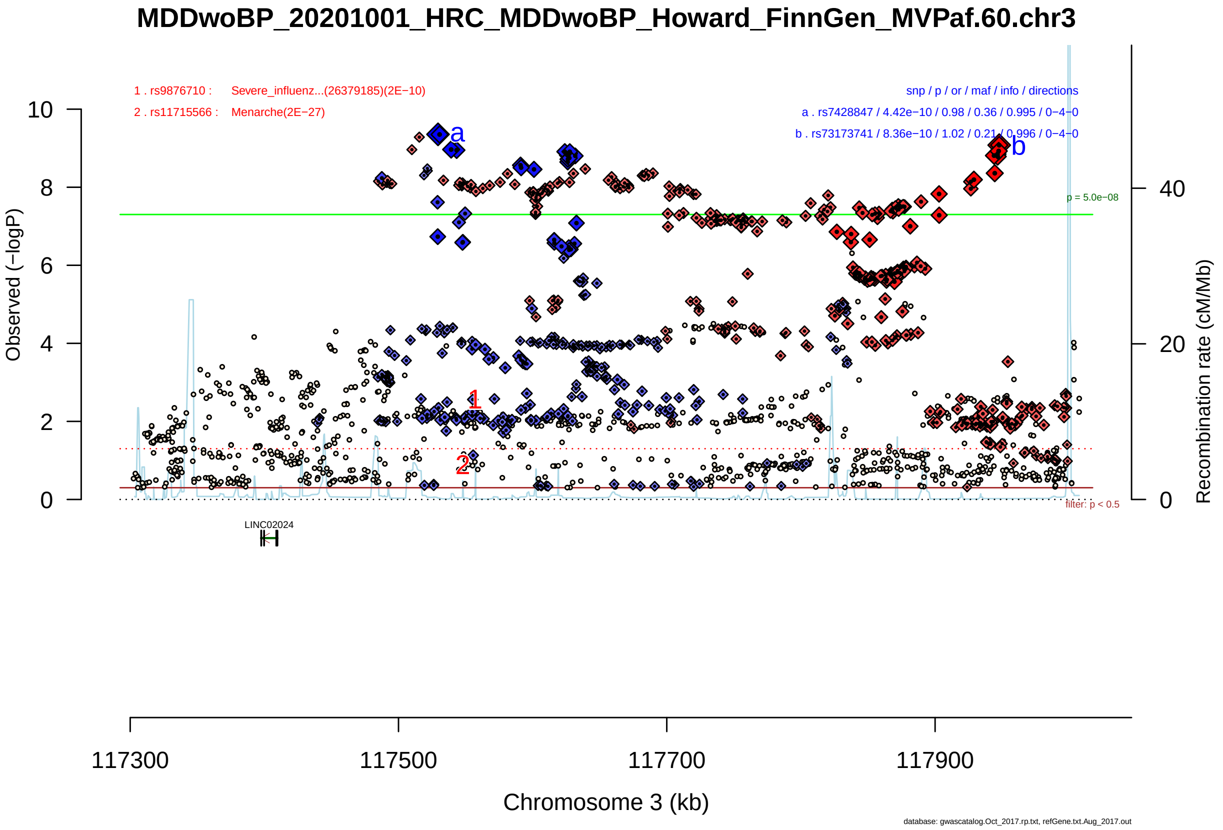

Figure S2-108: Regional plot of GWAS locus No. 108 , with rs7428847 as lead SNP.

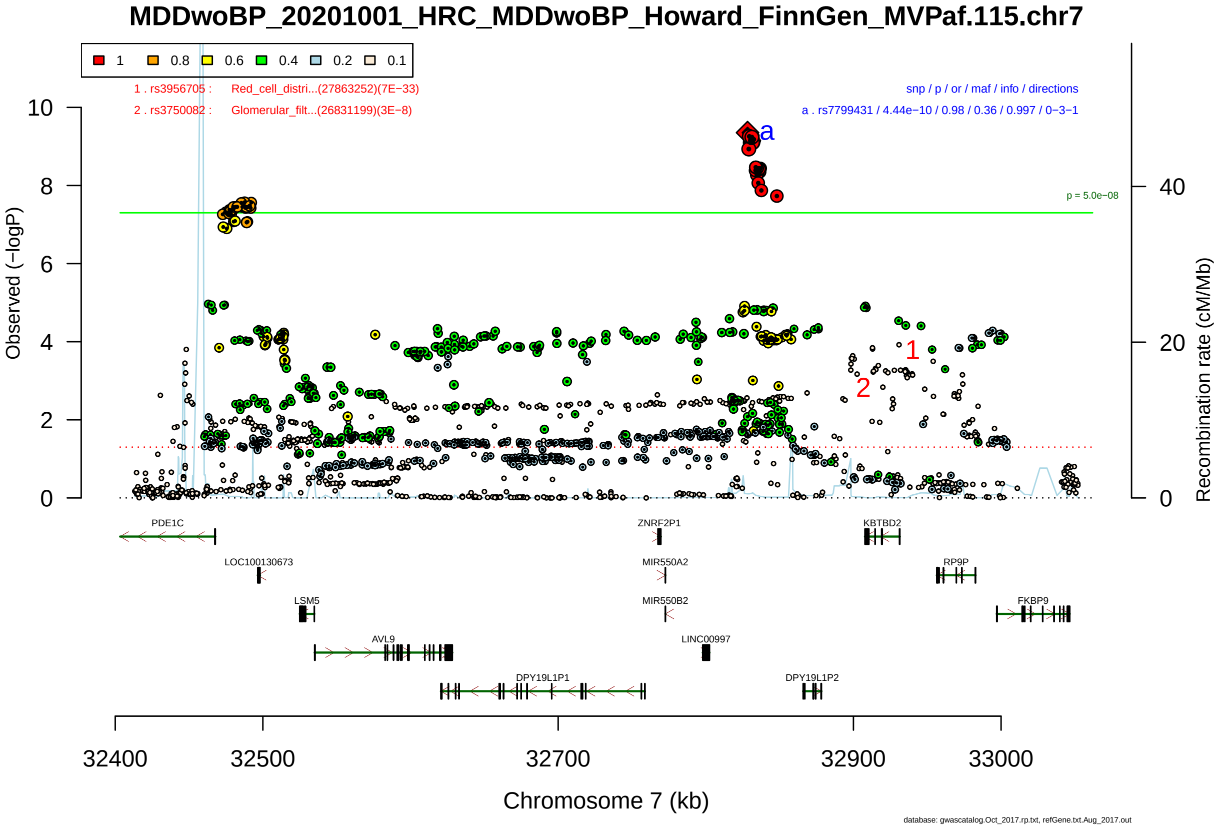

Figure S2-109: Regional plot of GWAS locus No. 109 , with rs7799431 as lead SNP.

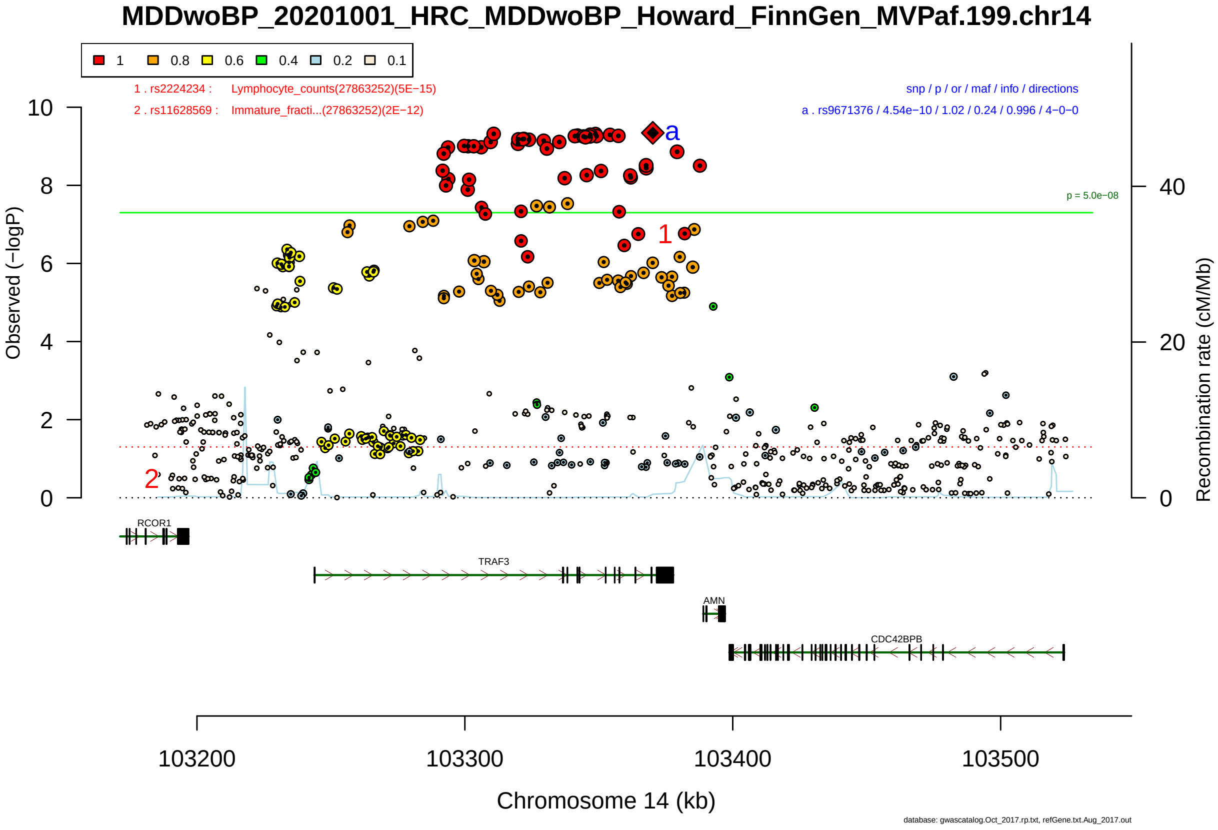

Figure S2-110: Regional plot of GWAS locus No. 110 , with rs9671376 as lead SNP.

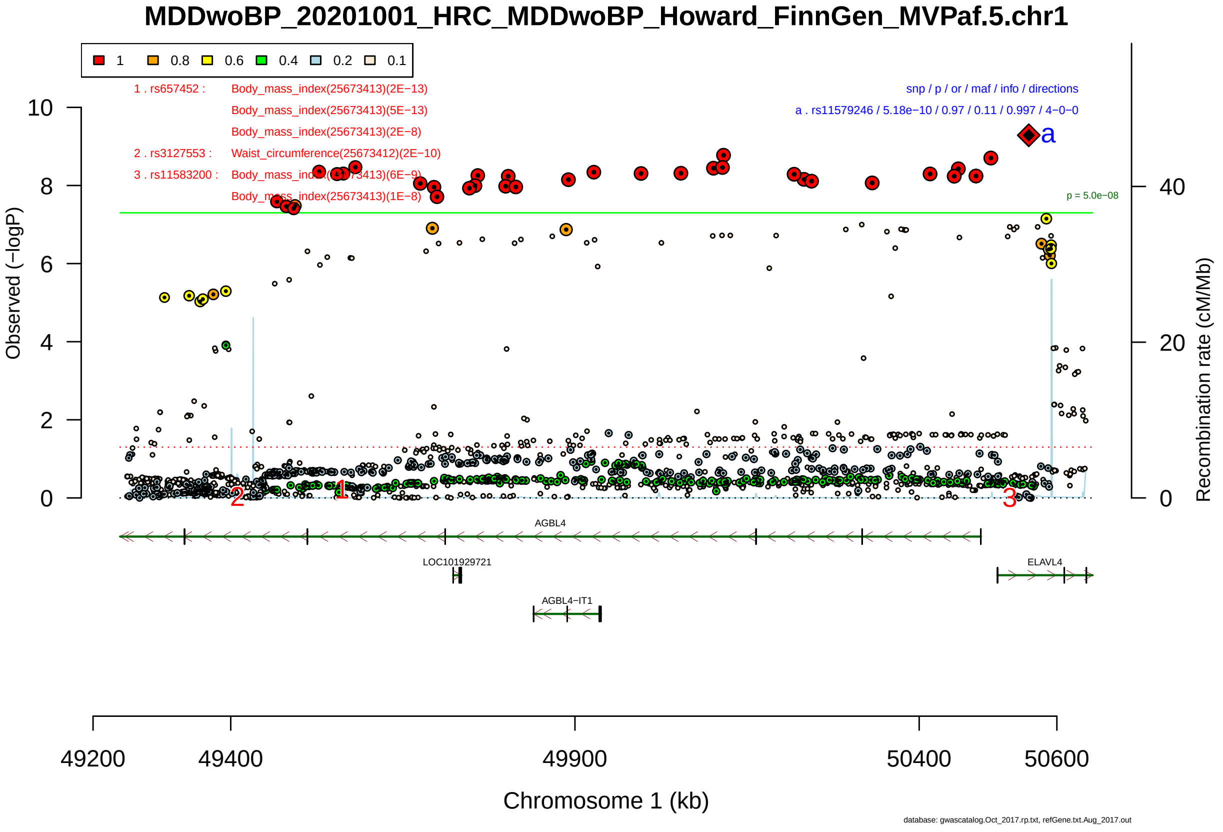

Figure S2-111: Regional plot of GWAS locus No. 111 , with rs11579246 as lead SNP.

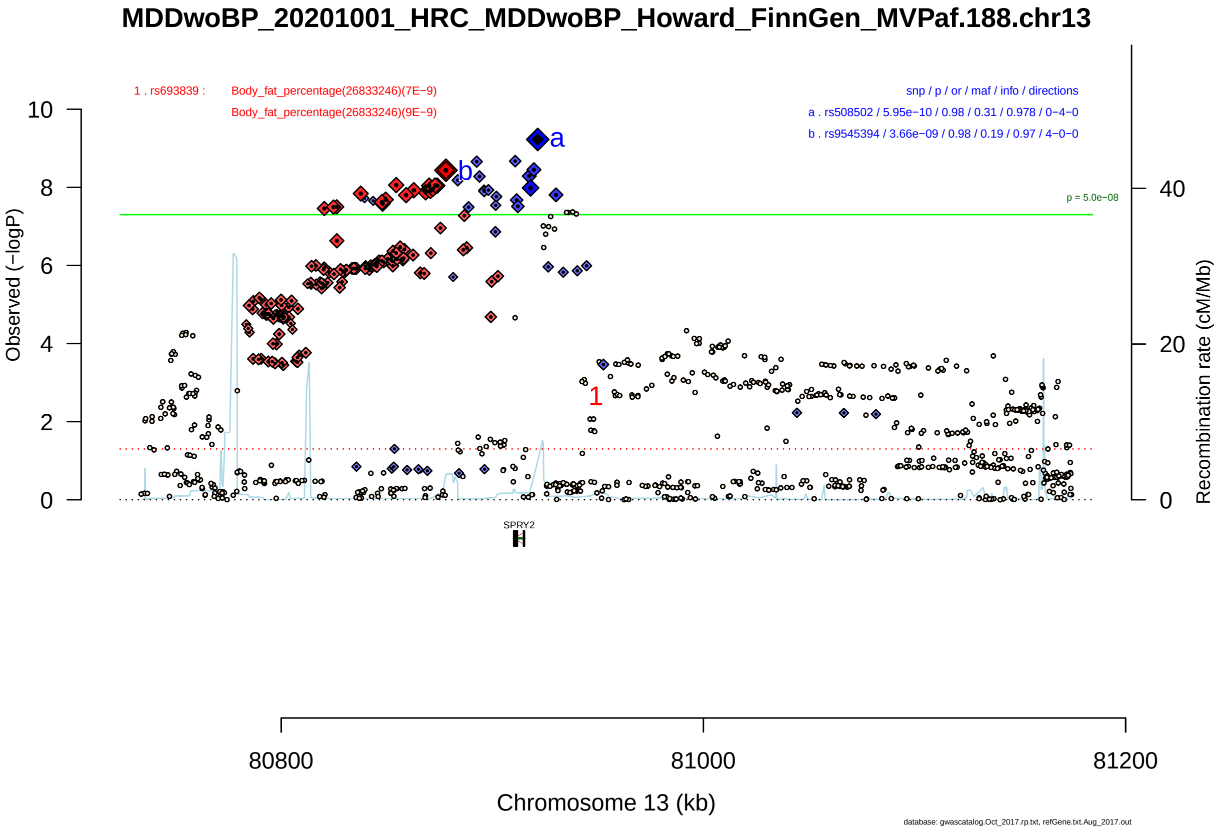

Figure S2-112: Regional plot of GWAS locus No. 112 , with rs508502 as lead SNP.

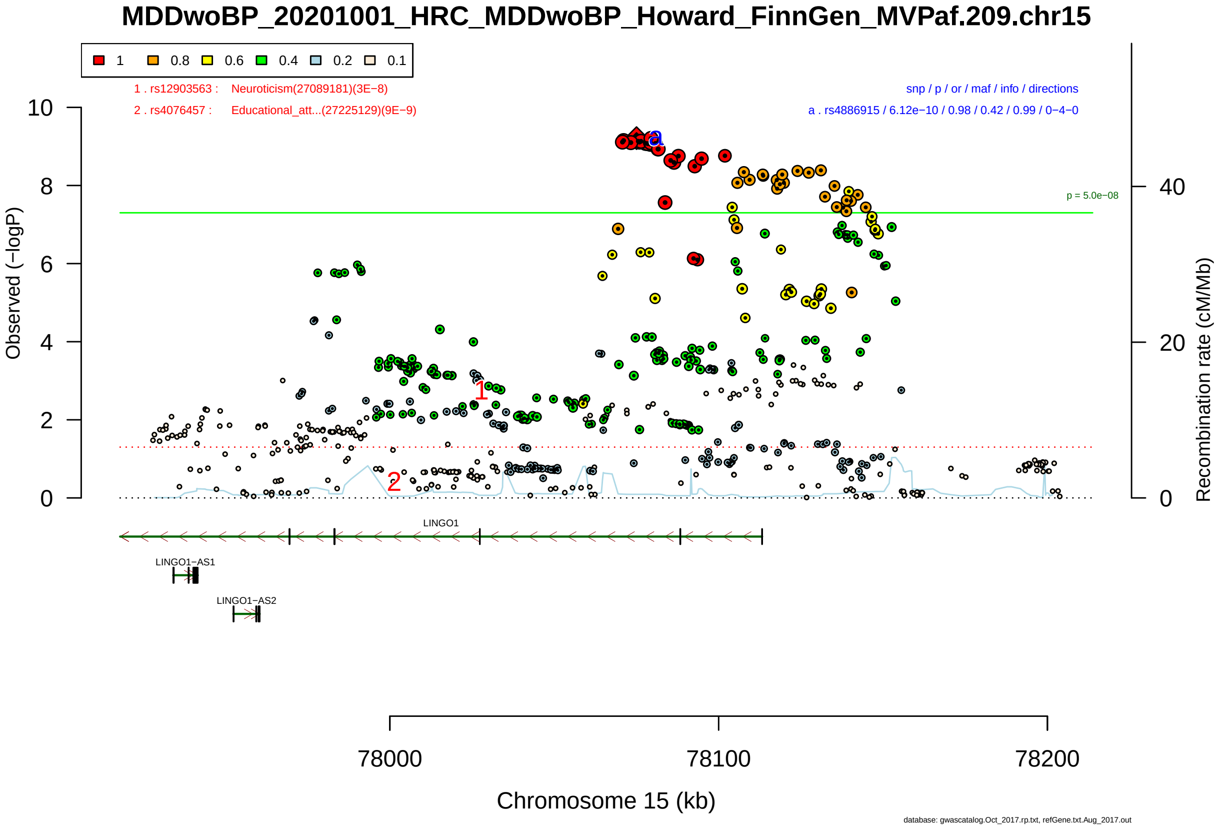

Figure S2-113: Regional plot of GWAS locus No. 113 , with rs4886915 as lead SNP.

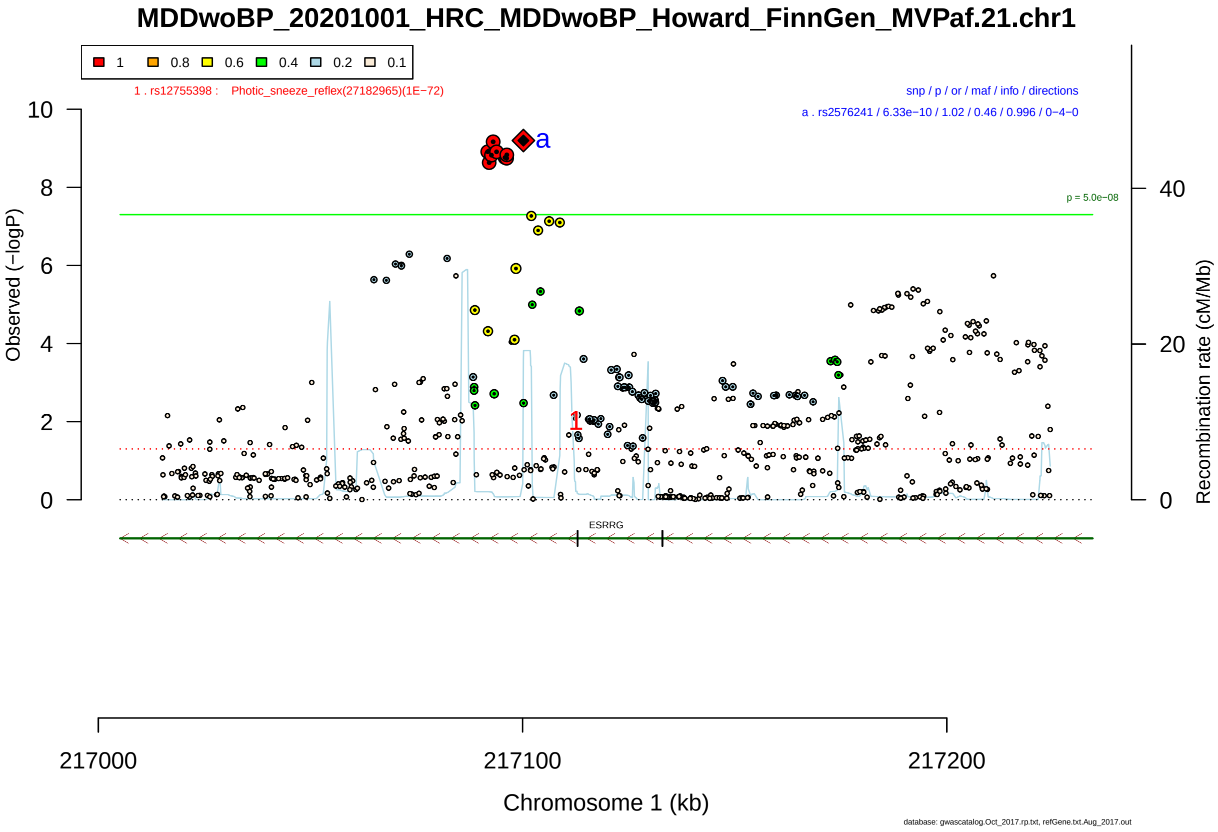

Figure S2-114: Regional plot of GWAS locus No. 114 , with rs2576241 as lead SNP.

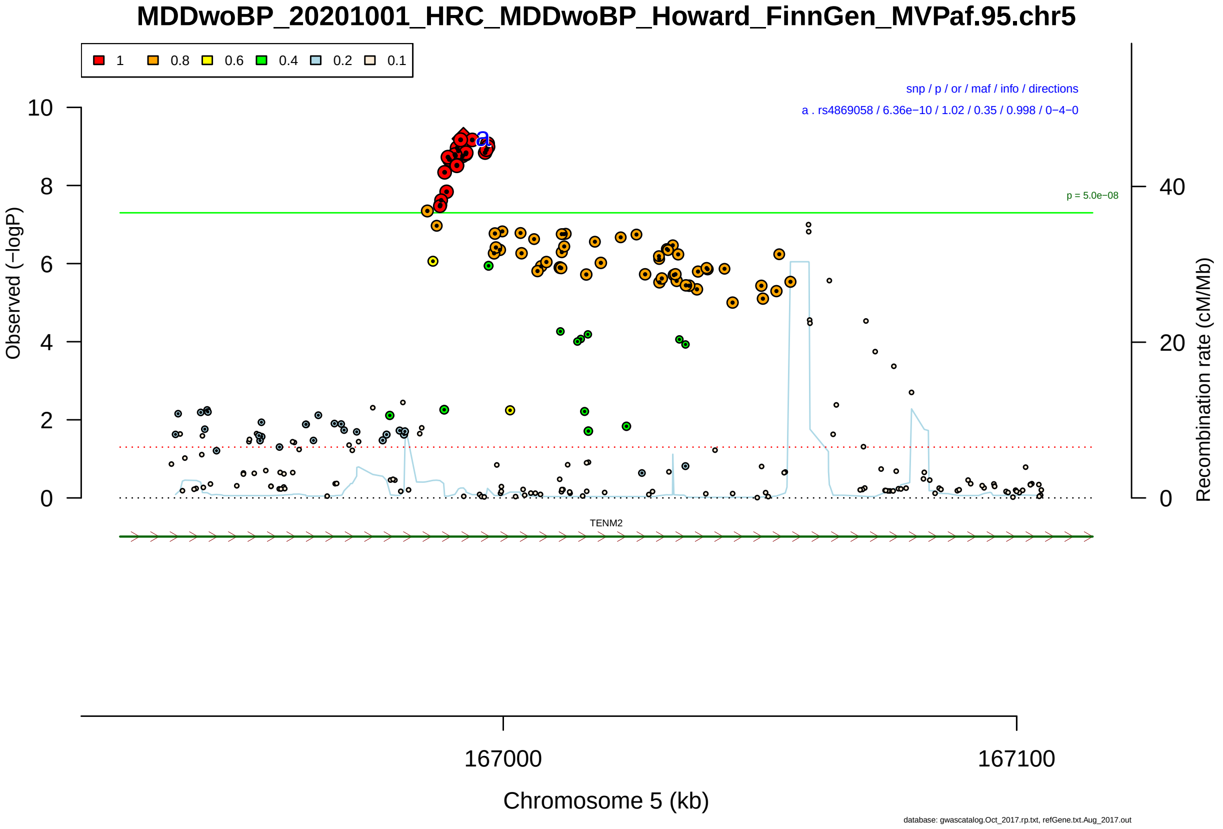

Figure S2-115: Regional plot of GWAS locus No. 115 , with rs4869058 as lead SNP.

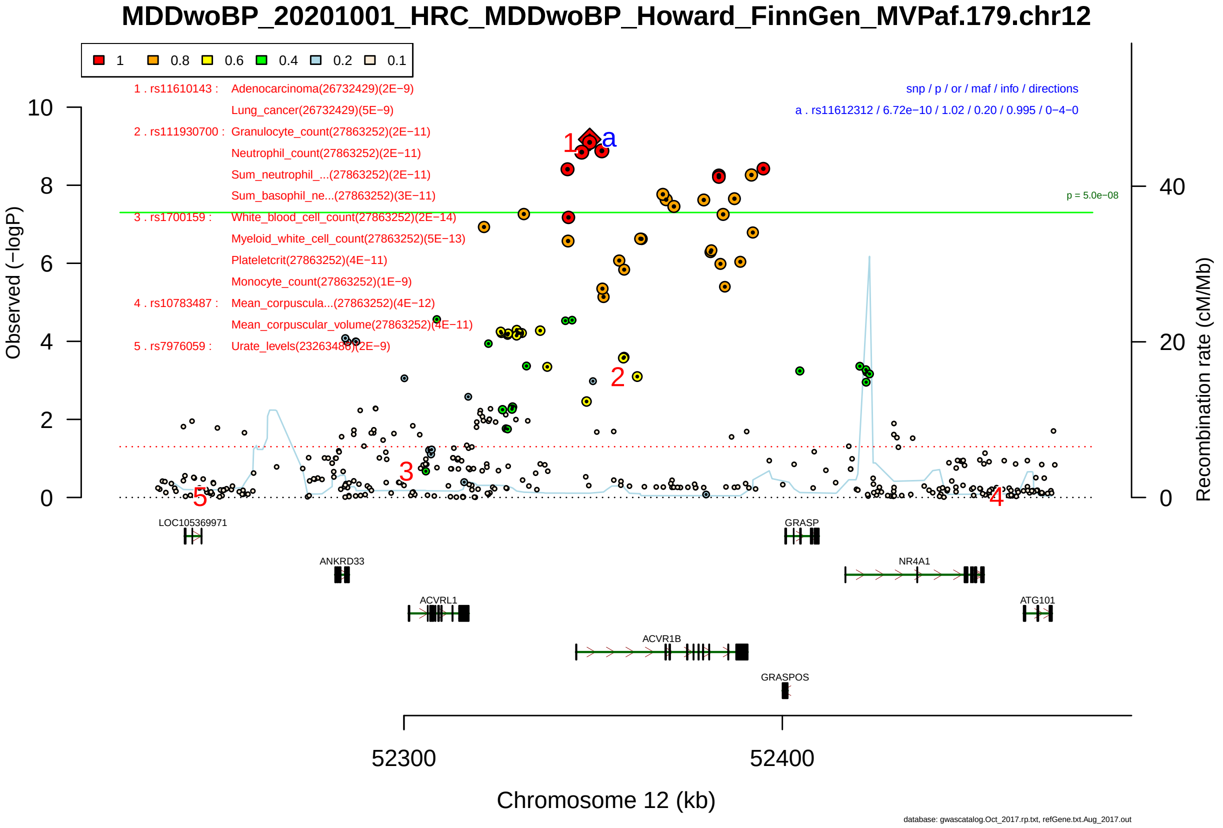

Figure S2-116: Regional plot of GWAS locus No. 116 , with rs11612312 as lead SNP.

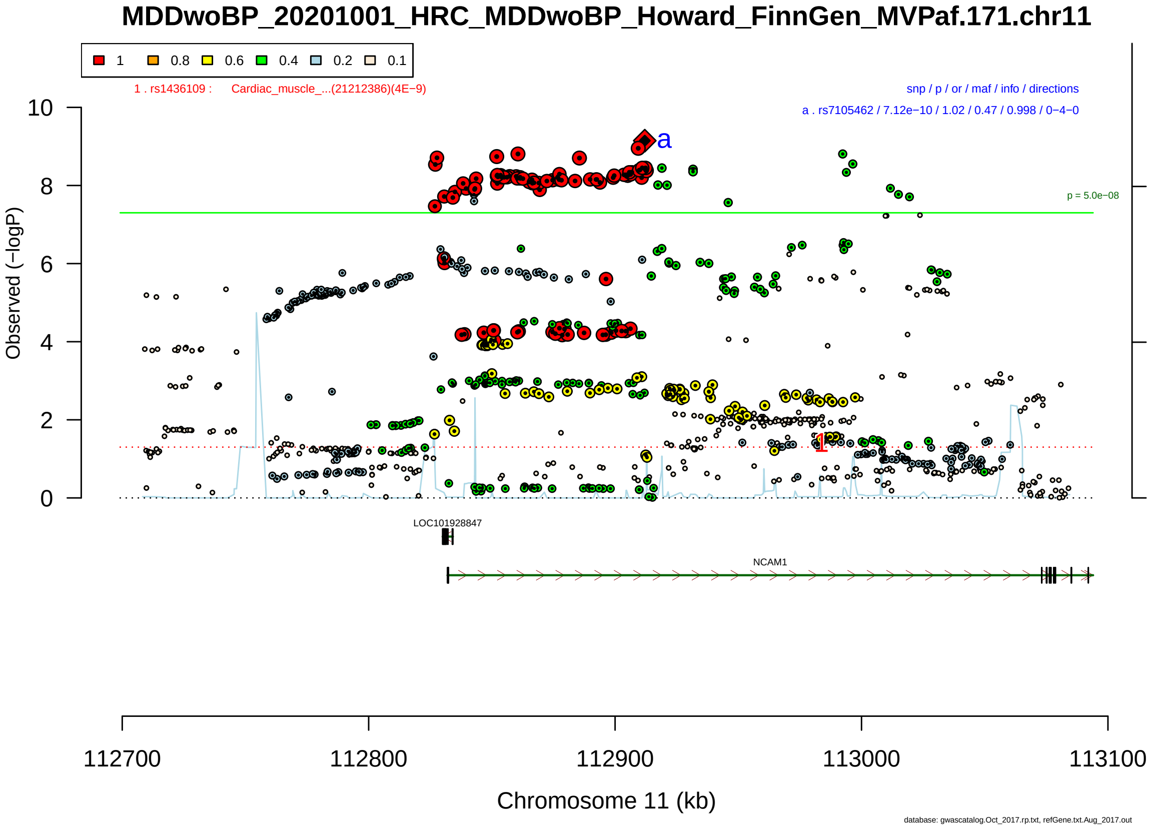

Figure S2-117: Regional plot of GWAS locus No. 117 , with rs967516 as lead SNP.

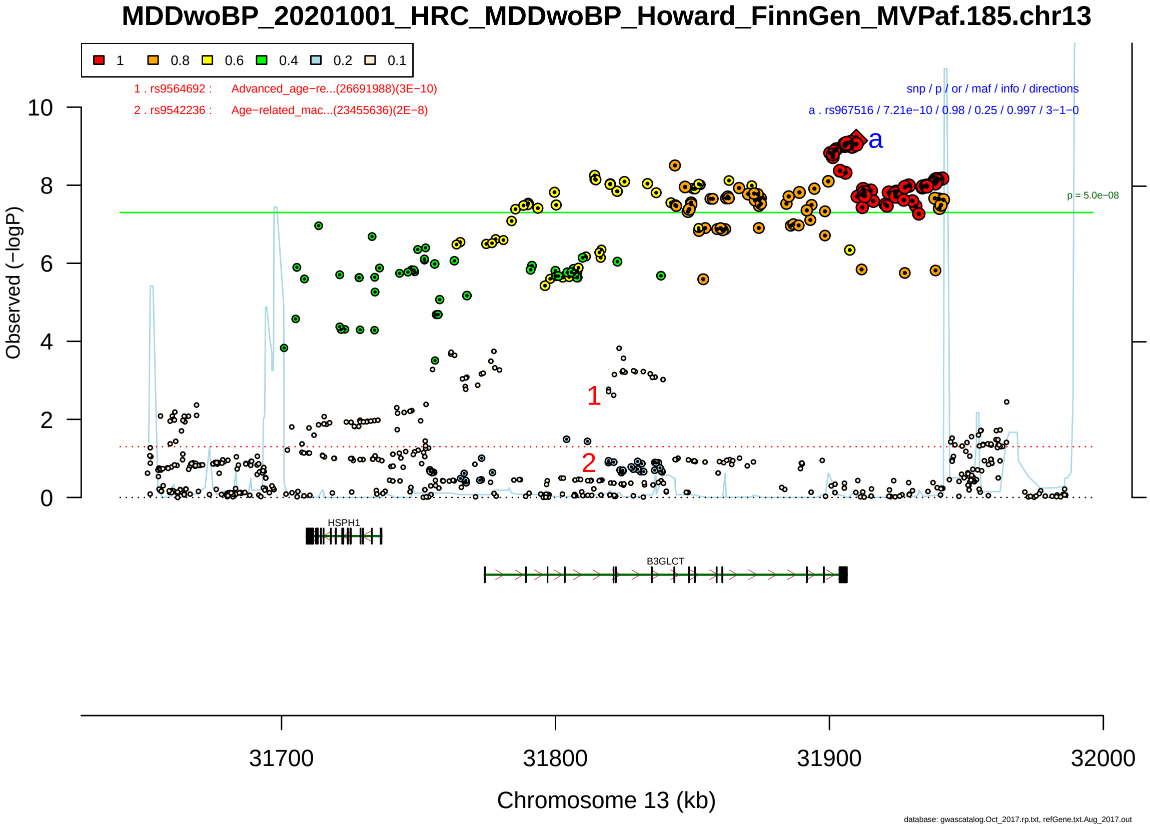

Figure S2-118: Regional plot of GWAS locus No. 118 , with rs7968921 as lead SNP.

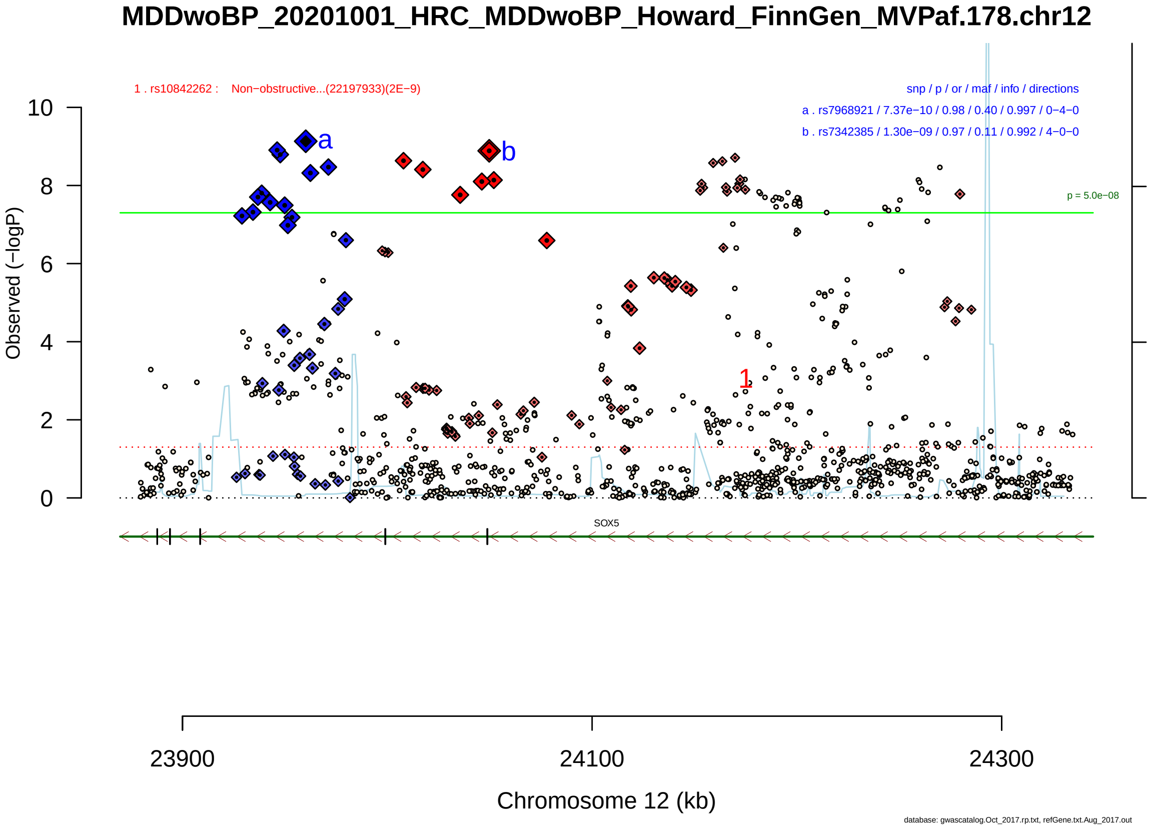

Figure S2-119: Regional plot of GWAS locus No. 119 , with rs2964003 as lead SNP.

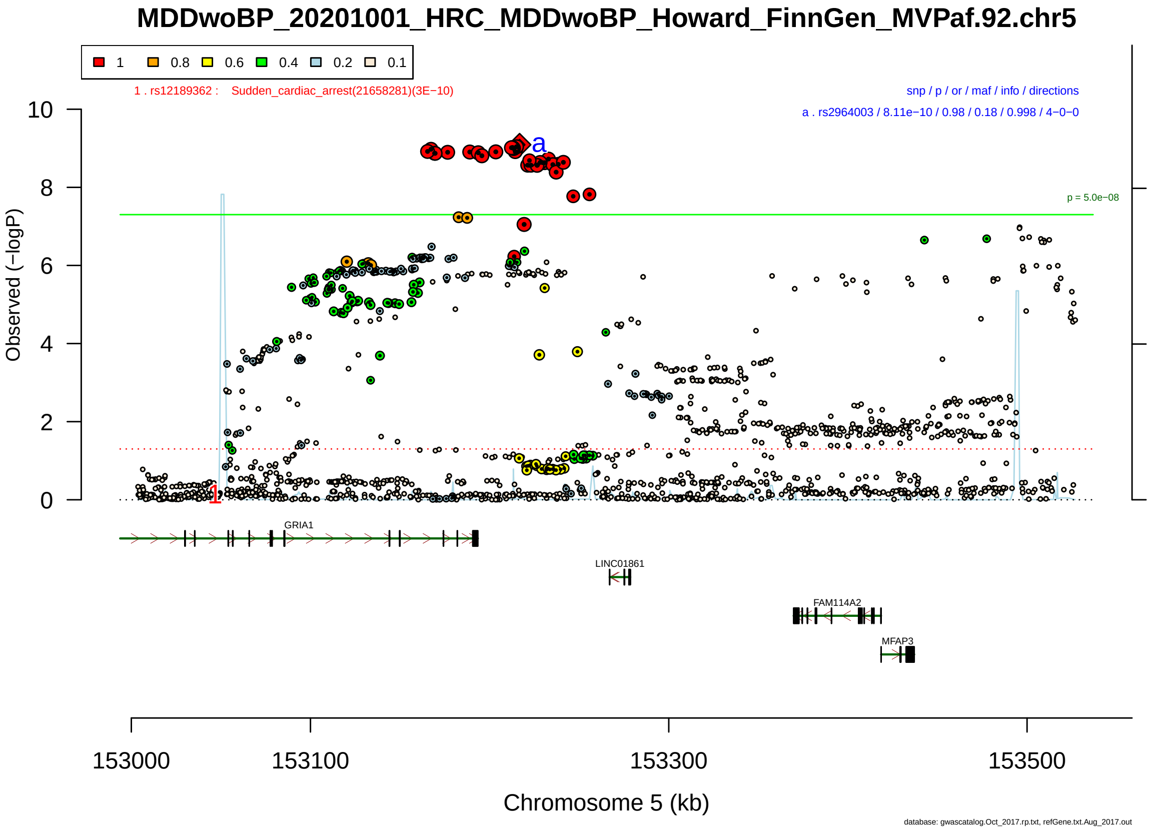

Figure S2-120: Regional plot of GWAS locus No. 120 , with rs1825119 as lead SNP.

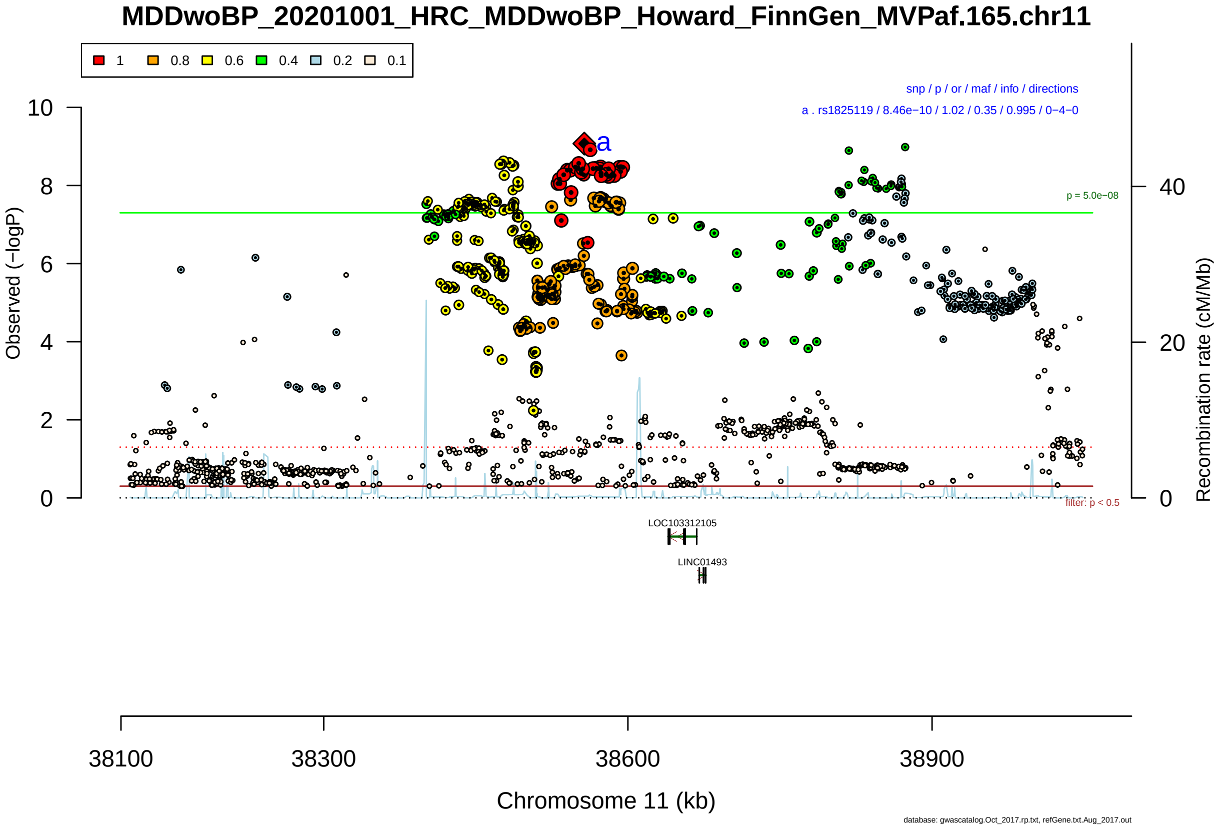

Figure S2-121: Regional plot of GWAS locus No. 121 , with rs2060337 as lead SNP.

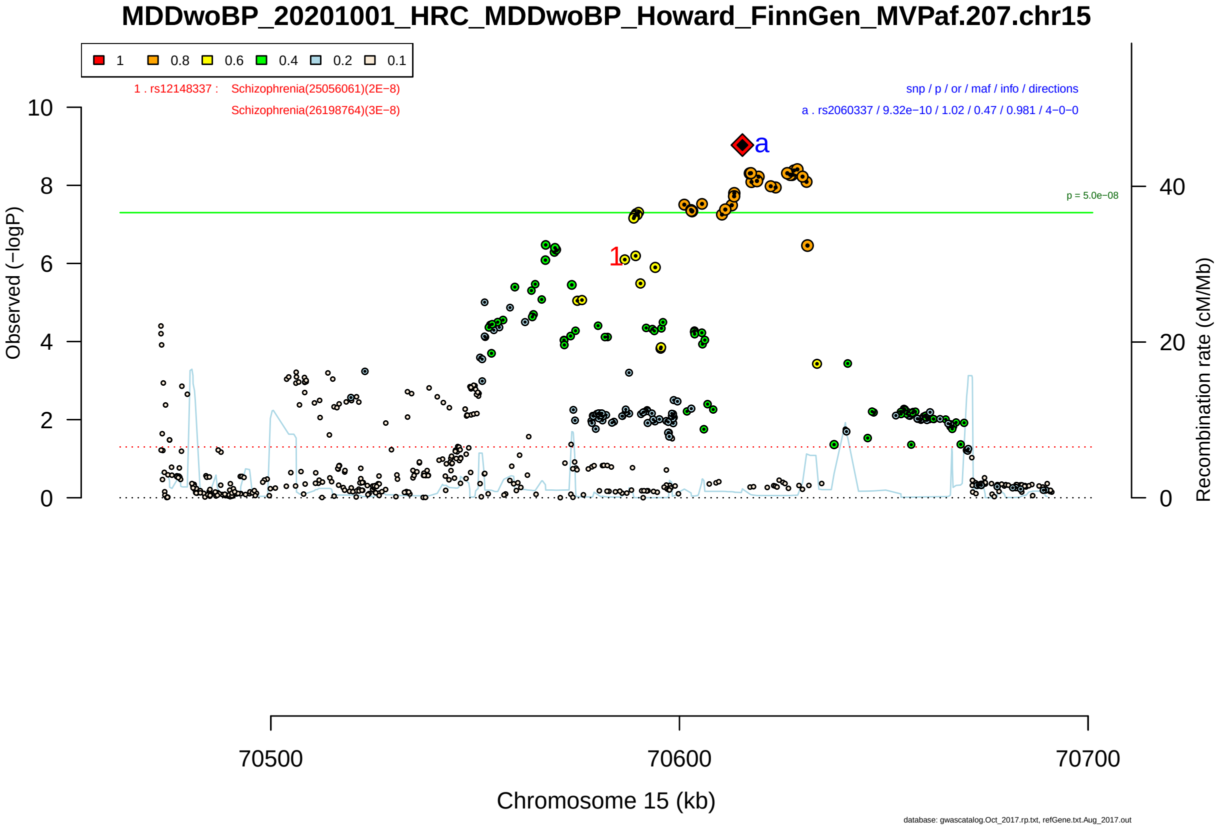

Figure S2-122: Regional plot of GWAS locus No. 122 , with rs1801251 as lead SNP.

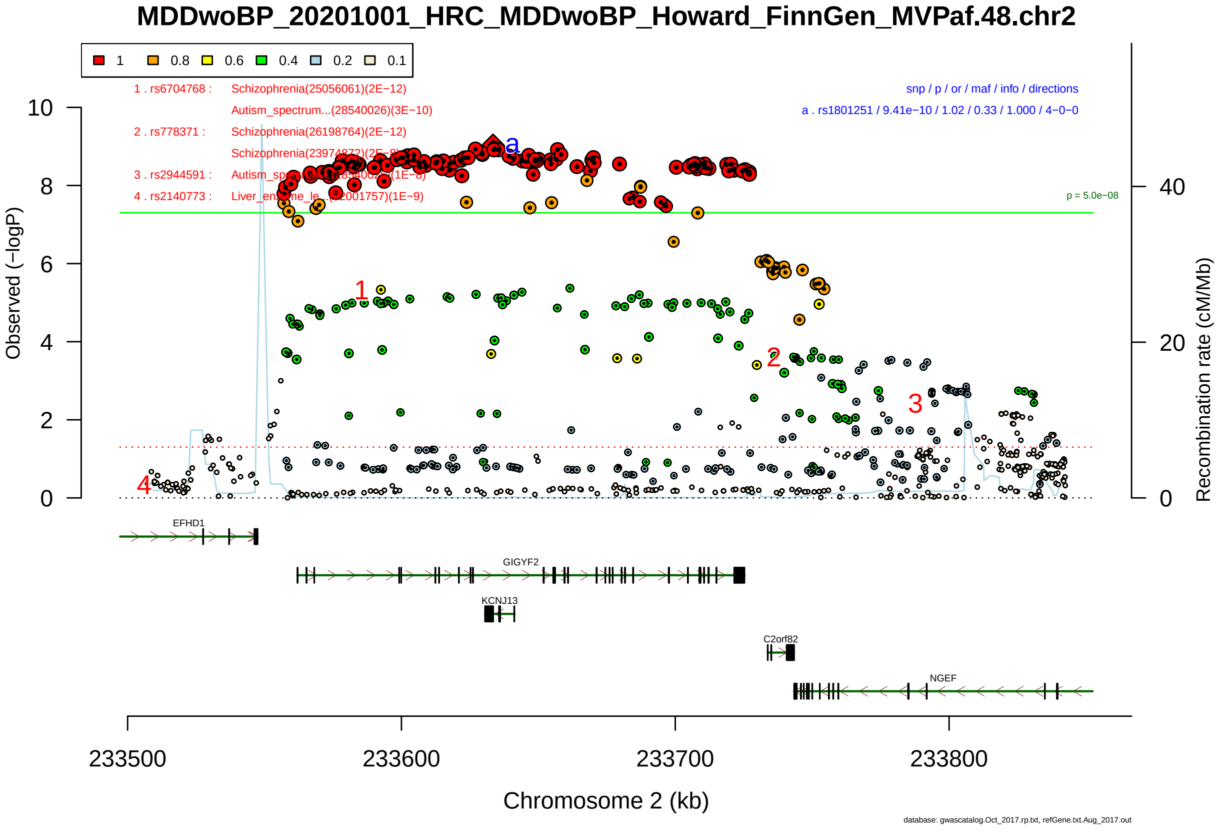

Figure S2-123: Regional plot of GWAS locus No. 123 , with rs4785307 as lead SNP.

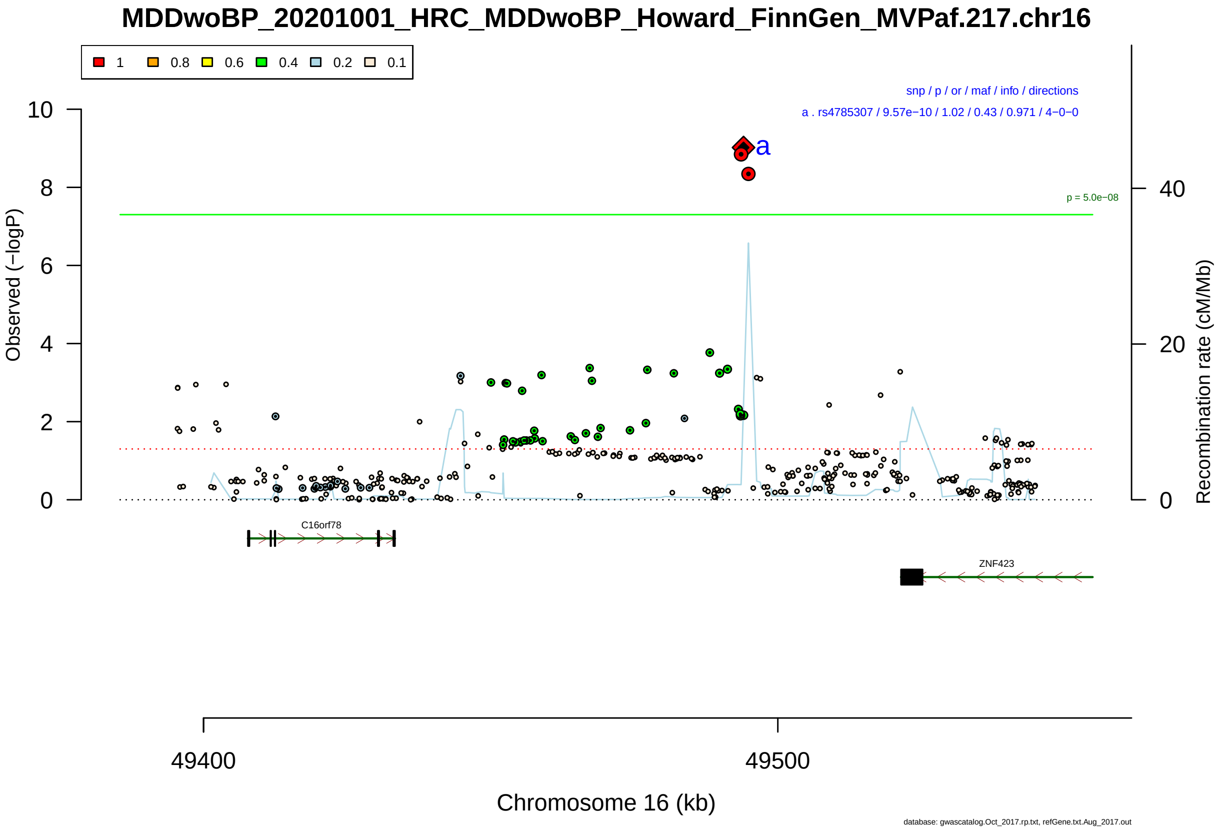

Figure S2-124: Regional plot of GWAS locus No. 124 , with rs80067584 as lead SNP.

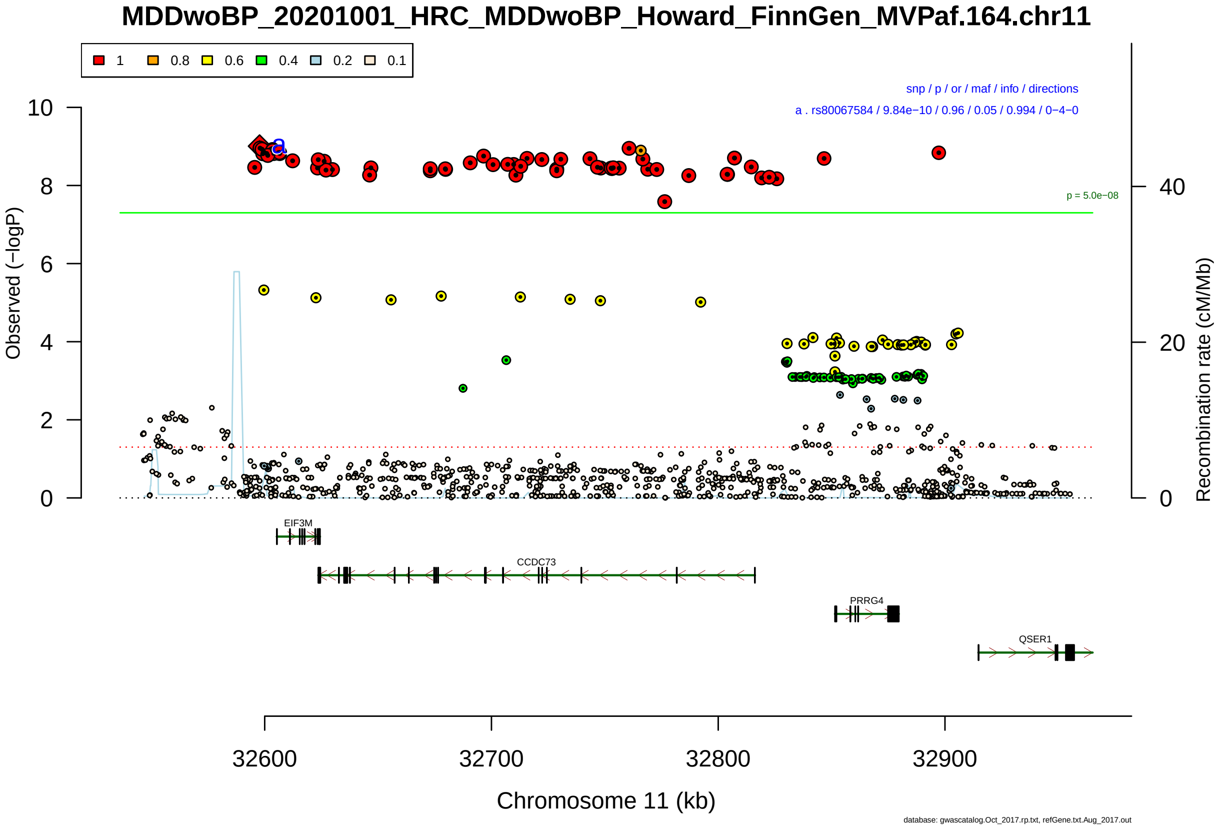

Figure S2-125: Regional plot of GWAS locus No. 125 , with rs4846898 as lead SNP.

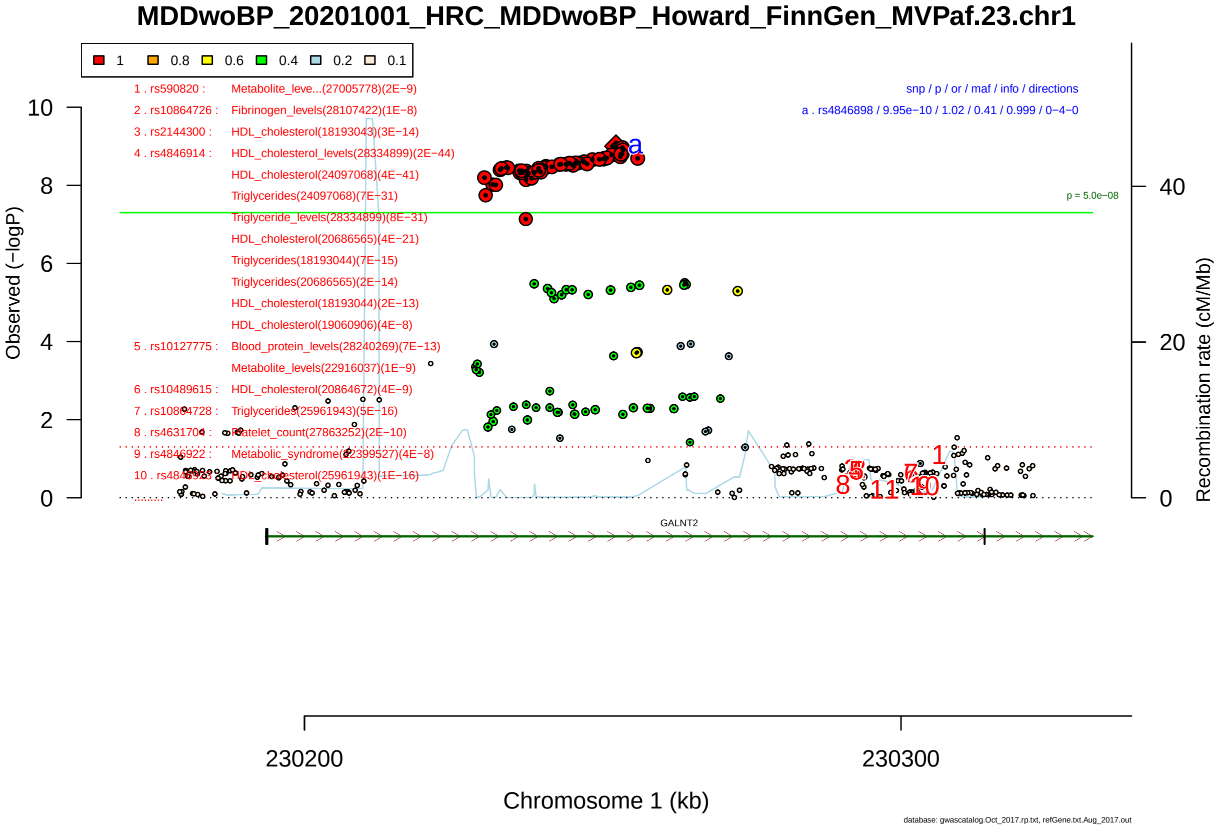

Figure S2-126: Regional plot of GWAS locus No. 126 , with rs11130821 as lead SNP.

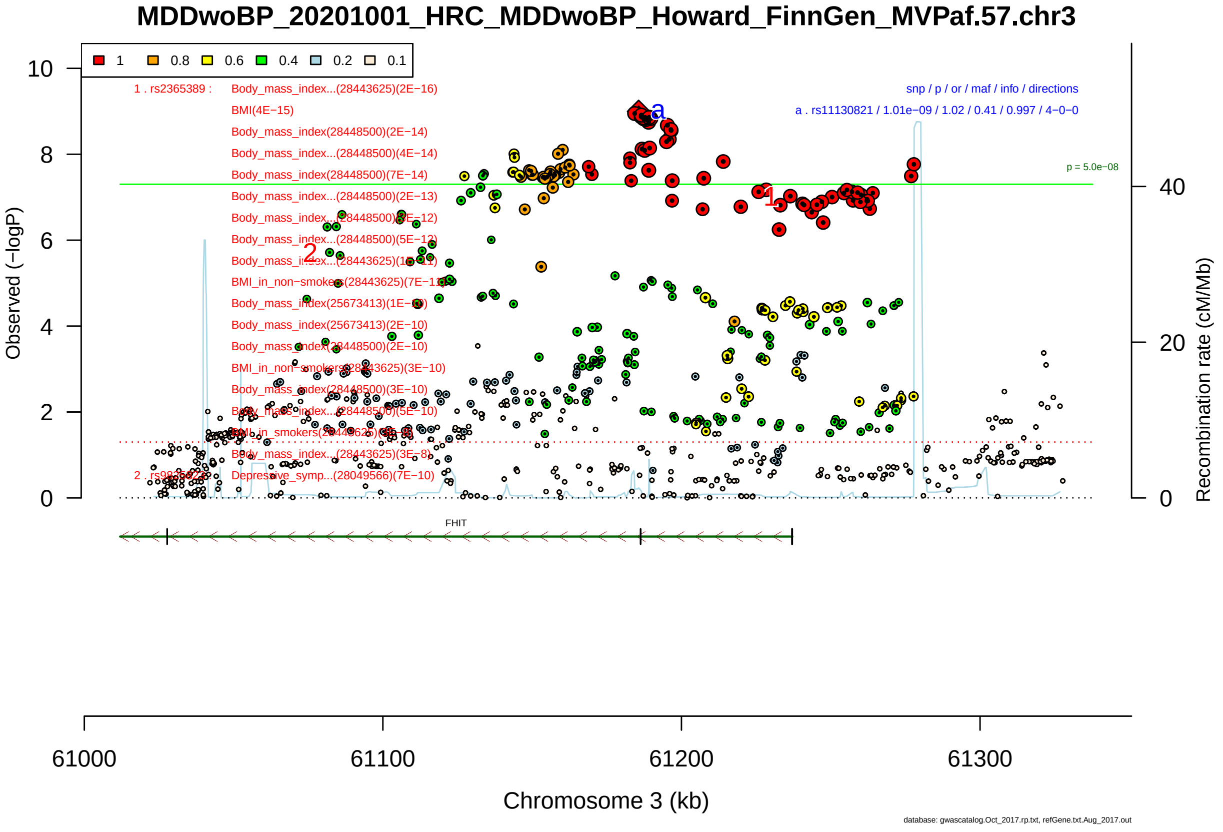

Figure S2-127: Regional plot of GWAS locus No. 127 , with rs112181005 as lead SNP.

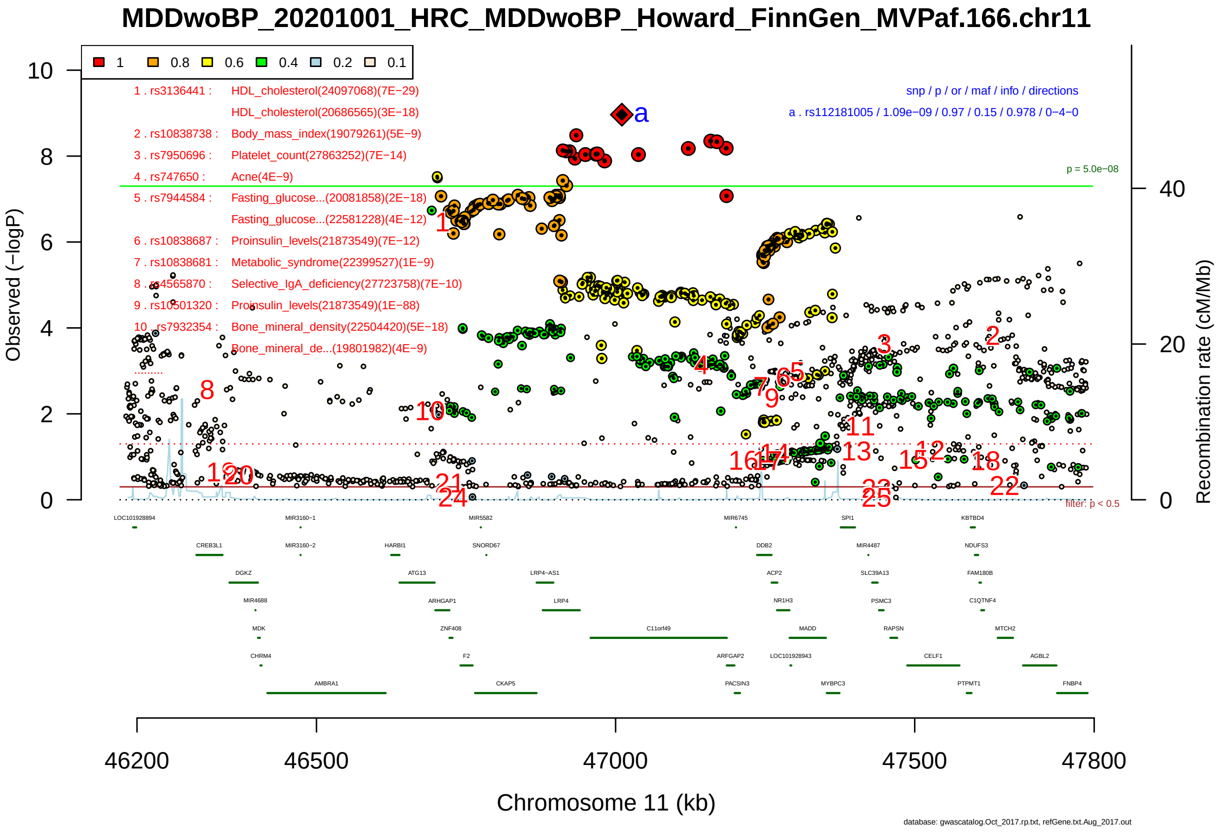

Figure S2-128: Regional plot of GWAS locus No. 128 , with rs10026036 as lead SNP.

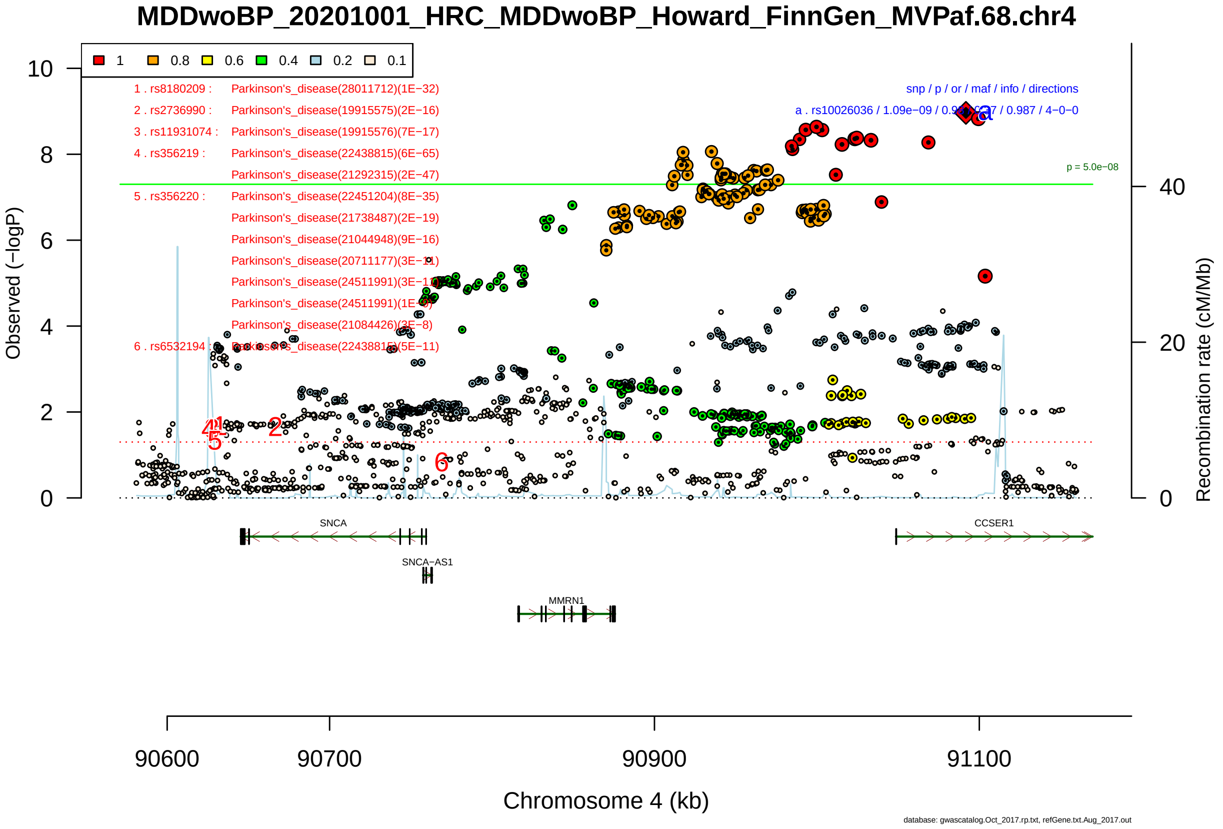

Figure S2-129: Regional plot of GWAS locus No. 129 , with rs56059718 as lead SNP.

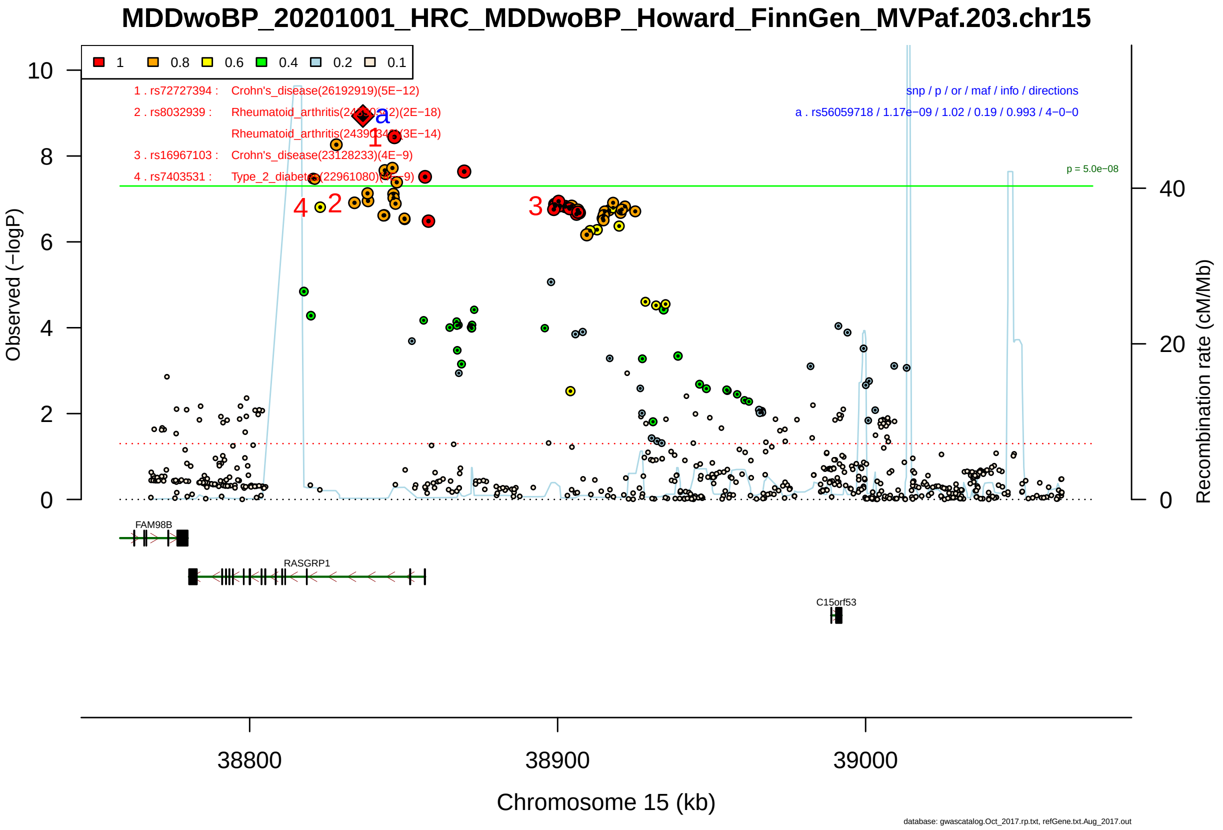

Figure S2-130: Regional plot of GWAS locus No. 130 , with rs3756335 as lead SNP.

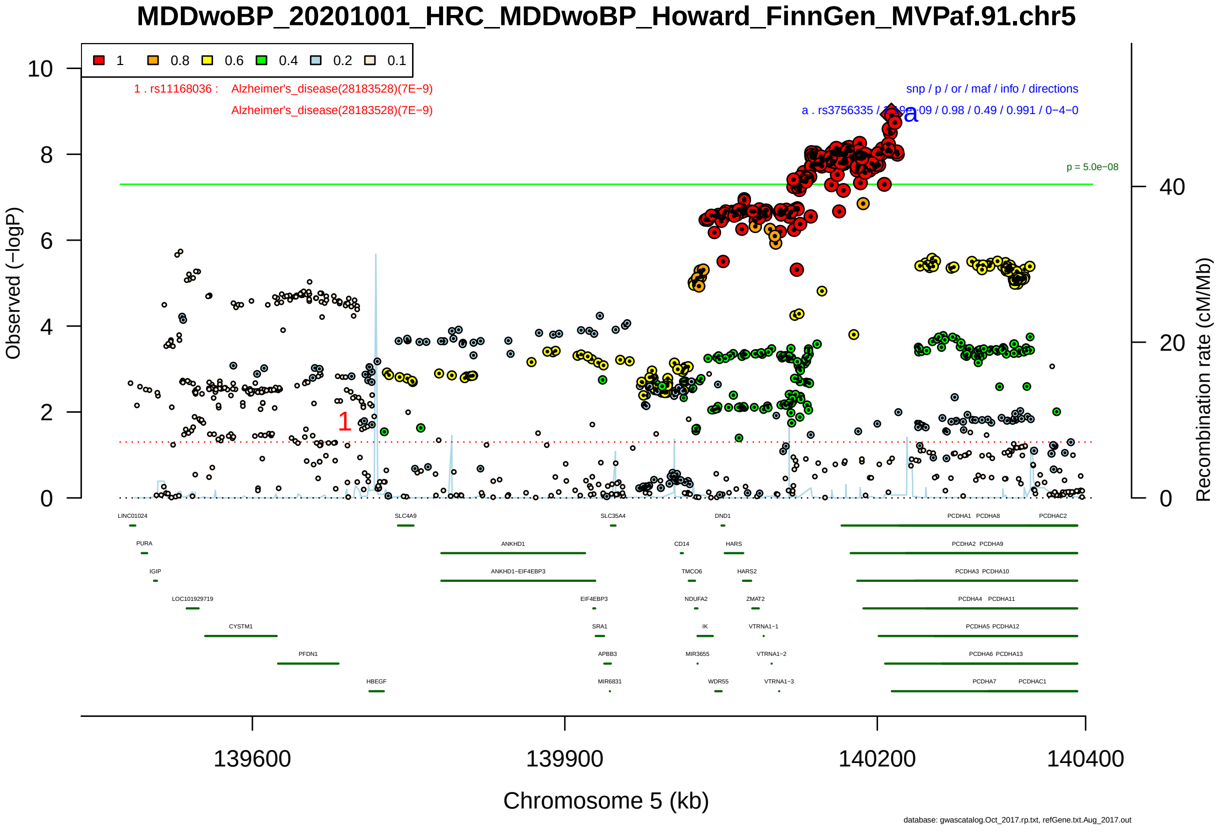

Figure S2-131: Regional plot of GWAS locus No. 131 , with rs7317645 as lead SNP.

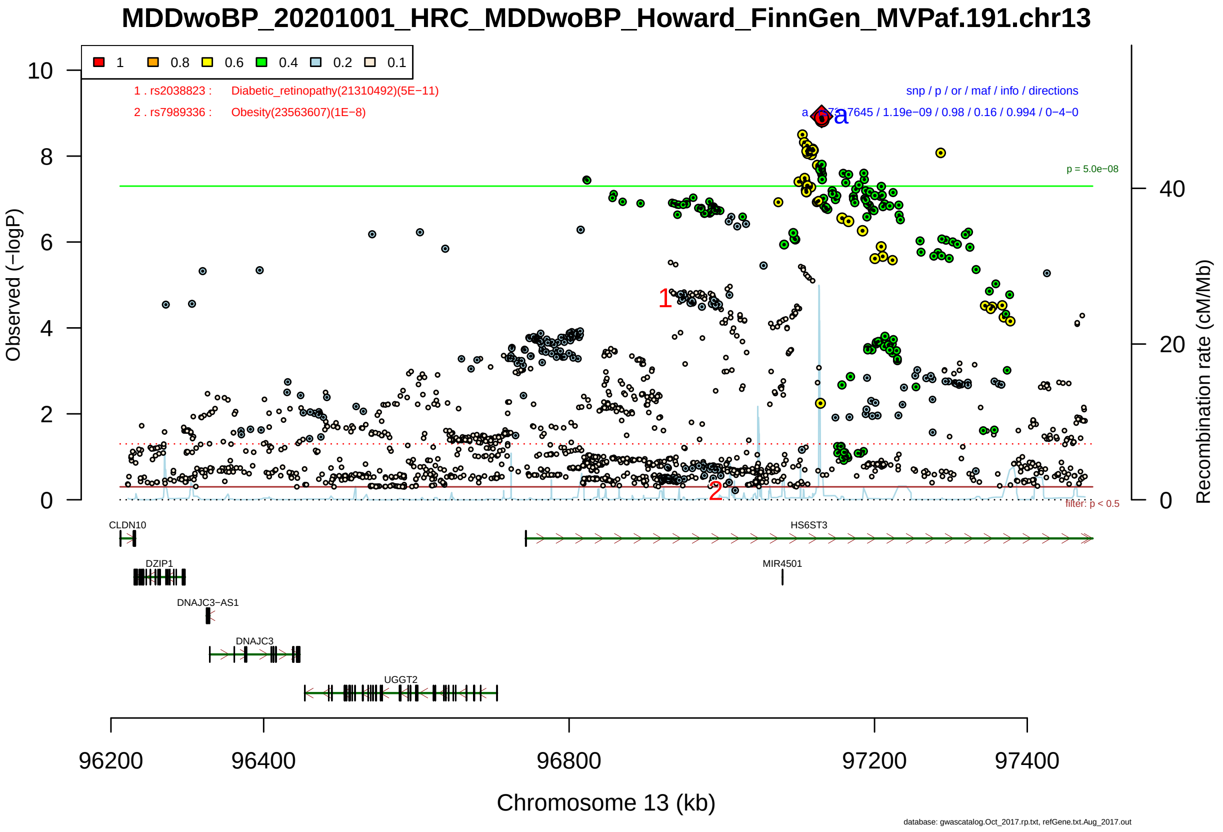

Figure S2-132: Regional plot of GWAS locus No. 132 , with rs6536630 as lead SNP.

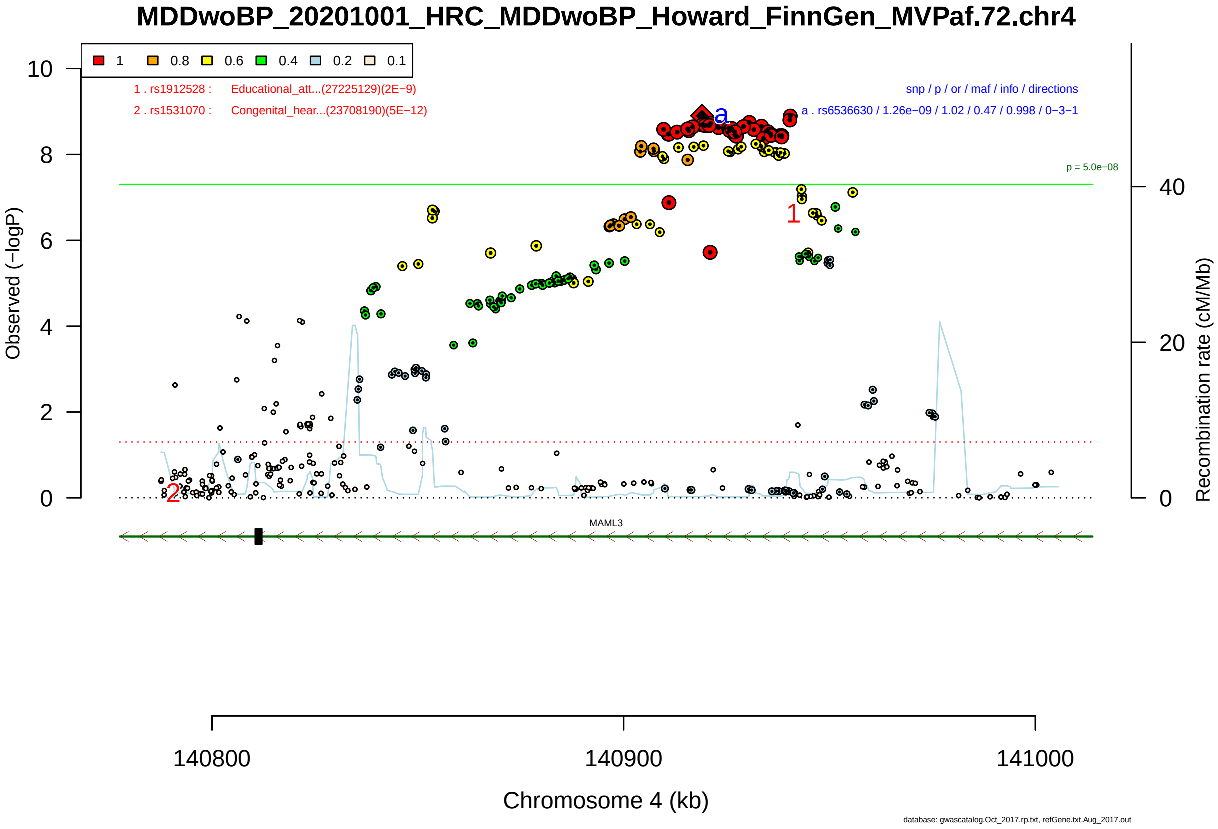

Figure S2-133: Regional plot of GWAS locus No. 133 , with rs4489042 as lead SNP.

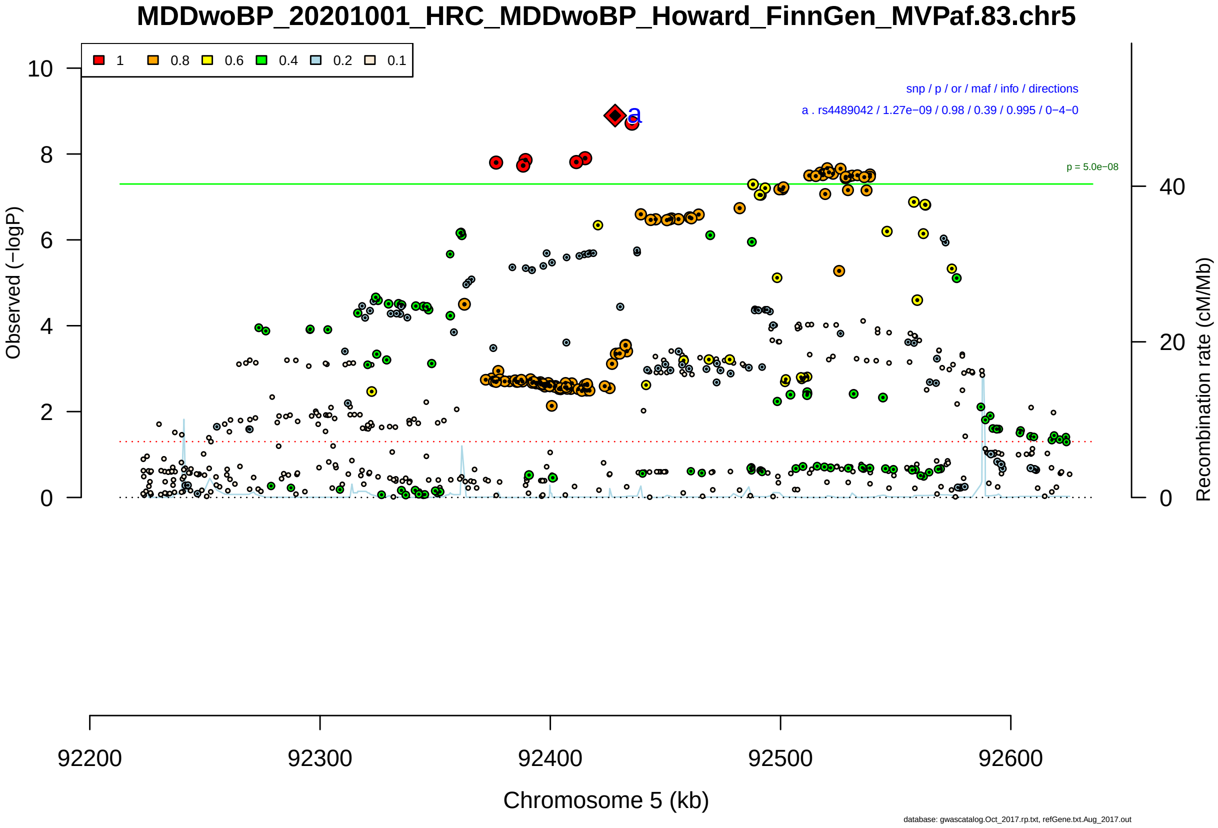

Figure S2-134: Regional plot of GWAS locus No. 134 , with rs6715105 as lead SNP.

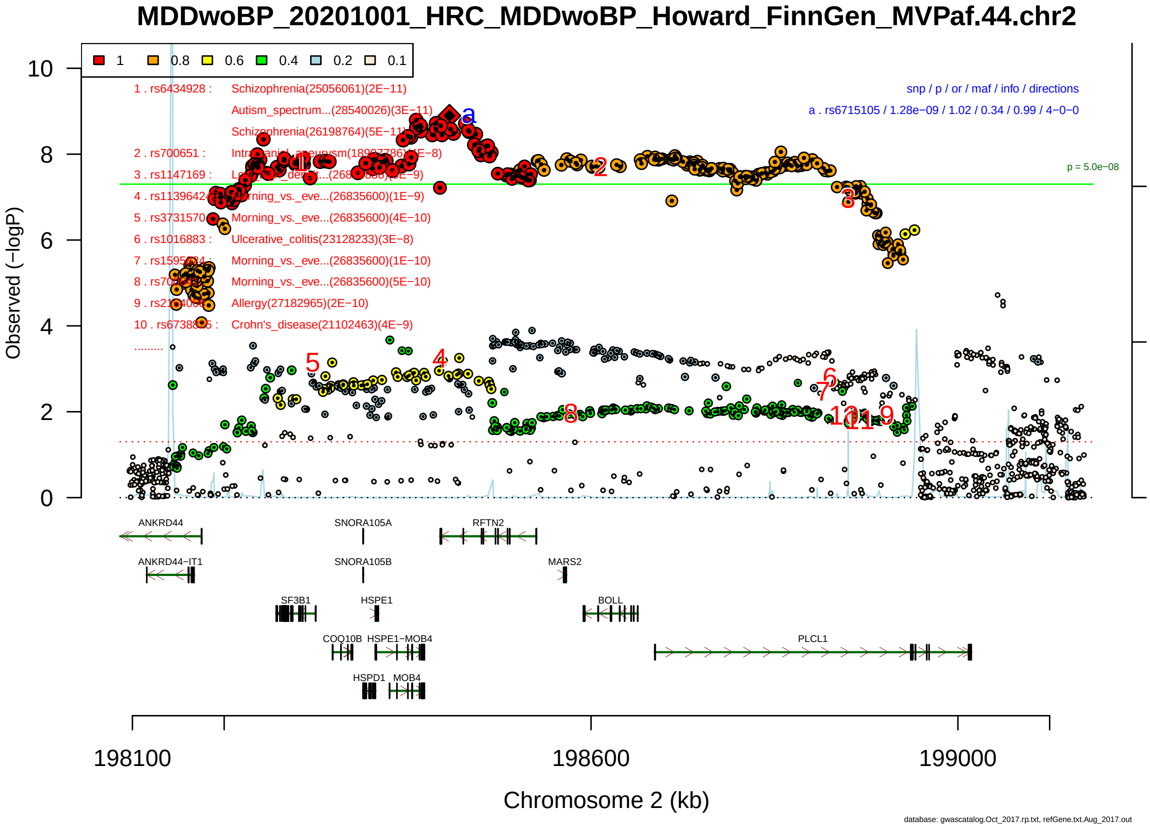

Figure S2-135: Regional plot of GWAS locus No. 135 , with rs913930 as lead SNP.

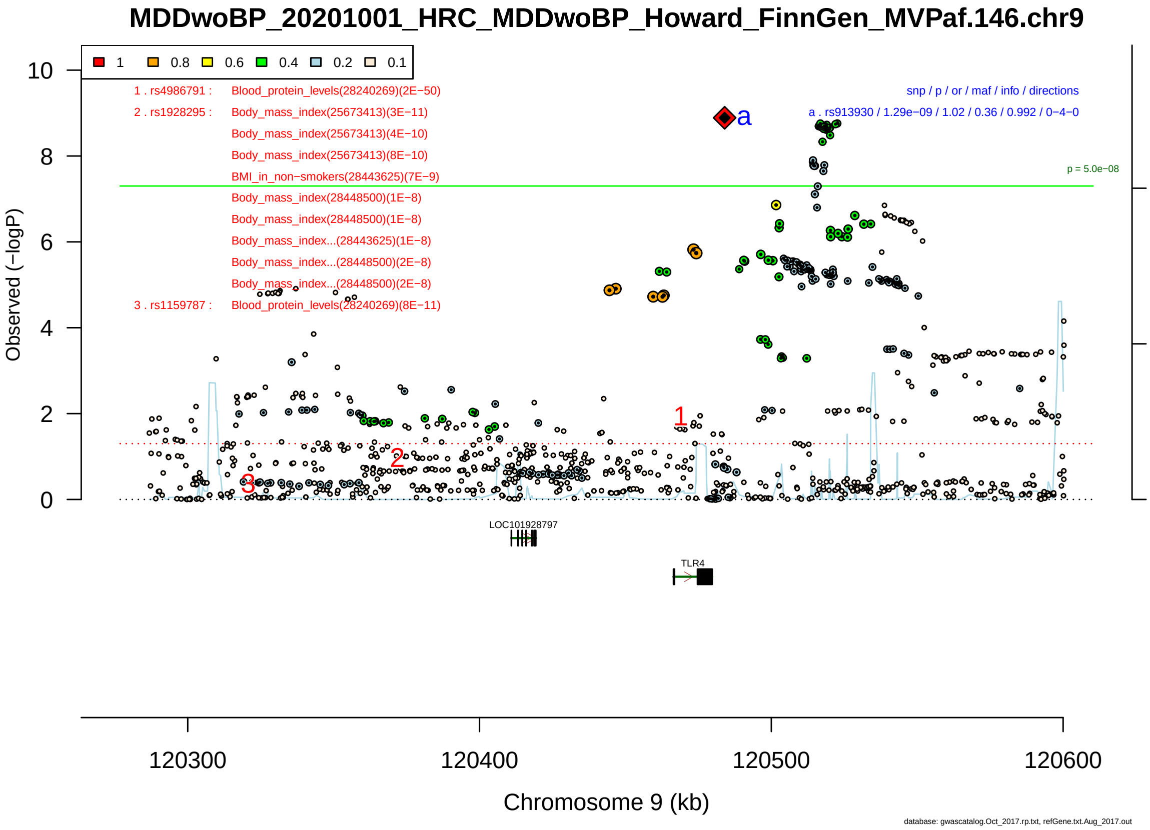

Figure S2-136: Regional plot of GWAS locus No. 136 , with rs957360 as lead SNP.

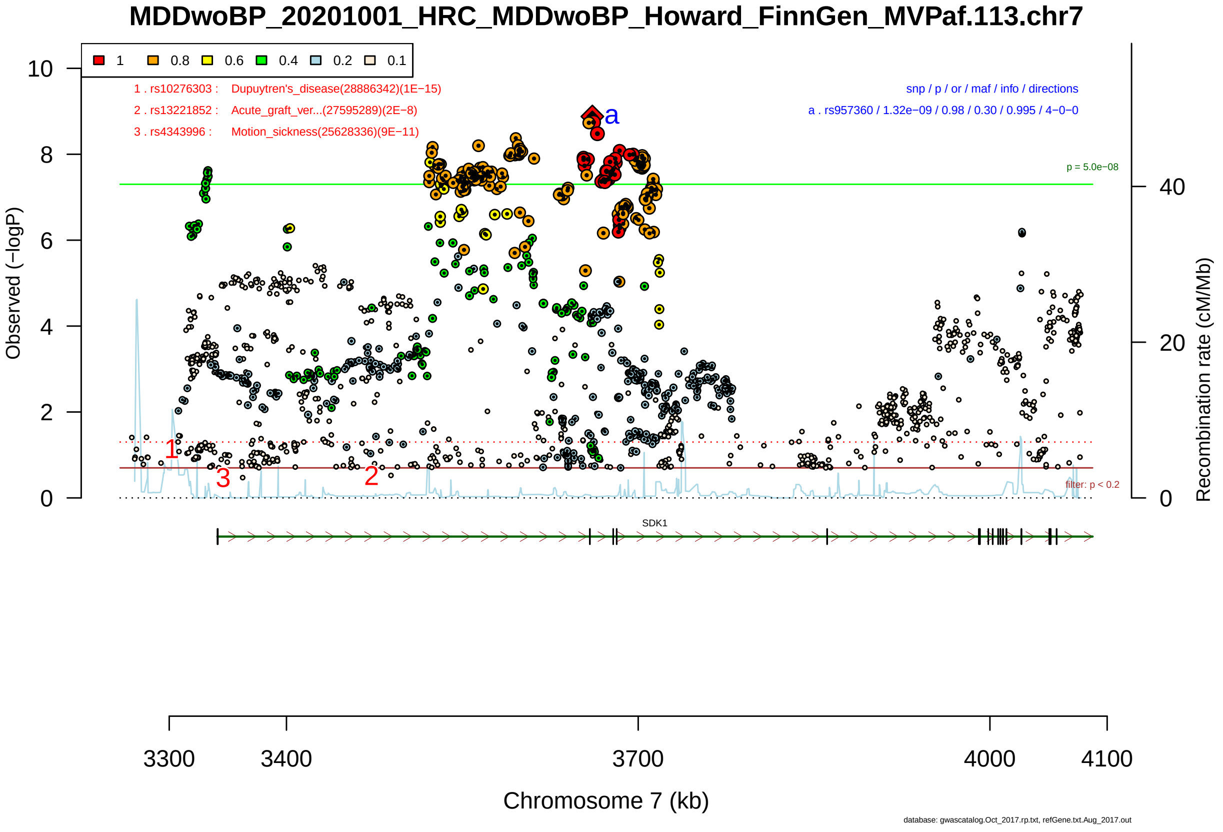

Figure S2-137: Regional plot of GWAS locus No. 137 , with rs182923402 as lead SNP.

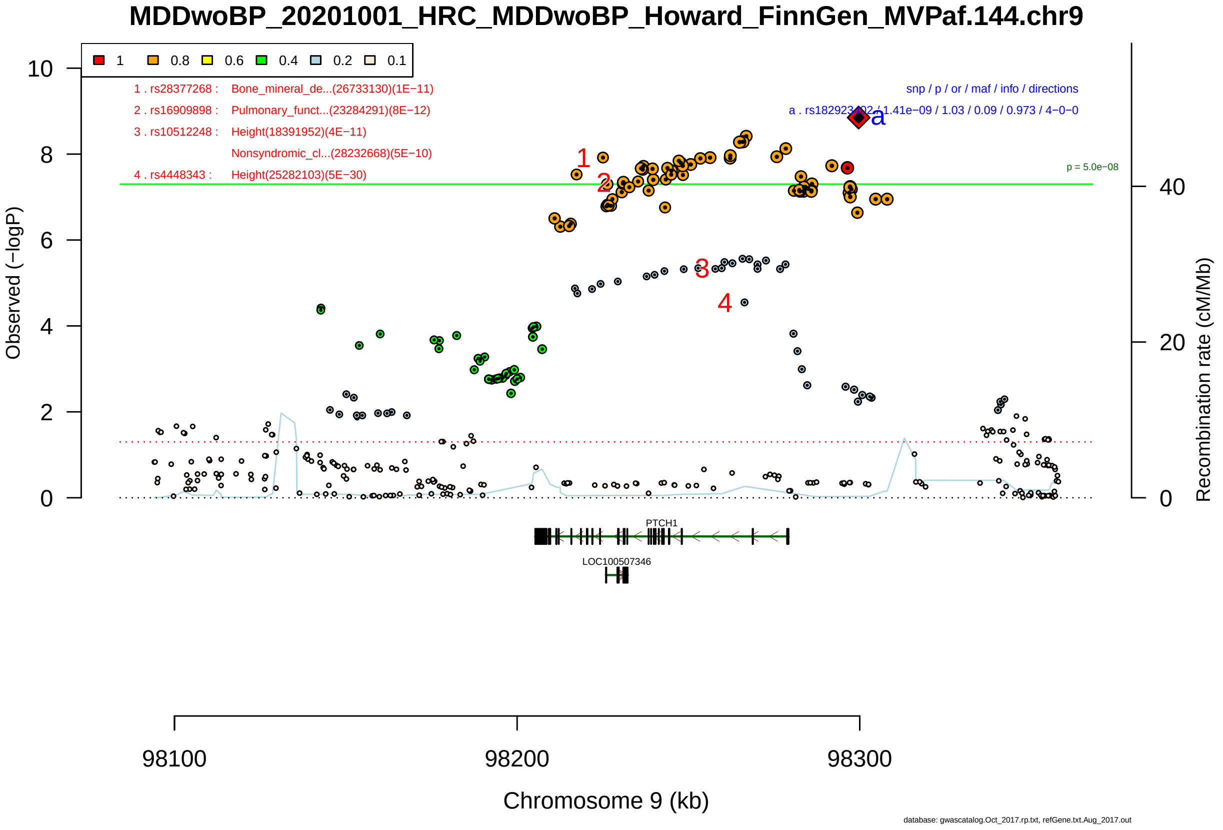

Figure S2-138: Regional plot of GWAS locus No. 138 , with rs62379847 as lead SNP.

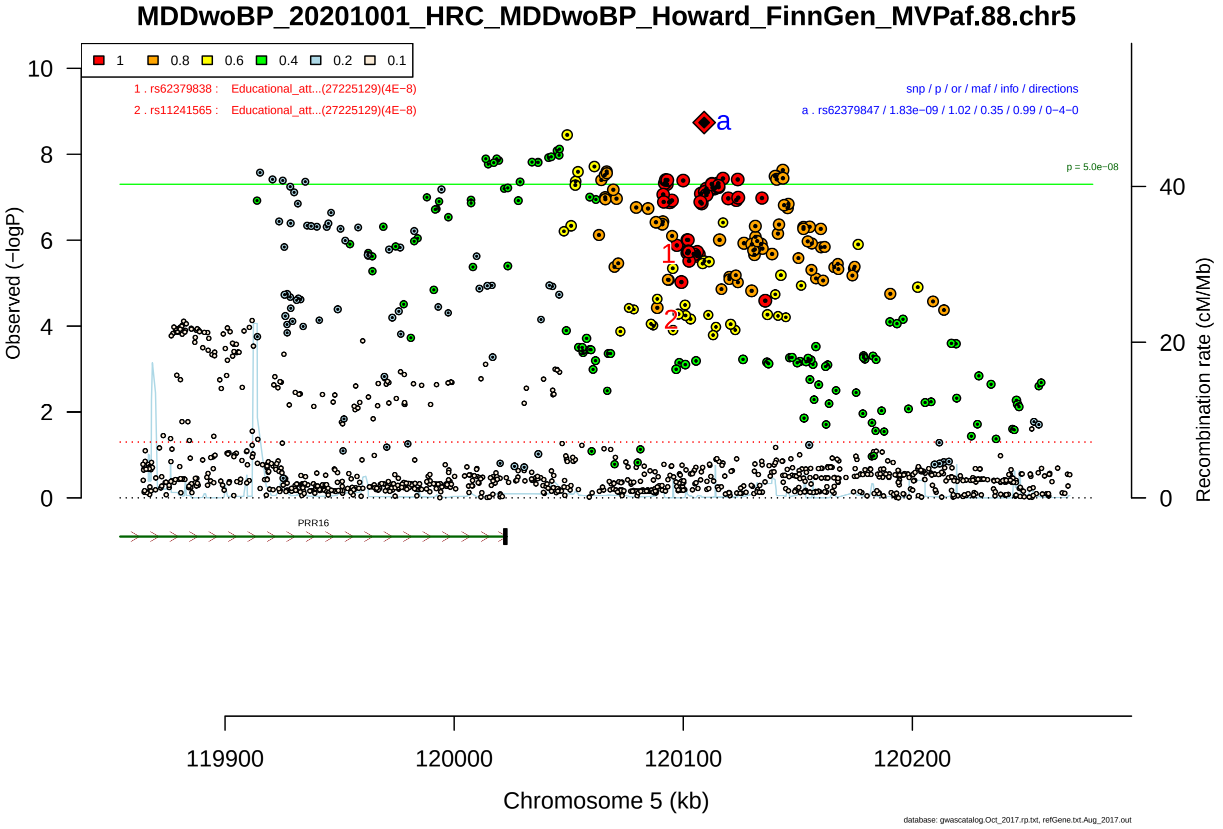

Figure S2-139: Regional plot of GWAS locus No. 139 , with rs7659414 as lead SNP.

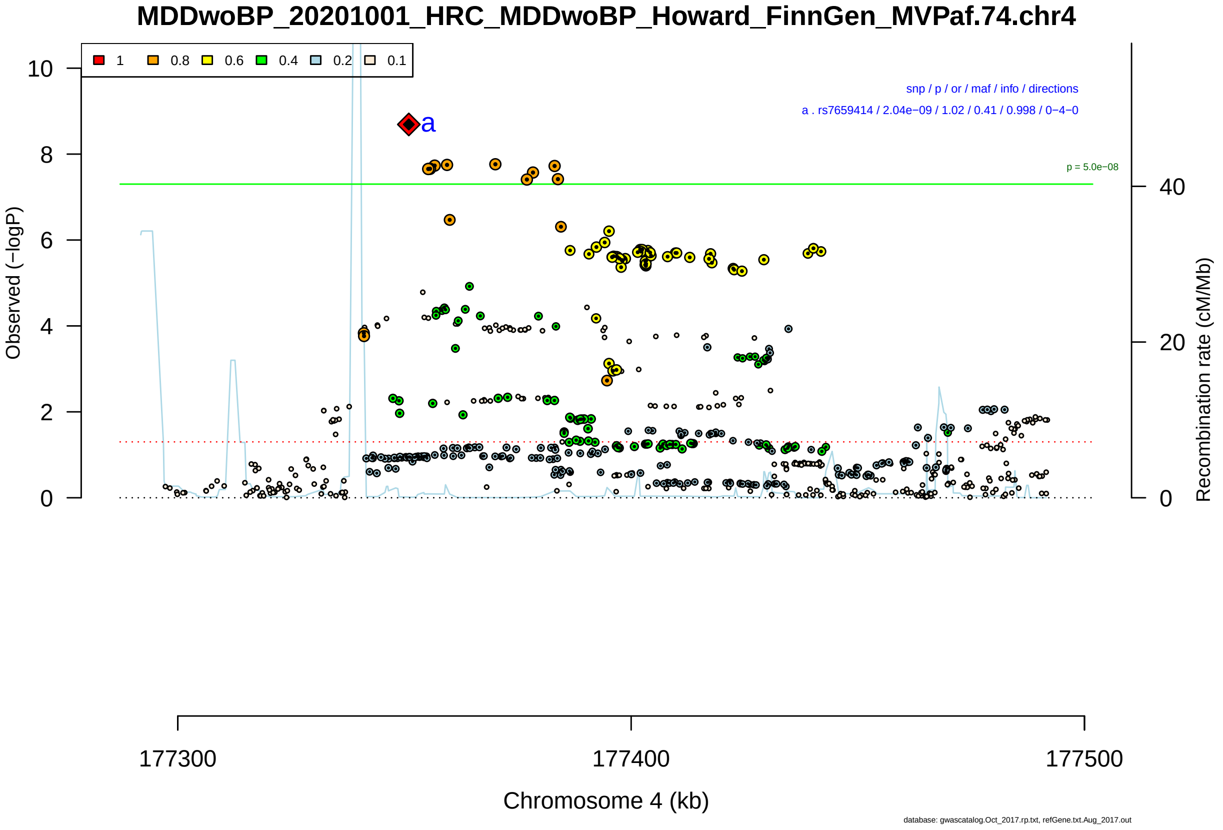

Figure S2-140: Regional plot of GWAS locus No. 140 , with rs3812281 as lead SNP.

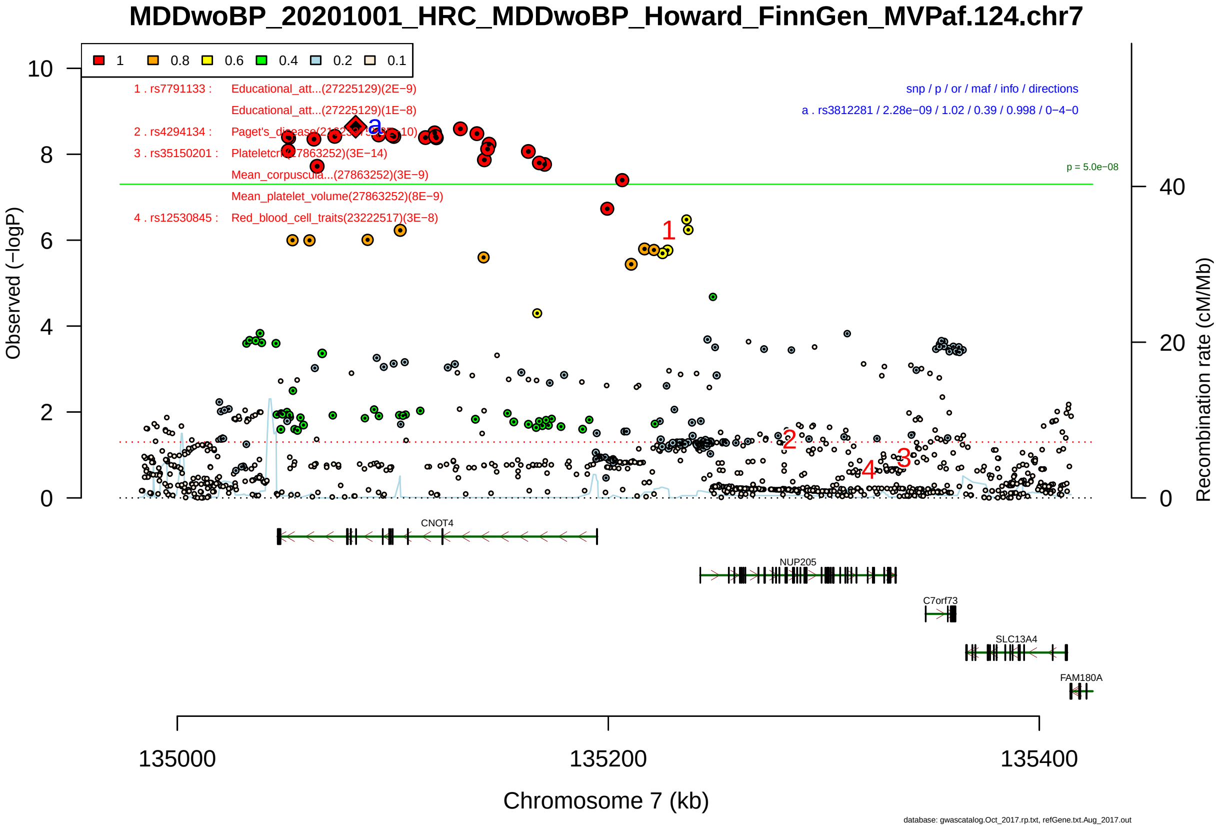

Figure S2-141: Regional plot of GWAS locus No. 141 , with rs4596421 as lead SNP.

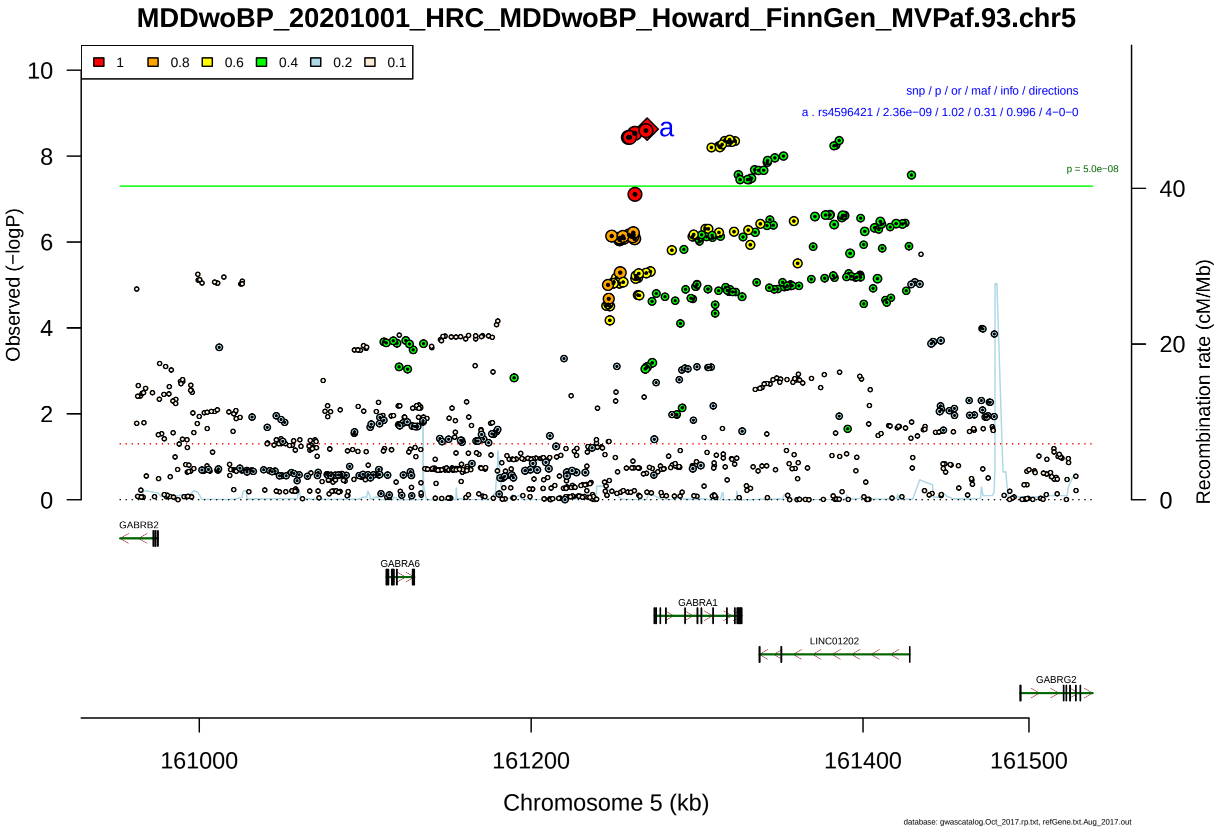

Figure S2-142: Regional plot of GWAS locus No. 142 , with rs35553410 as lead SNP.

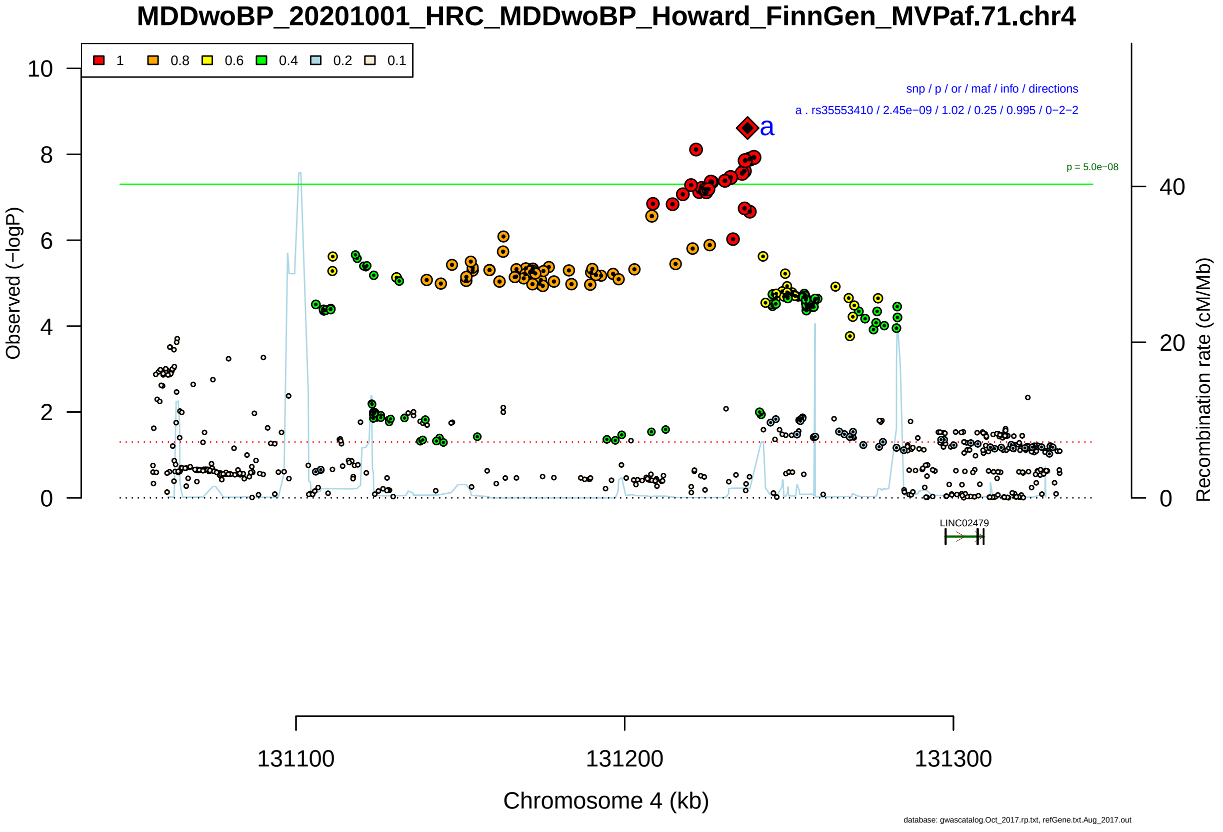

Figure S2-143: Regional plot of GWAS locus No. 143 , with rs56029819 as lead SNP.

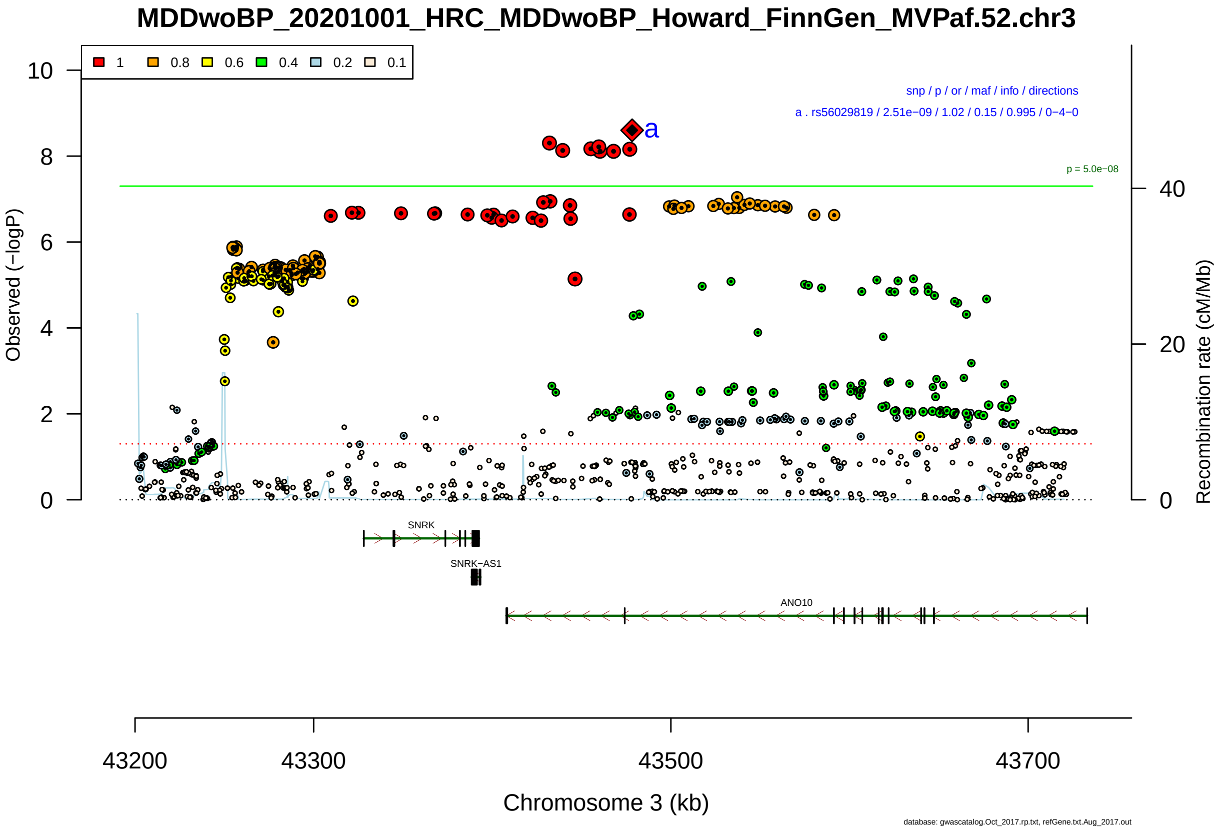

Figure S2-144: Regional plot of GWAS locus No. 144 , with rs7574611 as lead SNP.

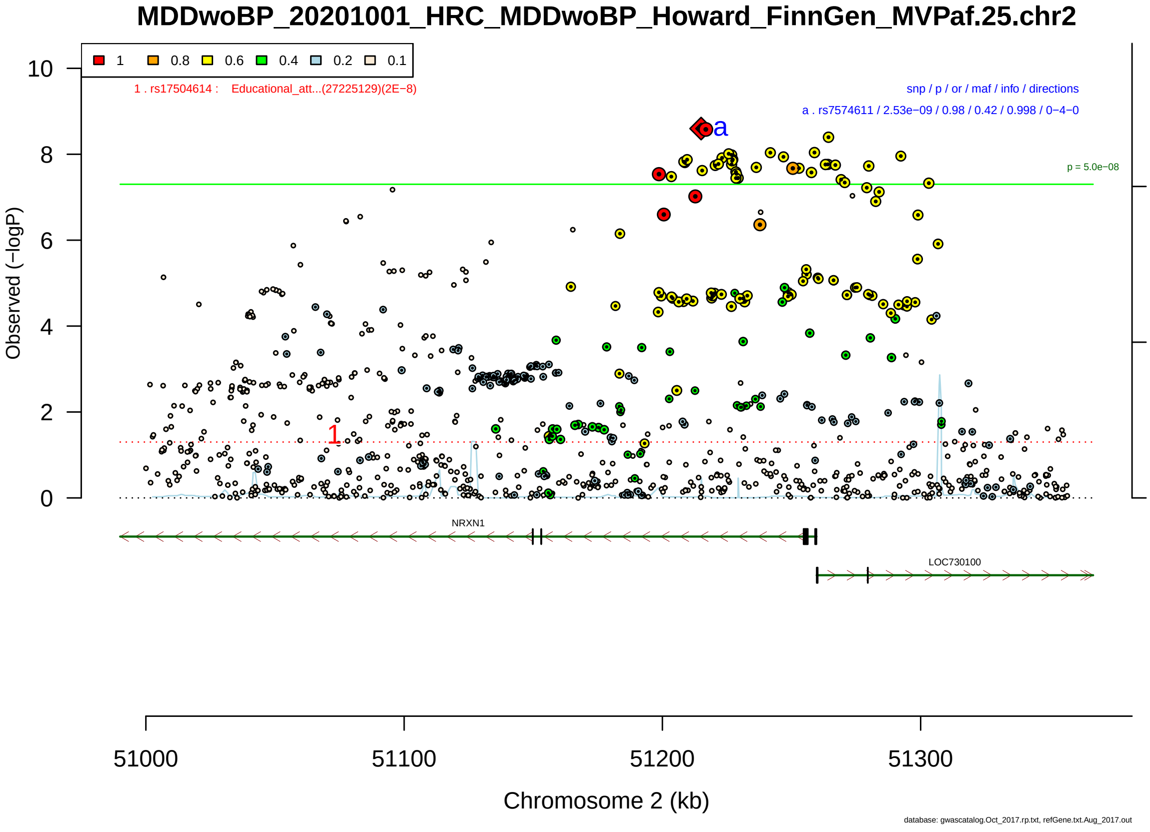

Figure S2-145: Regional plot of GWAS locus No. 145 , with rs10148598 as lead SNP.

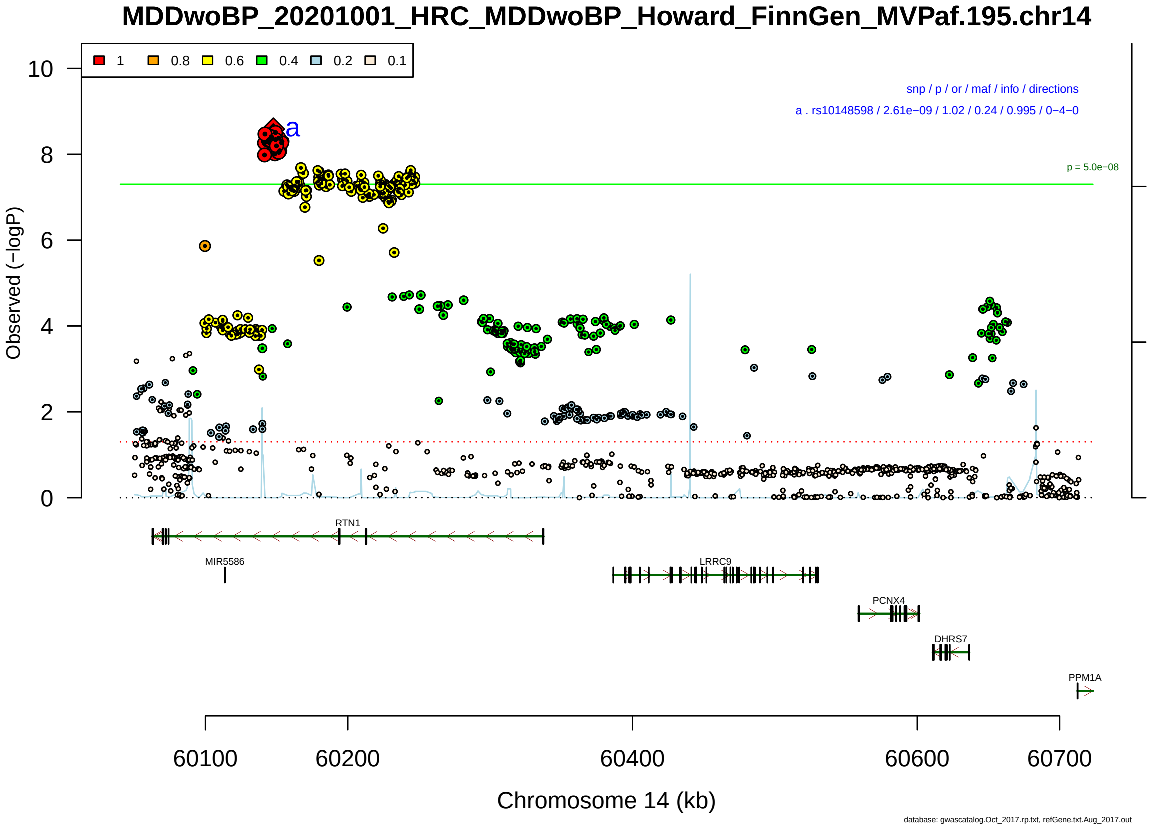

Figure S2-146: Regional plot of GWAS locus No. 146 , with rs1986692 as lead SNP.

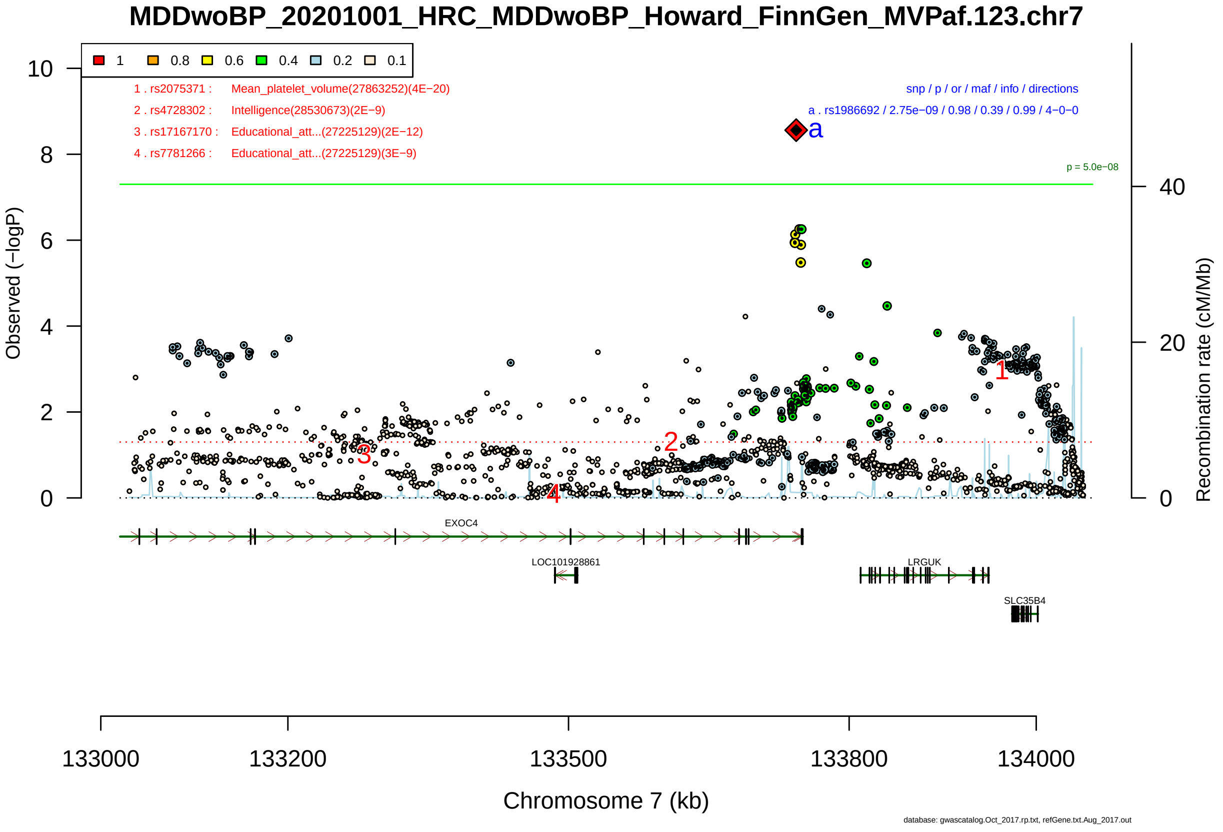

Figure S2-147: Regional plot of GWAS locus No. 147 , with rs1414592 as lead SNP.

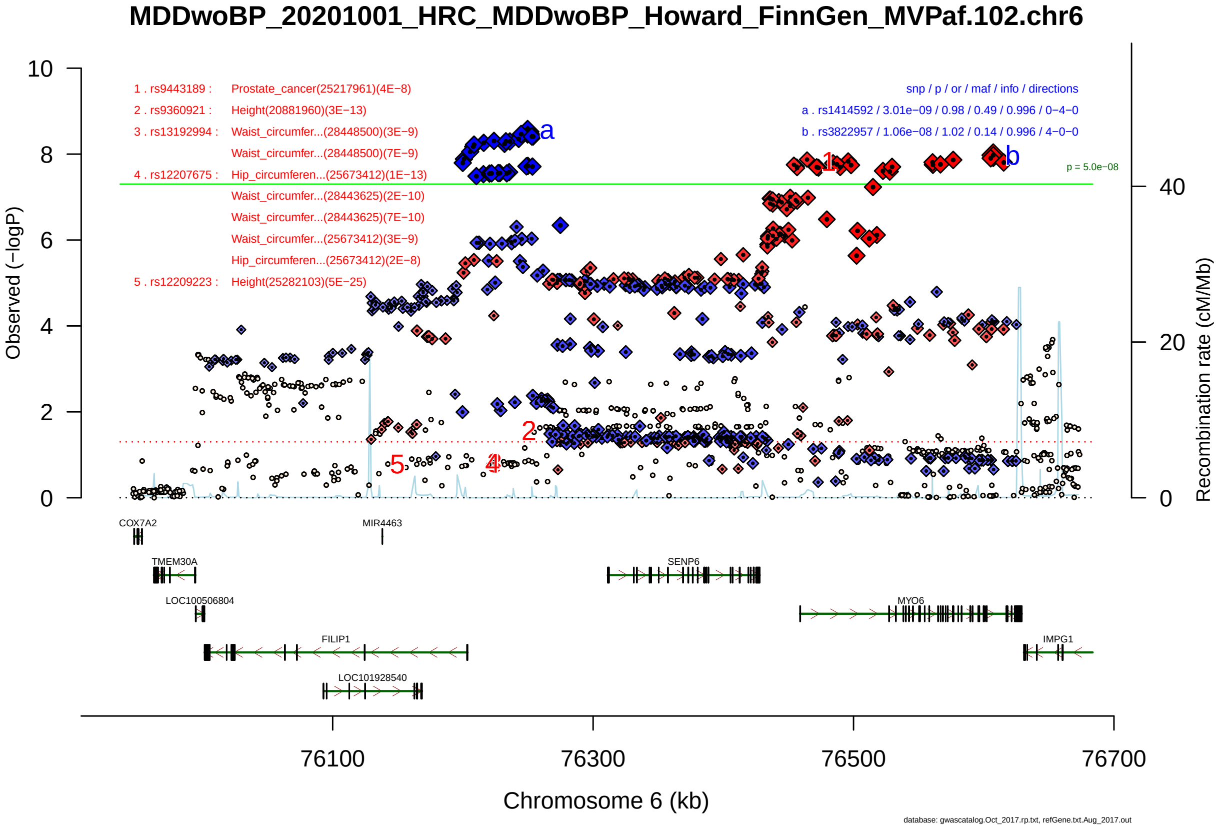

Figure S2-148: Regional plot of GWAS locus No. 148 , with rs4880918 as lead SNP.

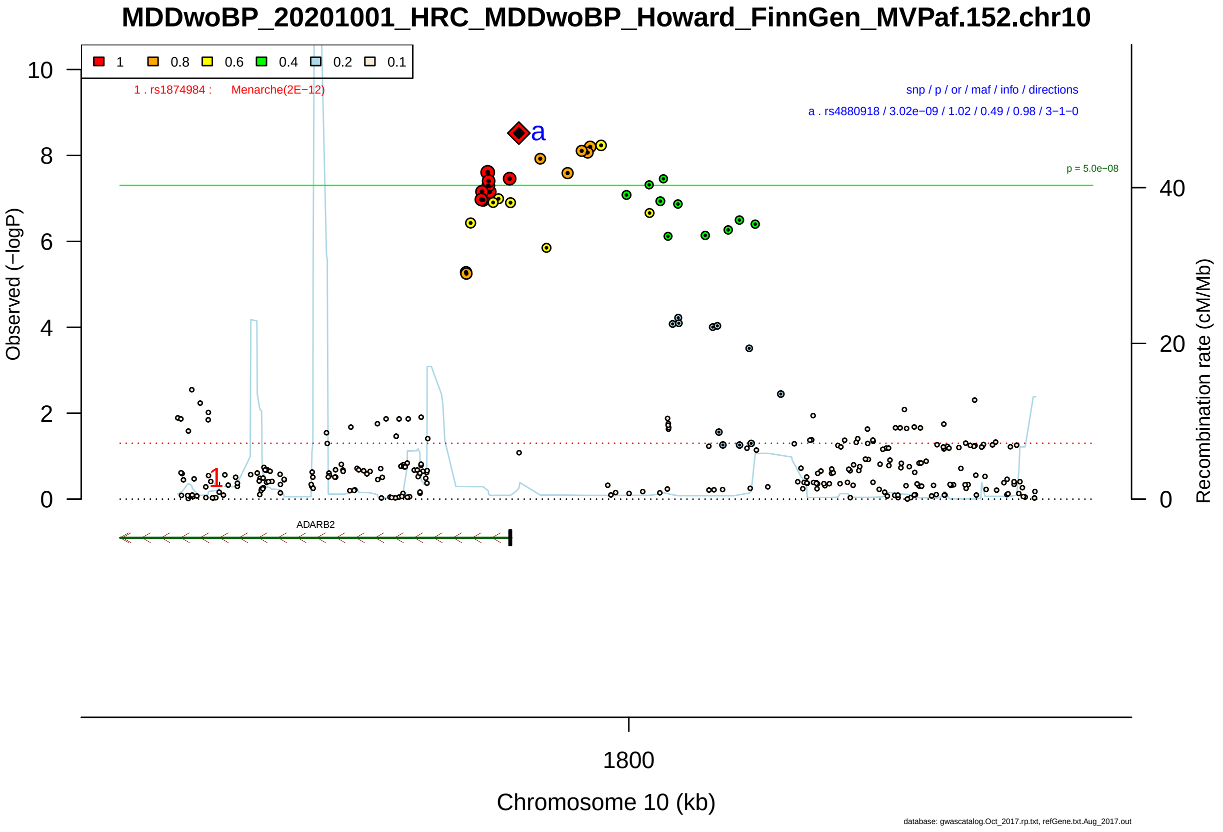

Figure S2-149: Regional plot of GWAS locus No. 149 , with rs1909696 as lead SNP.

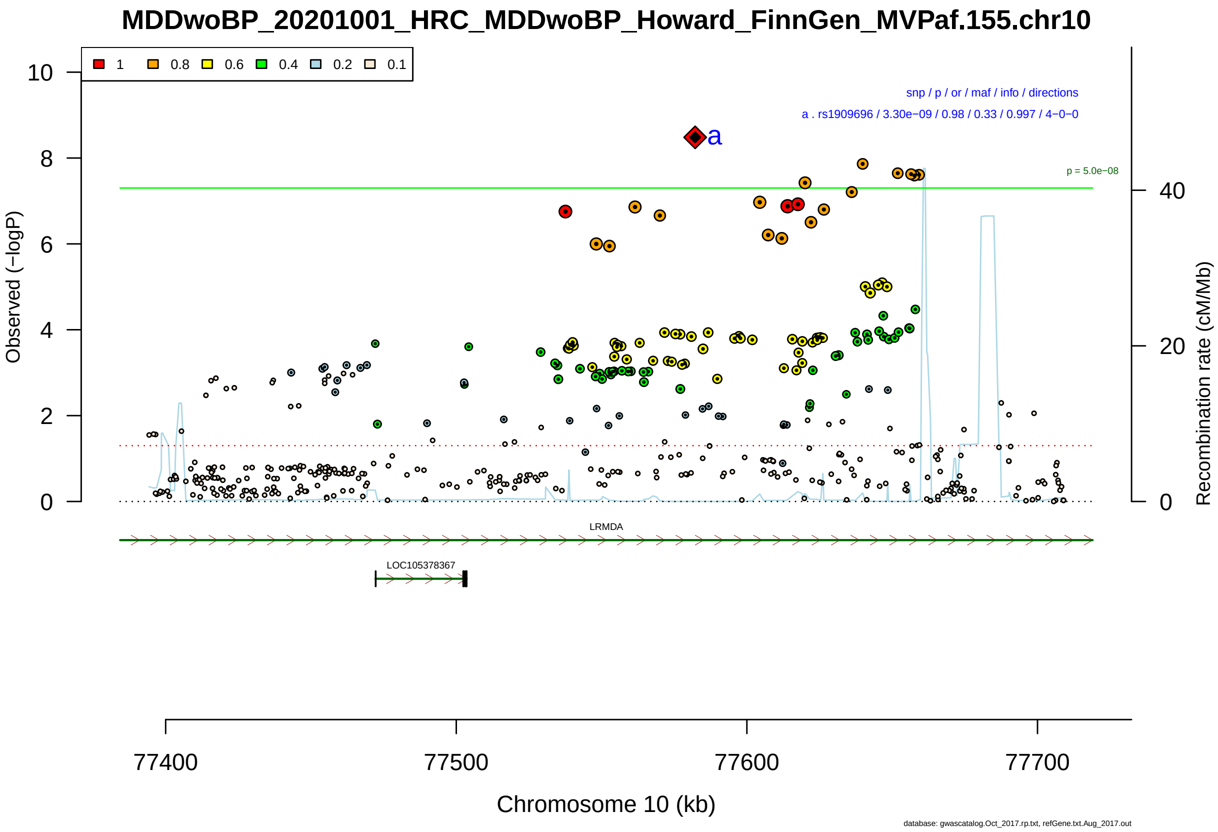

Figure S2-150: Regional plot of GWAS locus No. 150 , with rs7141014 as lead SNP.

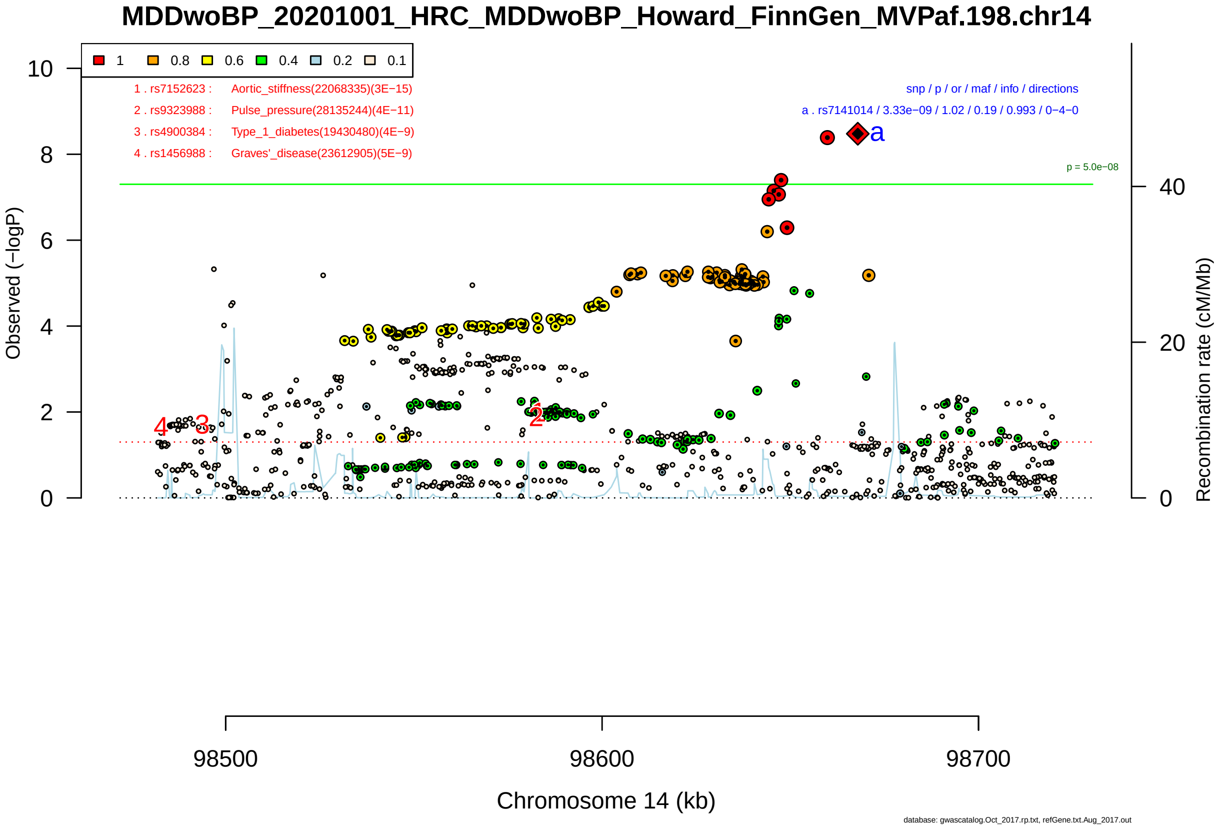

Figure S2-151: Regional plot of GWAS locus No. 151 , with rs74934992 as lead SNP.

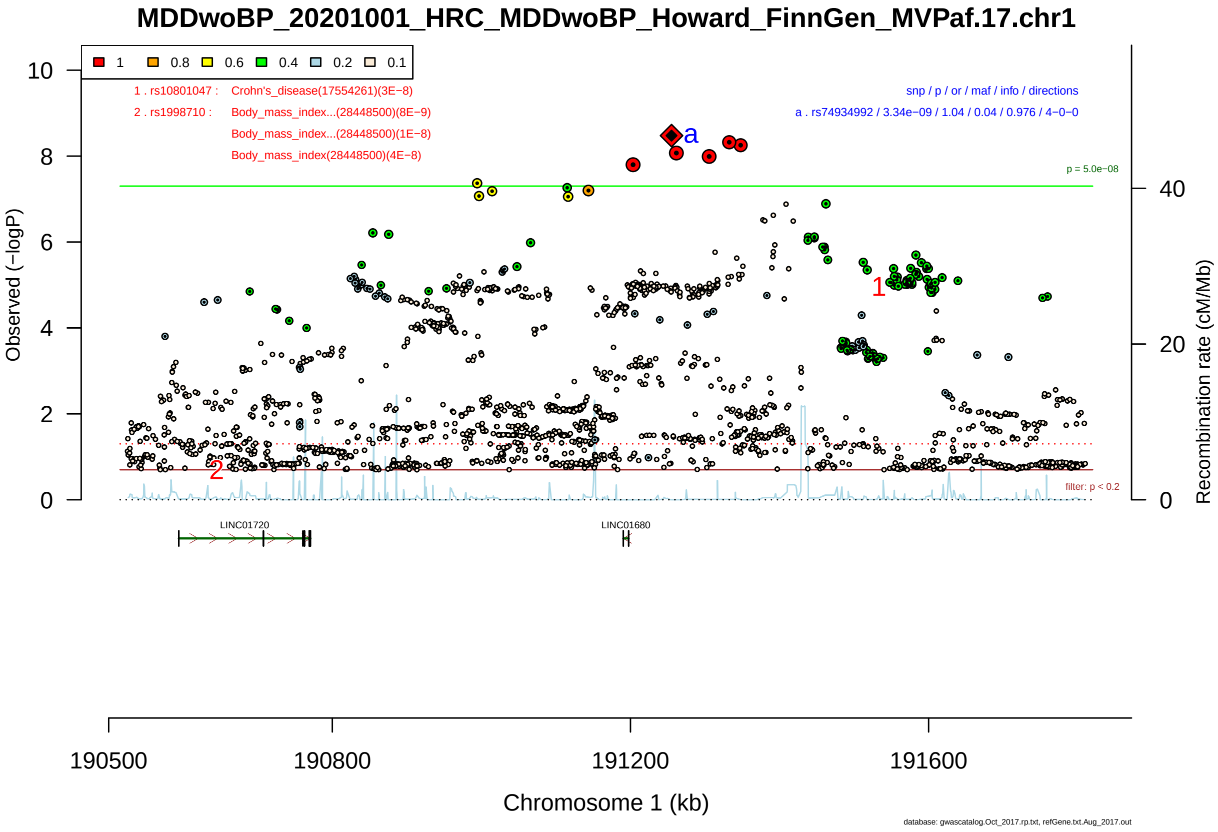

Figure S2-152: Regional plot of GWAS locus No. 152 , with rs9561331 as lead SNP.

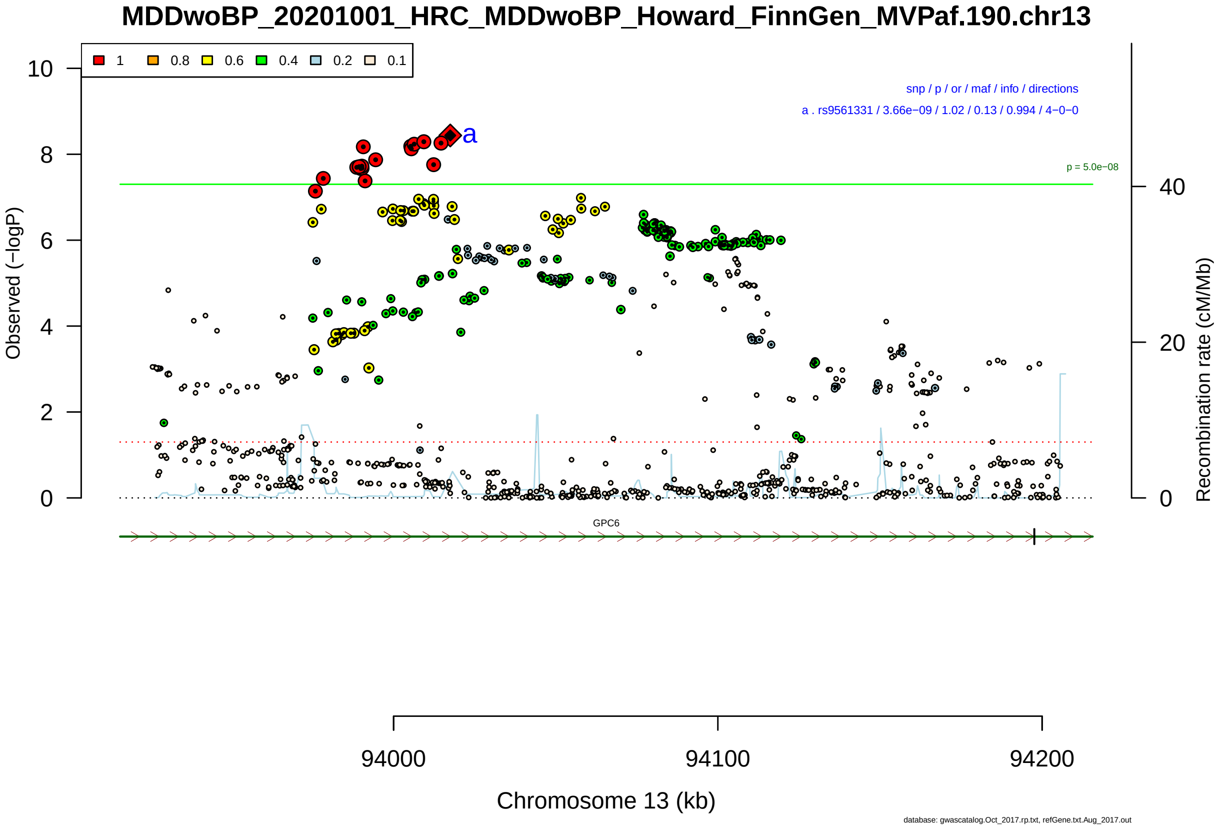

Figure S2-153: Regional plot of GWAS locus No. 153 , with rs7564151 as lead SNP.

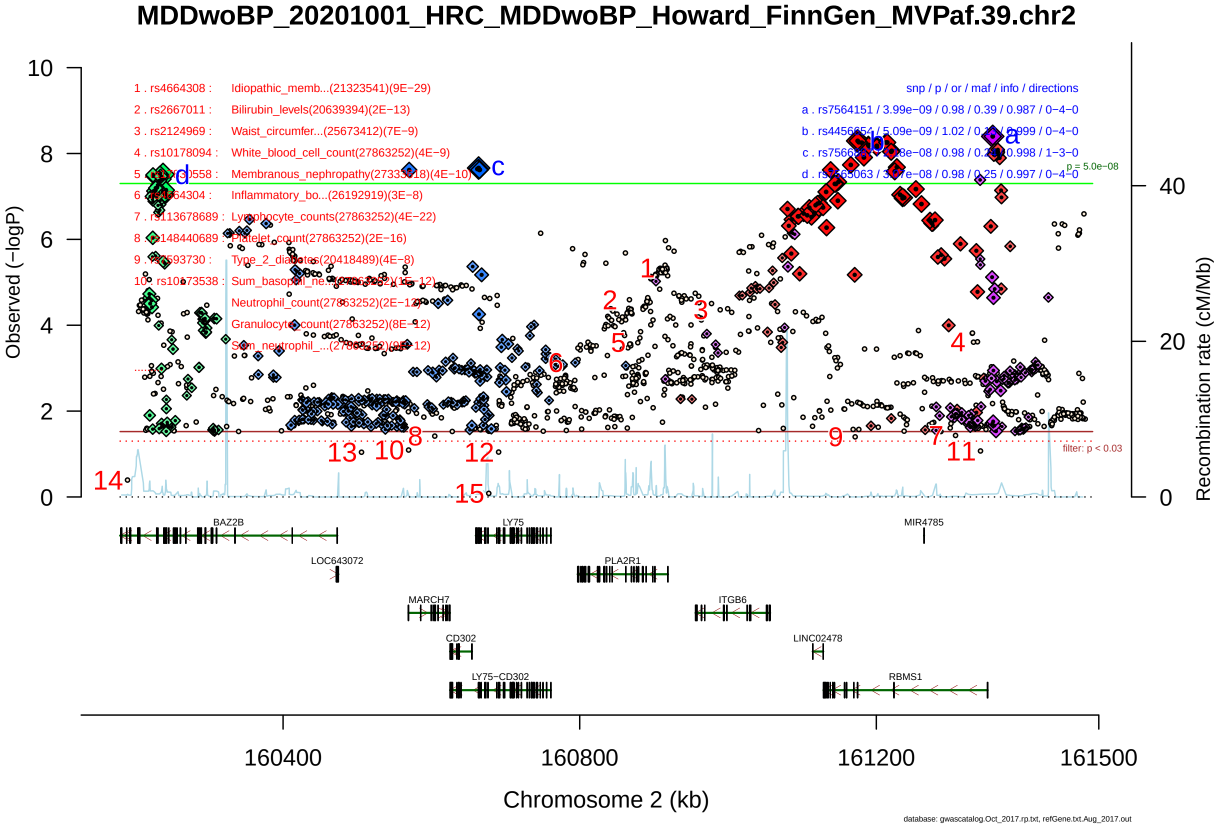

Figure S2-154: Regional plot of GWAS locus No. 154 , with rs4478545 as lead SNP.

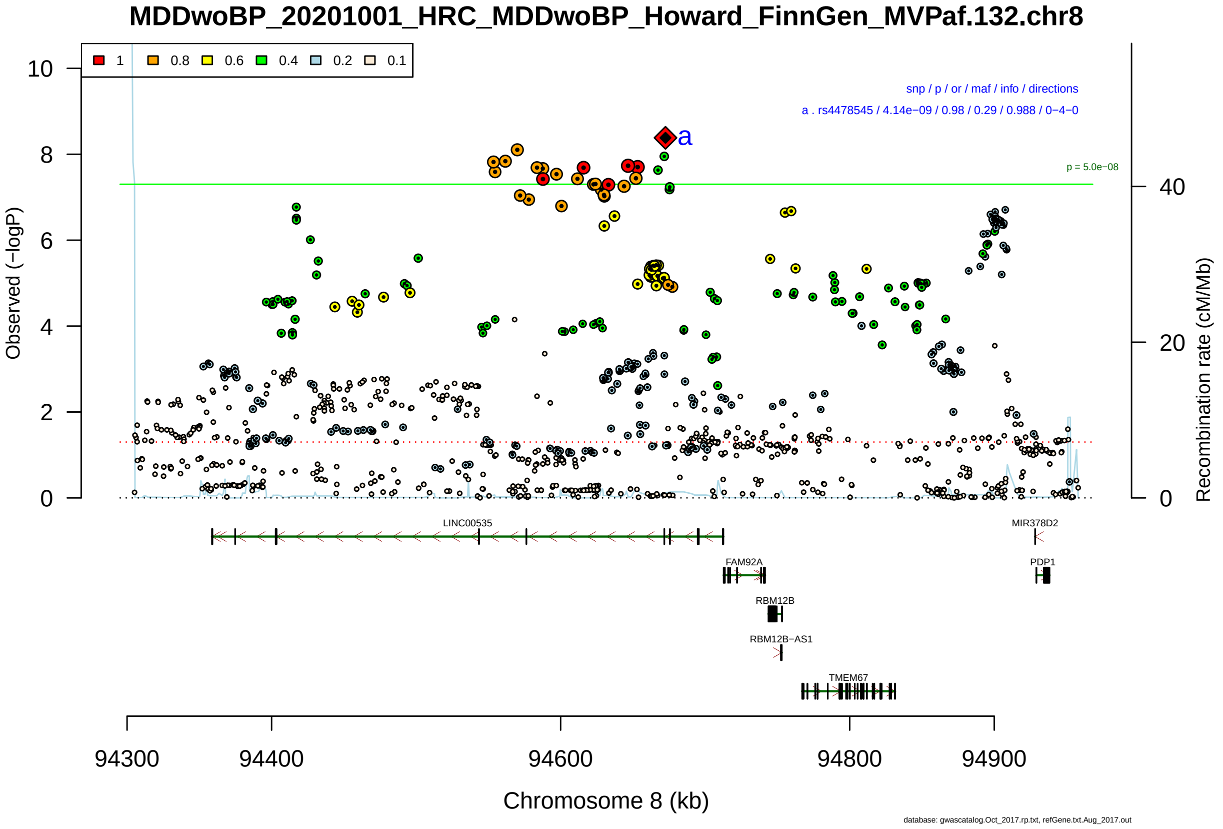

Figure S2-155: Regional plot of GWAS locus No. 155 , with rs4262121 as lead SNP.

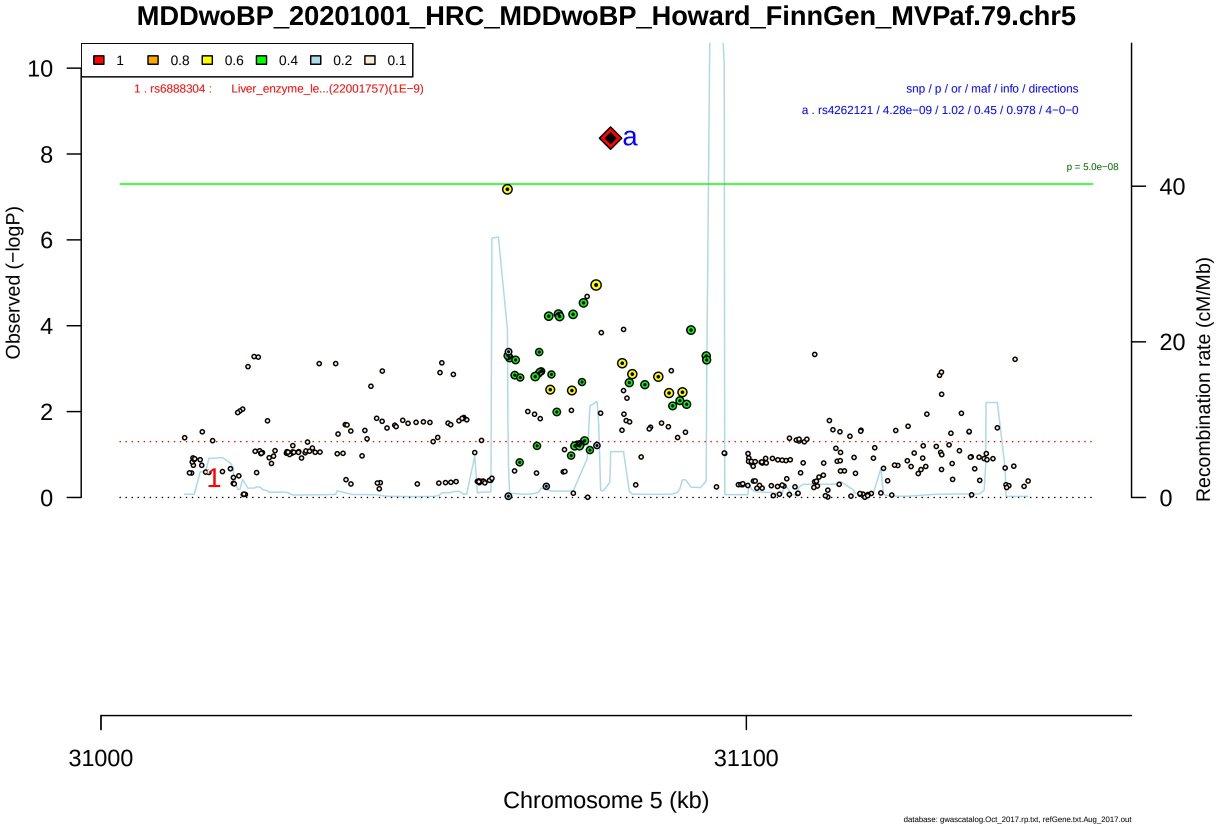

Figure S2-156: Regional plot of GWAS locus No. 156 , with rs7624336 as lead SNP.

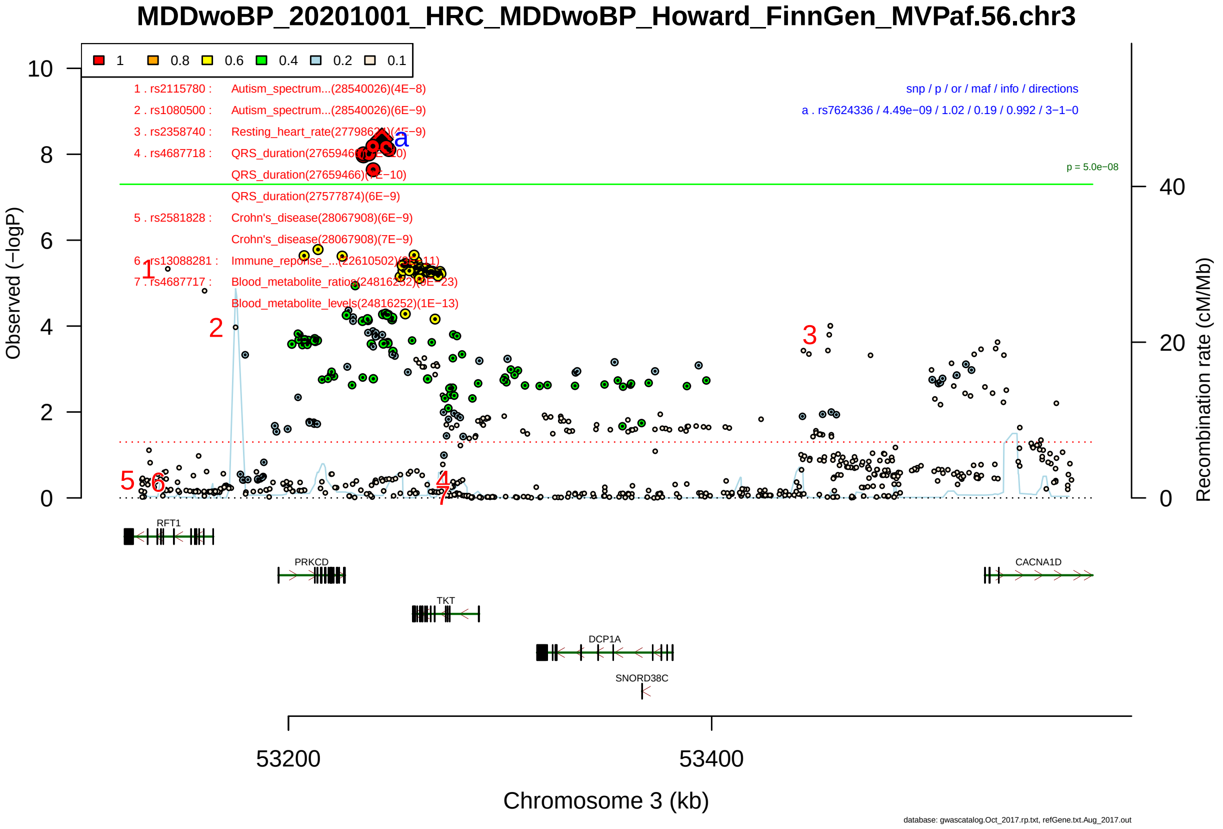

Figure S2-157: Regional plot of GWAS locus No. 157 , with rs2298969 as lead SNP.

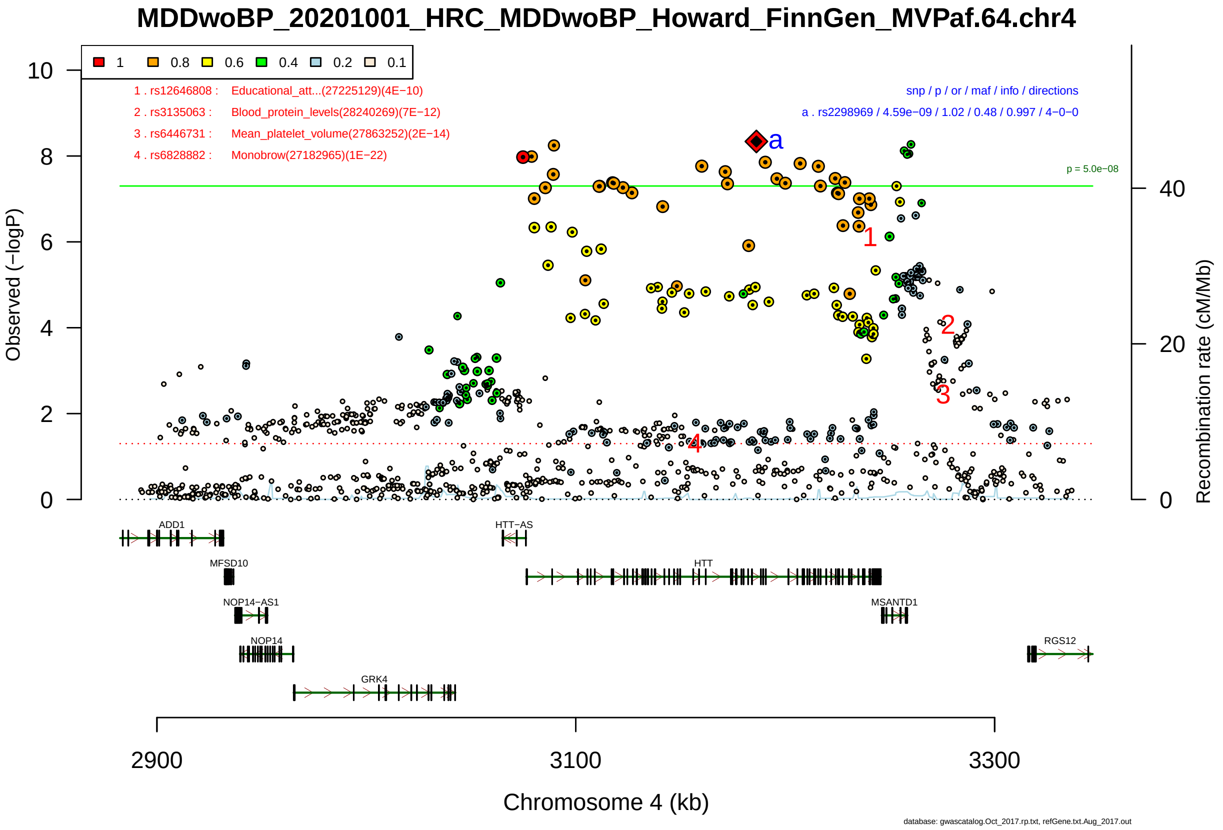

Figure S2-158: Regional plot of GWAS locus No. 158 , with rs667138 as lead SNP.

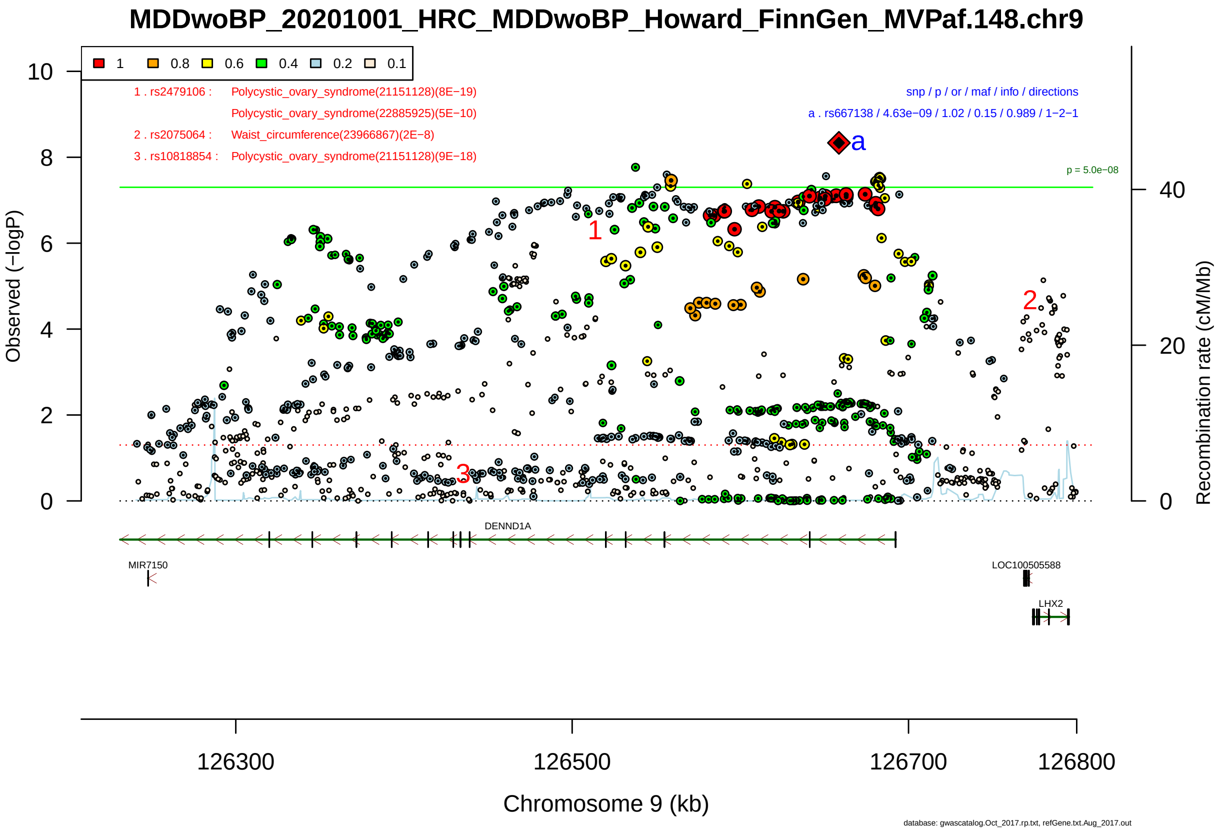

Figure S2-159: Regional plot of GWAS locus No. 159 , with rs1320138 as lead SNP.

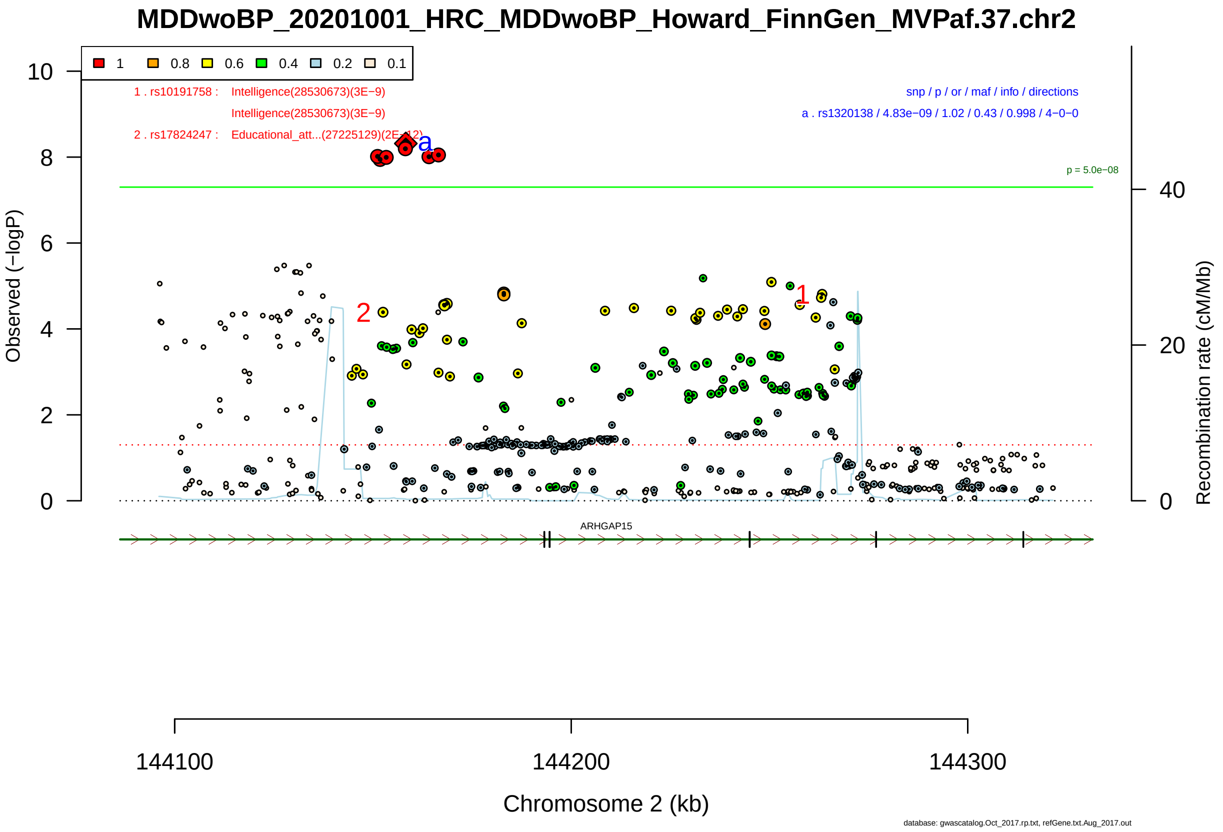

Figure S2-160: Regional plot of GWAS locus No. 160 , with rs4247258 as lead SNP.

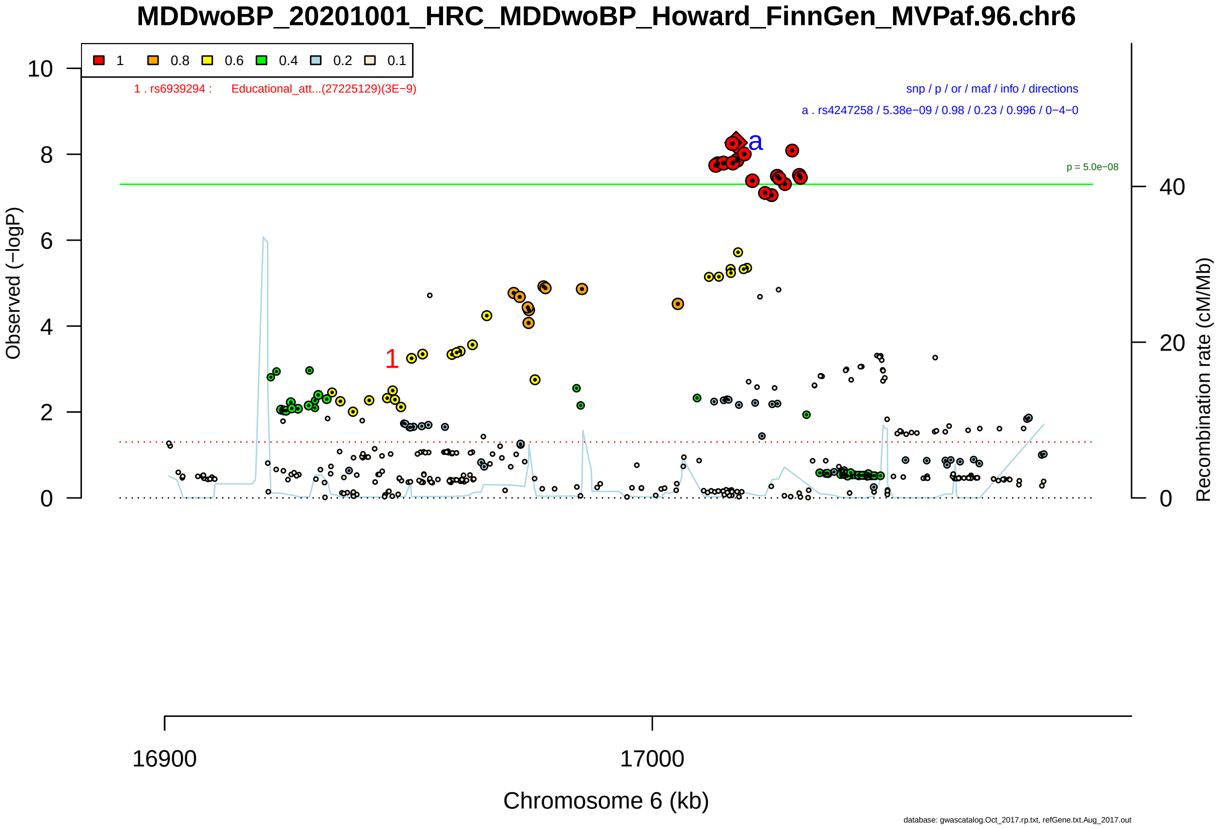

Figure S2-161: Regional plot of GWAS locus No. 161 , with rs1433965 as lead SNP.

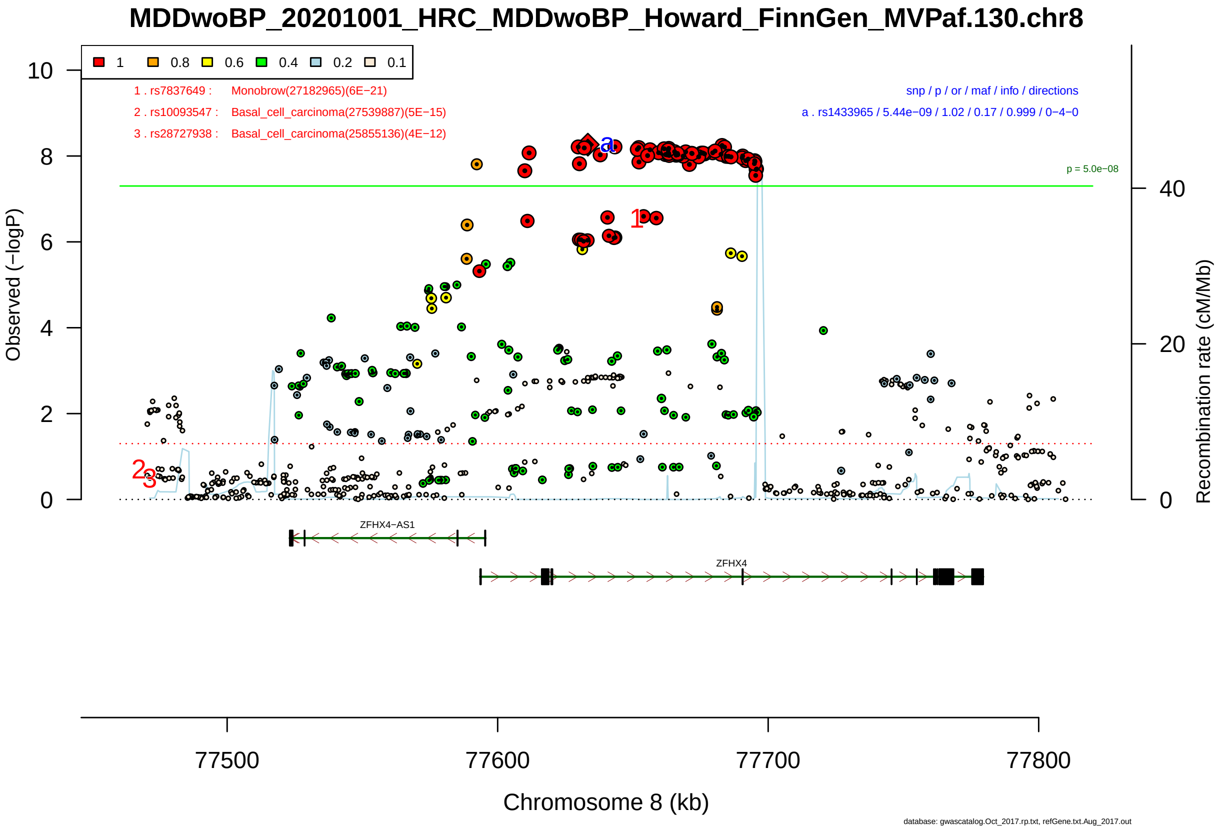

Figure S2-162: Regional plot of GWAS locus No. 162 , with rs7513962 as lead SNP.

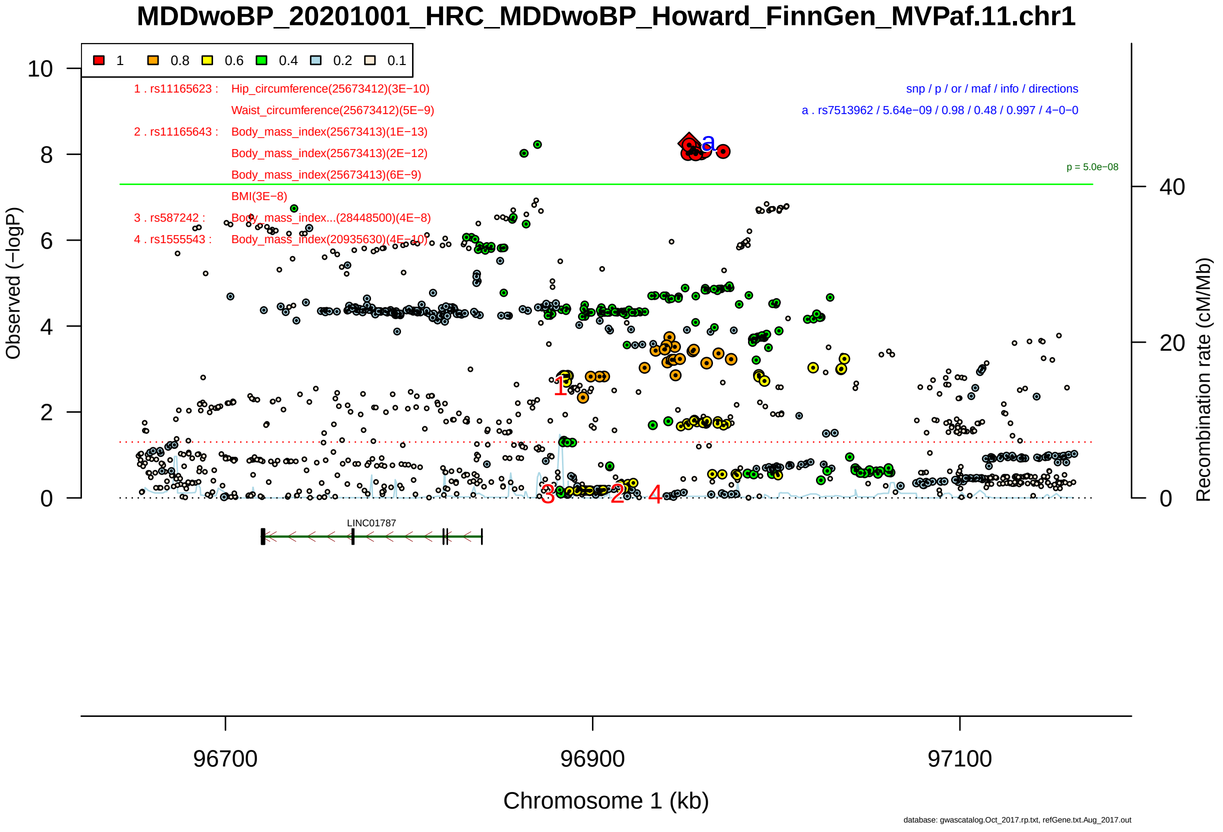

Figure S2-163: Regional plot of GWAS locus No. 163 , with rs73428490 as lead SNP.

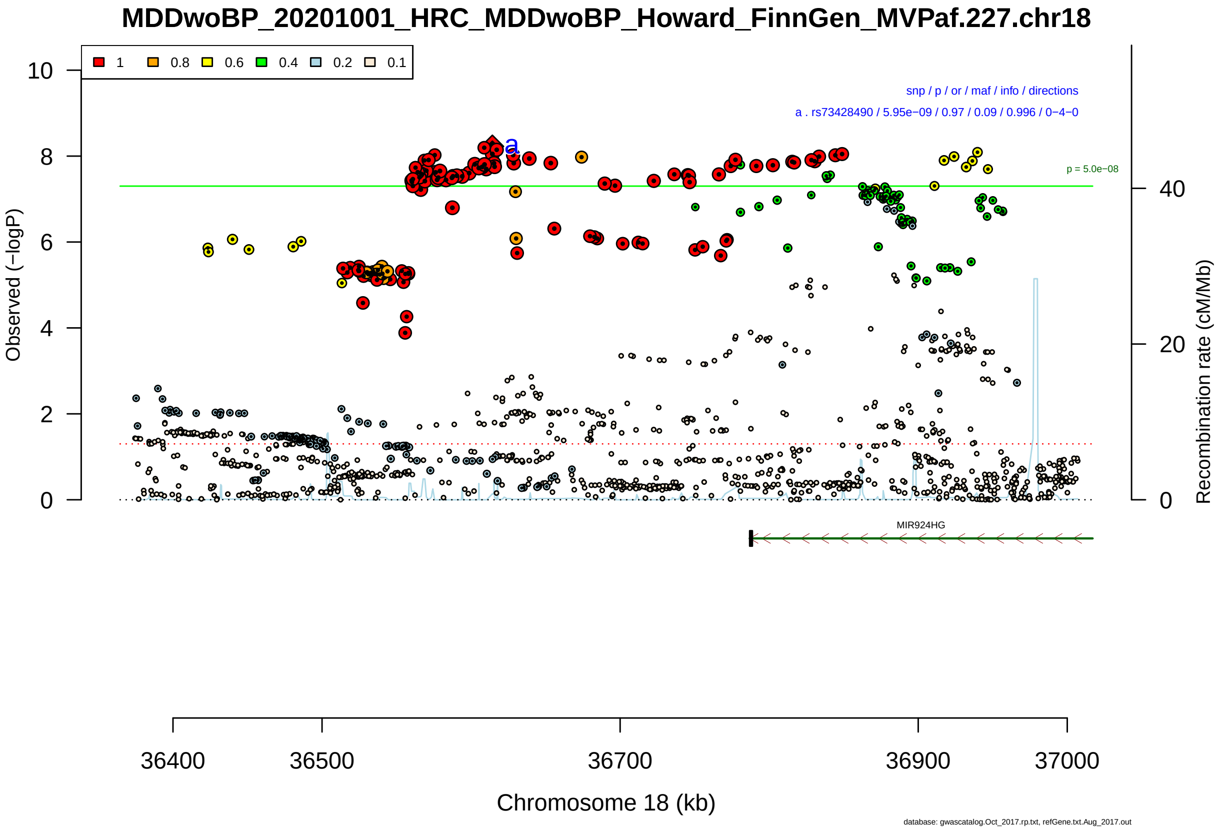

Figure S2-164: Regional plot of GWAS locus No. 164 , with rs10947690 as lead SNP.

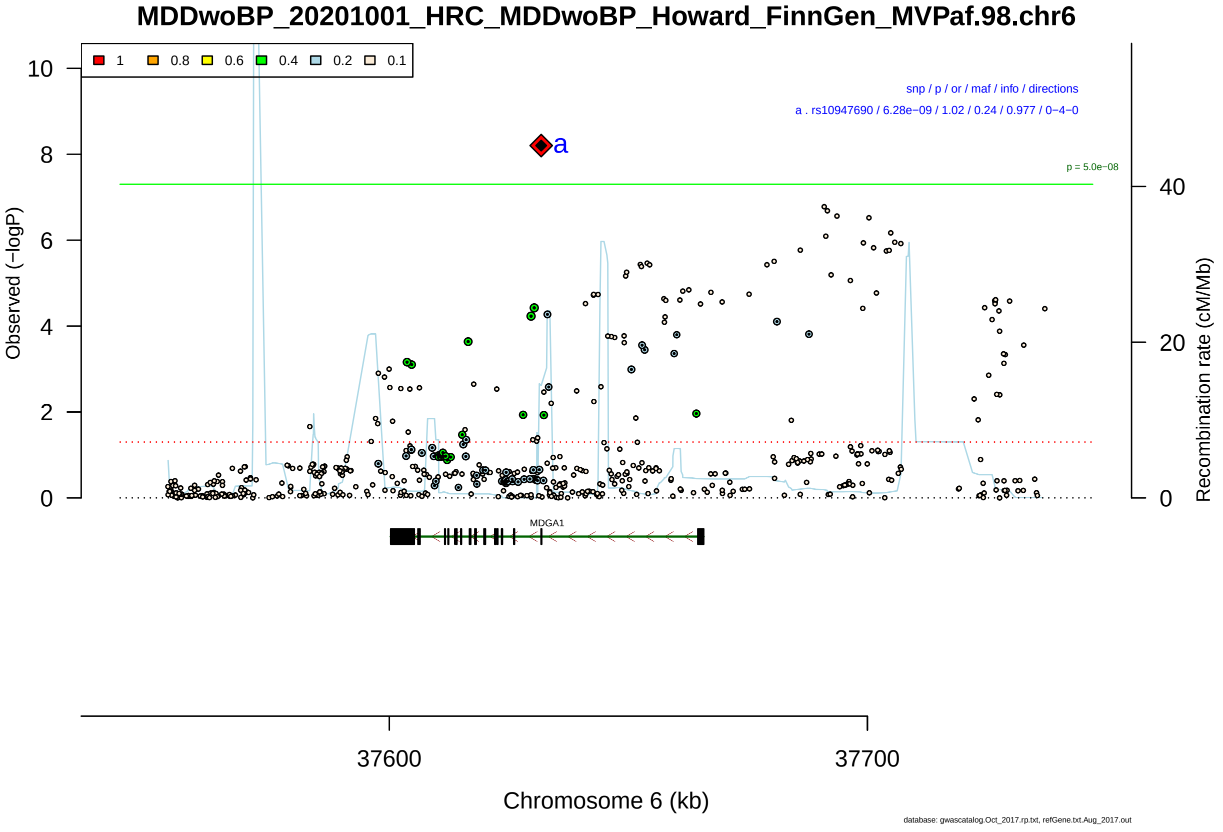

Figure S2-165: Regional plot of GWAS locus No. 165 , with rs10261857 as lead SNP.

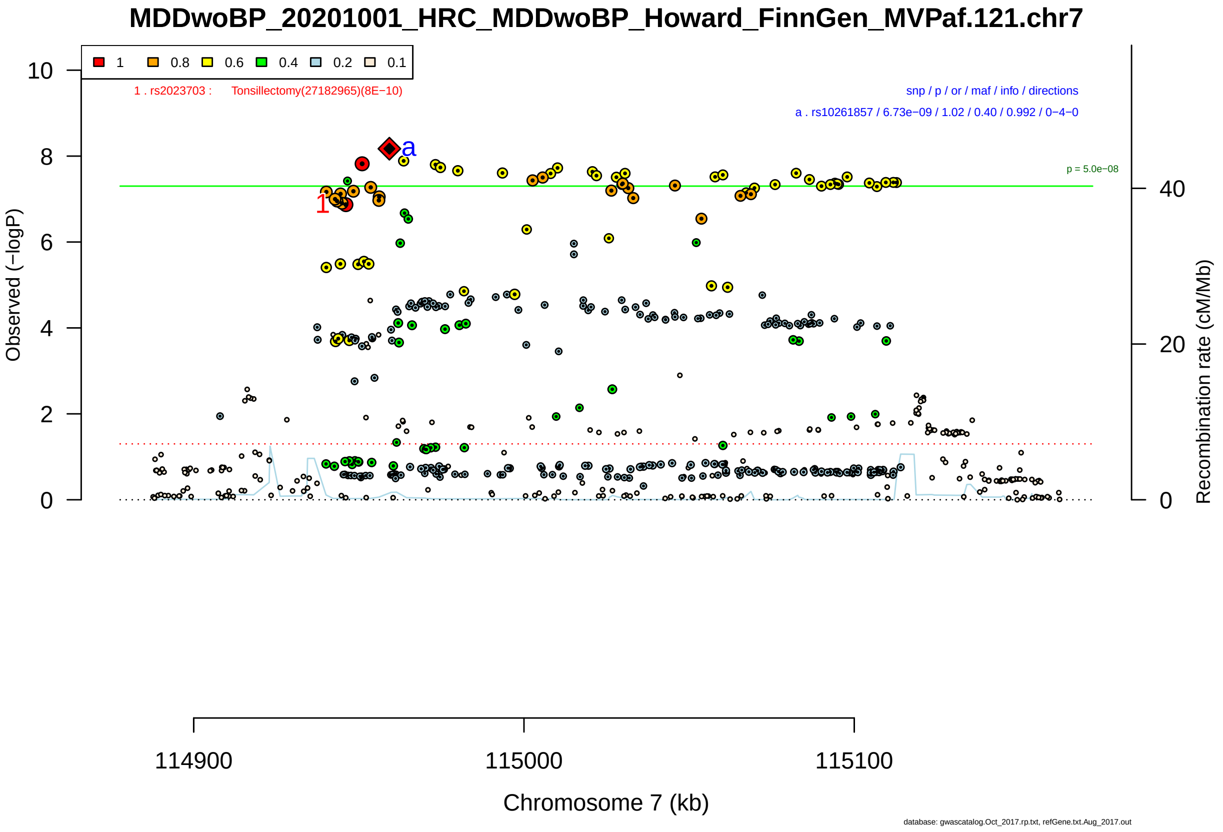

Figure S2-166: Regional plot of GWAS locus No. 166 , with rs11062170 as lead SNP.

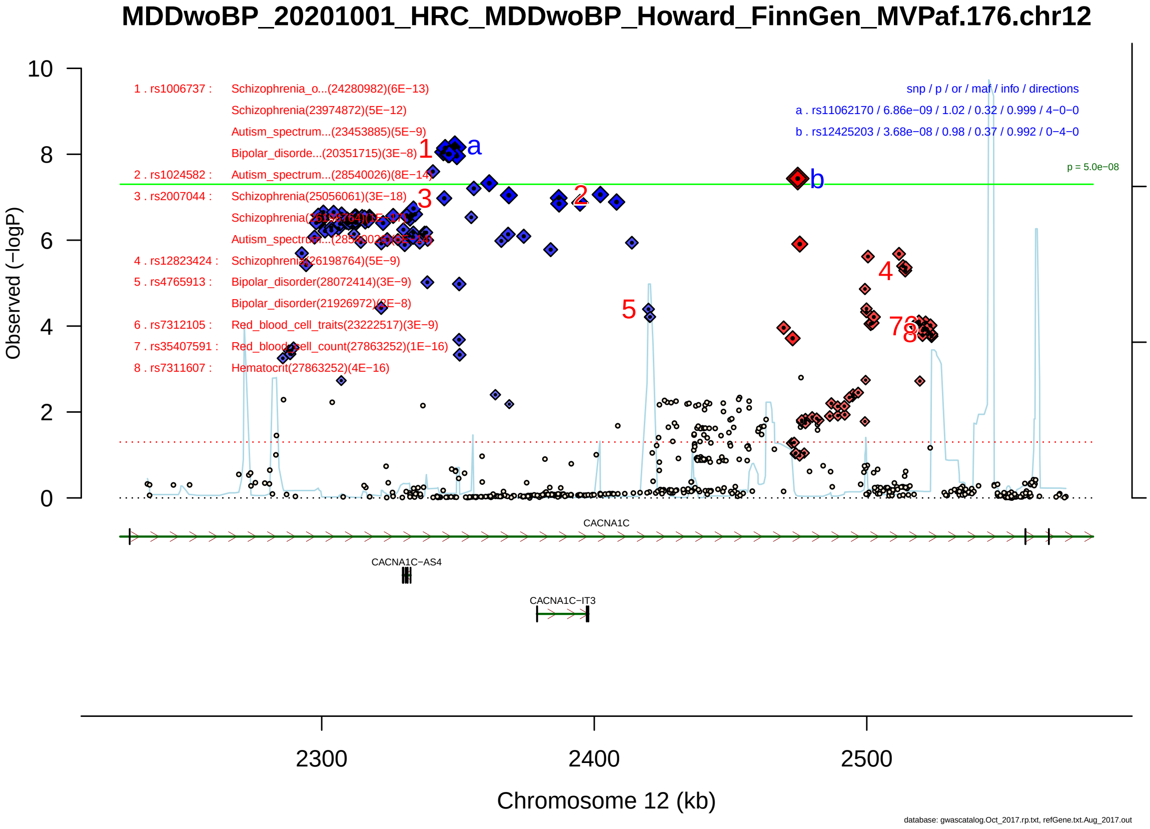

Figure S2-167: Regional plot of GWAS locus No. 167 , with rs7599039 as lead SNP.

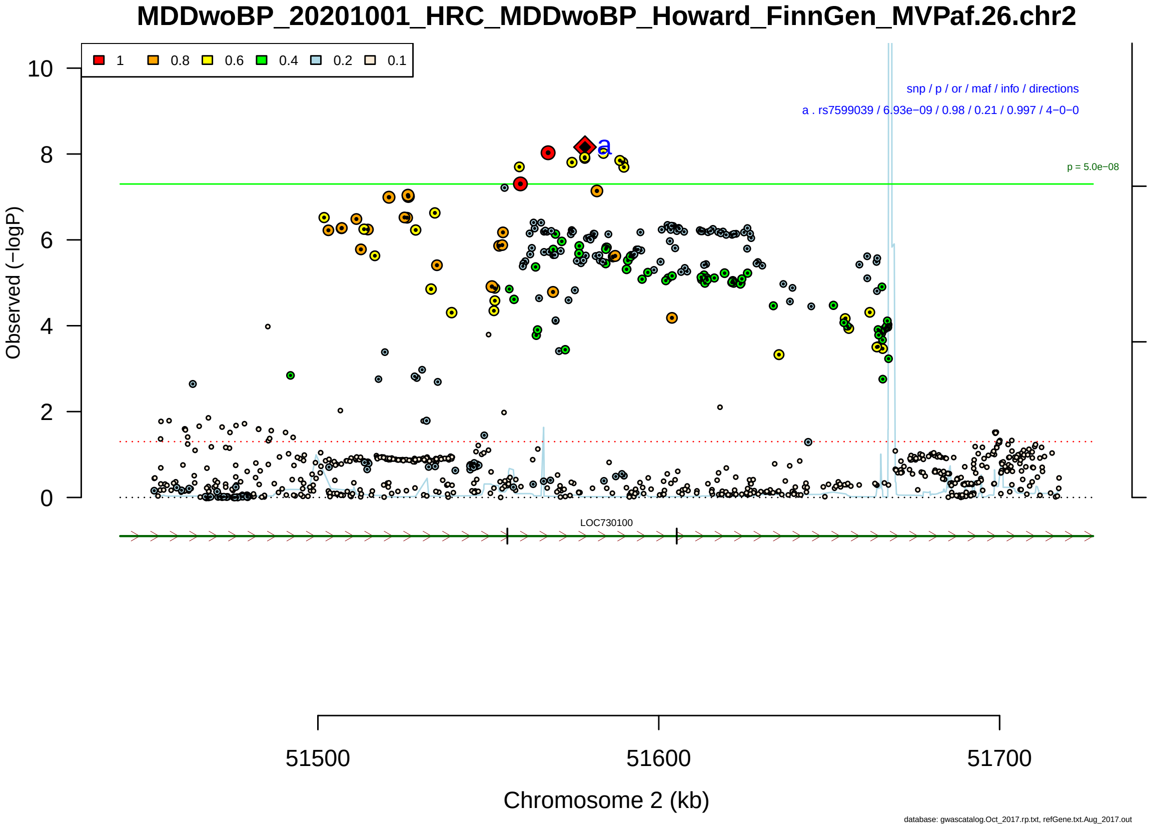

Figure S2-168: Regional plot of GWAS locus No. 168 , with rs10805794 as lead SNP.

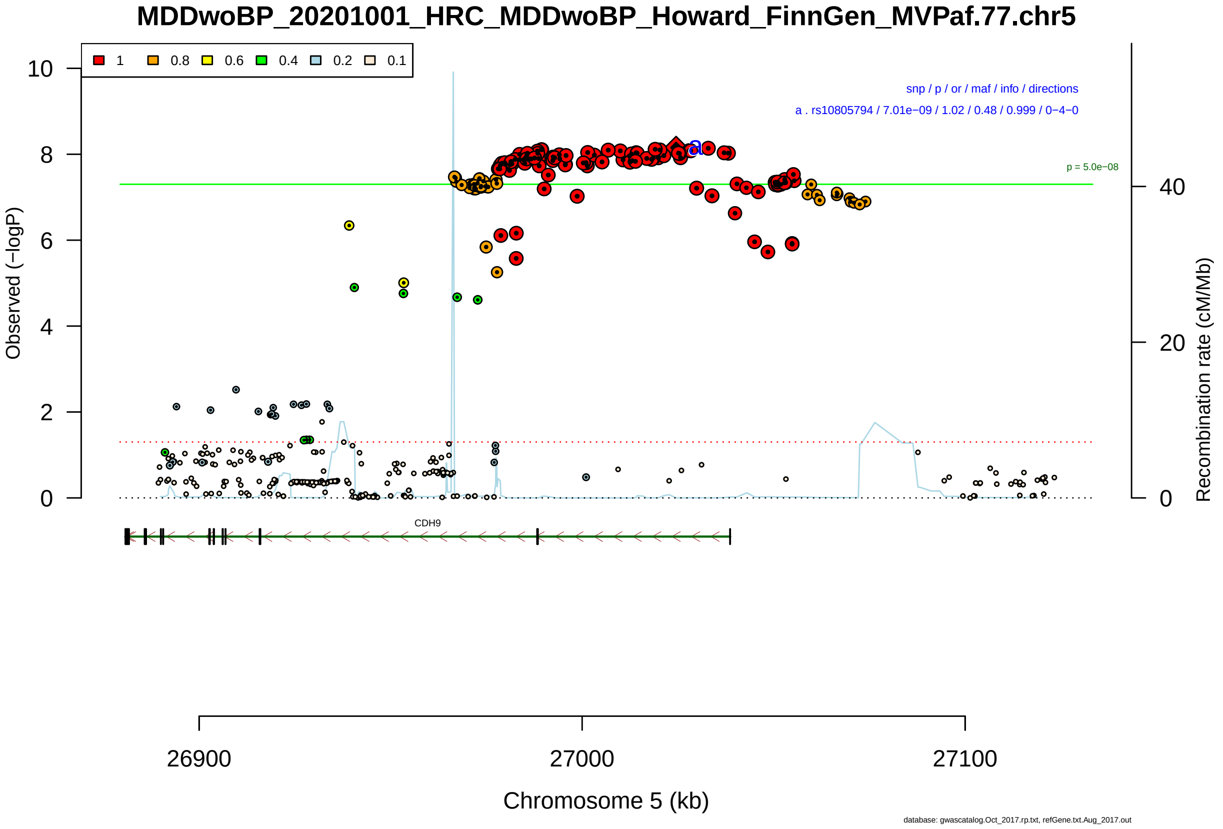

Figure S2-169: Regional plot of GWAS locus No. 169 , with rs3852788 as lead SNP.

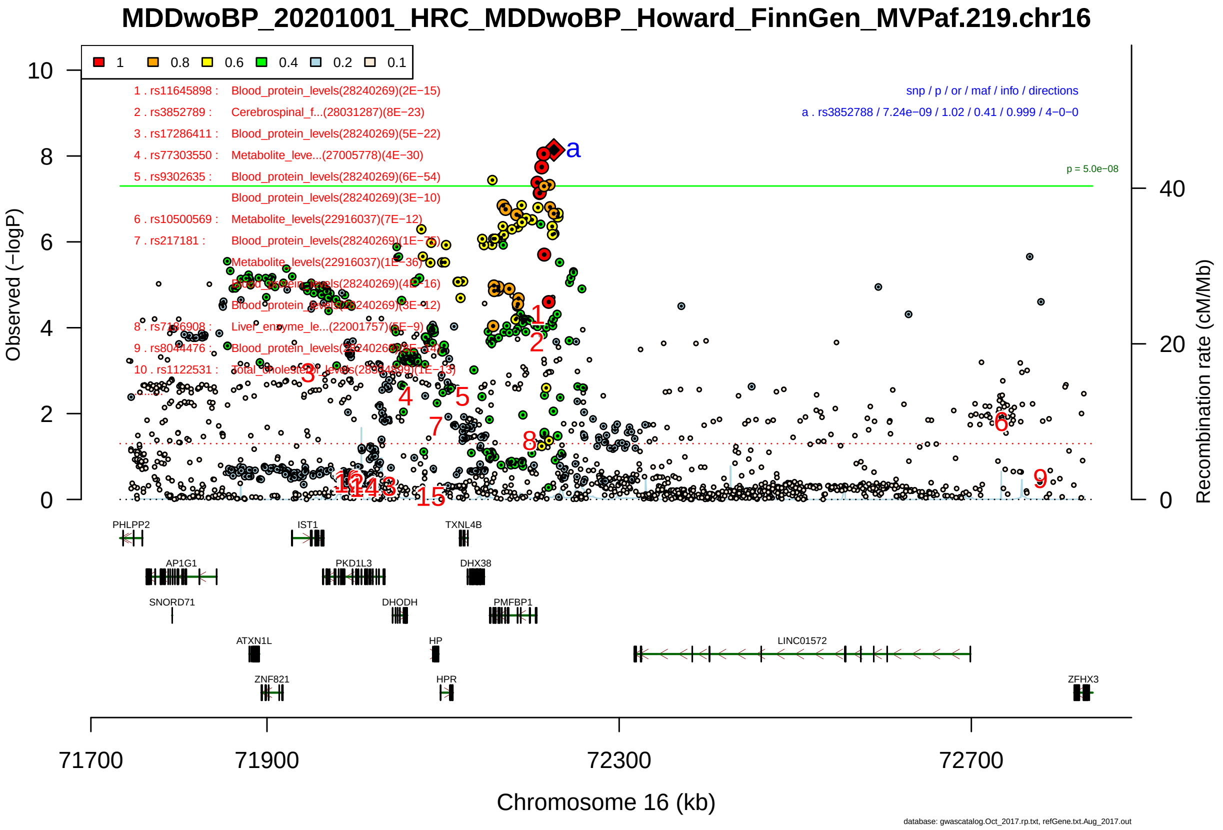

Figure S2-170: Regional plot of GWAS locus No. 170 , with rs75581564 as lead SNP.

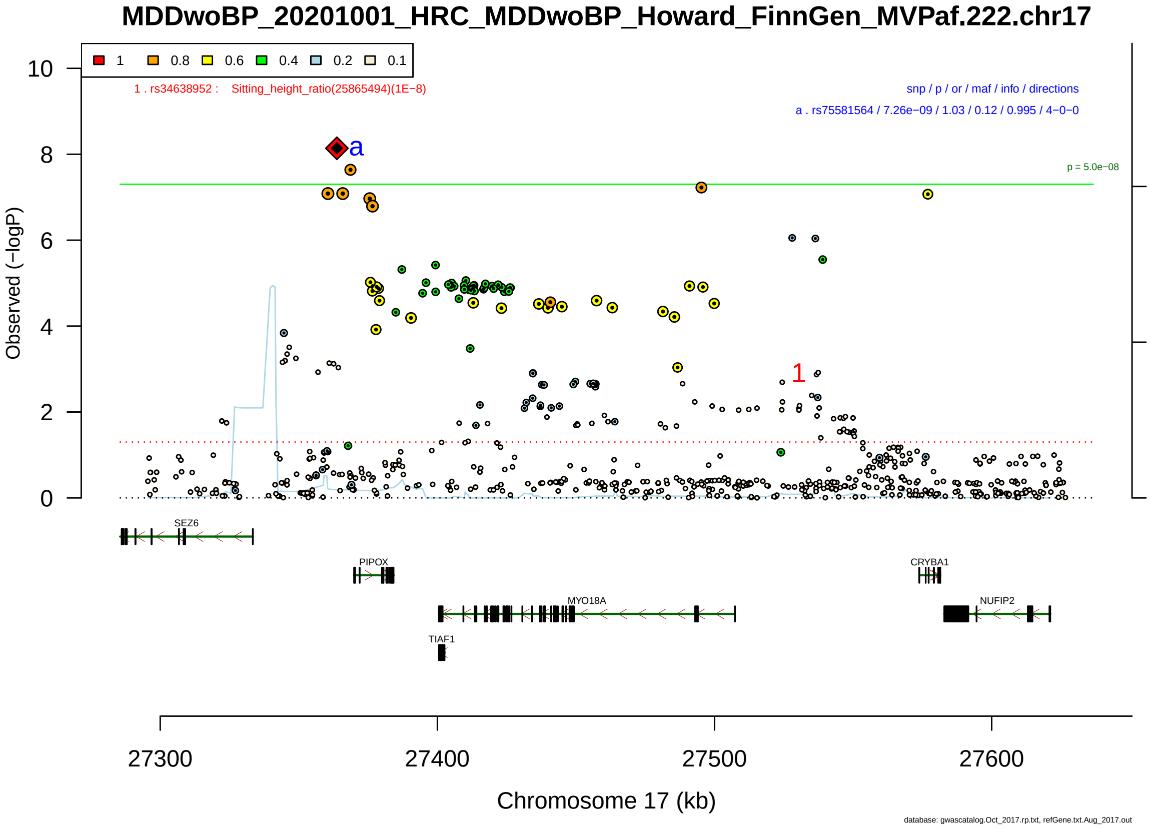

Figure S2-171: Regional plot of GWAS locus No. 171 , with rs2585399 as lead SNP.

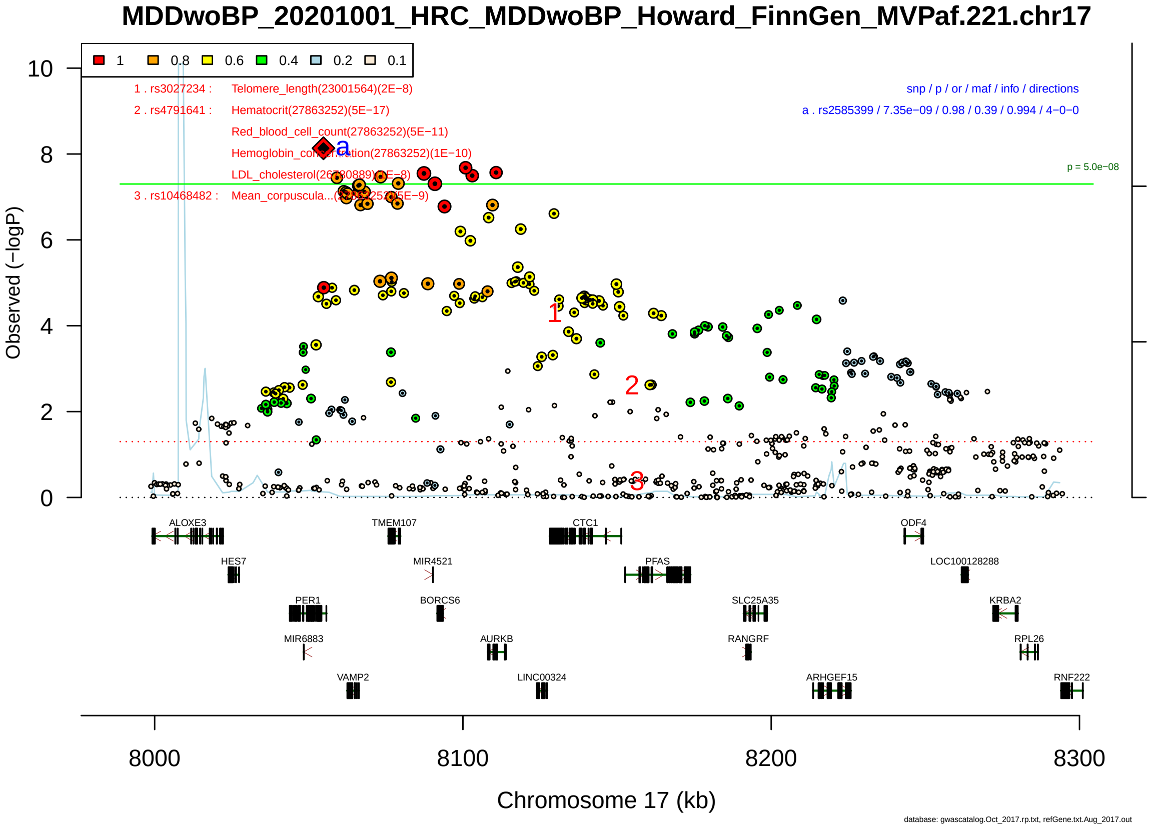

Figure S2-172: Regional plot of GWAS locus No. 172 , with rs1555132 as lead SNP.

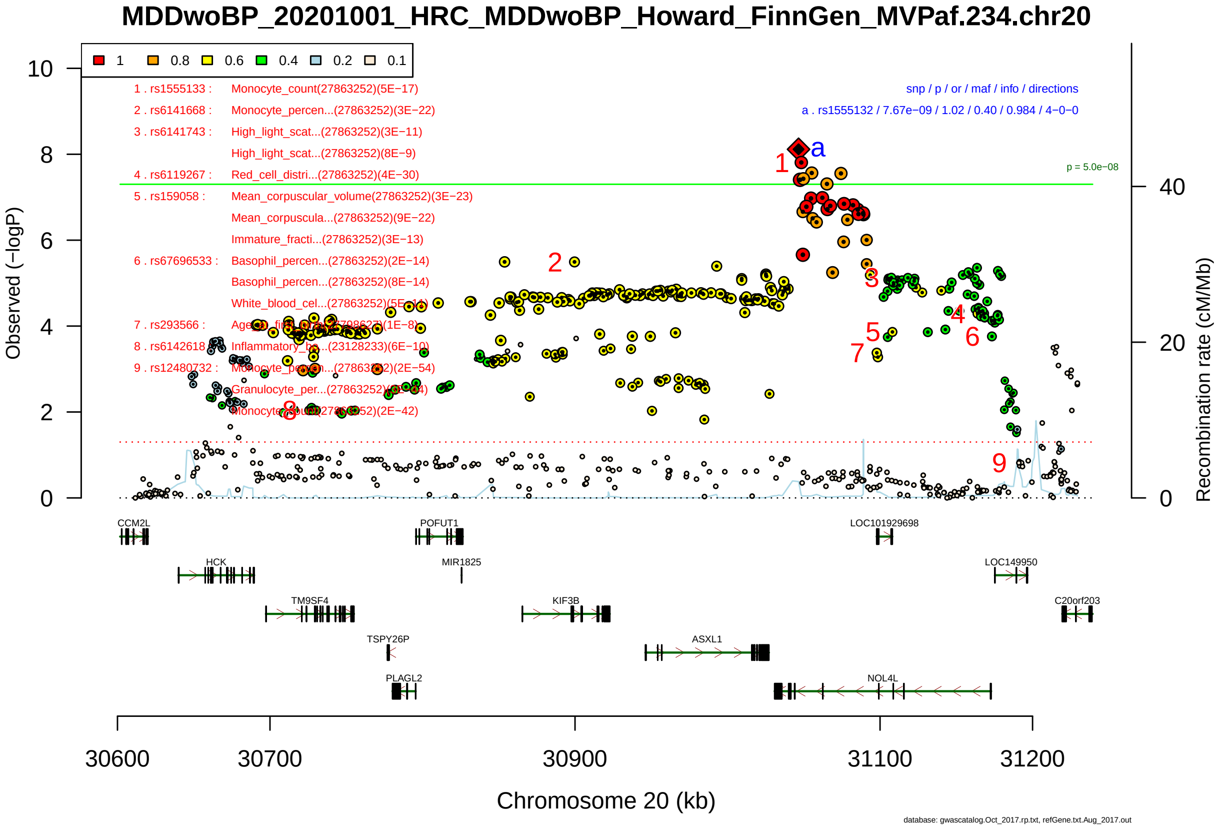

Figure S2-173: Regional plot of GWAS locus No. 173 , with rs12003380 as lead SNP.

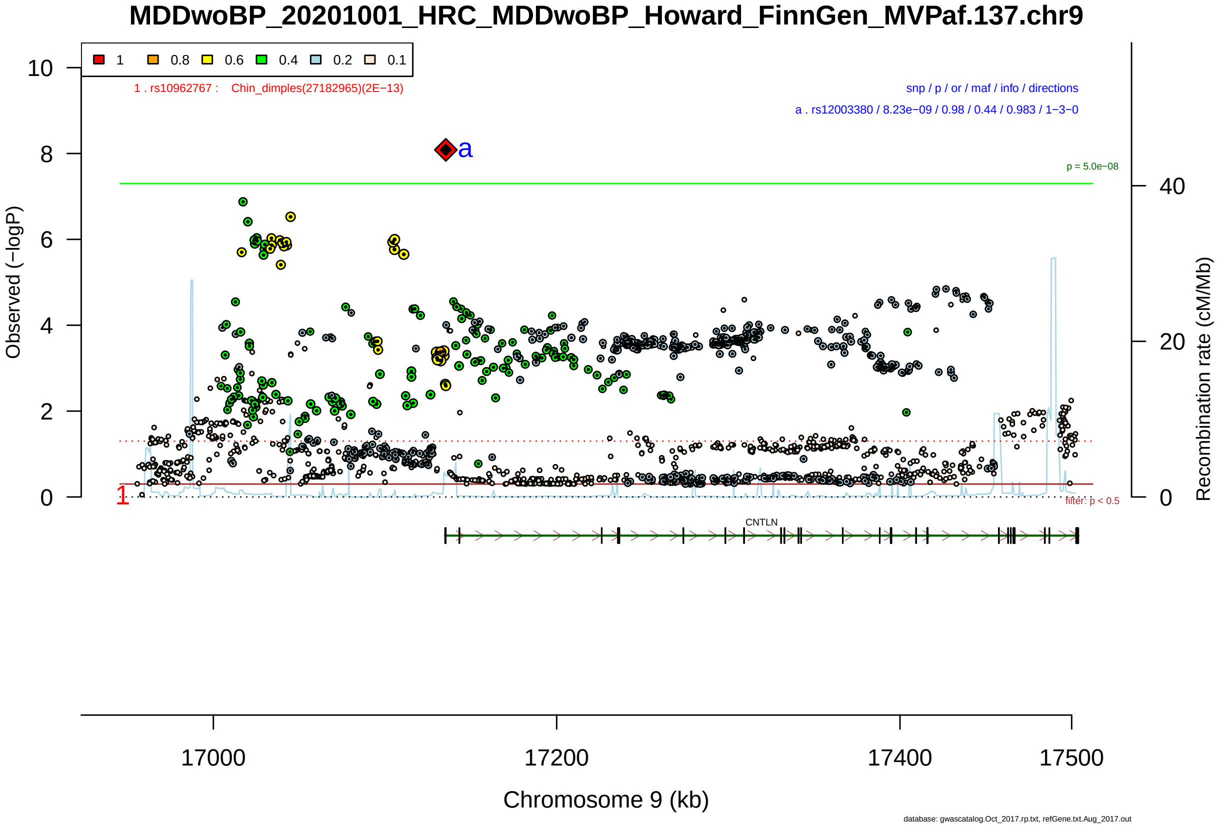

Figure S2-174: Regional plot of GWAS locus No. 174 , with rs17611770 as lead SNP.

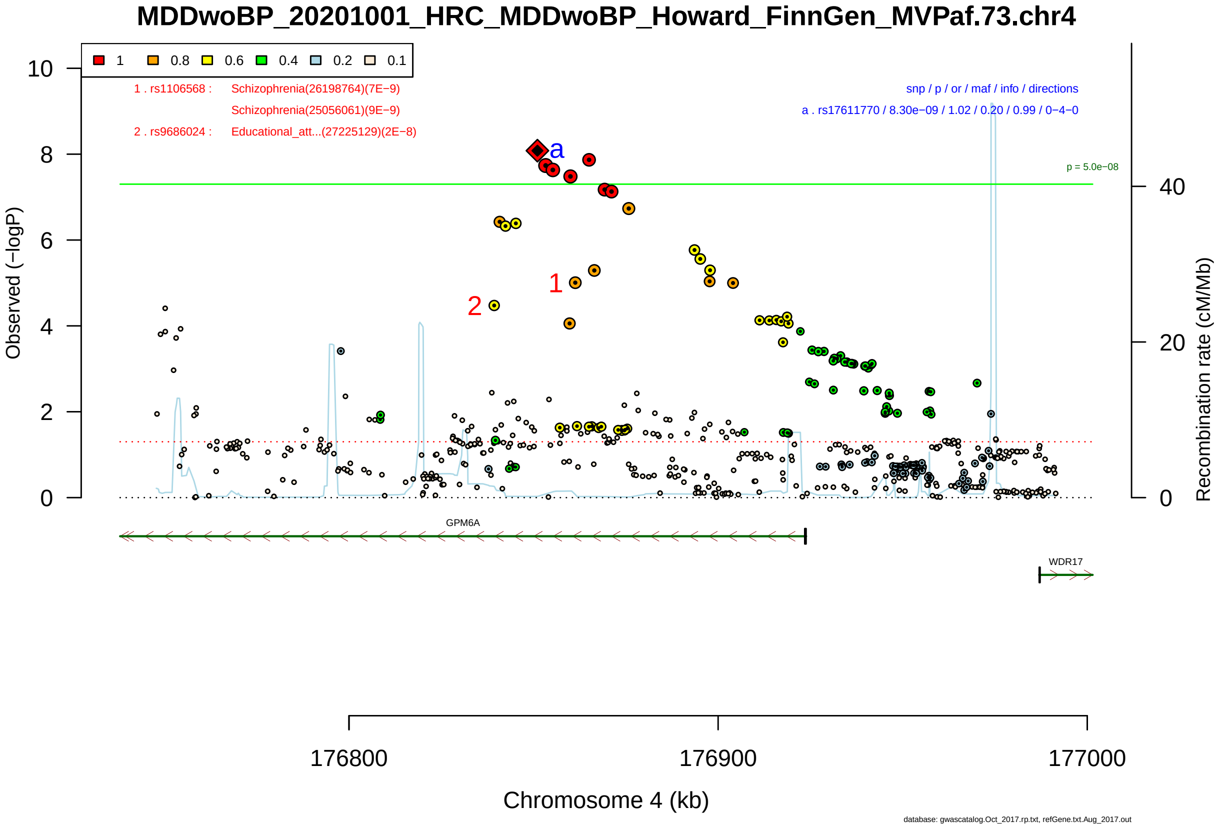

Figure S2-175: Regional plot of GWAS locus No. 175 , with rs12285422 as lead SNP.

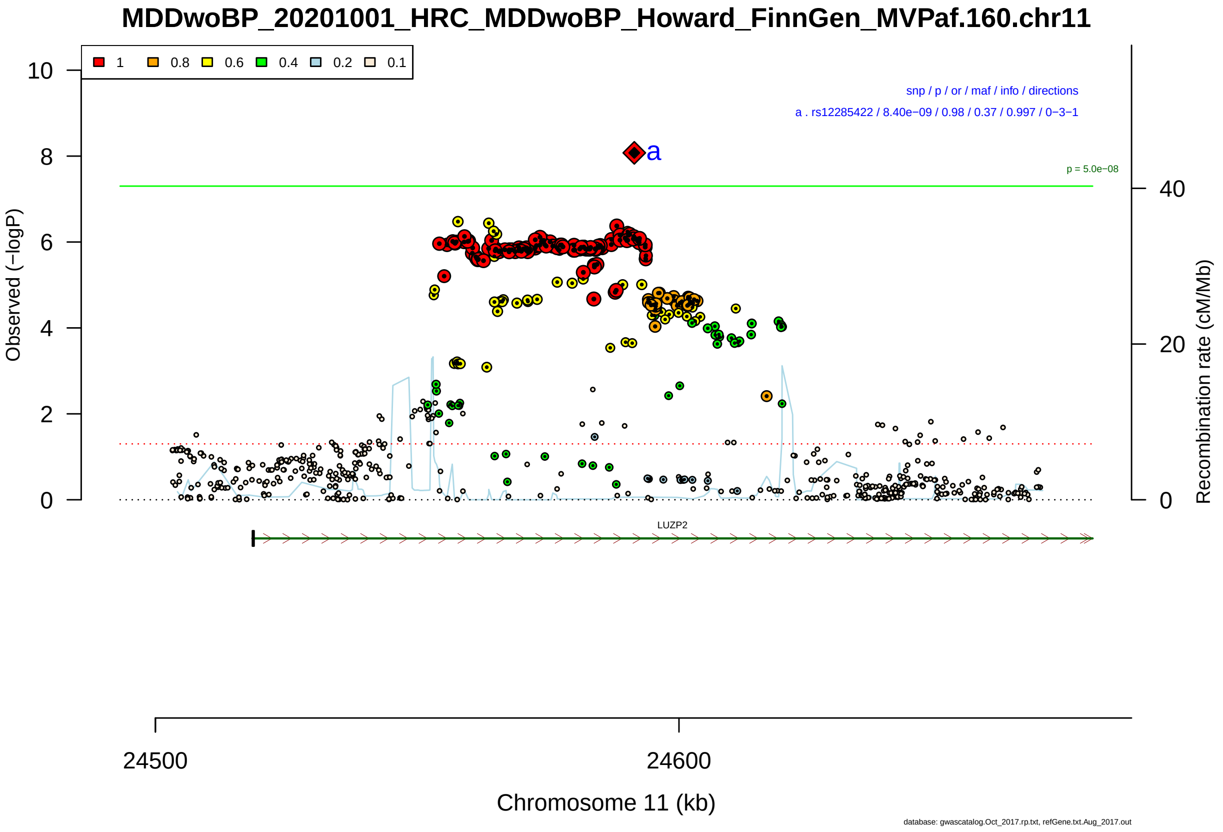

Figure S2-176: Regional plot of GWAS locus No. 176 , with rs9627391 as lead SNP.

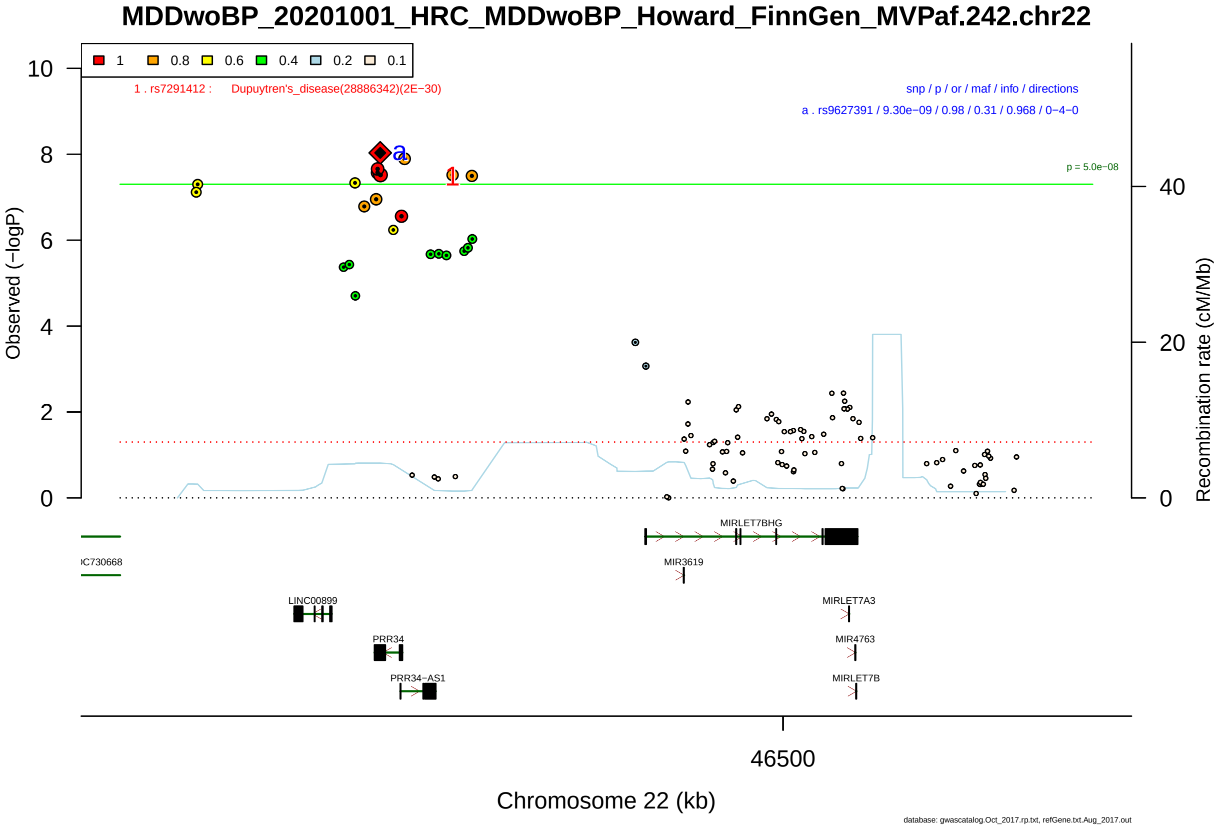

Figure S2-177: Regional plot of GWAS locus No. 177 , with rs57344483 as lead SNP.

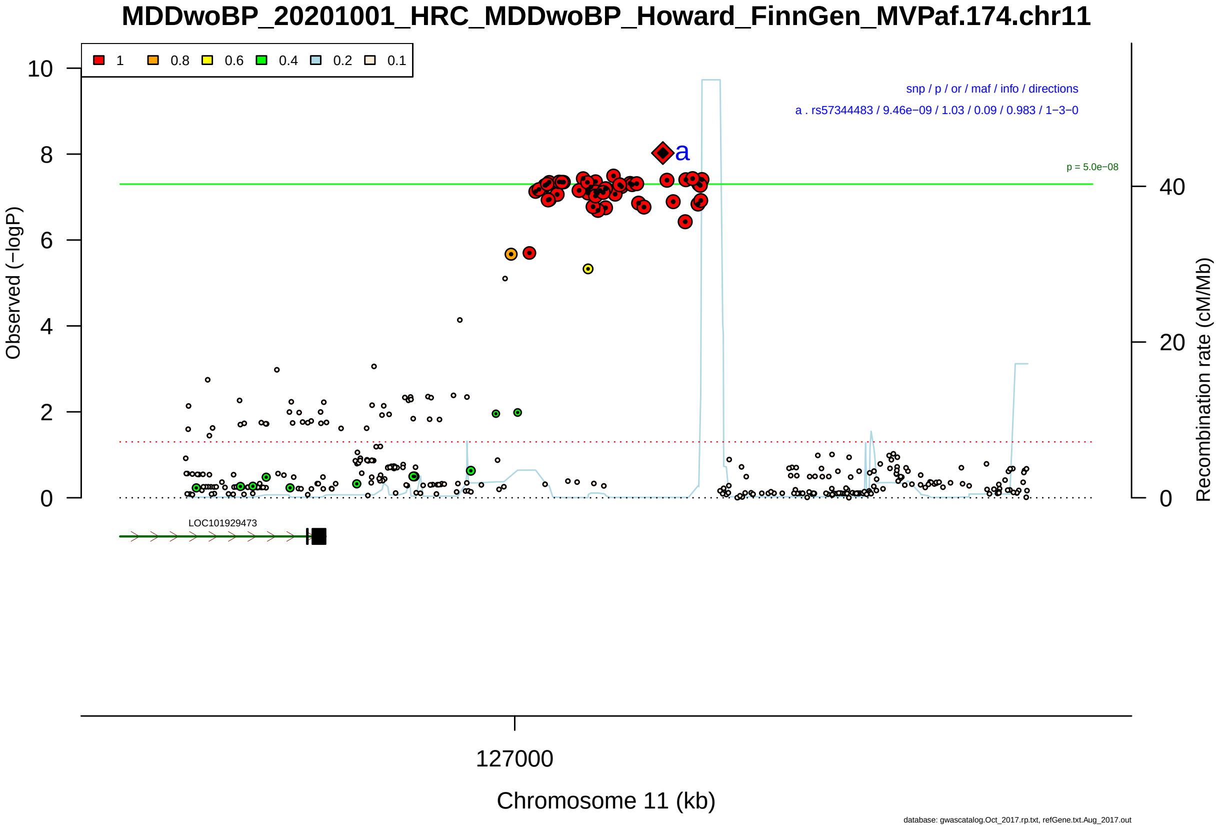

Figure S2-178: Regional plot of GWAS locus No. 178 , with rs10745389 as lead SNP.

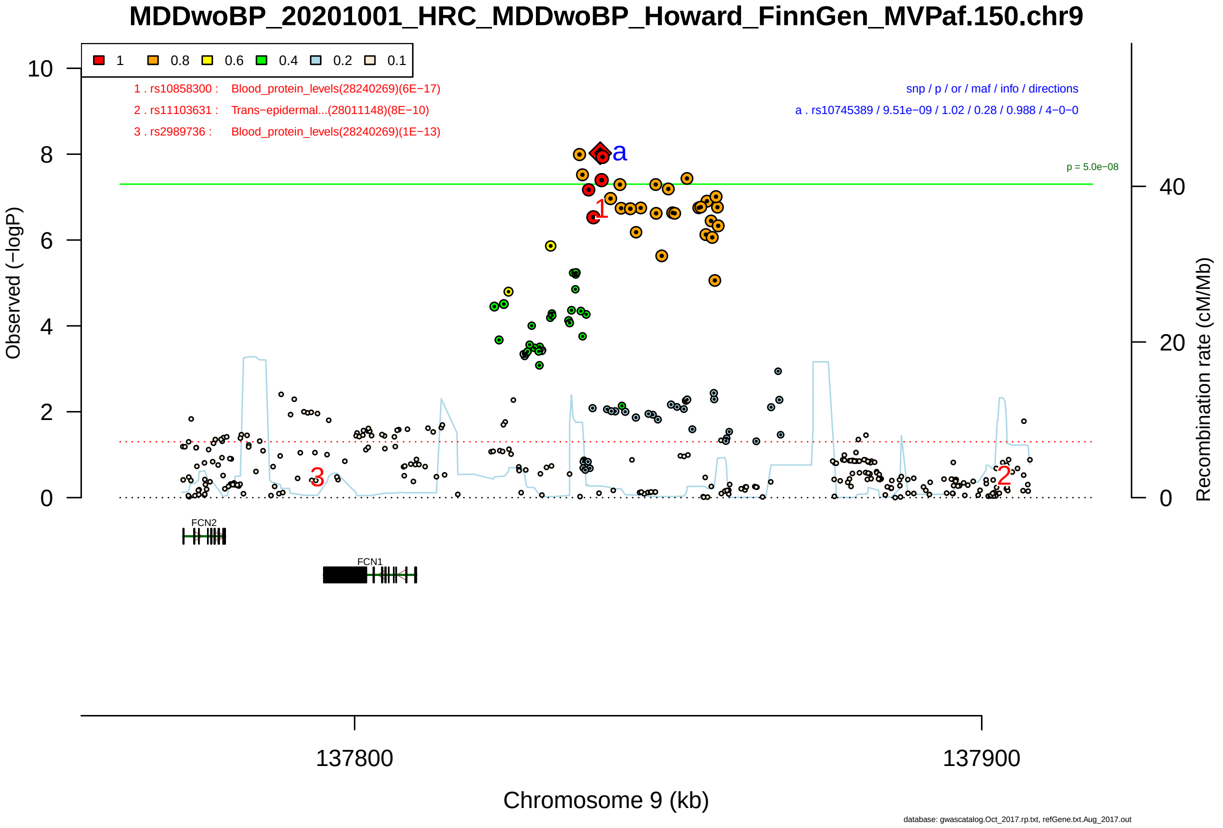

Figure S2-179: Regional plot of GWAS locus No. 179 , with rs57691884 as lead SNP.

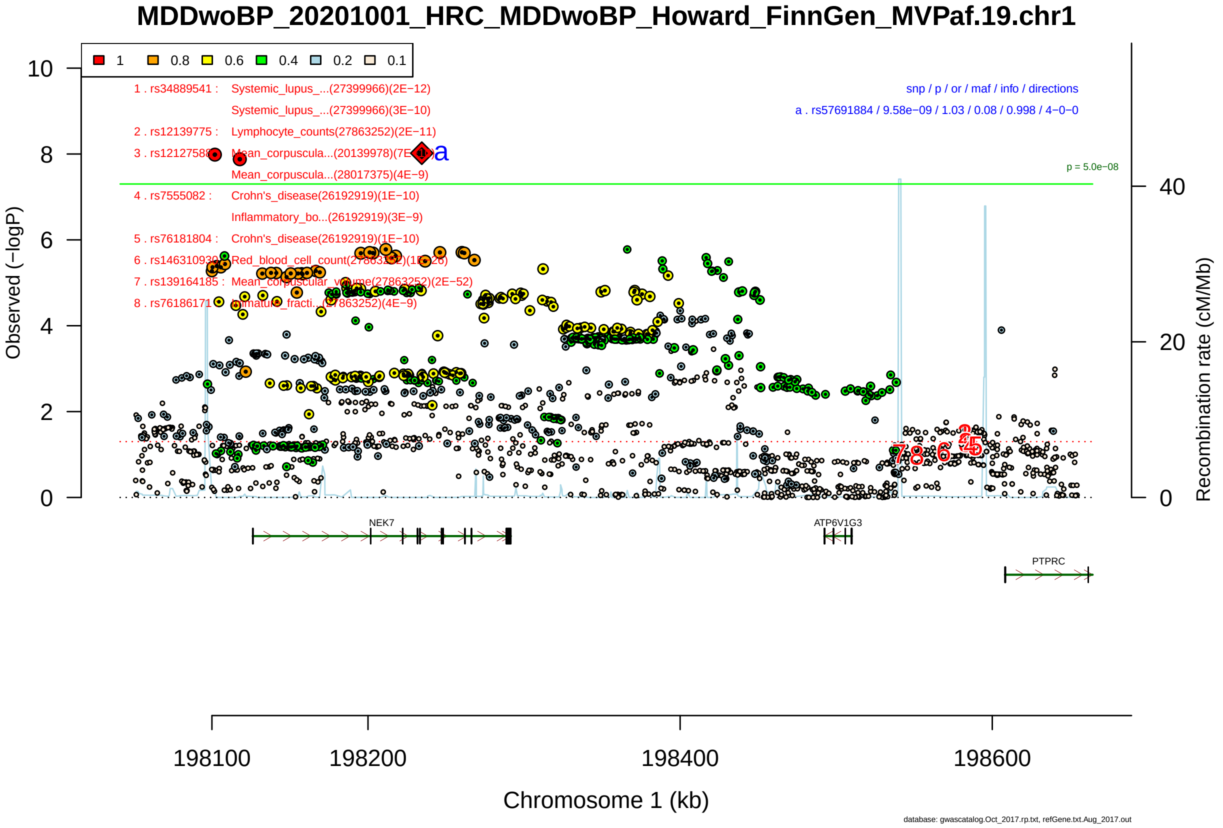

Figure S2-180: Regional plot of GWAS locus No. 180 , with rs12789028 as lead SNP.

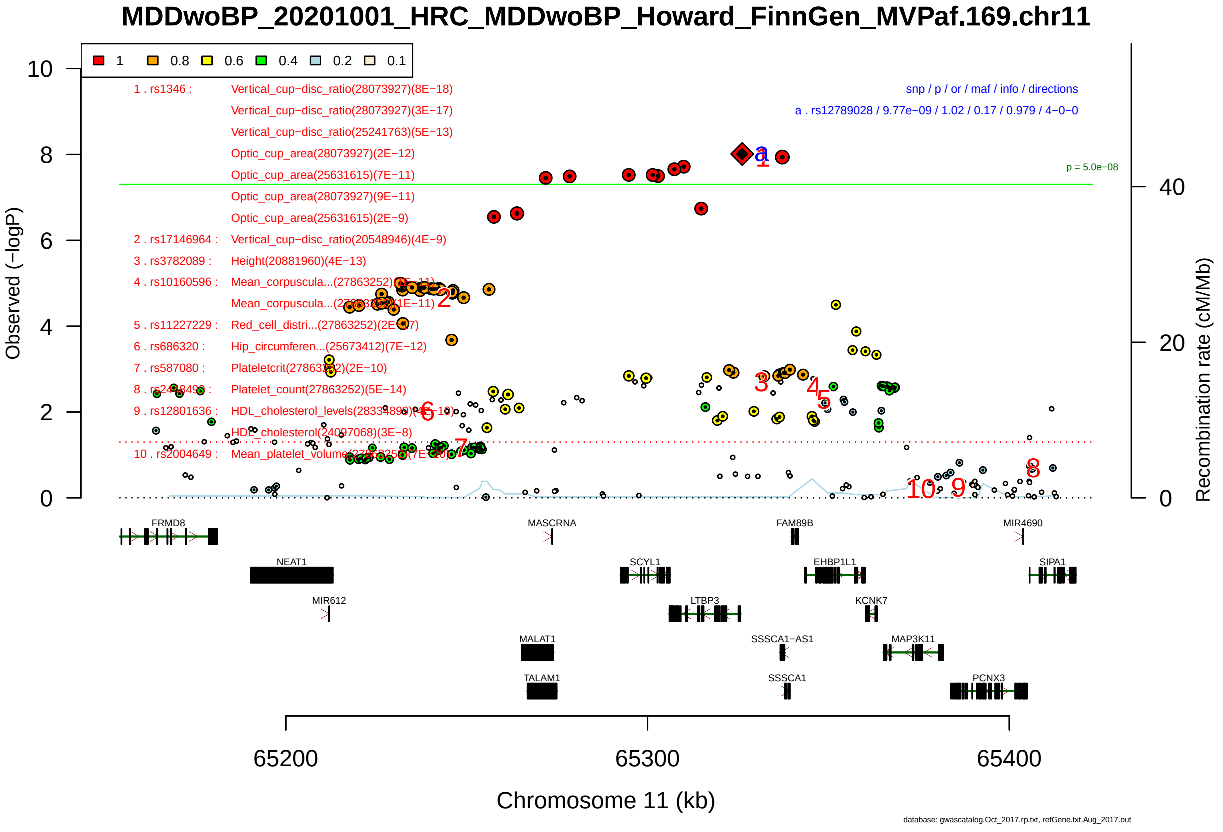

Figure S2-181: Regional plot of GWAS locus No. 181 , with rs34829058 as lead SNP.

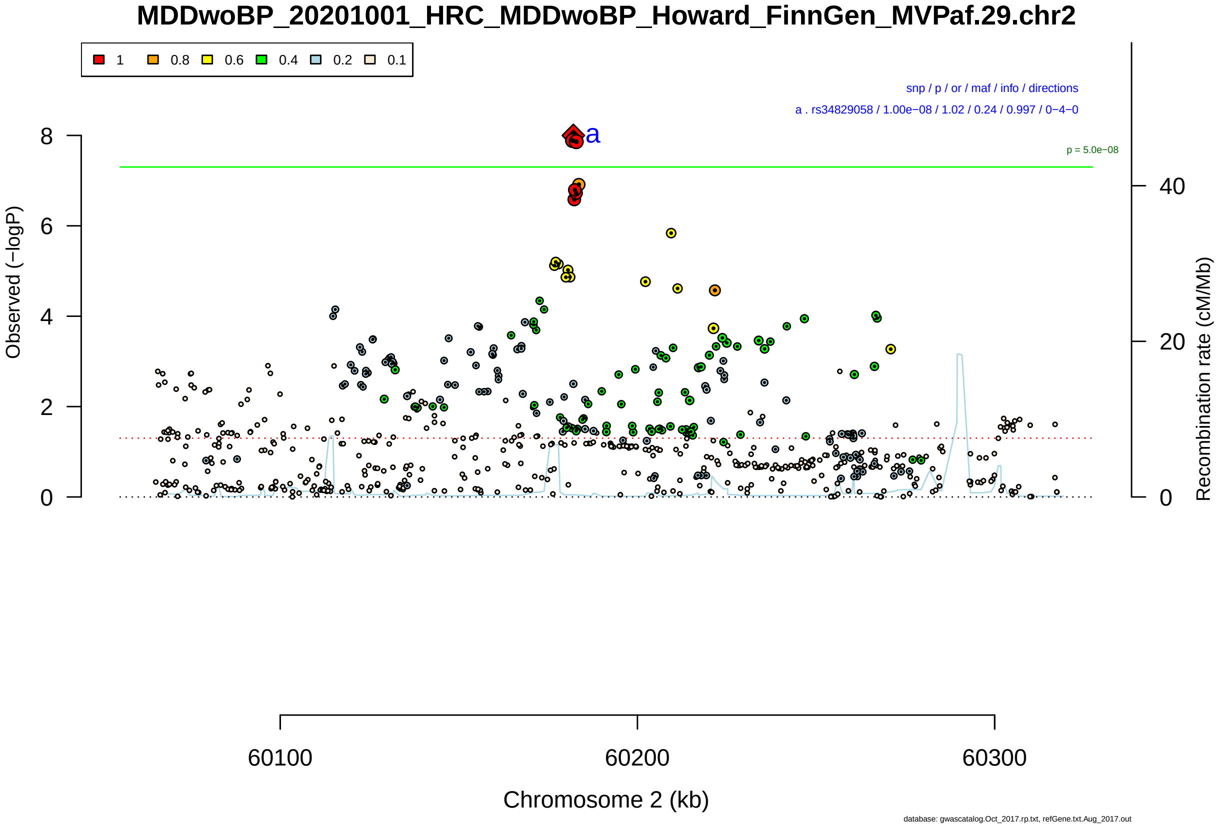

Figure S2-182: Regional plot of GWAS locus No. 182 , with rs330340 as lead SNP.

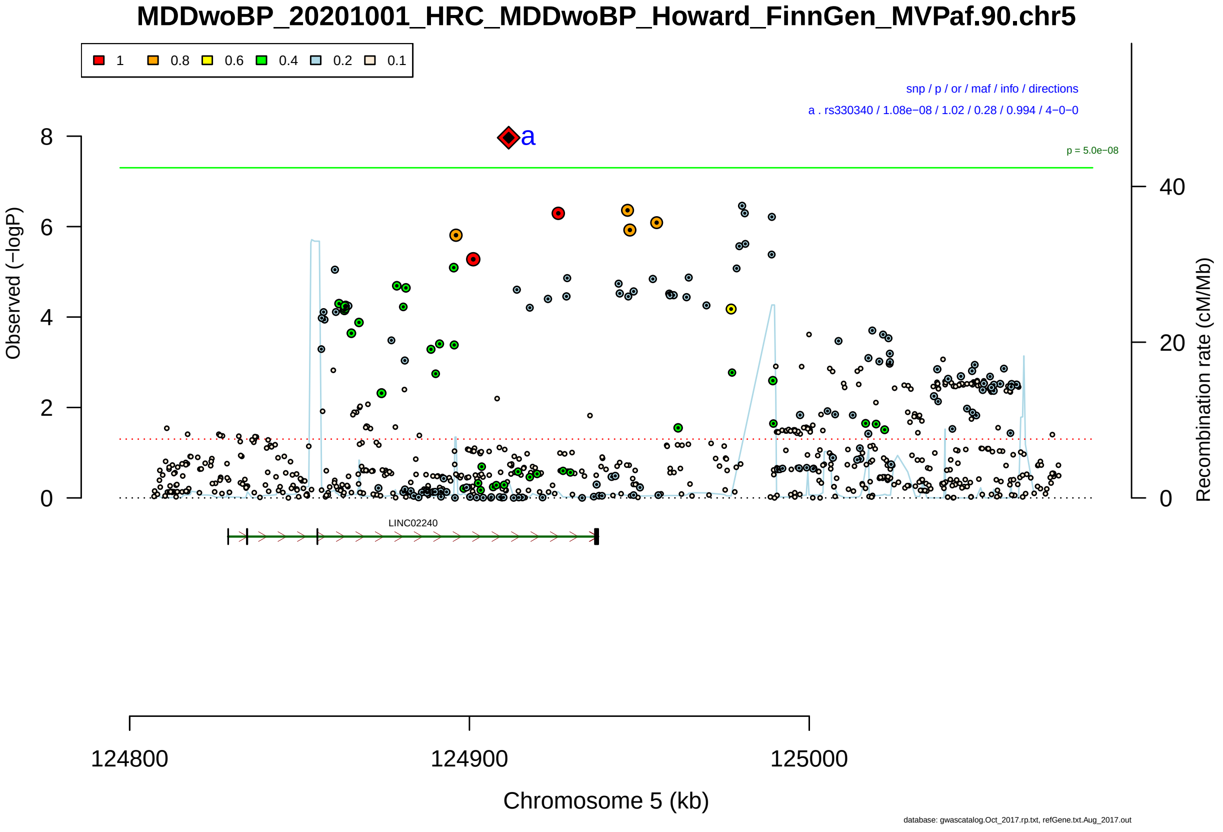

Figure S2-183: Regional plot of GWAS locus No. 183 , with rs11644513 as lead SNP.

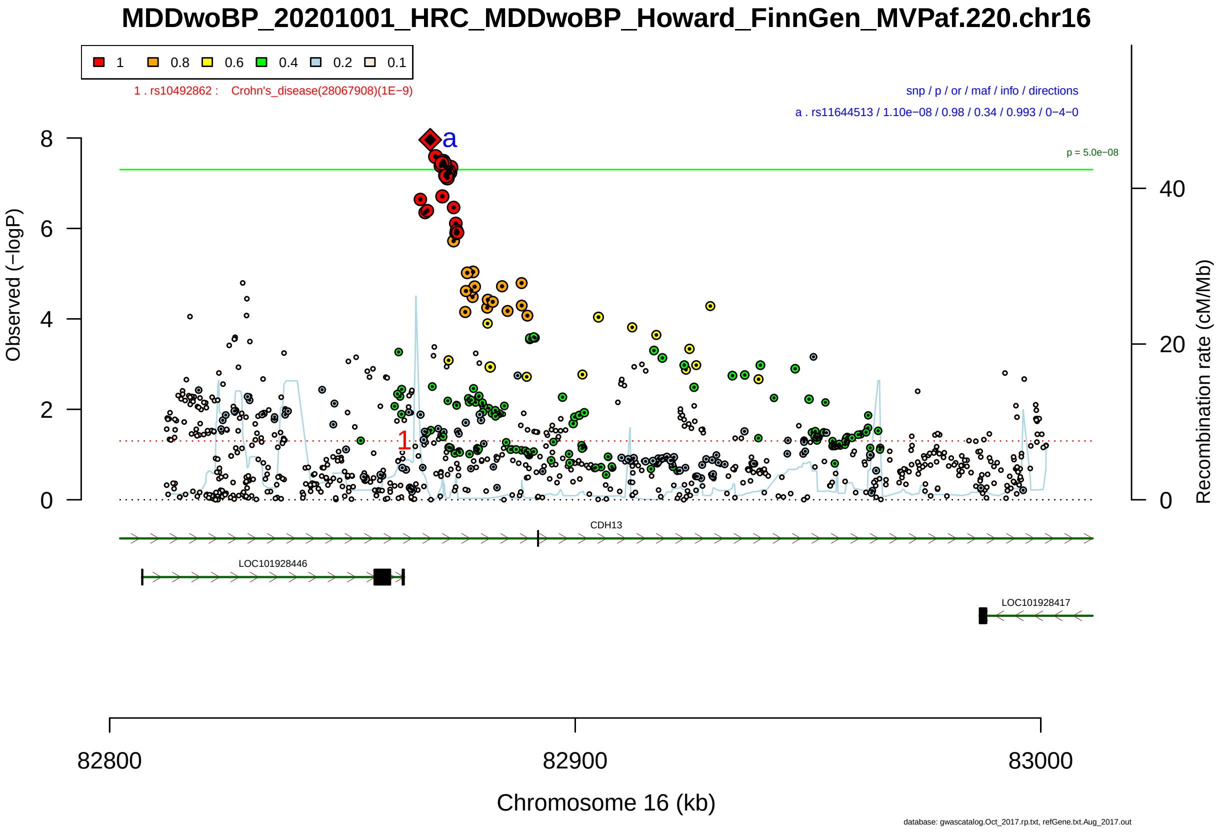

Figure S2-184: Regional plot of GWAS locus No. 184 , with rs12194317 as lead SNP.

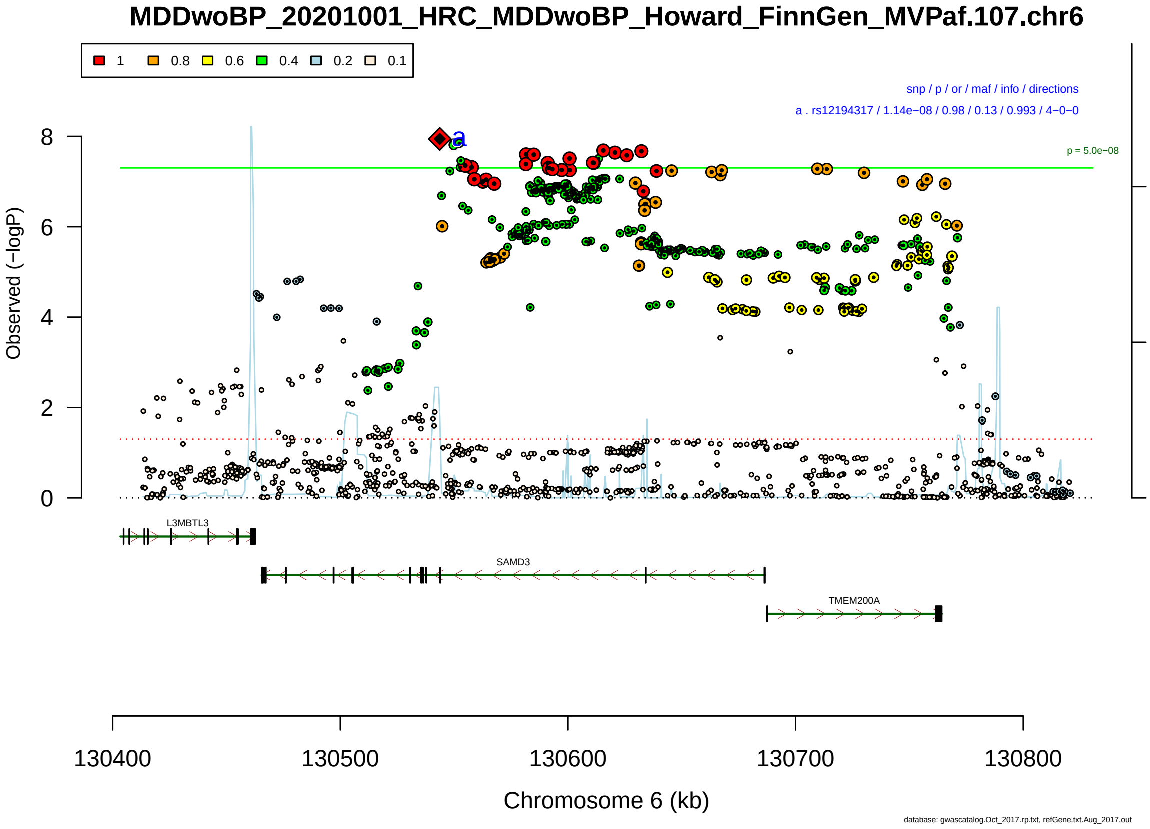

Figure S2-185: Regional plot of GWAS locus No. 185 , with rs6800583 as lead SNP.

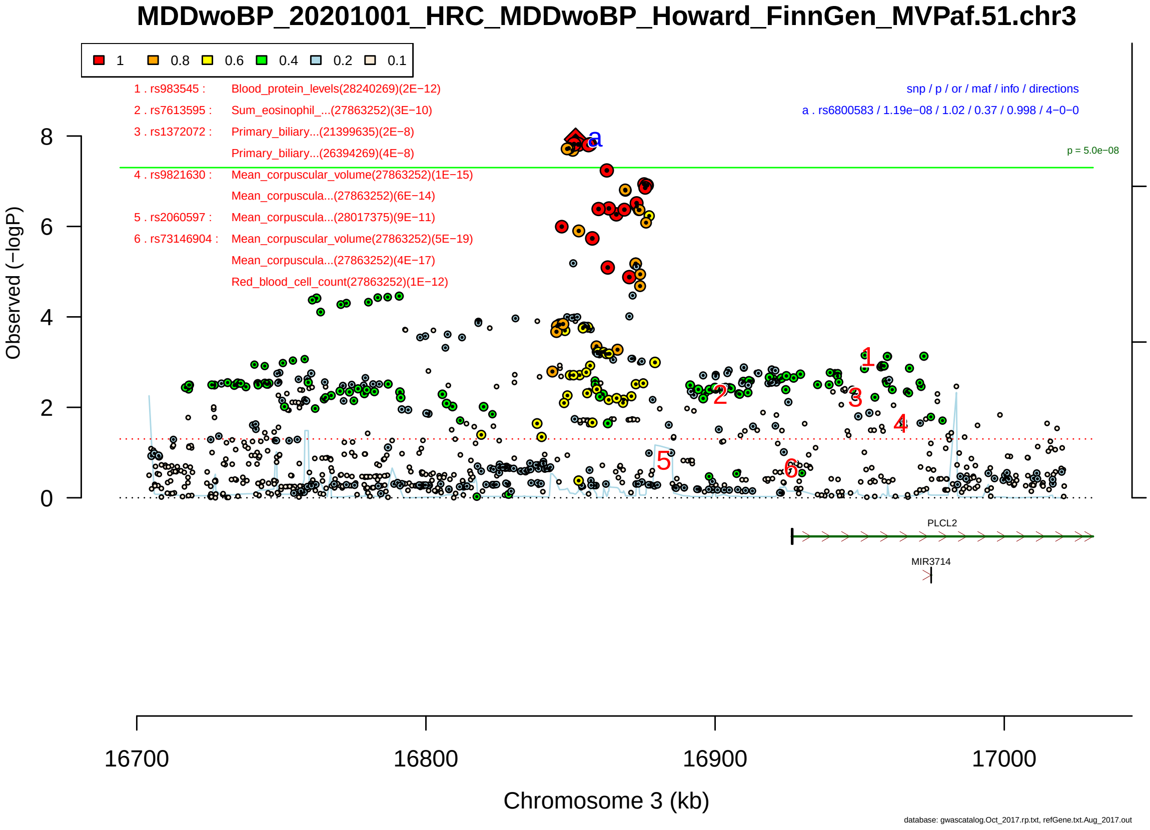

Figure S2-186: Regional plot of GWAS locus No. 186 , with rs7618264 as lead SNP.

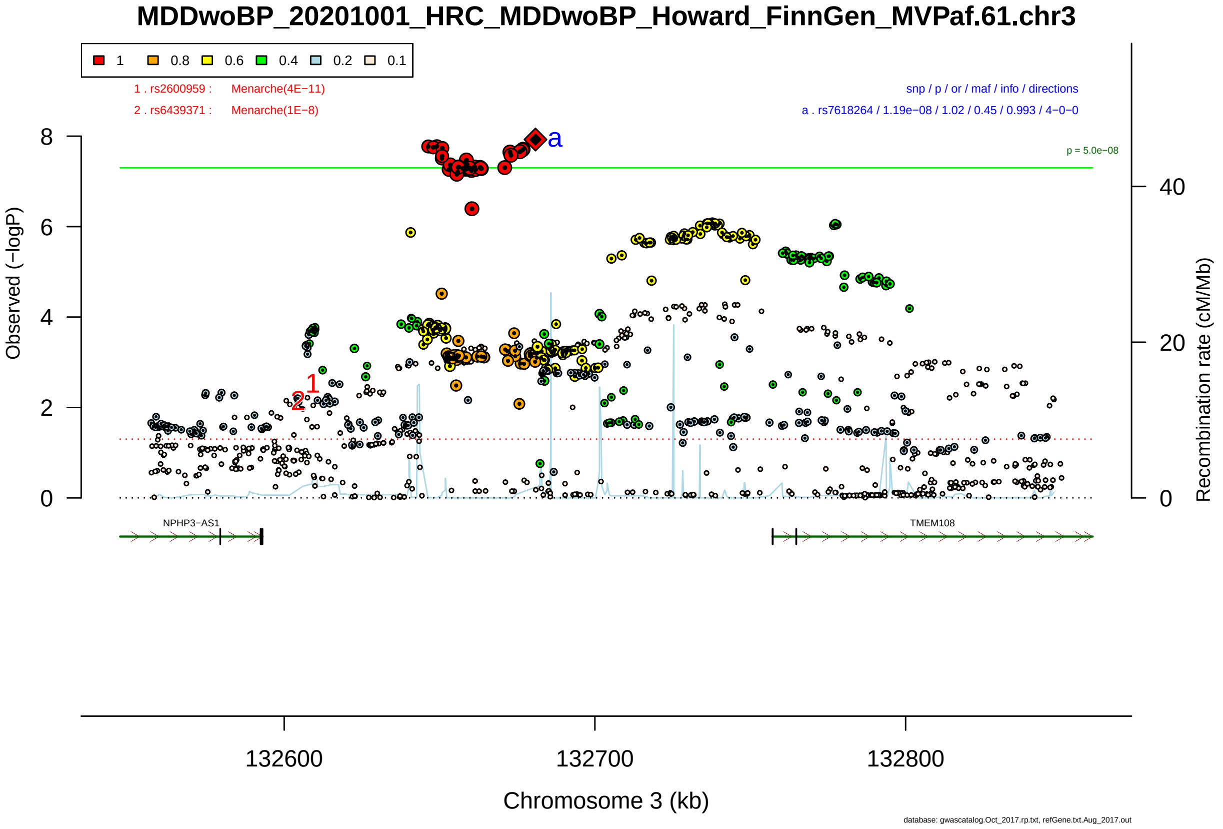

Figure S2-187: Regional plot of GWAS locus No. 187 , with rs61691222 as lead SNP.

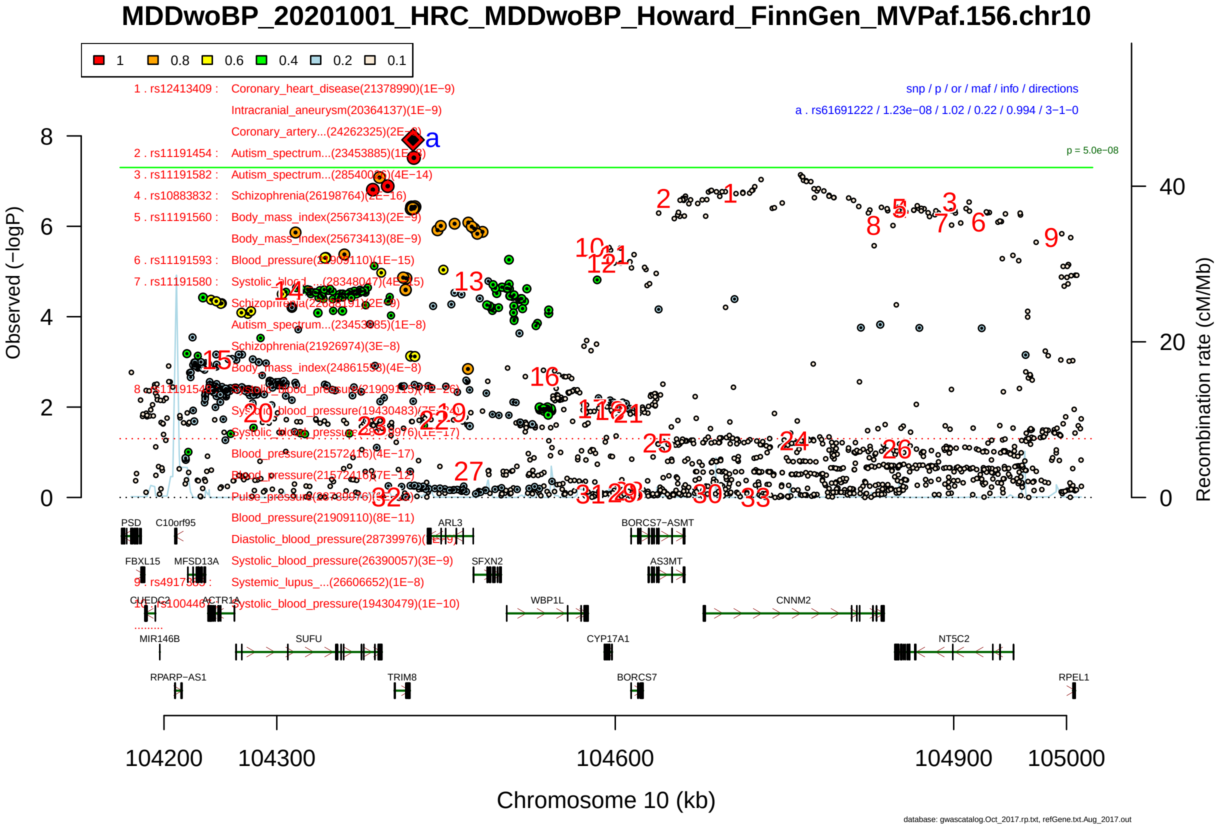

Figure S2-188: Regional plot of GWAS locus No. 188 , with rs116310555 as lead SNP.

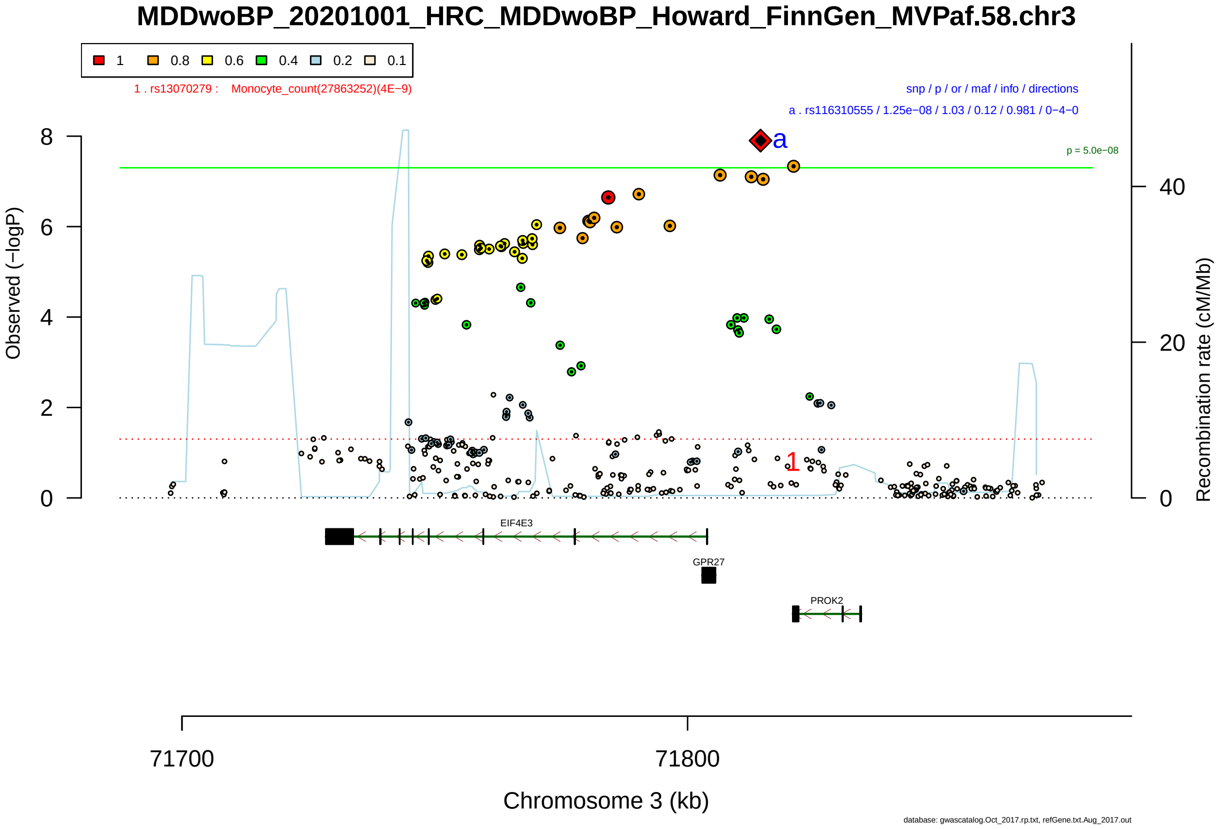

Figure S2-189: Regional plot of GWAS locus No. 189 , with rs111442587 as lead SNP.

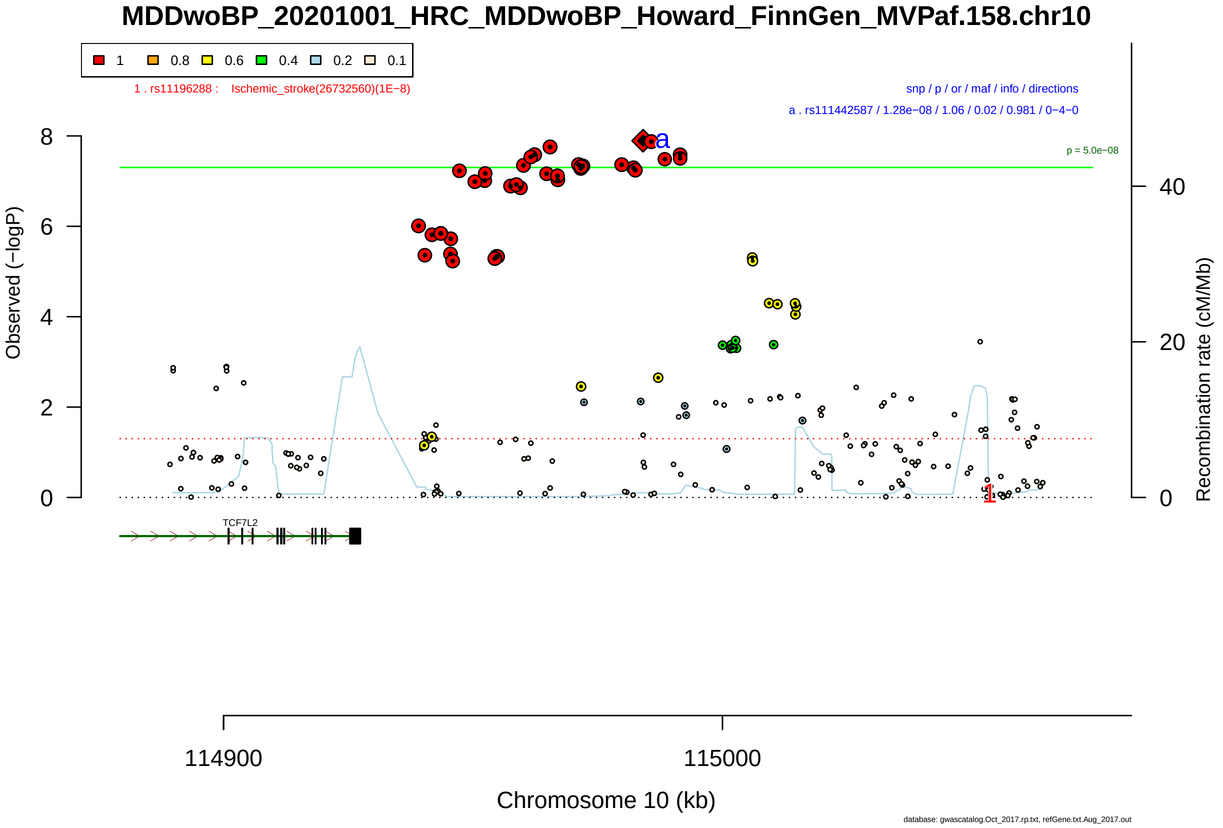

Figure S2-190: Regional plot of GWAS locus No. 190 , with rs657159 as lead SNP.

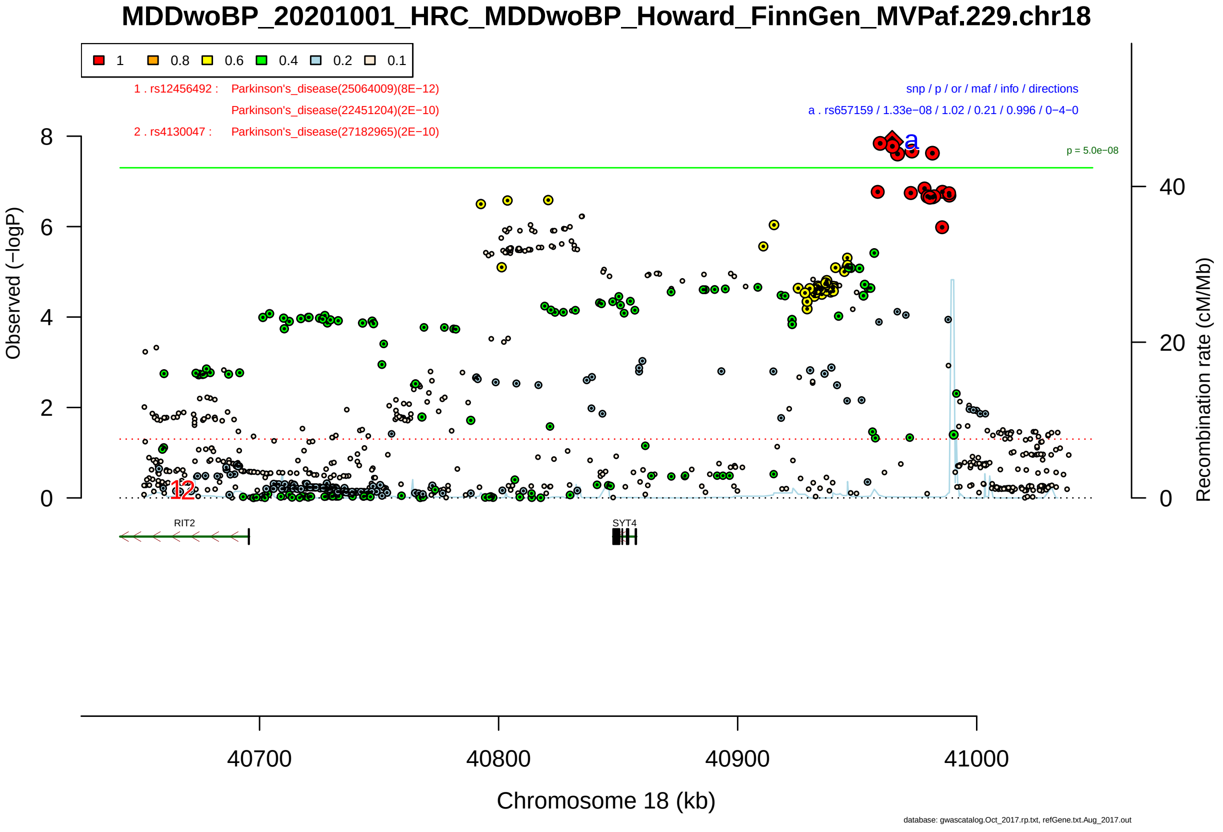

Figure S2-191: Regional plot of GWAS locus No. 191 , with rs13020607 as lead SNP.

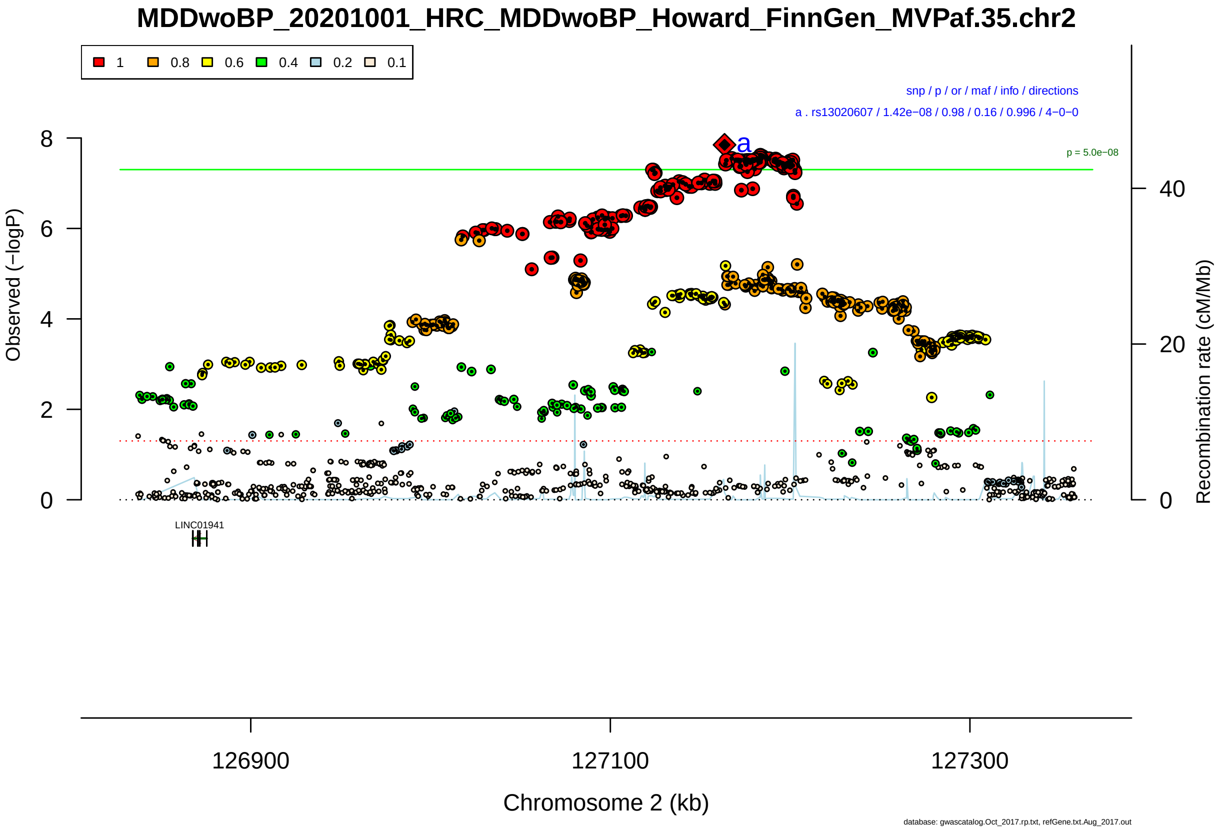

Figure S2-192: Regional plot of GWAS locus No. 192 , with rs7700453 as lead SNP.

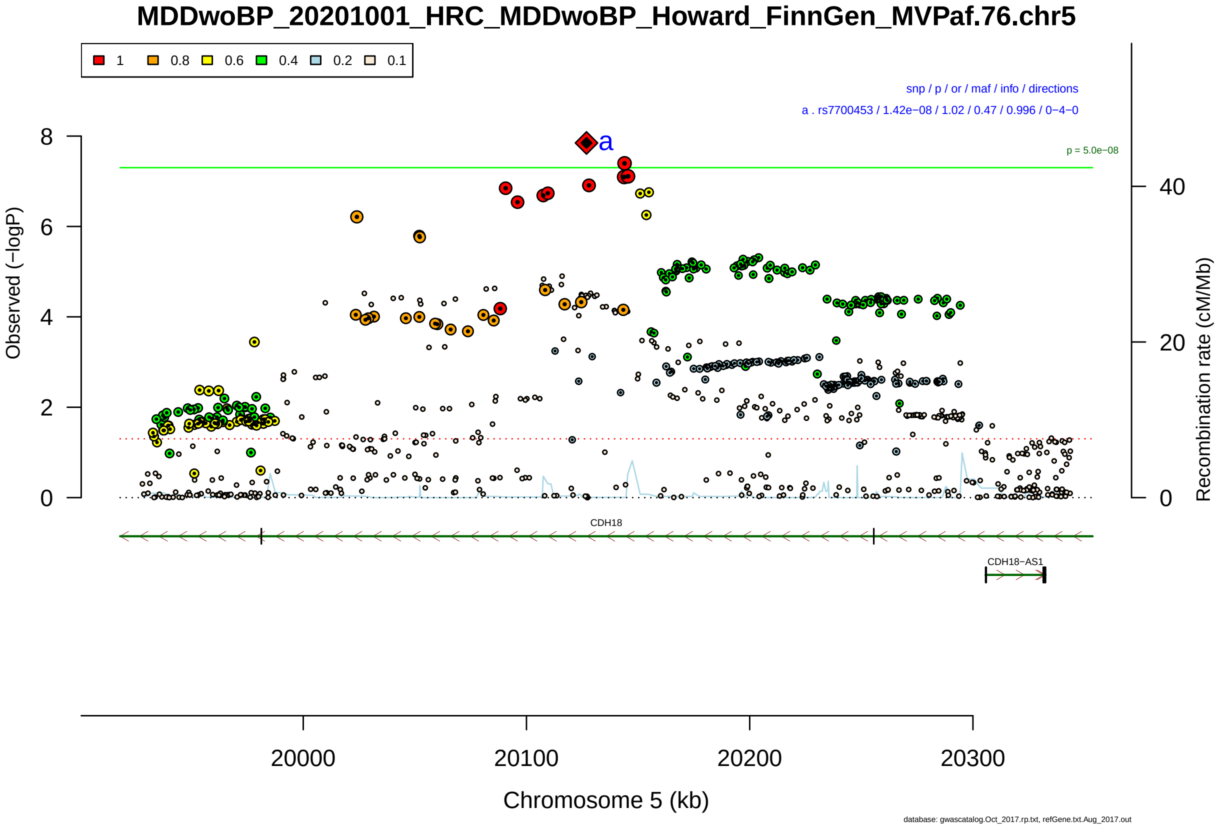

Figure S2-193: Regional plot of GWAS locus No. 193 , with rs57389877 as lead SNP.

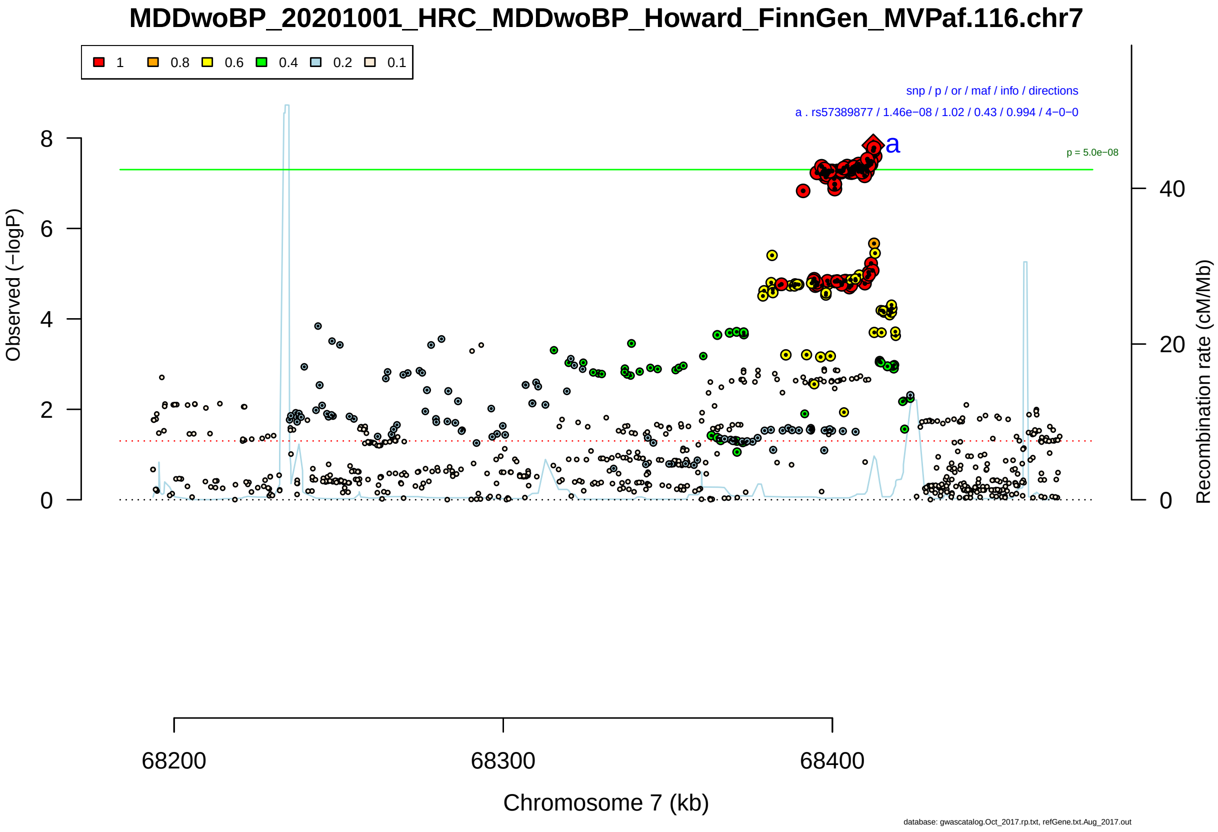

Figure S2-194: Regional plot of GWAS locus No. 194 , with rs7101595 as lead SNP.

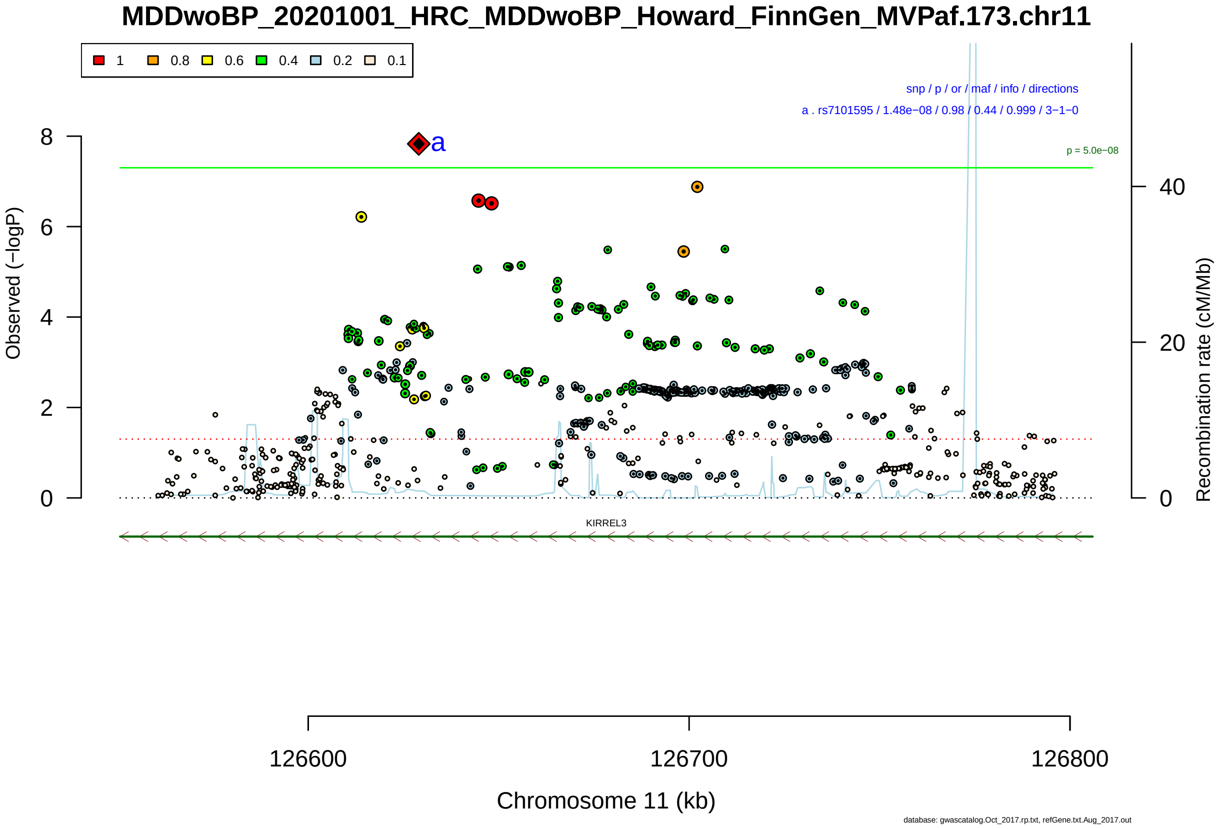

Figure S2-195: Regional plot of GWAS locus No. 195 , with rs4852252 as lead SNP.

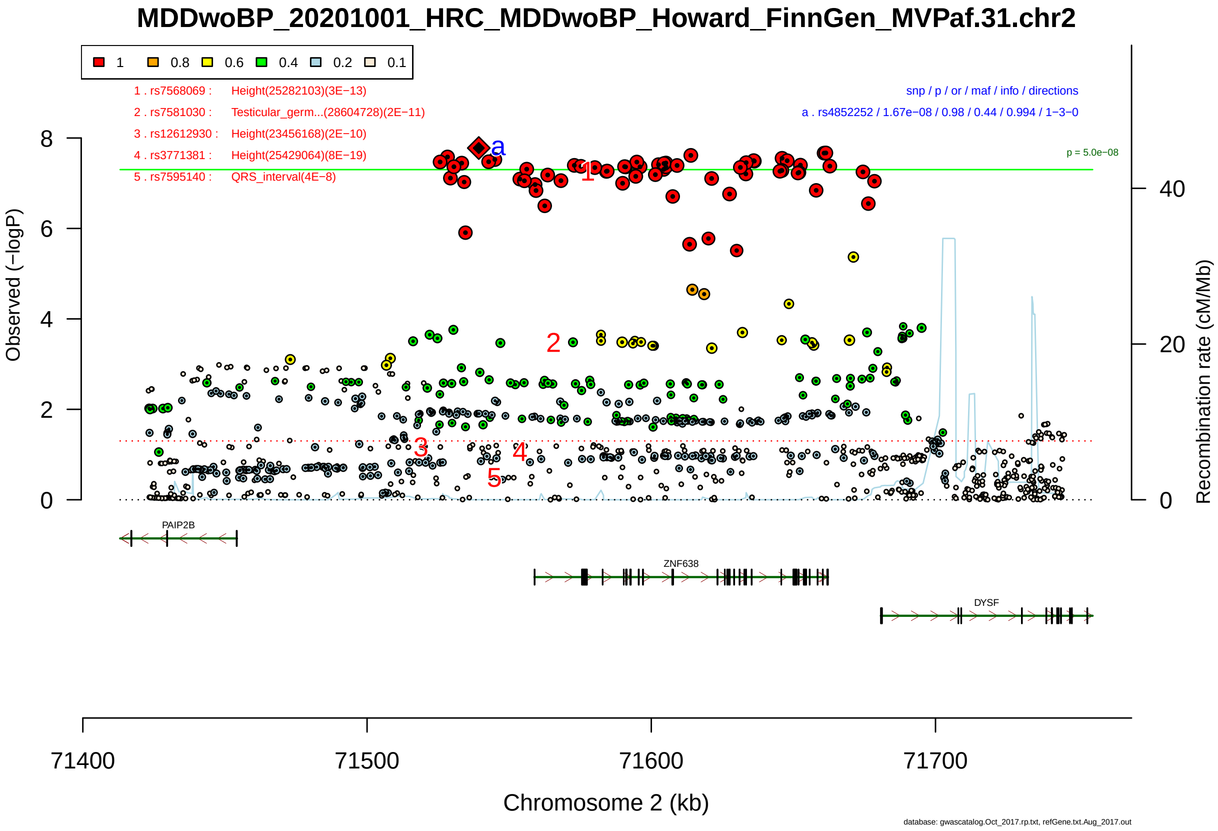

Figure S2-196: Regional plot of GWAS locus No. 196 , with rs7534271 as lead SNP.

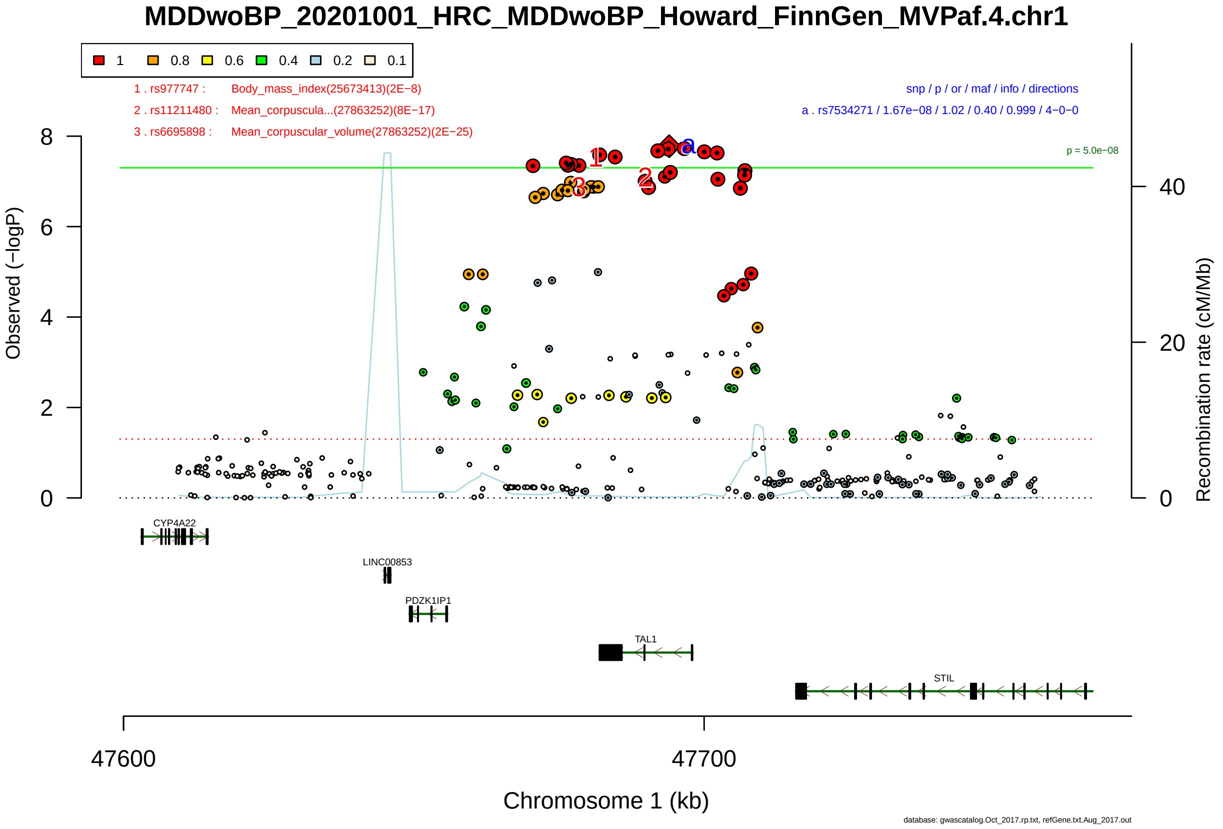

Figure S2-197: Regional plot of GWAS locus No. 197 , with rs4671458 as lead SNP.

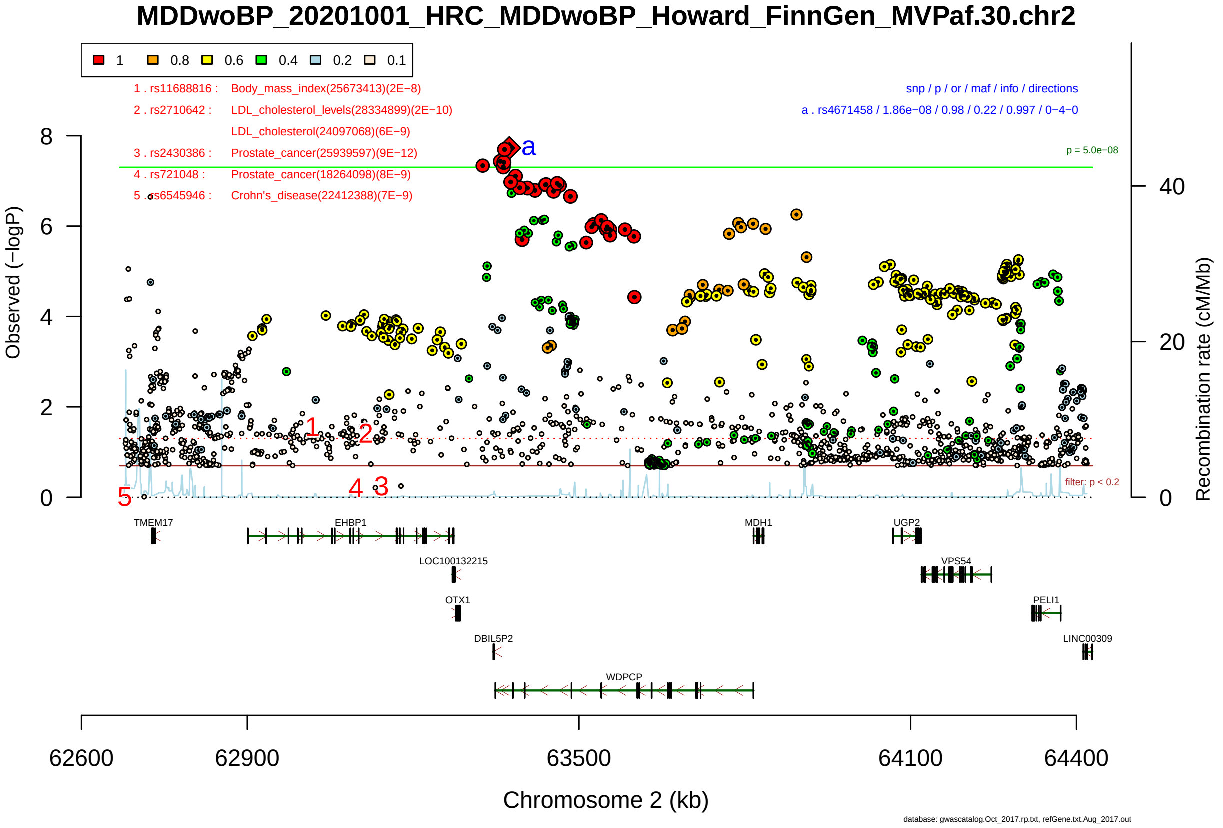

Figure S2-198: Regional plot of GWAS locus No. 198 , with rs1597076 as lead SNP.

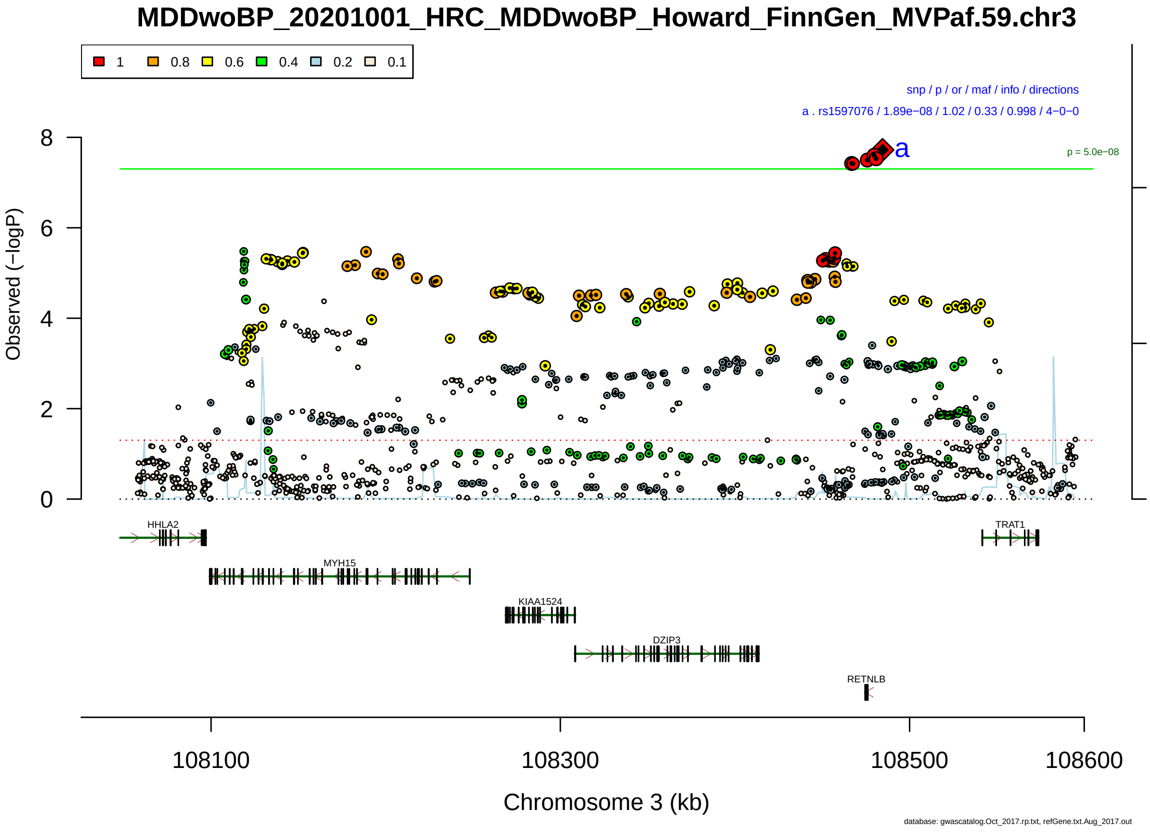

Figure S2-199: Regional plot of GWAS locus No. 199 , with rs1893562 as lead SNP.

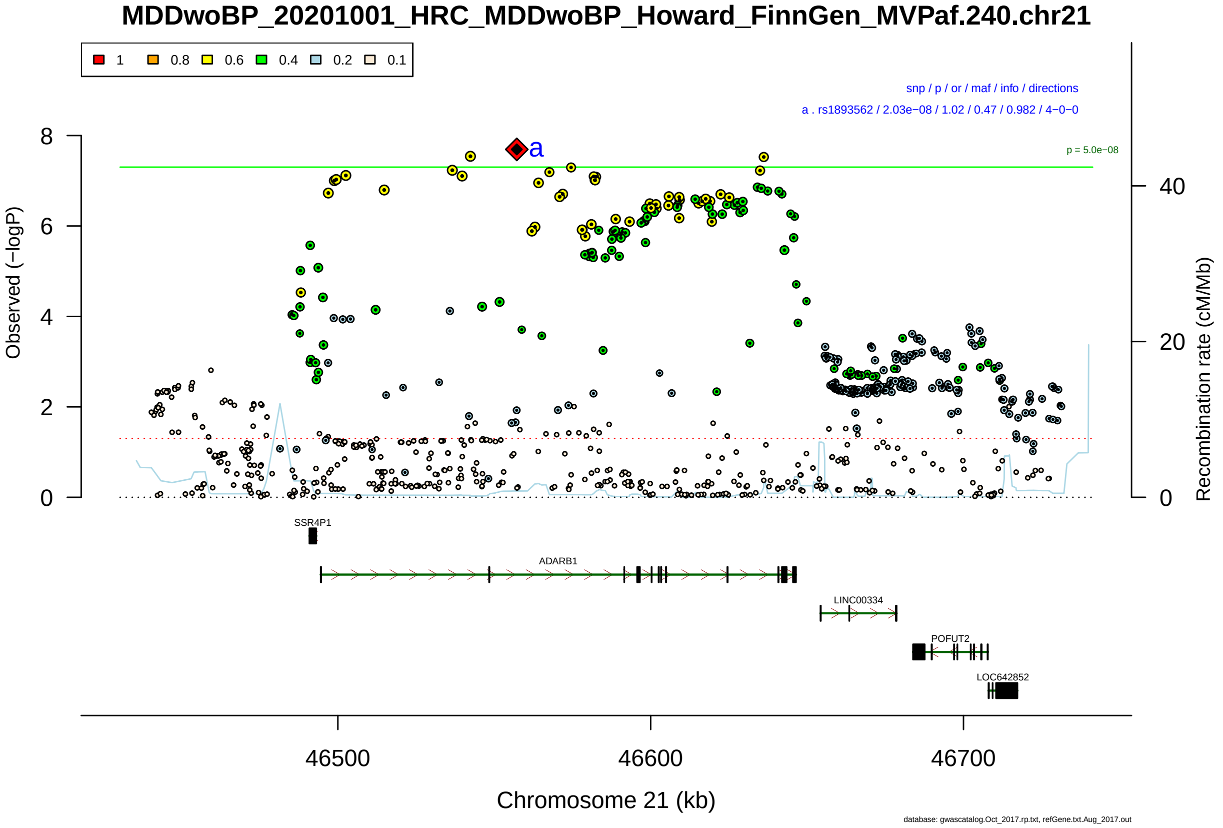

Figure S2-200: Regional plot of GWAS locus No. 200 , with rs1466887 as lead SNP.

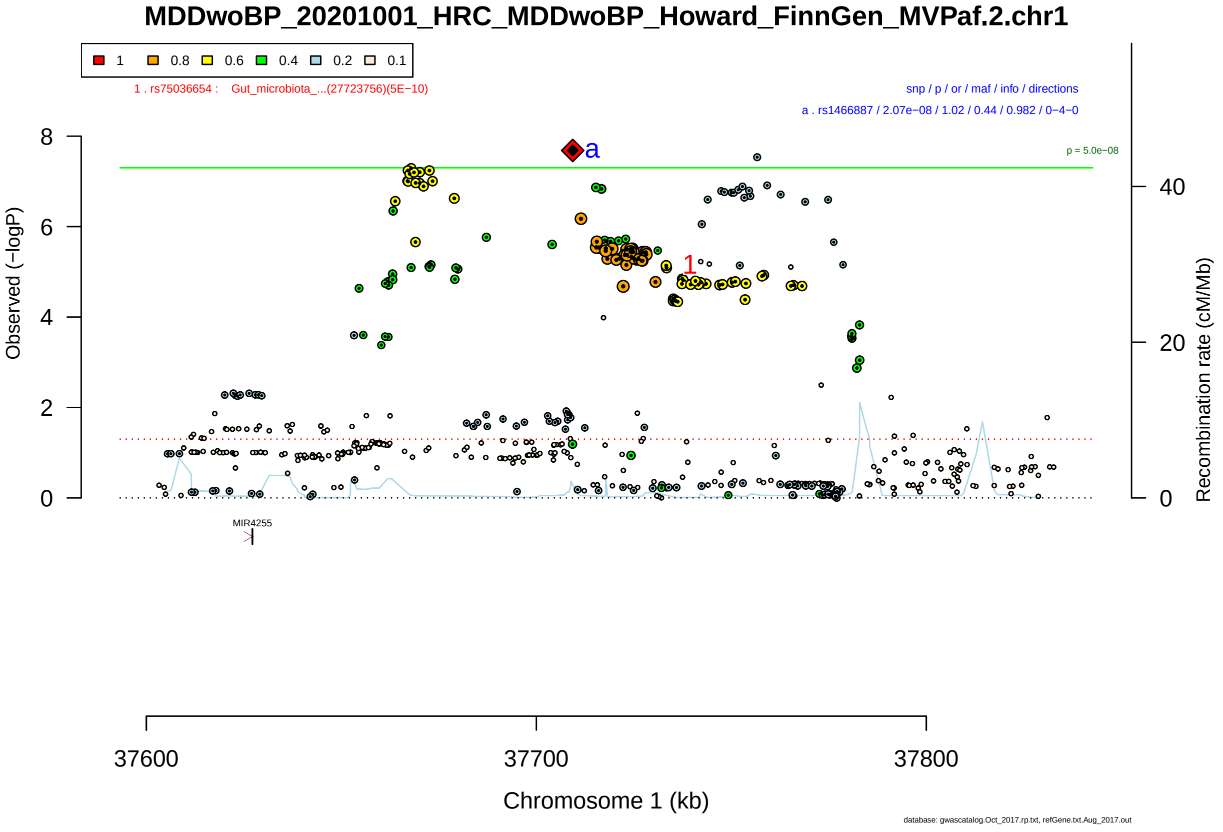

Figure S2-201: Regional plot of GWAS locus No. 201 , with rs4531131 as lead SNP.

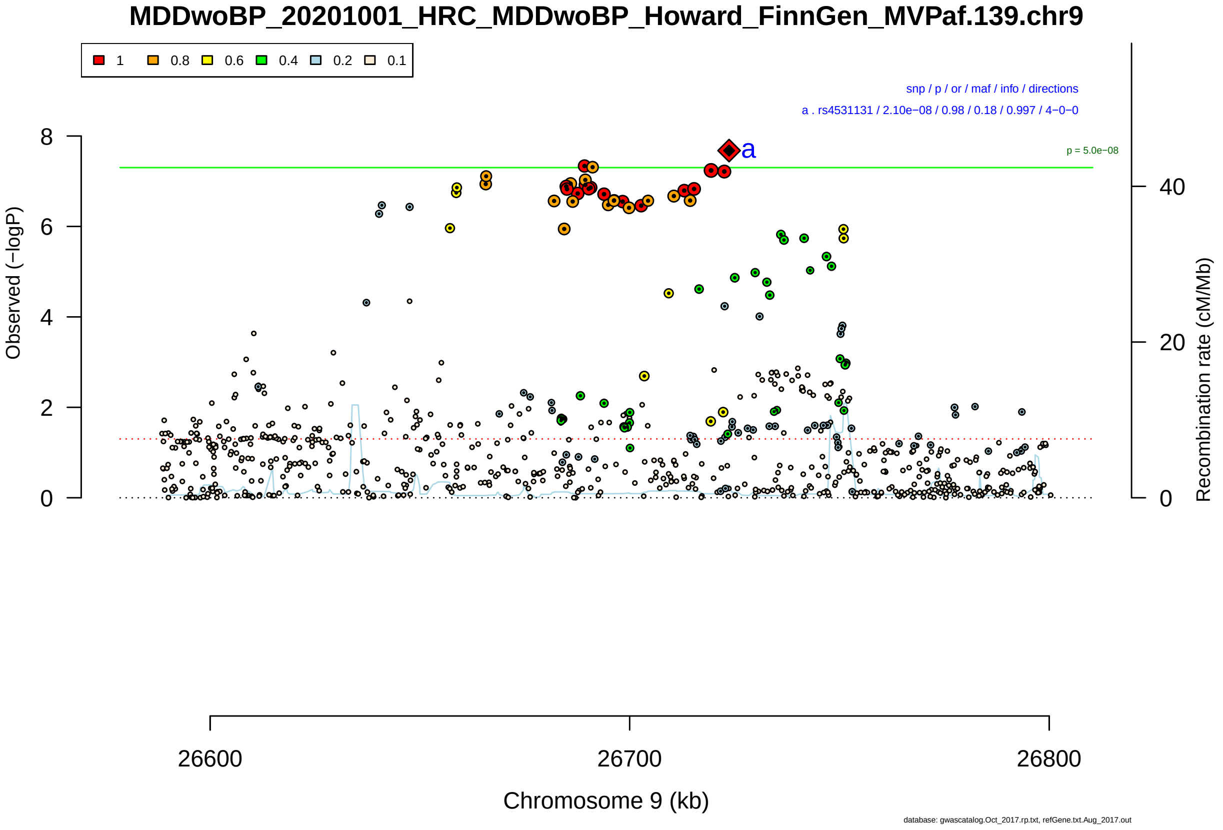

Figure S2-202: Regional plot of GWAS locus No. 202 , with rs1560195 as lead SNP.

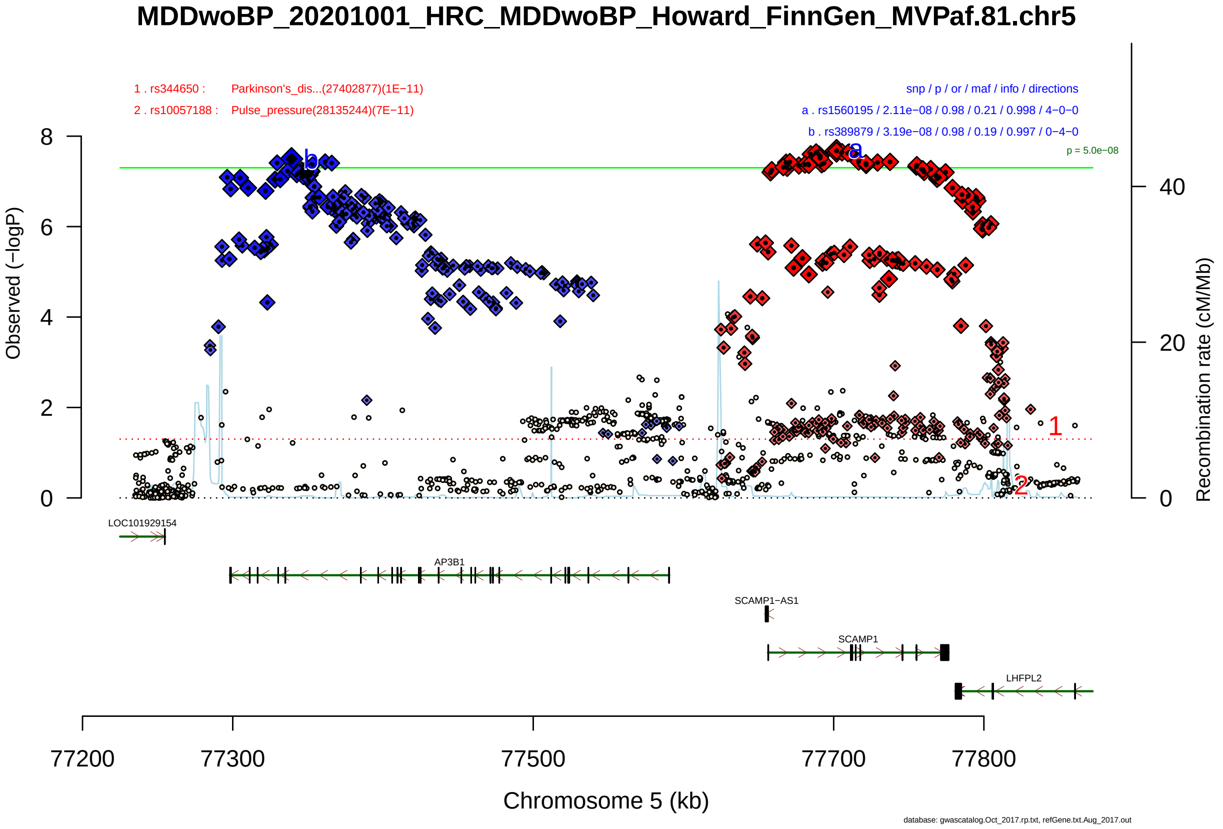

Figure S2-203: Regional plot of GWAS locus No. 203 , with rs169235 as lead SNP.

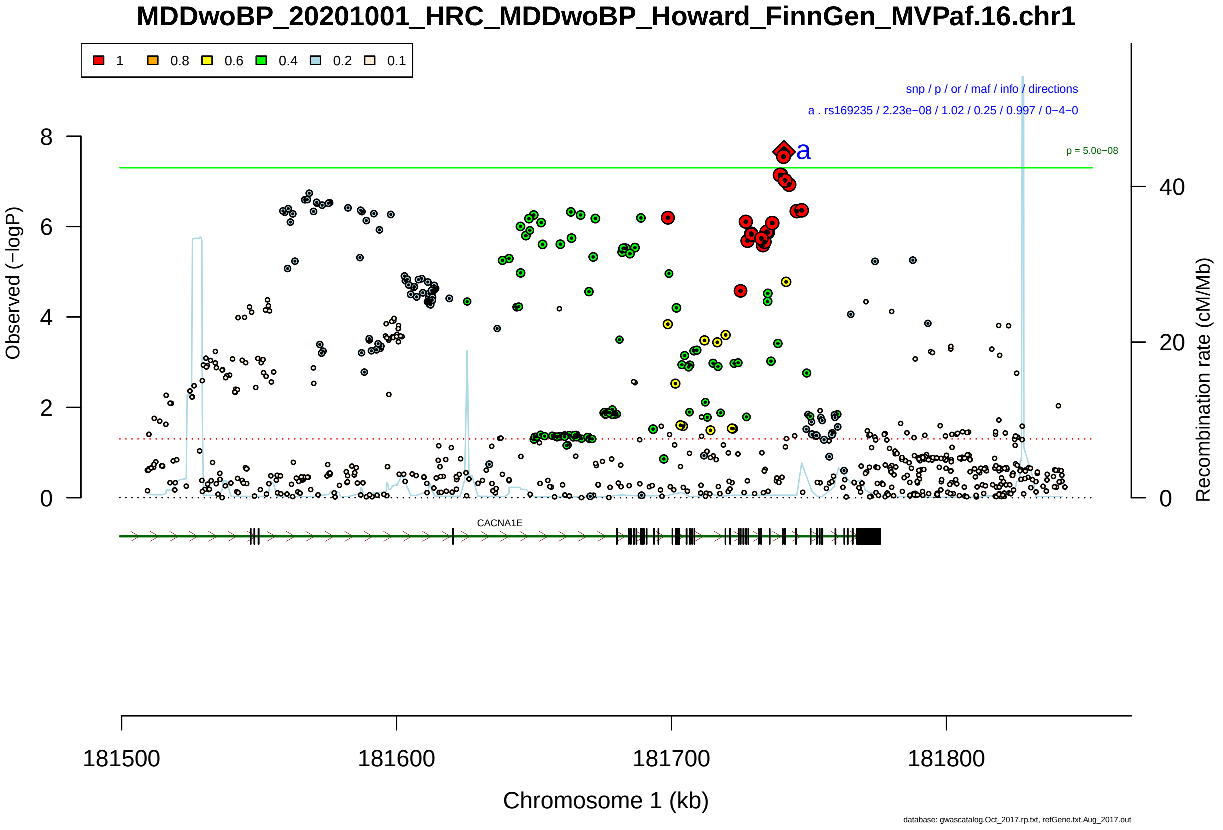

Figure S2-204: Regional plot of GWAS locus No. 204 , with rs59859947 as lead SNP.

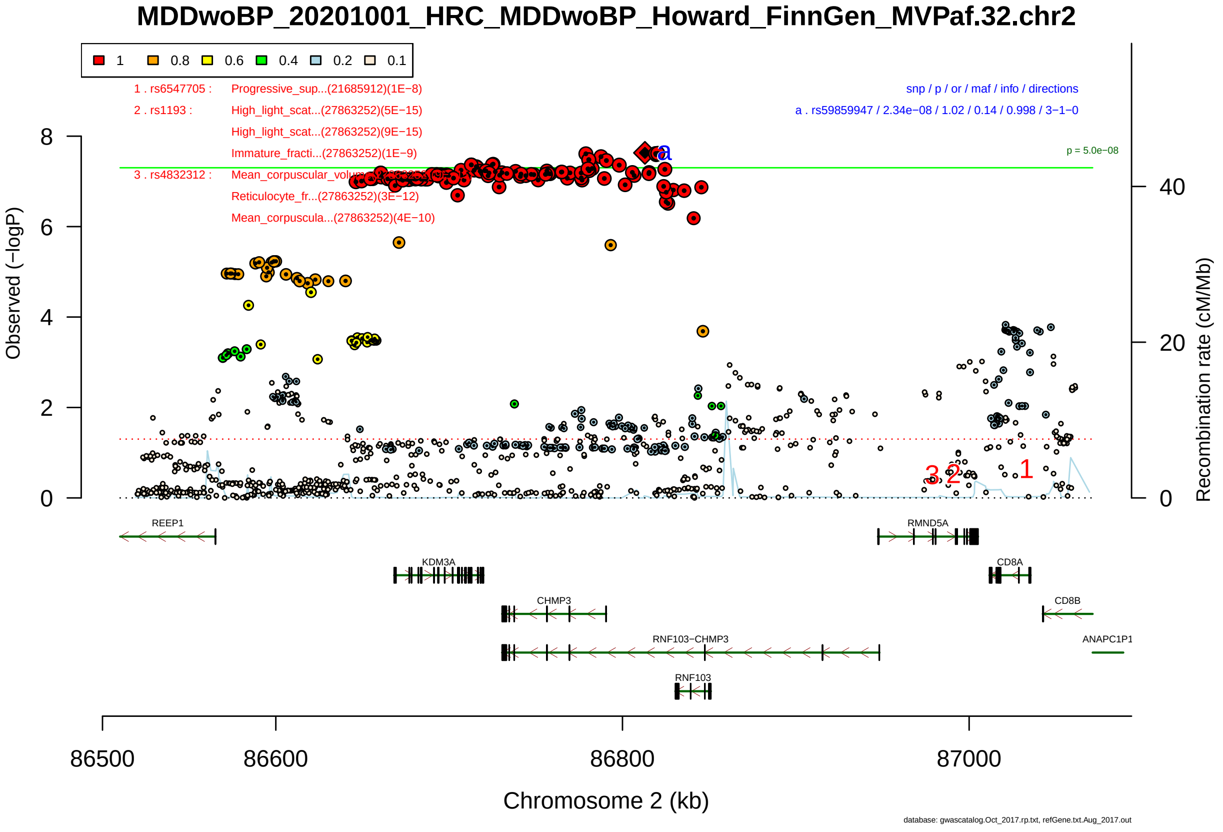

Figure S2-205: Regional plot of GWAS locus No. 205 , with rs521048 as lead SNP.

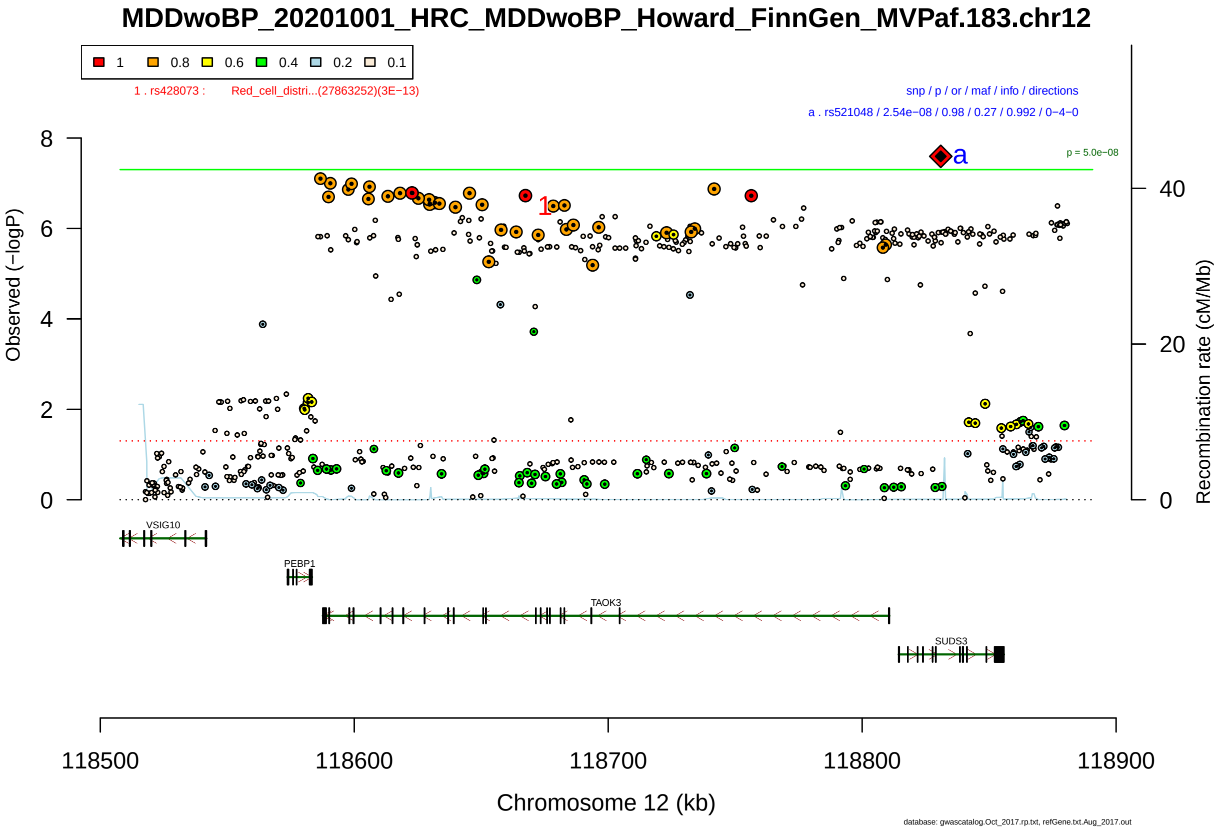

Figure S2-206: Regional plot of GWAS locus No. 206 , with rs6480132 as lead SNP.

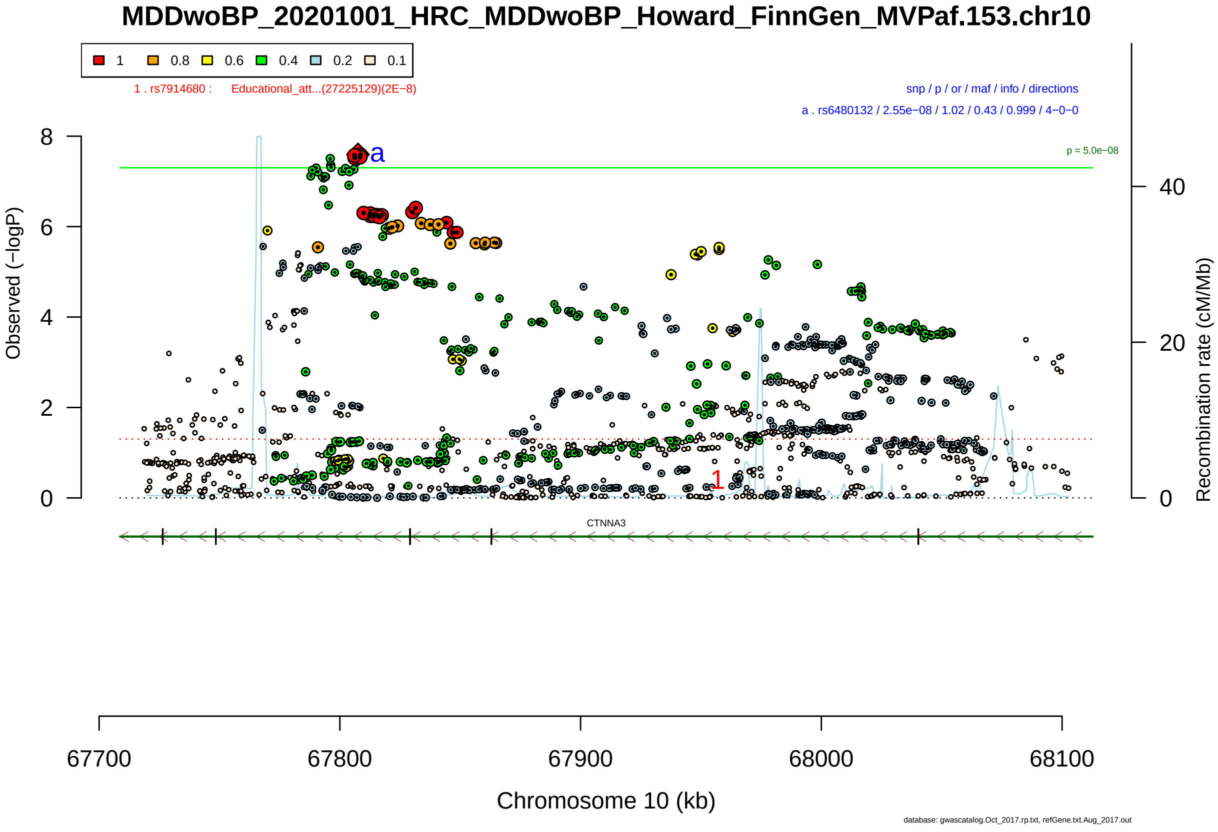

Figure S2-207: Regional plot of GWAS locus No. 207 , with rs4663414 as lead SNP.

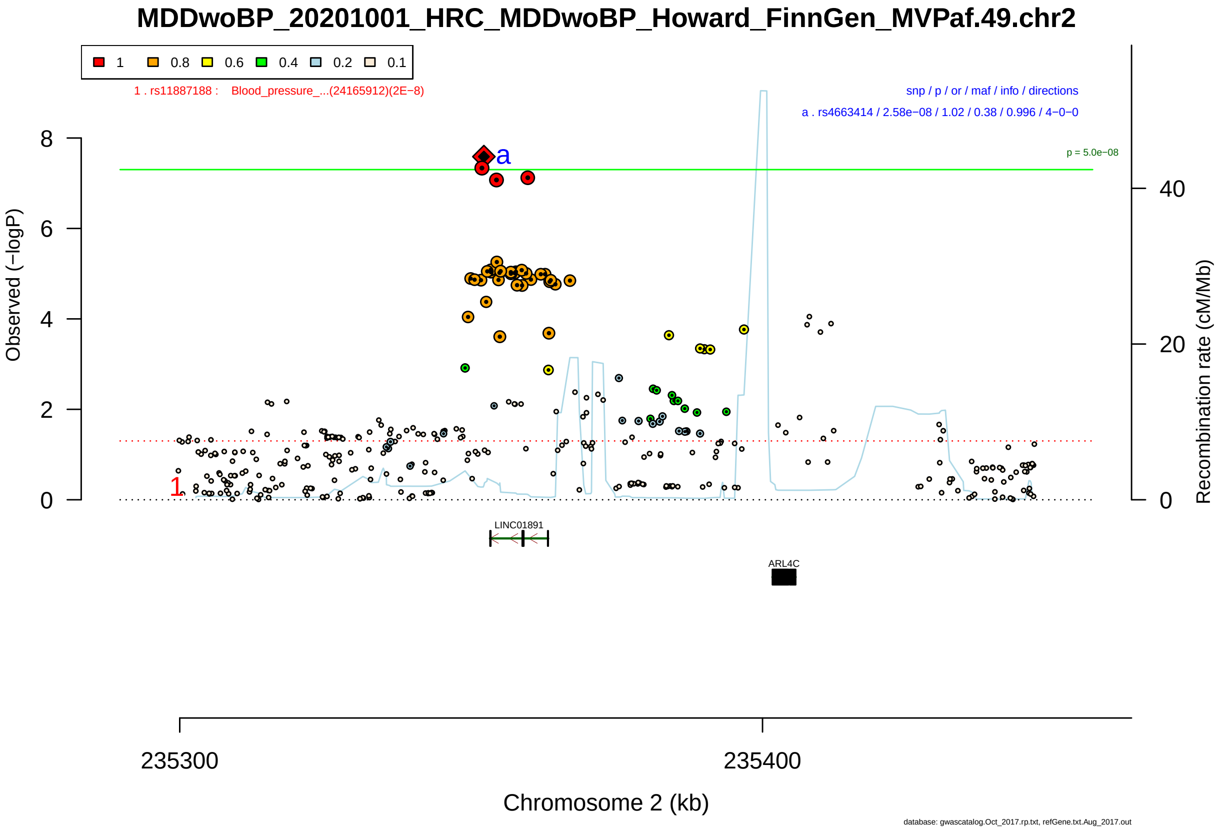

Figure S2-208: Regional plot of GWAS locus No. 208 , with rs2292996 as lead SNP.

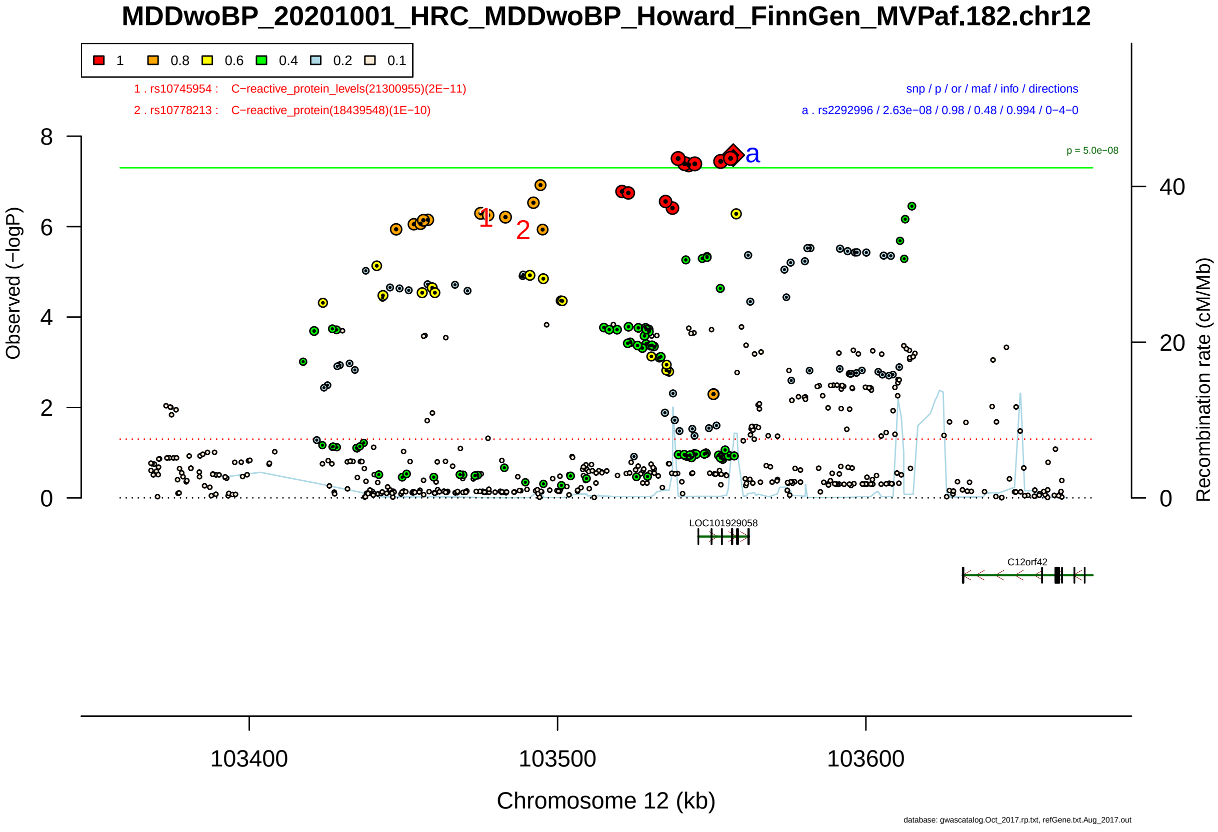

Figure S2-209: Regional plot of GWAS locus No. 209 , with rs9306914 as lead SNP.

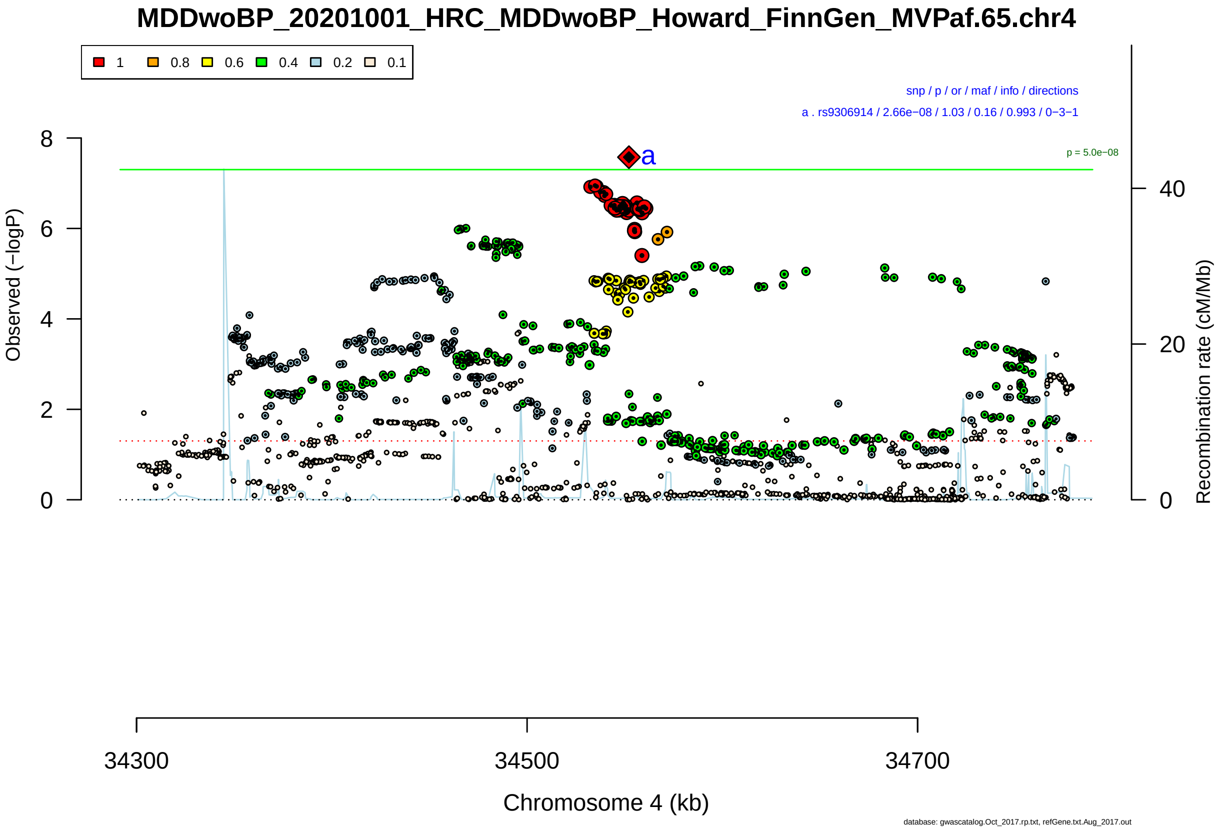

Figure S2-210: Regional plot of GWAS locus No. 210 , with rs908670 as lead SNP.

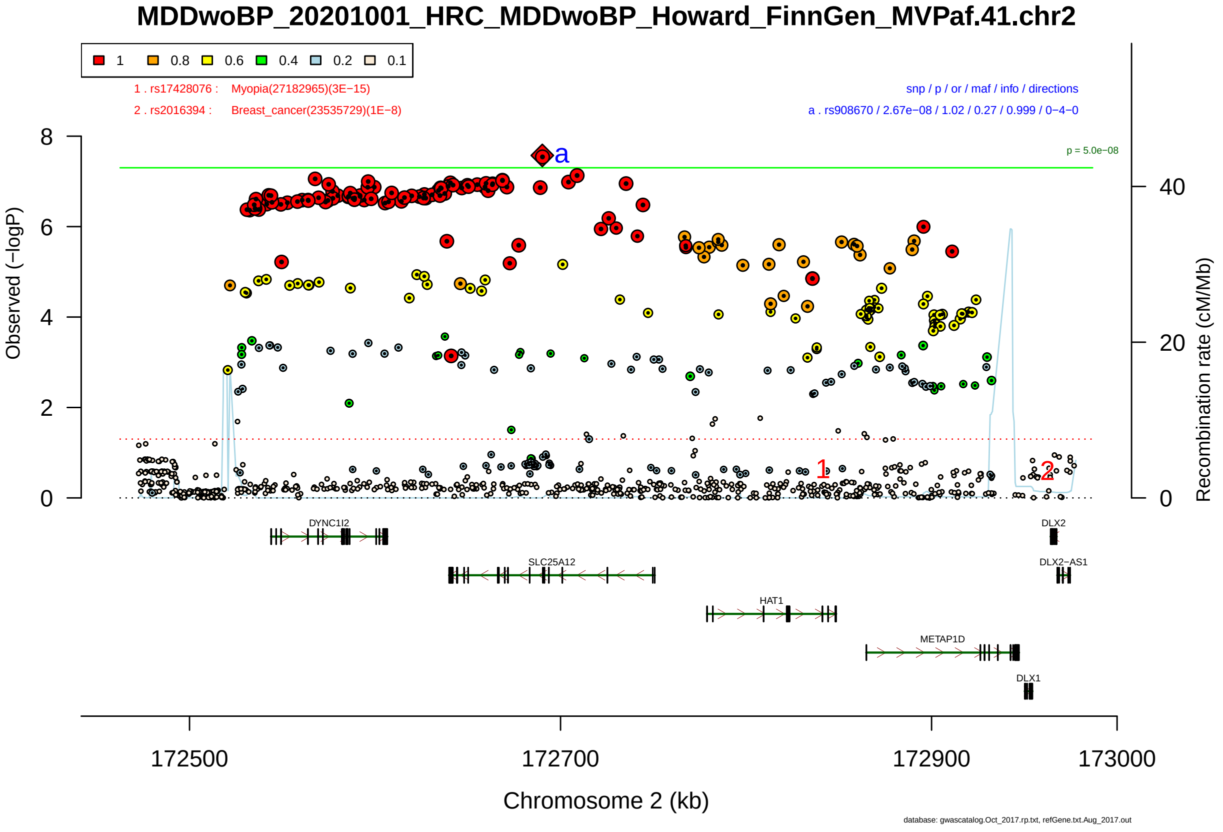

Figure S2-211: Regional plot of GWAS locus No. 211 , with rs6756747 as lead SNP.

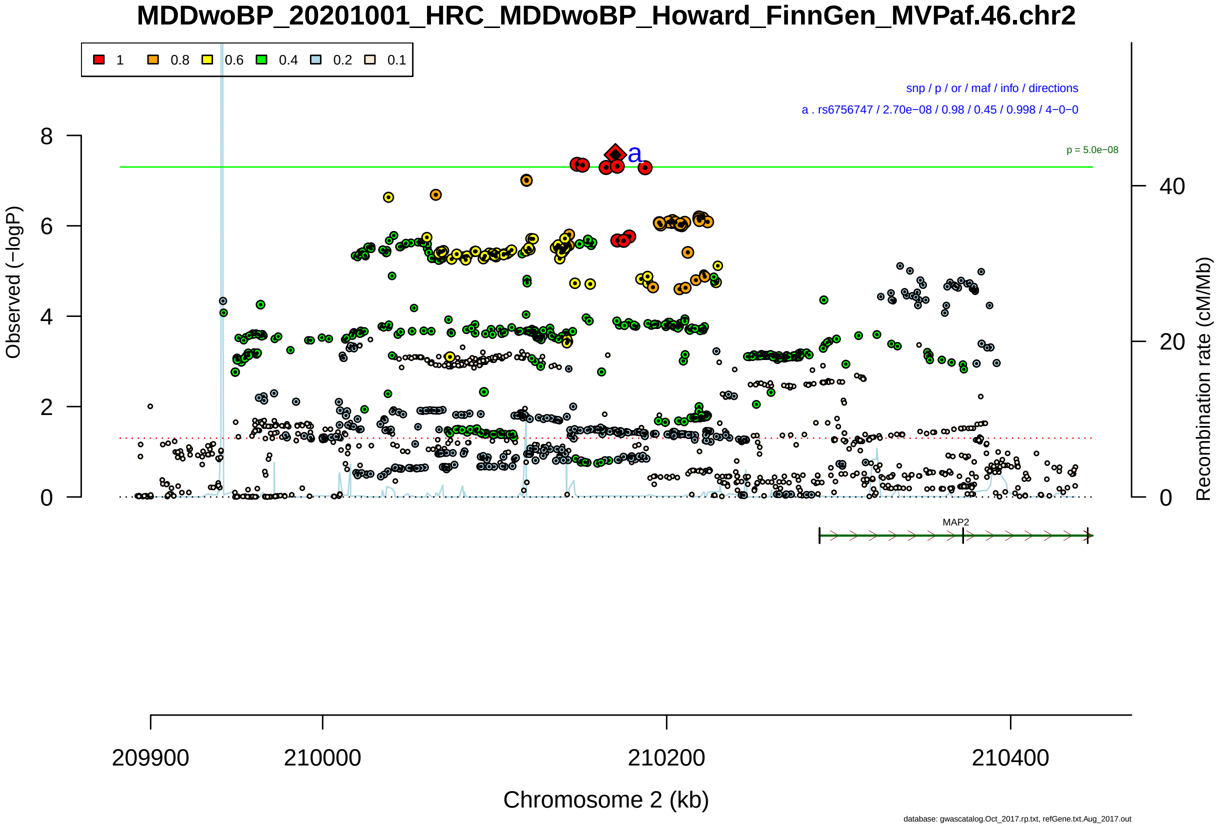

Figure S2-212: Regional plot of GWAS locus No. 212 , with rs10491990 as lead SNP.

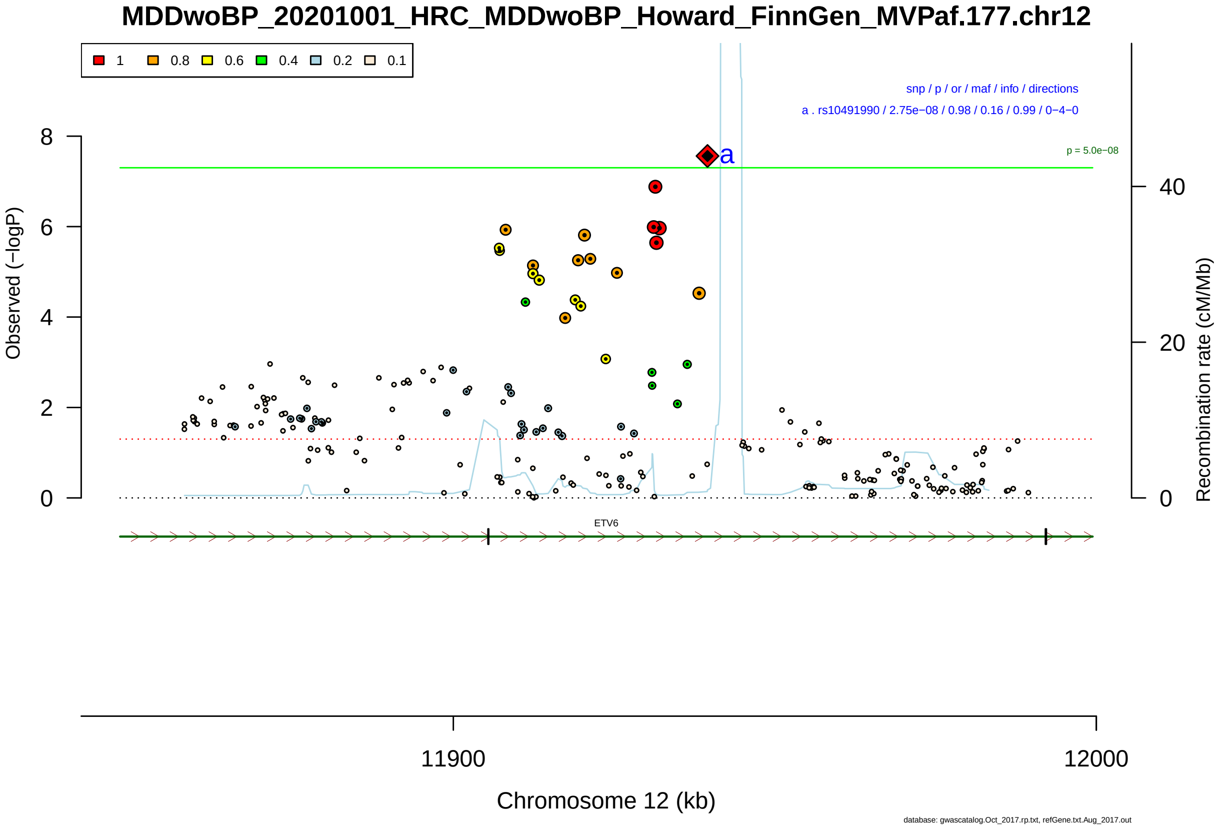

Figure S2-213: Regional plot of GWAS locus No. 213 , with rs11138500 as lead SNP.

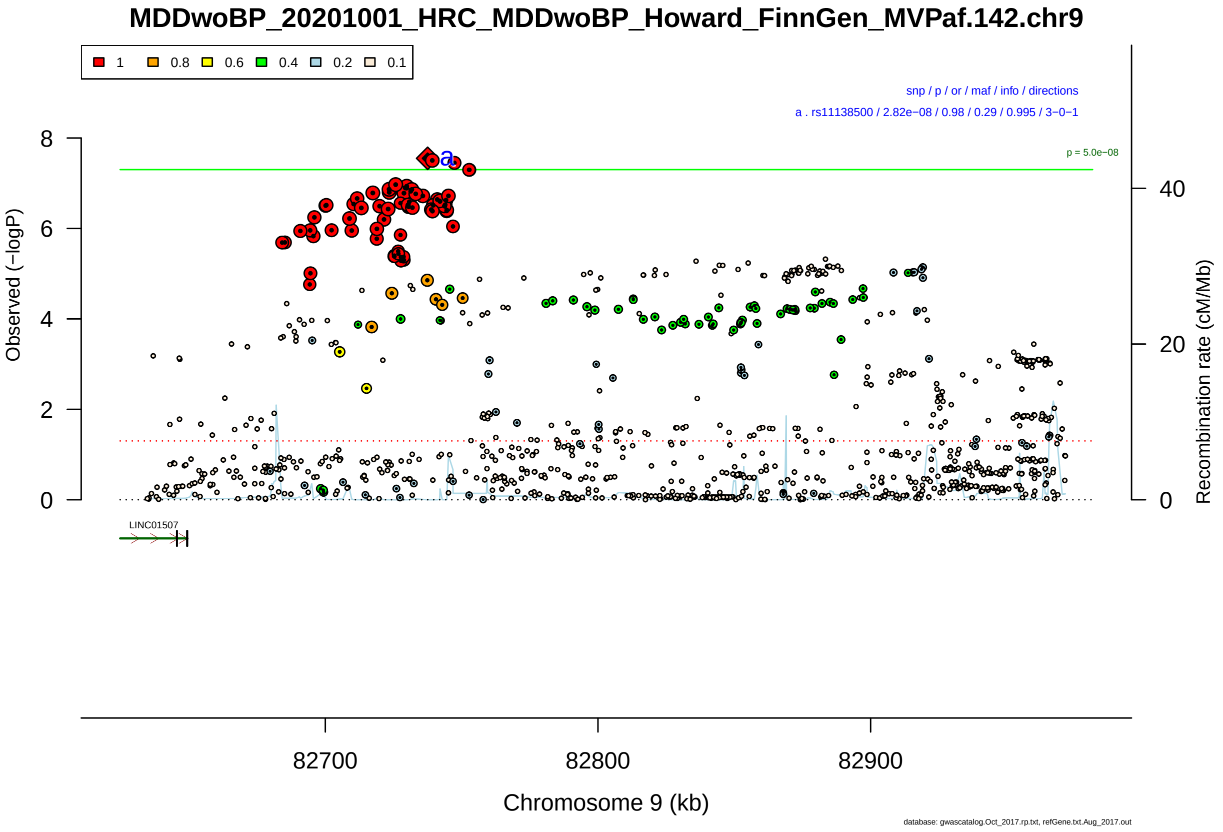

Figure S2-214: Regional plot of GWAS locus No. 214 , with rs17752199 as lead SNP.

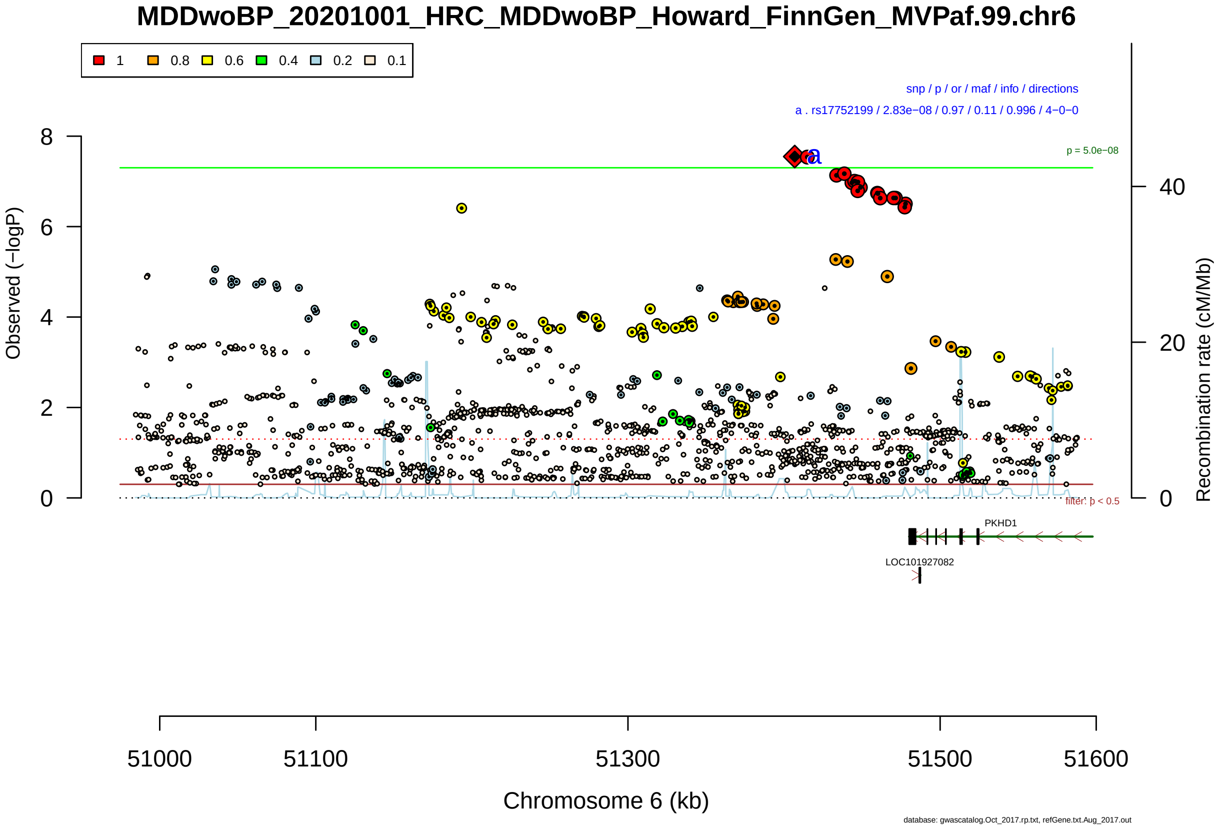

Figure S2-215: Regional plot of GWAS locus No. 215 , with rs4258186 as lead SNP.

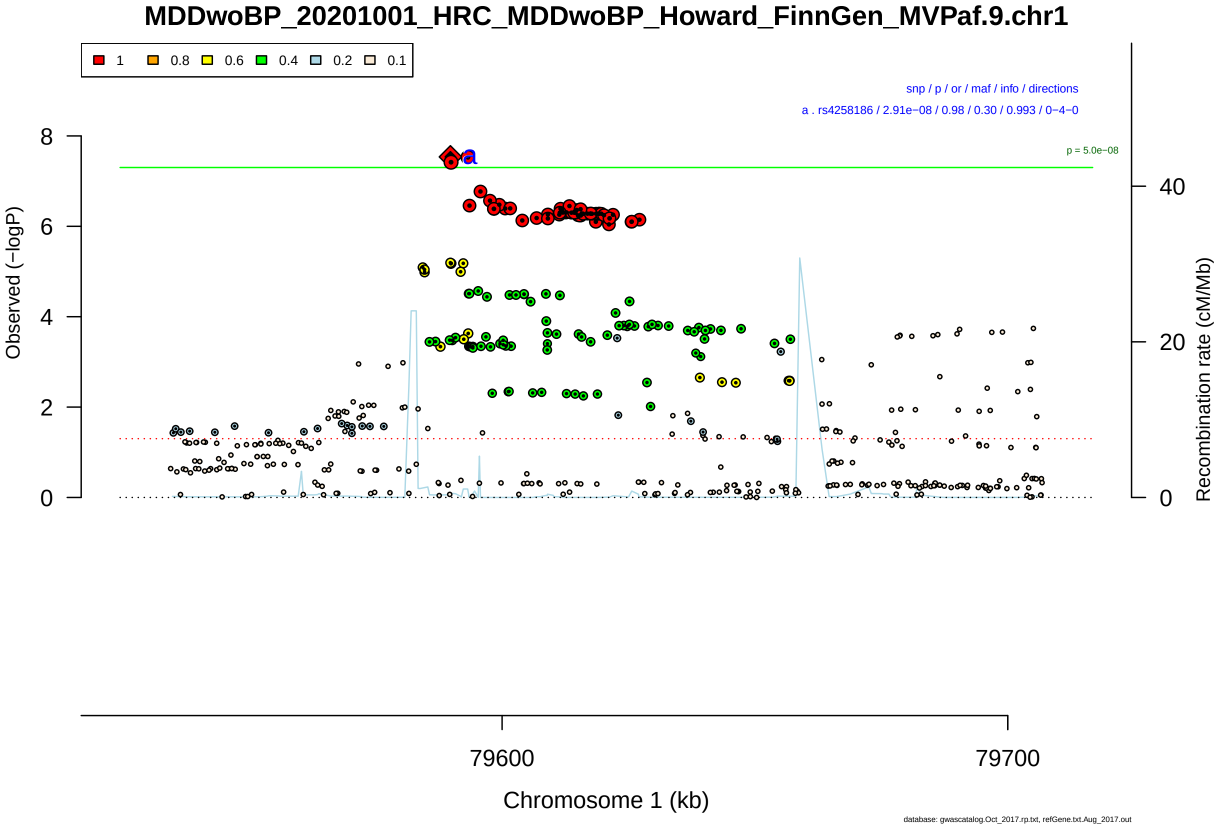

Figure S2-216: Regional plot of GWAS locus No. 216 , with rs73532287 as lead SNP.

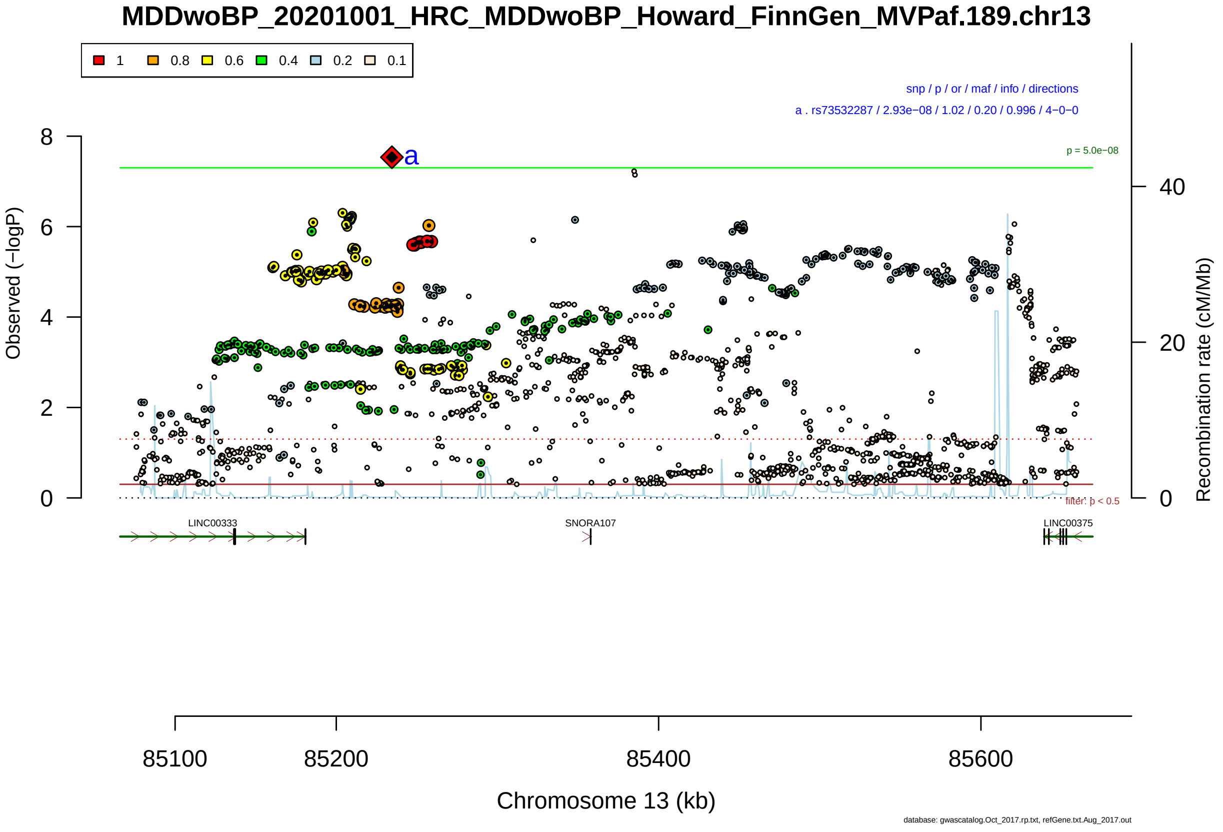

Figure S2-217: Regional plot of GWAS locus No. 217 , with rs11855600 as lead SNP.

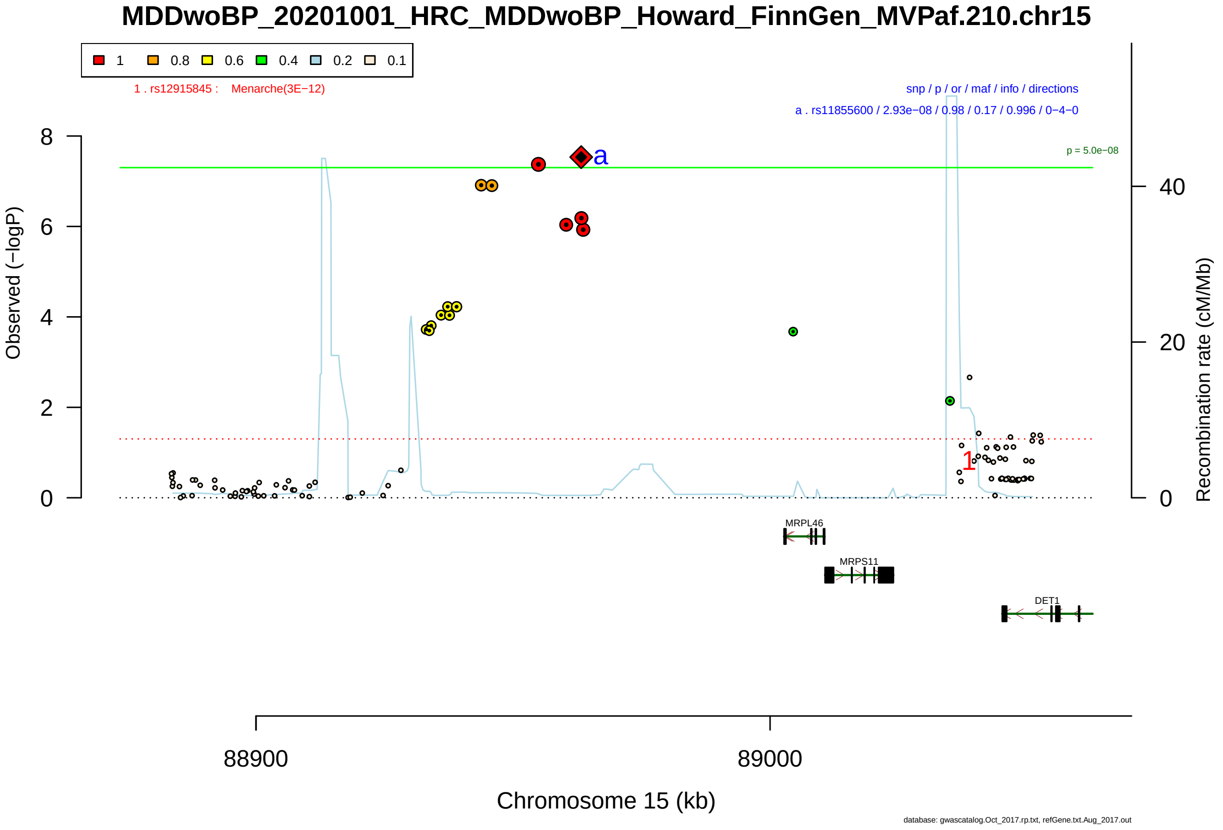

Figure S2-218: Regional plot of GWAS locus No. 218 , with rs11264103 as lead SNP.

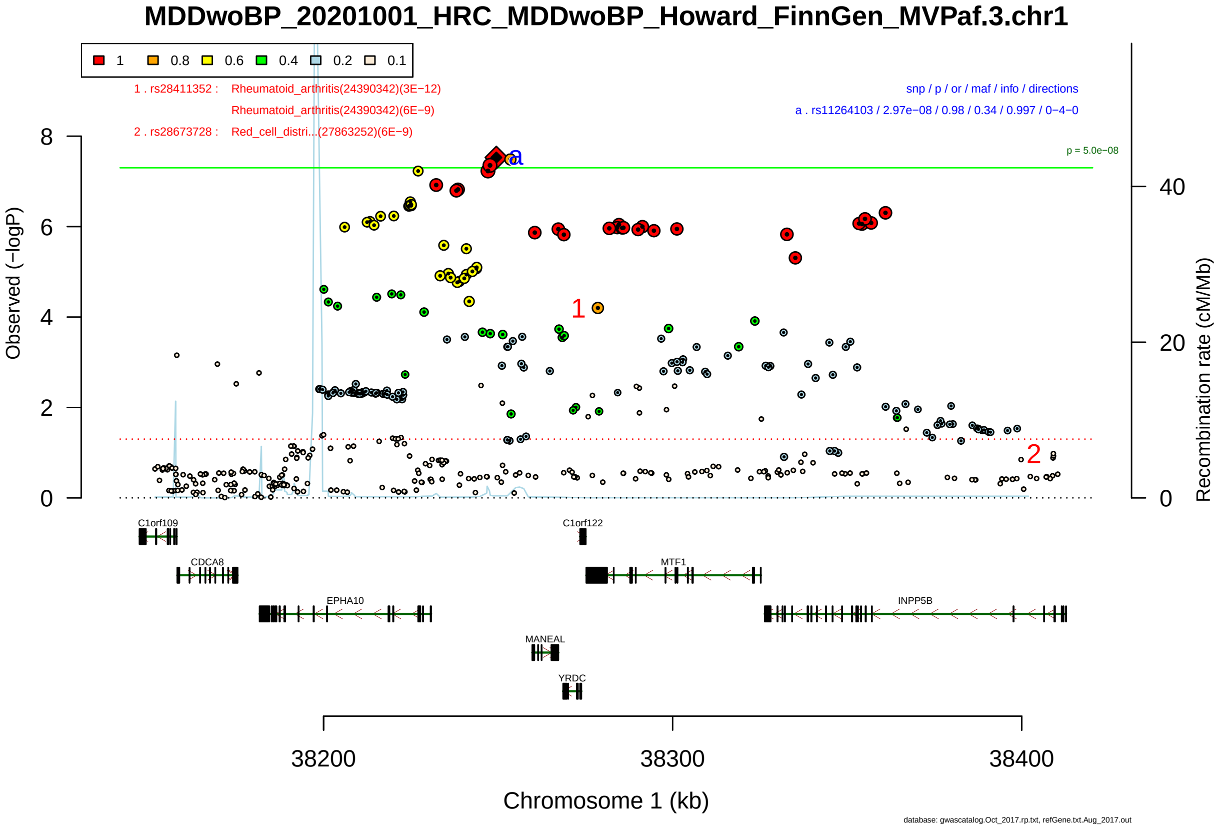

Figure S2-219: Regional plot of GWAS locus No. 219 , with rs62248938 as lead SNP.

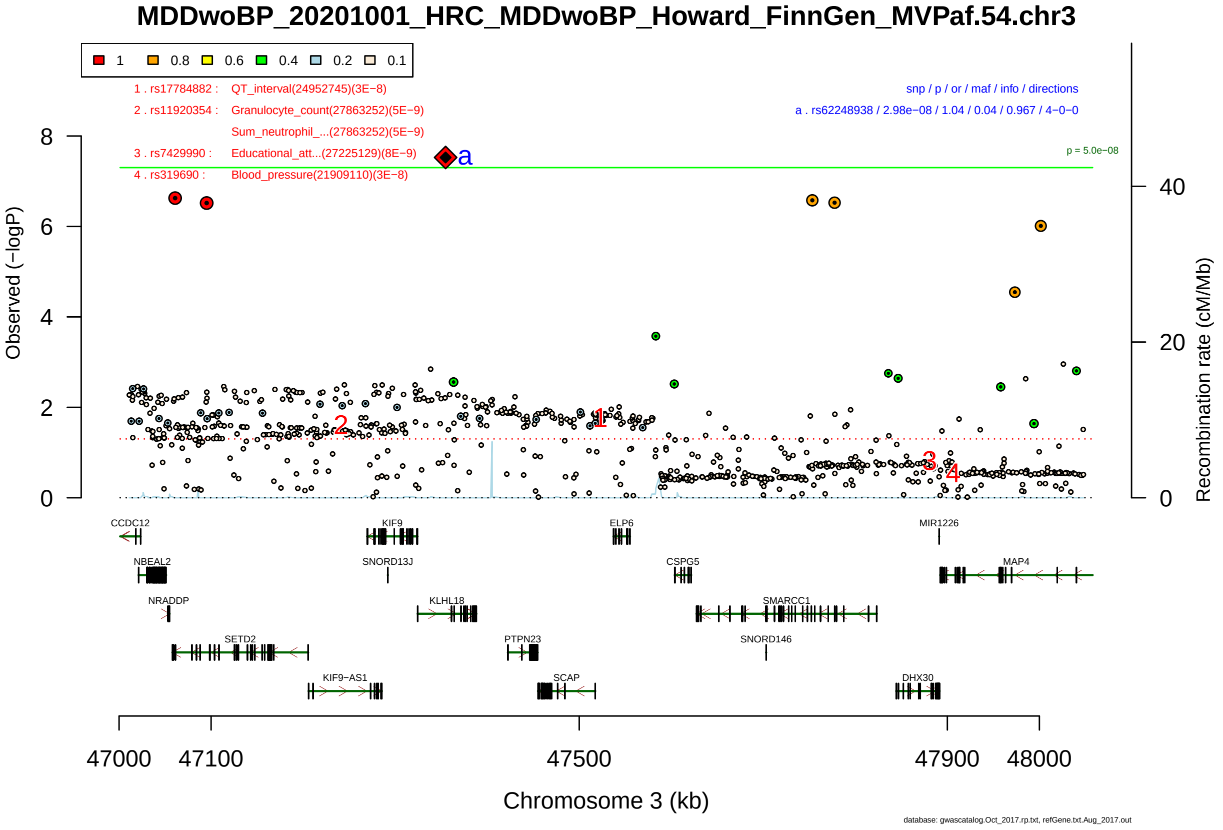

Figure S2-220: Regional plot of GWAS locus No. 220 , with rs10986606 as lead SNP.

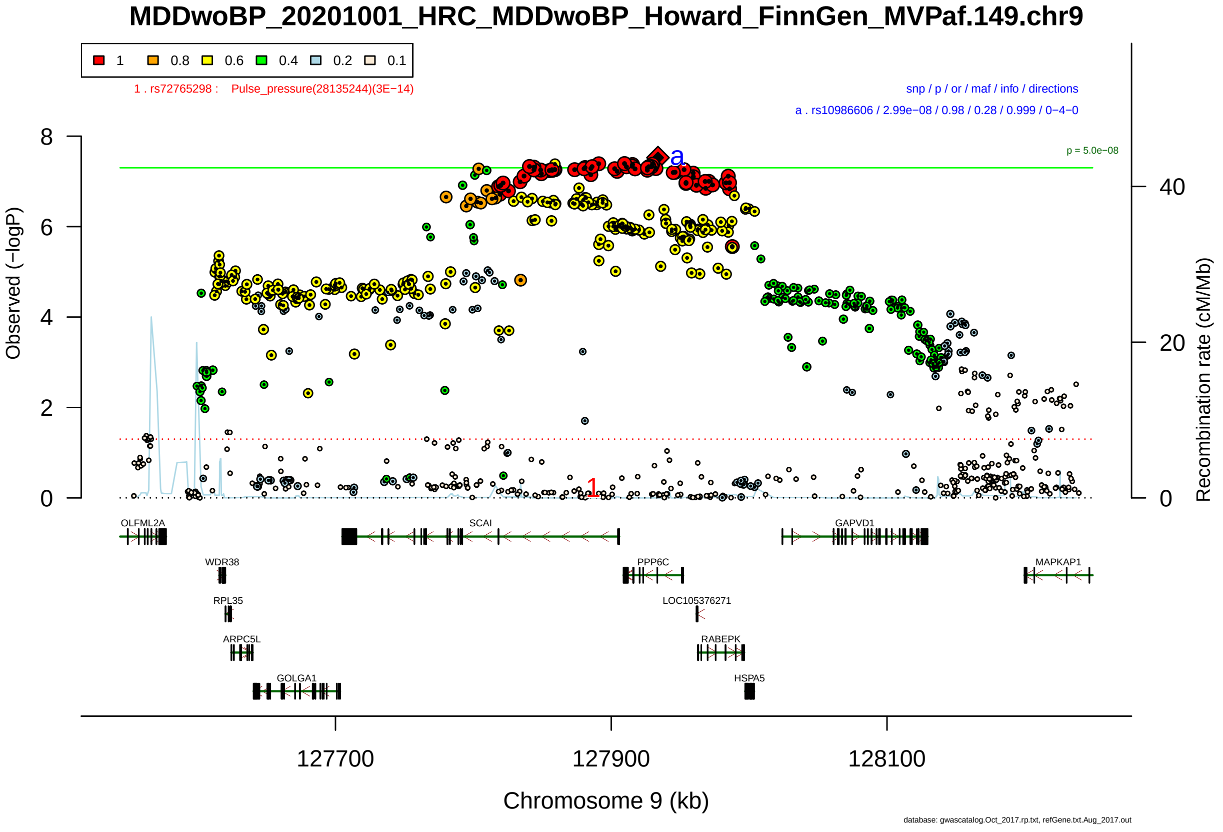

Figure S2-221: Regional plot of GWAS locus No. 221 , with rs544565 as lead SNP.

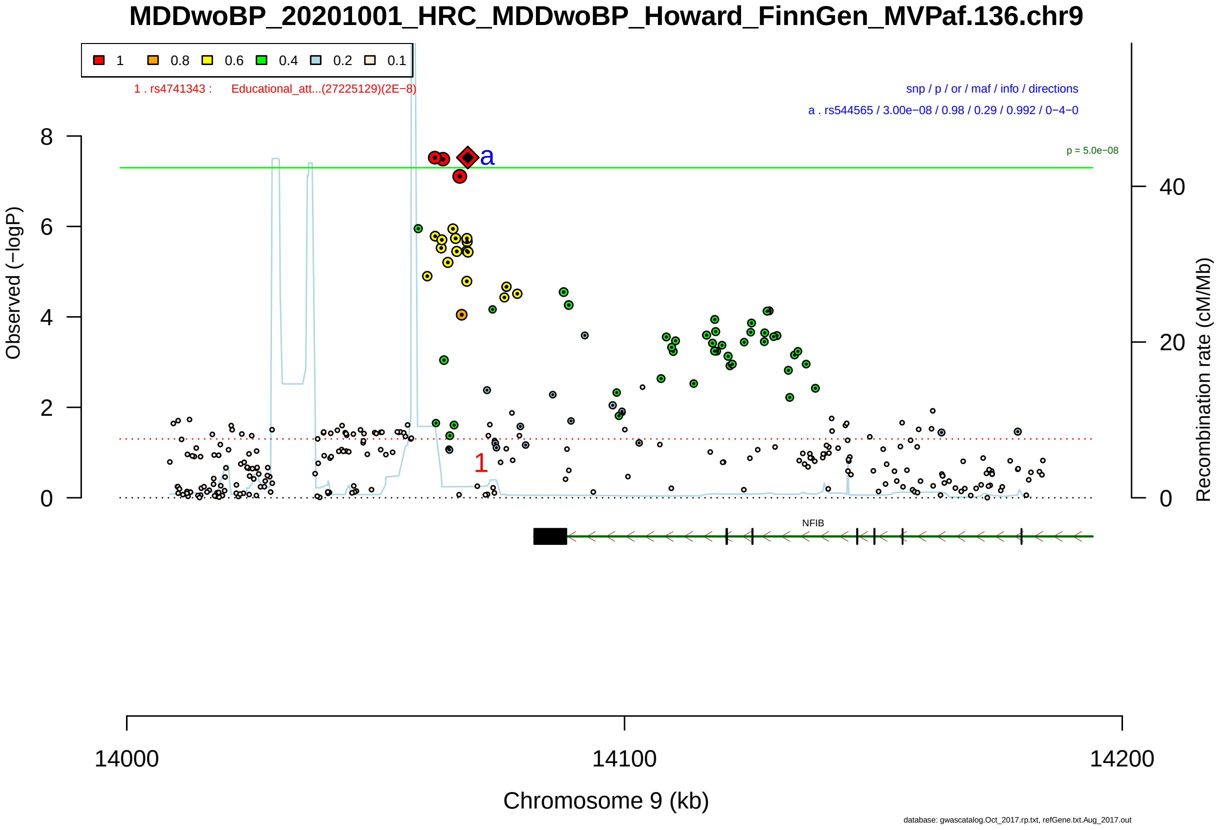

Figure S2-222: Regional plot of GWAS locus No. 222 , with rs12131852 as lead SNP.

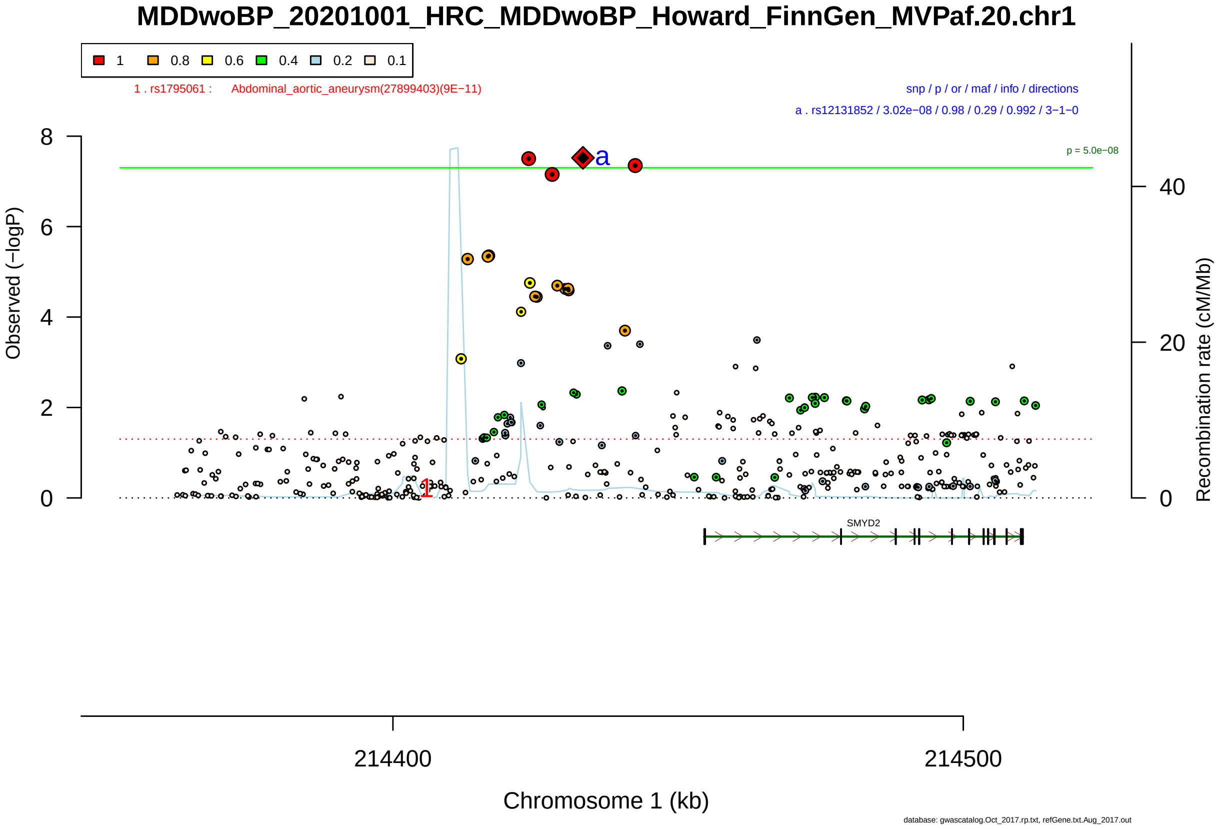

Figure S2-223: Regional plot of GWAS locus No. 223 , with rs288181 as lead SNP.

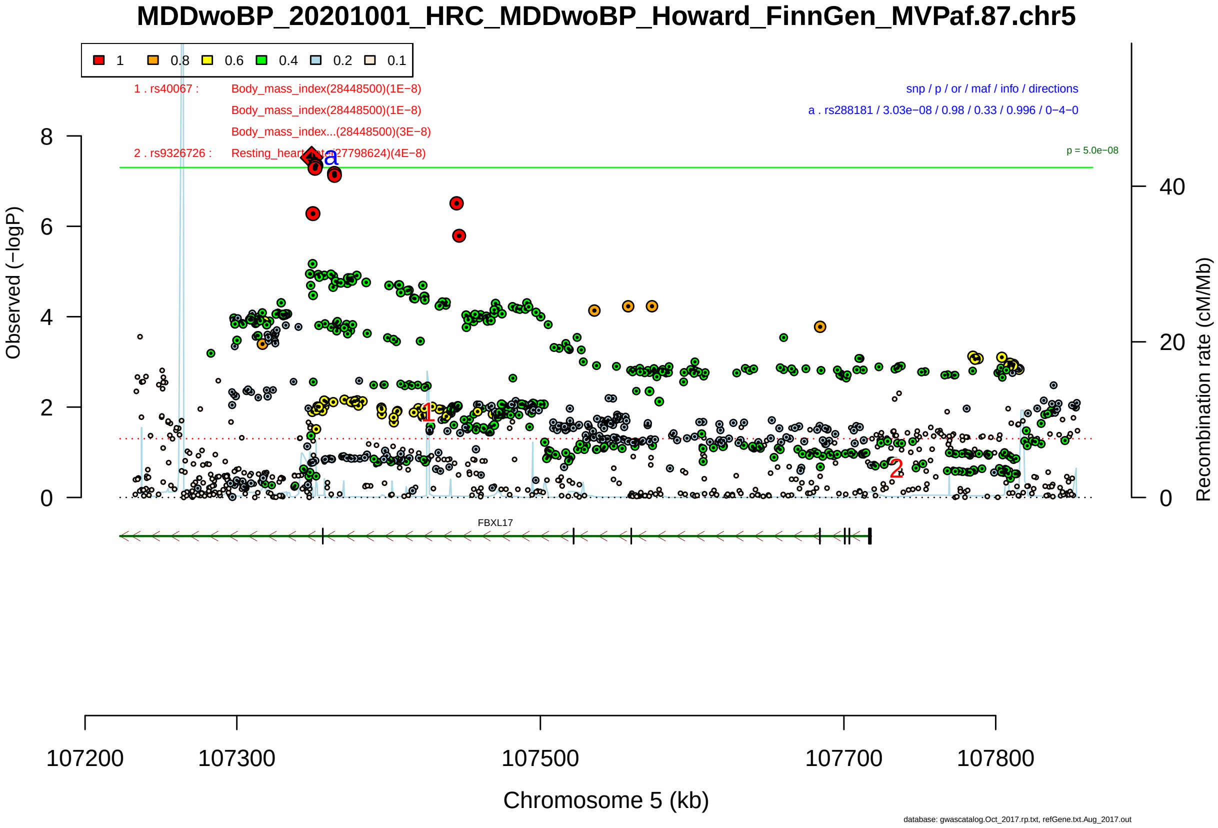

Figure S2-224: Regional plot of GWAS locus No. 224 , with rs7752418 as lead SNP.

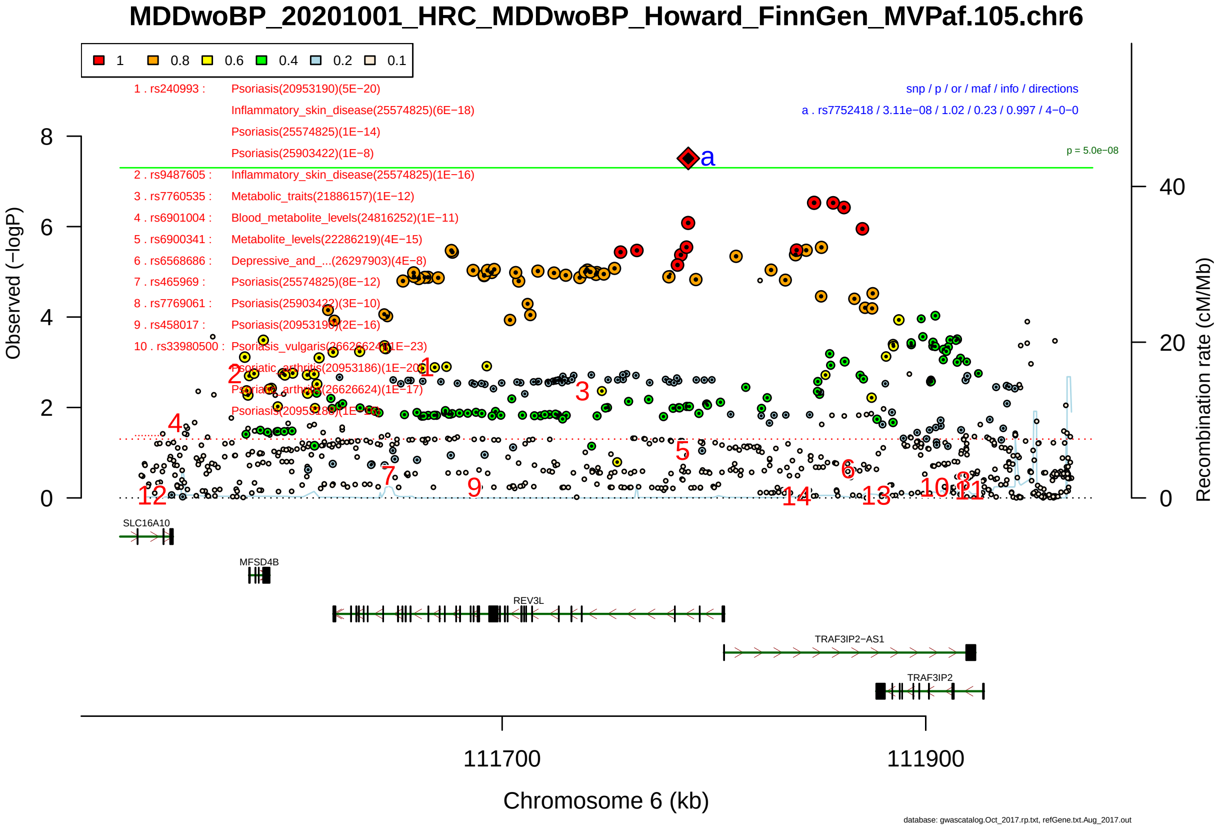

Figure S2-225: Regional plot of GWAS locus No. 225 , with rs211792 as lead SNP.

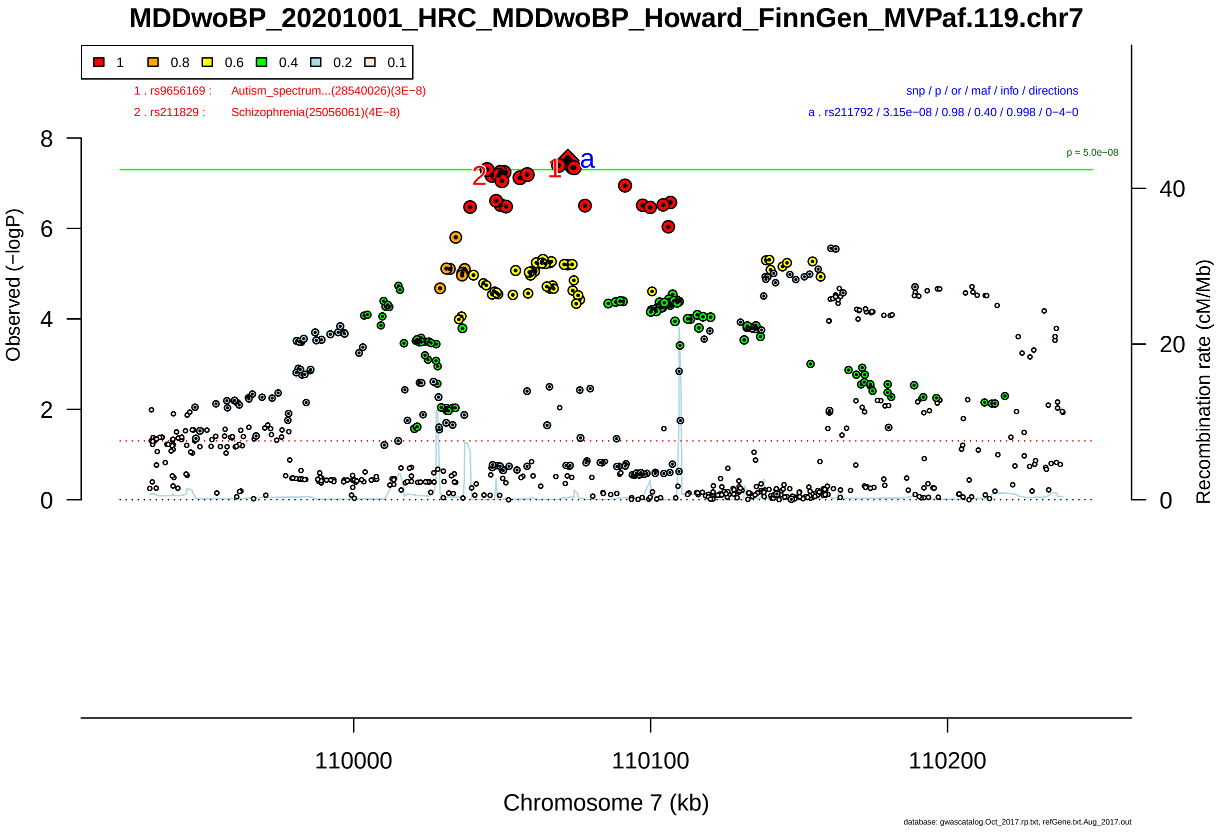

Figure S2-226: Regional plot of GWAS locus No. 226 , with rs6443178 as lead SNP.

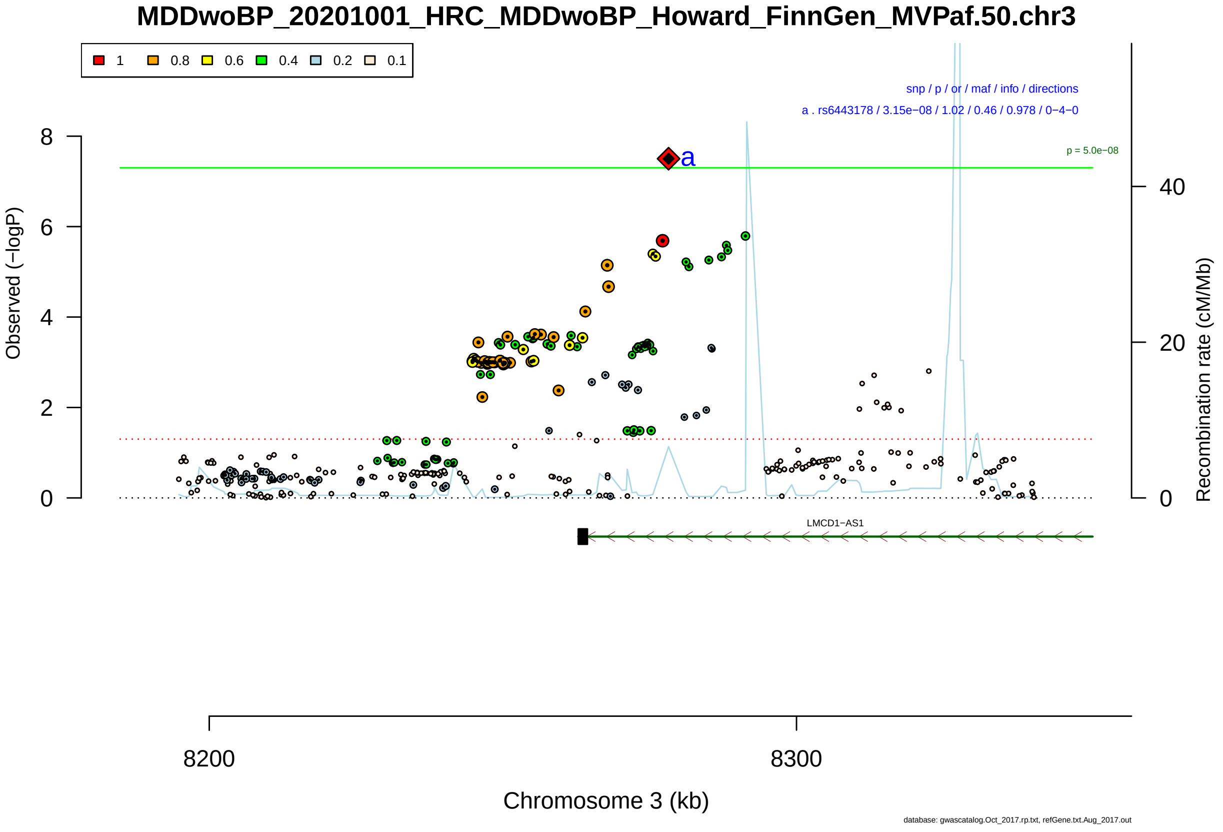

Figure S2-227: Regional plot of GWAS locus No. 227 , with rs6713469 as lead SNP.

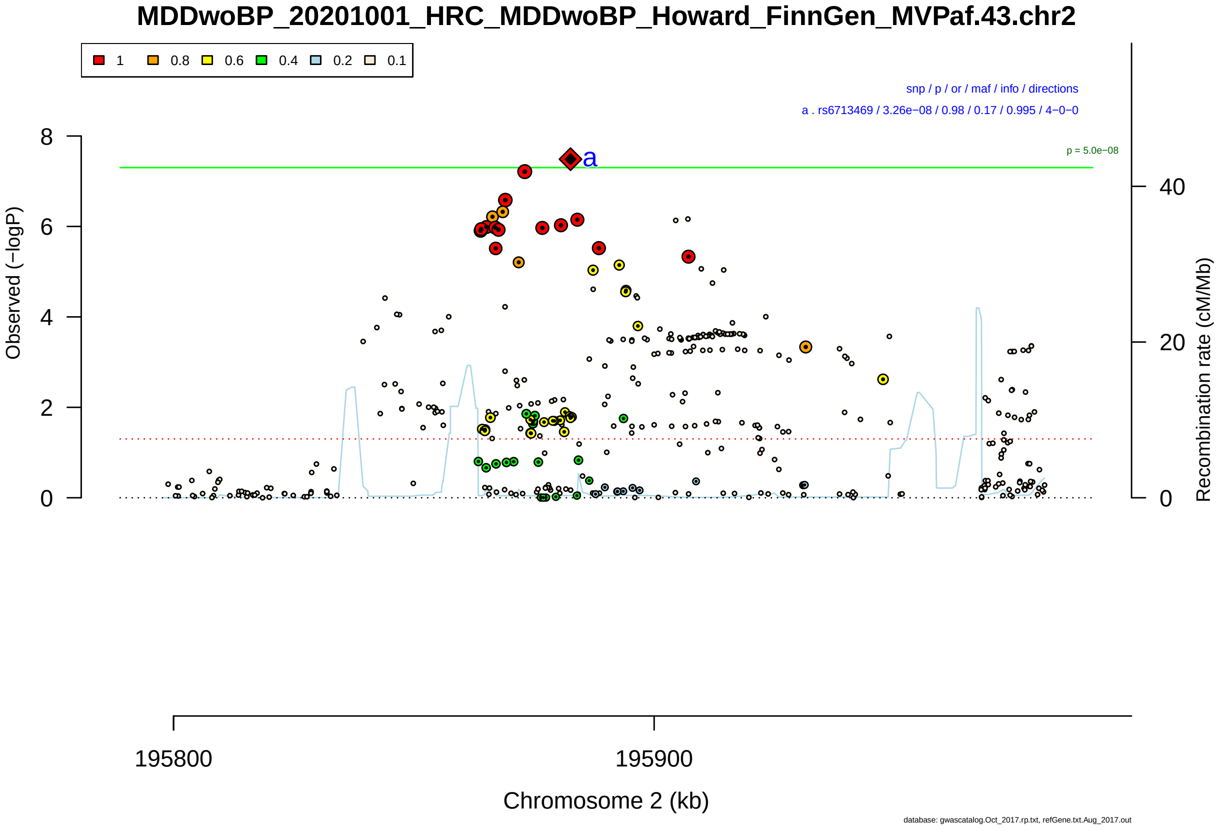

Figure S2-228: Regional plot of GWAS locus No. 228 , with rs1437336 as lead SNP.

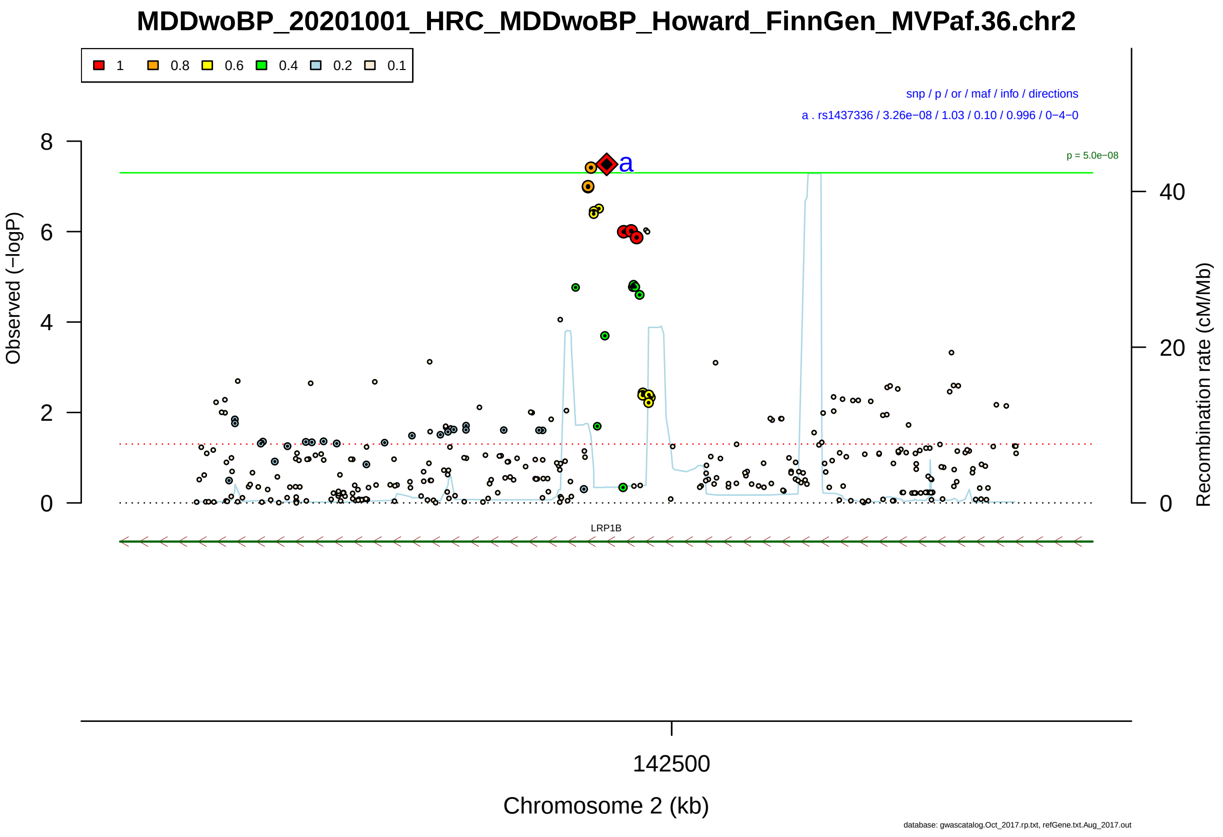

Figure S2-229: Regional plot of GWAS locus No. 229 , with rs520838 as lead SNP.

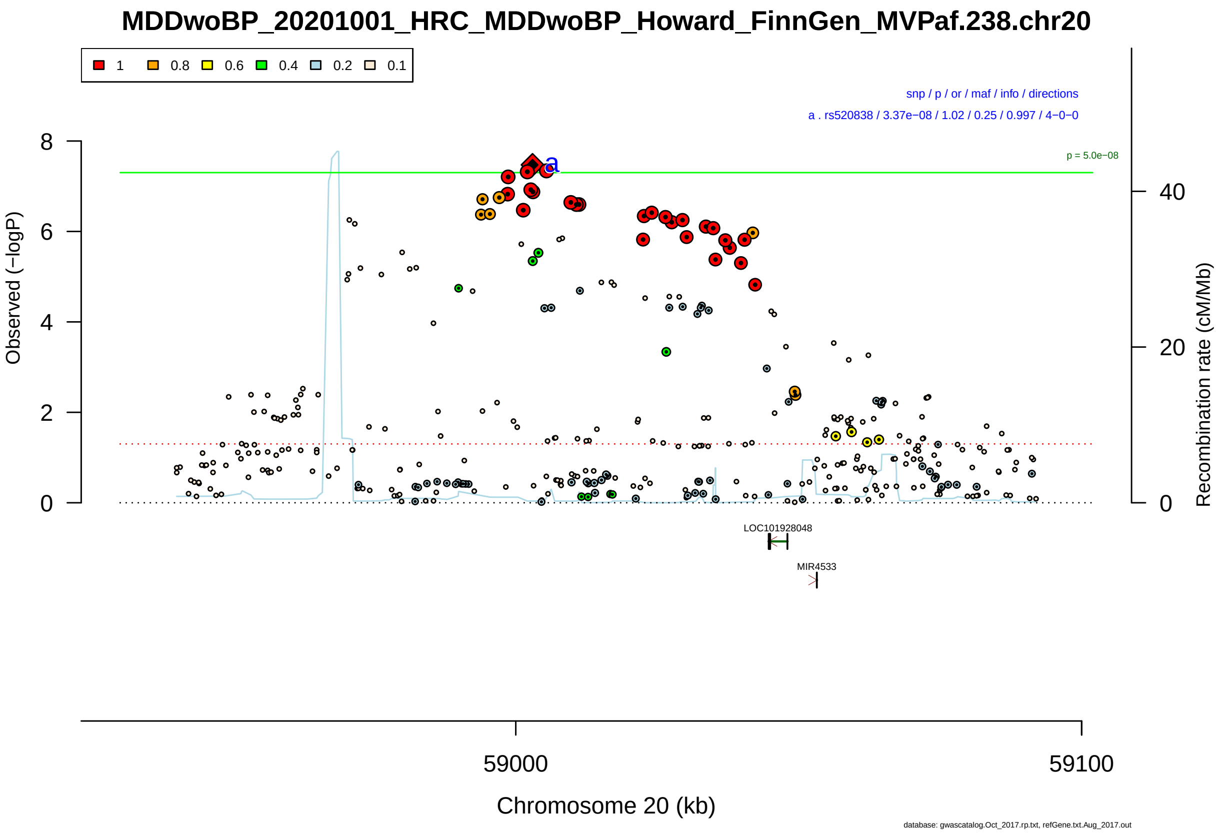

Figure S2-230: Regional plot of GWAS locus No. 230 , with rs10749240 as lead SNP.

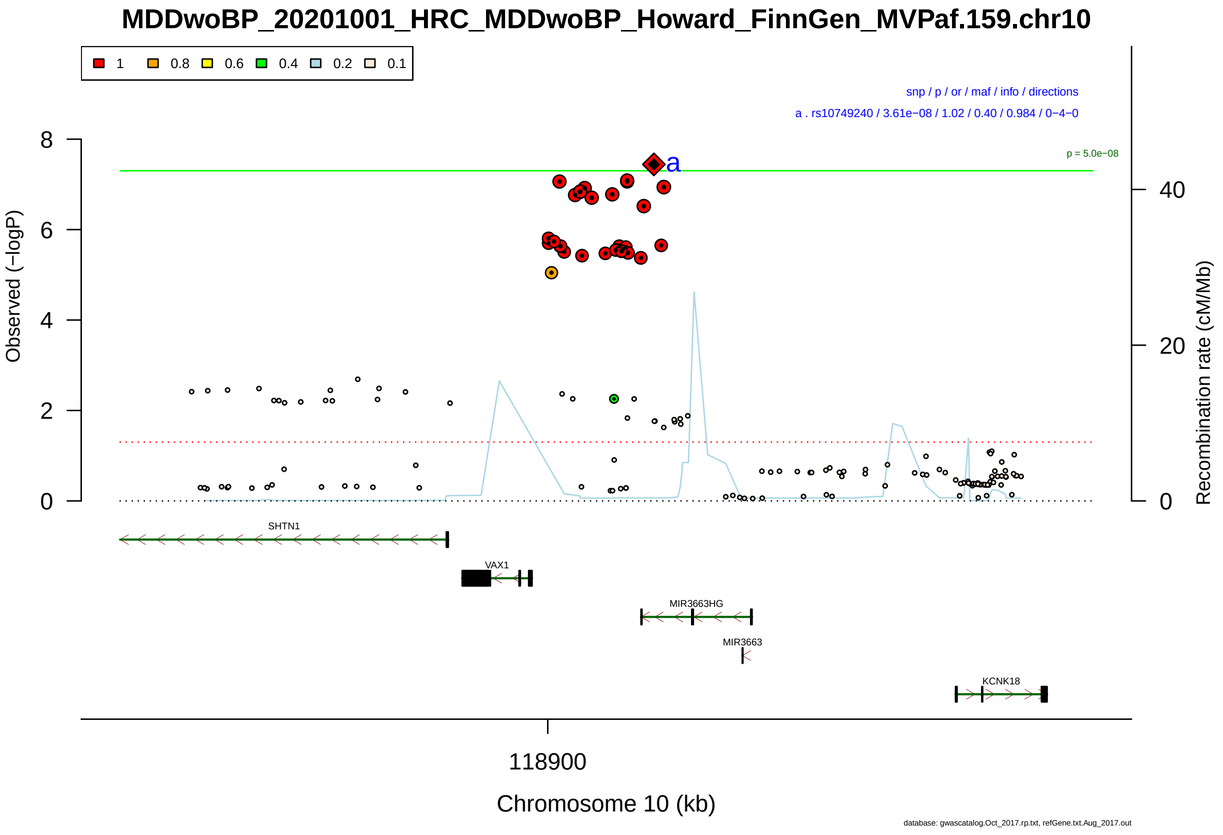

Figure S2-231: Regional plot of GWAS locus No. 231 , with rs4784296 as lead SNP.

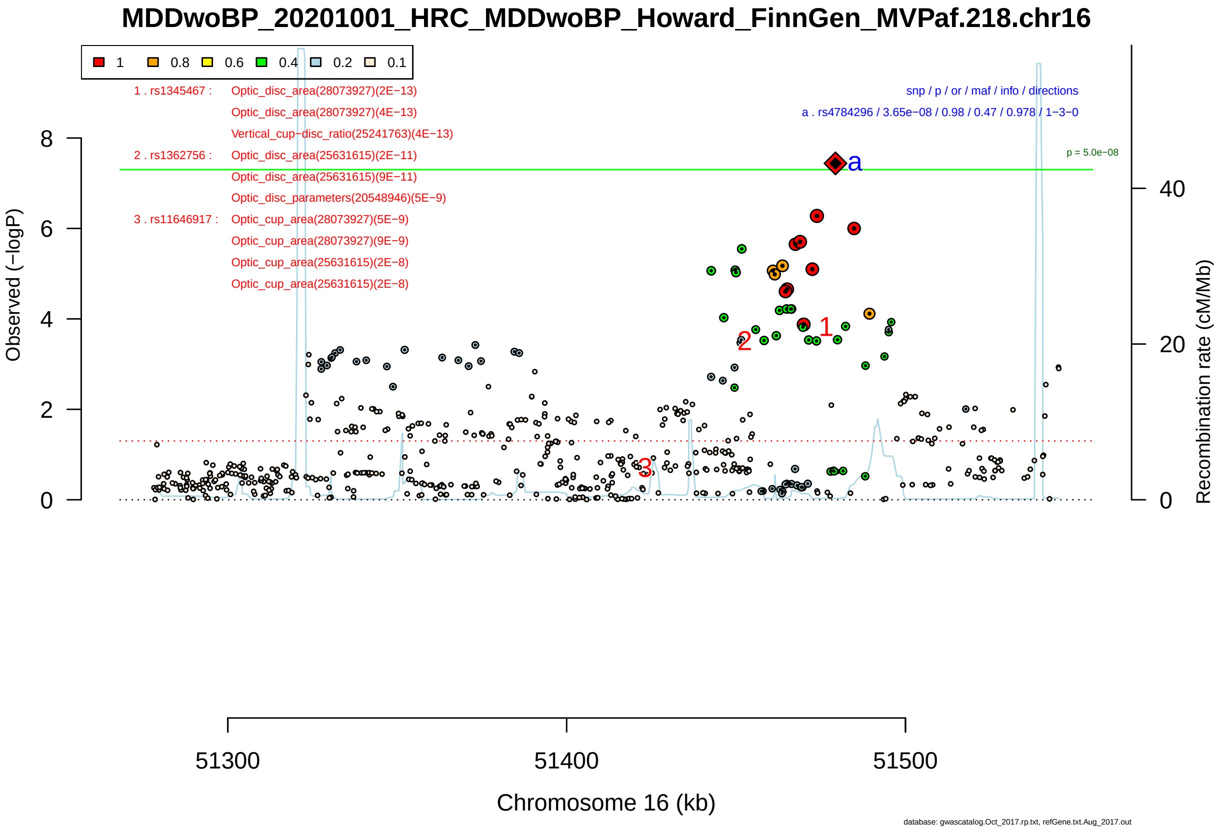

Figure S2-232: Regional plot of GWAS locus No. 232 , with rs73123350 as lead SNP.

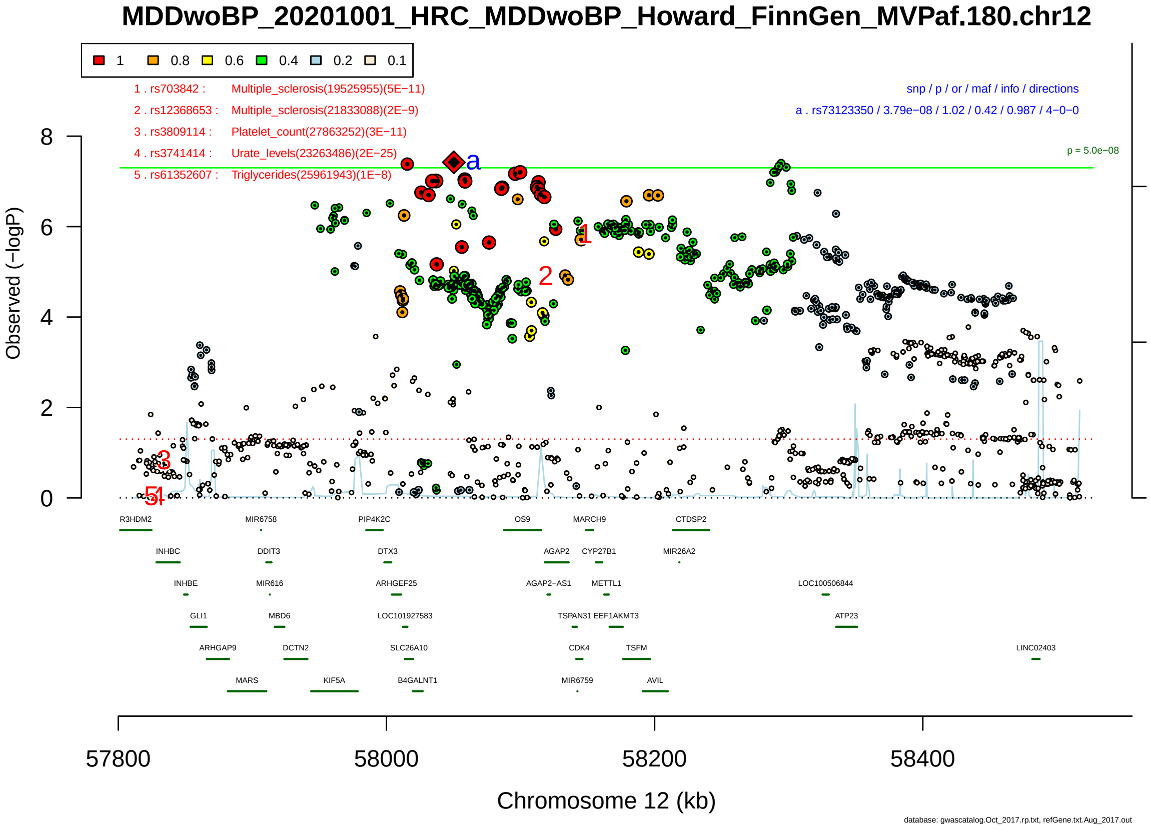

Figure S2-233: Regional plot of GWAS locus No. 233 , with rs12898464 as lead SNP.

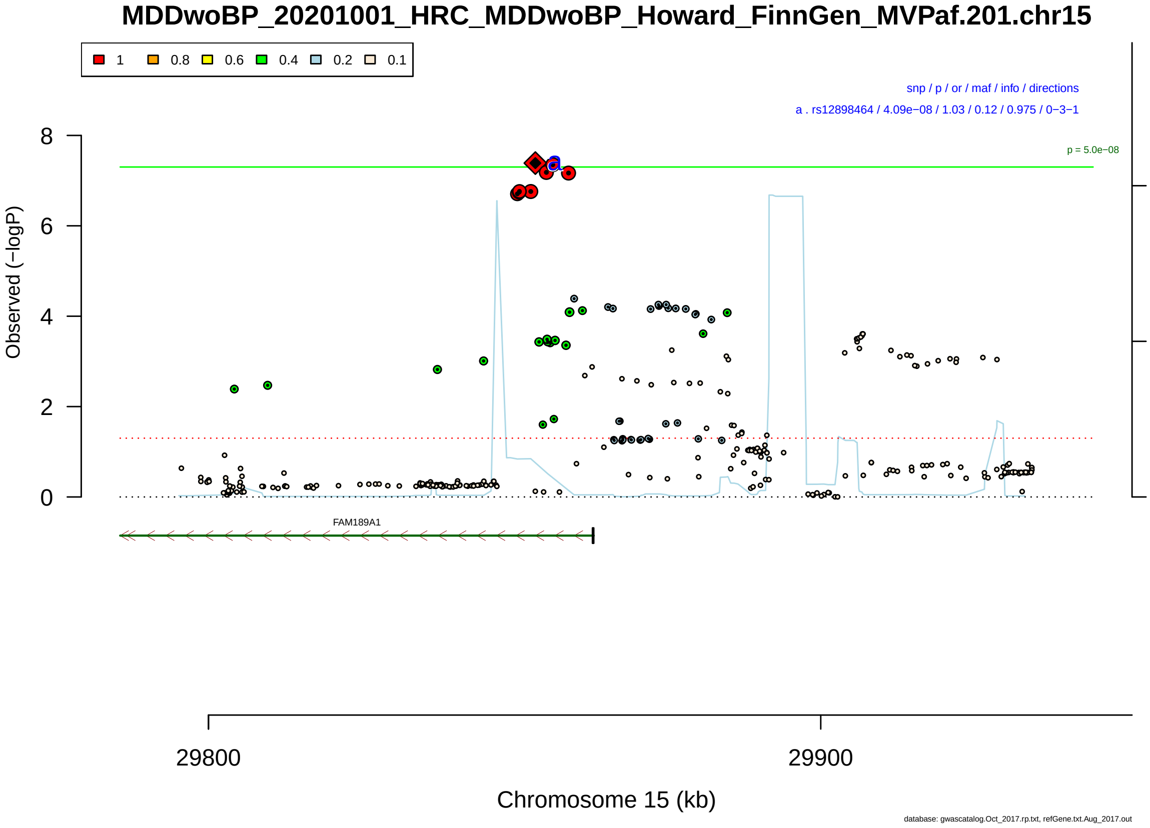

Figure S2-234: Regional plot of GWAS locus No. 234 , with rs2411044 as lead SNP.

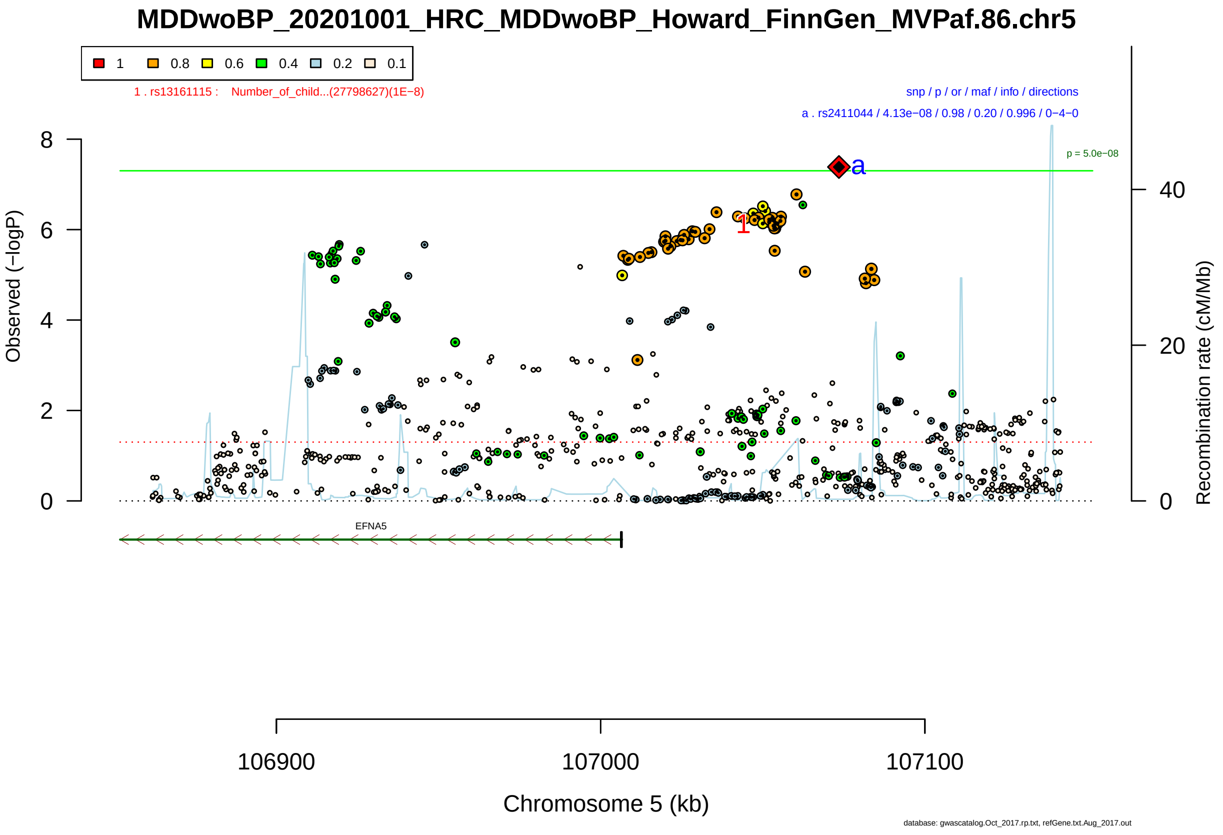

Figure S2-235: Regional plot of GWAS locus No. 235 , with rs2740792 as lead SNP.

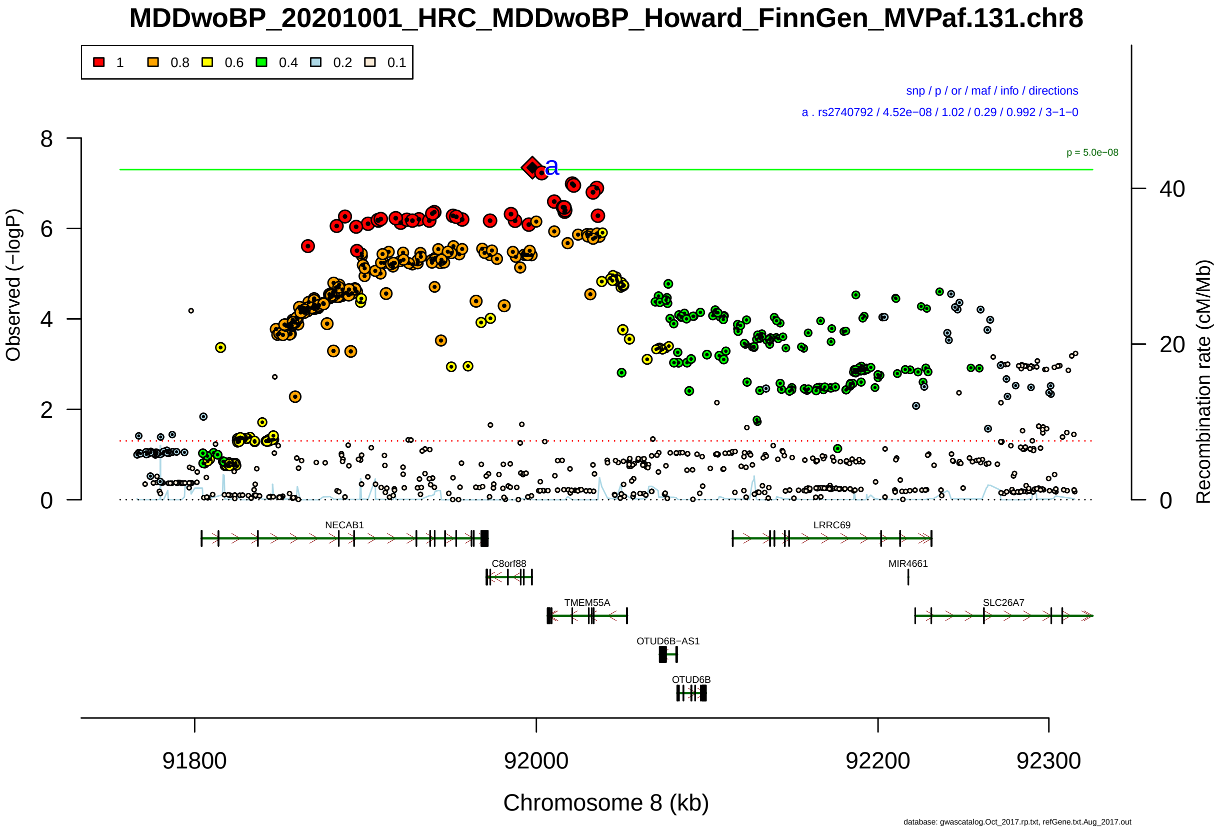

Figure S2-236: Regional plot of GWAS locus No. 236 , with rs9974713 as lead SNP.

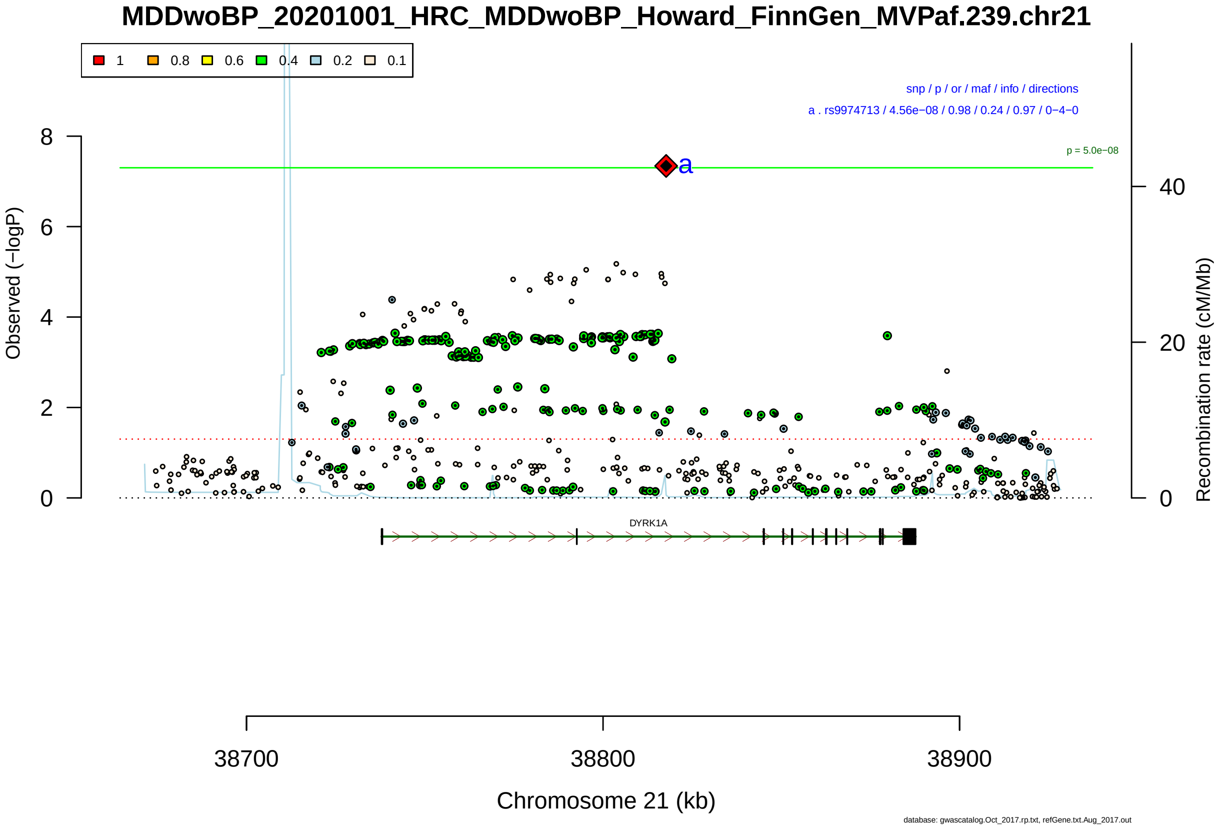

Figure S2-237: Regional plot of GWAS locus No. 237 , with rs11678860 as lead SNP.

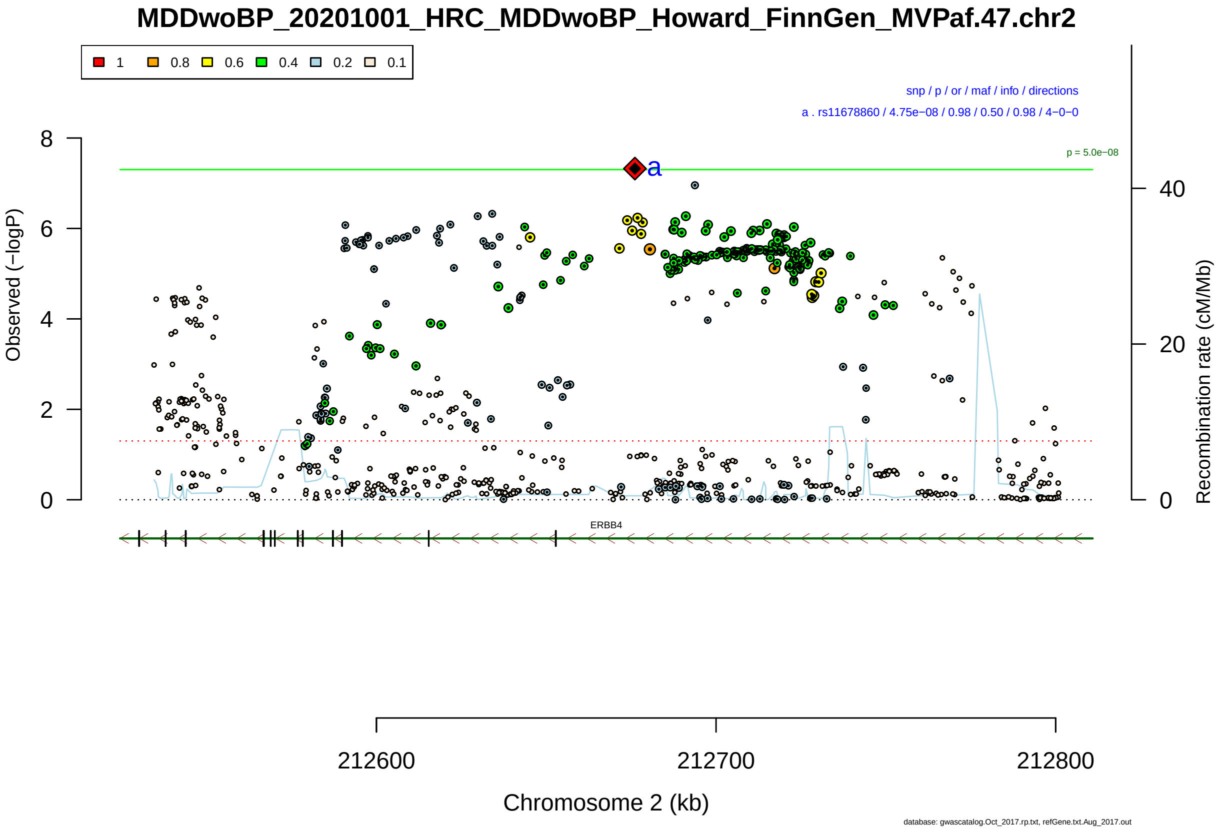

Figure S2-238: Regional plot of GWAS locus No. 238 , with rs2246221 as lead SNP.

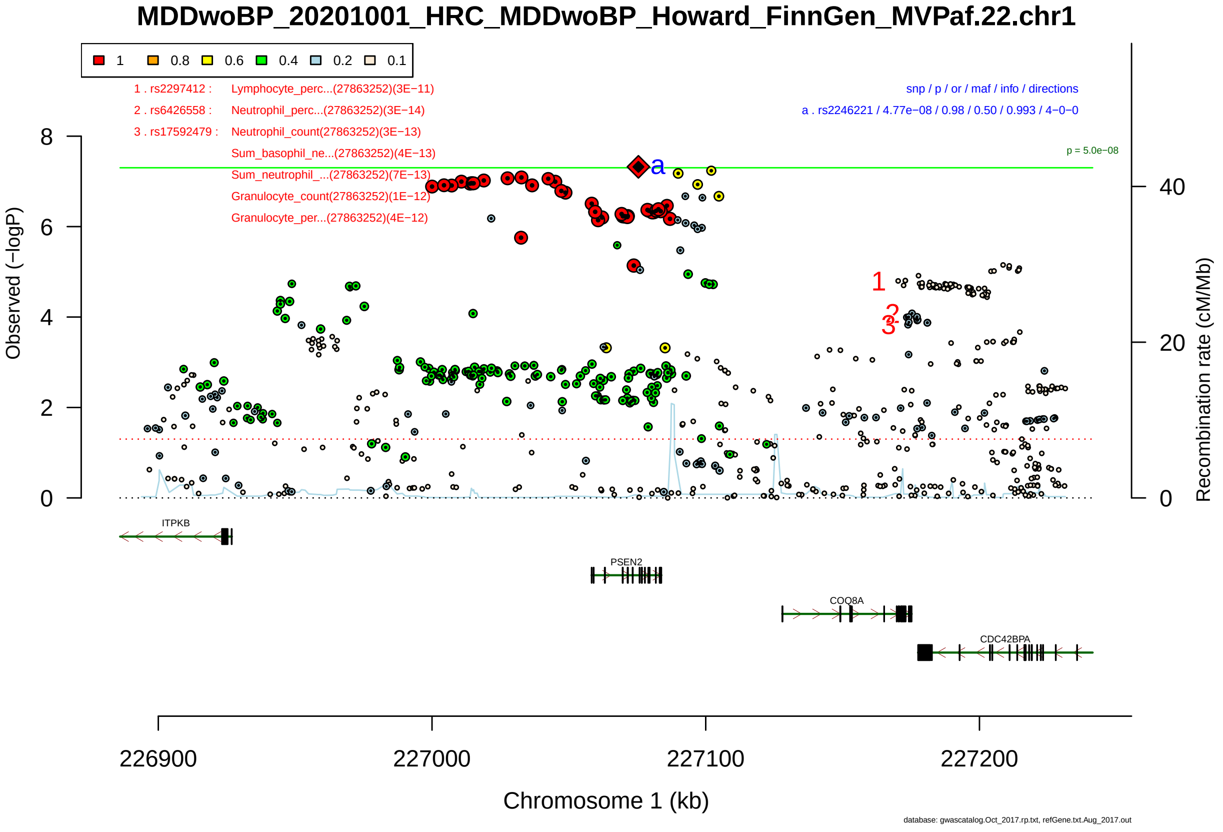

Figure S2-239: Regional plot of GWAS locus No. 239 , with rs6845263 as lead SNP.

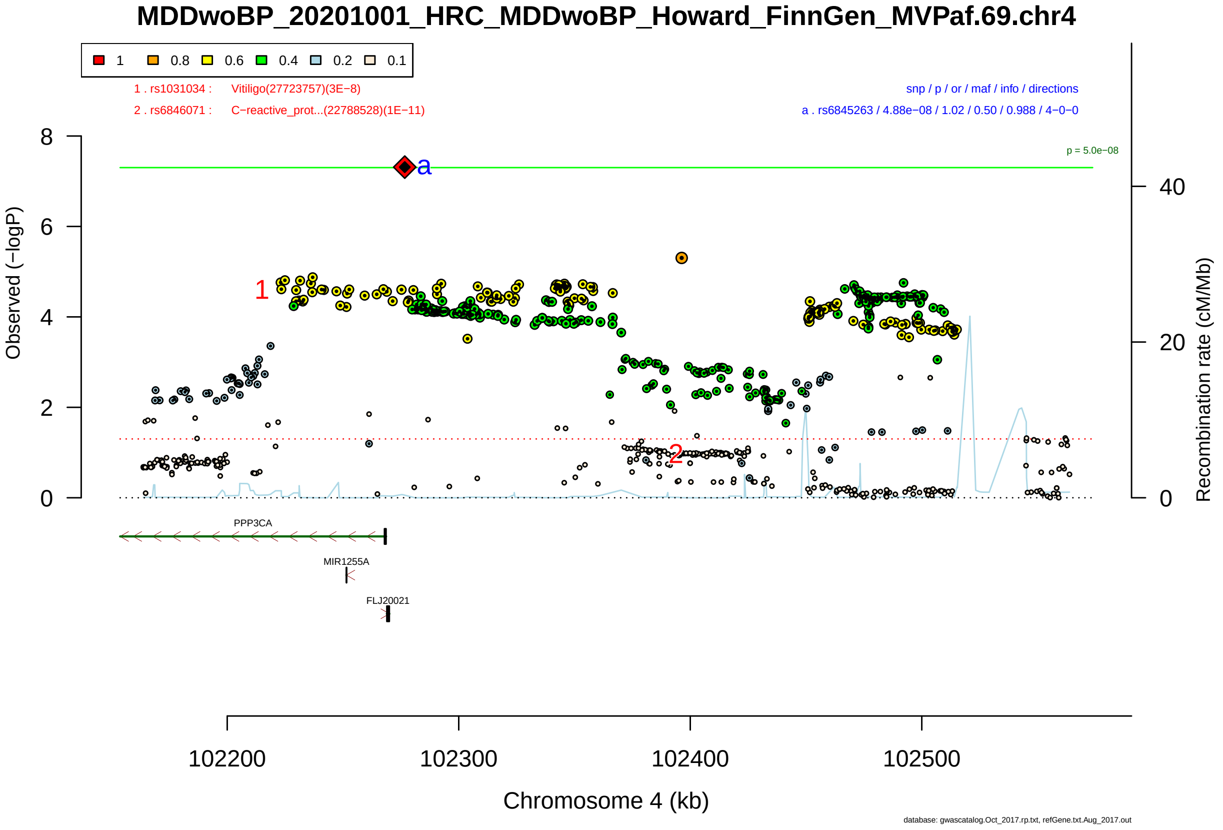

Figure S2-240: Regional plot of GWAS locus No. 240 , with rs11077206 as lead SNP.

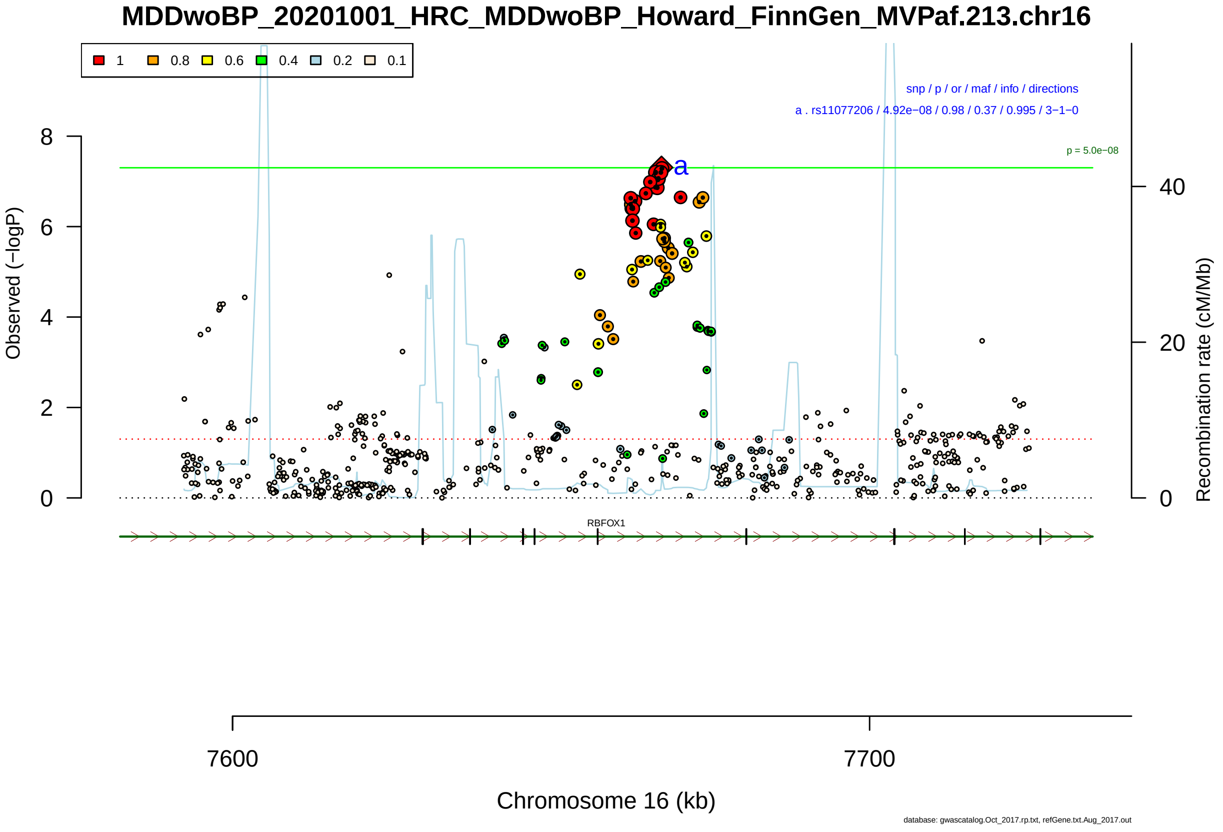

Figure S2-241: Regional plot of GWAS locus No. 241 , with rs2789444 as lead SNP.

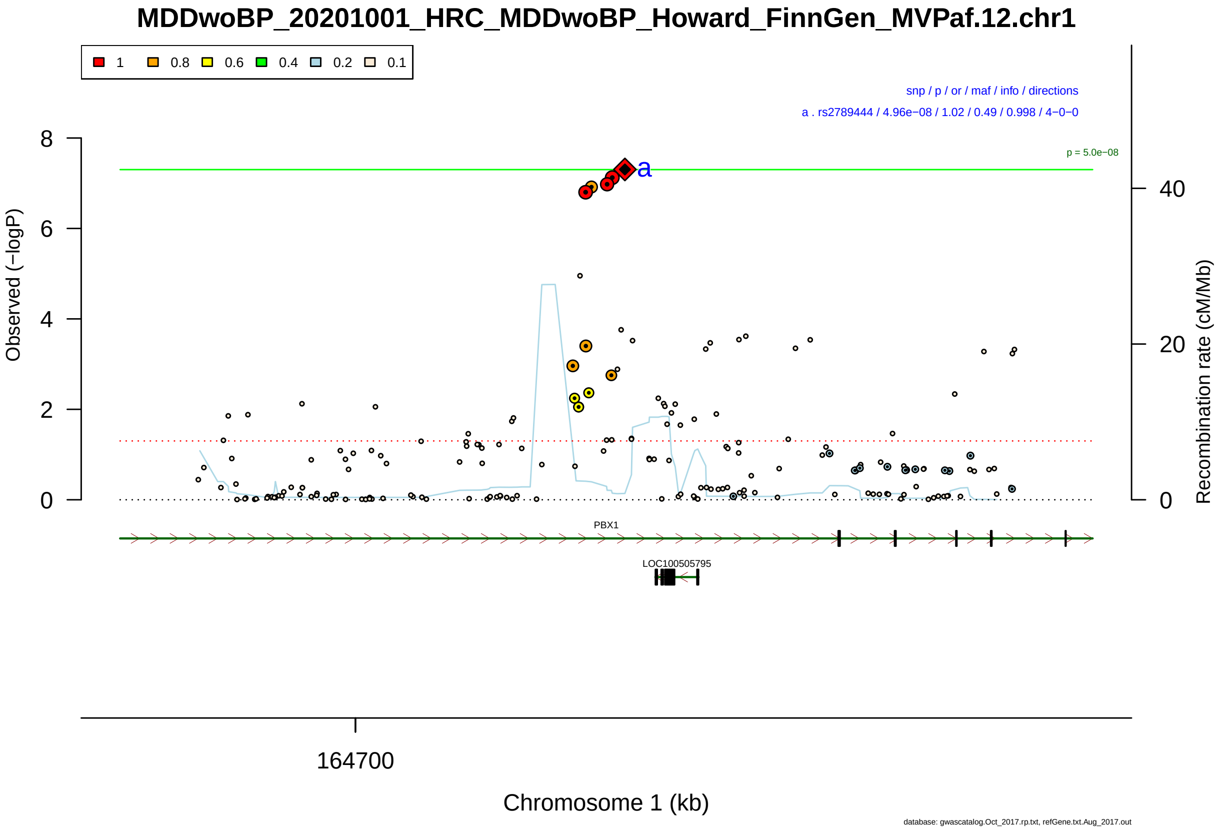

Figure S2-242: Regional plot of GWAS locus No. 242 , with rs11777872 as lead SNP.

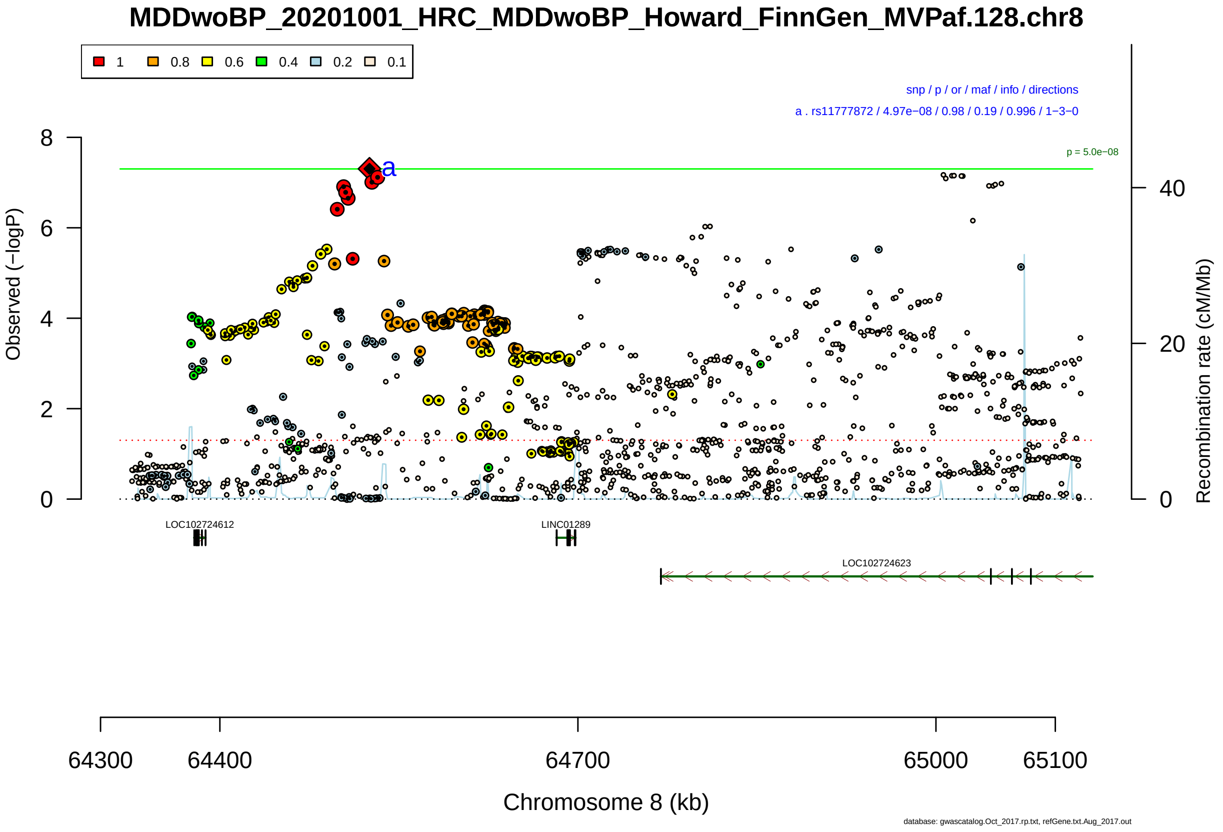

Figure S2-243: Regional plot of GWAS locus No. 243 , with NA as lead SNP.

**daner_MDDwoBP_20201001_HRC_MDDwoBP_Howard_FinnGen_MVPaf.het.gz.p3_GWA**

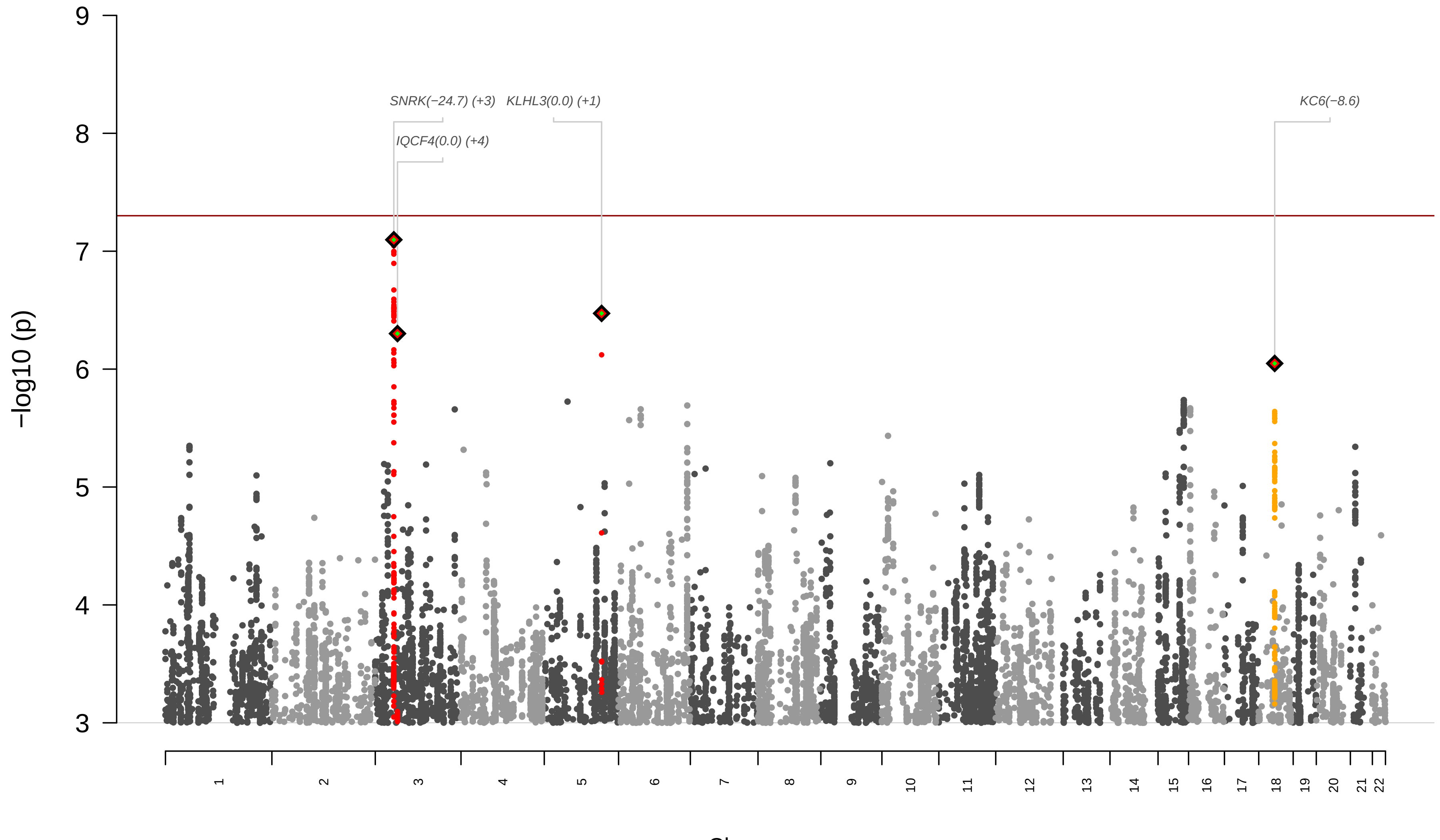

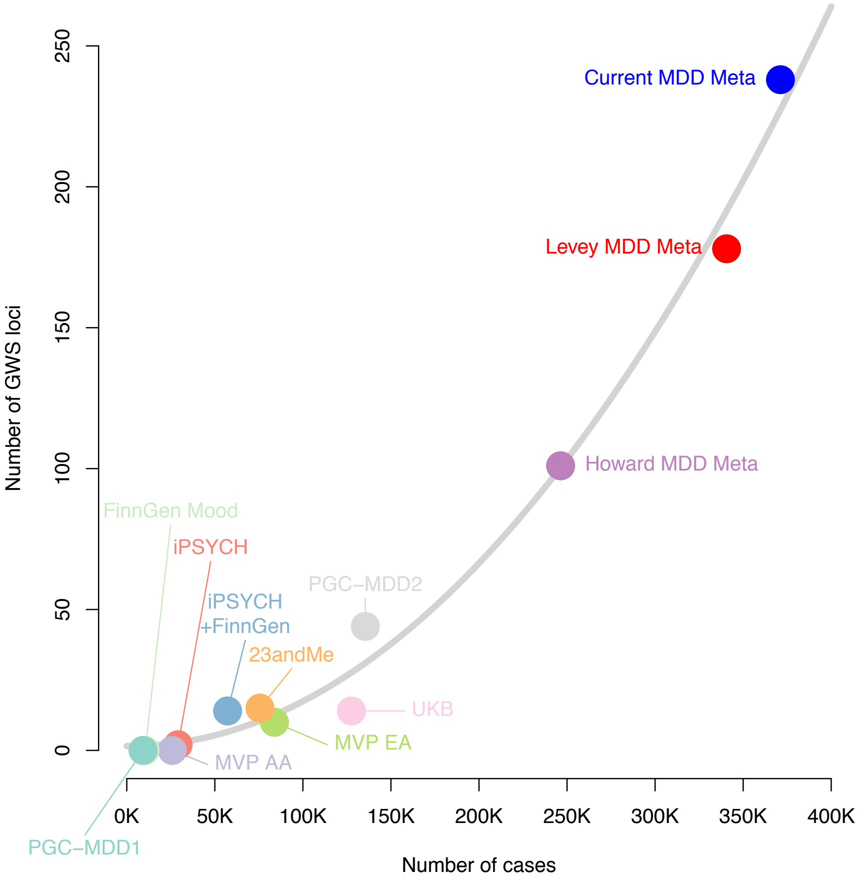

Figure S4: Number of loci as a function of the number of MD-cases. Figure adapted from Levey, D. F. et al. Nat Neurosci 24, 954-963, 2021.

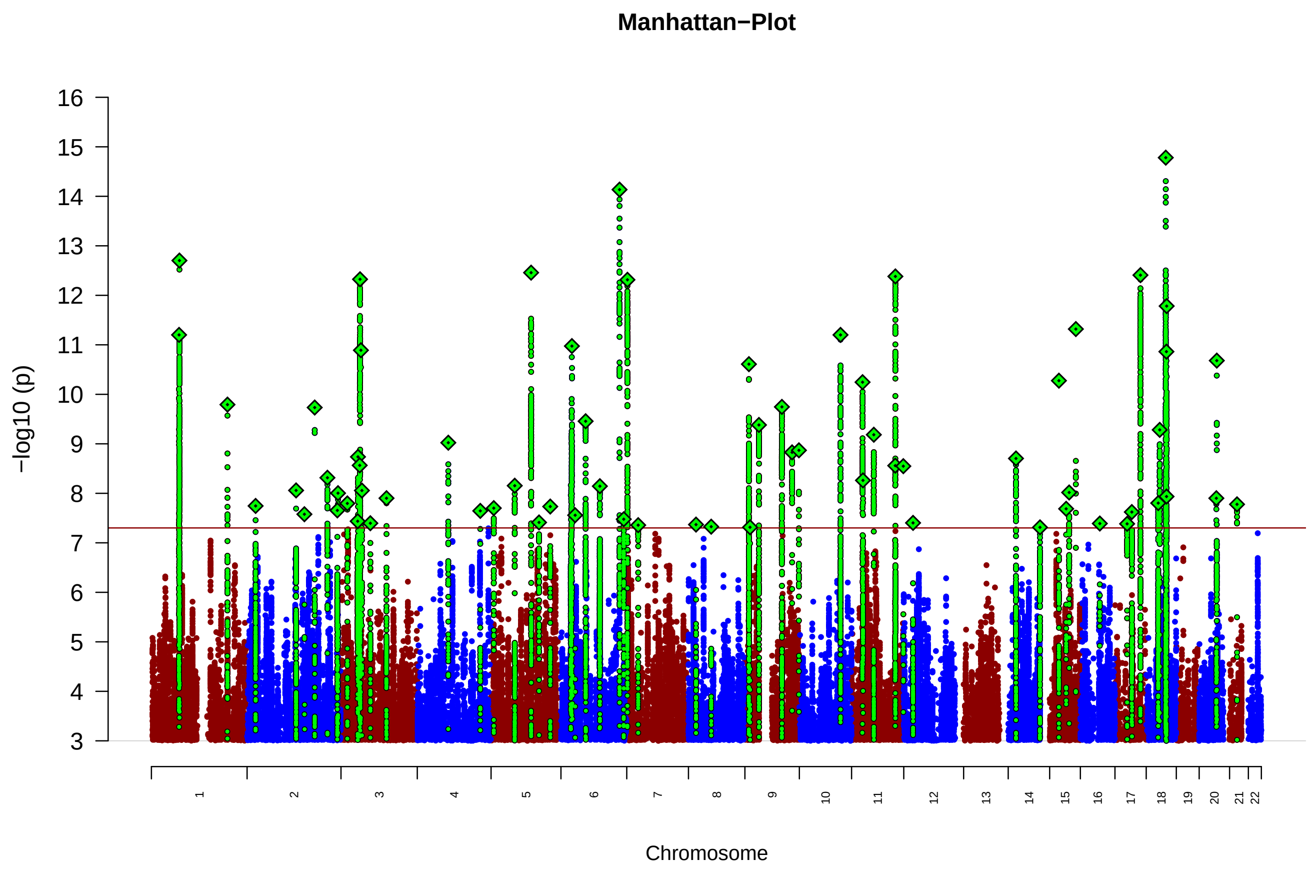

Figure S5: Manhattan plot of the meta analysis of narrow MD definition (N = 176143 cases and 528770 controls), with 67 genome-wide significant loci (76 independent SNPs)

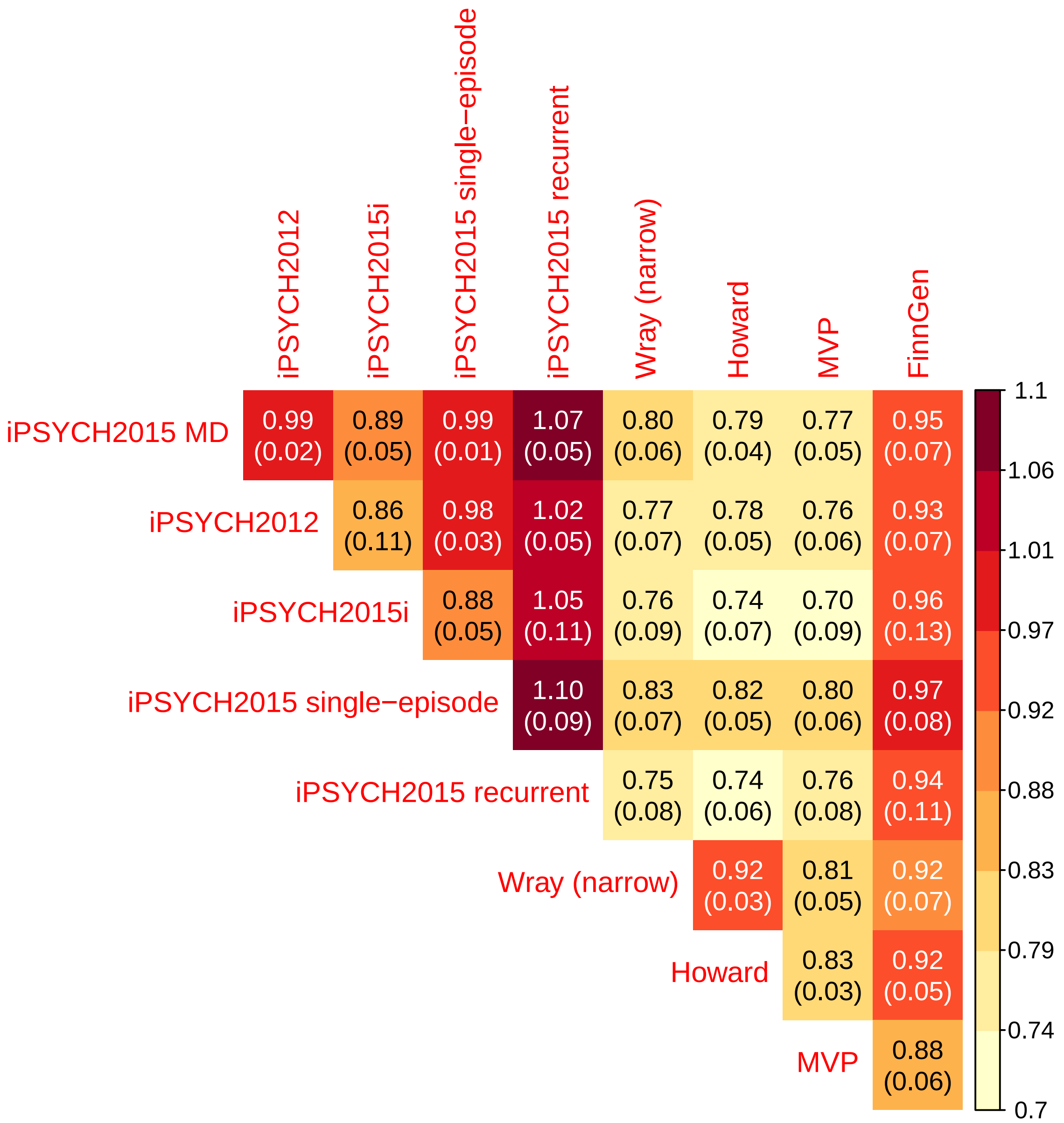

Figure S6: Genetic correlations among MD summary stats and MD subtypes within iPSYCH2015 using LD-score regression. $r_G(s.e.)$ is genetic correlation and standard error. iPSYCH2015 MD (N = 29158 cases and 38142 controls) is from a GWAS based on both the initial iPSYCH2012 (N = 20158 cases and 22750 controls) and the iPSYCH2015i extension (N = 10460 cases and 15450 controls). iPSYCH2015 single-episode is cases with only a single episode of depression (N = 23140 cases and 38142 controls), while the iPSYCH2015 recurrent GWAS is based on individuals that have experienced at least two seperate episodes of depression (N = 6018 cases and 38142 controls). Wray (narrow) is the PGC2 MD GWAS from Wray et al. 2018 excluding cohorts with self-reported MD-diagnoses (N = 35077 cases and 95406 controls). The $r_G$ of Howard is based on the GWAS published in Howard et al. 2019 (N = 230118 cases and 545339 controls). The $r_G$ of MVP is based on the Million Veteran Program MD-GWAS published in Levey et al. 2021 (N = 83810 cases and 166405 controls).

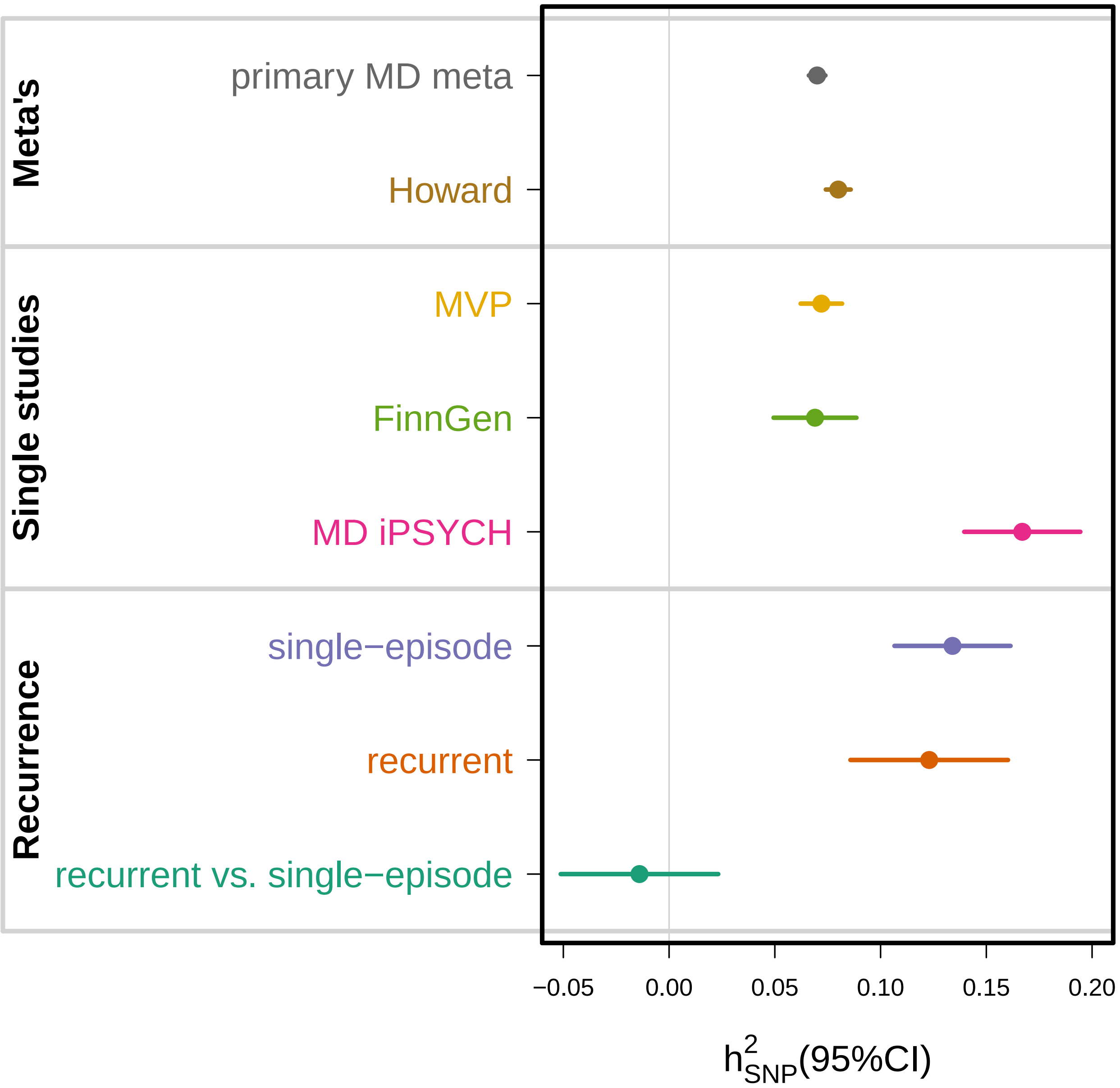

Figure S7: Heritability estimates ($h^2_{SNP}$) based on summary stats using LD-score regression. Error bars are 95% confidence intervals. The heritability estimate of the primary MD meta-analysis is based on (N = 371184 cases and 978703 controls). The $h^2_{SNP}$ of Howard is based on the GWAS published in Howard et al. 2019 (N = 230118 cases and 545339 controls). The $h^2_{SNP}$ of MVP is based on the Million Veteran Program MD-GWAS published in Levey et al. 2021 (N = 83810 cases and 166405 controls). MD iPSYCH2015 is based on 29158 cases and 38142 controls. iPSYCH2015 single-episode is cases with only a single episode of depression (N = 23140 cases and 38142 controls), while the iPSYCH2015 recurrent GWAS is based on individuals that have experienced at least two seperate episodes of depression (N = 6018 cases and 38142 controls). Recurrent vs. single-episode estimate is based on a GWAS considering individuals with recurrent depression as cases (N = 6018 ) and individuals with single-episode as controls (N = 23140 ) .

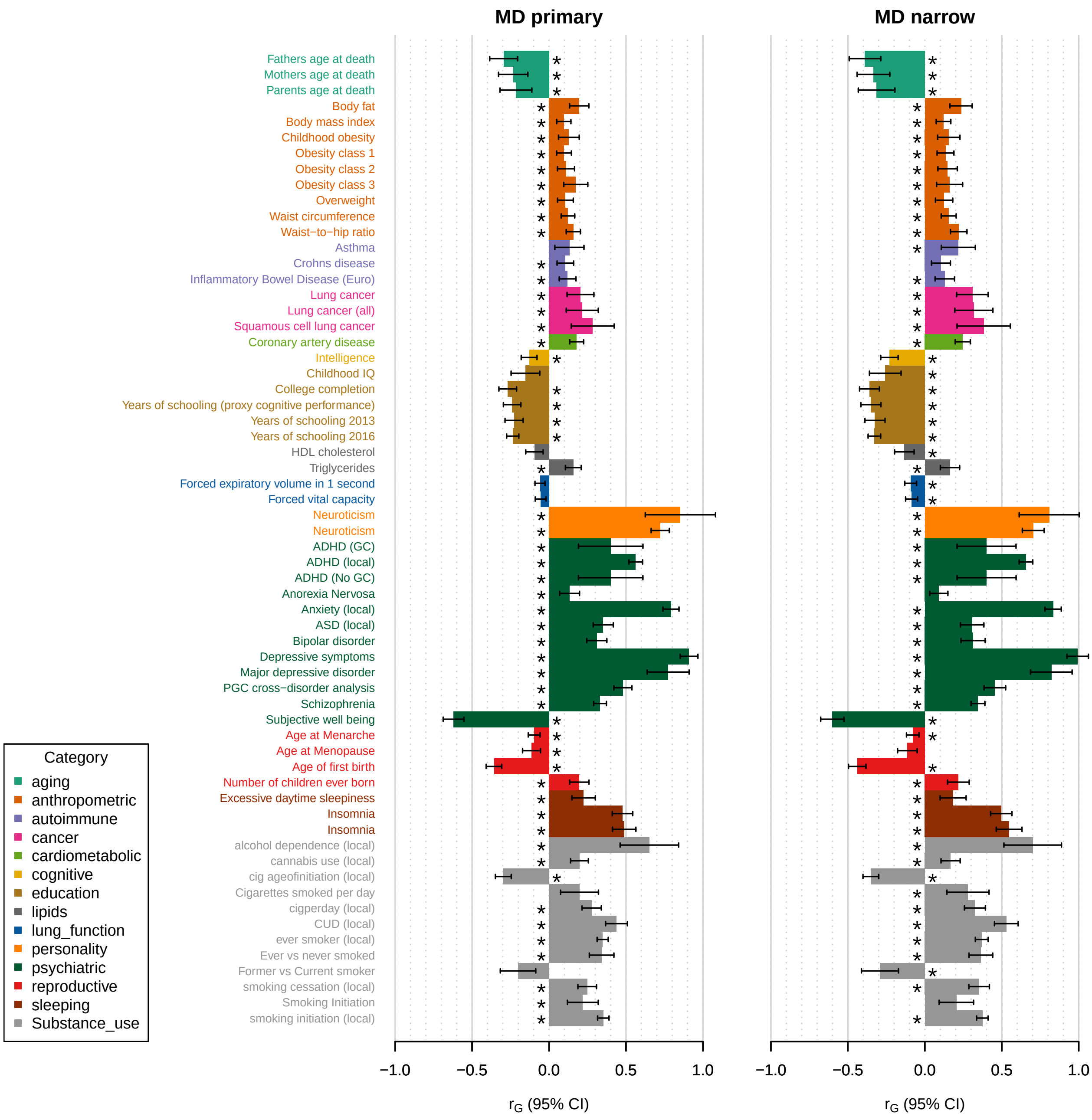

Figure S8A: Genetic correlations $r_G$ of the primary ($N_{cases}$ = 371184 and $N_{ctrls}$ = 978703) and narrow MD ($N_{cases}$ = 176143 and $N_{ctrls}$ = 528770) GWAS with other phenotypes (LD-hub). Non-UKBB data: A total of 55 (62) traits appeared to be significantly correlated with the MD (and/or MD narrow) phenotype. LD-hub traits were supplemented with updated or in-house GWAS summary statistics for ADHD, ASD{Grove, 2019 #3760}, cannabis use disorder{Johnson, 2020 #3800}, cannabis use{Pasman, 2018 #3804}, alcohol dependence{Walters, 2018 #3801}, drinks per week, smoking ever{Karlsson Linner, 2019 #3803}, age of initiation as a regular smoker, current versus a former smoker (smoking cessation), number of cigarettes per day, smoking initiation{Liu, 2019 #3802}

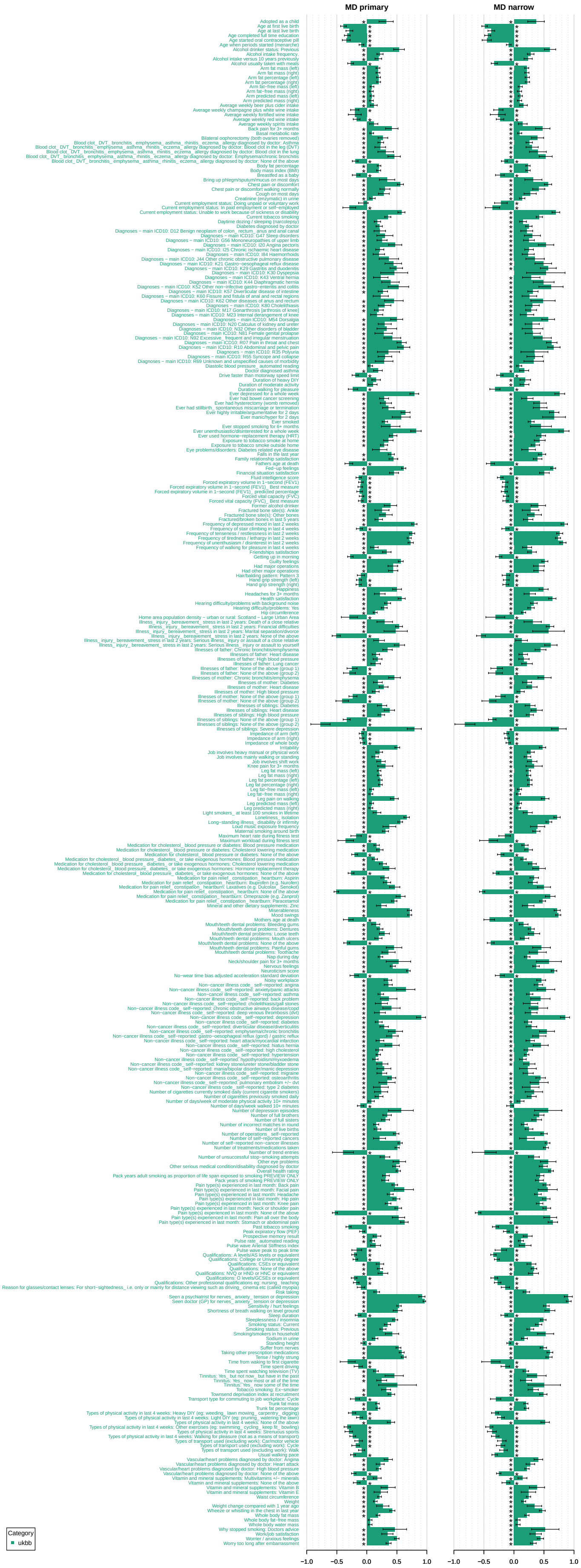

Figure S8B: Genetic correlations $r_G$ of the primary ($N_{cases}$ = 371184 and $N_{ctrls}$ = 978703) and narrow MD ($N_{cases}$ = 176143 and $N_{ctrls}$ = 528770) GWAS with other phenotypes (LD-hub). UKBB data: A total of 310 (328) traits appeared to be significantly correlated with the MD (and/or MD narrow) phenotype.

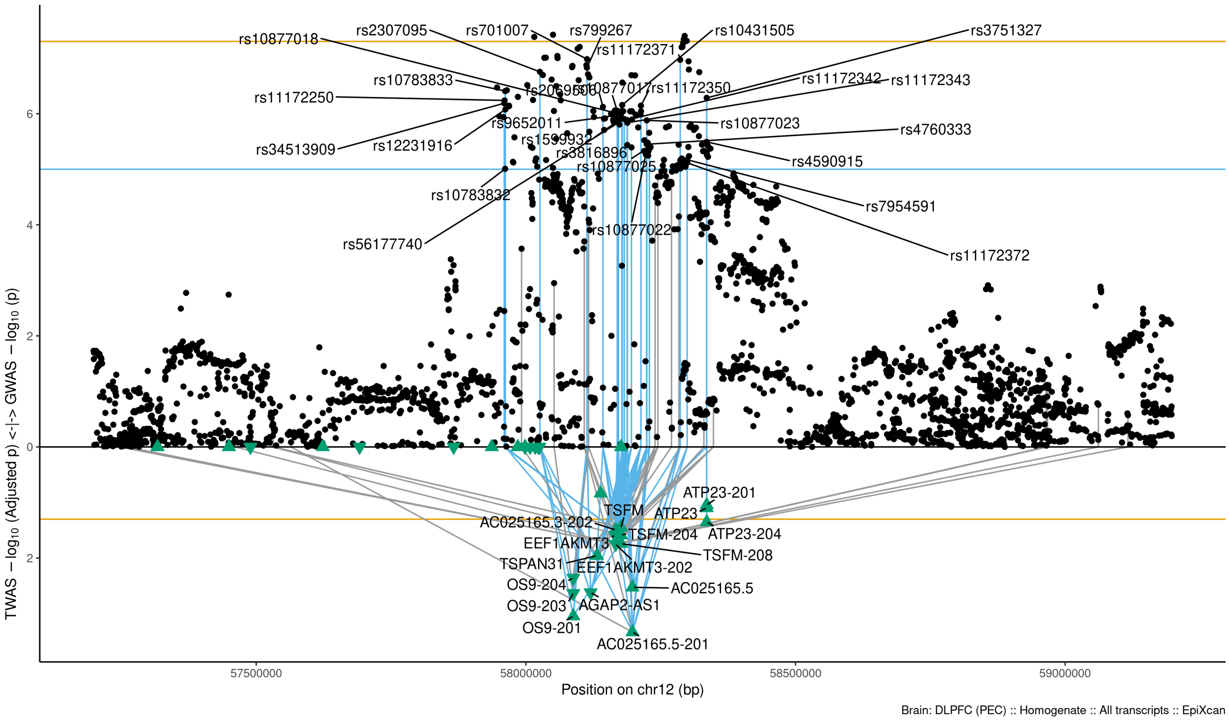

Figure S9-1: Regional Miami plots for AC025165201 gene/transcript : Regional Miami plots for the primary MD GWASs corresponding to the genomic region of the AC025165201 transcript (1Mbp window from start site). Please also refer to Supplemental Table SXX for details. Top panel shows the GWAS results (black dots); blue line corresponds to $p = 1 \times 10 - 5$, orange line to $p = 5 \times 10 - 8$ (genome-wide significance). Bottom panel shows the TWAS results (green triangles facing upwards or downwards for a positive or negative z-score respectively (up- or down-regultion); only the transcripts with Bonferroni-adjusted $p < 0.1$ are labelled for clarity) for different transcripts (genes are represented by both gene expression and isoform expression); orange line corresponds to Bonferroni-adjusted p = 0.05. Each transcript that is Bonferroni-significant in the region is connected with lines to the SNPs that contribute to its transcriptomic imputation model; lines are grey when the SNPs have a $p > 1 \times 10 - 5$, blue when $p < 1 \times 10 - 5$ but $> 510 - 8$ and orange when $p < 5 \times 10 - 8$. The SNPs that are above the blue line and contribute to the transcriptomic imputation models of significant transcripts are labelled.

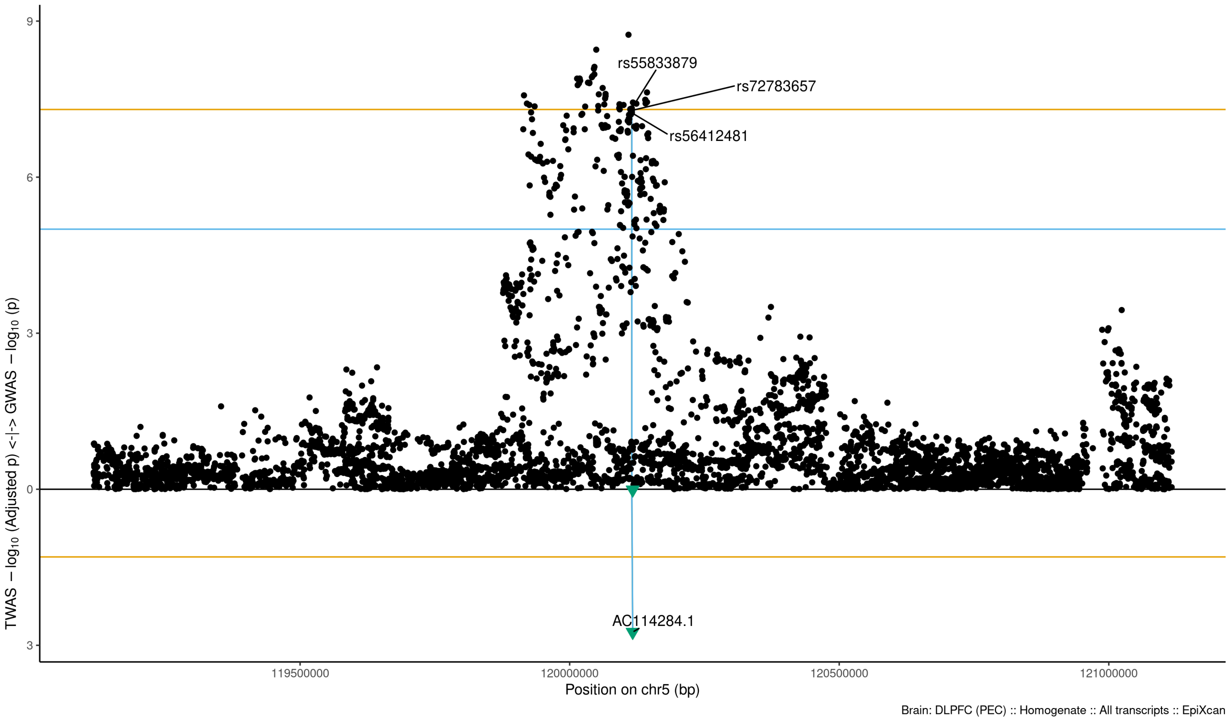

Figure S9-2: Regional Miami plots for AC114284 gene/transcript : Regional Miami plots for the primary MD GWASs corresponding to the genomic region of the AC114284 transcript (1Mbp window from start site). Please also refer to Supplemental Table SXX for details. Top panel shows the GWAS results (black dots); blue line corresponds to $p = 1 \times 10 - 5$, orange line to $p = 5 \times 10 - 8$ (genome-wide significance). Bottom panel shows the TWAS results (green triangles facing upwards or downwards for a positive or negative z-score respectively (up- or down-regultion); only the transcripts with Bonferroni-adjusted $p < 0.1$ are labelled for clarity) for different transcripts (genes are represented by both gene expression and isoform expression); orange line corresponds to Bonferroni-adjusted p = 0.05. Each transcript that is Bonferroni-significant in the region is connected with lines to the SNPs that contribute to its transcriptomic imputation model; lines are grey when the SNPs have a $p > 1 \times 10 - 5$, blue when $p < 1 \times 10 - 5$ but $> 510 - 8$ and orange when $p < 5 \times 10 - 8$. The SNPs that are above the blue line and contribute to the transcriptomic imputation models of significant transcripts are labelled.

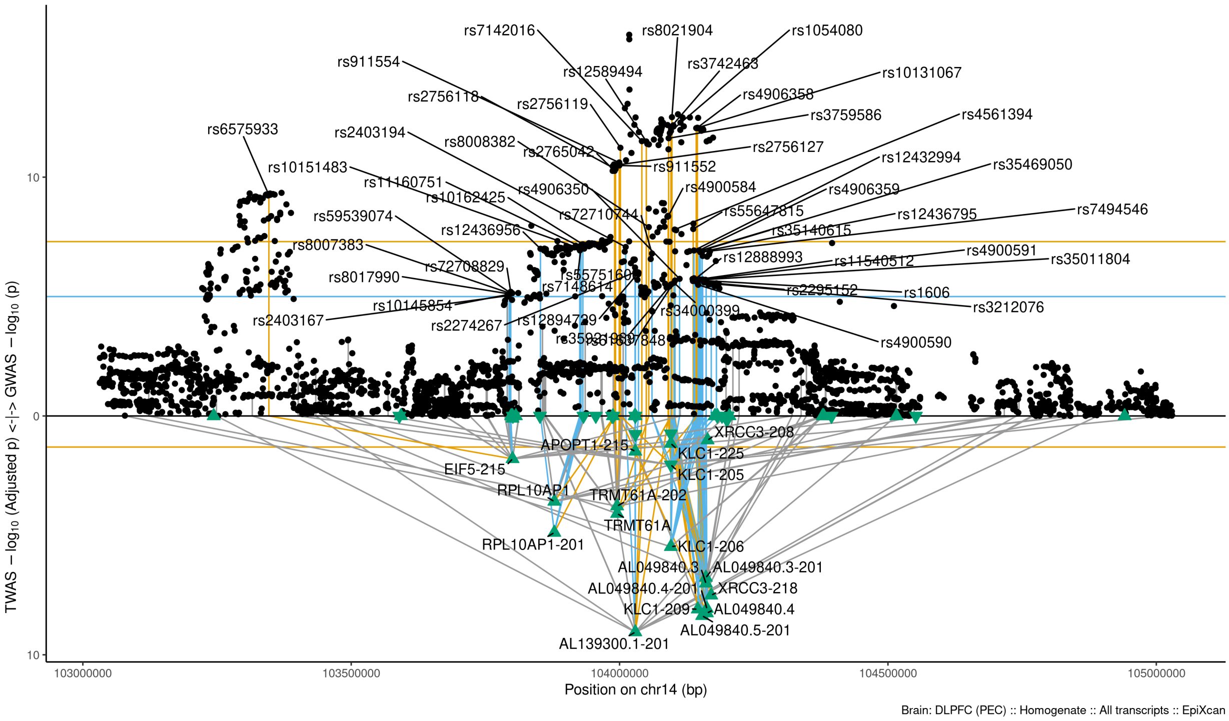

Figure S9-3: Regional Miami plots for AL139300201 gene/transcript : Regional Miami plots for the primary MD GWASs corresponding to the genomic region of the AL139300201 transcript (1Mbp window from start site). Please also refer to Supplemental Table SXX for details. Top panel shows the GWAS results (black dots); blue line corresponds to $p = 1 \times 10-5$, orange line to $p = 5 \times 10-8$ (genome-wide significance). Bottom panel shows the TWAS results (green triangles facing upwards or downwards for a positive or negative z-score respectively (up- or down-regultion); only the transcripts with Bonferroni-adjusted $p < 0.1$ are labelled for clarity) for different transcripts (genes are represented by both gene expression and isoform expression); orange line corresponds to Bonferroni-adjusted p = 0.05. Each transcript that is Bonferroni-significant in the region is connected with lines to the SNPs that contribute to its transcriptomic imputation model; lines are grey when the SNPs have a $p > 1 \times 10-5$, blue when $p < 1 \times 10-5$ but $> 510-8$ and orange when $p < 5 \times 10-8$. The SNPs that are above the blue line and contribute to the transcriptomic imputation models of significant transcripts are labelled.

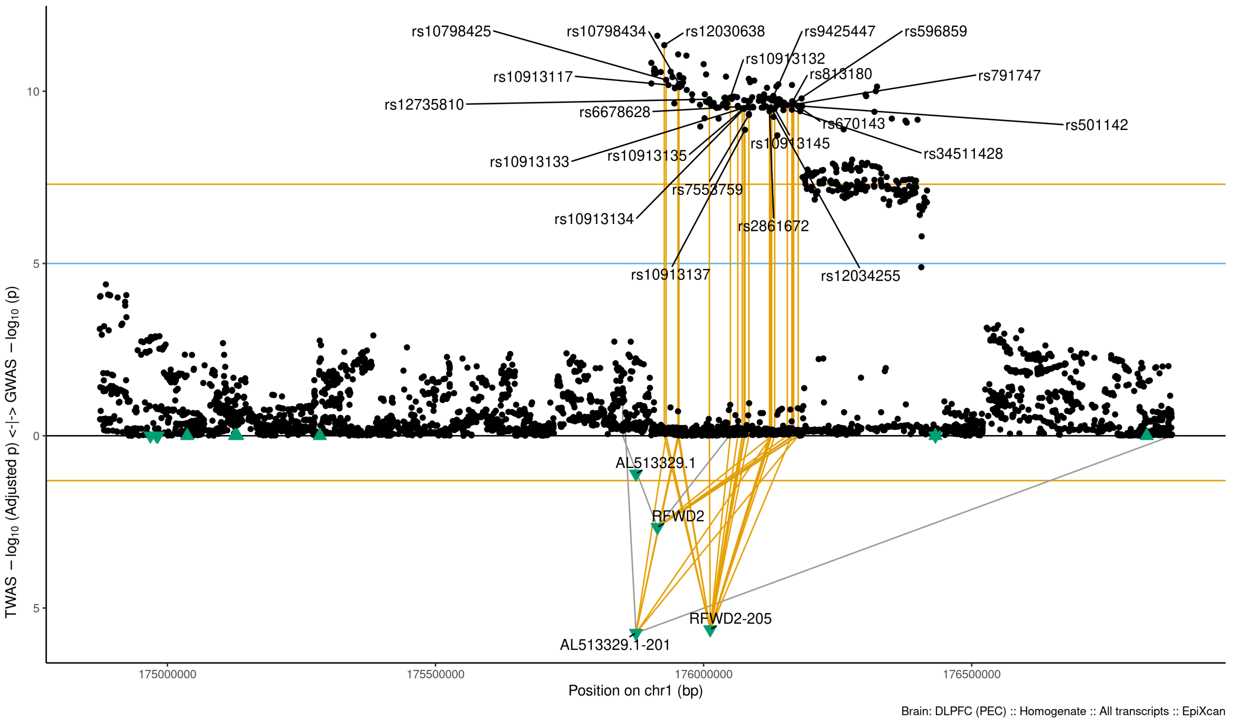

Figure S9-4: Regional Miami plots for AL513329201 gene/transcript : Regional Miami plots for the primary MD GWASs corresponding to the genomic region of the AL513329201 transcript (1Mbp window from start site). Please also refer to Supplemental Table SXX for details. Top panel shows the GWAS results (black dots); blue line corresponds to $p = 1 \times 10 - 5$, orange line to $p = 5 \times 10 - 8$ (genome-wide significance). Bottom panel shows the TWAS results (green triangles facing upwards or downwards for a positive or negative z-score respectively (up- or down-regultion); only the transcripts with Bonferroni-adjusted $p < 0.1$ are labelled for clarity) for different transcripts (genes are represented by both gene expression and isoform expression); orange line corresponds to Bonferroni-adjusted p = 0.05. Each transcript that is Bonferroni-significant in the region is connected with lines to the SNPs that contribute to its transcriptomic imputation model; lines are grey when the SNPs have a $p > 1 \times 10 - 5$, blue when $p < 1 \times 10 - 5$ but $> 510 - 8$ and orange when $p < 5 \times 10 - 8$. The SNPs that are above the blue line and contribute to the transcriptomic imputation models of significant transcripts are labelled.

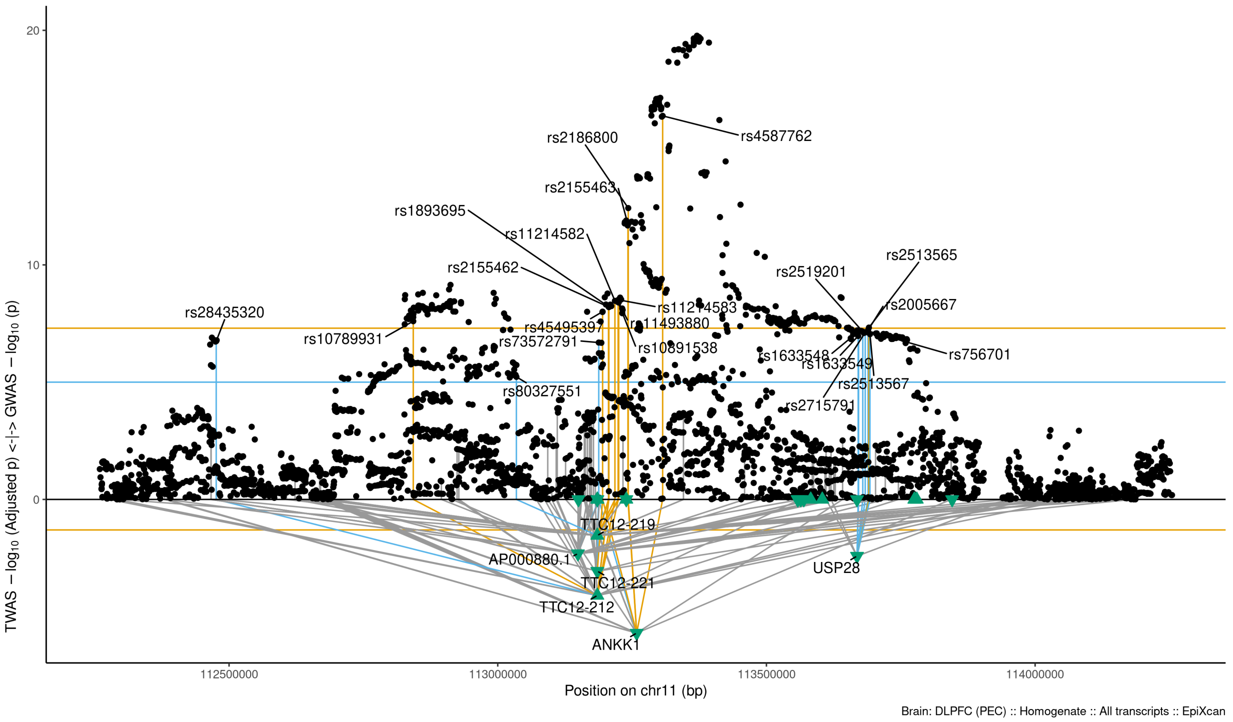

Figure S9-5: Regional Miami plots for ANKK1 gene/transcript : Regional Miami plots for the primary MD GWASs corresponding to the genomic region of the ANKK1 transcript (1Mbp window from start site). Please also refer to Supplemental Table SXX for details. Top panel shows the GWAS results (black dots); blue line corresponds to $p = 1 \times 10 - 5$, orange line to $p = 5 \times 10 - 8$ (genome-wide significance). Bottom panel shows the TWAS results (green triangles facing upwards or downwards for a positive or negative z-score respectively (up- or down-regultion); only the transcripts with Bonferroni-adjusted $p < 0.1$ are labelled for clarity) for different transcripts (genes are represented by both gene expression and isoform expression); orange line corresponds to Bonferroni-adjusted p = 0.05. Each transcript that is Bonferroni-significant in the region is connected with lines to the SNPs that contribute to its transcriptomic imputation model; lines are grey when the SNPs have a $p > 1 \times 10 - 5$, blue when $p < 1 \times 10 - 5$ but $> 510 - 8$ and orange when $p < 5 \times 10 - 8$. The SNPs that are above the blue line and contribute to the transcriptomic imputation models of significant transcripts are labelled.

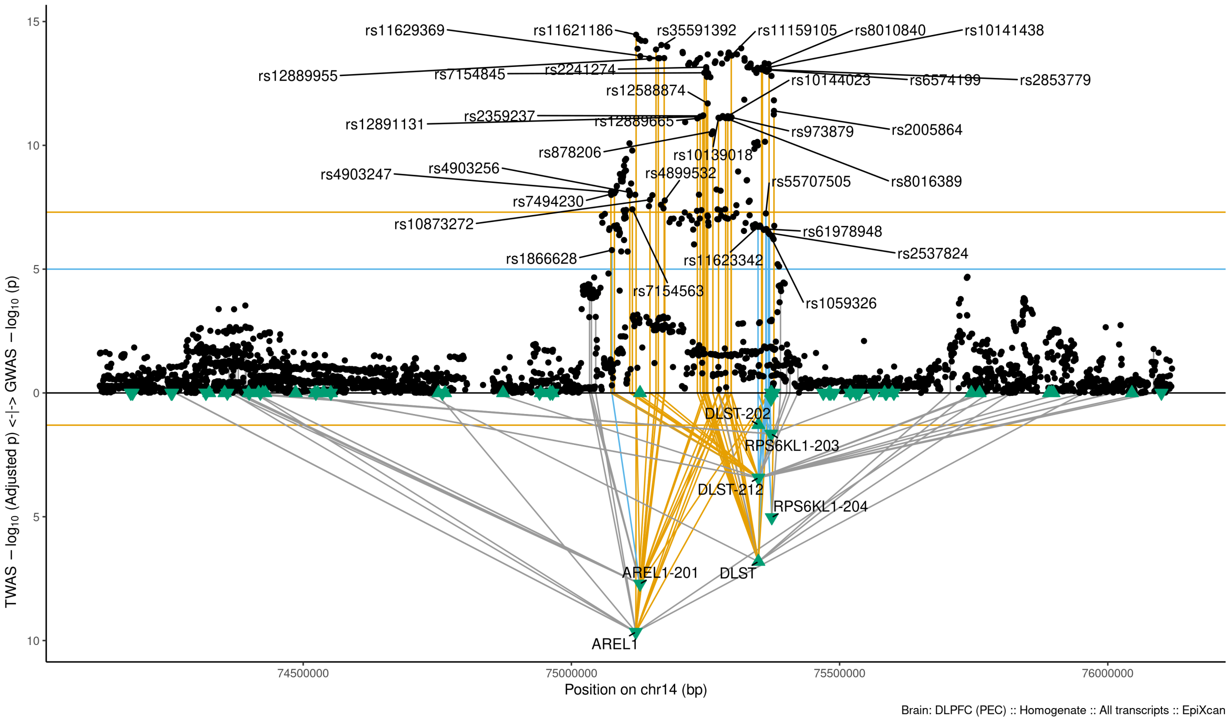

Figure S9-6: Regional Miami plots for AREL1 gene/transcript : Regional Miami plots for the primary MD GWASs corresponding to the genomic region of the AREL1 transcript (1Mbp window from start site). Please also refer to Supplemental Table SXX for details. Top panel shows the GWAS results (black dots); blue line corresponds to $p = 1 \times 10 - 5$, orange line to $p = 5 \times 10 - 8$ (genome-wide significance). Bottom panel shows the TWAS results (green triangles facing upwards or downwards for a positive or negative z-score respectively (up- or down-regultion); only the transcripts with Bonferroni-adjusted $p < 0.1$ are labelled for clarity) for different transcripts (genes are represented by both gene expression and isoform expression); orange line corresponds to Bonferroni-adjusted p = 0.05. Each transcript that is Bonferroni-significant in the region is connected with lines to the SNPs that contribute to its transcriptomic imputation model; lines are grey when the SNPs have a $p > 1 \times 10 - 5$, blue when $p < 1 \times 10 - 5$ but $> 510 - 8$ and orange when $p < 5 \times 10 - 8$. The SNPs that are above the blue line and contribute to the transcriptomic imputation models of significant transcripts are labelled.

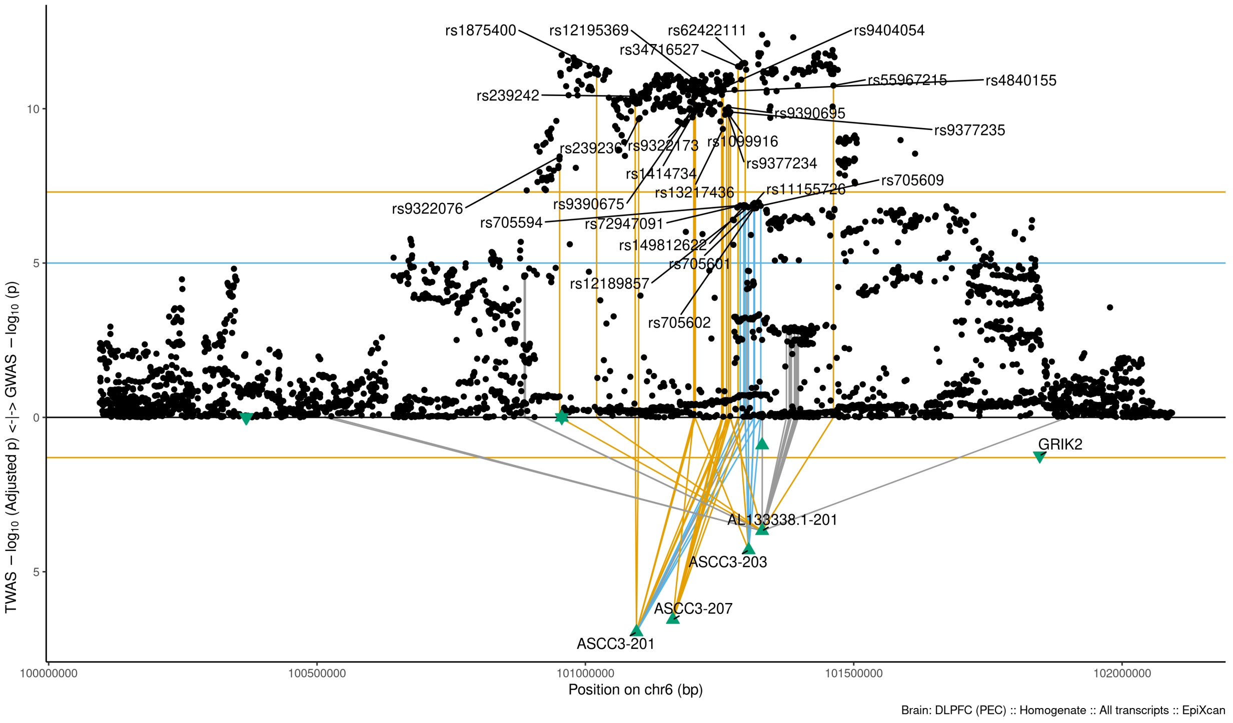

Figure S9-7: Regional Miami plots for ASCC3201 gene/transcript : Regional Miami plots for the primary MD GWASs corresponding to the genomic region of the ASCC3201 transcript (1Mbp window from start site). Please also refer to Supplemental Table SXX for details. Top panel shows the GWAS results (black dots); blue line corresponds to $p = 1 \times 10 - 5$, orange line to $p = 5 \times 10 - 8$ (genome-wide significance). Bottom panel shows the TWAS results (green triangles facing upwards or downwards for a positive or negative z-score respectively (up- or down-regultion); only the transcripts with Bonferroni-adjusted $p < 0.1$ are labelled for clarity) for different transcripts (genes are represented by both gene expression and isoform expression); orange line corresponds to Bonferroni-adjusted p = 0.05. Each transcript that is Bonferroni-significant in the region is connected with lines to the SNPs that contribute to its transcriptomic imputation model; lines are grey when the SNPs have a $p > 1 \times 10 - 5$, blue when $p < 1 \times 10 - 5$ but $> 510 - 8$ and orange when $p < 5 \times 10 - 8$. The SNPs that are above the blue line and contribute to the transcriptomic imputation models of significant transcripts are labelled.

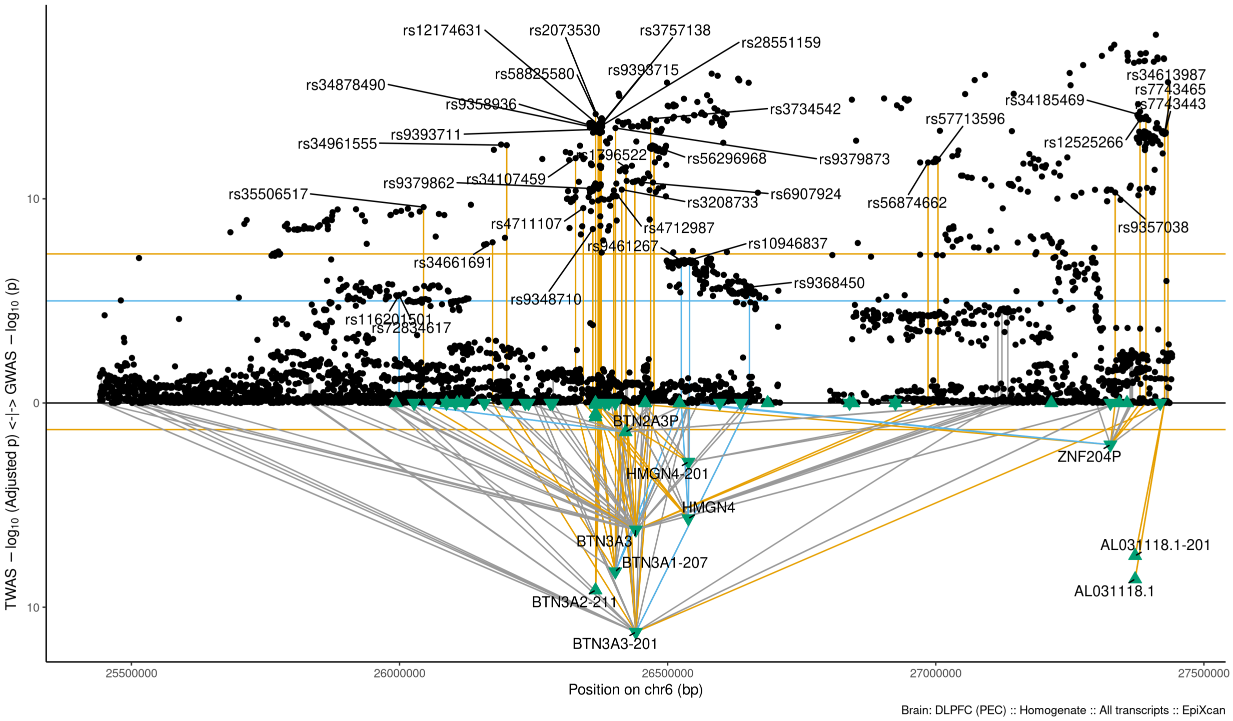

Figure S9-8: Regional Miami plots for BTN3A3201 gene/transcript : Regional Miami plots for the primary MD GWASs corresponding to the genomic region of the BTN3A3201 transcript (1Mbp window from start site). Please also refer to Supplemental Table SXX for details. Top panel shows the GWAS results (black dots); blue line corresponds to $p = 1 \times 10 - 5$, orange line to $p = 5 \times 10 - 8$ (genome-wide significance). Bottom panel shows the TWAS results (green triangles facing upwards or downwards for a positive or negative z-score respectively (up- or down-regultion); only the transcripts with Bonferroni-adjusted $p < 0.1$ are labelled for clarity) for different transcripts (genes are represented by both gene expression and isoform expression); orange line corresponds to Bonferroni-adjusted p = 0.05. Each transcript that is Bonferroni-significant in the region is connected with lines to the SNPs that contribute to its transcriptomic imputation model; lines are grey when the SNPs have a $p > 1 \times 10 - 5$, blue when $p < 1 \times 10 - 5$ but $> 510 - 8$ and orange when $p < 5 \times 10 - 8$. The SNPs that are above the blue line and contribute to the transcriptomic imputation models of significant transcripts are labelled.

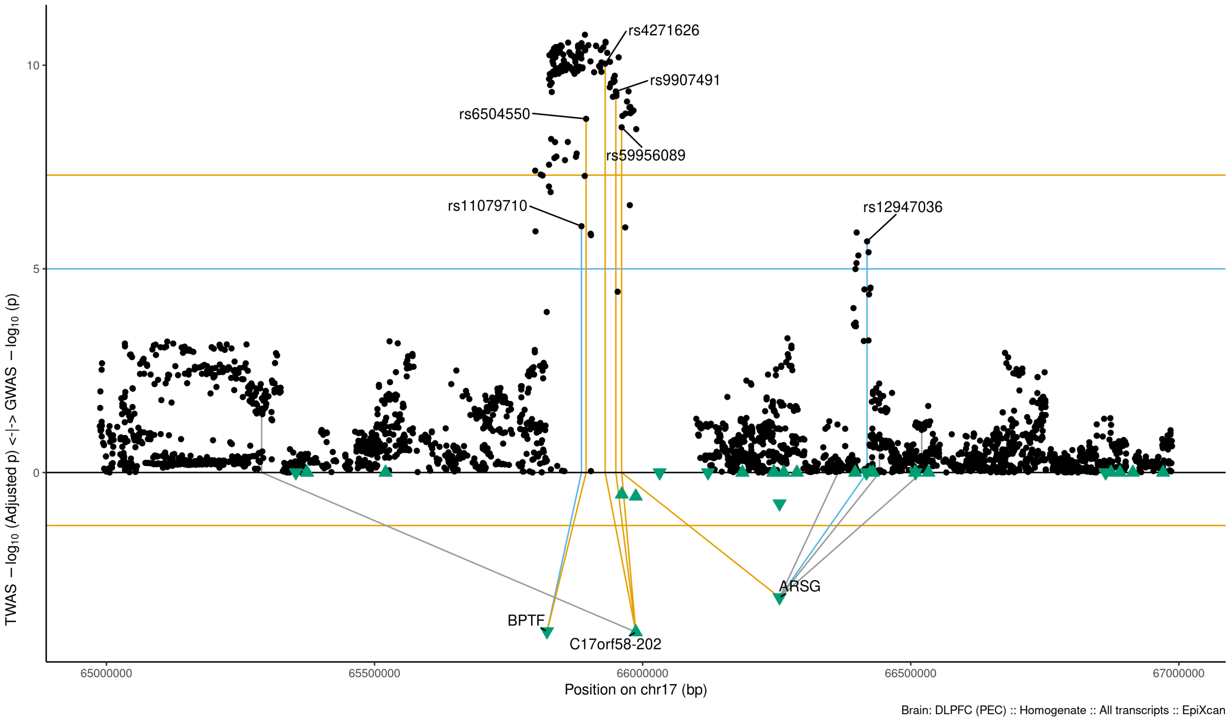

Figure S9-9: Regional Miami plots for C17orf58202 gene/transcript : Regional Miami plots for the primary MD GWASs corresponding to the genomic region of the C17orf58202 transcript (1Mbp window from start site). Please also refer to Supplemental Table SXX for details. Top panel shows the GWAS results (black dots); blue line corresponds to $p = 1 \times 10 - 5$, orange line to $p = 5 \times 10 - 8$ (genome-wide significance). Bottom panel shows the TWAS results (green triangles facing upwards or downwards for a positive or negative z-score respectively (up- or down-regultion); only the transcripts with Bonferroni-adjusted $p < 0.1$ are labelled for clarity) for different transcripts (genes are represented by both gene expression and isoform expression); orange line corresponds to Bonferroni-adjusted p = 0.05. Each transcript that is Bonferroni-significant in the region is connected with lines to the SNPs that contribute to its transcriptomic imputation model; lines are grey when the SNPs have a $p > 1 \times 10 - 5$, blue when $p < 1 \times 10 - 5$ but $> 510 - 8$ and orange when $p < 5 \times 10 - 8$. The SNPs that are above the blue line and contribute to the transcriptomic imputation models of significant transcripts are labelled.

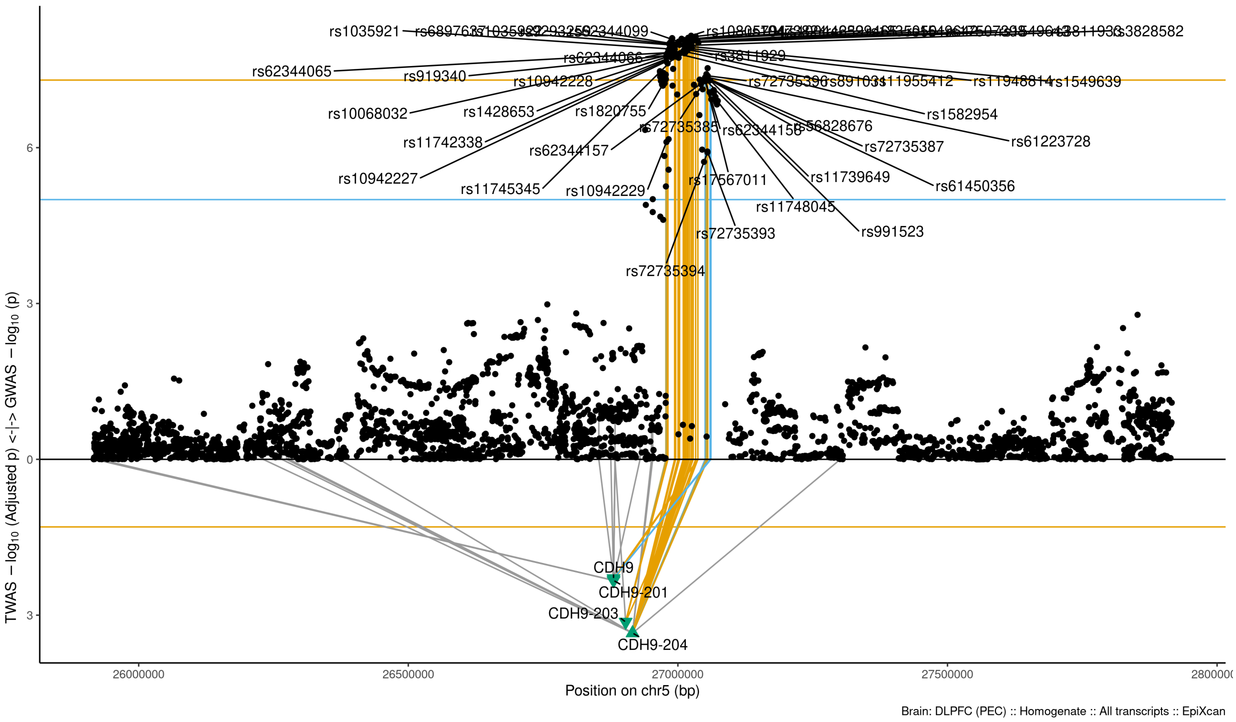

Figure S9-10: Regional Miami plots for CDH9204 gene/transcript : Regional Miami plots for the primary MD GWASs corresponding to the genomic region of the CDH9204 transcript (1Mbp window from start site). Please also refer to Supplemental Table SXX for details. Top panel shows the GWAS results (black dots); blue line corresponds to $p = 1 \times 10 - 5$, orange line to $p = 5 \times 10 - 8$ (genome-wide significance). Bottom panel shows the TWAS results (green triangles facing upwards or downwards for a positive or negative z-score respectively (up- or down-regultion); only the transcripts with Bonferroni-adjusted $p < 0.1$ are labelled for clarity) for different transcripts (genes are represented by both gene expression and isoform expression); orange line corresponds to Bonferroni-adjusted p = 0.05. Each transcript that is Bonferroni-significant in the region is connected with lines to the SNPs that contribute to its transcriptomic imputation model; lines are grey when the SNPs have a $p > 1 \times 10 - 5$, blue when $p < 1 \times 10 - 5$ but $> 510 - 8$ and orange when $p < 5 \times 10 - 8$. The SNPs that are above the blue line and contribute to the transcriptomic imputation models of significant transcripts are labelled.

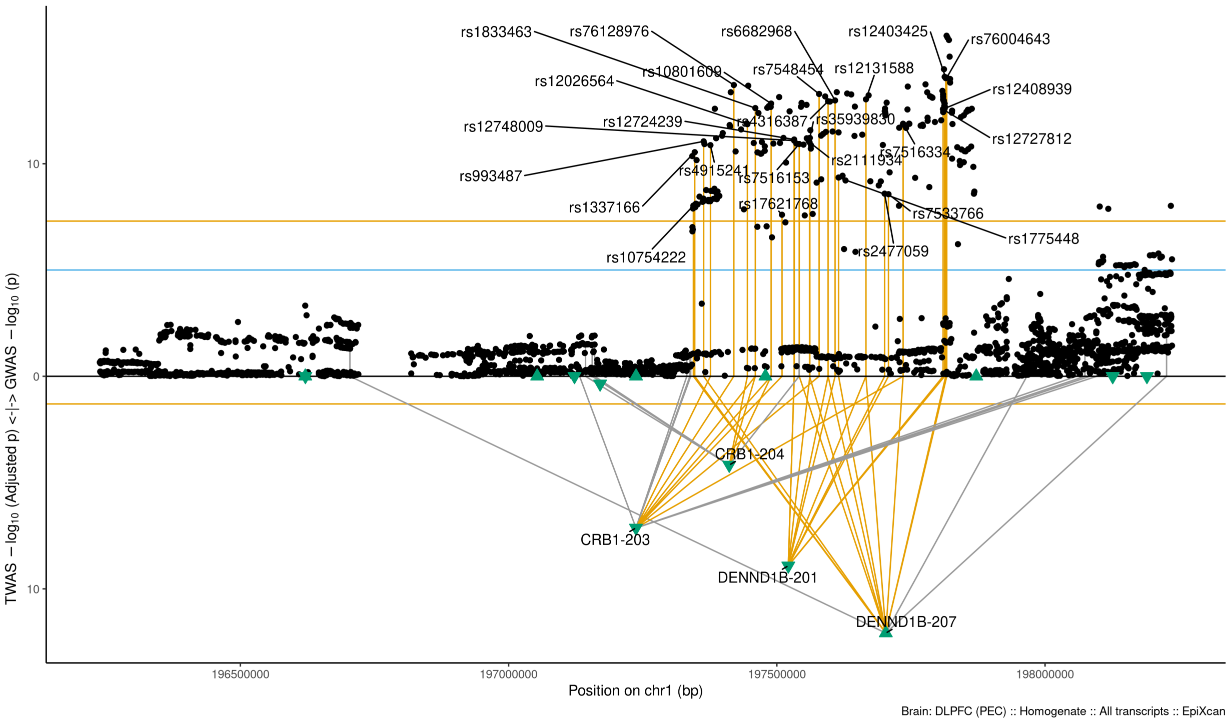

Figure S9-11: Regional Miami plots for CRB1203 gene/transcript : Regional Miami plots for the primary MD GWASs corresponding to the genomic region of the CRB1203 transcript (1Mbp window from start site). Please also refer to Supplemental Table SXX for details. Top panel shows the GWAS results (black dots); blue line corresponds to $p = 1 \times 10 - 5$, orange line to $p = 5 \times 10 - 8$ (genome-wide significance). Bottom panel shows the TWAS results (green triangles facing upwards or downwards for a positive or negative z-score respectively (up- or down-regultion); only the transcripts with Bonferroni-adjusted $p < 0.1$ are labelled for clarity) for different transcripts (genes are represented by both gene expression and isoform expression); orange line corresponds to Bonferroni-adjusted p = 0.05. Each transcript that is Bonferroni-significant in the region is connected with lines to the SNPs that contribute to its transcriptomic imputation model; lines are grey when the SNPs have a $p > 1 \times 10 - 5$, blue when $p < 1 \times 10 - 5$ but $> 510 - 8$ and orange when $p < 5 \times 10 - 8$. The SNPs that are above the blue line and contribute to the transcriptomic imputation models of significant transcripts are labelled.

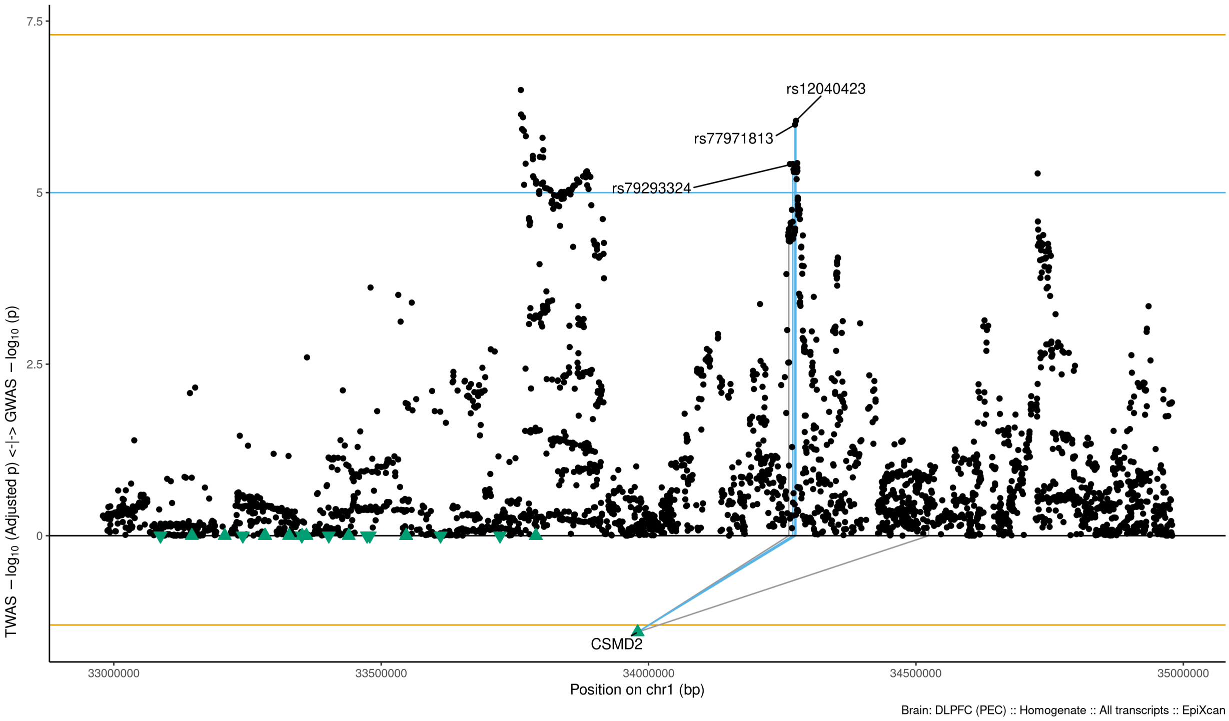

Figure S9-12: Regional Miami plots for CSMD2 gene/transcript : Regional Miami plots for the primary MD GWASs corresponding to the genomic region of the CSMD2 transcript (1Mbp window from start site). Please also refer to Supplemental Table SXX for details. Top panel shows the GWAS results (black dots); blue line corresponds to $p = 1 \times 10 - 5$, orange line to $p = 5 \times 10 - 8$ (genome-wide significance). Bottom panel shows the TWAS results (green triangles facing upwards or downwards for a positive or negative z-score respectively (up- or down-regultion); only the transcripts with Bonferroni-adjusted $p < 0.1$ are labelled for clarity) for different transcripts (genes are represented by both gene expression and isoform expression); orange line corresponds to Bonferroni-adjusted p = 0.05. Each transcript that is Bonferroni-significant in the region is connected with lines to the SNPs that contribute to its transcriptomic imputation model; lines are grey when the SNPs have a $p > 1 \times 10 - 5$, blue when $p < 1 \times 10 - 5$ but $> 510 - 8$ and orange when $p < 5 \times 10 - 8$. The SNPs that are above the blue line and contribute to the transcriptomic imputation models of significant transcripts are labelled.

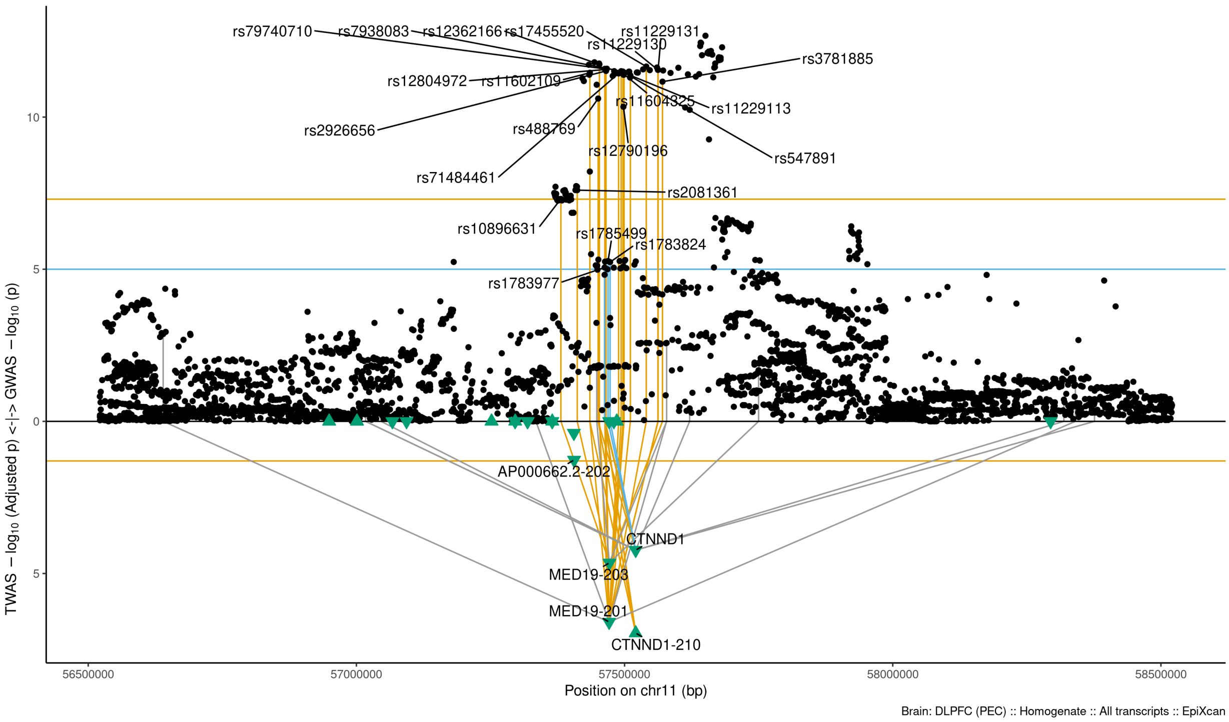

Figure S9-13: Regional Miami plots for CTNND1210 gene/transcript : Regional Miami plots for the primary MD GWASs corresponding to the genomic region of the CTNND1210 transcript (1Mbp window from start site). Please also refer to Supplemental Table SXX for details. Top panel shows the GWAS results (black dots); blue line corresponds to $p = 1 \times 10 - 5$, orange line to $p = 5 \times 10 - 8$ (genome-wide significance). Bottom panel shows the TWAS results (green triangles facing upwards or downwards for a positive or negative z-score respectively (up- or down-regultion); only the transcripts with Bonferroni-adjusted $p < 0.1$ are labelled for clarity) for different transcripts (genes are represented by both gene expression and isoform expression); orange line corresponds to Bonferroni-adjusted p = 0.05. Each transcript that is Bonferroni-significant in the region is connected with lines to the SNPs that contribute to its transcriptomic imputation model; lines are grey when the SNPs have a $p > 1 \times 10 - 5$, blue when $p < 1 \times 10 - 5$ but $> 510 - 8$ and orange when $p < 5 \times 10 - 8$. The SNPs that are above the blue line and contribute to the transcriptomic imputation models of significant transcripts are labelled.

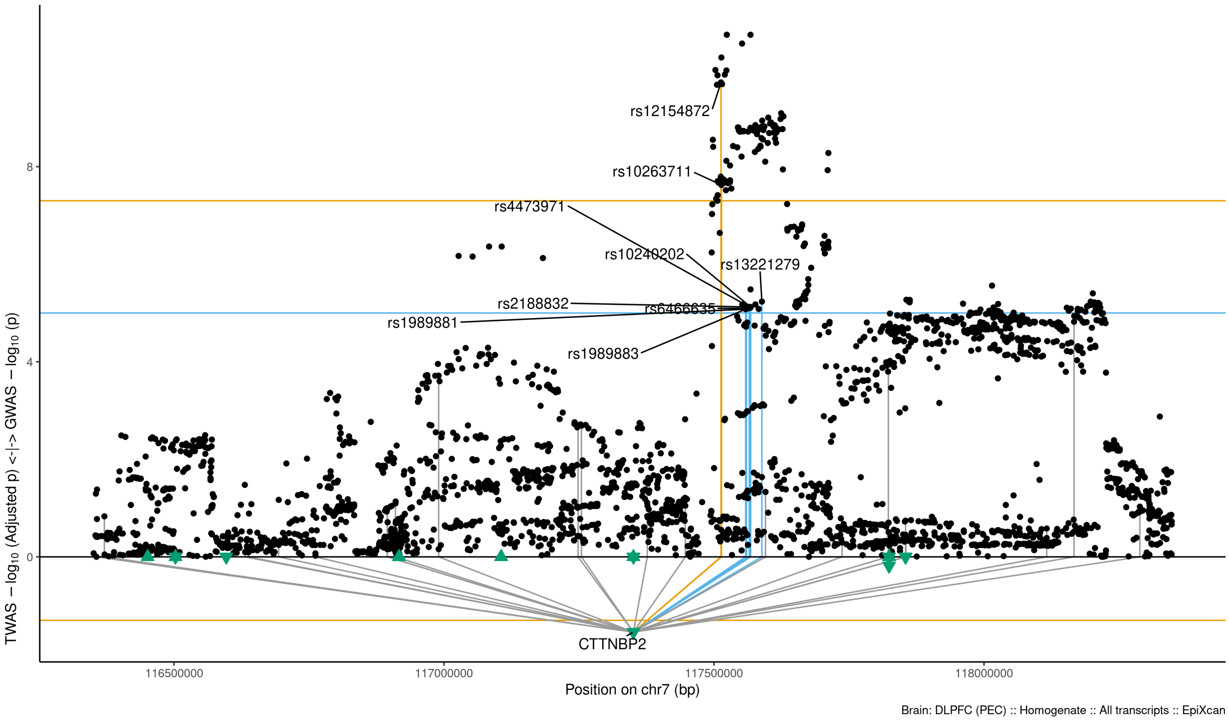

Figure S9-14: Regional Miami plots for CTTNBP2 gene/transcript : Regional Miami plots for the primary MD GWASs corresponding to the genomic region of the CTTNBP2 transcript (1Mbp window from start site). Please also refer to Supplemental Table SXX for details. Top panel shows the GWAS results (black dots); blue line corresponds to $p = 1 \times 10 - 5$, orange line to $p = 5 \times 10 - 8$ (genome-wide significance). Bottom panel shows the TWAS results (green triangles facing upwards or downwards for a positive or negative z-score respectively (up- or down-regultion); only the transcripts with Bonferroni-adjusted $p < 0.1$ are labelled for clarity) for different transcripts (genes are represented by both gene expression and isoform expression); orange line corresponds to Bonferroni-adjusted p = 0.05. Each transcript that is Bonferroni-significant in the region is connected with lines to the SNPs that contribute to its transcriptomic imputation model; lines are grey when the SNPs have a $p > 1 \times 10 - 5$, blue when $p < 1 \times 10 - 5$ but $> 510 - 8$ and orange when $p < 5 \times 10 - 8$. The SNPs that are above the blue line and contribute to the transcriptomic imputation models of significant transcripts are labelled.

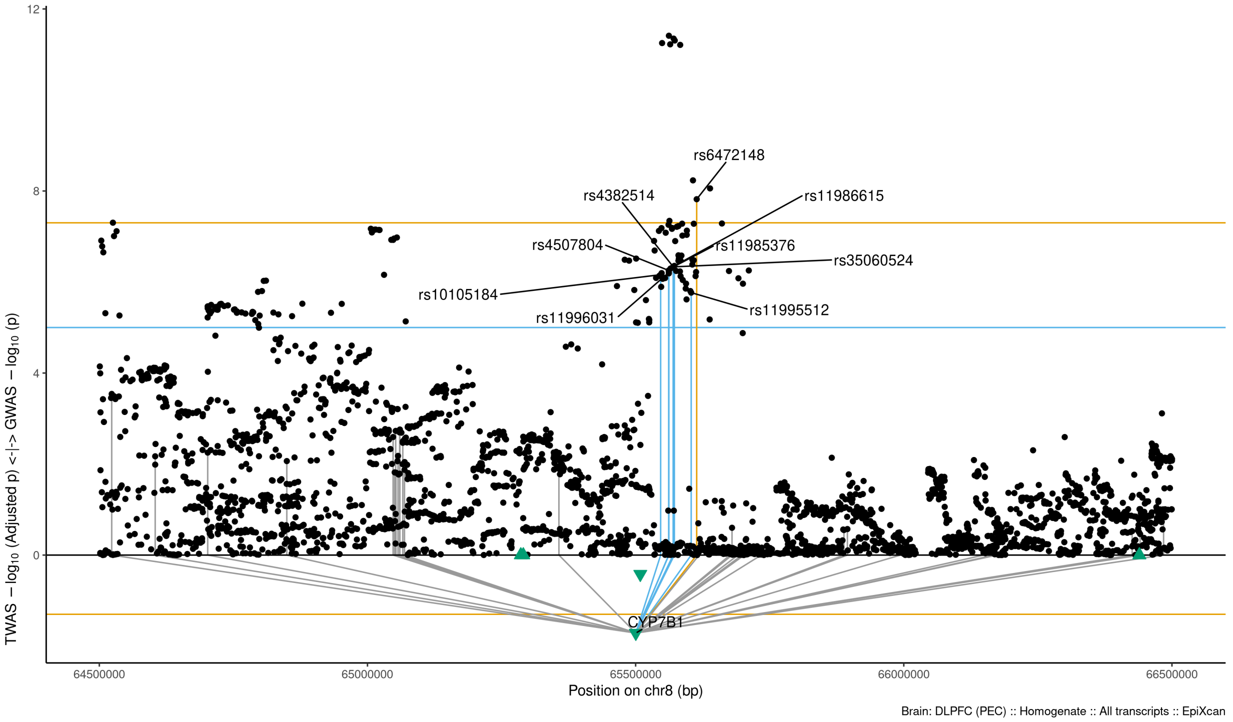

Figure S9-15: Regional Miami plots for CYP7B1 gene/transcript : Regional Miami plots for the primary MD GWASs corresponding to the genomic region of the CYP7B1 transcript (1Mbp window from start site). Please also refer to Supplemental Table SXX for details. Top panel shows the GWAS results (black dots); blue line corresponds to $p = 1 \times 10 - 5$, orange line to $p = 5 \times 10 - 8$ (genome-wide significance). Bottom panel shows the TWAS results (green triangles facing upwards or downwards for a positive or negative z-score respectively (up- or down-regultion); only the transcripts with Bonferroni-adjusted $p < 0.1$ are labelled for clarity) for different transcripts (genes are represented by both gene expression and isoform expression); orange line corresponds to Bonferroni-adjusted p = 0.05. Each transcript that is Bonferroni-significant in the region is connected with lines to the SNPs that contribute to its transcriptomic imputation model; lines are grey when the SNPs have a $p > 1 \times 10 - 5$, blue when $p < 1 \times 10 - 5$ but $> 510 - 8$ and orange when $p < 5 \times 10 - 8$. The SNPs that are above the blue line and contribute to the transcriptomic imputation models of significant transcripts are labelled.

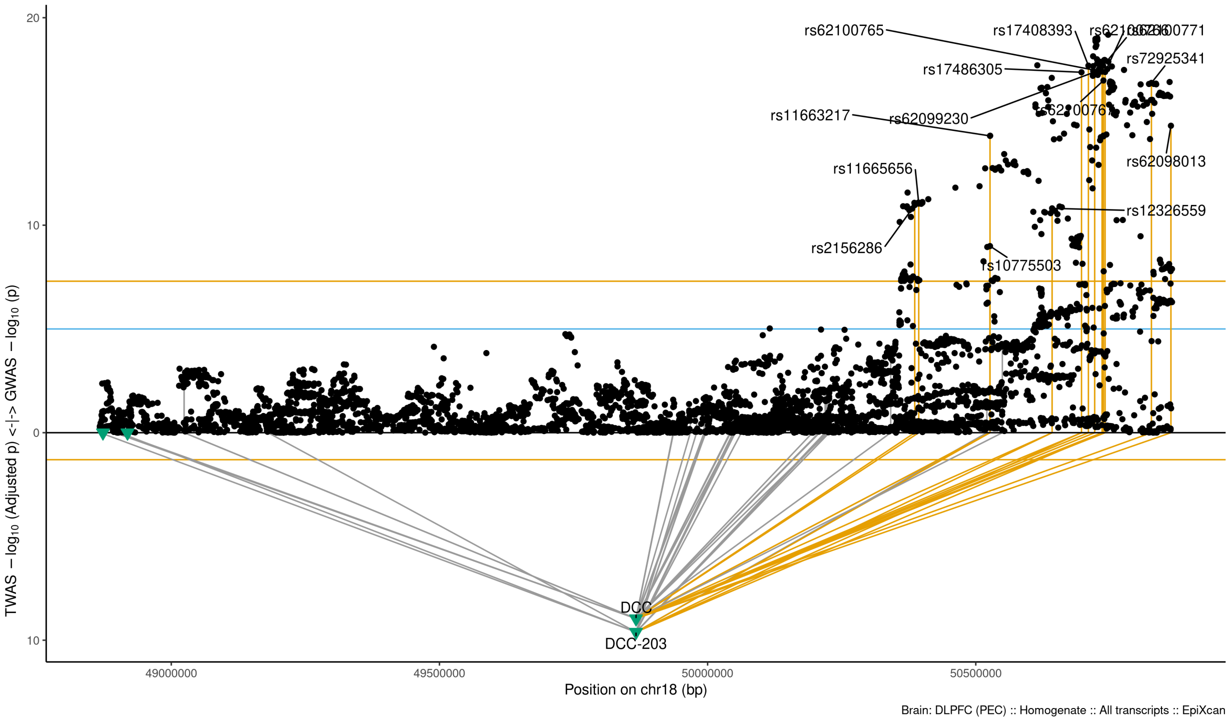

Figure S9-16: Regional Miami plots for DCC203 gene/transcript : Regional Miami plots for the primary MD GWASs corresponding to the genomic region of the DCC203 transcript (1Mbp window from start site). Please also refer to Supplemental Table SXX for details. Top panel shows the GWAS results (black dots); blue line corresponds to $p = 1 \times 10 - 5$, orange line to $p = 5 \times 10 - 8$ (genome-wide significance). Bottom panel shows the TWAS results (green triangles facing upwards or downwards for a positive or negative z-score respectively (up- or down-regultion); only the transcripts with Bonferroni-adjusted $p < 0.1$ are labelled for clarity) for different transcripts (genes are represented by both gene expression and isoform expression); orange line corresponds to Bonferroni-adjusted p = 0.05. Each transcript that is Bonferroni-significant in the region is connected with lines to the SNPs that contribute to its transcriptomic imputation model; lines are grey when the SNPs have a $p > 1 \times 10 - 5$, blue when $p < 1 \times 10 - 5$ but $> 510 - 8$ and orange when $p < 5 \times 10 - 8$. The SNPs that are above the blue line and contribute to the transcriptomic imputation models of significant transcripts are labelled.

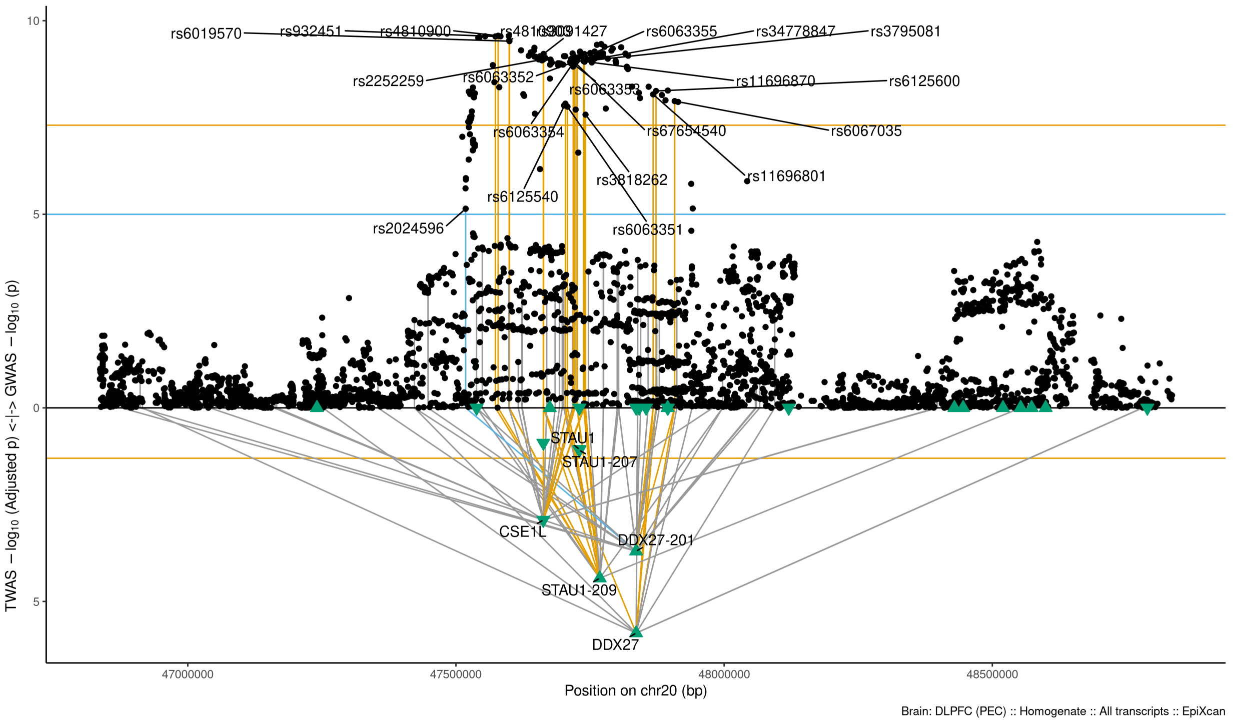

Figure S9-17: Regional Miami plots for DDX27 gene/transcript : Regional Miami plots for the primary MD GWASs corresponding to the genomic region of the DDX27 transcript (1Mbp window from start site). Please also refer to Supplemental Table SXX for details. Top panel shows the GWAS results (black dots); blue line corresponds to $p = 1 \times 10 - 5$, orange line to $p = 5 \times 10 - 8$ (genome-wide significance). Bottom panel shows the TWAS results (green triangles facing upwards or downwards for a positive or negative z-score respectively (up- or down-regultion); only the transcripts with Bonferroni-adjusted $p < 0.1$ are labelled for clarity) for different transcripts (genes are represented by both gene expression and isoform expression); orange line corresponds to Bonferroni-adjusted p = 0.05. Each transcript that is Bonferroni-significant in the region is connected with lines to the SNPs that contribute to its transcriptomic imputation model; lines are grey when the SNPs have a $p > 1 \times 10 - 5$, blue when $p < 1 \times 10 - 5$ but $> 510 - 8$ and orange when $p < 5 \times 10 - 8$. The SNPs that are above the blue line and contribute to the transcriptomic imputation models of significant transcripts are labelled.

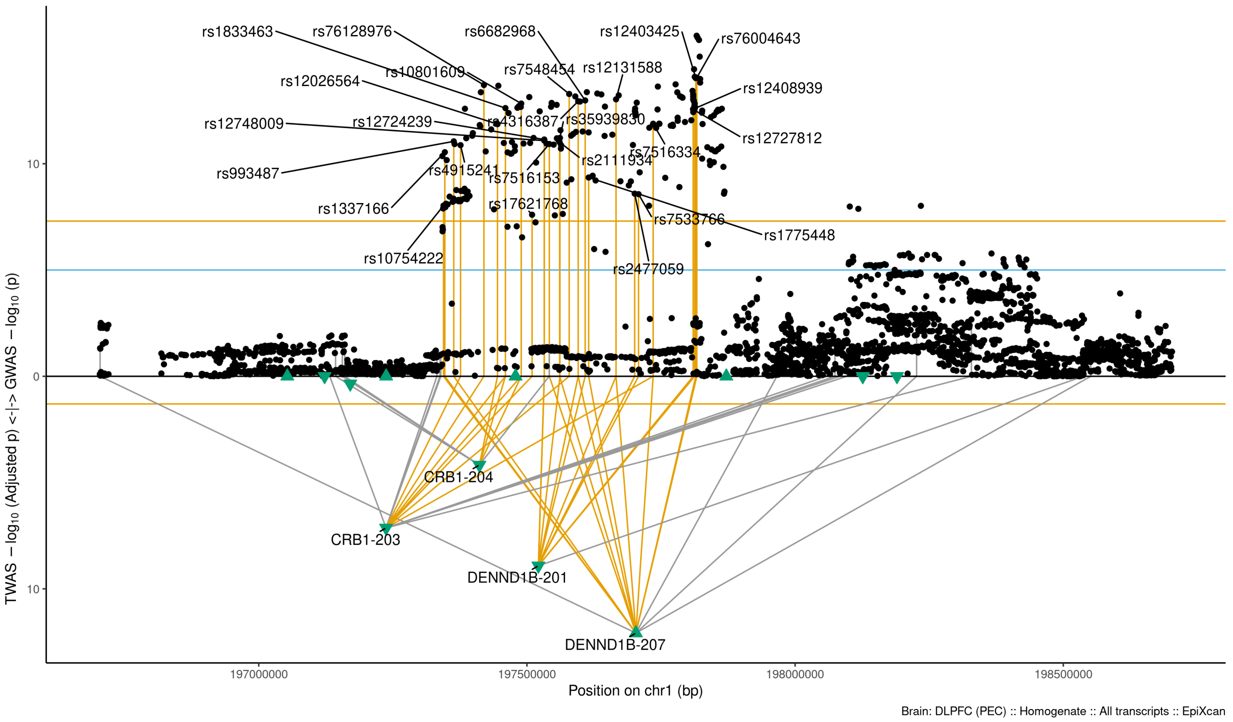

Figure S9-18: Regional Miami plots for DENND1B207 gene/transcript : Regional Miami plots for the primary MD GWASs corresponding to the genomic region of the DENND1B207 transcript (1Mbp window from start site). Please also refer to Supplemental Table SXX for details. Top panel shows the GWAS results (black dots); blue line corresponds to $p = 1 \times 10 - 5$, orange line to $p = 5 \times 10 - 8$ (genome-wide significance). Bottom panel shows the TWAS results (green triangles facing upwards or downwards for a positive or negative z-score respectively (up- or down-regultion); only the transcripts with Bonferroni-adjusted $p < 0.1$ are labelled for clarity) for different transcripts (genes are represented by both gene expression and isoform expression); orange line corresponds to Bonferroni-adjusted p = 0.05. Each transcript that is Bonferroni-significant in the region is connected with lines to the SNPs that contribute to its transcriptomic imputation model; lines are grey when the SNPs have a $p > 1 \times 10 - 5$, blue when $p < 1 \times 10 - 5$ but $> 510 - 8$ and orange when $p < 5 \times 10 - 8$. The SNPs that are above the blue line and contribute to the transcriptomic imputation models of significant transcripts are labelled.

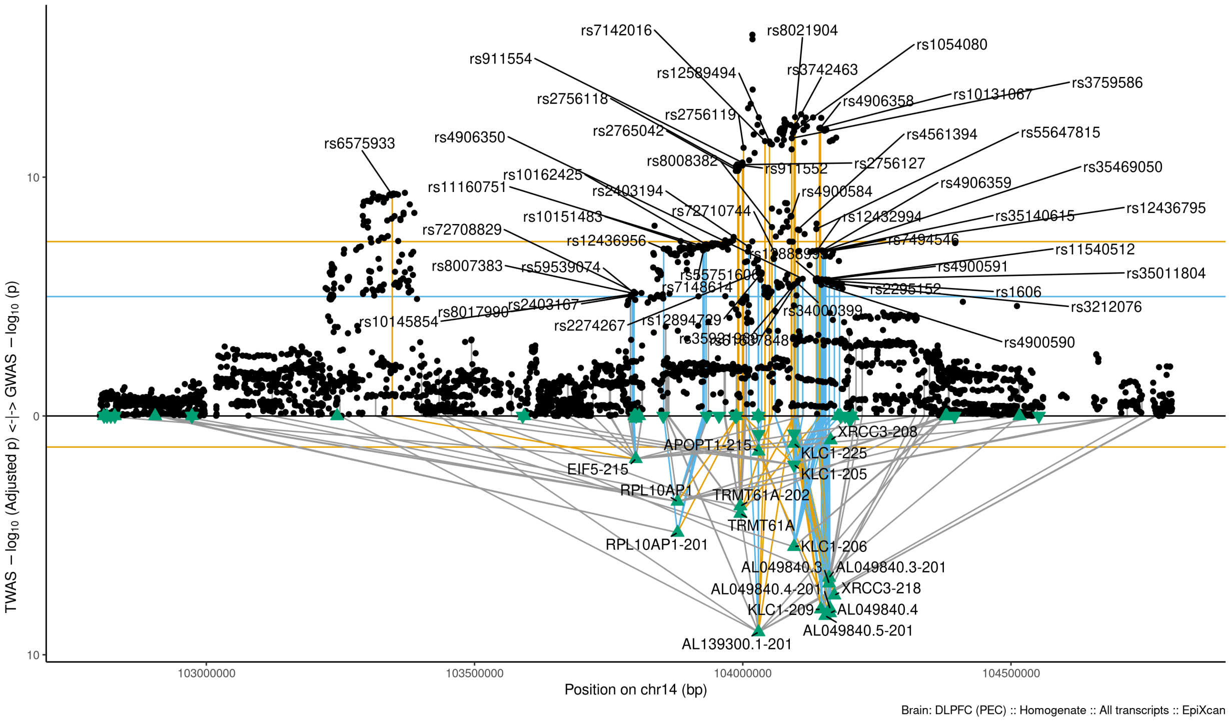

Figure S9-19: Regional Miami plots for EIF5215 gene/transcript : Regional Miami plots for the primary MD GWASs corresponding to the genomic region of the EIF5215 transcript (1Mbp window from start site). Please also refer to Supplemental Table SXX for details. Top panel shows the GWAS results (black dots); blue line corresponds to $p = 1 \times 10 - 5$, orange line to $p = 5 \times 10 - 8$ (genome-wide significance). Bottom panel shows the TWAS results (green triangles facing upwards or downwards for a positive or negative z-score respectively (up- or down-regultion); only the transcripts with Bonferroni-adjusted $p < 0.1$ are labelled for clarity) for different transcripts (genes are represented by both gene expression and isoform expression); orange line corresponds to Bonferroni-adjusted p = 0.05. Each transcript that is Bonferroni-significant in the region is connected with lines to the SNPs that contribute to its transcriptomic imputation model; lines are grey when the SNPs have a $p > 1 \times 10 - 5$, blue when $p < 1 \times 10 - 5$ but $> 510 - 8$ and orange when $p < 5 \times 10 - 8$. The SNPs that are above the blue line and contribute to the transcriptomic imputation models of significant transcripts are labelled.

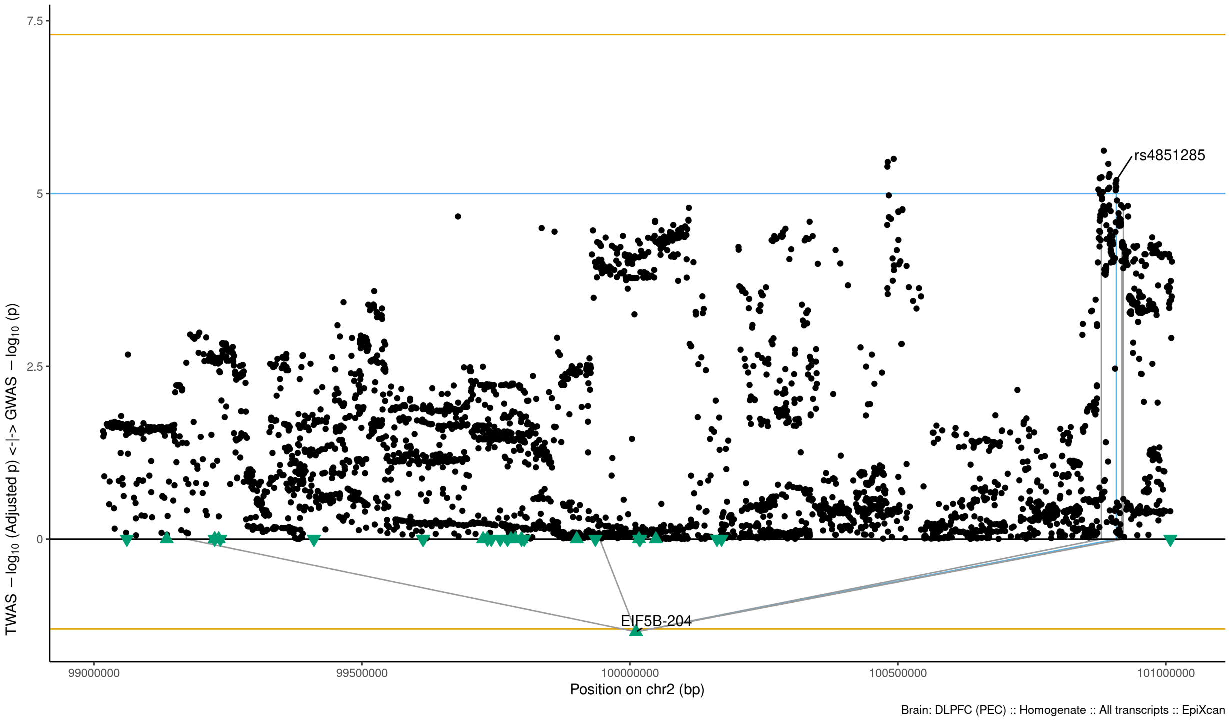

Figure S9-20: Regional Miami plots for EIF5B204 gene/transcript : Regional Miami plots for the primary MD GWASs corresponding to the genomic region of the EIF5B204 transcript (1Mbp window from start site). Please also refer to Supplemental Table SXX for details. Top panel shows the GWAS results (black dots); blue line corresponds to $p = 1 \times 10 - 5$, orange line to $p = 5 \times 10 - 8$ (genome-wide significance). Bottom panel shows the TWAS results (green triangles facing upwards or downwards for a positive or negative z-score respectively (up- or down-regultion); only the transcripts with Bonferroni-adjusted $p < 0.1$ are labelled for clarity) for different transcripts (genes are represented by both gene expression and isoform expression); orange line corresponds to Bonferroni-adjusted p = 0.05. Each transcript that is Bonferroni-significant in the region is connected with lines to the SNPs that contribute to its transcriptomic imputation model; lines are grey when the SNPs have a $p > 1 \times 10 - 5$, blue when $p < 1 \times 10 - 5$ but $> 510 - 8$ and orange when $p < 5 \times 10 - 8$. The SNPs that are above the blue line and contribute to the transcriptomic imputation models of significant transcripts are labelled.

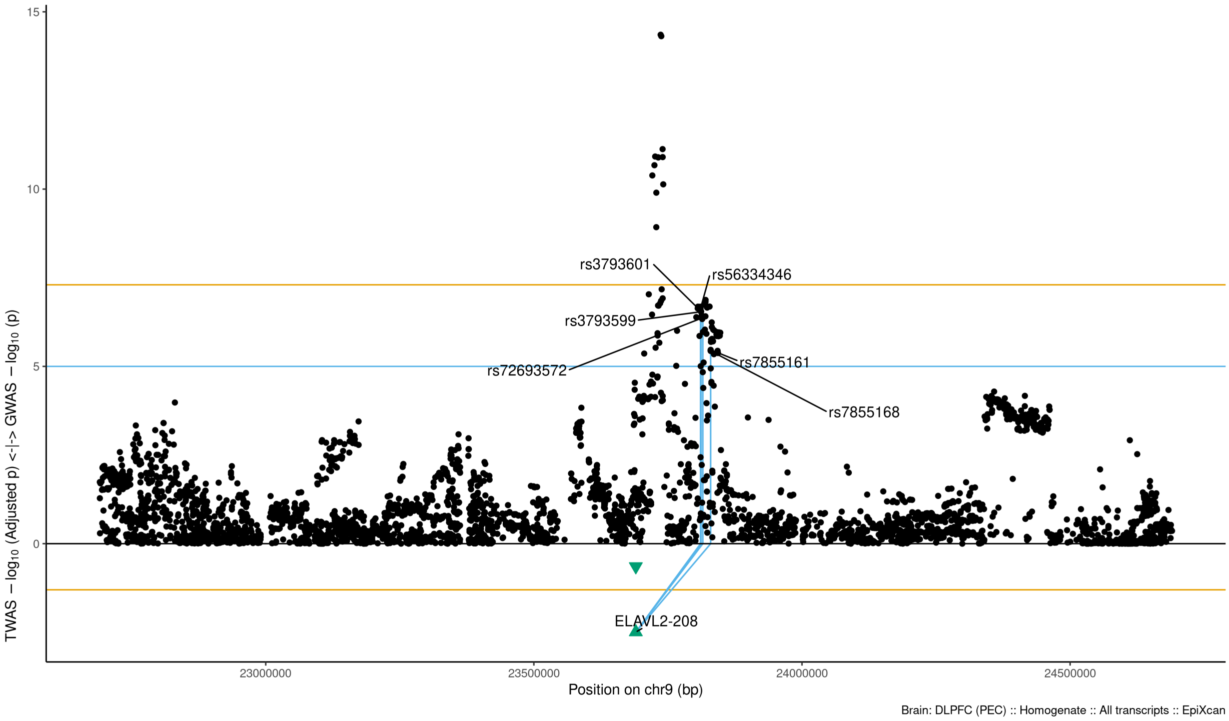

Figure S9-21: Regional Miami plots for ELAVL2208 gene/transcript : Regional Miami plots for the primary MD GWASs corresponding to the genomic region of the ELAVL2208 transcript (1Mbp window from start site). Please also refer to Supplemental Table SXX for details. Top panel shows the GWAS results (black dots); blue line corresponds to $p = 1 \times 10 - 5$, orange line to $p = 5 \times 10 - 8$ (genome-wide significance). Bottom panel shows the TWAS results (green triangles facing upwards or downwards for a positive or negative z-score respectively (up- or down-regultion); only the transcripts with Bonferroni-adjusted $p < 0.1$ are labelled for clarity) for different transcripts (genes are represented by both gene expression and isoform expression); orange line corresponds to Bonferroni-adjusted p = 0.05. Each transcript that is Bonferroni-significant in the region is connected with lines to the SNPs that contribute to its transcriptomic imputation model; lines are grey when the SNPs have a $p > 1 \times 10 - 5$, blue when $p < 1 \times 10 - 5$ but $> 510 - 8$ and orange when $p < 5 \times 10 - 8$. The SNPs that are above the blue line and contribute to the transcriptomic imputation models of significant transcripts are labelled.

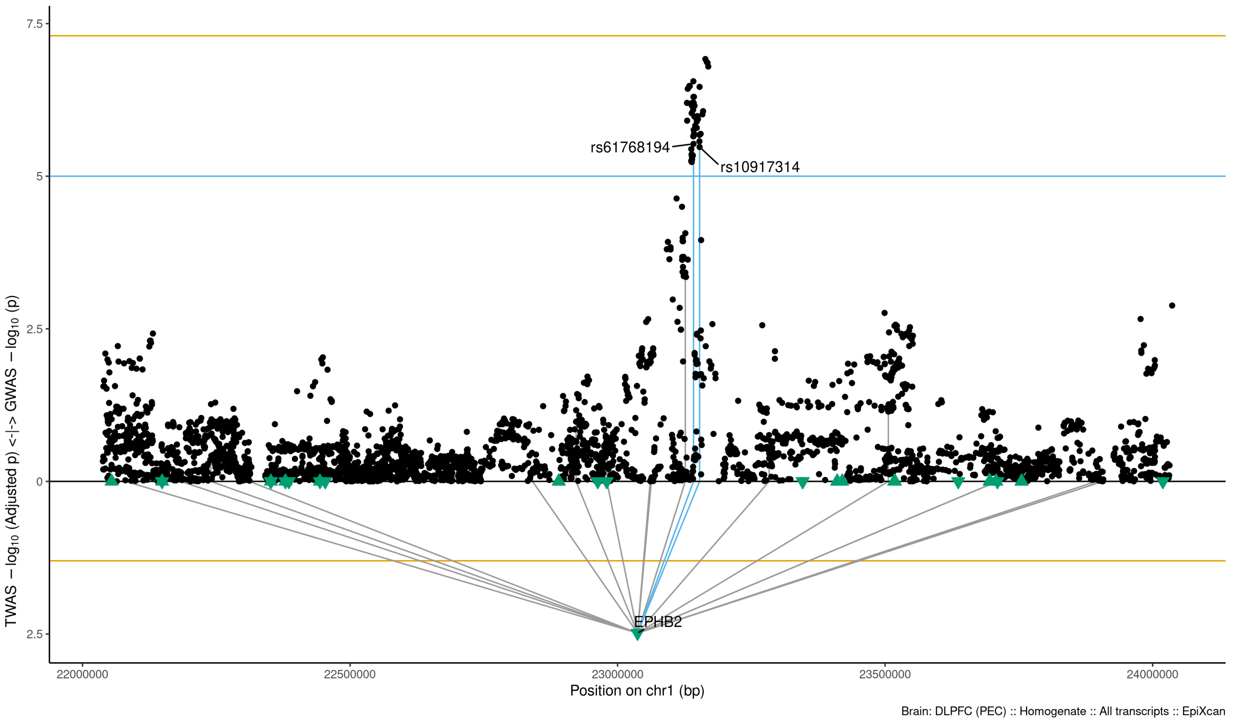

Figure S9-22: Regional Miami plots for EPHB2 gene/transcript : Regional Miami plots for the primary MD GWASs corresponding to the genomic region of the EPHB2 transcript (1Mbp window from start site). Please also refer to Supplemental Table SXX for details. Top panel shows the GWAS results (black dots); blue line corresponds to $p = 1 \times 10 - 5$, orange line to $p = 5 \times 10 - 8$ (genome-wide significance). Bottom panel shows the TWAS results (green triangles facing upwards or downwards for a positive or negative z-score respectively (up- or down-regultion); only the transcripts with Bonferroni-adjusted $p < 0.1$ are labelled for clarity) for different transcripts (genes are represented by both gene expression and isoform expression); orange line corresponds to Bonferroni-adjusted p = 0.05. Each transcript that is Bonferroni-significant in the region is connected with lines to the SNPs that contribute to its transcriptomic imputation model; lines are grey when the SNPs have a $p > 1 \times 10 - 5$, blue when $p < 1 \times 10 - 5$ but $> 510 - 8$ and orange when $p < 5 \times 10 - 8$. The SNPs that are above the blue line and contribute to the transcriptomic imputation models of significant transcripts are labelled.

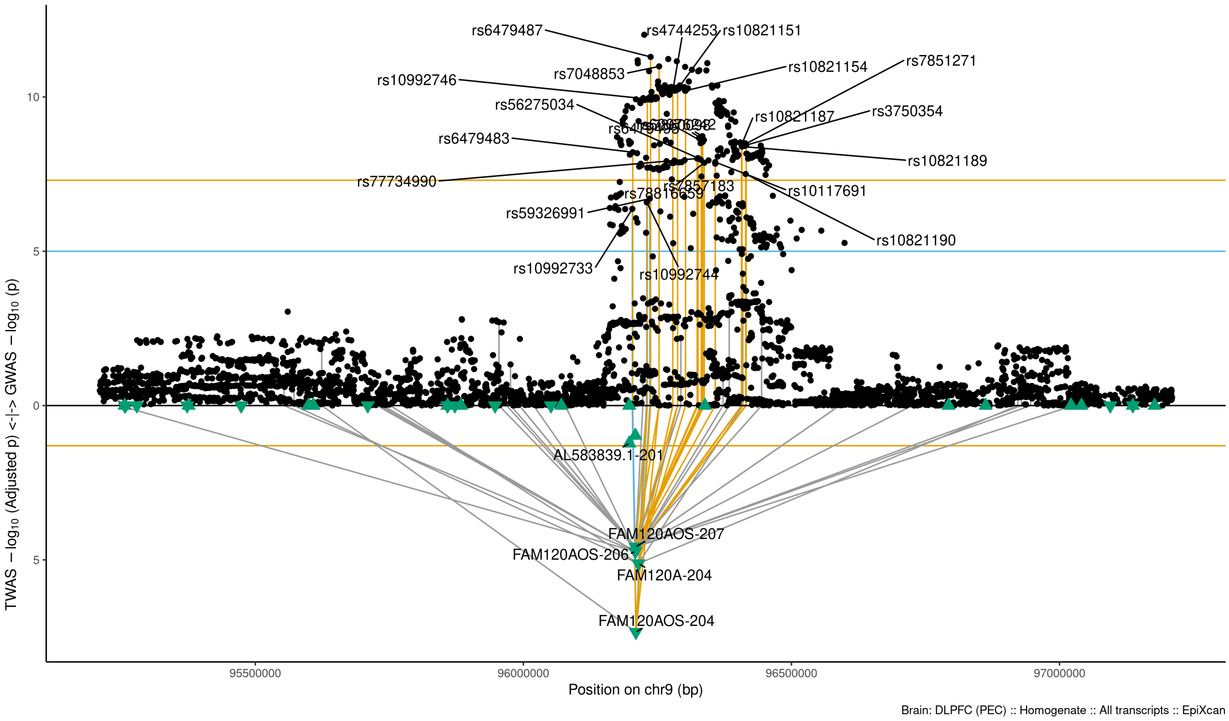

Figure S9-23: Regional Miami plots for FAM120AOS204 gene/transcript : Regional Miami plots for the primary MD GWASs corresponding to the genomic region of the FAM120AOS204 transcript (1Mbp window from start site). Please also refer to Supplemental Table SXX for details. Top panel shows the GWAS results (black dots); blue line corresponds to $p = 1 \times 10 - 5$, orange line to $p = 5 \times 10 - 8$ (genome-wide significance). Bottom panel shows the TWAS results (green triangles facing upwards or downwards for a positive or negative z-score respectively (up- or down-regultion); only the transcripts with Bonferroni-adjusted $p < 0.1$ are labelled for clarity) for different transcripts (genes are represented by both gene expression and isoform expression); orange line corresponds to Bonferroni-adjusted p = 0.05. Each transcript that is Bonferroni-significant in the region is connected with lines to the SNPs that contribute to its transcriptomic imputation model; lines are grey when the SNPs have a $p > 1 \times 10 - 5$, blue when $p < 1 \times 10 - 5$ but $> 510 - 8$ and orange when $p < 5 \times 10 - 8$. The SNPs that are above the blue line and contribute to the transcriptomic imputation models of significant transcripts are labelled.

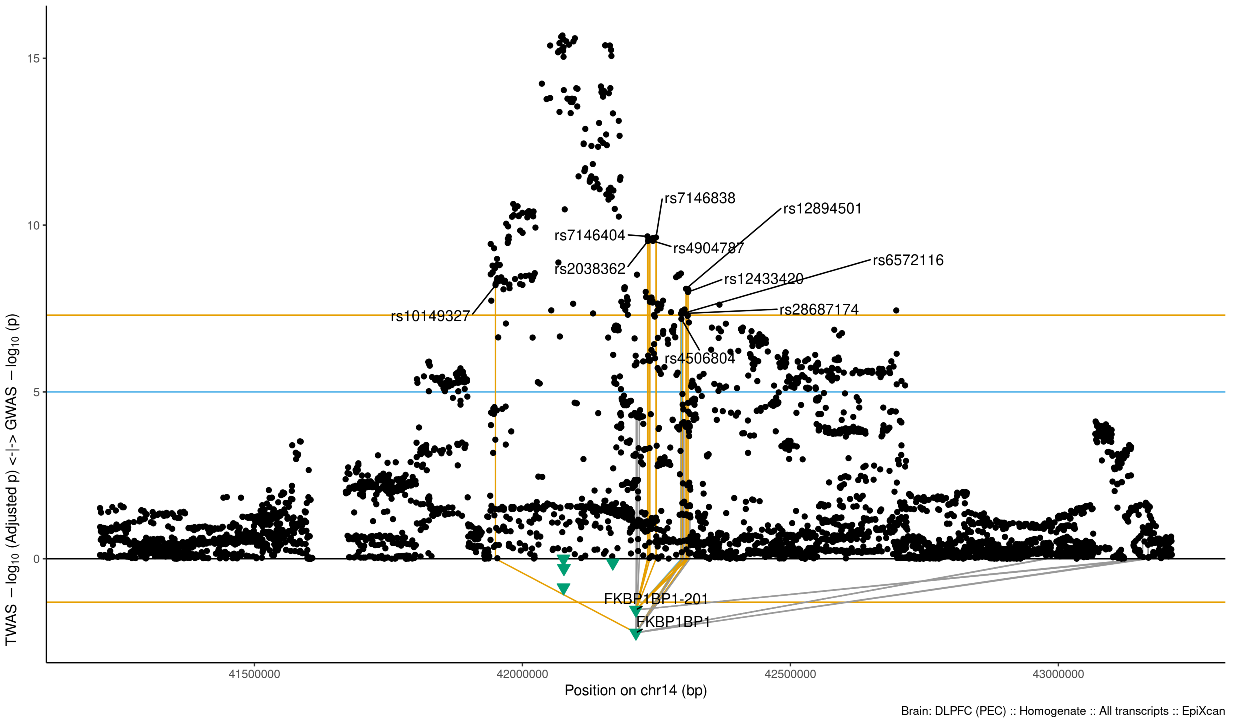

Figure S9-24: Regional Miami plots for FKBP1BP1 gene/transcript : Regional Miami plots for the primary MD GWASs corresponding to the genomic region of the FKBP1BP1 transcript (1Mbp window from start site). Please also refer to Supplemental Table SXX for details. Top panel shows the GWAS results (black dots); blue line corresponds to $p = 1 \times 10 - 5$, orange line to $p = 5 \times 10 - 8$ (genome-wide significance). Bottom panel shows the TWAS results (green triangles facing upwards or downwards for a positive or negative z-score respectively (up- or down-regultion); only the transcripts with Bonferroni-adjusted $p < 0.1$ are labelled for clarity) for different transcripts (genes are represented by both gene expression and isoform expression); orange line corresponds to Bonferroni-adjusted p = 0.05. Each transcript that is Bonferroni-significant in the region is connected with lines to the SNPs that contribute to its transcriptomic imputation model; lines are grey when the SNPs have a $p > 1 \times 10 - 5$, blue when $p < 1 \times 10 - 5$ but $> 510 - 8$ and orange when $p < 5 \times 10 - 8$. The SNPs that are above the blue line and contribute to the transcriptomic imputation models of significant transcripts are labelled.

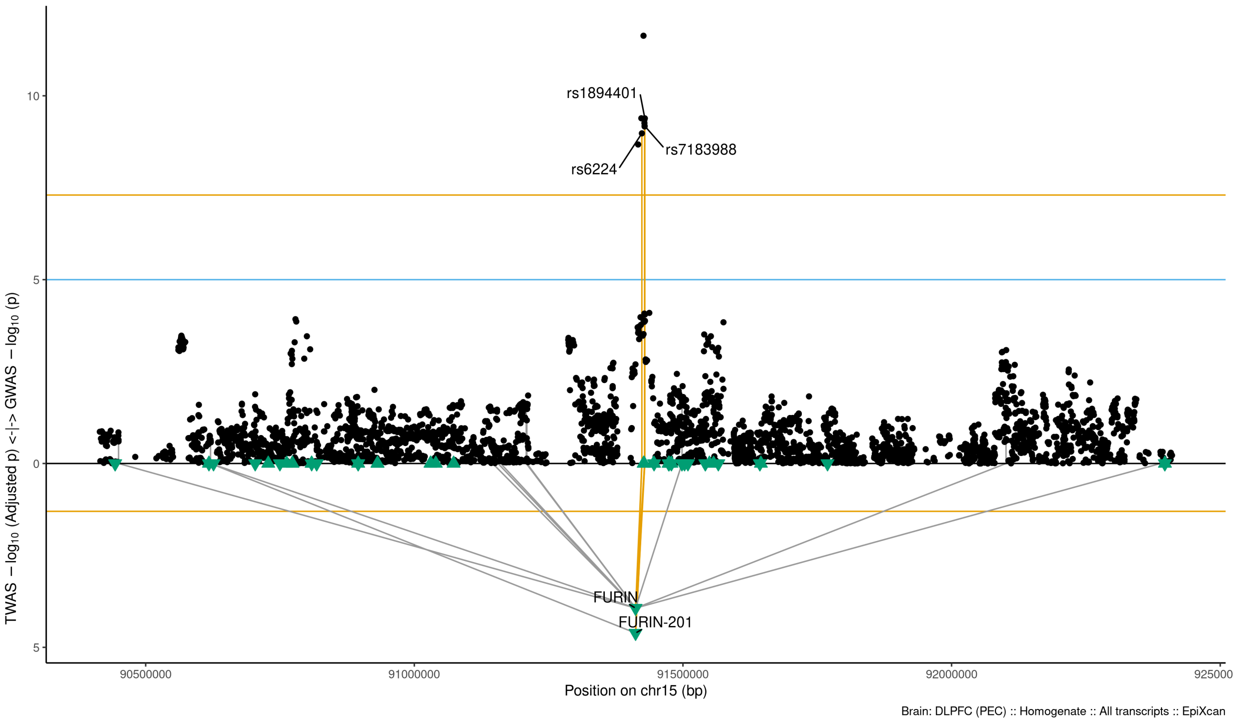

Figure S9-25: Regional Miami plots for FURIN201 gene/transcript : Regional Miami plots for the primary MD GWASs corresponding to the genomic region of the FURIN201 transcript (1Mbp window from start site). Please also refer to Supplemental Table SXX for details. Top panel shows the GWAS results (black dots); blue line corresponds to $p = 1 \times 10 - 5$, orange line to $p = 5 \times 10 - 8$ (genome-wide significance). Bottom panel shows the TWAS results (green triangles facing upwards or downwards for a positive or negative z-score respectively (up- or down-regultion); only the transcripts with Bonferroni-adjusted $p < 0.1$ are labelled for clarity) for different transcripts (genes are represented by both gene expression and isoform expression); orange line corresponds to Bonferroni-adjusted p = 0.05. Each transcript that is Bonferroni-significant in the region is connected with lines to the SNPs that contribute to its transcriptomic imputation model; lines are grey when the SNPs have a $p > 1 \times 10 - 5$, blue when $p < 1 \times 10 - 5$ but $> 510 - 8$ and orange when $p < 5 \times 10 - 8$. The SNPs that are above the blue line and contribute to the transcriptomic imputation models of significant transcripts are labelled.

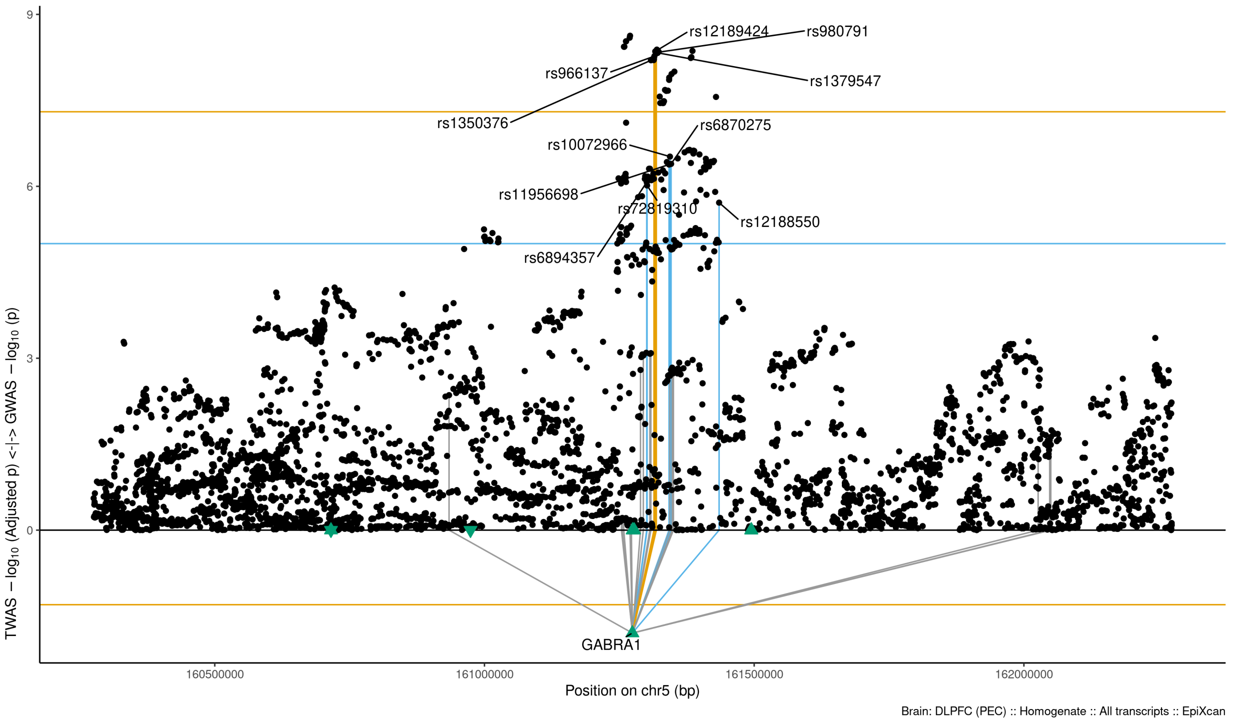

Figure S9-26: Regional Miami plots for GABRA1 gene/transcript : Regional Miami plots for the primary MD GWASs corresponding to the genomic region of the GABRA1 transcript (1Mbp window from start site). Please also refer to Supplemental Table SXX for details. Top panel shows the GWAS results (black dots); blue line corresponds to $p = 1 \times 10 - 5$, orange line to $p = 5 \times 10 - 8$ (genome-wide significance). Bottom panel shows the TWAS results (green triangles facing upwards or downwards for a positive or negative z-score respectively (up- or down-regultion); only the transcripts with Bonferroni-adjusted $p < 0.1$ are labelled for clarity) for different transcripts (genes are represented by both gene expression and isoform expression); orange line corresponds to Bonferroni-adjusted p = 0.05. Each transcript that is Bonferroni-significant in the region is connected with lines to the SNPs that contribute to its transcriptomic imputation model; lines are grey when the SNPs have a $p > 1 \times 10 - 5$, blue when $p < 1 \times 10 - 5$ but $> 510 - 8$ and orange when $p < 5 \times 10 - 8$. The SNPs that are above the blue line and contribute to the transcriptomic imputation models of significant transcripts are labelled.

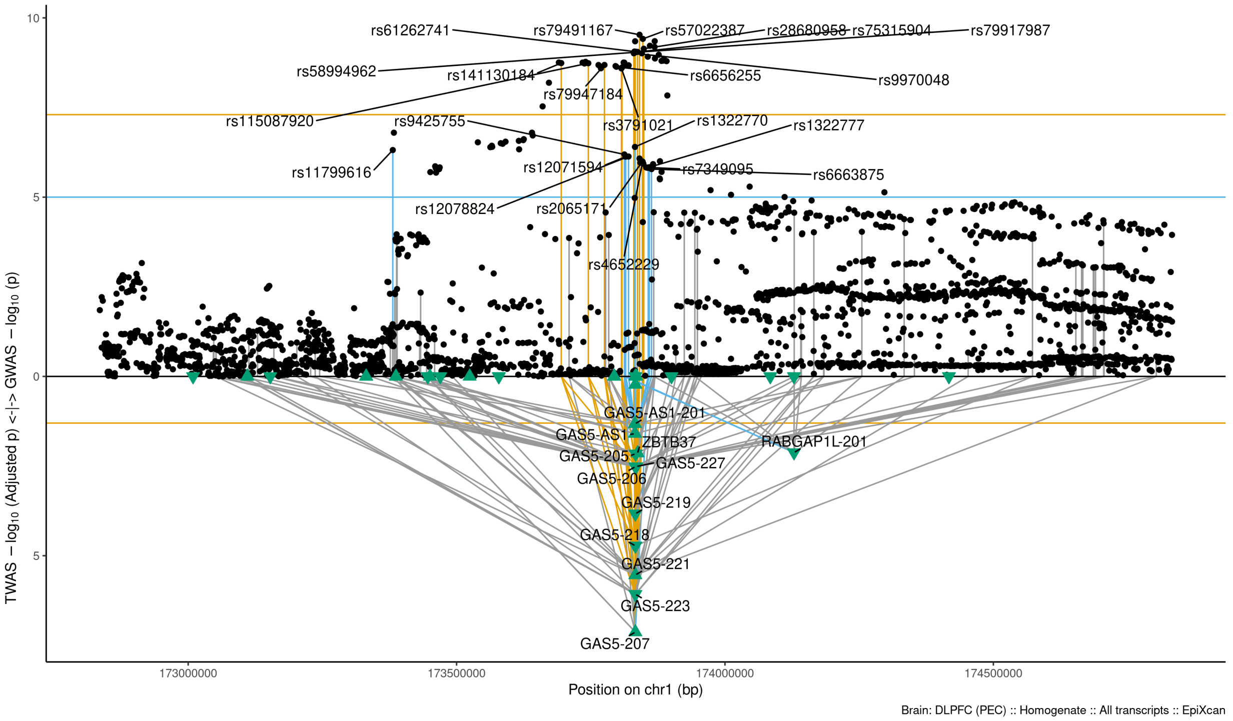

Figure S9-27: Regional Miami plots for GAS5207 gene/transcript : Regional Miami plots for the primary MD GWASs corresponding to the genomic region of the GAS5207 transcript (1Mbp window from start site). Please also refer to Supplemental Table SXX for details. Top panel shows the GWAS results (black dots); blue line corresponds to $p = 1 \times 10 - 5$, orange line to $p = 5 \times 10 - 8$ (genome-wide significance). Bottom panel shows the TWAS results (green triangles facing upwards or downwards for a positive or negative z-score respectively (up- or down-regultion); only the transcripts with Bonferroni-adjusted $p < 0.1$ are labelled for clarity) for different transcripts (genes are represented by both gene expression and isoform expression); orange line corresponds to Bonferroni-adjusted p = 0.05. Each transcript that is Bonferroni-significant in the region is connected with lines to the SNPs that contribute to its transcriptomic imputation model; lines are grey when the SNPs have a $p > 1 \times 10 - 5$, blue when $p < 1 \times 10 - 5$ but $> 510 - 8$ and orange when $p < 5 \times 10 - 8$. The SNPs that are above the blue line and contribute to the transcriptomic imputation models of significant transcripts are labelled.

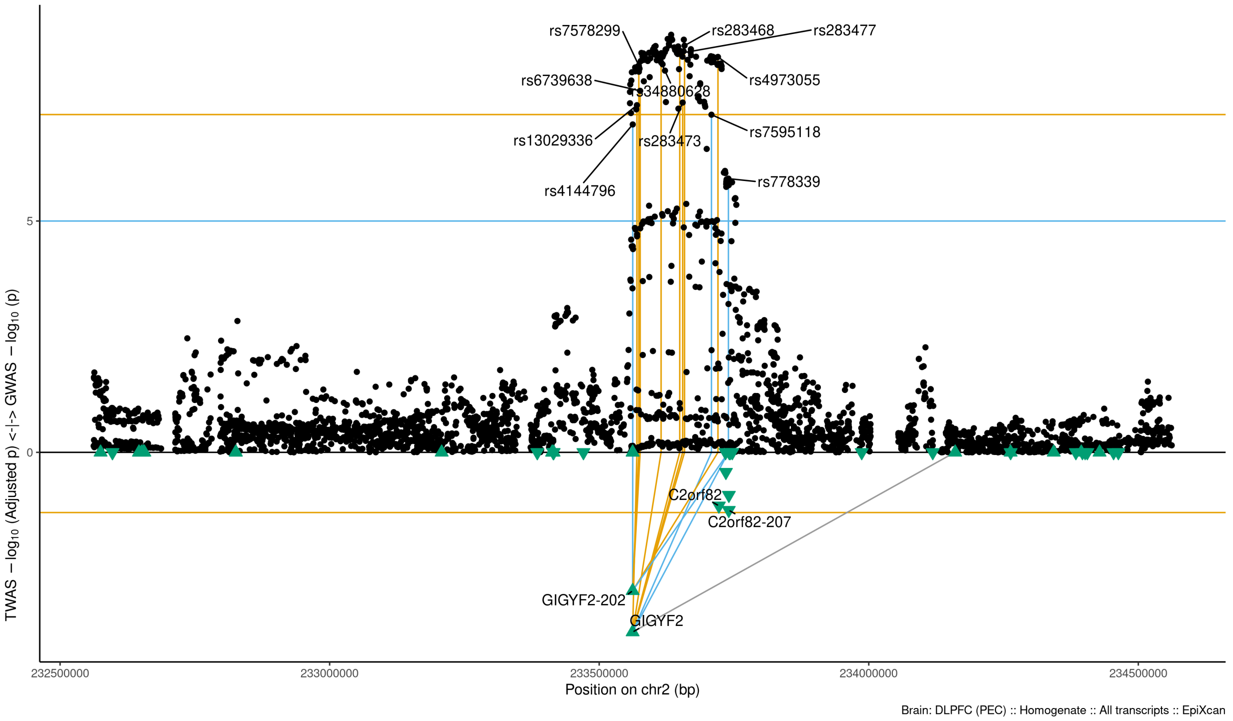

Figure S9-28: Regional Miami plots for GIGYF2 gene/transcript : Regional Miami plots for the primary MD GWASs corresponding to the genomic region of the GIGYF2 transcript (1Mbp window from start site). Please also refer to Supplemental Table SXX for details. Top panel shows the GWAS results (black dots); blue line corresponds to $p = 1 \times 10 - 5$, orange line to $p = 5 \times 10 - 8$ (genome-wide significance). Bottom panel shows the TWAS results (green triangles facing upwards or downwards for a positive or negative z-score respectively (up- or down-regultion); only the transcripts with Bonferroni-adjusted $p < 0.1$ are labelled for clarity) for different transcripts (genes are represented by both gene expression and isoform expression); orange line corresponds to Bonferroni-adjusted p = 0.05. Each transcript that is Bonferroni-significant in the region is connected with lines to the SNPs that contribute to its transcriptomic imputation model; lines are grey when the SNPs have a $p > 1 \times 10 - 5$, blue when $p < 1 \times 10 - 5$ but $> 510 - 8$ and orange when $p < 5 \times 10 - 8$. The SNPs that are above the blue line and contribute to the transcriptomic imputation models of significant transcripts are labelled.

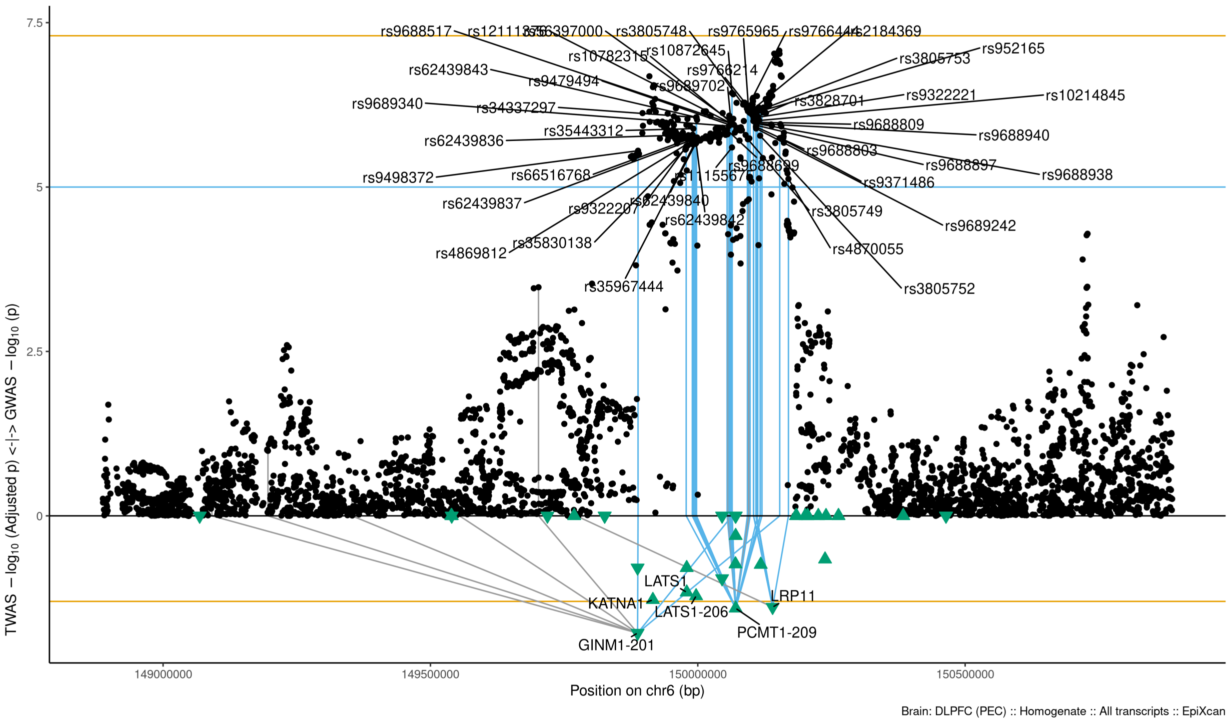

Figure S9-29: Regional Miami plots for GINM1201 gene/transcript : Regional Miami plots for the primary MD GWASs corresponding to the genomic region of the GINM1201 transcript (1Mbp window from start site). Please also refer to Supplemental Table SXX for details. Top panel shows the GWAS results (black dots); blue line corresponds to $p = 1 \times 10 - 5$, orange line to $p = 5 \times 10 - 8$ (genome-wide significance). Bottom panel shows the TWAS results (green triangles facing upwards or downwards for a positive or negative z-score respectively (up- or down-regultion); only the transcripts with Bonferroni-adjusted $p < 0.1$ are labelled for clarity) for different transcripts (genes are represented by both gene expression and isoform expression); orange line corresponds to Bonferroni-adjusted p = 0.05. Each transcript that is Bonferroni-significant in the region is connected with lines to the SNPs that contribute to its transcriptomic imputation model; lines are grey when the SNPs have a $p > 1 \times 10 - 5$, blue when $p < 1 \times 10 - 5$ but $> 510 - 8$ and orange when $p < 5 \times 10 - 8$. The SNPs that are above the blue line and contribute to the transcriptomic imputation models of significant transcripts are labelled.

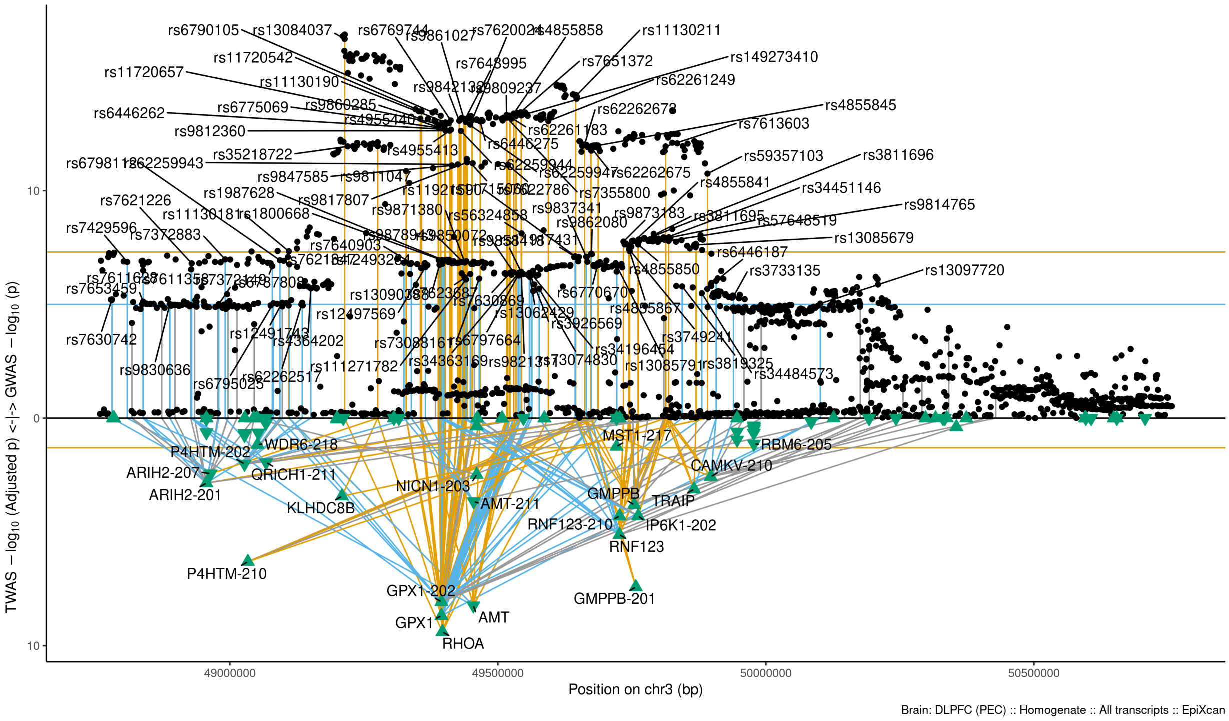

Figure S9-30: Regional Miami plots for GMPPB201 gene/transcript : Regional Miami plots for the primary MD GWASs corresponding to the genomic region of the GMPPB201 transcript (1Mbp window from start site). Please also refer to Supplemental Table SXX for details. Top panel shows the GWAS results (black dots); blue line corresponds to $p = 1 \times 10 - 5$, orange line to $p = 5 \times 10 - 8$ (genome-wide significance). Bottom panel shows the TWAS results (green triangles facing upwards or downwards for a positive or negative z-score respectively (up- or down-regultion); only the transcripts with Bonferroni-adjusted $p < 0.1$ are labelled for clarity) for different transcripts (genes are represented by both gene expression and isoform expression); orange line corresponds to Bonferroni-adjusted p = 0.05. Each transcript that is Bonferroni-significant in the region is connected with lines to the SNPs that contribute to its transcriptomic imputation model; lines are grey when the SNPs have a $p > 1 \times 10 - 5$, blue when $p < 1 \times 10 - 5$ but $> 510 - 8$ and orange when $p < 5 \times 10 - 8$. The SNPs that are above the blue line and contribute to the transcriptomic imputation models of significant transcripts are labelled.

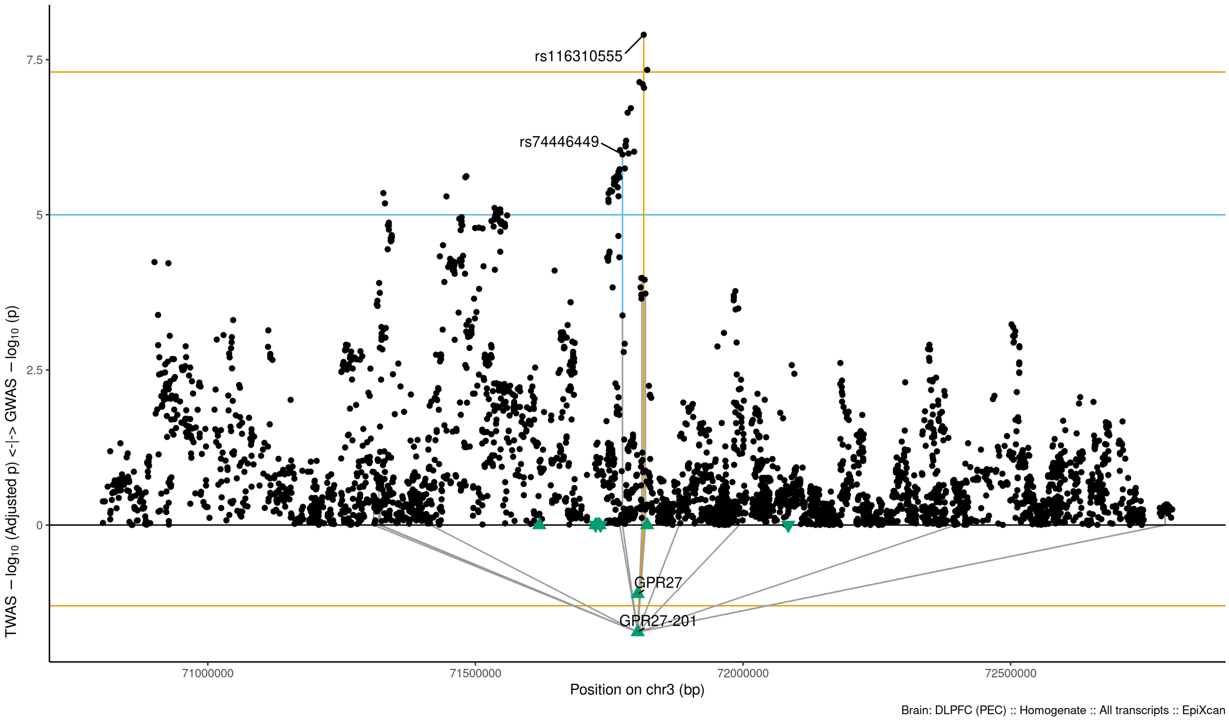

Figure S9-31: Regional Miami plots for GPR27201 gene/transcript : Regional Miami plots for the primary MD GWASs corresponding to the genomic region of the GPR27201 transcript (1Mbp window from start site). Please also refer to Supplemental Table SXX for details. Top panel shows the GWAS results (black dots); blue line corresponds to $p = 1 \times 10 - 5$, orange line to $p = 5 \times 10 - 8$ (genome-wide significance). Bottom panel shows the TWAS results (green triangles facing upwards or downwards for a positive or negative z-score respectively (up- or down-regultion); only the transcripts with Bonferroni-adjusted $p < 0.1$ are labelled for clarity) for different transcripts (genes are represented by both gene expression and isoform expression); orange line corresponds to Bonferroni-adjusted p = 0.05. Each transcript that is Bonferroni-significant in the region is connected with lines to the SNPs that contribute to its transcriptomic imputation model; lines are grey when the SNPs have a $p > 1 \times 10 - 5$, blue when $p < 1 \times 10 - 5$ but $> 510 - 8$ and orange when $p < 5 \times 10 - 8$. The SNPs that are above the blue line and contribute to the transcriptomic imputation models of significant transcripts are labelled.

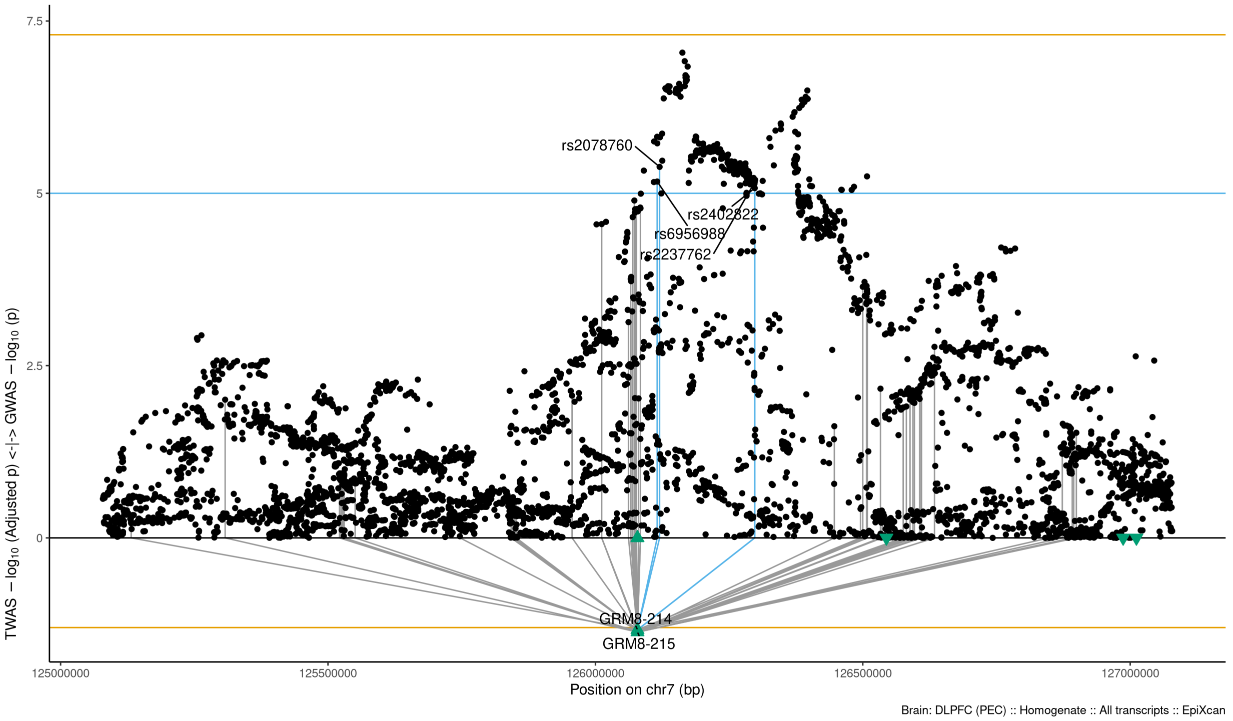

Figure S9-32: Regional Miami plots for GRM8215 gene/transcript : Regional Miami plots for the primary MD GWASs corresponding to the genomic region of the GRM8215 transcript (1Mbp window from start site). Please also refer to Supplemental Table SXX for details. Top panel shows the GWAS results (black dots); blue line corresponds to $p = 1 \times 10 - 5$, orange line to $p = 5 \times 10 - 8$ (genome-wide significance). Bottom panel shows the TWAS results (green triangles facing upwards or downwards for a positive or negative z-score respectively (up- or down-regultion); only the transcripts with Bonferroni-adjusted $p < 0.1$ are labelled for clarity) for different transcripts (genes are represented by both gene expression and isoform expression); orange line corresponds to Bonferroni-adjusted p = 0.05. Each transcript that is Bonferroni-significant in the region is connected with lines to the SNPs that contribute to its transcriptomic imputation model; lines are grey when the SNPs have a $p > 1 \times 10 - 5$, blue when $p < 1 \times 10 - 5$ but $> 510 - 8$ and orange when $p < 5 \times 10 - 8$. The SNPs that are above the blue line and contribute to the transcriptomic imputation models of significant transcripts are labelled.

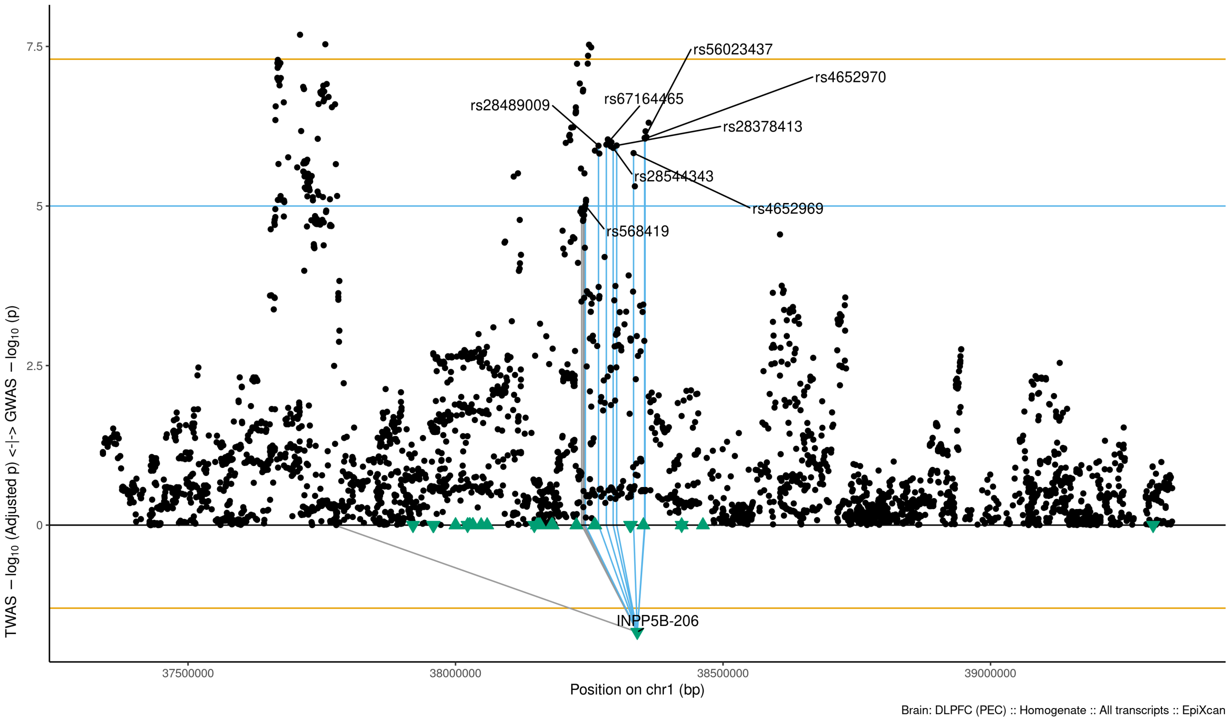

Figure S9-33: Regional Miami plots for INPP5B206 gene/transcript : Regional Miami plots for the primary MD GWASs corresponding to the genomic region of the INPP5B206 transcript (1Mbp window from start site). Please also refer to Supplemental Table SXX for details. Top panel shows the GWAS results (black dots); blue line corresponds to $p = 1 \times 10 - 5$, orange line to $p = 5 \times 10 - 8$ (genome-wide significance). Bottom panel shows the TWAS results (green triangles facing upwards or downwards for a positive or negative z-score respectively (up- or down-regultion); only the transcripts with Bonferroni-adjusted $p < 0.1$ are labelled for clarity) for different transcripts (genes are represented by both gene expression and isoform expression); orange line corresponds to Bonferroni-adjusted p = 0.05. Each transcript that is Bonferroni-significant in the region is connected with lines to the SNPs that contribute to its transcriptomic imputation model; lines are grey when the SNPs have a $p > 1 \times 10 - 5$, blue when $p < 1 \times 10 - 5$ but $> 510 - 8$ and orange when $p < 5 \times 10 - 8$. The SNPs that are above the blue line and contribute to the transcriptomic imputation models of significant transcripts are labelled.

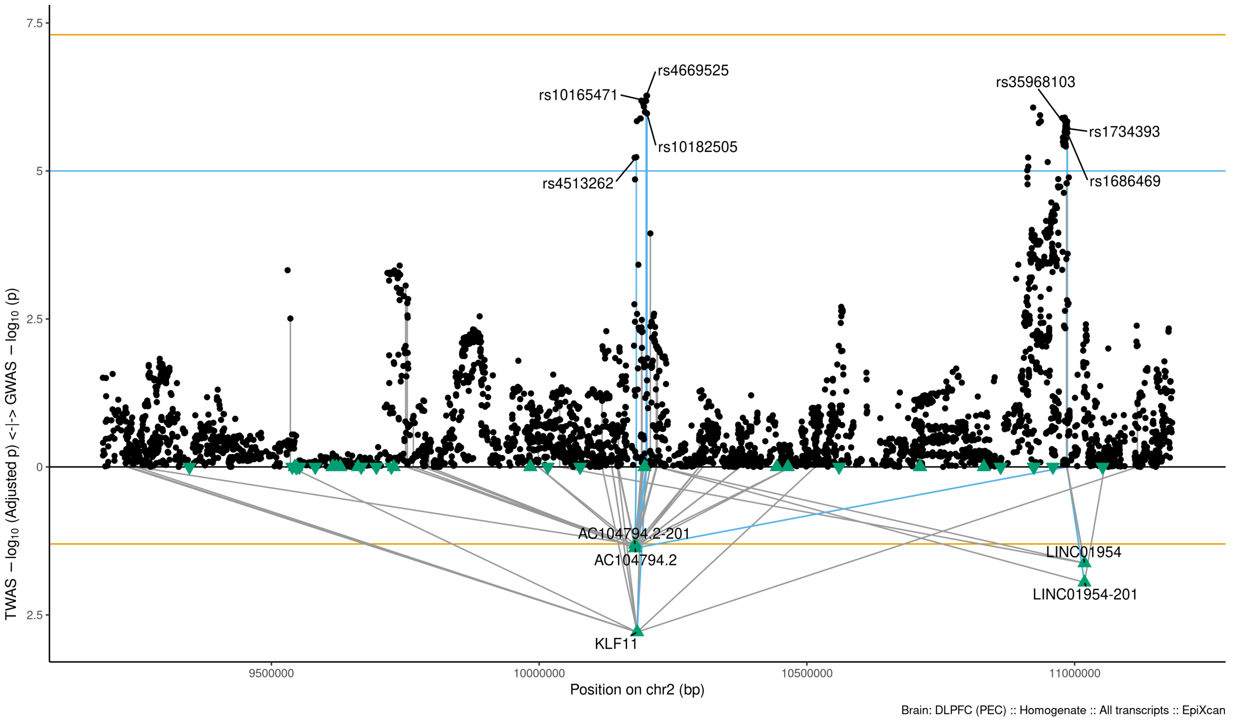

Figure S9-34: Regional Miami plots for KLF11 gene/transcript : Regional Miami plots for the primary MD GWASs corresponding to the genomic region of the KLF11 transcript (1Mbp window from start site). Please also refer to Supplemental Table SXX for details. Top panel shows the GWAS results (black dots); blue line corresponds to $p = 1 \times 10 - 5$, orange line to $p = 5 \times 10 - 8$ (genome-wide significance). Bottom panel shows the TWAS results (green triangles facing upwards or downwards for a positive or negative z-score respectively (up- or down-regultion); only the transcripts with Bonferroni-adjusted $p < 0.1$ are labelled for clarity) for different transcripts (genes are represented by both gene expression and isoform expression); orange line corresponds to Bonferroni-adjusted p = 0.05. Each transcript that is Bonferroni-significant in the region is connected with lines to the SNPs that contribute to its transcriptomic imputation model; lines are grey when the SNPs have a $p > 1 \times 10 - 5$, blue when $p < 1 \times 10 - 5$ but $> 510 - 8$ and orange when $p < 5 \times 10 - 8$. The SNPs that are above the blue line and contribute to the transcriptomic imputation models of significant transcripts are labelled.

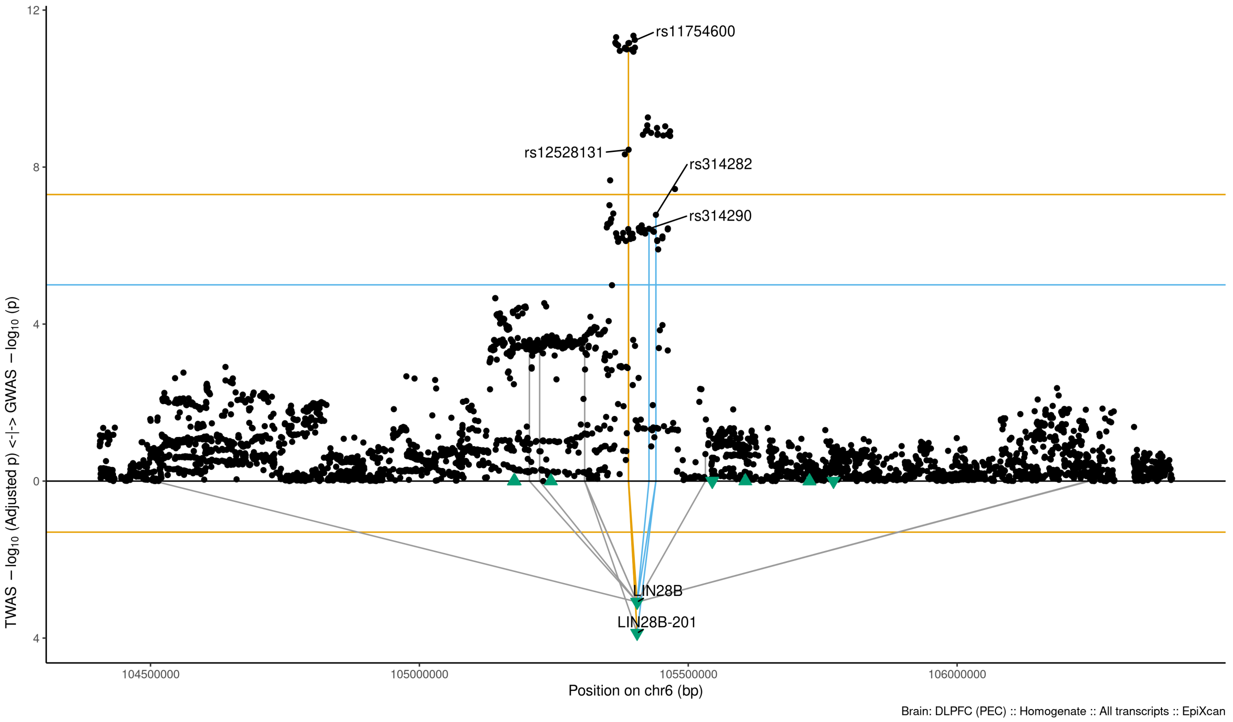

Figure S9-35: Regional Miami plots for LIN28B201 gene/transcript : Regional Miami plots for the primary MD GWASs corresponding to the genomic region of the LIN28B201 transcript (1Mbp window from start site). Please also refer to Supplemental Table SXX for details. Top panel shows the GWAS results (black dots); blue line corresponds to $p = 1 \times 10 - 5$, orange line to $p = 5 \times 10 - 8$ (genome-wide significance). Bottom panel shows the TWAS results (green triangles facing upwards or downwards for a positive or negative z-score respectively (up- or down-regultion); only the transcripts with Bonferroni-adjusted $p < 0.1$ are labelled for clarity) for different transcripts (genes are represented by both gene expression and isoform expression); orange line corresponds to Bonferroni-adjusted p = 0.05. Each transcript that is Bonferroni-significant in the region is connected with lines to the SNPs that contribute to its transcriptomic imputation model; lines are grey when the SNPs have a $p > 1 \times 10 - 5$, blue when $p < 1 \times 10 - 5$ but $> 510 - 8$ and orange when $p < 5 \times 10 - 8$. The SNPs that are above the blue line and contribute to the transcriptomic imputation models of significant transcripts are labelled.

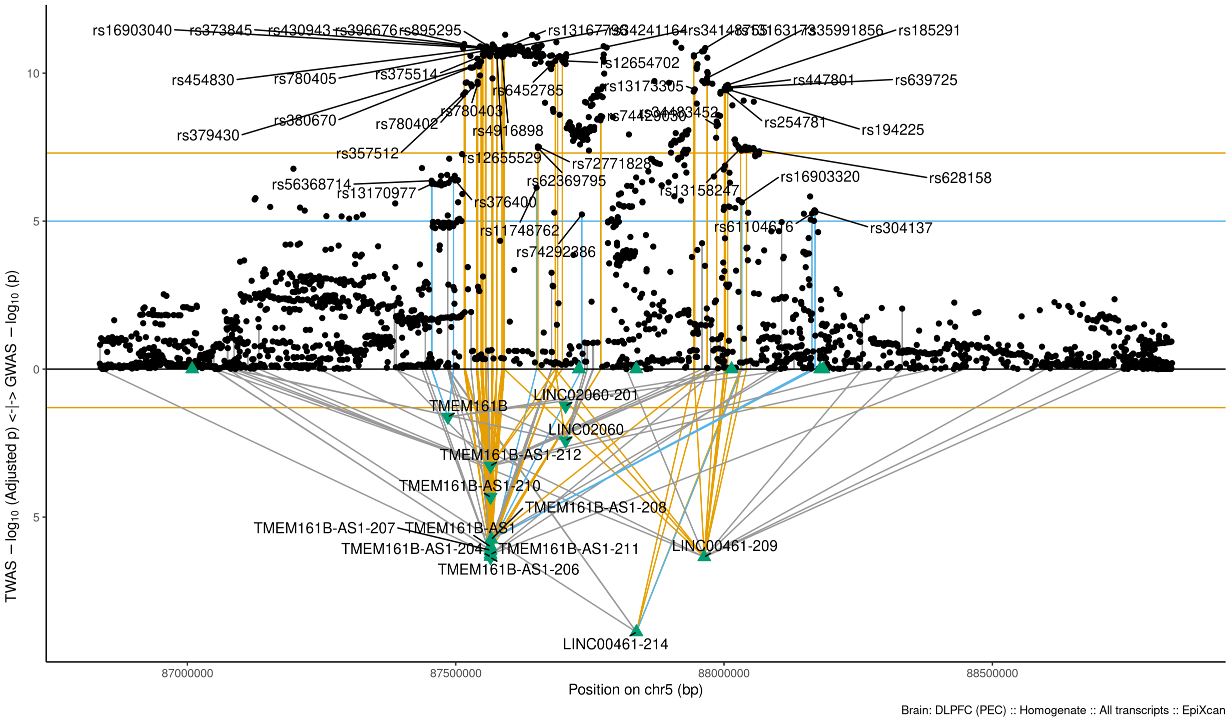

Figure S9-36: Regional Miami plots for LINC00461214 gene/transcript : Regional Miami plots for the primary MD GWASs corresponding to the genomic region of the LINC00461214 transcript (1Mbp window from start site). Please also refer to Supplemental Table SXX for details. Top panel shows the GWAS results (black dots); blue line corresponds to $p = 1 \times 10 - 5$, orange line to $p = 5 \times 10 - 8$ (genome-wide significance). Bottom panel shows the TWAS results (green triangles facing upwards or downwards for a positive or negative z-score respectively (up- or down-regultion); only the transcripts with Bonferroni-adjusted $p < 0.1$ are labelled for clarity) for different transcripts (genes are represented by both gene expression and isoform expression); orange line corresponds to Bonferroni-adjusted p = 0.05. Each transcript that is Bonferroni-significant in the region is connected with lines to the SNPs that contribute to its transcriptomic imputation model; lines are grey when the SNPs have a $p > 1 \times 10 - 5$, blue when $p < 1 \times 10 - 5$ but $> 510 - 8$ and orange when $p < 5 \times 10 - 8$. The SNPs that are above the blue line and contribute to the transcriptomic imputation models of significant transcripts are labelled.

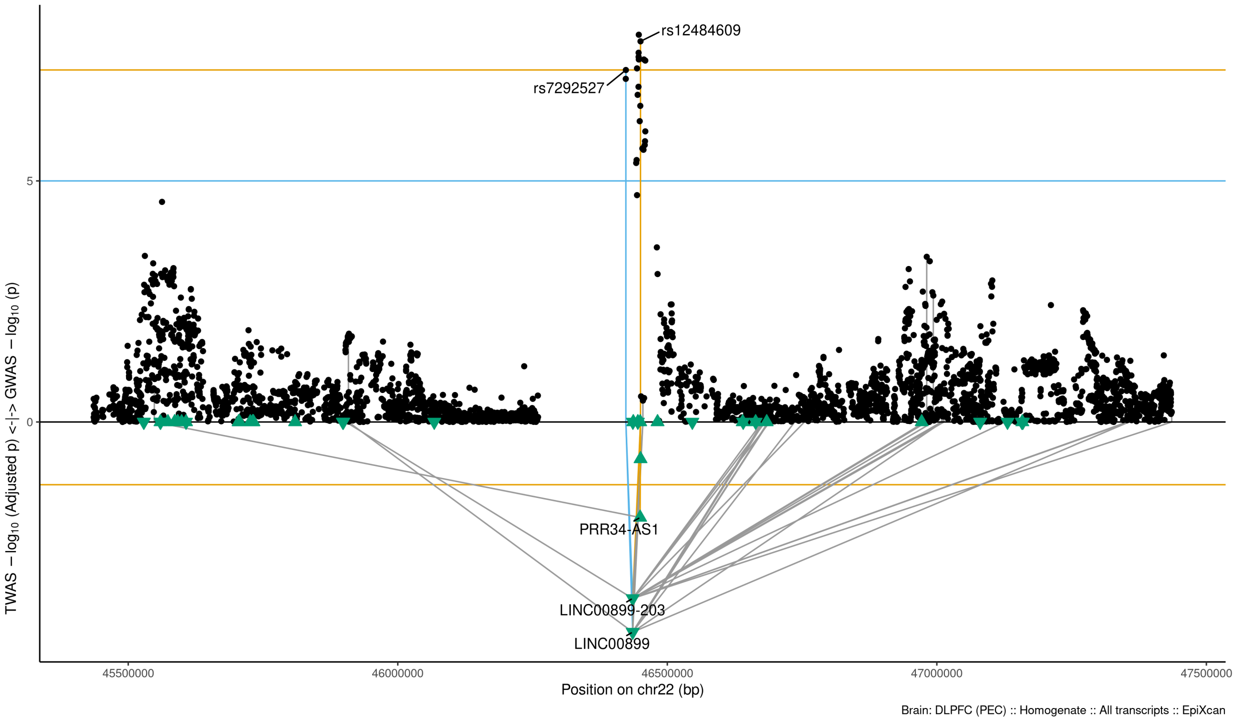

Figure S9-37: Regional Miami plots for LINC00899 gene/transcript : Regional Miami plots for the primary MD GWASs corresponding to the genomic region of the LINC00899 transcript (1Mbp window from start site). Please also refer to Supplemental Table SXX for details. Top panel shows the GWAS results (black dots); blue line corresponds to $p = 1 \times 10 - 5$, orange line to $p = 5 \times 10 - 8$ (genome-wide significance). Bottom panel shows the TWAS results (green triangles facing upwards or downwards for a positive or negative z-score respectively (up- or down-regultion); only the transcripts with Bonferroni-adjusted $p < 0.1$ are labelled for clarity) for different transcripts (genes are represented by both gene expression and isoform expression); orange line corresponds to Bonferroni-adjusted p = 0.05. Each transcript that is Bonferroni-significant in the region is connected with lines to the SNPs that contribute to its transcriptomic imputation model; lines are grey when the SNPs have a $p > 1 \times 10 - 5$, blue when $p < 1 \times 10 - 5$ but $> 510 - 8$ and orange when $p < 5 \times 10 - 8$. The SNPs that are above the blue line and contribute to the transcriptomic imputation models of significant transcripts are labelled.

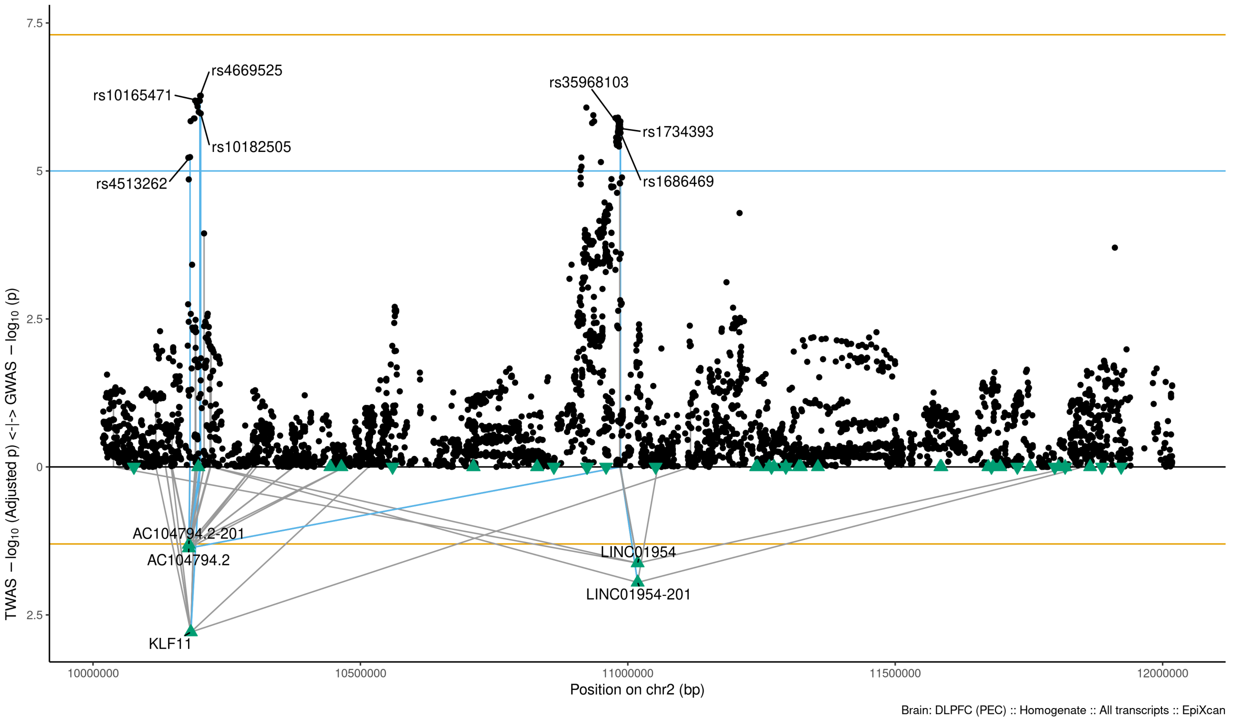

Figure S9-38: Regional Miami plots for LINC01954201 gene/transcript : Regional Miami plots for the primary MD GWASs corresponding to the genomic region of the LINC01954201 transcript (1Mbp window from start site). Please also refer to Supplemental Table SXX for details. Top panel shows the GWAS results (black dots); blue line corresponds to $p = 1 \times 10 - 5$, orange line to $p = 5 \times 10 - 8$ (genome-wide significance). Bottom panel shows the TWAS results (green triangles facing upwards or downwards for a positive or negative z-score respectively (up- or down-regultion); only the transcripts with Bonferroni-adjusted $p < 0.1$ are labelled for clarity) for different transcripts (genes are represented by both gene expression and isoform expression); orange line corresponds to Bonferroni-adjusted p = 0.05. Each transcript that is Bonferroni-significant in the region is connected with lines to the SNPs that contribute to its transcriptomic imputation model; lines are grey when the SNPs have a $p > 1 \times 10 - 5$, blue when $p < 1 \times 10 - 5$ but $> 510 - 8$ and orange when $p < 5 \times 10 - 8$. The SNPs that are above the blue line and contribute to the transcriptomic imputation models of significant transcripts are labelled.

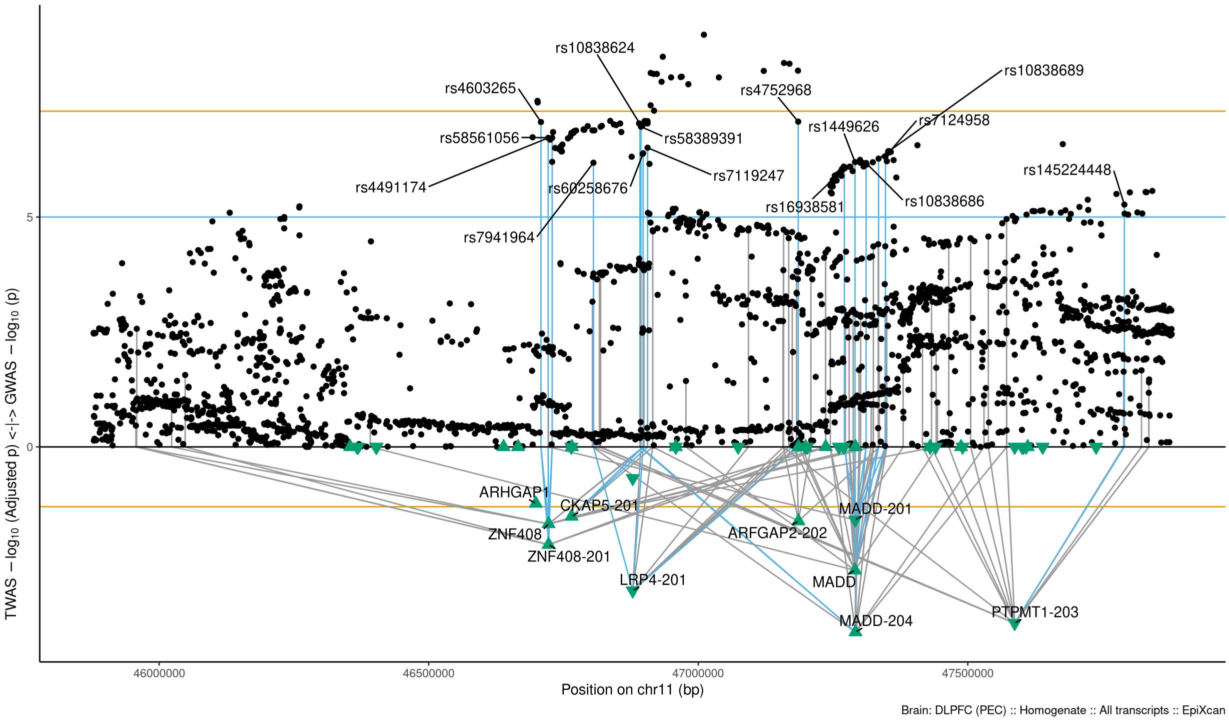

Figure S9-39: Regional Miami plots for LRP4201 gene/transcript : Regional Miami plots for the primary MD GWASs corresponding to the genomic region of the LRP4201 transcript (1Mbp window from start site). Please also refer to Supplemental Table SXX for details. Top panel shows the GWAS results (black dots); blue line corresponds to $p = 1 \times 10 - 5$, orange line to $p = 5 \times 10 - 8$ (genome-wide significance). Bottom panel shows the TWAS results (green triangles facing upwards or downwards for a positive or negative z-score respectively (up- or down-regultion); only the transcripts with Bonferroni-adjusted $p < 0.1$ are labelled for clarity) for different transcripts (genes are represented by both gene expression and isoform expression); orange line corresponds to Bonferroni-adjusted p = 0.05. Each transcript that is Bonferroni-significant in the region is connected with lines to the SNPs that contribute to its transcriptomic imputation model; lines are grey when the SNPs have a $p > 1 \times 10 - 5$, blue when $p < 1 \times 10 - 5$ but $> 510 - 8$ and orange when $p < 5 \times 10 - 8$. The SNPs that are above the blue line and contribute to the transcriptomic imputation models of significant transcripts are labelled.

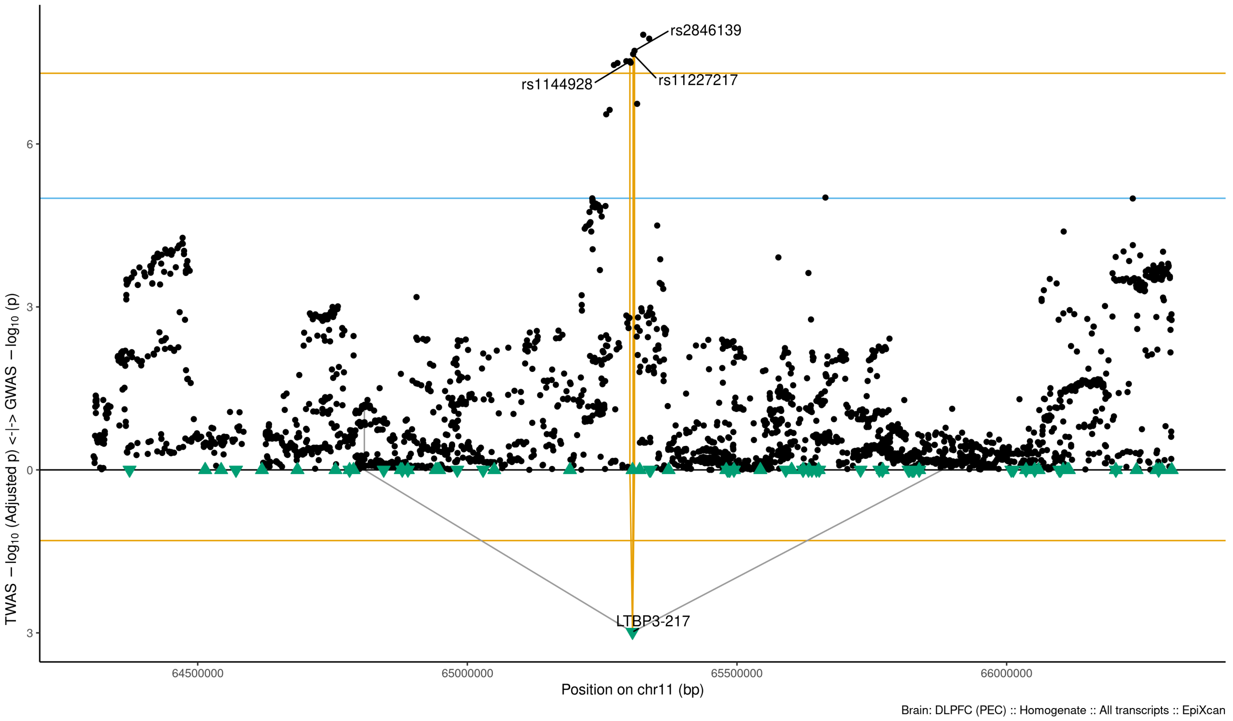

Figure S9-40: Regional Miami plots for LTBP3217 gene/transcript : Regional Miami plots for the primary MD GWASs corresponding to the genomic region of the LTBP3217 transcript (1Mbp window from start site). Please also refer to Supplemental Table SXX for details. Top panel shows the GWAS results (black dots); blue line corresponds to $p = 1 \times 10 - 5$, orange line to $p = 5 \times 10 - 8$ (genome-wide significance). Bottom panel shows the TWAS results (green triangles facing upwards or downwards for a positive or negative z-score respectively (up- or down-regultion); only the transcripts with Bonferroni-adjusted $p < 0.1$ are labelled for clarity) for different transcripts (genes are represented by both gene expression and isoform expression); orange line corresponds to Bonferroni-adjusted p = 0.05. Each transcript that is Bonferroni-significant in the region is connected with lines to the SNPs that contribute to its transcriptomic imputation model; lines are grey when the SNPs have a $p > 1 \times 10 - 5$, blue when $p < 1 \times 10 - 5$ but $> 510 - 8$ and orange when $p < 5 \times 10 - 8$. The SNPs that are above the blue line and contribute to the transcriptomic imputation models of significant transcripts are labelled.

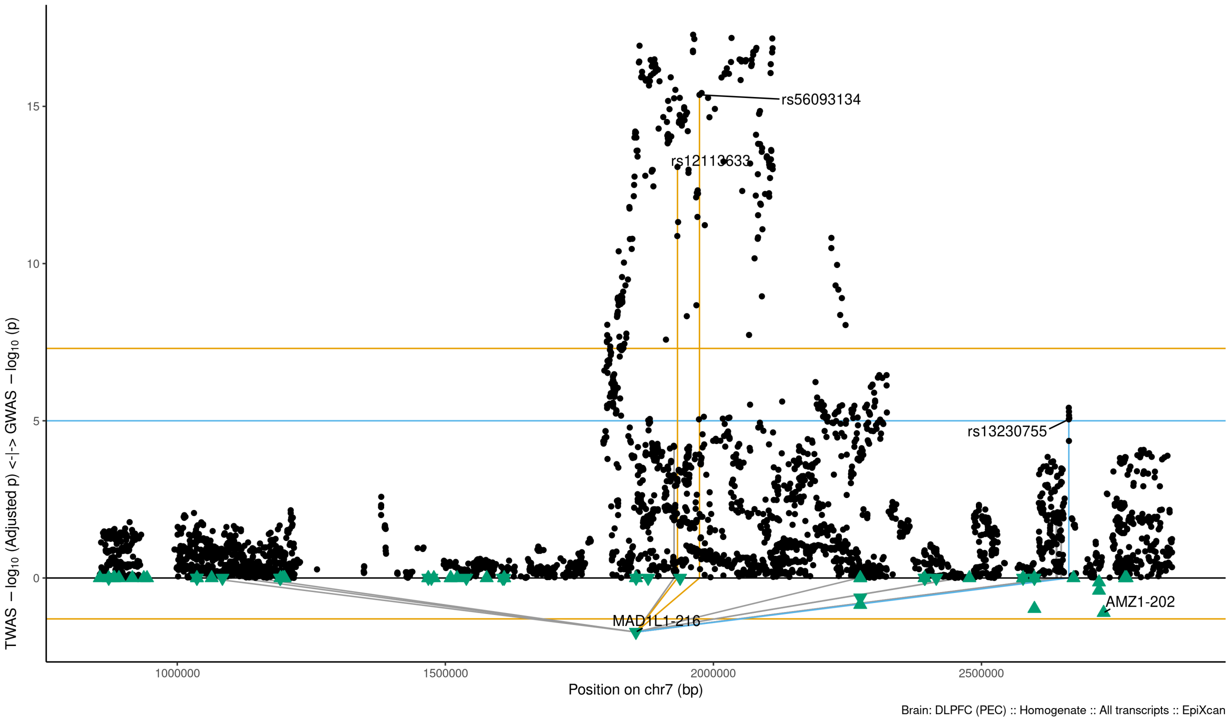

Figure S9-41: Regional Miami plots for MAD1L1216 gene/transcript : Regional Miami plots for the primary MD GWASs corresponding to the genomic region of the MAD1L1216 transcript (1Mbp window from start site). Please also refer to Supplemental Table SXX for details. Top panel shows the GWAS results (black dots); blue line corresponds to $p = 1 \times 10 - 5$, orange line to $p = 5 \times 10 - 8$ (genome-wide significance). Bottom panel shows the TWAS results (green triangles facing upwards or downwards for a positive or negative z-score respectively (up- or down-regultion); only the transcripts with Bonferroni-adjusted $p < 0.1$ are labelled for clarity) for different transcripts (genes are represented by both gene expression and isoform expression); orange line corresponds to Bonferroni-adjusted p = 0.05. Each transcript that is Bonferroni-significant in the region is connected with lines to the SNPs that contribute to its transcriptomic imputation model; lines are grey when the SNPs have a $p > 1 \times 10 - 5$, blue when $p < 1 \times 10 - 5$ but $> 510 - 8$ and orange when $p < 5 \times 10 - 8$. The SNPs that are above the blue line and contribute to the transcriptomic imputation models of significant transcripts are labelled.

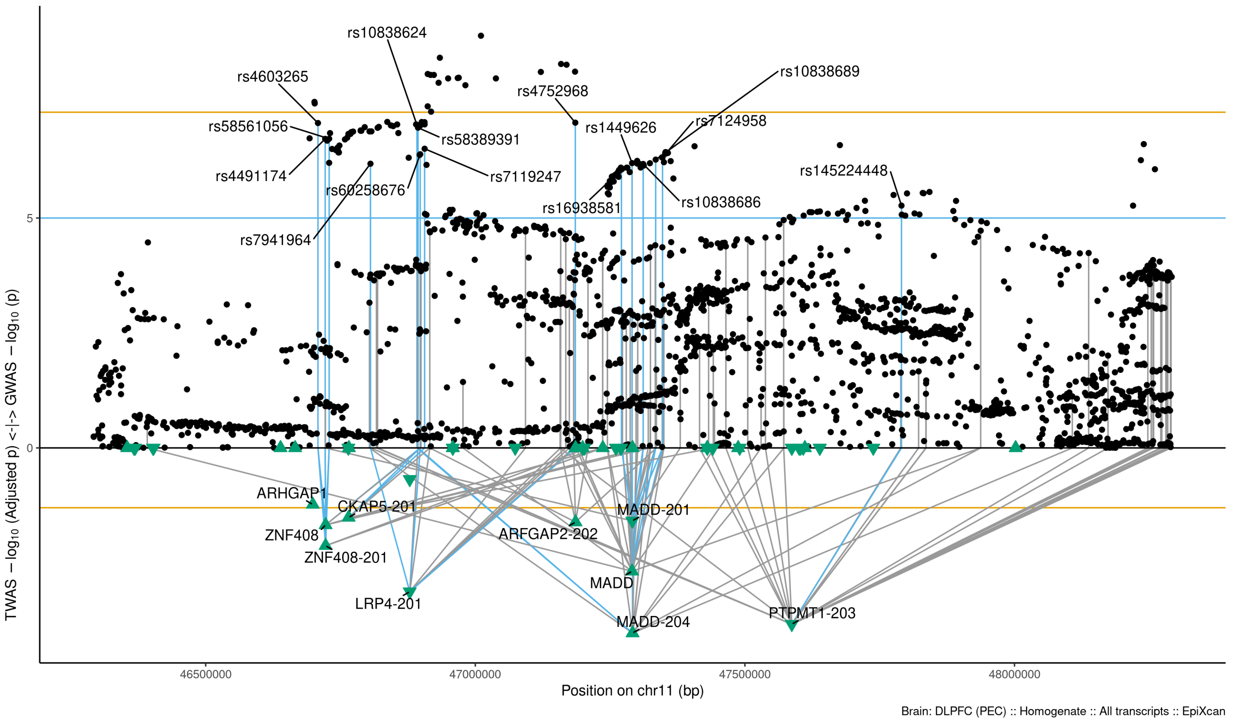

Figure S9-42: Regional Miami plots for MADD204 gene/transcript : Regional Miami plots for the primary MD GWASs corresponding to the genomic region of the MADD204 transcript (1Mbp window from start site). Please also refer to Supplemental Table SXX for details. Top panel shows the GWAS results (black dots); blue line corresponds to $p = 1 \times 10 - 5$, orange line to $p = 5 \times 10 - 8$ (genome-wide significance). Bottom panel shows the TWAS results (green triangles facing upwards or downwards for a positive or negative z-score respectively (up- or down-regultion); only the transcripts with Bonferroni-adjusted $p < 0.1$ are labelled for clarity) for different transcripts (genes are represented by both gene expression and isoform expression); orange line corresponds to Bonferroni-adjusted p = 0.05. Each transcript that is Bonferroni-significant in the region is connected with lines to the SNPs that contribute to its transcriptomic imputation model; lines are grey when the SNPs have a $p > 1 \times 10 - 5$, blue when $p < 1 \times 10 - 5$ but $> 510 - 8$ and orange when $p < 5 \times 10 - 8$. The SNPs that are above the blue line and contribute to the transcriptomic imputation models of significant transcripts are labelled.

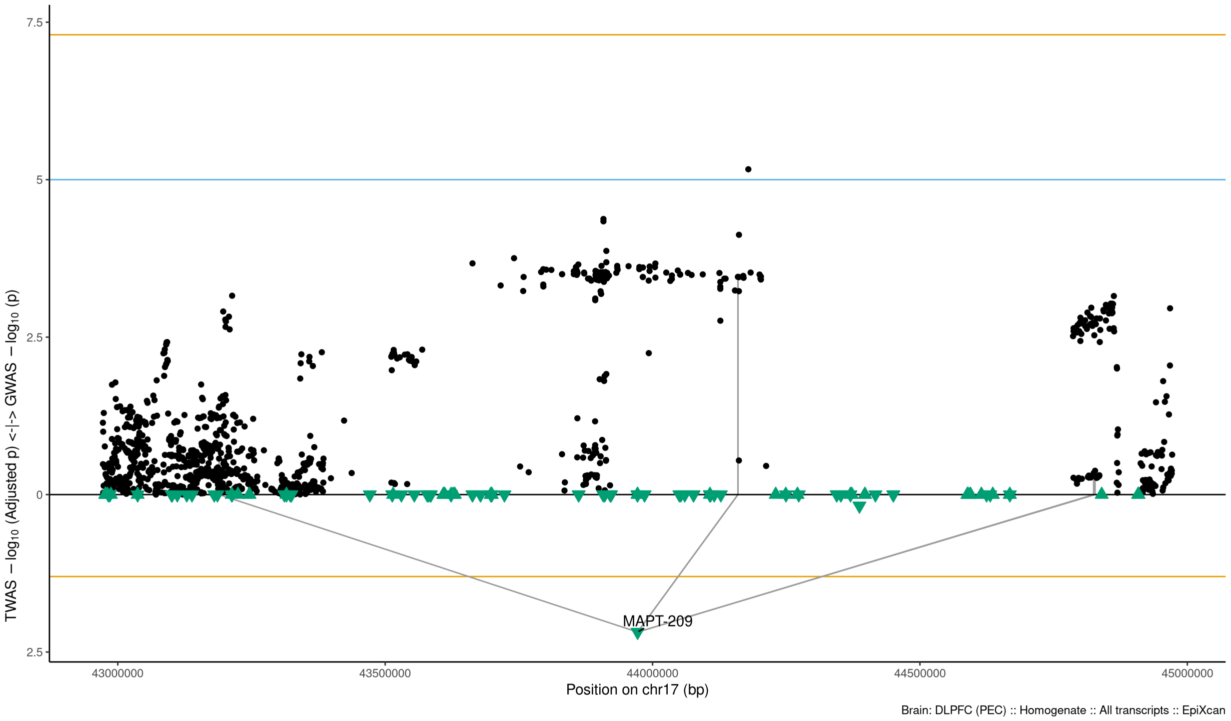

Figure S9-43: Regional Miami plots for MAPT209 gene/transcript : Regional Miami plots for the primary MD GWASs corresponding to the genomic region of the MAPT209 transcript (1Mbp window from start site). Please also refer to Supplemental Table SXX for details. Top panel shows the GWAS results (black dots); blue line corresponds to $p = 1 \times 10 - 5$, orange line to $p = 5 \times 10 - 8$ (genome-wide significance). Bottom panel shows the TWAS results (green triangles facing upwards or downwards for a positive or negative z-score respectively (up- or down-regultion); only the transcripts with Bonferroni-adjusted $p < 0.1$ are labelled for clarity) for different transcripts (genes are represented by both gene expression and isoform expression); orange line corresponds to Bonferroni-adjusted p = 0.05. Each transcript that is Bonferroni-significant in the region is connected with lines to the SNPs that contribute to its transcriptomic imputation model; lines are grey when the SNPs have a $p > 1 \times 10 - 5$, blue when $p < 1 \times 10 - 5$ but $> 510 - 8$ and orange when $p < 5 \times 10 - 8$. The SNPs that are above the blue line and contribute to the transcriptomic imputation models of significant transcripts are labelled.

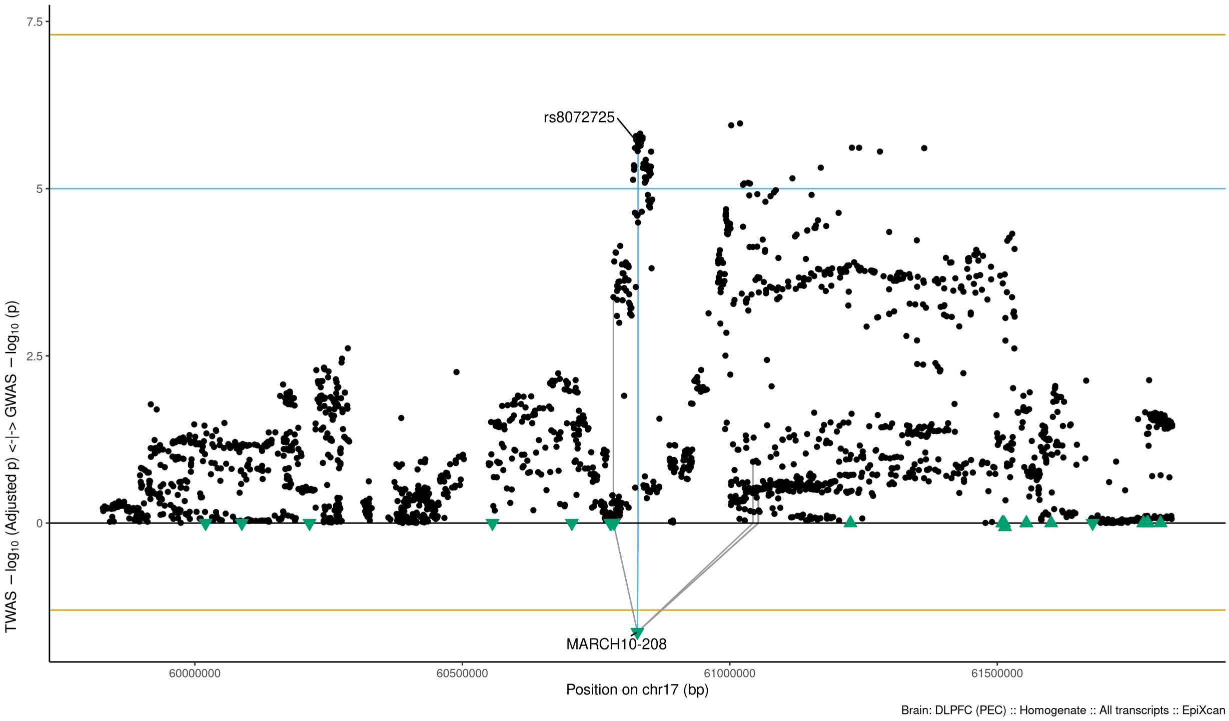

Figure S9-44: Regional Miami plots for MARCH10208 gene/transcript : Regional Miami plots for the primary MD GWASs corresponding to the genomic region of the MARCH10208 transcript (1Mbp window from start site). Please also refer to Supplemental Table SXX for details. Top panel shows the GWAS results (black dots); blue line corresponds to $p = 1 \times 10 - 5$, orange line to $p = 5 \times 10 - 8$ (genome-wide significance). Bottom panel shows the TWAS results (green triangles facing upwards or downwards for a positive or negative z-score respectively (up- or down-regultion); only the transcripts with Bonferroni-adjusted $p < 0.1$ are labelled for clarity) for different transcripts (genes are represented by both gene expression and isoform expression); orange line corresponds to Bonferroni-adjusted p = 0.05. Each transcript that is Bonferroni-significant in the region is connected with lines to the SNPs that contribute to its transcriptomic imputation model; lines are grey when the SNPs have a $p > 1 \times 10 - 5$, blue when $p < 1 \times 10 - 5$ but $> 510 - 8$ and orange when $p < 5 \times 10 - 8$. The SNPs that are above the blue line and contribute to the transcriptomic imputation models of significant transcripts are labelled.

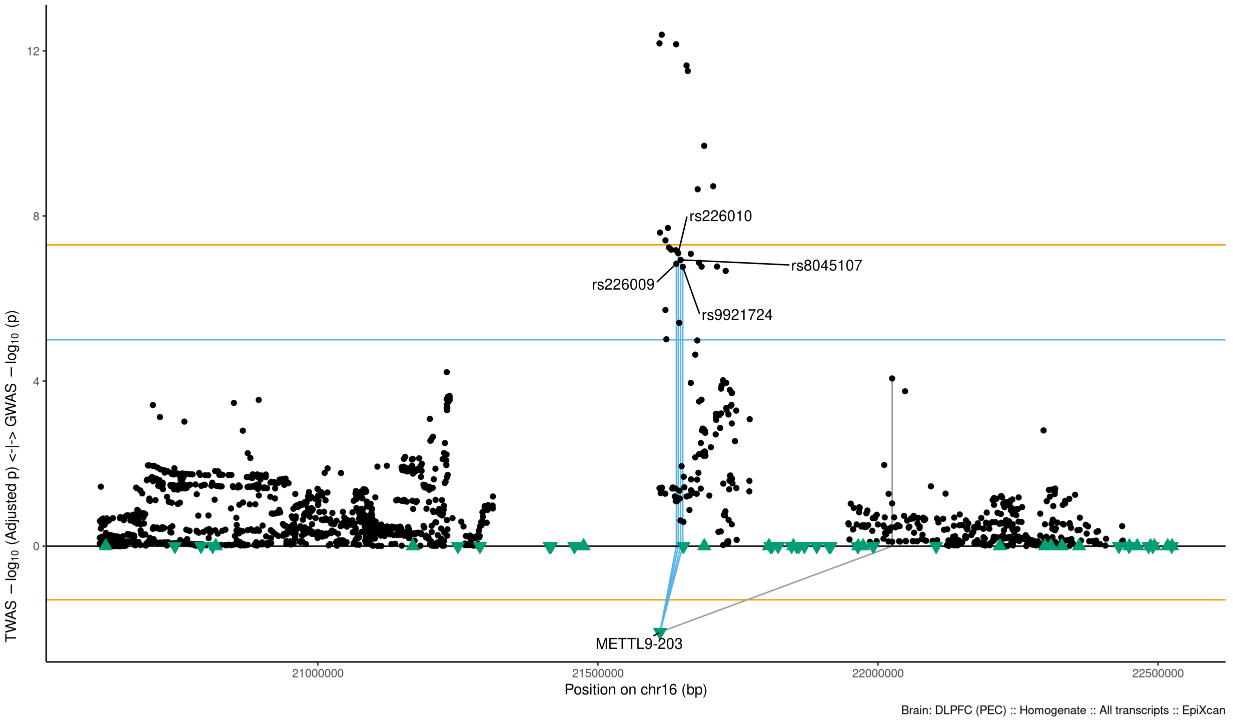

Figure S9-45: Regional Miami plots for METTL9203 gene/transcript : Regional Miami plots for primary MD GWASs corresponding to the genomic region of the METTL9203 transcript (1Mbp window from start site). Please also refer to Supplemental Table SXX for details. Top panel shows the GWAS results (black dots); blue line corresponds to $p = 1 \times 10 - 5$, orange line to $p = 5 \times 10 - 8$ (genome-wide significance). Bottom panel shows the TWAS results (green triangles facing upwards or downwards for a positive or negative z-score respectively (up- or down-regultion); only the transcripts with Bonferroni-adjusted $p < 0.1$ are labelled for clarity) for different transcripts (genes are represented by both gene expression and isoform expression); orange line corresponds to Bonferroni-adjusted p = 0.05. Each transcript that is Bonferroni-significant in the region is connected with lines to the SNPs that contribute to its transcriptomic imputation model; lines are grey when the SNPs have a $p > 1 \times 10 - 5$, blue when $p < 1 \times 10 - 5$ but $> 510 - 8$ and orange when $p < 5 \times 10 - 8$. The SNPs that are above the blue line and contribute to the transcriptomic imputation models of significant transcripts are labelled.

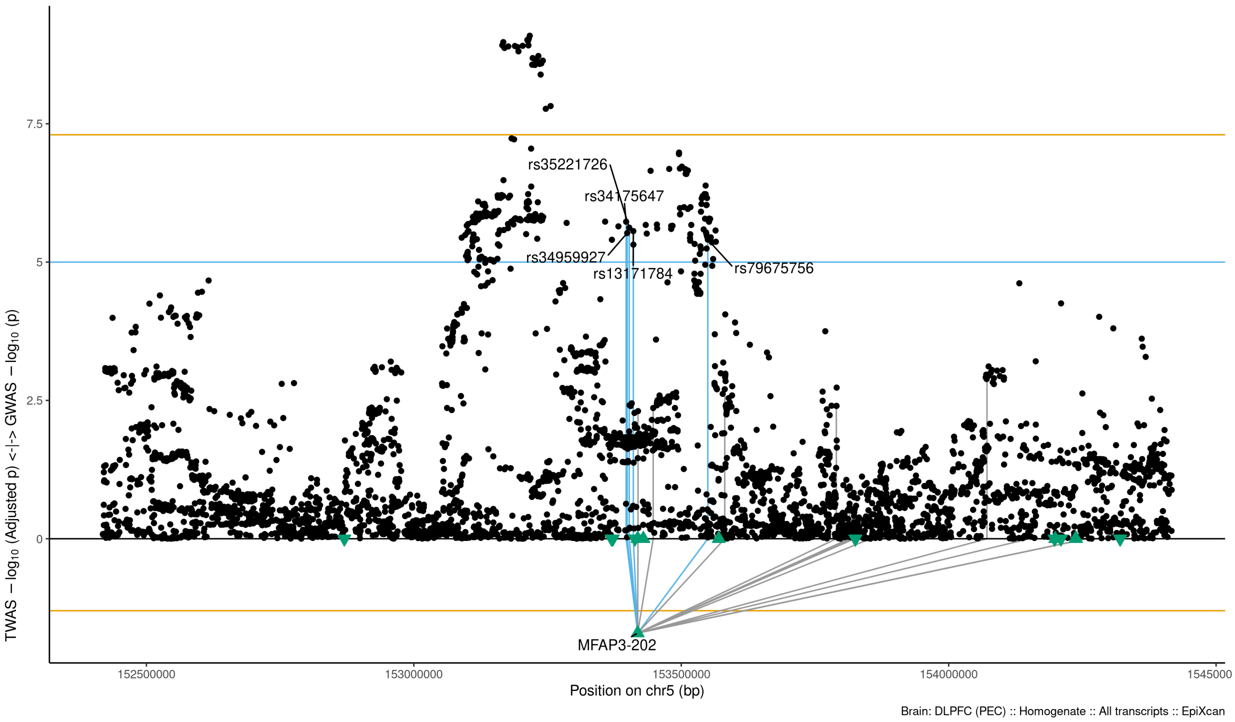

Figure S9-46: Regional Miami plots for MFAP3202 gene/transcript : Regional Miami plots for primary MD GWASs corresponding to the genomic region of the MFAP3202 transcript (1Mbp window from start site). Please also refer to Supplemental Table SXX for details. Top panel shows the GWAS results (black dots); blue line corresponds to $p = 1 \times 10 - 5$, orange line to $p = 5 \times 10 - 8$ (genome-wide significance). Bottom panel shows the TWAS results (green triangles facing upwards or downwards for a positive or negative z-score respectively (up- or down-regultion); only the transcripts with Bonferroni-adjusted $p < 0.1$ are labelled for clarity) for different transcripts (genes are represented by both gene expression and isoform expression); orange line corresponds to Bonferroni-adjusted p = 0.05. Each transcript that is Bonferroni-significant in the region is connected with lines to the SNPs that contribute to its transcriptomic imputation model; lines are grey when the SNPs have a $p > 1 \times 10 - 5$, blue when $p < 1 \times 10 - 5$ but $> 510 - 8$ and orange when $p < 5 \times 10 - 8$. The SNPs that are above the blue line and contribute to the transcriptomic imputation models of significant transcripts are labelled.

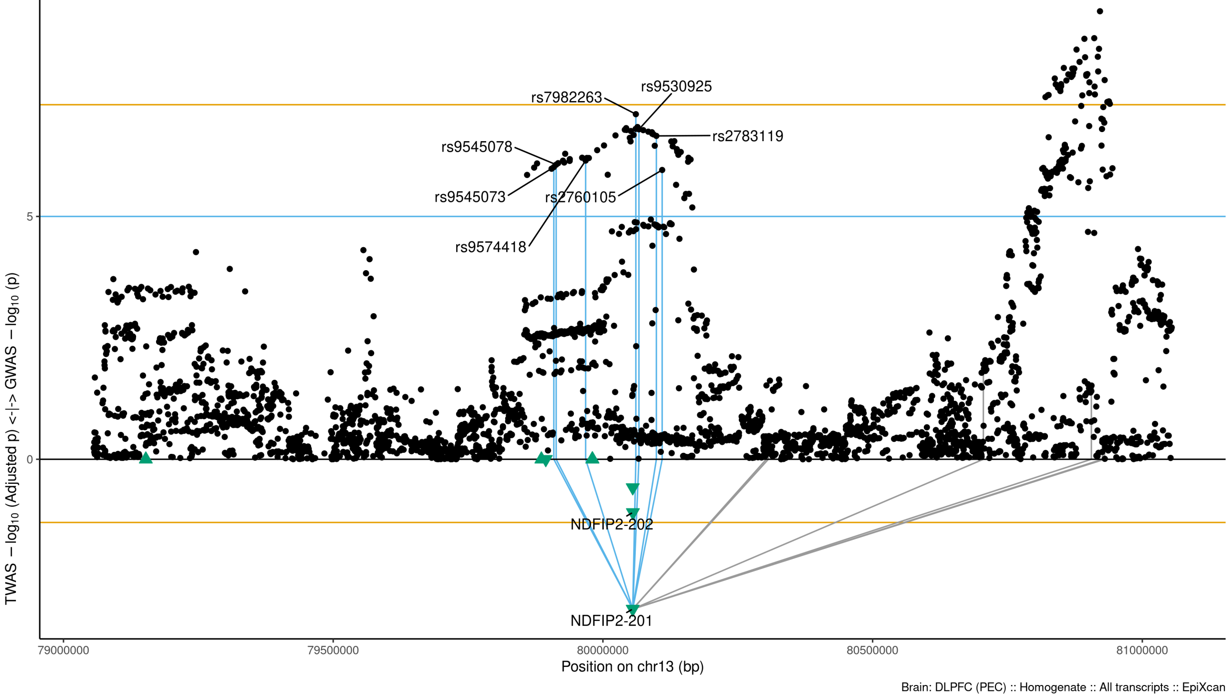

Figure S9-47: Regional Miami plots for NDFIP2201 gene/transcript : Regional Miami plots for primary MD GWASs corresponding to the genomic region of the NDFIP2201 transcript (1Mbp window from start site). Please also refer to Supplemental Table SXX for details. Top panel shows the GWAS results (black dots); blue line corresponds to $p = 1 \times 10 - 5$, orange line to $p = 5 \times 10 - 8$ (genome-wide significance). Bottom panel shows the TWAS results (green triangles facing upwards or downwards for a positive or negative z-score respectively (up- or down-regultion); only the transcripts with Bonferroni-adjusted $p < 0.1$ are labelled for clarity) for different transcripts (genes are represented by both gene expression and isoform expression); orange line corresponds to Bonferroni-adjusted p = 0.05. Each transcript that is Bonferroni-significant in the region is connected with lines to the SNPs that contribute to its transcriptomic imputation model; lines are grey when the SNPs have a $p > 1 \times 10 - 5$, blue when $p < 1 \times 10 - 5$ but $> 510 - 8$ and orange when $p < 5 \times 10 - 8$. The SNPs that are above the blue line and contribute to the transcriptomic imputation models of significant transcripts are labelled.

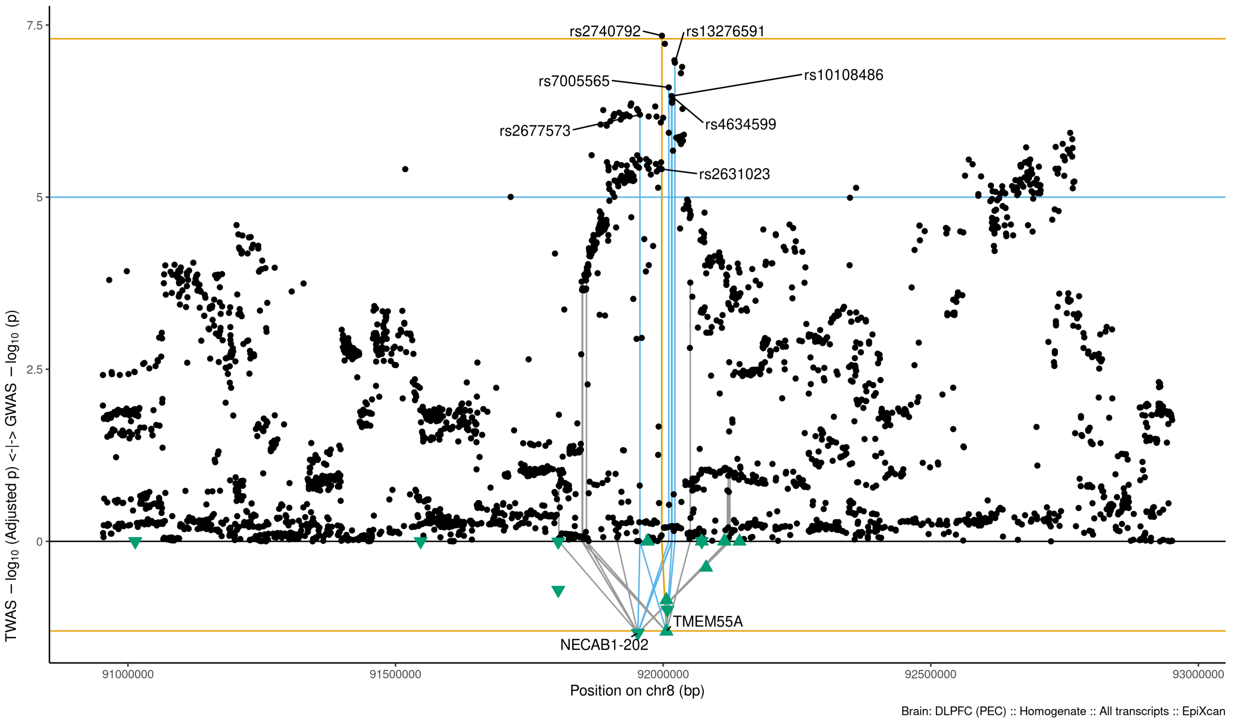

Figure S9-48: Regional Miami plots for NECAB1202 gene/transcript : Regional Miami plots for primary MD GWASs corresponding to the genomic region of the NECAB1202 transcript (1Mbp window from start site). Please also refer to Supplemental Table SXX for details. Top panel shows the GWAS results (black dots); blue line corresponds to $p = 1 \times 10 - 5$, orange line to $p = 5 \times 10 - 8$ (genome-wide significance). Bottom panel shows the TWAS results (green triangles facing upwards or downwards for a positive or negative z-score respectively (up- or down-regultion); only the transcripts with Bonferroni-adjusted $p < 0.1$ are labelled for clarity) for different transcripts (genes are represented by both gene expression and isoform expression); orange line corresponds to Bonferroni-adjusted p = 0.05. Each transcript that is Bonferroni-significant in the region is connected with lines to the SNPs that contribute to its transcriptomic imputation model; lines are grey when the SNPs have a $p > 1 \times 10 - 5$, blue when $p < 1 \times 10 - 5$ but $> 510 - 8$ and orange when $p < 5 \times 10 - 8$. The SNPs that are above the blue line and contribute to the transcriptomic imputation models of significant transcripts are labelled.

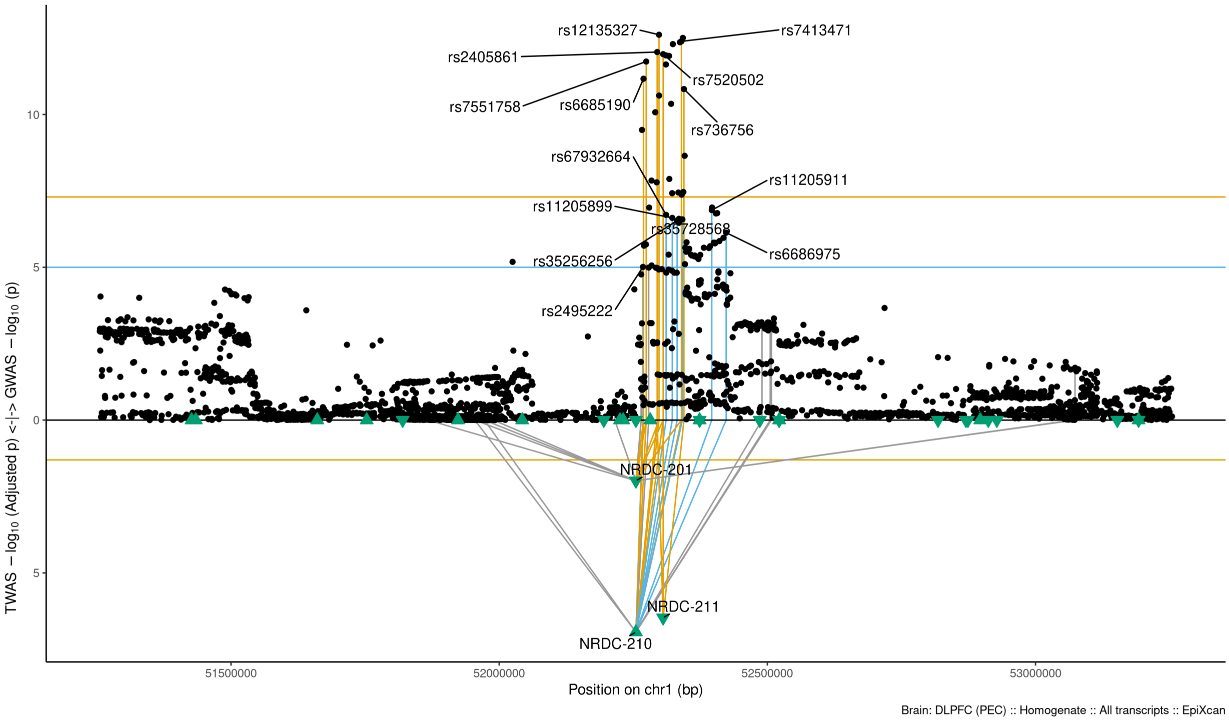

Figure S9-49: Regional Miami plots for NRDC210 gene/transcript : Regional Miami plots for primary MD GWASs corresponding to the genomic region of the NRDC210 transcript (1Mbp window from start site). Please also refer to Supplemental Table SXX for details. Top panel shows the GWAS results (black dots); blue line corresponds to $p = 1 \times 10 - 5$, orange line to $p = 5 \times 10 - 8$ (genome-wide significance). Bottom panel shows the TWAS results (green triangles facing upwards or downwards for a positive or negative z-score respectively (up- or down-regultion); only the transcripts with Bonferroni-adjusted $p < 0.1$ are labelled for clarity) for different transcripts (genes are represented by both gene expression and isoform expression); orange line corresponds to Bonferroni-adjusted p = 0.05. Each transcript that is Bonferroni-significant in the region is connected with lines to the SNPs that contribute to its transcriptomic imputation model; lines are grey when the SNPs have a $p > 1 \times 10 - 5$, blue when $p < 1 \times 10 - 5$ but $> 510 - 8$ and orange when $p < 5 \times 10 - 8$. The SNPs that are above the blue line and contribute to the transcriptomic imputation models of significant transcripts are labelled.

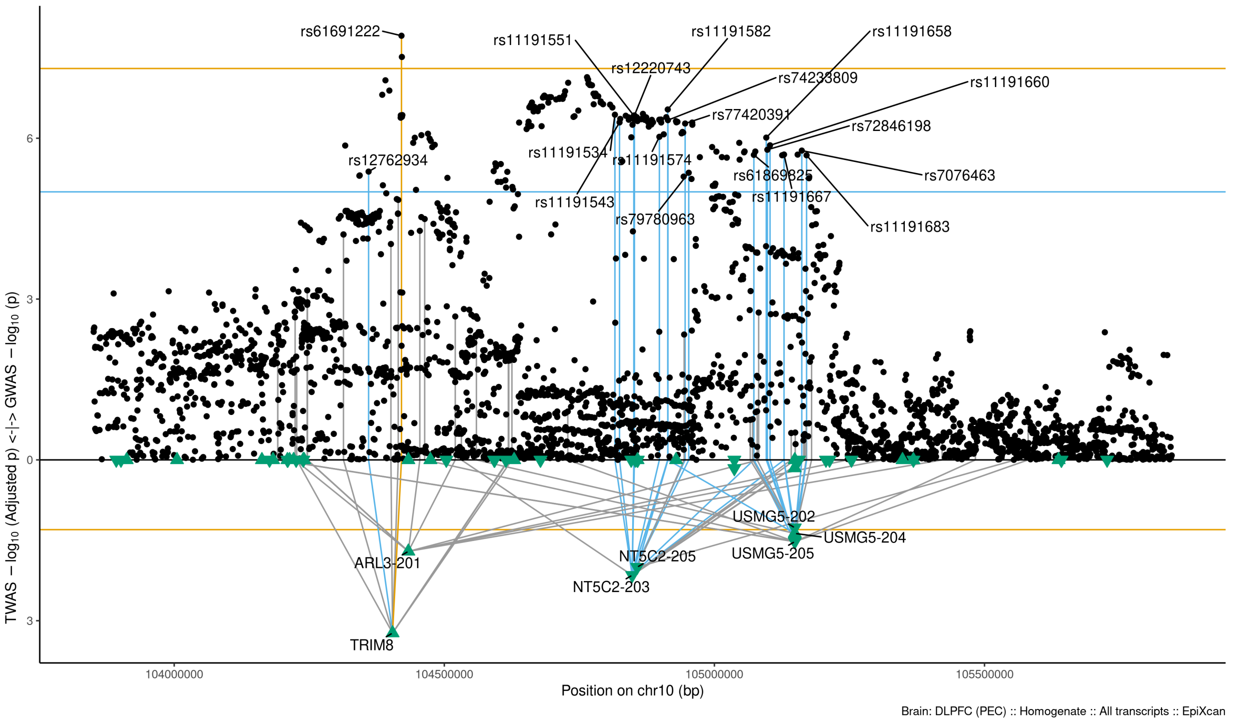

Figure S9-50: Regional Miami plots for NT5C2203 gene/transcript : Regional Miami plots for primary MD GWASs corresponding to the genomic region of the NT5C2203 transcript (1Mbp window from start site). Please also refer to Supplemental Table SXX for details. Top panel shows the GWAS results (black dots); blue line corresponds to $p = 1 \times 10 - 5$, orange line to $p = 5 \times 10 - 8$ (genome-wide significance). Bottom panel shows the TWAS results (green triangles facing upwards or downwards for a positive or negative z-score respectively (up- or down-regultion); only the transcripts with Bonferroni-adjusted $p < 0.1$ are labelled for clarity) for different transcripts (genes are represented by both gene expression and isoform expression); orange line corresponds to Bonferroni-adjusted p = 0.05. Each transcript that is Bonferroni-significant in the region is connected with lines to the SNPs that contribute to its transcriptomic imputation model; lines are grey when the SNPs have a $p > 1 \times 10 - 5$, blue when $p < 1 \times 10 - 5$ but $> 510 - 8$ and orange when $p < 5 \times 10 - 8$. The SNPs that are above the blue line and contribute to the transcriptomic imputation models of significant transcripts are labelled.

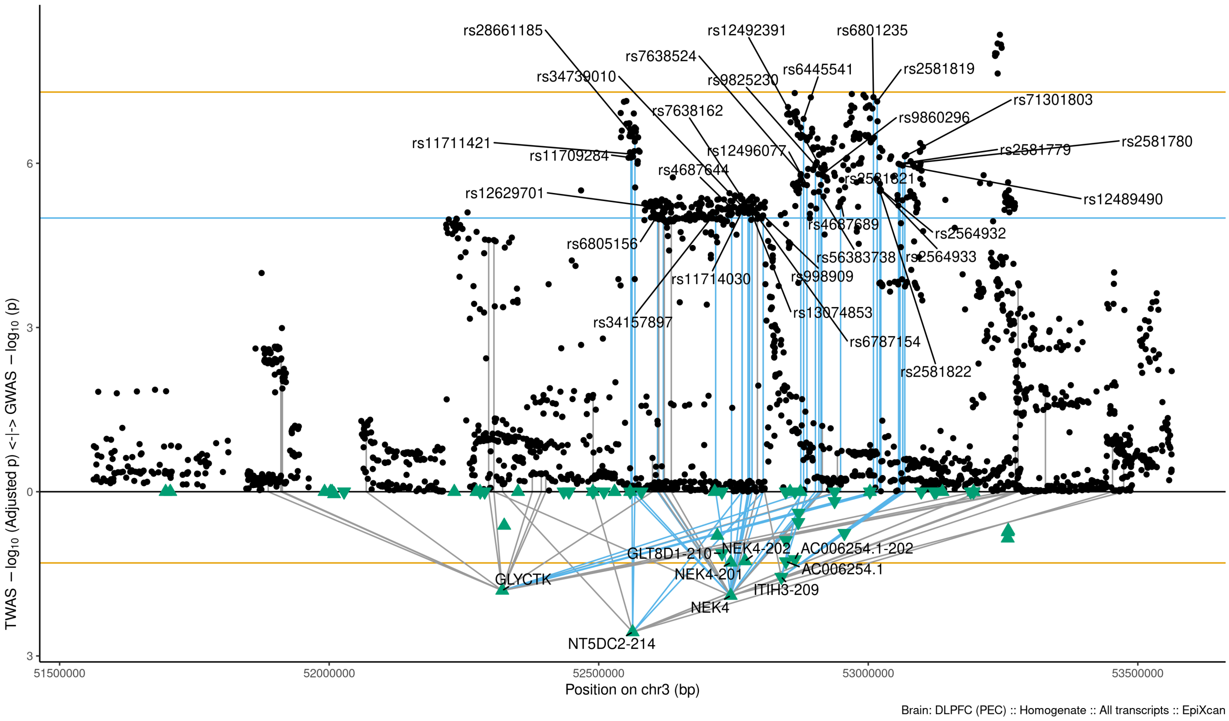

Figure S9-51: Regional Miami plots for NT5DC2214 gene/transcript : Regional Miami plots for primary MD GWASs corresponding to the genomic region of the NT5DC2214 transcript (1Mbp window from start site). Please also refer to Supplemental Table SXX for details. Top panel shows the GWAS results (black dots); blue line corresponds to $p = 1 \times 10 - 5$, orange line to $p = 5 \times 10 - 8$ (genome-wide significance). Bottom panel shows the TWAS results (green triangles facing upwards or downwards for a positive or negative z-score respectively (up- or down-regultion); only the transcripts with Bonferroni-adjusted $p < 0.1$ are labelled for clarity) for different transcripts (genes are represented by both gene expression and isoform expression); orange line corresponds to Bonferroni-adjusted p = 0.05. Each transcript that is Bonferroni-significant in the region is connected with lines to the SNPs that contribute to its transcriptomic imputation model; lines are grey when the SNPs have a $p > 1 \times 10 - 5$, blue when $p < 1 \times 10 - 5$ but $> 510 - 8$ and orange when $p < 5 \times 10 - 8$. The SNPs that are above the blue line and contribute to the transcriptomic imputation models of significant transcripts are labelled.

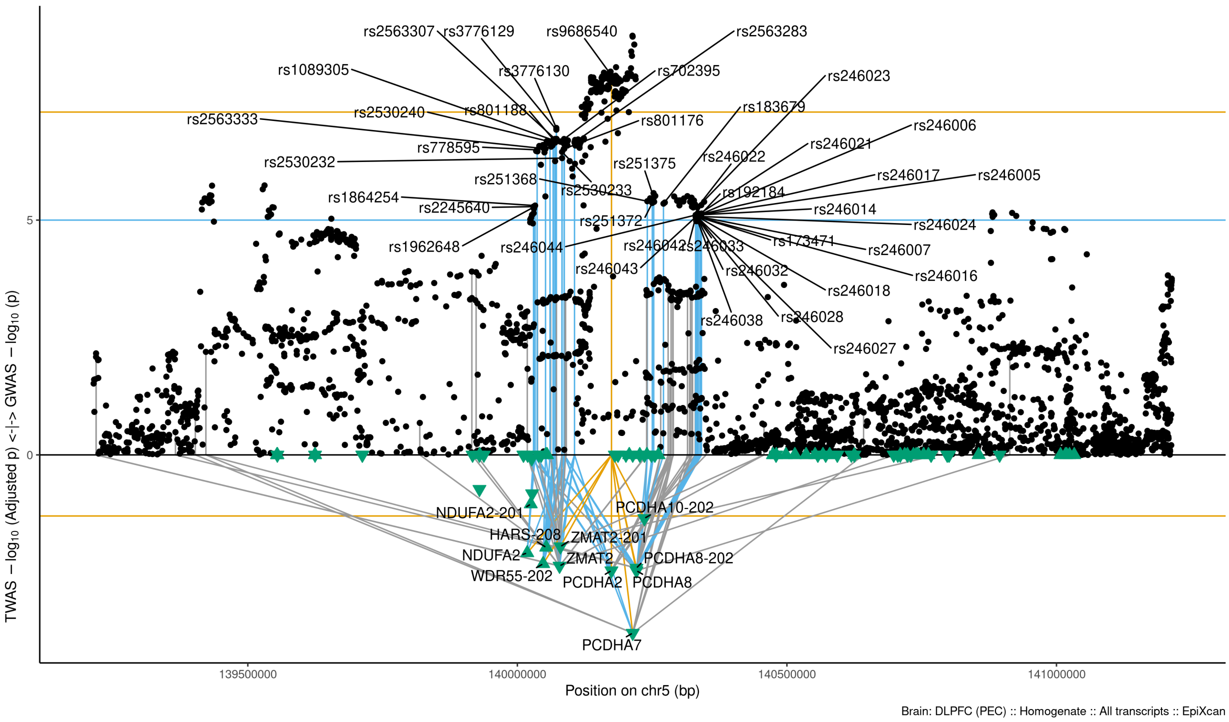

Figure S9-52: Regional Miami plots for PCDHA7 gene/transcript : Regional Miami plots for primary MD GWASs corresponding to the genomic region of the PCDHA7 transcript (1Mbp window from start site). Please also refer to Supplemental Table SXX for details. Top panel shows the GWAS results (black dots); blue line corresponds to $p = 1 \times 10 - 5$, orange line to $p = 5 \times 10 - 8$ (genome-wide significance). Bottom panel shows the TWAS results (green triangles facing upwards or downwards for a positive or negative z-score respectively (up- or down-regultion); only the transcripts with Bonferroni-adjusted $p < 0.1$ are labelled for clarity) for different transcripts (genes are represented by both gene expression and isoform expression); orange line corresponds to Bonferroni-adjusted p = 0.05. Each transcript that is Bonferroni-significant in the region is connected with lines to the SNPs that contribute to its transcriptomic imputation model; lines are grey when the SNPs have a $p > 1 \times 10 - 5$, blue when $p < 1 \times 10 - 5$ but $> 510 - 8$ and orange when $p < 5 \times 10 - 8$. The SNPs that are above the blue line and contribute to the transcriptomic imputation models of significant transcripts are labelled.

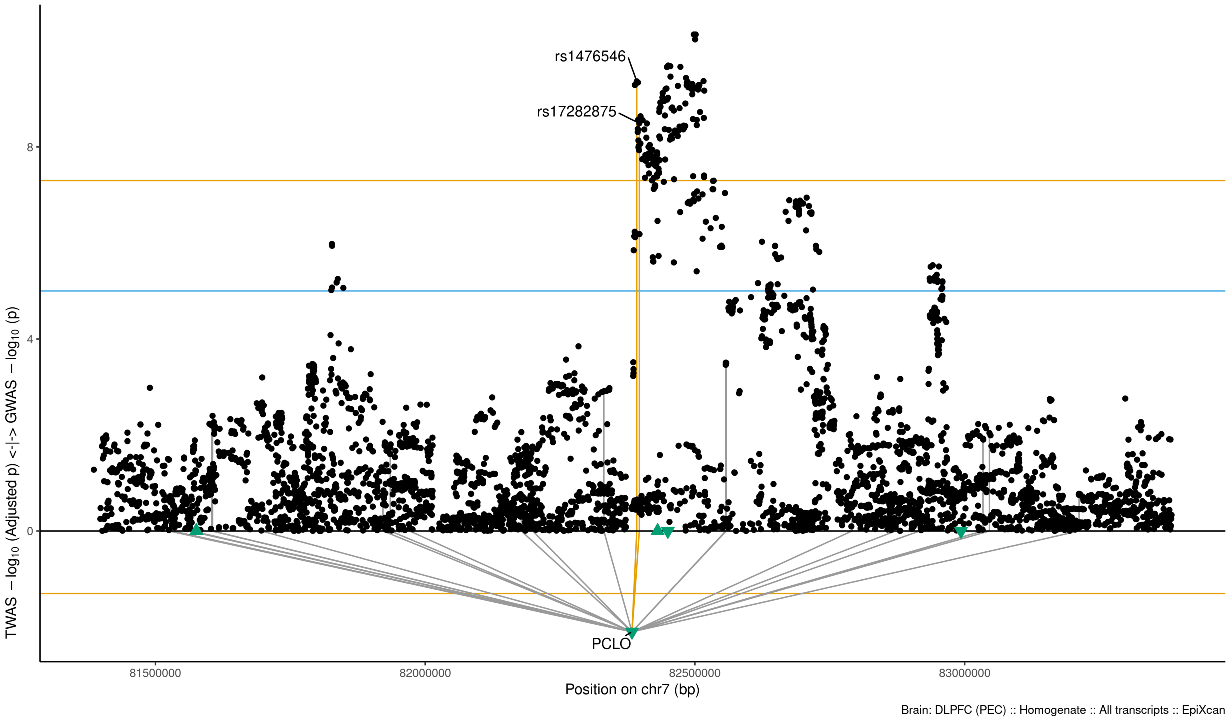

Figure S9-53: Regional Miami plots for PCLO gene/transcript : Regional Miami plots for primary MD GWASs corresponding to the genomic region of the PCLO transcript (1Mbp window from start site). Please also refer to Supplemental Table SXX for details. Top panel shows the GWAS results (black dots); blue line corresponds to $p = 1 \times 10 - 5$, orange line to $p = 5 \times 10 - 8$ (genome-wide significance). Bottom panel shows the TWAS results (green triangles facing upwards or downwards for a positive or negative z-score respectively (up- or down-regultion); only the transcripts with Bonferroni-adjusted $p < 0.1$ are labelled for clarity) for different transcripts (genes are represented by both gene expression and isoform expression); orange line corresponds to Bonferroni-adjusted p = 0.05. Each transcript that is Bonferroni-significant in the region is connected with lines to the SNPs that contribute to its transcriptomic imputation model; lines are grey when the SNPs have a $p > 1 \times 10 - 5$, blue when $p < 1 \times 10 - 5$ but $> 510 - 8$ and orange when $p < 5 \times 10 - 8$. The SNPs that are above the blue line and contribute to the transcriptomic imputation models of significant transcripts are labelled.

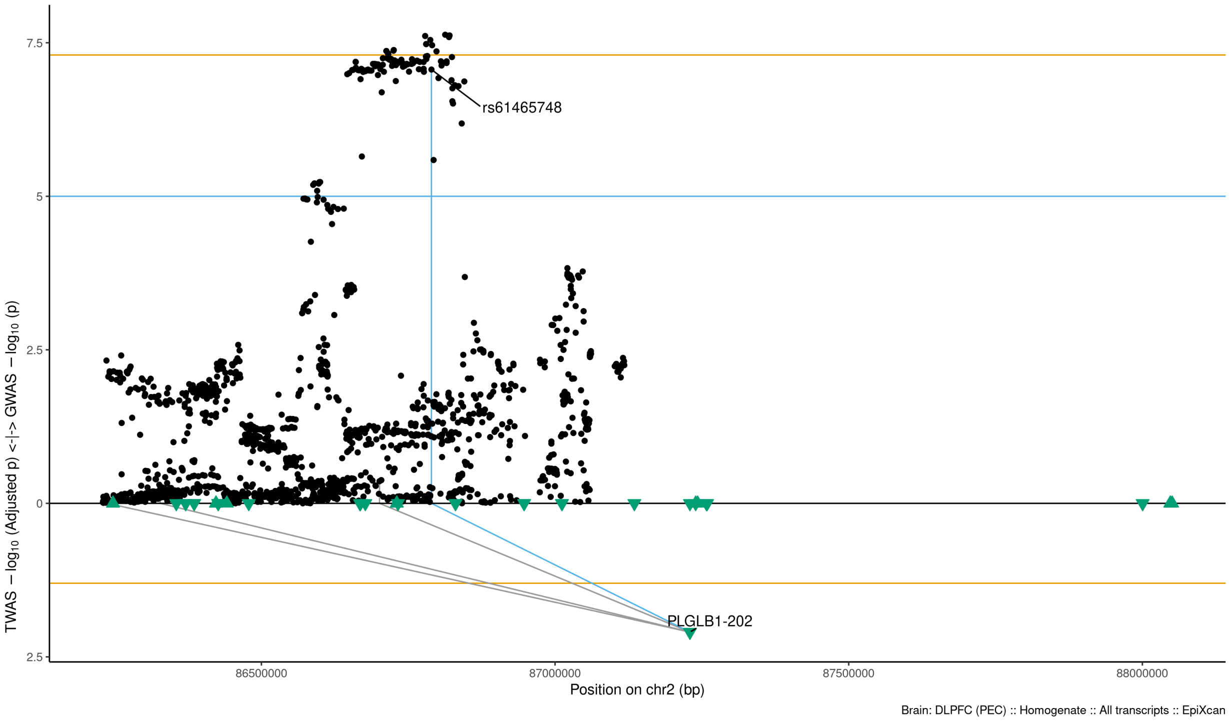

Figure S9-54: Regional Miami plots for PLGLB1202 gene/transcript : Regional Miami plots for primary MD GWASs corresponding to the genomic region of the PLGLB1202 transcript (1Mbp window from start site). Please also refer to Supplemental Table SXX for details. Top panel shows the GWAS results (black dots); blue line corresponds to $p = 1 \times 10 - 5$, orange line to $p = 5 \times 10 - 8$ (genome-wide significance). Bottom panel shows the TWAS results (green triangles facing upwards or downwards for a positive or negative z-score respectively (up- or down-regultion); only the transcripts with Bonferroni-adjusted $p < 0.1$ are labelled for clarity) for different transcripts (genes are represented by both gene expression and isoform expression); orange line corresponds to Bonferroni-adjusted p = 0.05. Each transcript that is Bonferroni-significant in the region is connected with lines to the SNPs that contribute to its transcriptomic imputation model; lines are grey when the SNPs have a $p > 1 \times 10 - 5$, blue when $p < 1 \times 10 - 5$ but $> 510 - 8$ and orange when $p < 5 \times 10 - 8$. The SNPs that are above the blue line and contribute to the transcriptomic imputation models of significant transcripts are labelled.

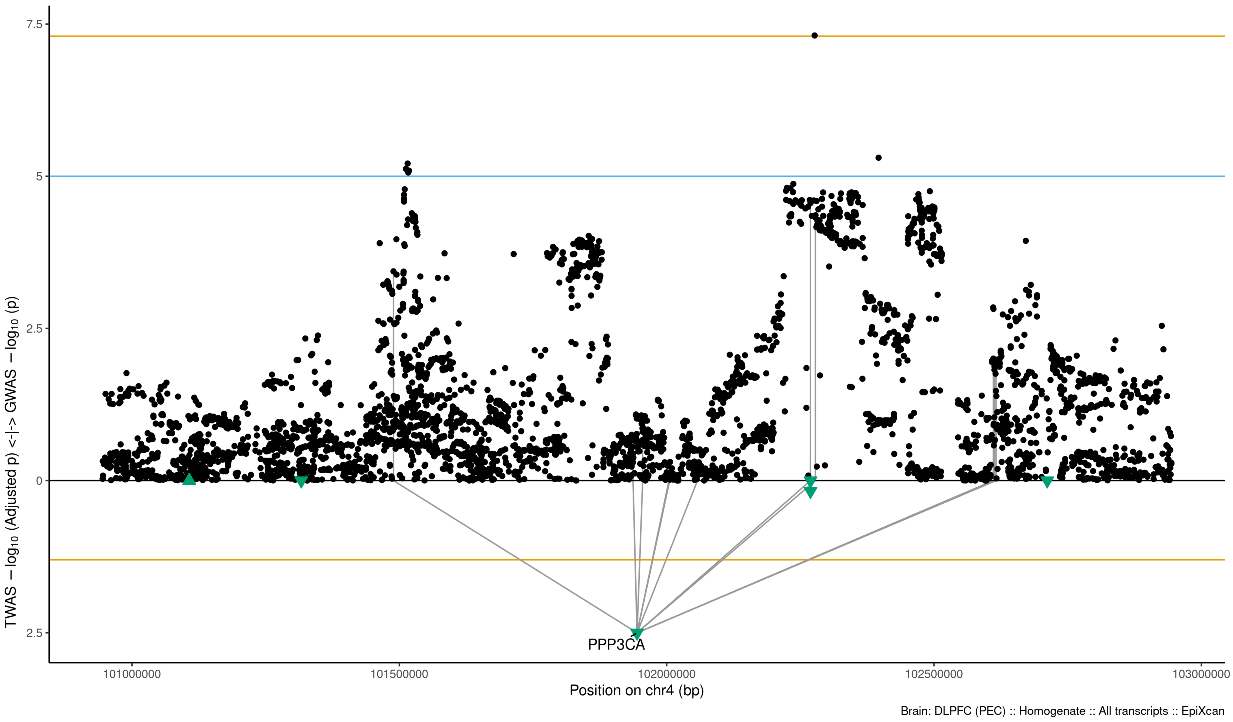

Figure S9-55: Regional Miami plots for PPP3CA gene/transcript : Regional Miami plots for primary MD GWASs corresponding to the genomic region of the PPP3CA transcript (1Mbp window from start site). Please also refer to Supplemental Table SXX for details. Top panel shows the GWAS results (black dots); blue line corresponds to $p = 1 \times 10 - 5$, orange line to $p = 5 \times 10 - 8$ (genome-wide significance). Bottom panel shows the TWAS results (green triangles facing upwards or downwards for a positive or negative z-score respectively (up- or down-regultion); only the transcripts with Bonferroni-adjusted $p < 0.1$ are labelled for clarity) for different transcripts (genes are represented by both gene expression and isoform expression); orange line corresponds to Bonferroni-adjusted p = 0.05. Each transcript that is Bonferroni-significant in the region is connected with lines to the SNPs that contribute to its transcriptomic imputation model; lines are grey when the SNPs have a $p > 1 \times 10 - 5$, blue when $p < 1 \times 10 - 5$ but $> 510 - 8$ and orange when $p < 5 \times 10 - 8$. The SNPs that are above the blue line and contribute to the transcriptomic imputation models of significant transcripts are labelled.

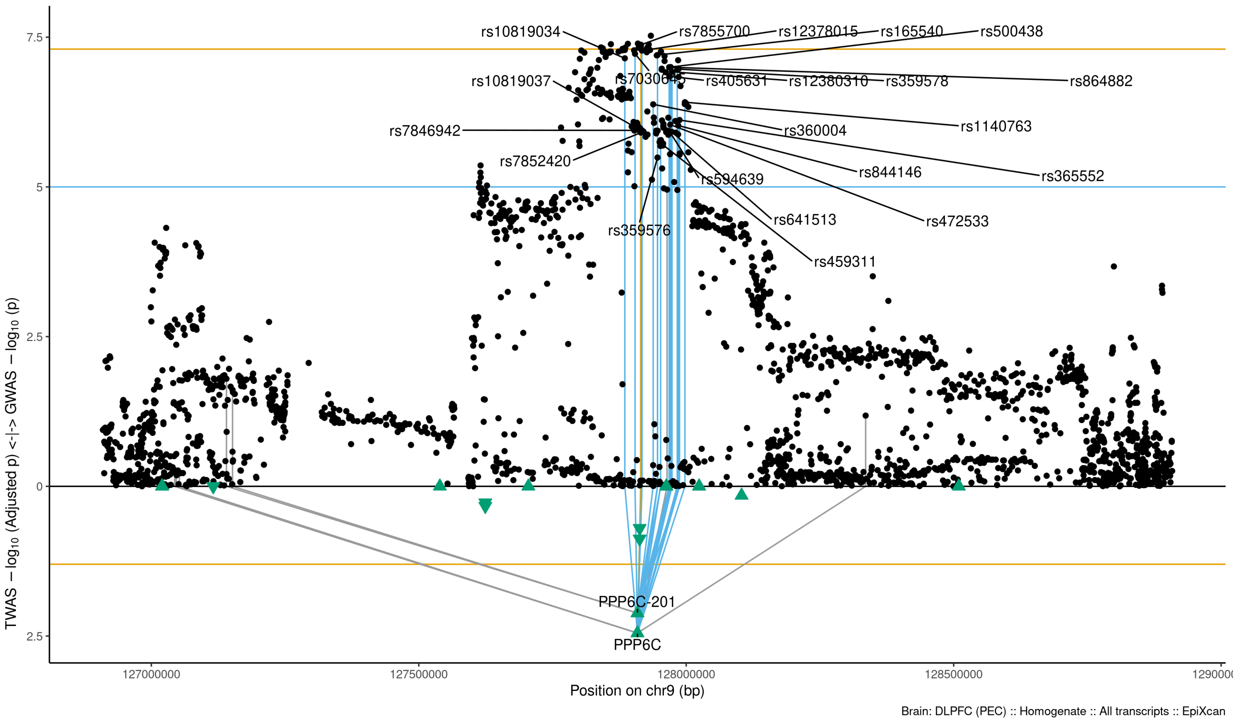

Figure S9-56: Regional Miami plots for PPP6C gene/transcript : Regional Miami plots for primary MD GWASs corresponding to the genomic region of the PPP6C transcript (1Mbp window from start site). Please also refer to Supplemental Table SXX for details. Top panel shows the GWAS results (black dots); blue line corresponds to $p = 1 \times 10 - 5$, orange line to $p = 5 \times 10 - 8$ (genome-wide significance). Bottom panel shows the TWAS results (green triangles facing upwards or downwards for a positive or negative z-score respectively (up- or down-regultion); only the transcripts with Bonferroni-adjusted $p < 0.1$ are labelled for clarity) for different transcripts (genes are represented by both gene expression and isoform expression); orange line corresponds to Bonferroni-adjusted p = 0.05. Each transcript that is Bonferroni-significant in the region is connected with lines to the SNPs that contribute to its transcriptomic imputation model; lines are grey when the SNPs have a $p > 1 \times 10 - 5$, blue when $p < 1 \times 10 - 5$ but $> 510 - 8$ and orange when $p < 5 \times 10 - 8$. The SNPs that are above the blue line and contribute to the transcriptomic imputation models of significant transcripts are labelled.

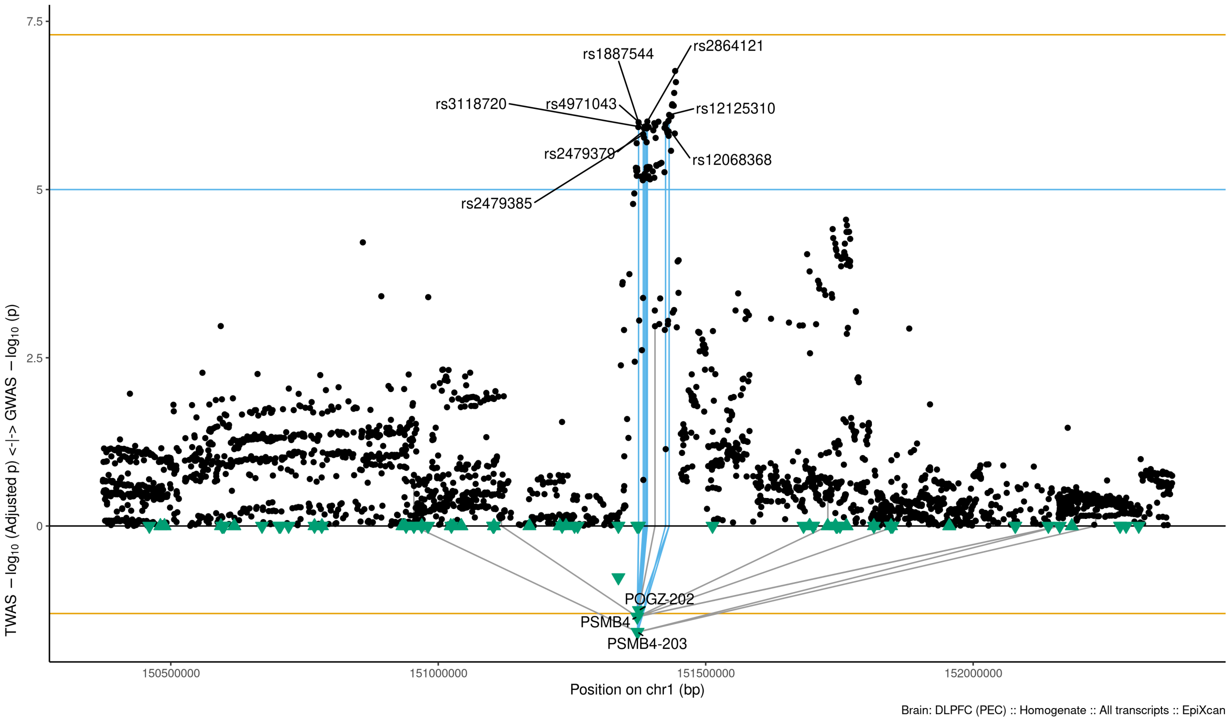

Figure S9-57: Regional Miami plots for PSMB4203 gene/transcript : Regional Miami plots for primary MD GWASs corresponding to the genomic region of the PSMB4203 transcript (1Mbp window from start site). Please also refer to Supplemental Table SXX for details. Top panel shows the GWAS results (black dots); blue line corresponds to $p = 1 \times 10 - 5$, orange line to $p = 5 \times 10 - 8$ (genome-wide significance). Bottom panel shows the TWAS results (green triangles facing upwards or downwards for a positive or negative z-score respectively (up- or down-regultion); only the transcripts with Bonferroni-adjusted $p < 0.1$ are labelled for clarity) for different transcripts (genes are represented by both gene expression and isoform expression); orange line corresponds to Bonferroni-adjusted p = 0.05. Each transcript that is Bonferroni-significant in the region is connected with lines to the SNPs that contribute to its transcriptomic imputation model; lines are grey when the SNPs have a $p > 1 \times 10 - 5$, blue when $p < 1 \times 10 - 5$ but $> 510 - 8$ and orange when $p < 5 \times 10 - 8$. The SNPs that are above the blue line and contribute to the transcriptomic imputation models of significant transcripts are labelled.

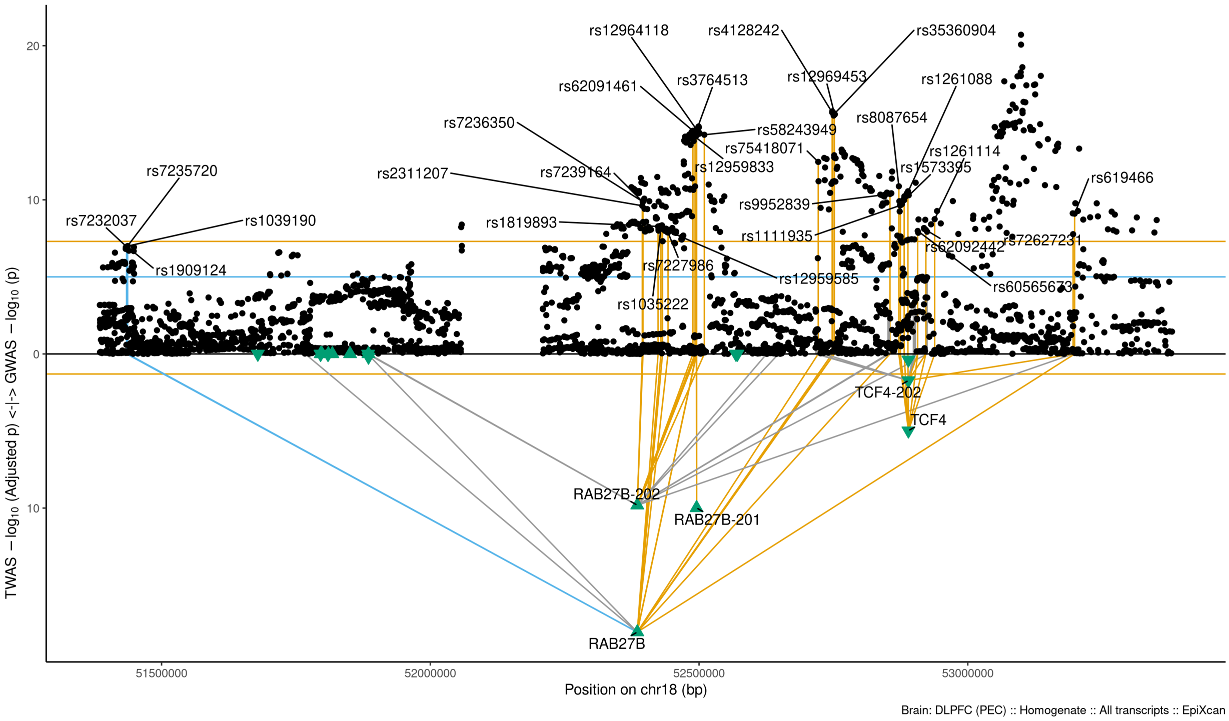

Figure S9-58: Regional Miami plots for RAB27B gene/transcript : Regional Miami plots for primary MD GWASs corresponding to the genomic region of the RAB27B transcript (1Mbp window from start site). Please also refer to Supplemental Table SXX for details. Top panel shows the GWAS results (black dots); blue line corresponds to $p = 1 \times 10 - 5$, orange line to $p = 5 \times 10 - 8$ (genome-wide significance). Bottom panel shows the TWAS results (green triangles facing upwards or downwards for a positive or negative z-score respectively (up- or down-regultion); only the transcripts with Bonferroni-adjusted $p < 0.1$ are labelled for clarity) for different transcripts (genes are represented by both gene expression and isoform expression); orange line corresponds to Bonferroni-adjusted p = 0.05. Each transcript that is Bonferroni-significant in the region is connected with lines to the SNPs that contribute to its transcriptomic imputation model; lines are grey when the SNPs have a $p > 1 \times 10 - 5$, blue when $p < 1 \times 10 - 5$ but $> 510 - 8$ and orange when $p < 5 \times 10 - 8$. The SNPs that are above the blue line and contribute to the transcriptomic imputation models of significant transcripts are labelled.

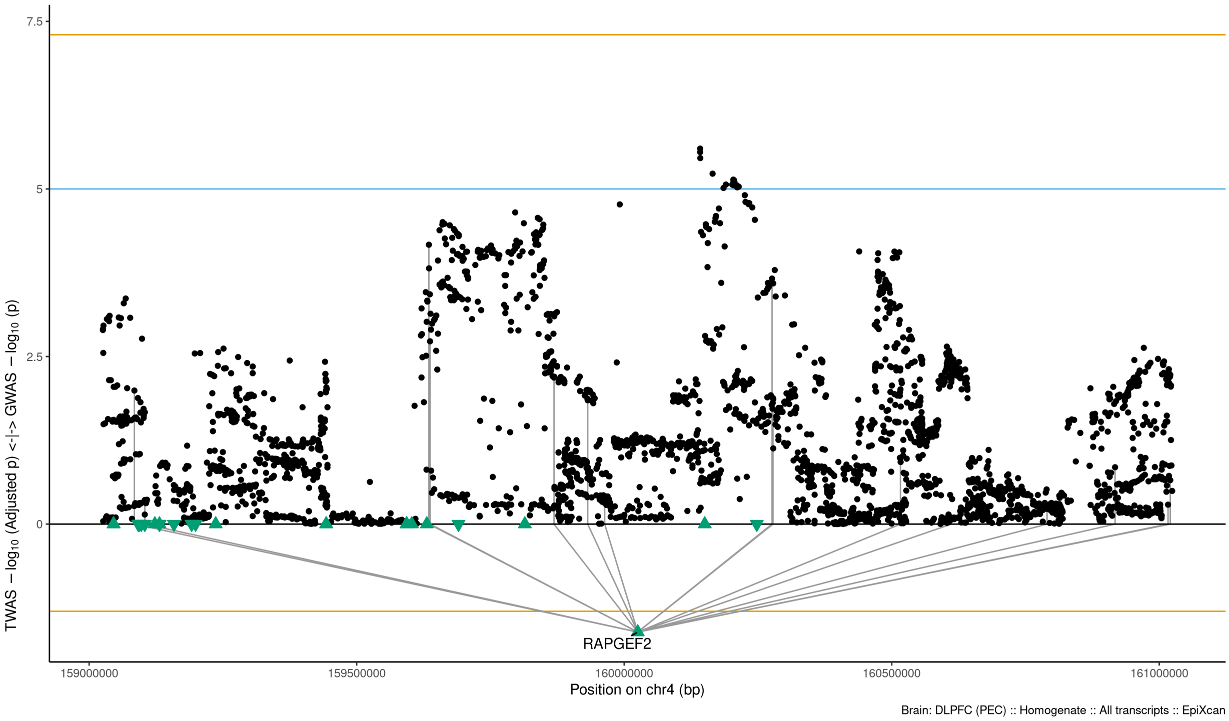

Figure S9-59: Regional Miami plots for RAPGEF2 gene/transcript : Regional Miami plots for primary MD GWASs corresponding to the genomic region of the RAPGEF2 transcript (1Mbp window from start site). Please also refer to Supplemental Table SXX for details. Top panel shows the GWAS results (black dots); blue line corresponds to $p = 1 \times 10 - 5$, orange line to $p = 5 \times 10 - 8$ (genome-wide significance). Bottom panel shows the TWAS results (green triangles facing upwards or downwards for a positive or negative z-score respectively (up- or down-regultion); only the transcripts with Bonferroni-adjusted $p < 0.1$ are labelled for clarity) for different transcripts (genes are represented by both gene expression and isoform expression); orange line corresponds to Bonferroni-adjusted p = 0.05. Each transcript that is Bonferroni-significant in the region is connected with lines to the SNPs that contribute to its transcriptomic imputation model; lines are grey when the SNPs have a $p > 1 \times 10 - 5$, blue when $p < 1 \times 10 - 5$ but $> 510 - 8$ and orange when $p < 5 \times 10 - 8$. The SNPs that are above the blue line and contribute to the transcriptomic imputation models of significant transcripts are labelled.

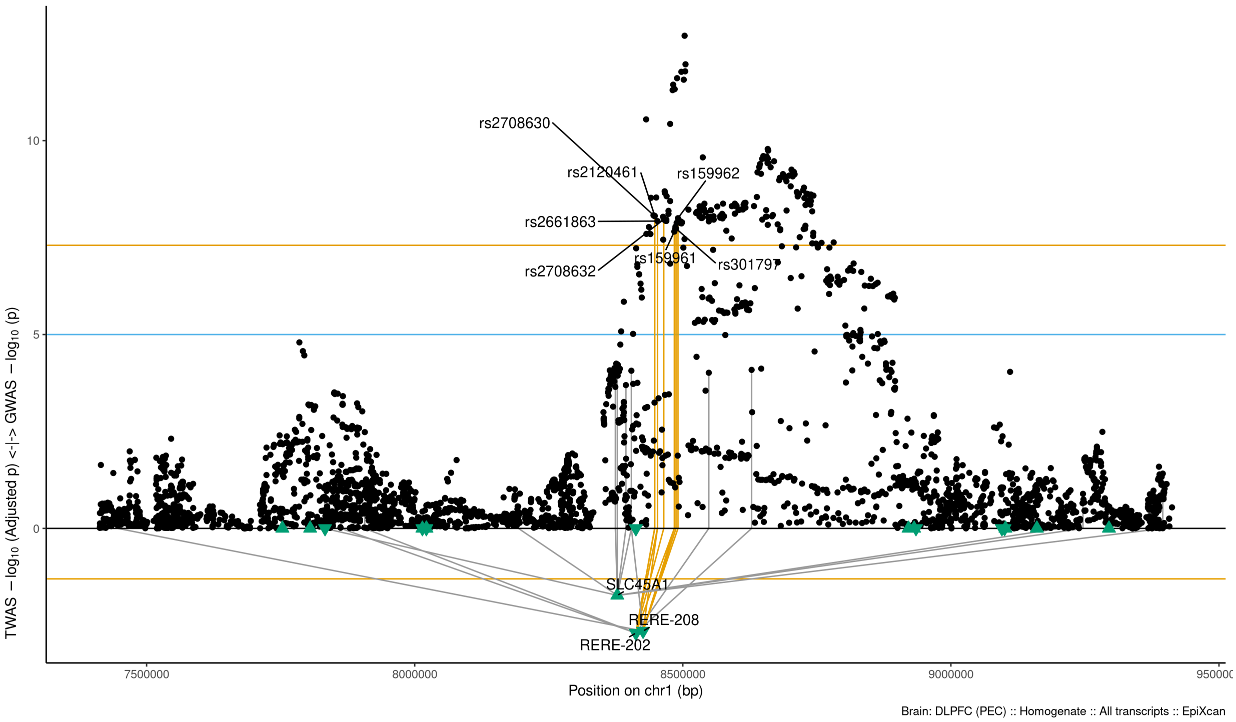

Figure S9-60: Regional Miami plots for RERE202 gene/transcript : Regional Miami plots for primary MD GWASs corresponding to the genomic region of the RERE202 transcript (1Mbp window from start site). Please also refer to Supplemental Table SXX for details. Top panel shows the GWAS results (black dots); blue line corresponds to $p = 1 \times 10 - 5$, orange line to $p = 5 \times 10 - 8$ (genome-wide significance). Bottom panel shows the TWAS results (green triangles facing upwards or downwards for a positive or negative z-score respectively (up- or down-regultion); only the transcripts with Bonferroni-adjusted $p < 0.1$ are labelled for clarity) for different transcripts (genes are represented by both gene expression and isoform expression); orange line corresponds to Bonferroni-adjusted p = 0.05. Each transcript that is Bonferroni-significant in the region is connected with lines to the SNPs that contribute to its transcriptomic imputation model; lines are grey when the SNPs have a $p > 1 \times 10 - 5$, blue when $p < 1 \times 10 - 5$ but $> 510 - 8$ and orange when $p < 5 \times 10 - 8$. The SNPs that are above the blue line and contribute to the transcriptomic imputation models of significant transcripts are labelled.

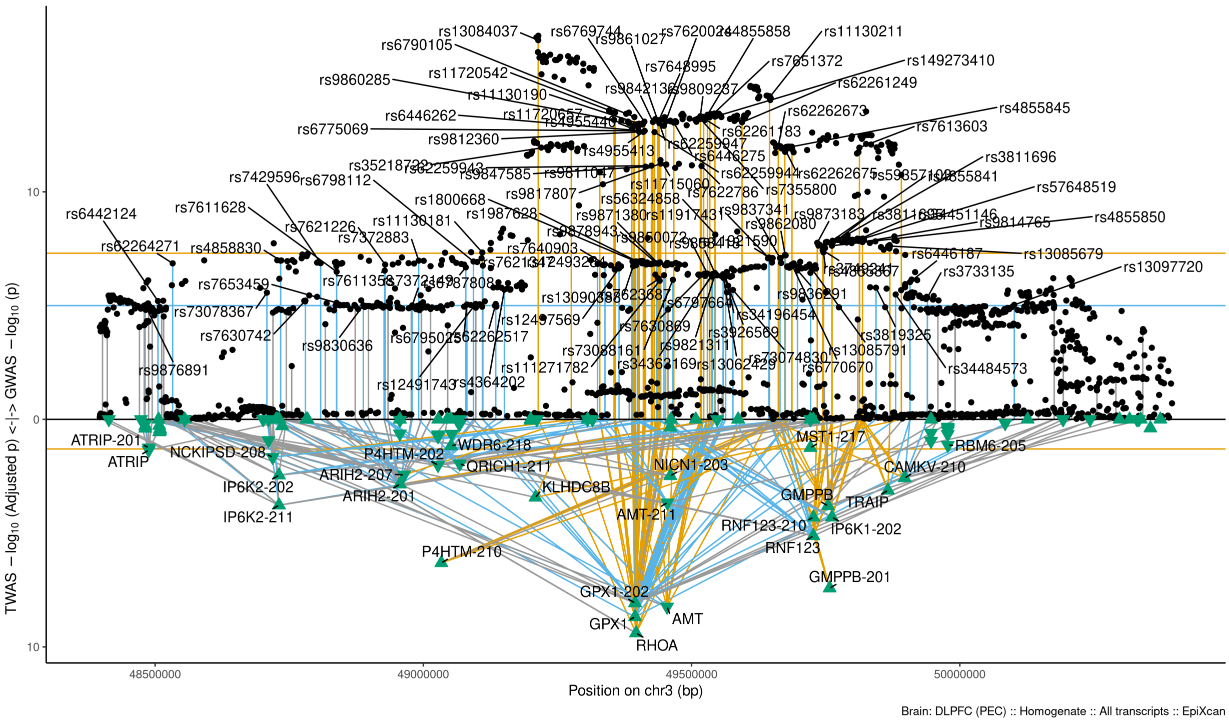

Figure S9-61: Regional Miami plots for RHOA gene/transcript : Regional Miami plots for primary MD GWASs corresponding to the genomic region of the RHOA transcript (1Mbp window from start site). Please also refer to Supplemental Table SXX for details. Top panel shows the GWAS results (black dots); blue line corresponds to $p = 1 \times 10 - 5$, orange line to $p = 5 \times 10 - 8$ (genome-wide significance). Bottom panel shows the TWAS results (green triangles facing upwards or downwards for a positive or negative z-score respectively (up- or down-regultion); only the transcripts with Bonferroni-adjusted $p < 0.1$ are labelled for clarity) for different transcripts (genes are represented by both gene expression and isoform expression); orange line corresponds to Bonferroni-adjusted p = 0.05. Each transcript that is Bonferroni-significant in the region is connected with lines to the SNPs that contribute to its transcriptomic imputation model; lines are grey when the SNPs have a $p > 1 \times 10 - 5$, blue when $p < 1 \times 10 - 5$ but $> 510 - 8$ and orange when $p < 5 \times 10 - 8$. The SNPs that are above the blue line and contribute to the transcriptomic imputation models of significant transcripts are labelled.

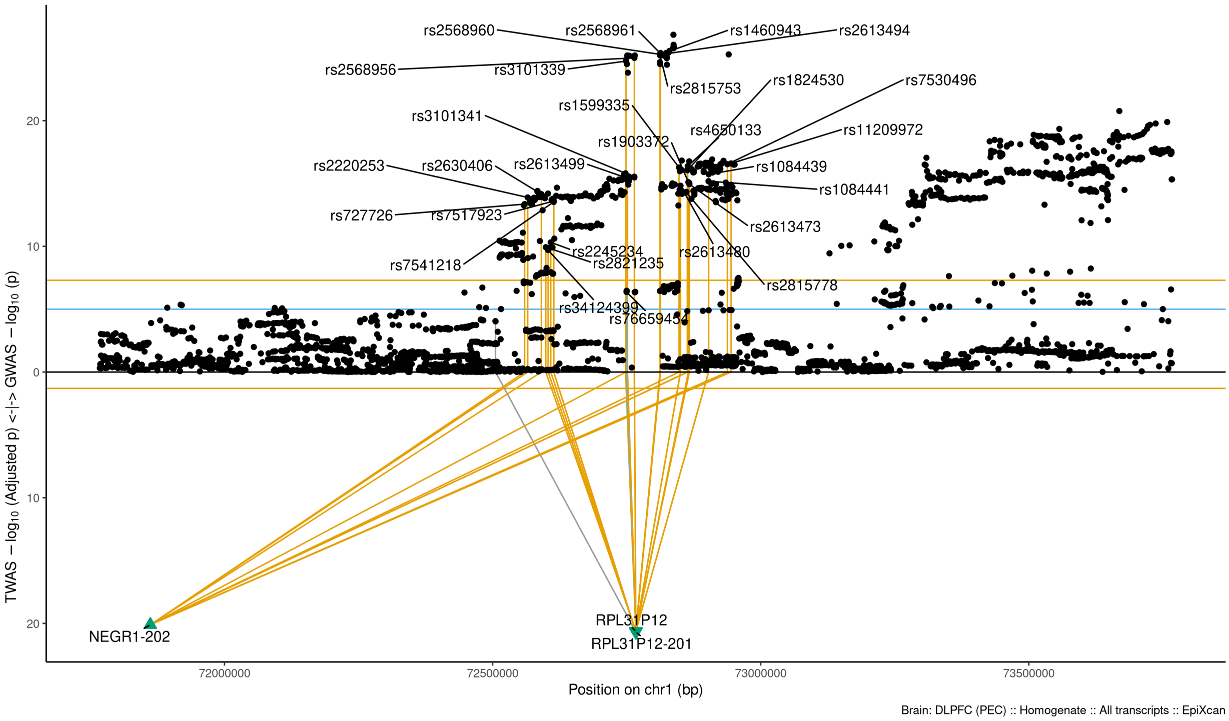

Figure S9-62: Regional Miami plots for RPL31P12201 gene/transcript : Regional Miami plots for primary MD GWASs corresponding to the genomic region of the RPL31P12201 transcript (1Mbp window from start site). Please also refer to Supplemental Table SXX for details. Top panel shows the GWAS results (black dots); blue line corresponds to $p = 1 \times 10 - 5$, orange line to $p = 5 \times 10 - 8$ (genome-wide significance). Bottom panel shows the TWAS results (green triangles facing upwards or downwards for a positive or negative z-score respectively (up- or down-regultion); only the transcripts with Bonferroni-adjusted $p < 0.1$ are labelled for clarity) for different transcripts (genes are represented by both gene expression and isoform expression); orange line corresponds to Bonferroni-adjusted p = 0.05. Each transcript that is Bonferroni-significant in the region is connected with lines to the SNPs that contribute to its transcriptomic imputation model; lines are grey when the SNPs have a $p > 1 \times 10 - 5$, blue when $p < 1 \times 10 - 5$ but $> 510 - 8$ and orange when $p < 5 \times 10 - 8$. The SNPs that are above the blue line and contribute to the transcriptomic imputation models of significant transcripts are labelled.

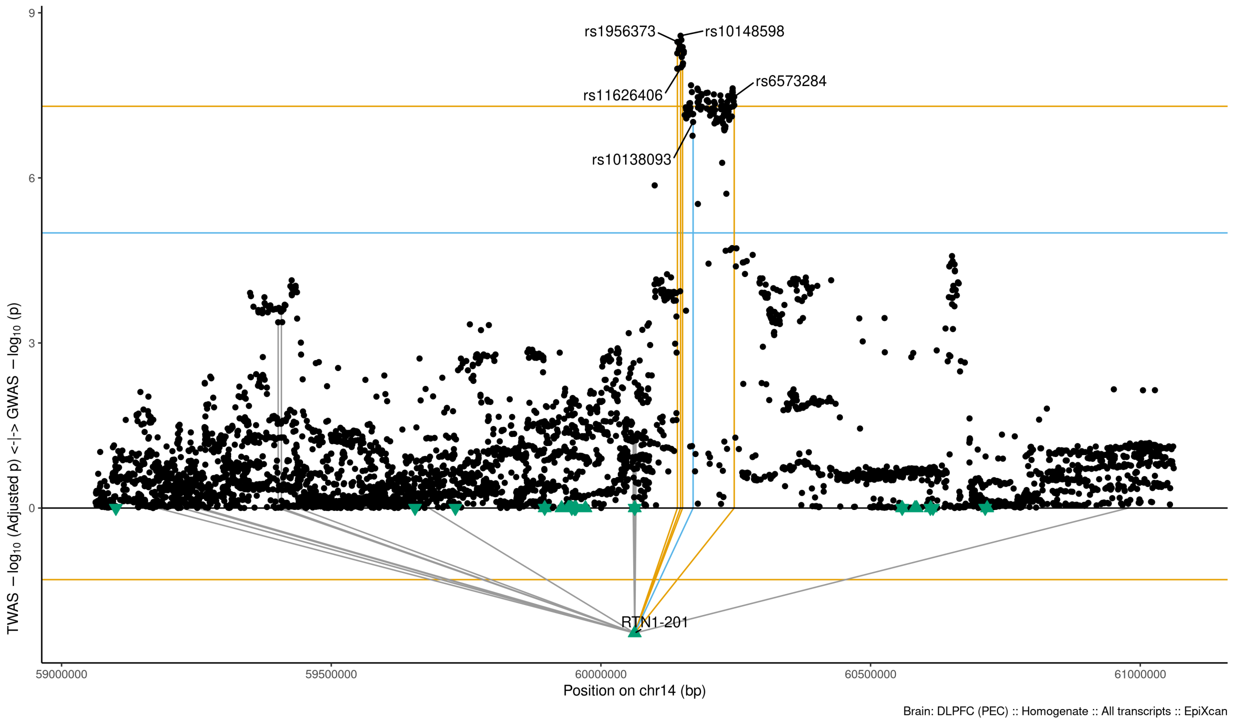

Figure S9-63: Regional Miami plots for RTN1201 gene/transcript : Regional Miami plots for primary MD GWASs corresponding to the genomic region of the RTN1201 transcript (1Mbp window from start site). Please also refer to Supplemental Table SXX for details. Top panel shows the GWAS results (black dots); blue line corresponds to $p = 1 \times 10 - 5$, orange line to $p = 5 \times 10 - 8$ (genome-wide significance). Bottom panel shows the TWAS results (green triangles facing upwards or downwards for a positive or negative z-score respectively (up- or down-regultion); only the transcripts with Bonferroni-adjusted $p < 0.1$ are labelled for clarity) for different transcripts (genes are represented by both gene expression and isoform expression); orange line corresponds to Bonferroni-adjusted p = 0.05. Each transcript that is Bonferroni-significant in the region is connected with lines to the SNPs that contribute to its transcriptomic imputation model; lines are grey when the SNPs have a $p > 1 \times 10 - 5$, blue when $p < 1 \times 10 - 5$ but $> 510 - 8$ and orange when $p < 5 \times 10 - 8$. The SNPs that are above the blue line and contribute to the transcriptomic imputation models of significant transcripts are labelled.

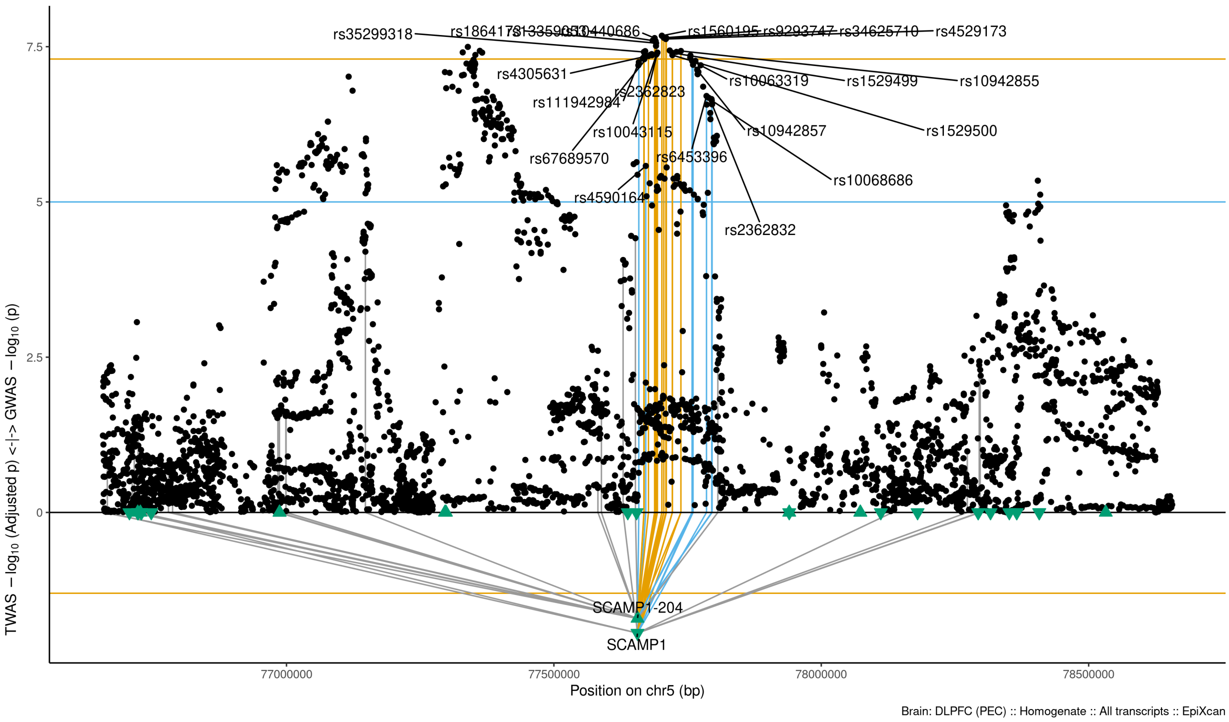

Figure S9-64: Regional Miami plots for SCAMP1 gene/transcript : Regional Miami plots for primary MD GWASs corresponding to the genomic region of the SCAMP1 transcript (1Mbp window from start site). Please also refer to Supplemental Table SXX for details. Top panel shows the GWAS results (black dots); blue line corresponds to $p = 1 \times 10 - 5$, orange line to $p = 5 \times 10 - 8$ (genome-wide significance). Bottom panel shows the TWAS results (green triangles facing upwards or downwards for a positive or negative z-score respectively (up- or down-regultion); only the transcripts with Bonferroni-adjusted $p < 0.1$ are labelled for clarity) for different transcripts (genes are represented by both gene expression and isoform expression); orange line corresponds to Bonferroni-adjusted p = 0.05. Each transcript that is Bonferroni-significant in the region is connected with lines to the SNPs that contribute to its transcriptomic imputation model; lines are grey when the SNPs have a $p > 1 \times 10 - 5$, blue when $p < 1 \times 10 - 5$ but $> 510 - 8$ and orange when $p < 5 \times 10 - 8$. The SNPs that are above the blue line and contribute to the transcriptomic imputation models of significant transcripts are labelled.

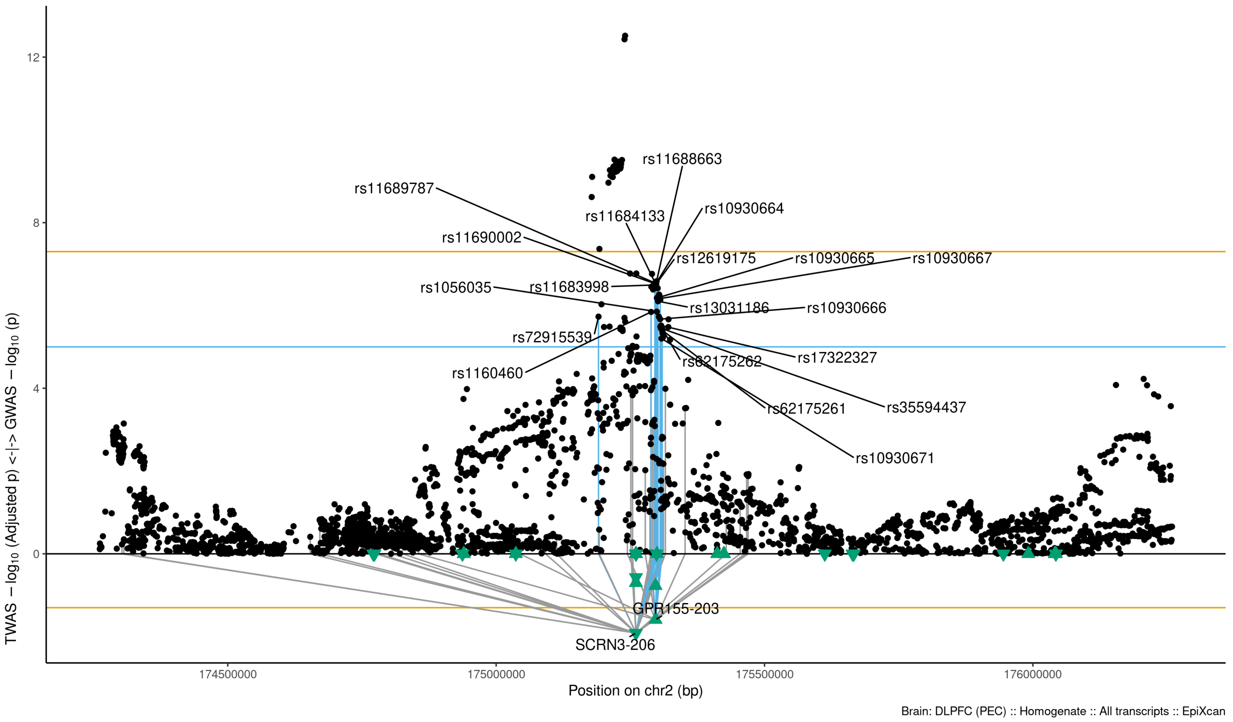

Figure S9-65: Regional Miami plots for SCRN3206 gene/transcript : Regional Miami plots for primary MD GWASs corresponding to the genomic region of the SCRN3206 transcript (1Mbp window from start site). Please also refer to Supplemental Table SXX for details. Top panel shows the GWAS results (black dots); blue line corresponds to $p = 1 \times 10 - 5$, orange line to $p = 5 \times 10 - 8$ (genome-wide significance). Bottom panel shows the TWAS results (green triangles facing upwards or downwards for a positive or negative z-score respectively (up- or down-regultion); only the transcripts with Bonferroni-adjusted $p < 0.1$ are labelled for clarity) for different transcripts (genes are represented by both gene expression and isoform expression); orange line corresponds to Bonferroni-adjusted p = 0.05. Each transcript that is Bonferroni-significant in the region is connected with lines to the SNPs that contribute to its transcriptomic imputation model; lines are grey when the SNPs have a $p > 1 \times 10 - 5$, blue when $p < 1 \times 10 - 5$ but $> 510 - 8$ and orange when $p < 5 \times 10 - 8$. The SNPs that are above the blue line and contribute to the transcriptomic imputation models of significant transcripts are labelled.

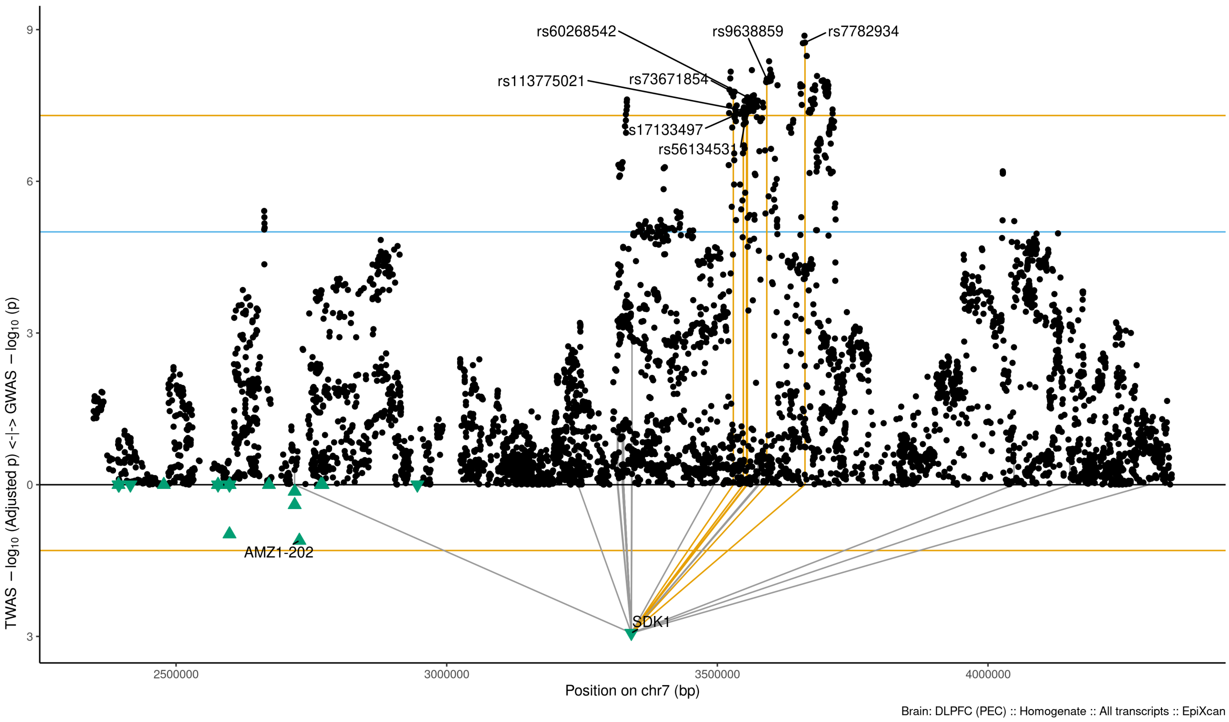

Figure S9-66: Regional Miami plots for SDK1 gene/transcript : Regional Miami plots for primary MD GWASs corresponding to the genomic region of the SDK1 transcript (1Mbp window from start site). Please also refer to Supplemental Table SXX for details. Top panel shows the GWAS results (black dots); blue line corresponds to $p = 1 \times 10 - 5$, orange line to $p = 5 \times 10 - 8$ (genome-wide significance). Bottom panel shows the TWAS results (green triangles facing upwards or downwards for a positive or negative z-score respectively (up- or down-regultion); only the transcripts with Bonferroni-adjusted $p < 0.1$ are labelled for clarity) for different transcripts (genes are represented by both gene expression and isoform expression); orange line corresponds to Bonferroni-adjusted p = 0.05. Each transcript that is Bonferroni-significant in the region is connected with lines to the SNPs that contribute to its transcriptomic imputation model; lines are grey when the SNPs have a $p > 1 \times 10 - 5$, blue when $p < 1 \times 10 - 5$ but $> 510 - 8$ and orange when $p < 5 \times 10 - 8$. The SNPs that are above the blue line and contribute to the transcriptomic imputation models of significant transcripts are labelled.

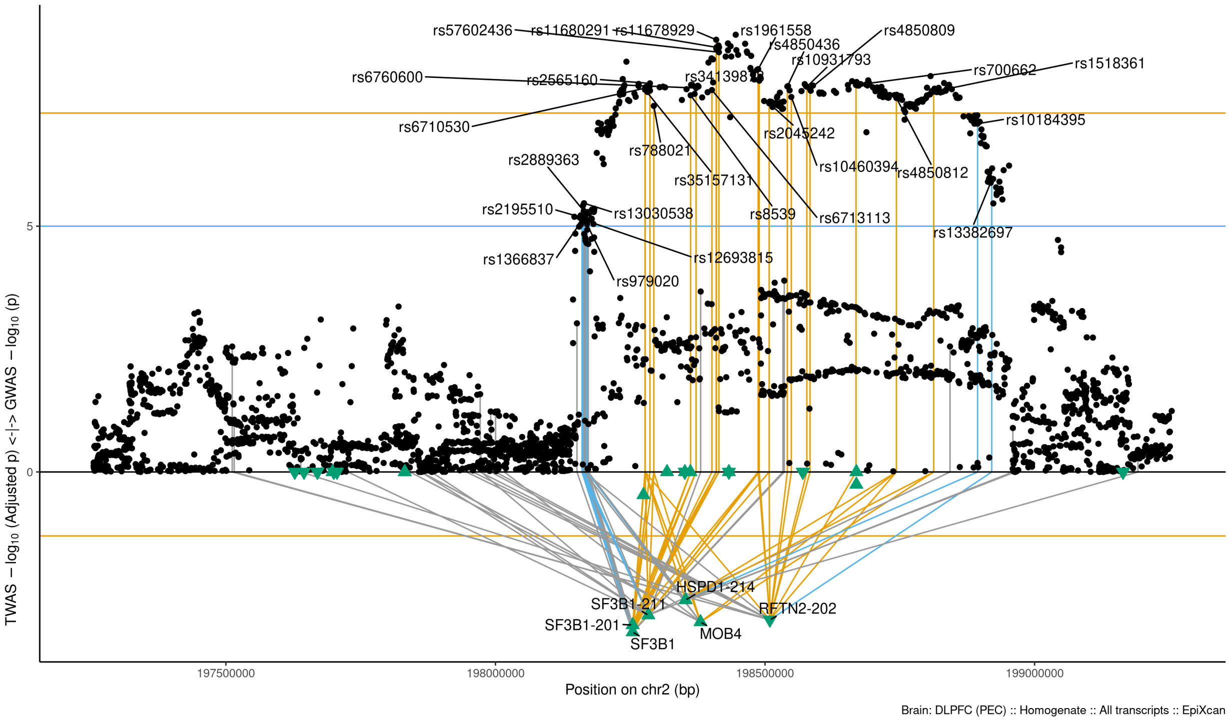

Figure S9-67: Regional Miami plots for SF3B1 gene/transcript : Regional Miami plots for primary MD GWASs corresponding to the genomic region of the SF3B1 transcript (1Mbp window from start site). Please also refer to Supplemental Table SXX for details. Top panel shows the GWAS results (black dots); blue line corresponds to $p = 1 \times 10 - 5$, orange line to $p = 5 \times 10 - 8$ (genome-wide significance). Bottom panel shows the TWAS results (green triangles facing upwards or downwards for a positive or negative z-score respectively (up- or down-regultion); only the transcripts with Bonferroni-adjusted $p < 0.1$ are labelled for clarity) for different transcripts (genes are represented by both gene expression and isoform expression); orange line corresponds to Bonferroni-adjusted p = 0.05. Each transcript that is Bonferroni-significant in the region is connected with lines to the SNPs that contribute to its transcriptomic imputation model; lines are grey when the SNPs have a $p > 1 \times 10 - 5$, blue when $p < 1 \times 10 - 5$ but $> 510 - 8$ and orange when $p < 5 \times 10 - 8$. The SNPs that are above the blue line and contribute to the transcriptomic imputation models of significant transcripts are labelled.

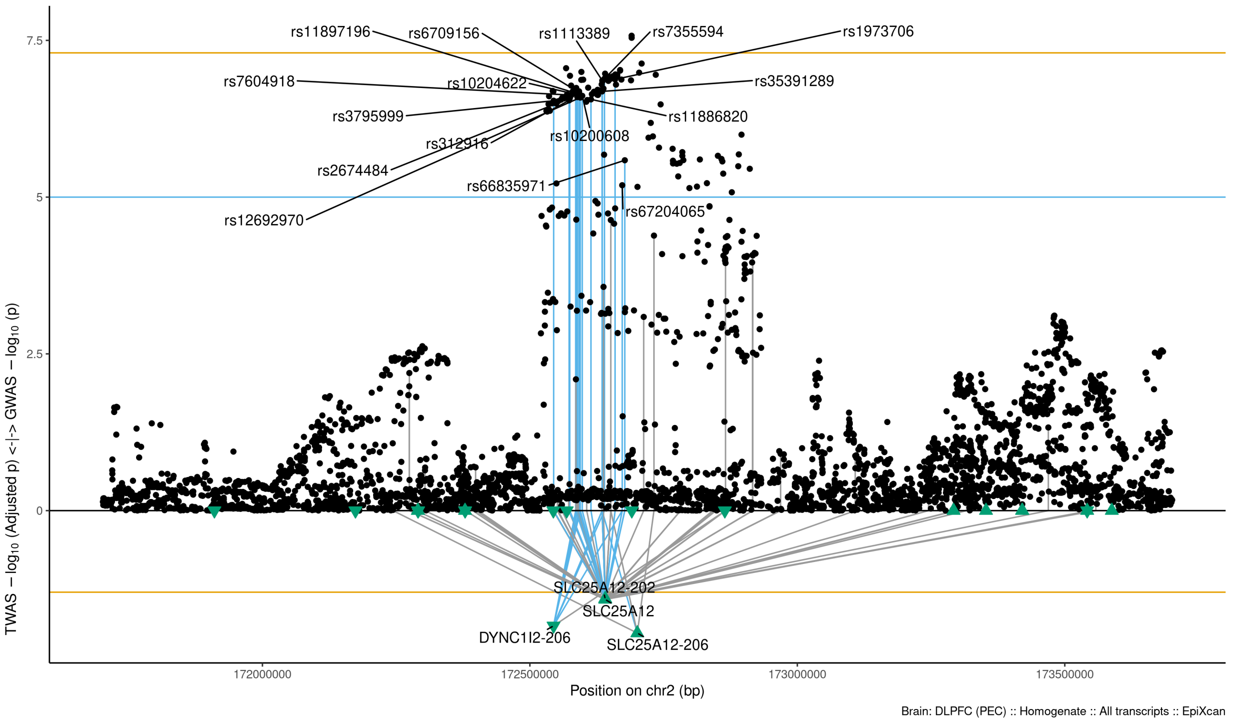

Figure S9-68: Regional Miami plots for SLC25A12206 gene/transcript : Regional Miami plots for primary MD GWASs corresponding to the genomic region of the SLC25A12206 transcript (1Mbp window from start site). Please also refer to Supplemental Table SXX for details. Top panel shows the GWAS results (black dots); blue line corresponds to $p = 1 \times 10 - 5$, orange line to $p = 5 \times 10 - 8$ (genome-wide significance). Bottom panel shows the TWAS results (green triangles facing upwards or downwards for a positive or negative z-score respectively (up- or down-regultion); only the transcripts with Bonferroni-adjusted $p < 0.1$ are labelled for clarity) for different transcripts (genes are represented by both gene expression and isoform expression); orange line corresponds to Bonferroni-adjusted p = 0.05. Each transcript that is Bonferroni-significant in the region is connected with lines to the SNPs that contribute to its transcriptomic imputation model; lines are grey when the SNPs have a $p > 1 \times 10 - 5$, blue when $p < 1 \times 10 - 5$ but $> 510 - 8$ and orange when $p < 5 \times 10 - 8$. The SNPs that are above the blue line and contribute to the transcriptomic imputation models of significant transcripts are labelled.

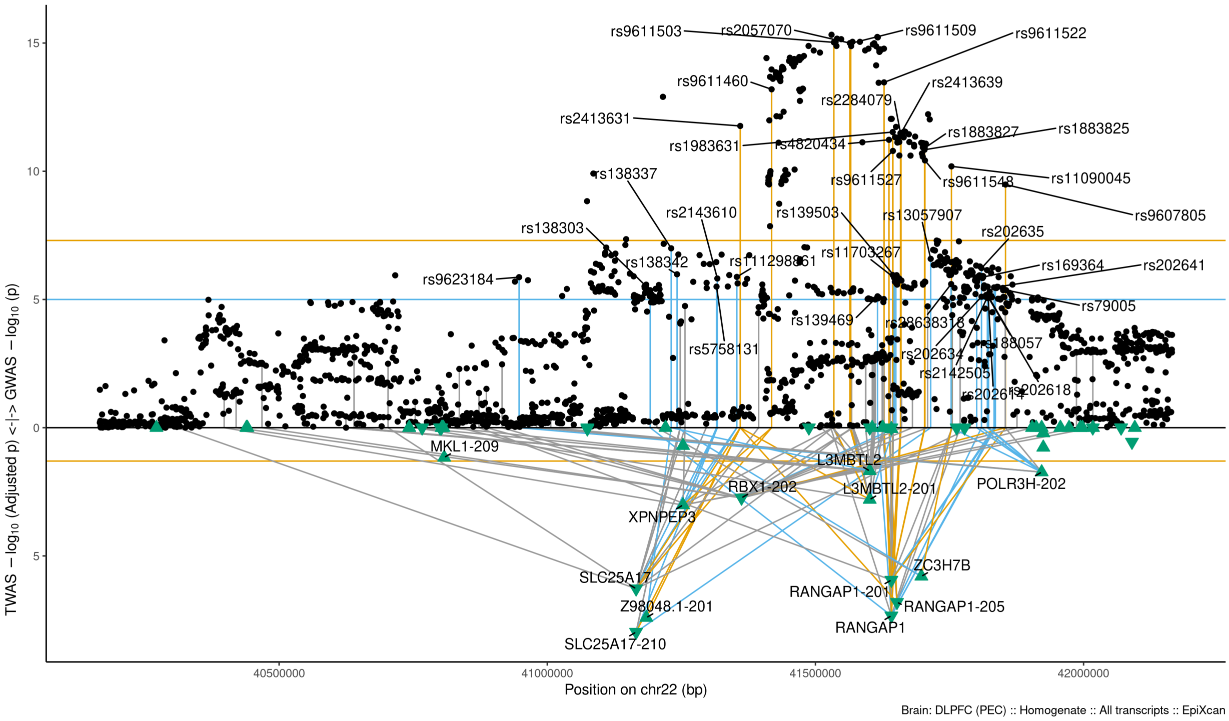

Figure S9-69: Regional Miami plots for SLC25A17210 gene/transcript : Regional Miami plots for primary MD GWASs corresponding to the genomic region of the SLC25A17210 transcript (1Mbp window from start site). Please also refer to Supplemental Table SXX for details. Top panel shows the GWAS results (black dots); blue line corresponds to $p = 1 \times 10 - 5$, orange line to $p = 5 \times 10 - 8$ (genome-wide significance). Bottom panel shows the TWAS results (green triangles facing upwards or downwards for a positive or negative z-score respectively (up- or down-regultion); only the transcripts with Bonferroni-adjusted $p < 0.1$ are labelled for clarity) for different transcripts (genes are represented by both gene expression and isoform expression); orange line corresponds to Bonferroni-adjusted p = 0.05. Each transcript that is Bonferroni-significant in the region is connected with lines to the SNPs that contribute to its transcriptomic imputation model; lines are grey when the SNPs have a $p > 1 \times 10 - 5$, blue when $p < 1 \times 10 - 5$ but $> 510 - 8$ and orange when $p < 5 \times 10 - 8$. The SNPs that are above the blue line and contribute to the transcriptomic imputation models of significant transcripts are labelled.

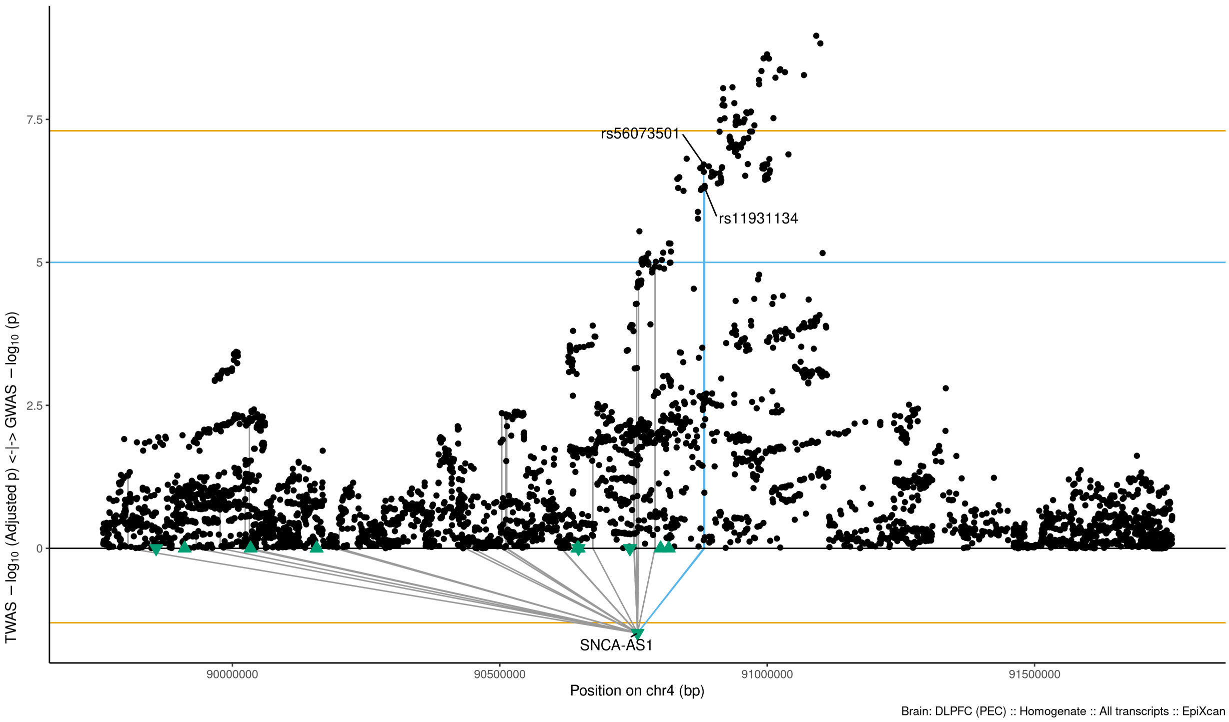

Figure S9-70: Regional Miami plots for SNCAAS1 gene/transcript : Regional Miami plots for primary MD GWASs corresponding to the genomic region of the SNCAAS1 transcript (1Mbp window from start site). Please also refer to Supplemental Table SXX for details. Top panel shows the GWAS results (black dots); blue line corresponds to $p = 1 \times 10 - 5$, orange line to $p = 5 \times 10 - 8$ (genome-wide significance). Bottom panel shows the TWAS results (green triangles facing upwards or downwards for a positive or negative z-score respectively (up- or down-regultion); only the transcripts with Bonferroni-adjusted $p < 0.1$ are labelled for clarity) for different transcripts (genes are represented by both gene expression and isoform expression); orange line corresponds to Bonferroni-adjusted p = 0.05. Each transcript that is Bonferroni-significant in the region is connected with lines to the SNPs that contribute to its transcriptomic imputation model; lines are grey when the SNPs have a $p > 1 \times 10 - 5$, blue when $p < 1 \times 10 - 5$ but $> 510 - 8$ and orange when $p < 5 \times 10 - 8$. The SNPs that are above the blue line and contribute to the transcriptomic imputation models of significant transcripts are labelled.

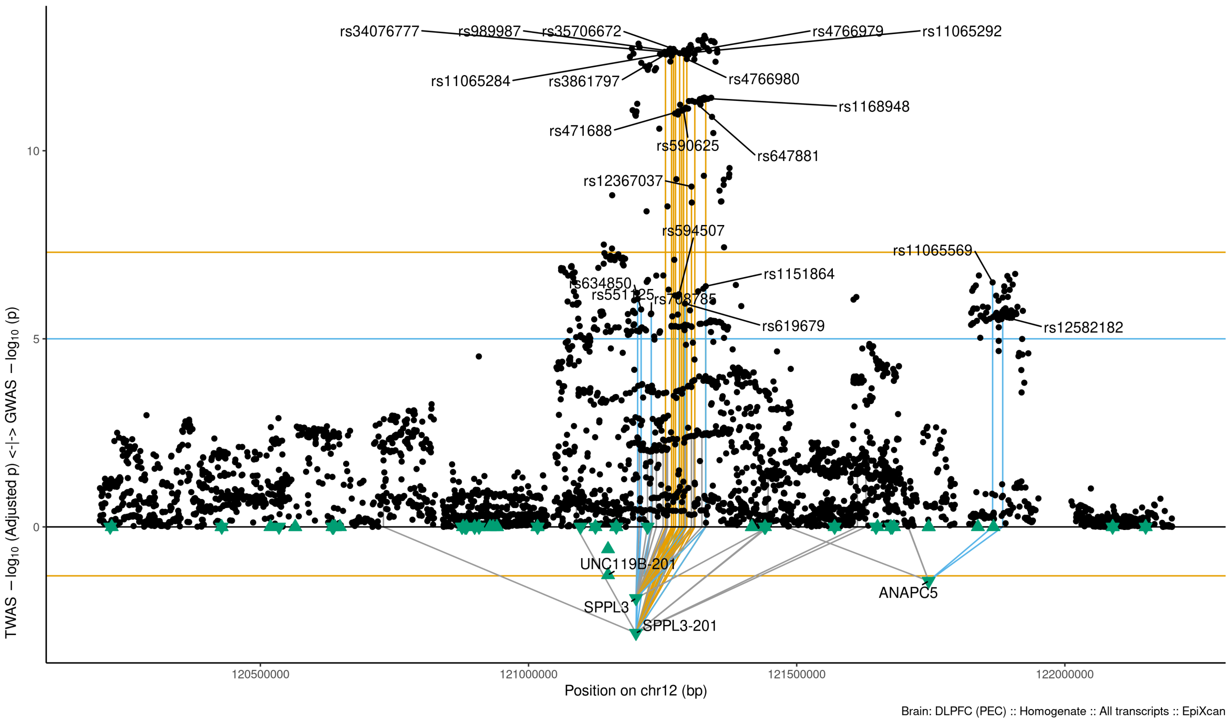

Figure S9-71: Regional Miami plots for SPPL3201 gene/transcript : Regional Miami plots for primary MD GWASs corresponding to the genomic region of the SPPL3201 transcript (1Mbp window from start site). Please also refer to Supplemental Table SXX for details. Top panel shows the GWAS results (black dots); blue line corresponds to $p = 1 \times 10 - 5$, orange line to $p = 5 \times 10 - 8$ (genome-wide significance). Bottom panel shows the TWAS results (green triangles facing upwards or downwards for a positive or negative z-score respectively (up- or down-regultion); only the transcripts with Bonferroni-adjusted $p < 0.1$ are labelled for clarity) for different transcripts (genes are represented by both gene expression and isoform expression); orange line corresponds to Bonferroni-adjusted p = 0.05. Each transcript that is Bonferroni-significant in the region is connected with lines to the SNPs that contribute to its transcriptomic imputation model; lines are grey when the SNPs have a $p > 1 \times 10 - 5$, blue when $p < 1 \times 10 - 5$ but $> 510 - 8$ and orange when $p < 5 \times 10 - 8$. The SNPs that are above the blue line and contribute to the transcriptomic imputation models of significant transcripts are labelled.

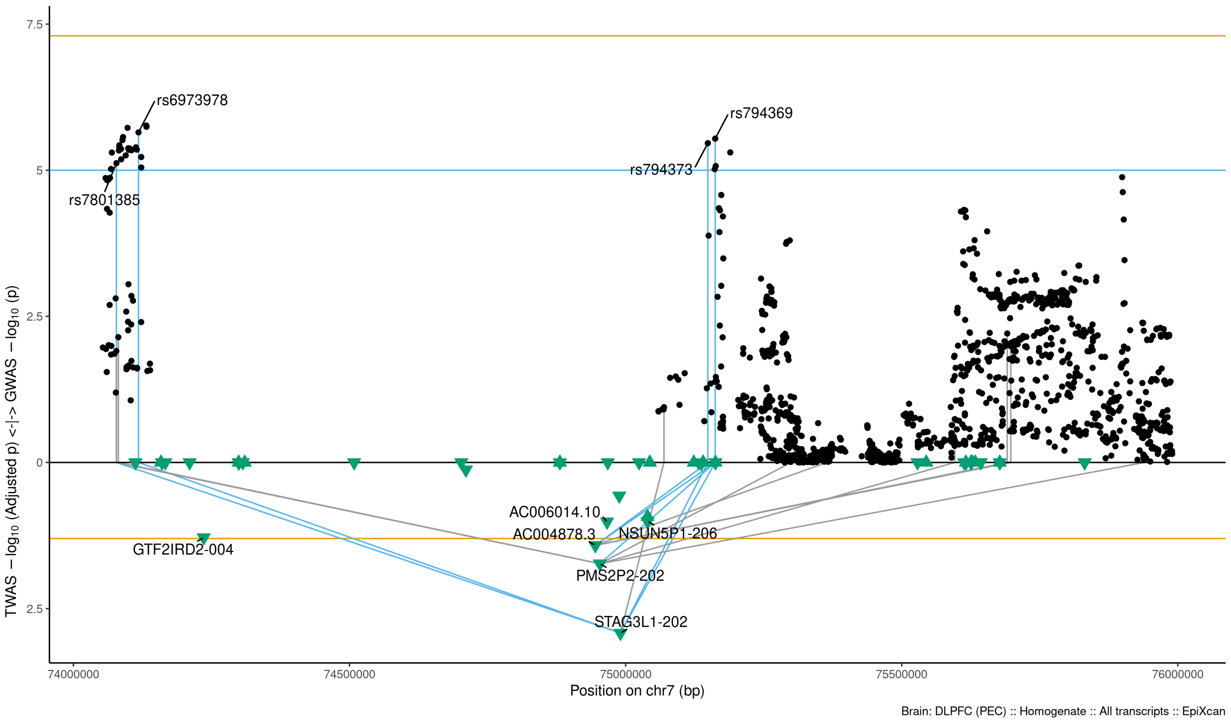

Figure S9-72: Regional Miami plots for STAG3L1202 gene/transcript : Regional Miami plots for primary MD GWASs corresponding to the genomic region of the STAG3L1202 transcript (1Mbp window from start site). Please also refer to Supplemental Table SXX for details. Top panel shows the GWAS results (black dots); blue line corresponds to $p = 1 \times 10 - 5$, orange line to $p = 5 \times 10 - 8$ (genome-wide significance). Bottom panel shows the TWAS results (green triangles facing upwards or downwards for a positive or negative z-score respectively (up- or down-regultion); only the transcripts with Bonferroni-adjusted $p < 0.1$ are labelled for clarity) for different transcripts (genes are represented by both gene expression and isoform expression); orange line corresponds to Bonferroni-adjusted p = 0.05. Each transcript that is Bonferroni-significant in the region is connected with lines to the SNPs that contribute to its transcriptomic imputation model; lines are grey when the SNPs have a $p > 1 \times 10 - 5$, blue when $p < 1 \times 10 - 5$ but $> 510 - 8$ and orange when $p < 5 \times 10 - 8$. The SNPs that are above the blue line and contribute to the transcriptomic imputation models of significant transcripts are labelled.

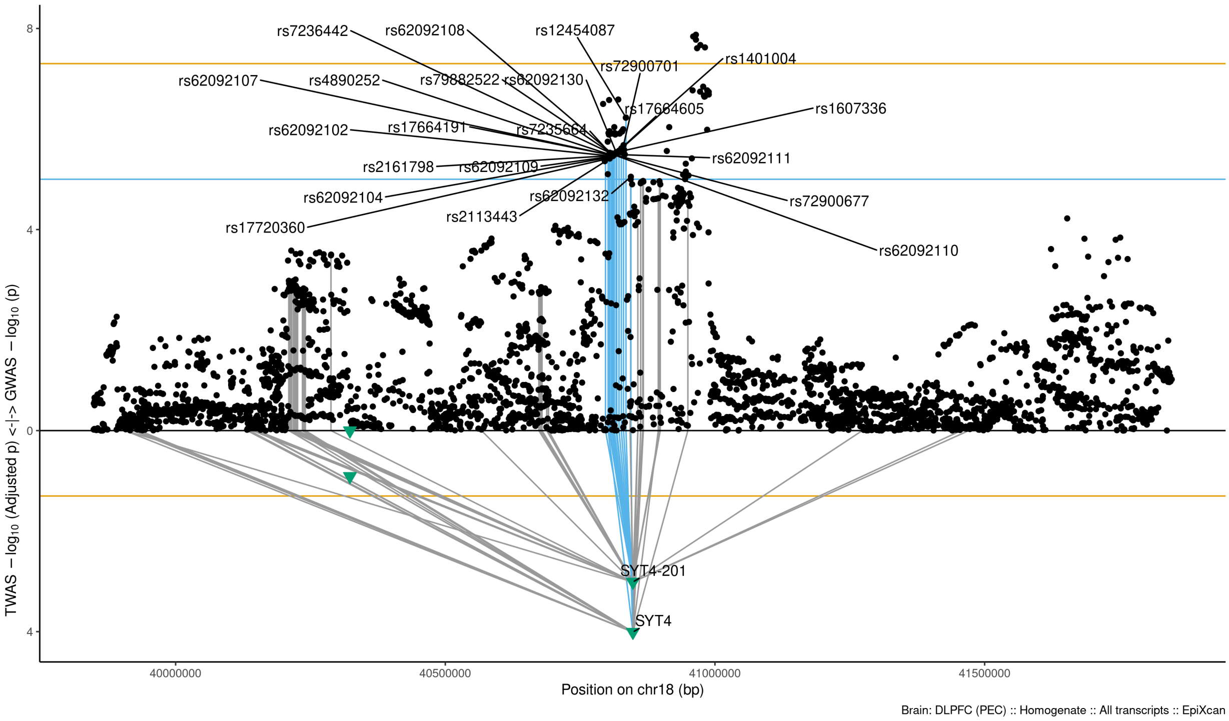

Figure S9-73: Regional Miami plots for SYT4 gene/transcript : Regional Miami plots for primary MD GWASs corresponding to the genomic region of the SYT4 transcript (1Mbp window from start site). Please also refer to Supplemental Table SXX for details. Top panel shows the GWAS results (black dots); blue line corresponds to $p = 1 \times 10 - 5$, orange line to $p = 5 \times 10 - 8$ (genome-wide significance). Bottom panel shows the TWAS results (green triangles facing upwards or downwards for a positive or negative z-score respectively (up- or down-regultion); only the transcripts with Bonferroni-adjusted $p < 0.1$ are labelled for clarity) for different transcripts (genes are represented by both gene expression and isoform expression); orange line corresponds to Bonferroni-adjusted $\mathrm{p} = 0.05$. Each transcript that is Bonferroni-significant in the region is connected with lines to the SNPs that contribute to its transcriptomic imputation model; lines are grey when the SNPs have a $p > 1 \times 10 - 5$, blue when $p < 1 \times 10 - 5$ but $> 510 - 8$ and orange when $p < 5 \times 10 - 8$. The SNPs that are above the blue line and contribute to the transcriptomic imputation models of significant transcripts are labelled.

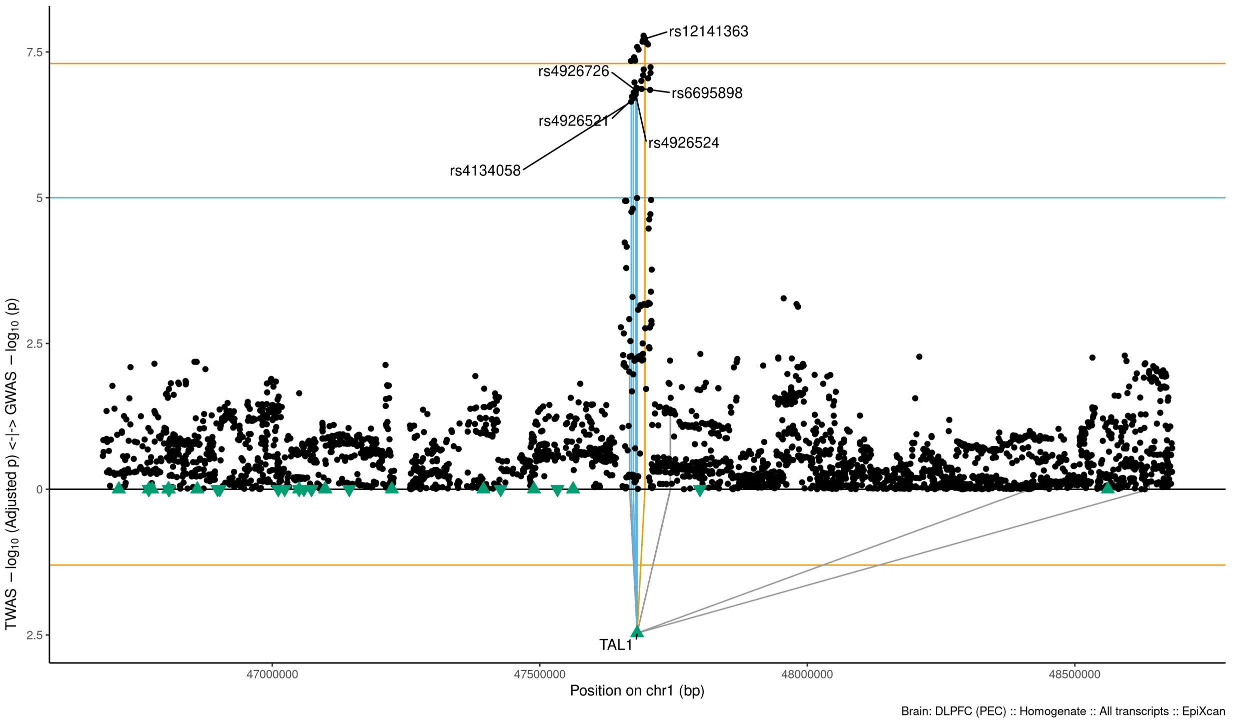

Figure S9-74: Regional Miami plots for TAL1 gene/transcript : Regional Miami plots for primary MD GWASs corresponding to the genomic region of the TAL1 transcript (1Mbp window from start site). Please also refer to Supplemental Table SXX for details. Top panel shows the GWAS results (black dots); blue line corresponds to $p = 1 \times 10 - 5$, orange line to $p = 5 \times 10 - 8$ (genome-wide significance). Bottom panel shows the TWAS results (green triangles facing upwards or downwards for a positive or negative z-score respectively (up- or down-regultion); only the transcripts with Bonferroni-adjusted $p < 0.1$ are labelled for clarity) for different transcripts (genes are represented by both gene expression and isoform expression); orange line corresponds to Bonferroni-adjusted p = 0.05. Each transcript that is Bonferroni-significant in the region is connected with lines to the SNPs that contribute to its transcriptomic imputation model; lines are grey when the SNPs have a $p > 1 \times 10 - 5$, blue when $p < 1 \times 10 - 5$ but $> 510 - 8$ and orange when $p < 5 \times 10 - 8$. The SNPs that are above the blue line and contribute to the transcriptomic imputation models of significant transcripts are labelled.

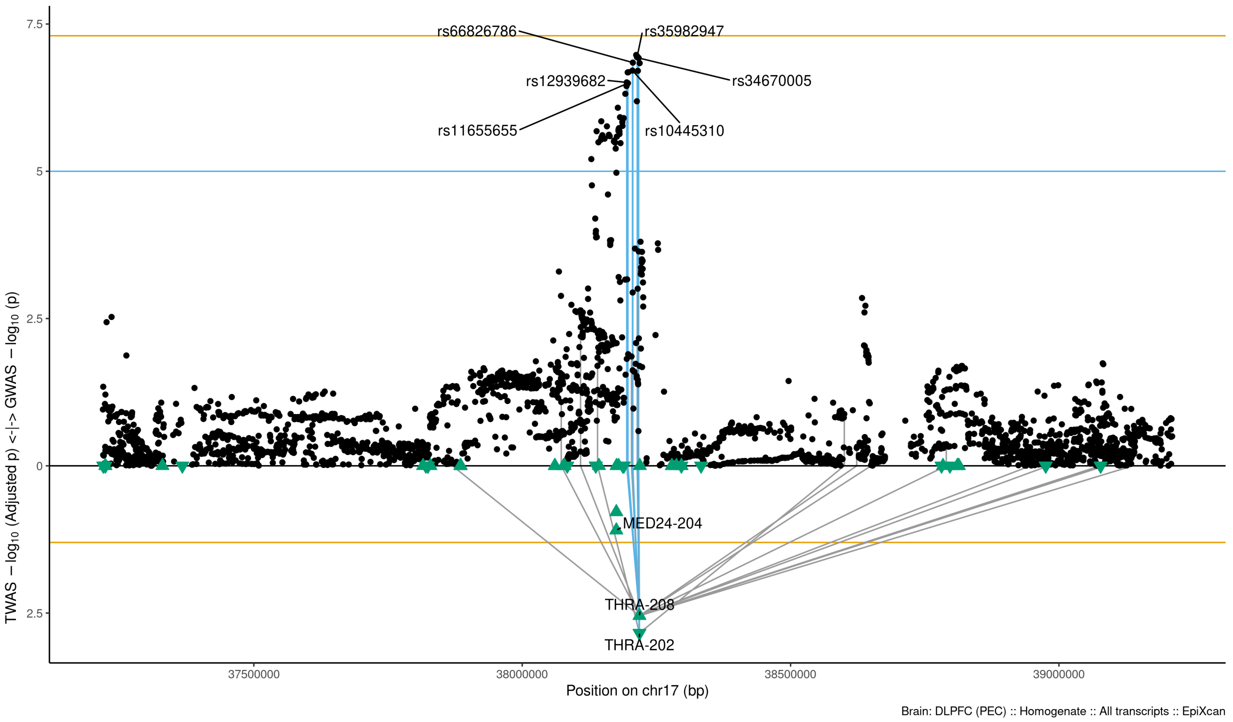

Figure S9-75: Regional Miami plots for THRA202 gene/transcript : Regional Miami plots for primary MD GWASs corresponding to the genomic region of the THRA202 transcript (1Mbp window from start site). Please also refer to Supplemental Table SXX for details. Top panel shows the GWAS results (black dots); blue line corresponds to $p = 1 \times 10 - 5$, orange line to $p = 5 \times 10 - 8$ (genome-wide significance). Bottom panel shows the TWAS results (green triangles facing upwards or downwards for a positive or negative z-score respectively (up- or down-regultion); only the transcripts with Bonferroni-adjusted $p < 0.1$ are labelled for clarity) for different transcripts (genes are represented by both gene expression and isoform expression); orange line corresponds to Bonferroni-adjusted p = 0.05. Each transcript that is Bonferroni-significant in the region is connected with lines to the SNPs that contribute to its transcriptomic imputation model; lines are grey when the SNPs have a $p > 1 \times 10 - 5$, blue when $p < 1 \times 10 - 5$ but $> 510 - 8$ and orange when $p < 5 \times 10 - 8$. The SNPs that are above the blue line and contribute to the transcriptomic imputation models of significant transcripts are labelled.

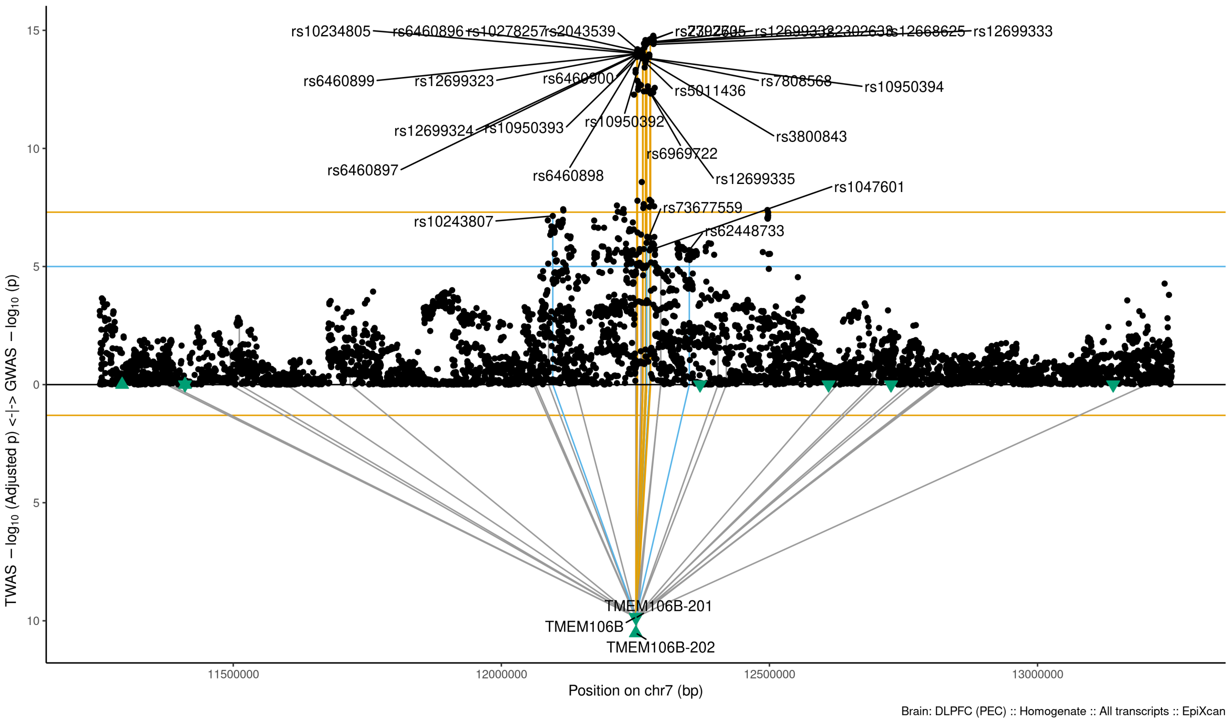

Figure S9-76: Regional Miami plots for TMEM106B202 gene/transcript : Regional Miami plots for primary MD GWASs corresponding to the genomic region of the TMEM106B202 transcript (1Mbp window from start site). Please also refer to Supplemental Table SXX for details. Top panel shows the GWAS results (black dots); blue line corresponds to $p = 1 \times 10 - 5$, orange line to $p = 5 \times 10 - 8$ (genome-wide significance). Bottom panel shows the TWAS results (green triangles facing upwards or downwards for a positive or negative z-score respectively (up- or down-regultion); only the transcripts with Bonferroni-adjusted $p < 0.1$ are labelled for clarity) for different transcripts (genes are represented by both gene expression and isoform expression); orange line corresponds to Bonferroni-adjusted p = 0.05. Each transcript that is Bonferroni-significant in the region is connected with lines to the SNPs that contribute to its transcriptomic imputation model; lines are grey when the SNPs have a $p > 1 \times 10 - 5$, blue when $p < 1 \times 10 - 5$ but $> 510 - 8$ and orange when $p < 5 \times 10 - 8$. The SNPs that are above the blue line and contribute to the transcriptomic imputation models of significant transcripts are labelled.

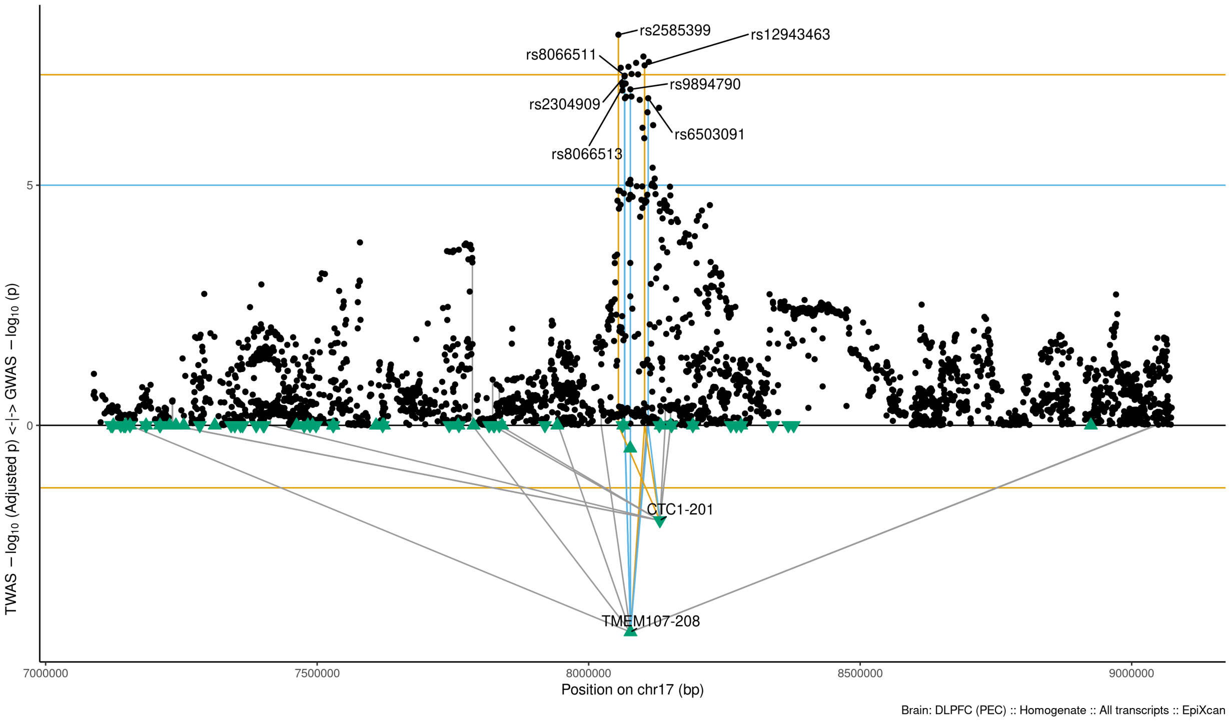

Figure S9-77: Regional Miami plots for TMEM107208 gene/transcript : Regional Miami plots for primary MD GWASs corresponding to the genomic region of the TMEM107208 transcript (1Mbp window from start site). Please also refer to Supplemental Table SXX for details. Top panel shows the GWAS results (black dots); blue line corresponds to $p = 1 \times 10 - 5$, orange line to $p = 5 \times 10 - 8$ (genome-wide significance). Bottom panel shows the TWAS results (green triangles facing upwards or downwards for a positive or negative z-score respectively (up- or down-regultion); only the transcripts with Bonferroni-adjusted $p < 0.1$ are labelled for clarity) for different transcripts (genes are represented by both gene expression and isoform expression); orange line corresponds to Bonferroni-adjusted p = 0.05. Each transcript that is Bonferroni-significant in the region is connected with lines to the SNPs that contribute to its transcriptomic imputation model; lines are grey when the SNPs have a $p > 1 \times 10 - 5$, blue when $p < 1 \times 10 - 5$ but $> 510 - 8$ and orange when $p < 5 \times 10 - 8$. The SNPs that are above the blue line and contribute to the transcriptomic imputation models of significant transcripts are labelled.

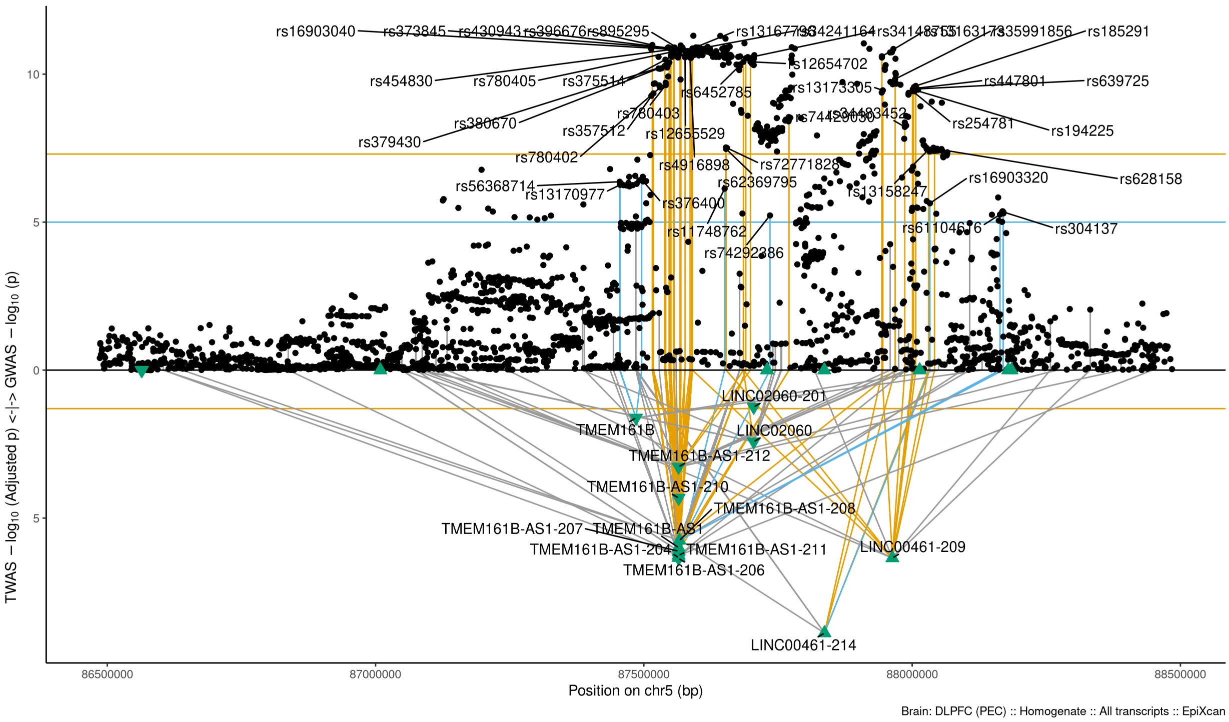

Figure S9-78: Regional Miami plots for TMEM161B gene/transcript : Regional Miami plots for primary MD GWASs corresponding to the genomic region of the TMEM161B transcript (1Mbp window from start site). Please also refer to Supplemental Table SXX for details. Top panel shows the GWAS results (black dots); blue line corresponds to $p = 1 \times 10 - 5$, orange line to $p = 5 \times 10 - 8$ (genome-wide significance). Bottom panel shows the TWAS results (green triangles facing upwards or downwards for a positive or negative z-score respectively (up- or down-regultion); only the transcripts with Bonferroni-adjusted $p < 0.1$ are labelled for clarity) for different transcripts (genes are represented by both gene expression and isoform expression); orange line corresponds to Bonferroni-adjusted p = 0.05. Each transcript that is Bonferroni-significant in the region is connected with lines to the SNPs that contribute to its transcriptomic imputation model; lines are grey when the SNPs have a $p > 1 \times 10 - 5$, blue when $p < 1 \times 10 - 5$ but $> 510 - 8$ and orange when $p < 5 \times 10 - 8$. The SNPs that are above the blue line and contribute to the transcriptomic imputation models of significant transcripts are labelled.

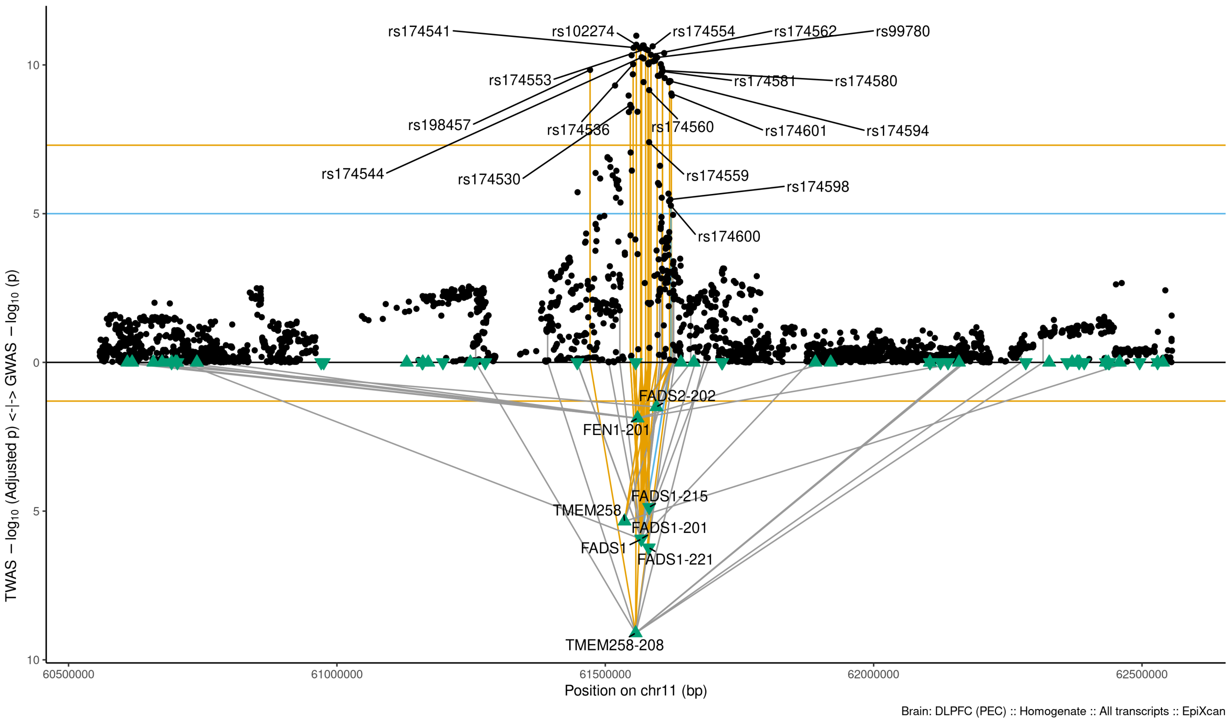

Figure S9-79: Regional Miami plots for TMEM258208 gene/transcript : Regional Miami plots for primary MD GWASs corresponding to the genomic region of the TMEM258208 transcript (1Mbp window from start site). Please also refer to Supplemental Table SXX for details. Top panel shows the GWAS results (black dots); blue line corresponds to $p = 1 \times 10 - 5$, orange line to $p = 5 \times 10 - 8$ (genome-wide significance). Bottom panel shows the TWAS results (green triangles facing upwards or downwards for a positive or negative z-score respectively (up- or down-regultion); only the transcripts with Bonferroni-adjusted $p < 0.1$ are labelled for clarity) for different transcripts (genes are represented by both gene expression and isoform expression); orange line corresponds to Bonferroni-adjusted p = 0.05. Each transcript that is Bonferroni-significant in the region is connected with lines to the SNPs that contribute to its transcriptomic imputation model; lines are grey when the SNPs have a $p > 1 \times 10 - 5$, blue when $p < 1 \times 10 - 5$ but $> 510 - 8$ and orange when $p < 5 \times 10 - 8$. The SNPs that are above the blue line and contribute to the transcriptomic imputation models of significant transcripts are labelled.

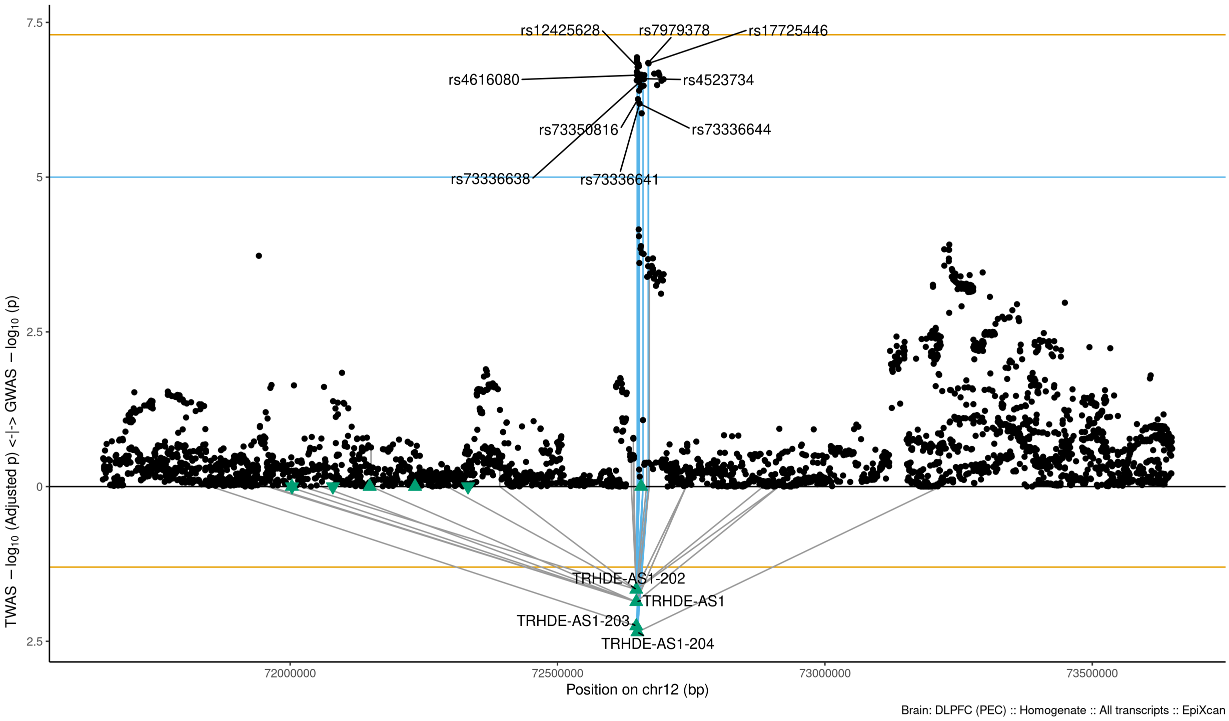

Figure S9-80: Regional Miami plots for TRHDEAS1204 gene/transcript : Regional Miami plots for primary MD GWASs corresponding to the genomic region of the TRHDEAS1204 transcript (1Mbp window from start site). Please also refer to Supplemental Table SXX for details. Top panel shows the GWAS results (black dots); blue line corresponds to $p = 1 \times 10 - 5$, orange line to $p = 5 \times 10 - 8$ (genome-wide significance). Bottom panel shows the TWAS results (green triangles facing upwards or downwards for a positive or negative z-score respectively (up- or down-regultion); only the transcripts with Bonferroni-adjusted $p < 0.1$ are labelled for clarity) for different transcripts (genes are represented by both gene expression and isoform expression); orange line corresponds to Bonferroni-adjusted p = 0.05. Each transcript that is Bonferroni-significant in the region is connected with lines to the SNPs that contribute to its transcriptomic imputation model; lines are grey when the SNPs have a $p > 1 \times 10 - 5$, blue when $p < 1 \times 10 - 5$ but $> 510 - 8$ and orange when $p < 5 \times 10 - 8$. The SNPs that are above the blue line and contribute to the transcriptomic imputation models of significant transcripts are labelled.

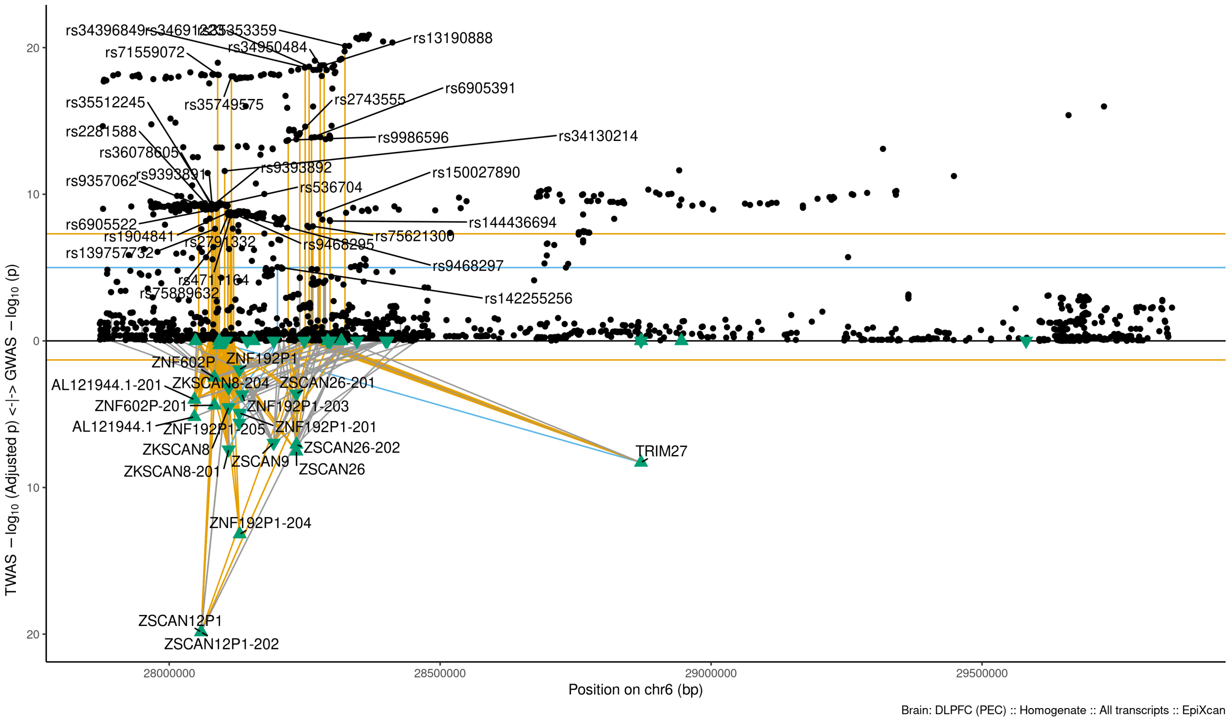

Figure S9-81: Regional Miami plots for TRIM27 gene/transcript : Regional Miami plots for primary MD GWASs corresponding to the genomic region of the TRIM27 transcript (1Mbp window from start site). Please also refer to Supplemental Table SXX for details. Top panel shows the GWAS results (black dots); blue line corresponds to $p = 1 \times 10 - 5$, orange line to $p = 5 \times 10 - 8$ (genome-wide significance). Bottom panel shows the TWAS results (green triangles facing upwards or downwards for a positive or negative z-score respectively (up- or down-regultion); only the transcripts with Bonferroni-adjusted $p < 0.1$ are labelled for clarity) for different transcripts (genes are represented by both gene expression and isoform expression); orange line corresponds to Bonferroni-adjusted p = 0.05. Each transcript that is Bonferroni-significant in the region is connected with lines to the SNPs that contribute to its transcriptomic imputation model; lines are grey when the SNPs have a $p > 1 \times 10 - 5$, blue when $p < 1 \times 10 - 5$ but $> 510 - 8$ and orange when $p < 5 \times 10 - 8$. The SNPs that are above the blue line and contribute to the transcriptomic imputation models of significant transcripts are labelled.

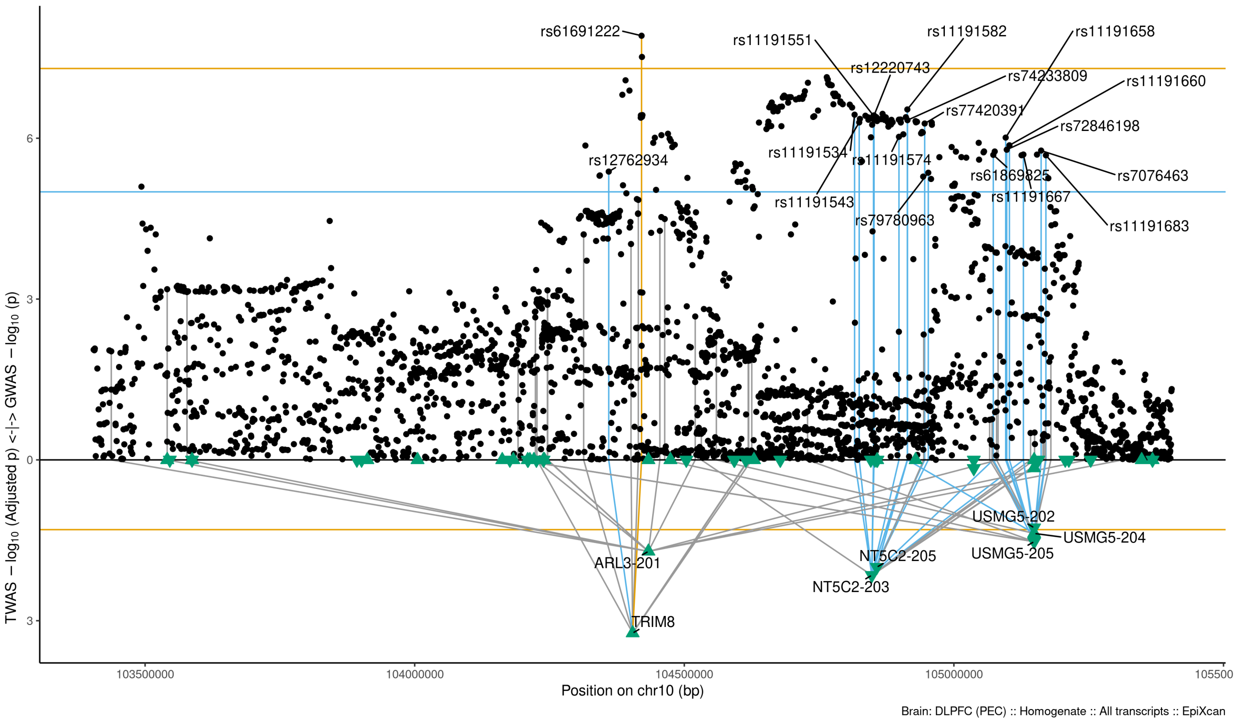

Figure S9-82: Regional Miami plots for TRIM8 gene/transcript : Regional Miami plots for primary MD GWASs corresponding to the genomic region of the TRIM8 transcript (1Mbp window from start site). Please also refer to Supplemental Table SXX for details. Top panel shows the GWAS results (black dots); blue line corresponds to $p = 1 \times 10 - 5$, orange line to $p = 5 \times 10 - 8$ (genome-wide significance). Bottom panel shows the TWAS results (green triangles facing upwards or downwards for a positive or negative z-score respectively (up- or down-regultion); only the transcripts with Bonferroni-adjusted $p < 0.1$ are labelled for clarity) for different transcripts (genes are represented by both gene expression and isoform expression); orange line corresponds to Bonferroni-adjusted p = 0.05. Each transcript that is Bonferroni-significant in the region is connected with lines to the SNPs that contribute to its transcriptomic imputation model; lines are grey when the SNPs have a $p > 1 \times 10 - 5$, blue when $p < 1 \times 10 - 5$ but $> 510 - 8$ and orange when $p < 5 \times 10 - 8$. The SNPs that are above the blue line and contribute to the transcriptomic imputation models of significant transcripts are labelled.

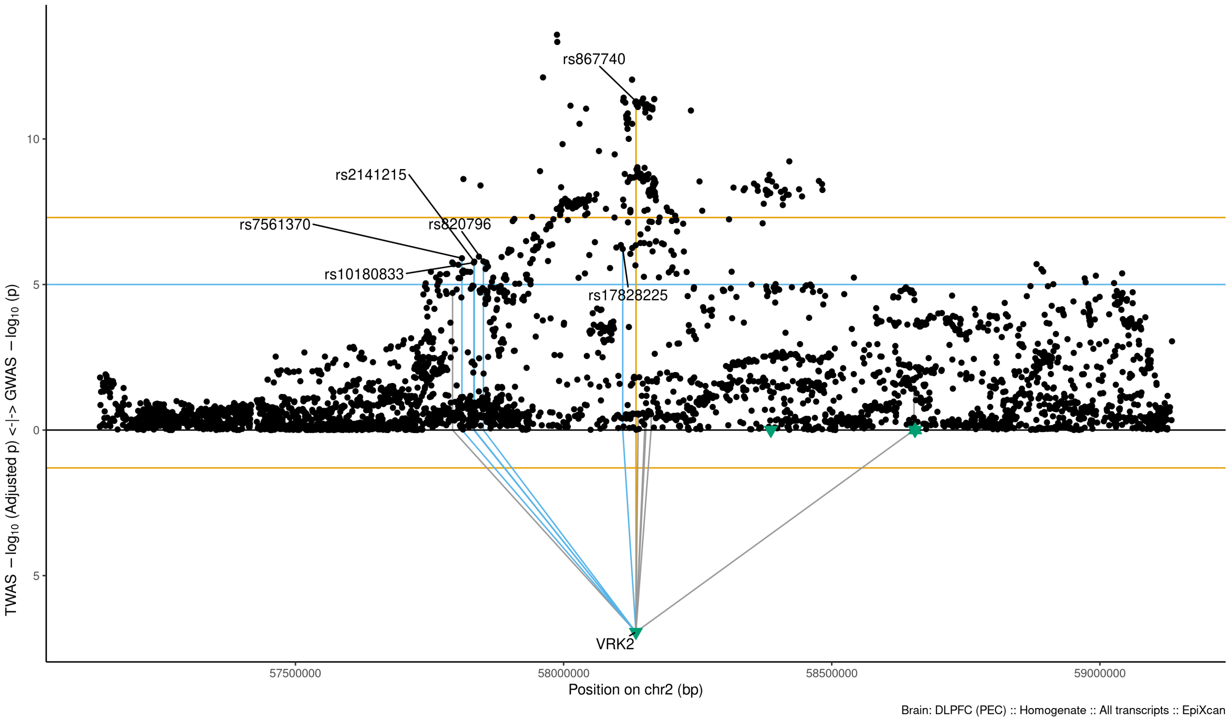

Figure S9-83: Regional Miami plots for VRK2 gene/transcript : Regional Miami plots for primary MD GWASs corresponding to the genomic region of the VRK2 transcript (1Mbp window from start site). Please also refer to Supplemental Table SXX for details. Top panel shows the GWAS results (black dots); blue line corresponds to $p = 1 \times 10 - 5$, orange line to $p = 5 \times 10 - 8$ (genome-wide significance). Bottom panel shows the TWAS results (green triangles facing upwards or downwards for a positive or negative z-score respectively (up- or down-regultion); only the transcripts with Bonferroni-adjusted $p < 0.1$ are labelled for clarity) for different transcripts (genes are represented by both gene expression and isoform expression); orange line corresponds to Bonferroni-adjusted p = 0.05. Each transcript that is Bonferroni-significant in the region is connected with lines to the SNPs that contribute to its transcriptomic imputation model; lines are grey when the SNPs have a $p > 1 \times 10 - 5$, blue when $p < 1 \times 10 - 5$ but $> 510 - 8$ and orange when $p < 5 \times 10 - 8$. The SNPs that are above the blue line and contribute to the transcriptomic imputation models of significant transcripts are labelled.

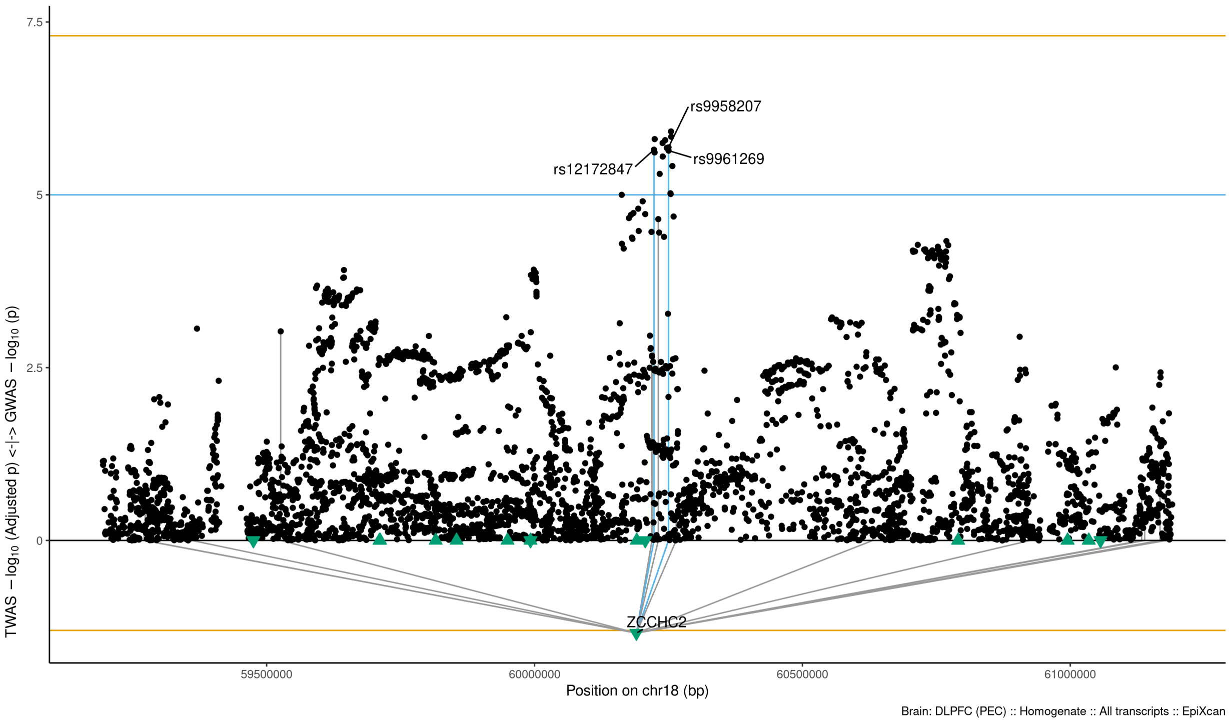

Figure S9-84: Regional Miami plots for ZCCHC2 gene/transcript : Regional Miami plots for primary MD GWASs corresponding to the genomic region of the ZCCHC2 transcript (1Mbp window from start site). Please also refer to Supplemental Table SXX for details. Top panel shows the GWAS results (black dots); blue line corresponds to $p = 1 \times 10 - 5$, orange line to $p = 5 \times 10 - 8$ (genome-wide significance). Bottom panel shows the TWAS results (green triangles facing upwards or downwards for a positive or negative z-score respectively (up- or down-regultion); only the transcripts with Bonferroni-adjusted $p < 0.1$ are labelled for clarity) for different transcripts (genes are represented by both gene expression and isoform expression); orange line corresponds to Bonferroni-adjusted p = 0.05. Each transcript that is Bonferroni-significant in the region is connected with lines to the SNPs that contribute to its transcriptomic imputation model; lines are grey when the SNPs have a $p > 1 \times 10 - 5$, blue when $p < 1 \times 10 - 5$ but $> 510 - 8$ and orange when $p < 5 \times 10 - 8$. The SNPs that are above the blue line and contribute to the transcriptomic imputation models of significant transcripts are labelled.

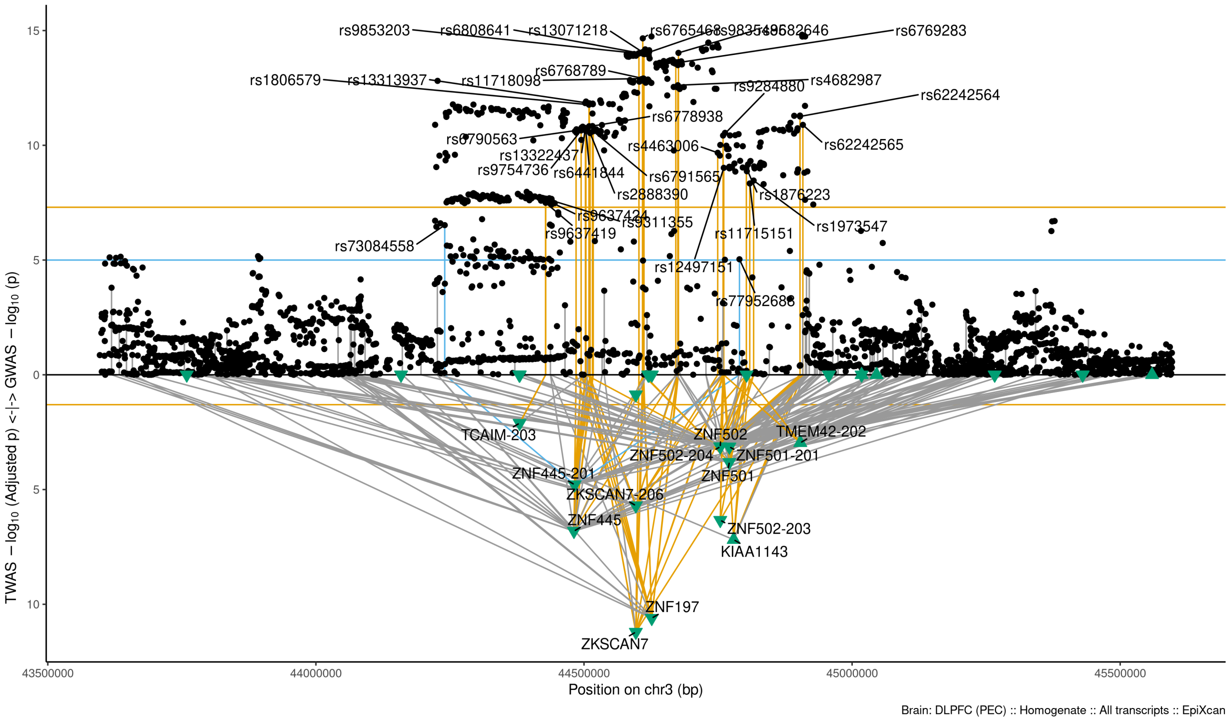

Figure S9-85: Regional Miami plots for ZKSCAN7 gene/transcript : Regional Miami plots for primary MD GWASs corresponding to the genomic region of the ZKSCAN7 transcript (1Mbp window from start site). Please also refer to Supplemental Table SXX for details. Top panel shows the GWAS results (black dots); blue line corresponds to $p = 1 \times 10 - 5$, orange line to $p = 5 \times 10 - 8$ (genome-wide significance). Bottom panel shows the TWAS results (green triangles facing upwards or downwards for a positive or negative z-score respectively (up- or down-regultion); only the transcripts with Bonferroni-adjusted $p < 0.1$ are labelled for clarity) for different transcripts (genes are represented by both gene expression and isoform expression); orange line corresponds to Bonferroni-adjusted p = 0.05. Each transcript that is Bonferroni-significant in the region is connected with lines to the SNPs that contribute to its transcriptomic imputation model; lines are grey when the SNPs have a $p > 1 \times 10 - 5$, blue when $p < 1 \times 10 - 5$ but $> 510 - 8$ and orange when $p < 5 \times 10 - 8$. The SNPs that are above the blue line and contribute to the transcriptomic imputation models of significant transcripts are labelled.

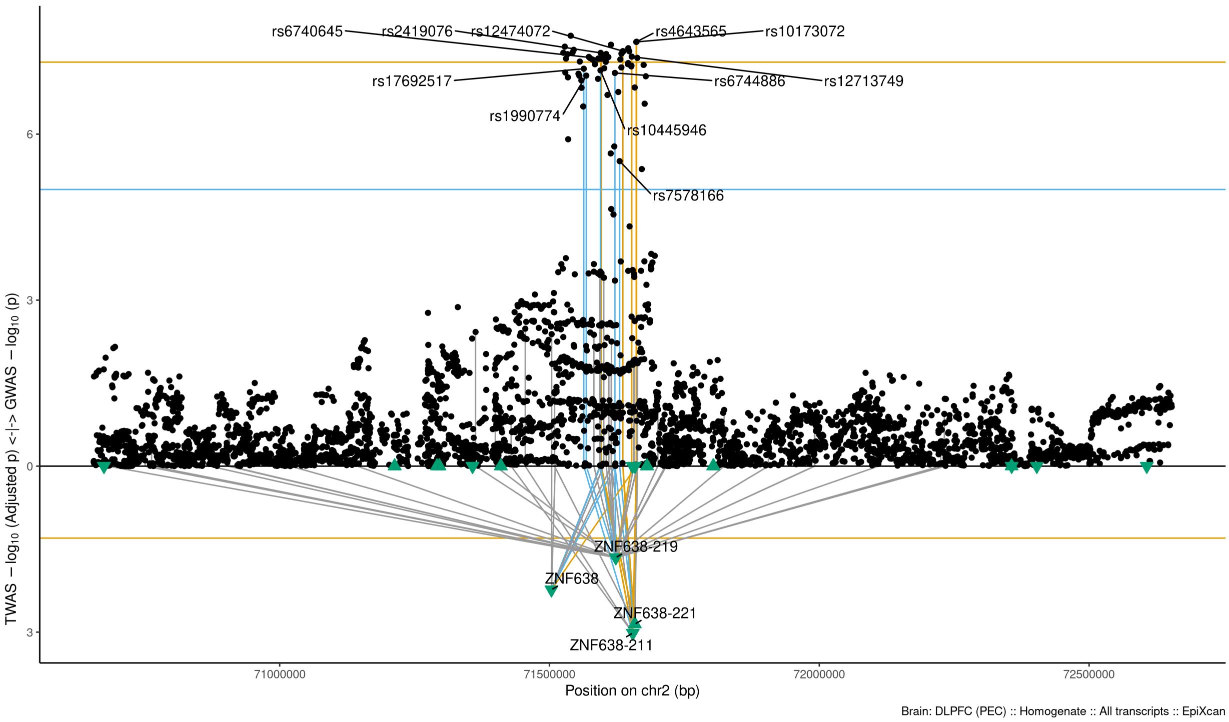

Figure S9-86: Regional Miami plots for ZNF638211 gene/transcript : Regional Miami plots for primary MD GWASs corresponding to the genomic region of the ZNF638211 transcript (1Mbp window from start site). Please also refer to Supplemental Table SXX for details. Top panel shows the GWAS results (black dots); blue line corresponds to $p = 1 \times 10 - 5$, orange line to $p = 5 \times 10 - 8$ (genome-wide significance). Bottom panel shows the TWAS results (green triangles facing upwards or downwards for a positive or negative z-score respectively (up- or down-regultion); only the transcripts with Bonferroni-adjusted $p < 0.1$ are labelled for clarity) for different transcripts (genes are represented by both gene expression and isoform expression); orange line corresponds to Bonferroni-adjusted p = 0.05. Each transcript that is Bonferroni-significant in the region is connected with lines to the SNPs that contribute to its transcriptomic imputation model; lines are grey when the SNPs have a $p > 1 \times 10 - 5$, blue when $p < 1 \times 10 - 5$ but $> 510 - 8$ and orange when $p < 5 \times 10 - 8$. The SNPs that are above the blue line and contribute to the transcriptomic imputation models of significant transcripts are labelled.

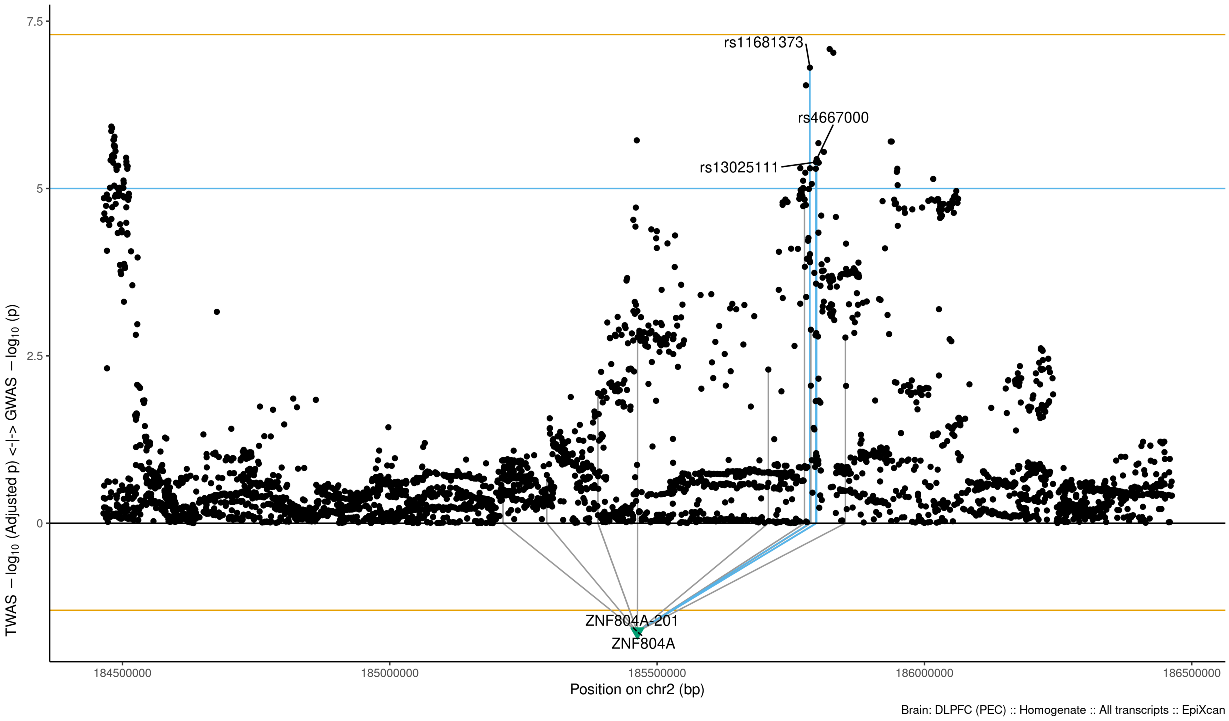

Figure S9-87: Regional Miami plots for ZNF804A gene/transcript : Regional Miami plots for primary MD GWASs corresponding to the genomic region of the ZNF804A transcript (1Mbp window from start site). Please also refer to Supplemental Table SXX for details. Top panel shows the GWAS results (black dots); blue line corresponds to $p = 1 \times 10 - 5$, orange line to $p = 5 \times 10 - 8$ (genome-wide significance). Bottom panel shows the TWAS results (green triangles facing upwards or downwards for a positive or negative z-score respectively (up- or down-regultion); only the transcripts with Bonferroni-adjusted $p < 0.1$ are labelled for clarity) for different transcripts (genes are represented by both gene expression and isoform expression); orange line corresponds to Bonferroni-adjusted p = 0.05. Each transcript that is Bonferroni-significant in the region is connected with lines to the SNPs that contribute to its transcriptomic imputation model; lines are grey when the SNPs have a $p > 1 \times 10 - 5$, blue when $p < 1 \times 10 - 5$ but $> 510 - 8$ and orange when $p < 5 \times 10 - 8$. The SNPs that are above the blue line and contribute to the transcriptomic imputation models of significant transcripts are labelled.

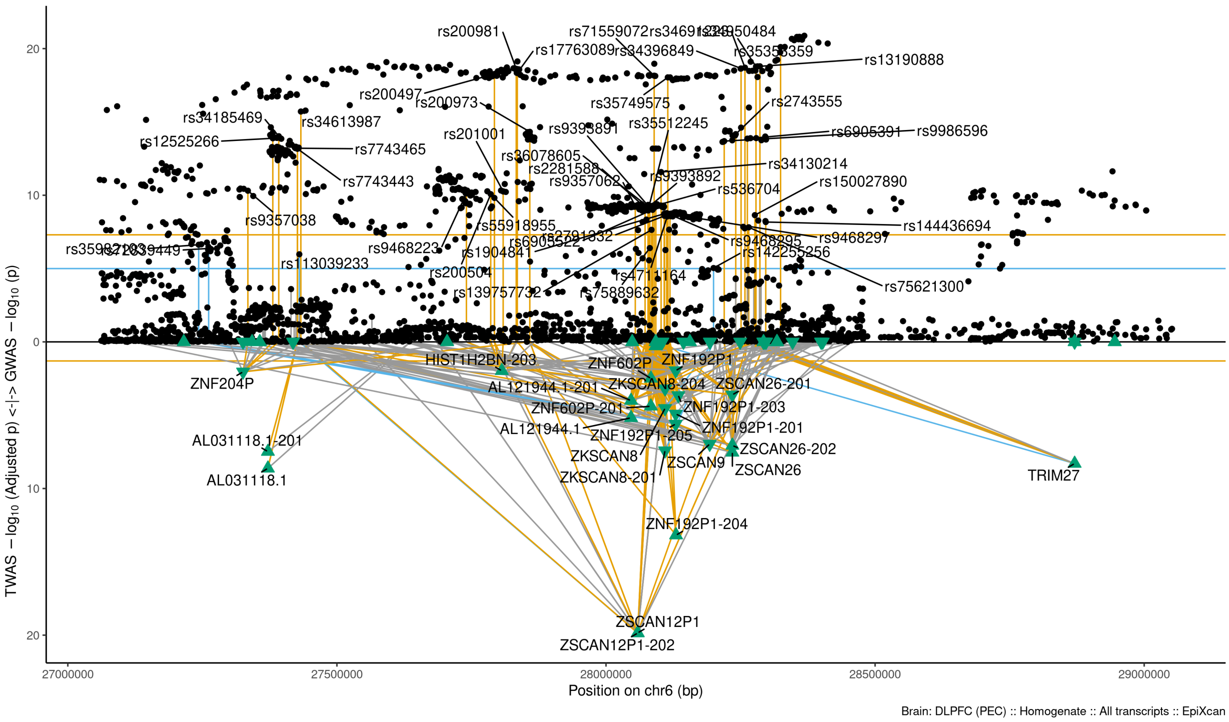

Figure S9-88: Regional Miami plots for ZSCAN12P1202 gene/transcript : Regional Miami plots for primary MD GWASs corresponding to the genomic region of the ZSCAN12P1202 transcript (1Mbp window from start site). Please also refer to Supplemental Table SXX for details. Top panel shows the GWAS results (black dots); blue line corresponds to $p = 1 \times 10 - 5$, orange line to $p = 5 \times 10 - 8$ (genome-wide significance). Bottom panel shows the TWAS results (green triangles facing upwards or downwards for a positive or negative z-score respectively (up- or down-regultion); only the transcripts with Bonferroni-adjusted $p < 0.1$ are labelled for clarity) for different transcripts (genes are represented by both gene expression and isoform expression); orange line corresponds to Bonferroni-adjusted p = 0.05. Each transcript that is Bonferroni-significant in the region is connected with lines to the SNPs that contribute to its transcriptomic imputation model; lines are grey when the SNPs have a $p > 1 \times 10 - 5$, blue when $p < 1 \times 10 - 5$ but $> 510 - 8$ and orange when $p < 5 \times 10 - 8$. The SNPs that are above the blue line and contribute to the transcriptomic imputation models of significant transcripts are labelled.

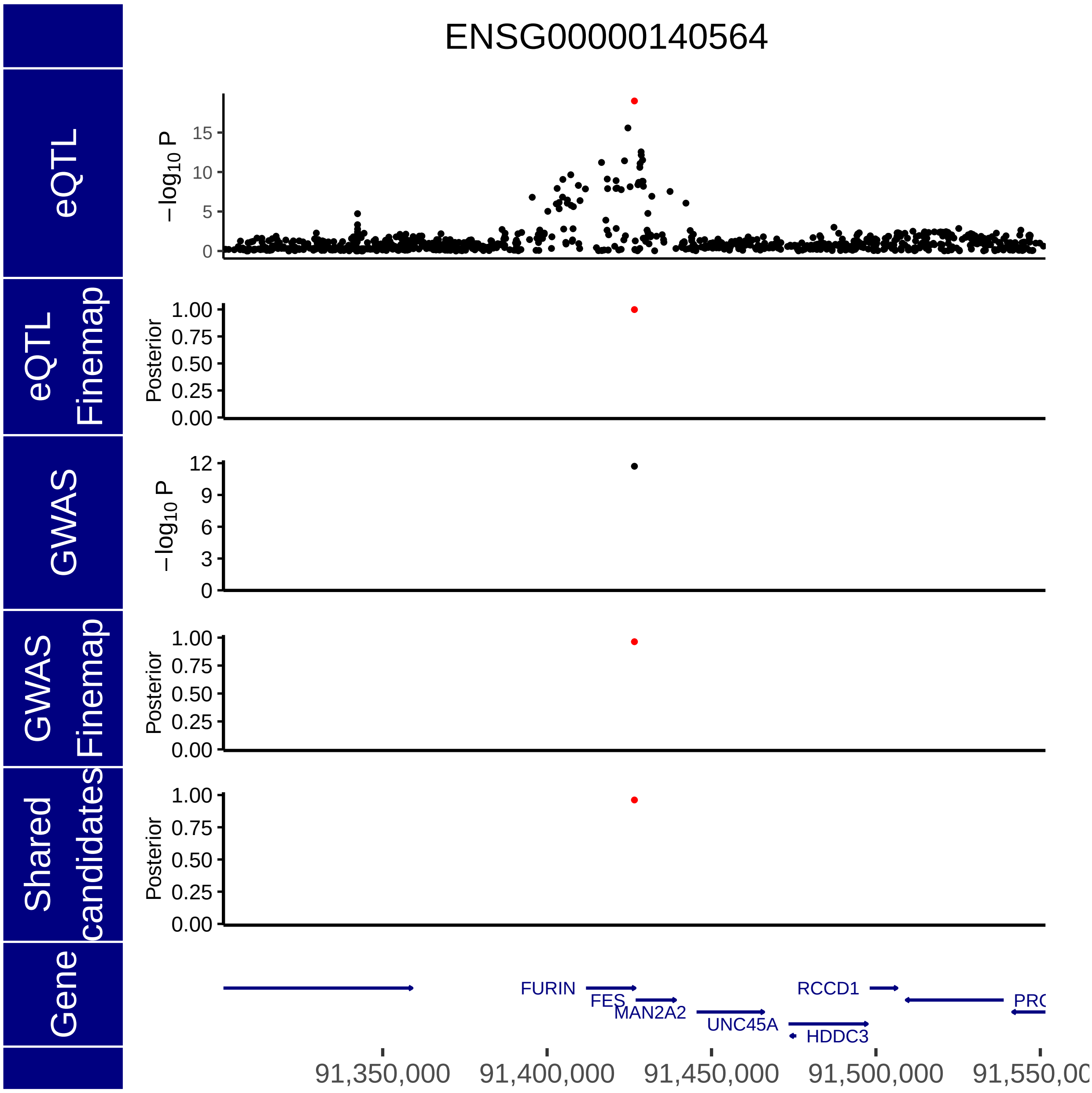

Figure S10-1: eCAVIAR - Gene 1: *FURIN*. eQTL and GWAS CAusal Variants Identification in Associated Regions (eCAVIAR) approach. Genomic position is shown on the x-axis, while the y-axis show $-\log_{10}(P)$ for eQTL analysis, the posterior probability of the eQTL finemapping, $-\log_{10}(P)$ from the GWAS, posterior probability from the GWAS fine mapping and the posterior probability for the shared candidates from both analyses.

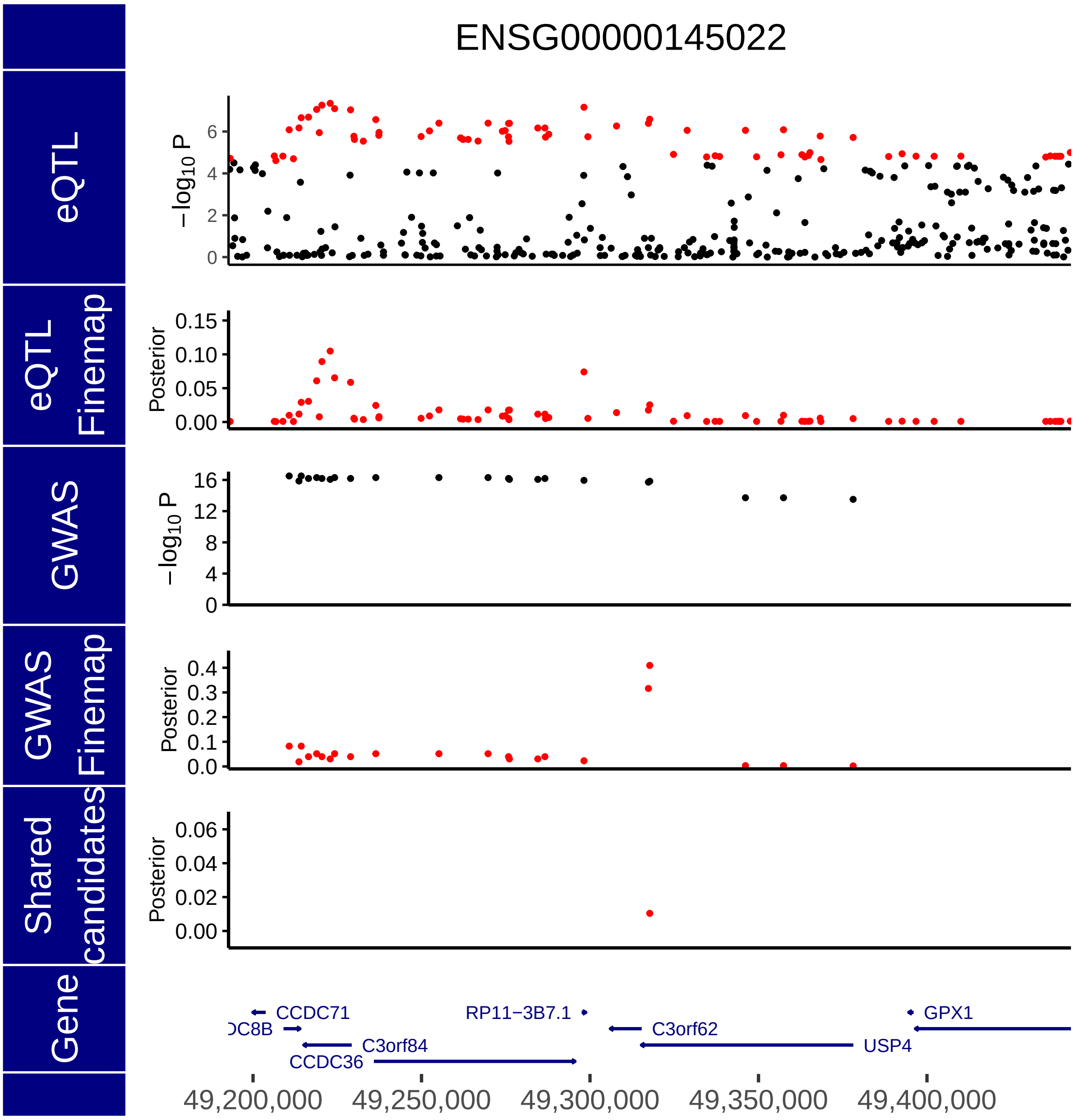

Figure S10-2: eCAVIAR - Gene 2: *TCTA*. eQTL and GWAS CAusal Variants Identification in Associated Regions (eCAVIAR) approach. Genomic position is shown on the x-axis, while the y-axis show $-\log_{10}(P)$ for eQTL analysis, the posterior probability of the eQTL finemapping, $-\log_{10}(P)$ from the GWAS, posterior probability from the GWAS fine mapping and the posterior probability for the shared candidates from both analyses.

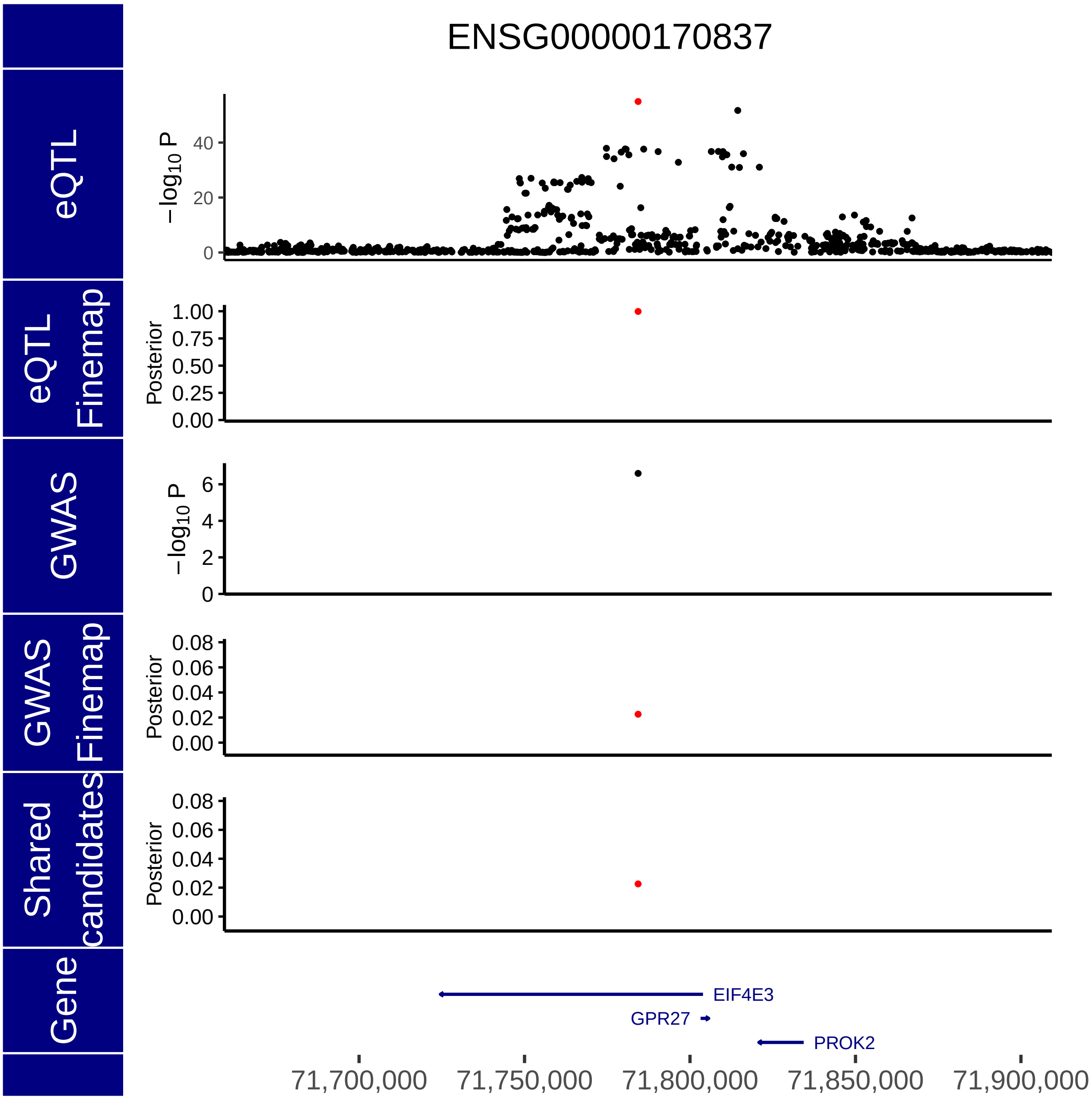

Figure S10-3: eCAVIAR - Gene 3: *GRP27*. eQTL and GWAS CAusal Variants Identification in Associated Regions (eCAVIAR) approach. Genomic position is shown on the x-axis, while the y-axis show $-\log_{10}(P)$ for eQTL analysis, the posterior probability of the eQTL finemapping, $-\log_{10}(P)$ from the GWAS, posterior probability from the GWAS fine mapping and the posterior probability for the shared candidates from both analyses.

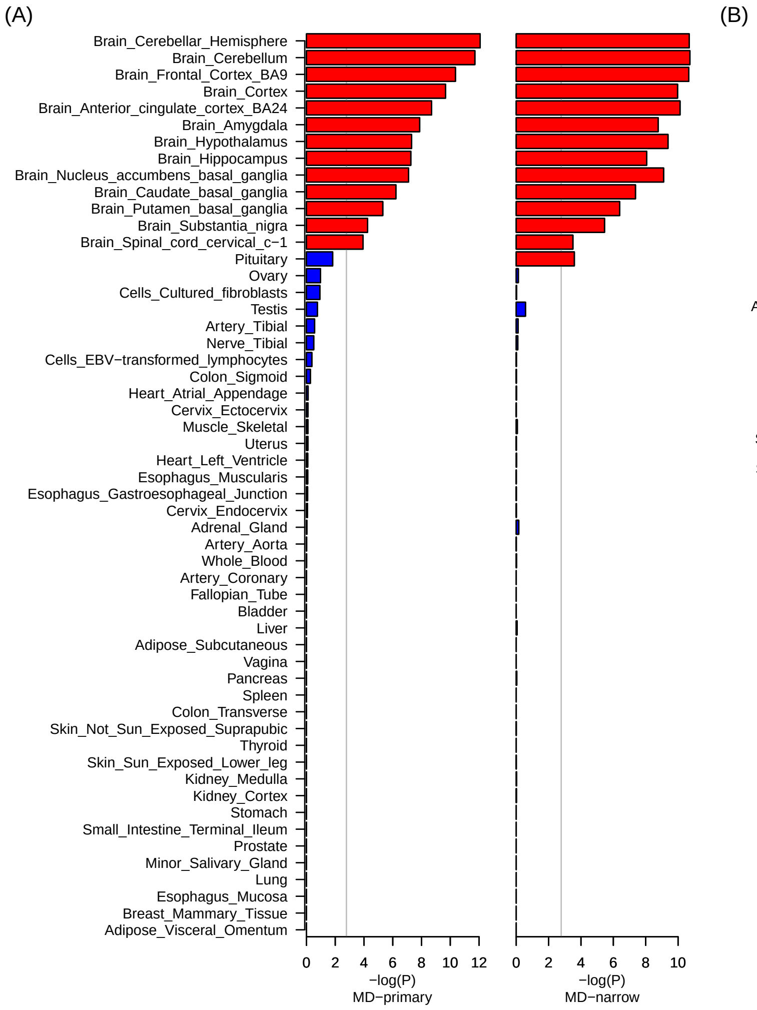

Figure S11: Results of gene-property analysis in MAGMA as implemented in FUMA, for the primary ($N_{cases}$ = 371184 and $N_{ctrls}$ = 978703) and narrow MD phenotype ($N_{cases}$ = 176143 and $N_{ctrls}$ = 528770). Tissue specific data is obtained from the GTEx v8 dataset (https://www.gtexportal.org)8. Tissues with red bar surpass experiment-wide significance. (A) Individual tissue type analyses with 54 tissue types. (B) General tissue type analysis with 30 tissues.

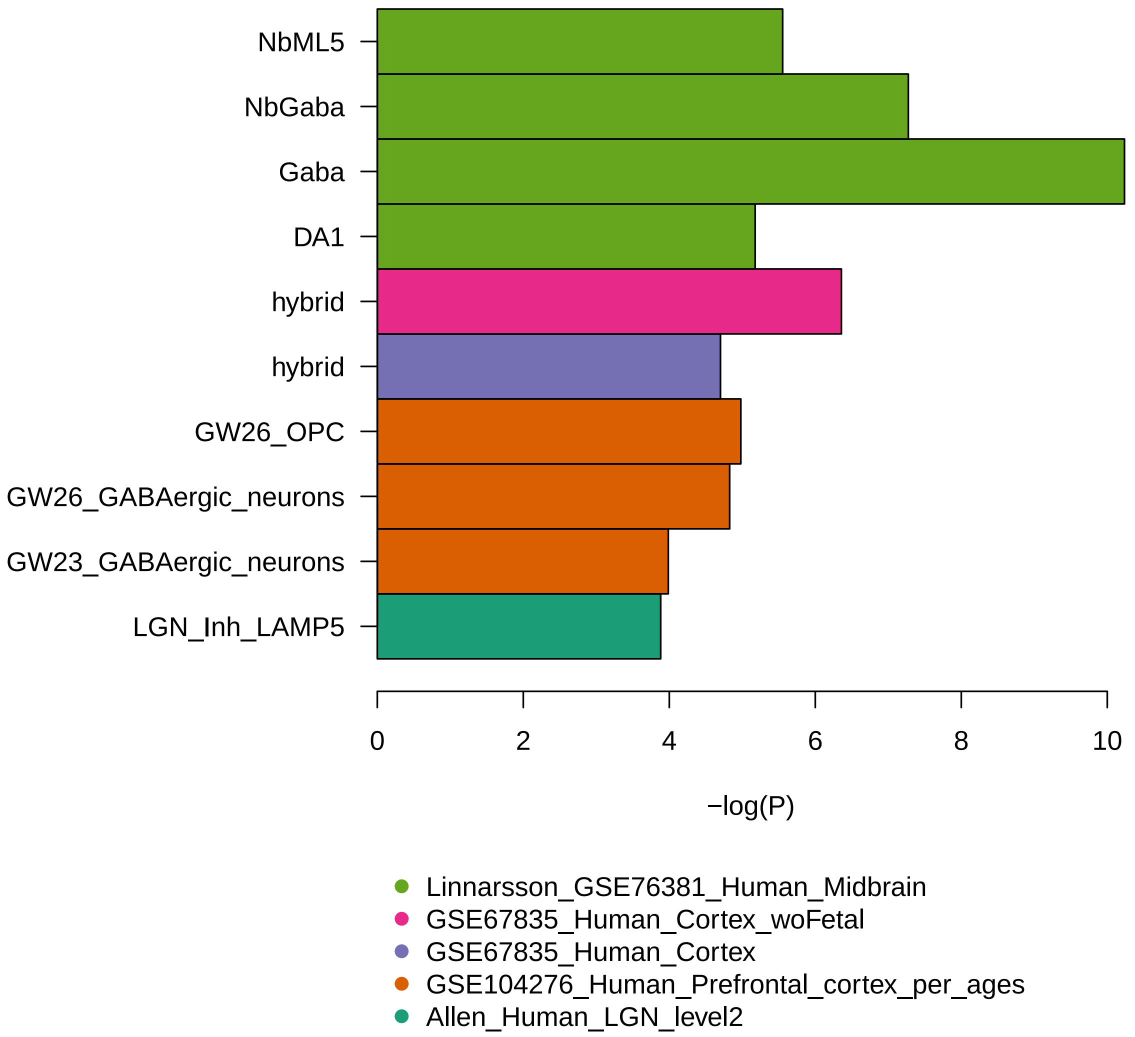

Figure S12: Nominal p-values for FUMA cell-type analysis significant across datasets, i.e. after experiment wide bonferroni correction. The primary MD GWAS ($N_{cases}$ = 371184 and $N_{ctrls}$ = 978703) was used as input for this analysis.

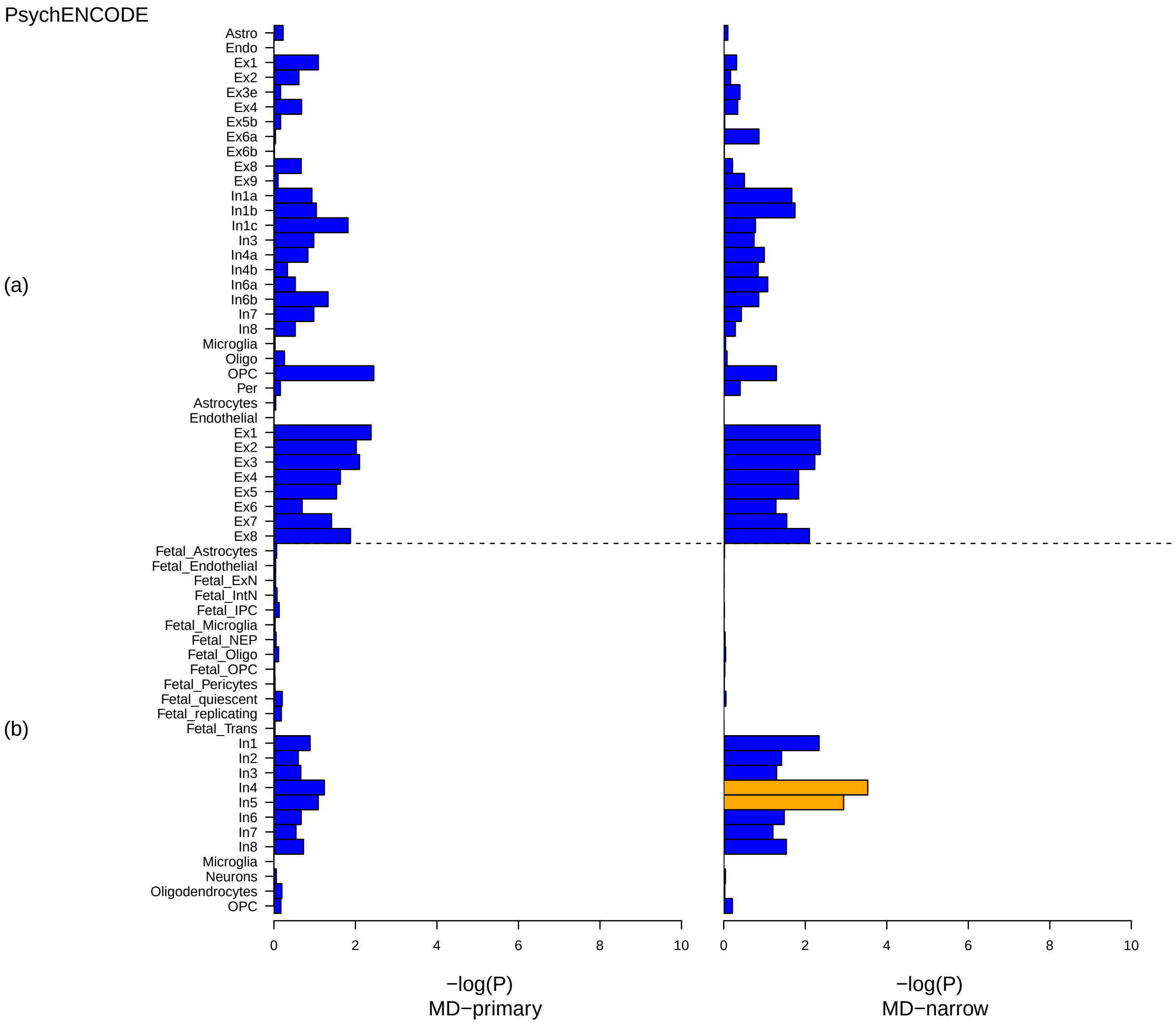

Figure S13-1: FUMA single-cell analysis for MD GWAS. The primary ($Ncases = 371184$ and $Nctrls = 978703$) and narrow MD ($Ncases = 176143$ $Nctrls = 528770$) GWAS was used as input for this analysis.. Cell types with blue bars are not significant, bars in yellow are significant within the tested dataset, while bars in red are significant after correction for multiple testing across datasets. Per-dataset cell type specificity: (a) PsychENCODE Developmental, (b) PscyhENCODE Adult

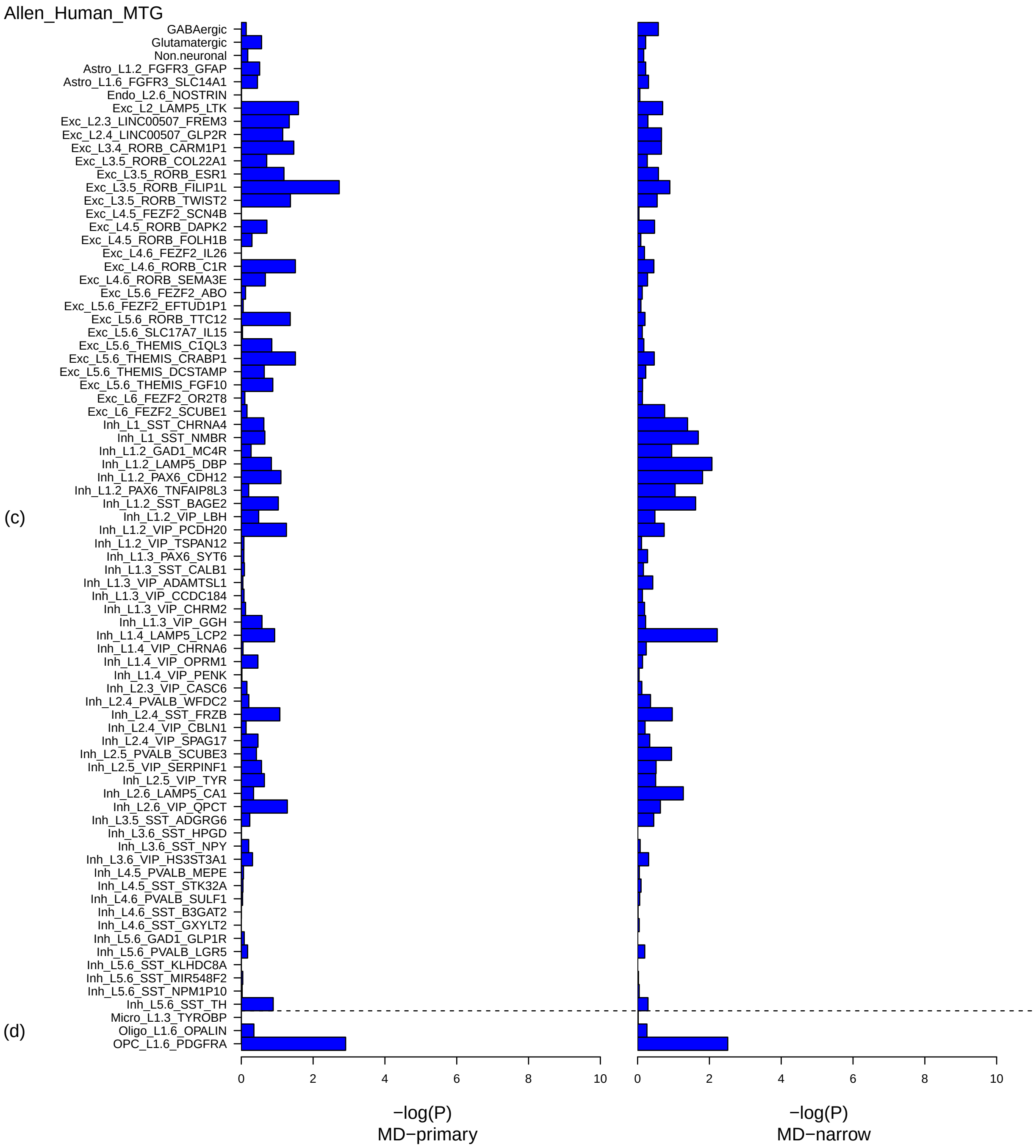

Figure S13-2: FUMA single-cell analysis for MD GWAS. The primary ($Ncases = 371184$ and $Nctrls = 978703$) and narrow MD ($Ncases = 176143$ $Nctrls = 528770$) GWAS was used as input for this analysis.. Cell types with blue bars are not significant, bars in yellow are significant within the tested dataset, while bars in red are significant after correction for multiple testing across datasets. Per-dataset cell type specificity: (c) Allen Human MTG level1, (d) Allen Human MTG level2

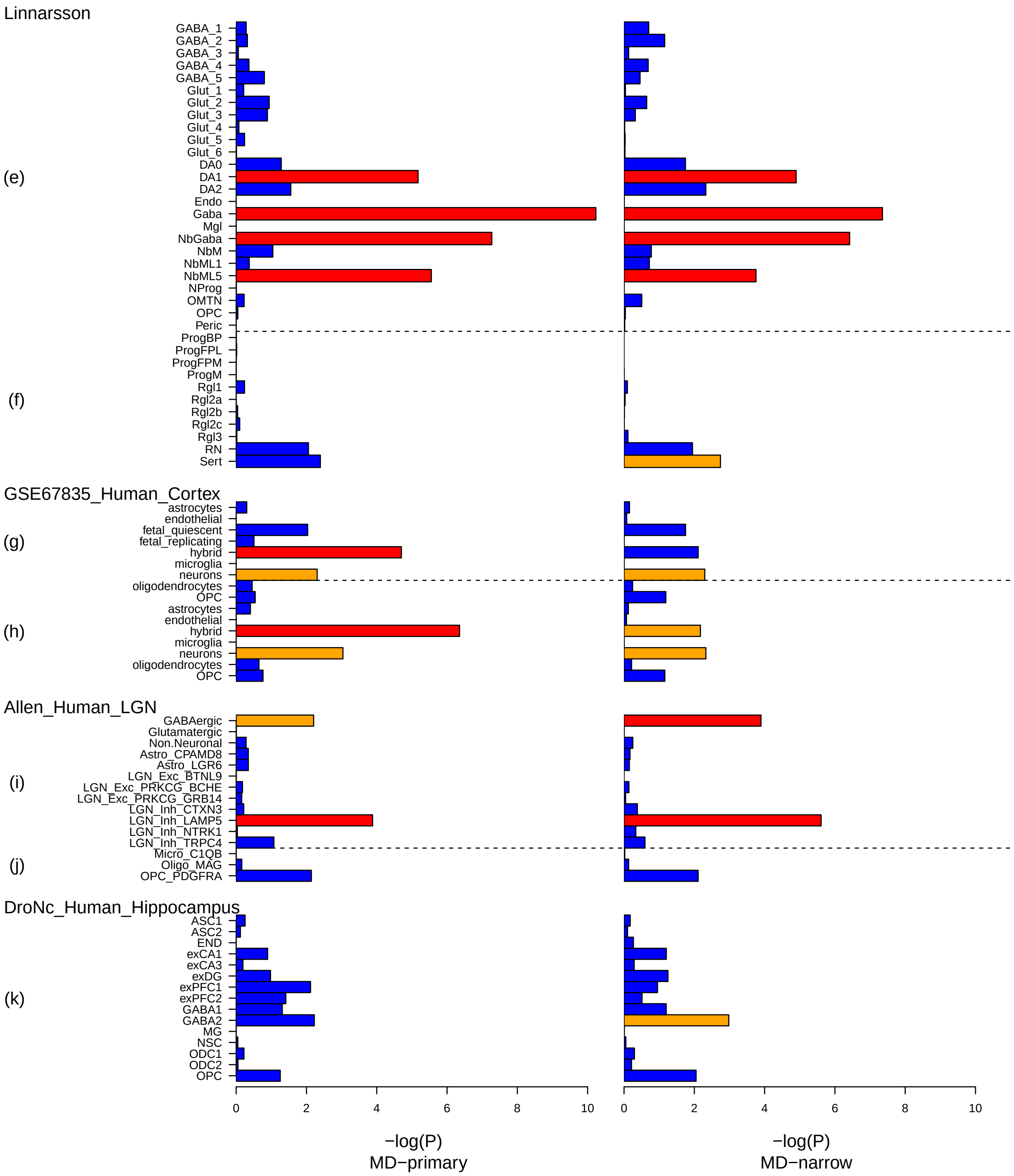

Figure S13-3: FUMA single-cell analysis for MD GWAS. The primary ($Ncases = 371184$ and $Nctrls = 978703$) and narrow MD ($Ncases = 176143$ $Nctrls = 528770$) GWAS was used as input for this analysis.. Cell types with blue bars are not significant, bars in yellow are significant within the tested dataset, while bars in red are significant after correction for multiple testing across datasets. Per-dataset cell type specificity: (e) Linnarsson GSE76381 Human Midbrain, (f) Linnarsson GSE101601 Human Temporal cortex, (g) GSE67835 Human Cortex woFetal, (h) GSE67835 Human Cortex, (i) Allen Human LGN level2, (j) Allen Human LGN level1, (k) DroNc Human Hippocampus

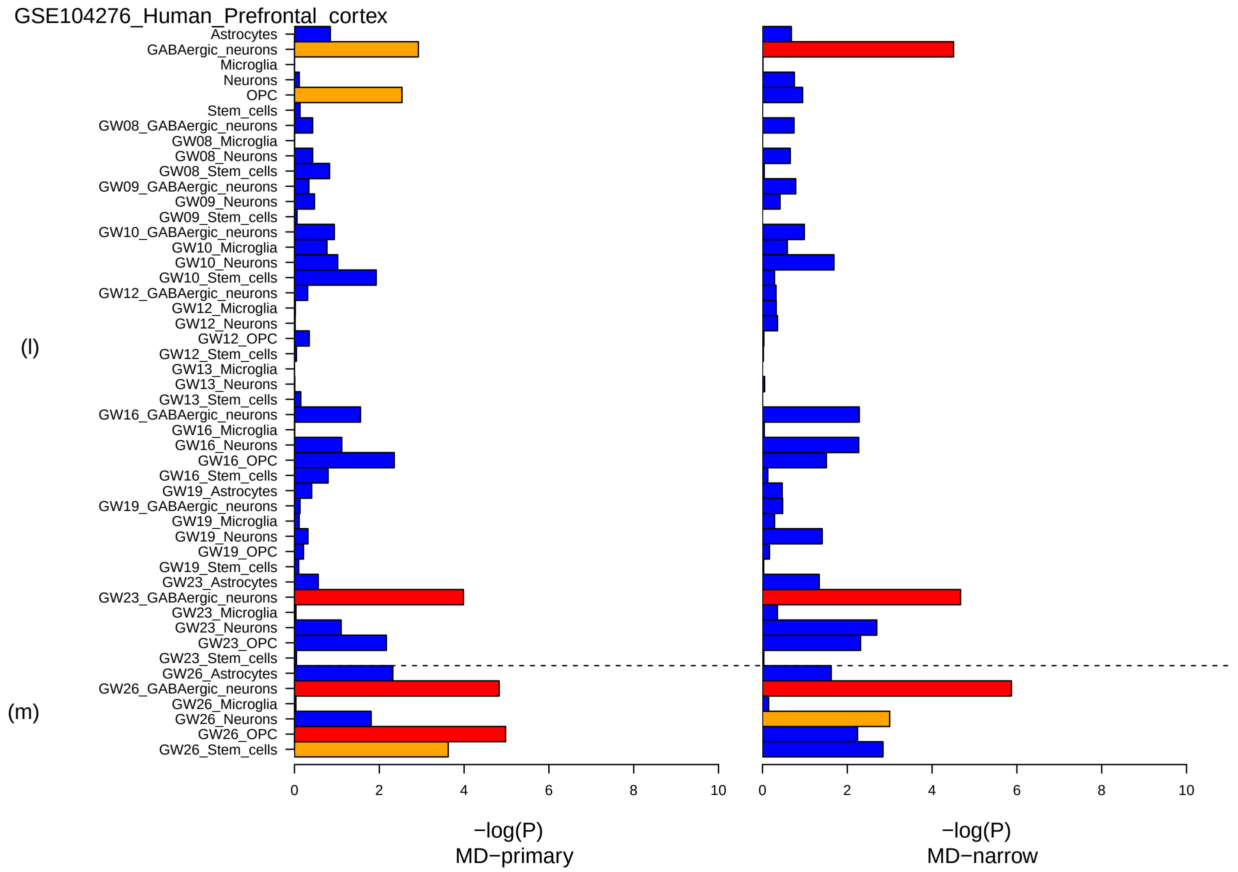

Figure S13-4: FUMA single-cell analysis for MD GWAS. The primary ($Ncases = 371184$ and $Nctrls = 978703$) and narrow MD ($Ncases = 176143$ $Nctrls = 528770$) GWAS was used as input for this analysis.. Cell types with blue bars are not significant, bars in yellow are significant within the tested dataset, while bars in red are significant after correction for multiple testing across datasets. Per-dataset cell type specificity: (l) GSE104276 Human Prefrontal cortex per ages, (m) GSE104276 Human Prefrontal cortex all ages-

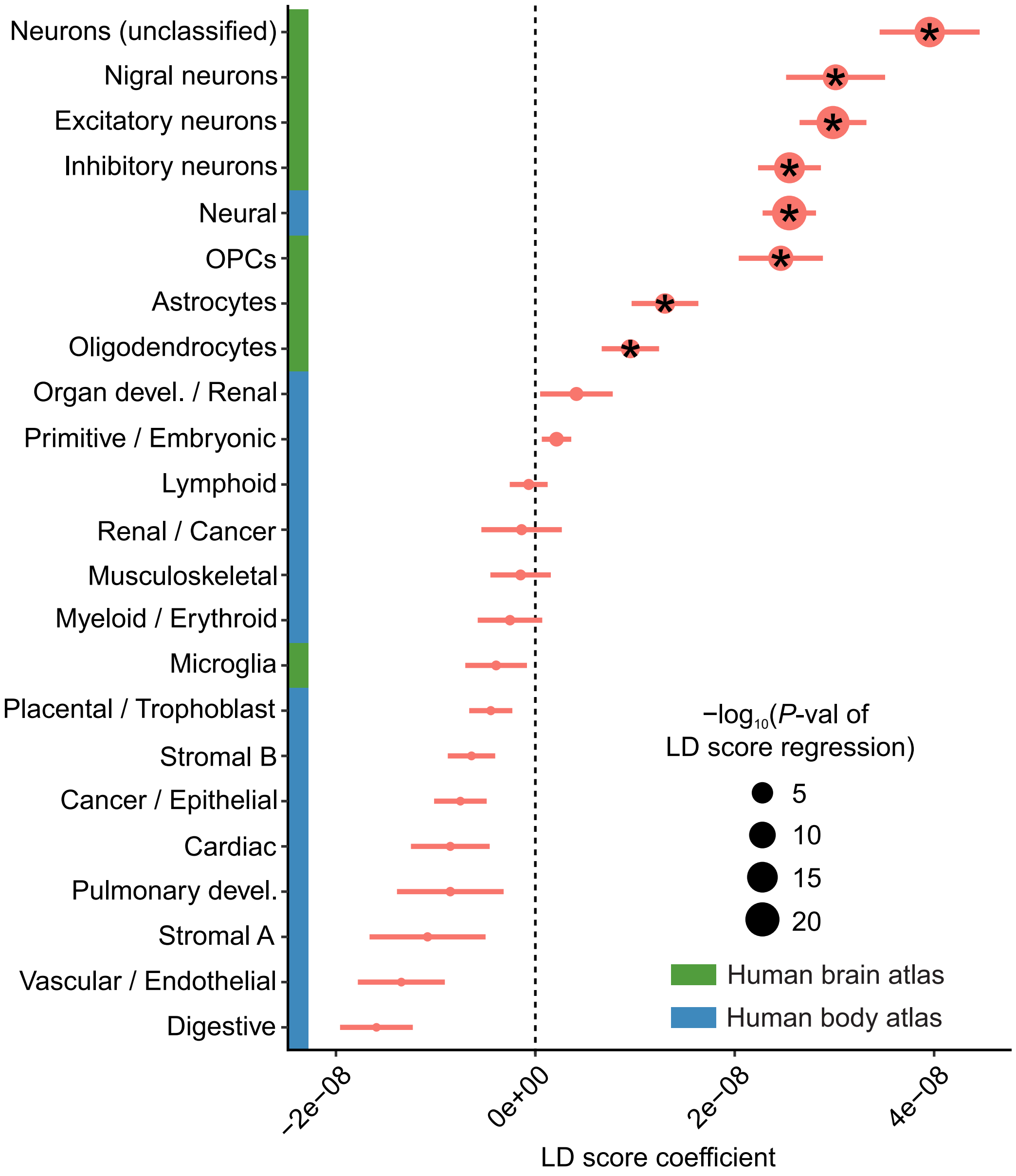

Figure S14: Co-localization of common MD risk variants with cell-specific open chromatin regions. LD score coefficients with standard error (x-axis) for various cell-types (y-axis). A positive LD score coefficient signifies enrichment in heritability. Dot size reflects P-value of LD score regression, '*' denotes test-wide significant associations. The primary MD GWAS ($N_{cases} = 371184$ and $N_{ctrls} = 978703$) was used as input for this analysis

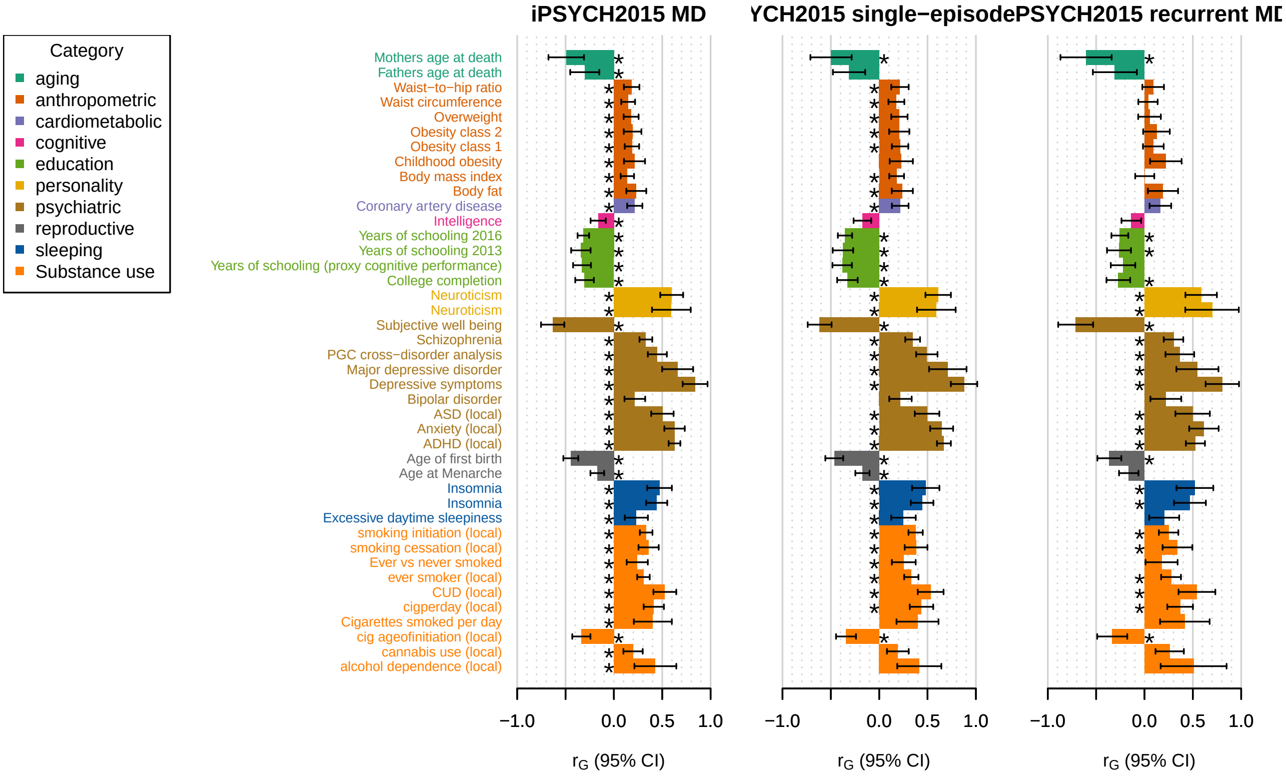

Figure S15A: $r_G$ of MD, single-episode and recurrent depresion with non-UKB phenotypes A total of 42 non-UKBB traits showed significant genetic correlation with either MD ($N_{cases}$ = 29158 and $N_{ctrls}$ = 38142), single-episode ($N_{cases}$ = 23140 and $N_{ctrls}$ = 38142) or recurrent depression ($N_{cases}$ = 6018 and $N_{ctrls}$ = 38142) within the iPSYCH2015 cohors. ASD data not available on LDhub, but at genomeDK.

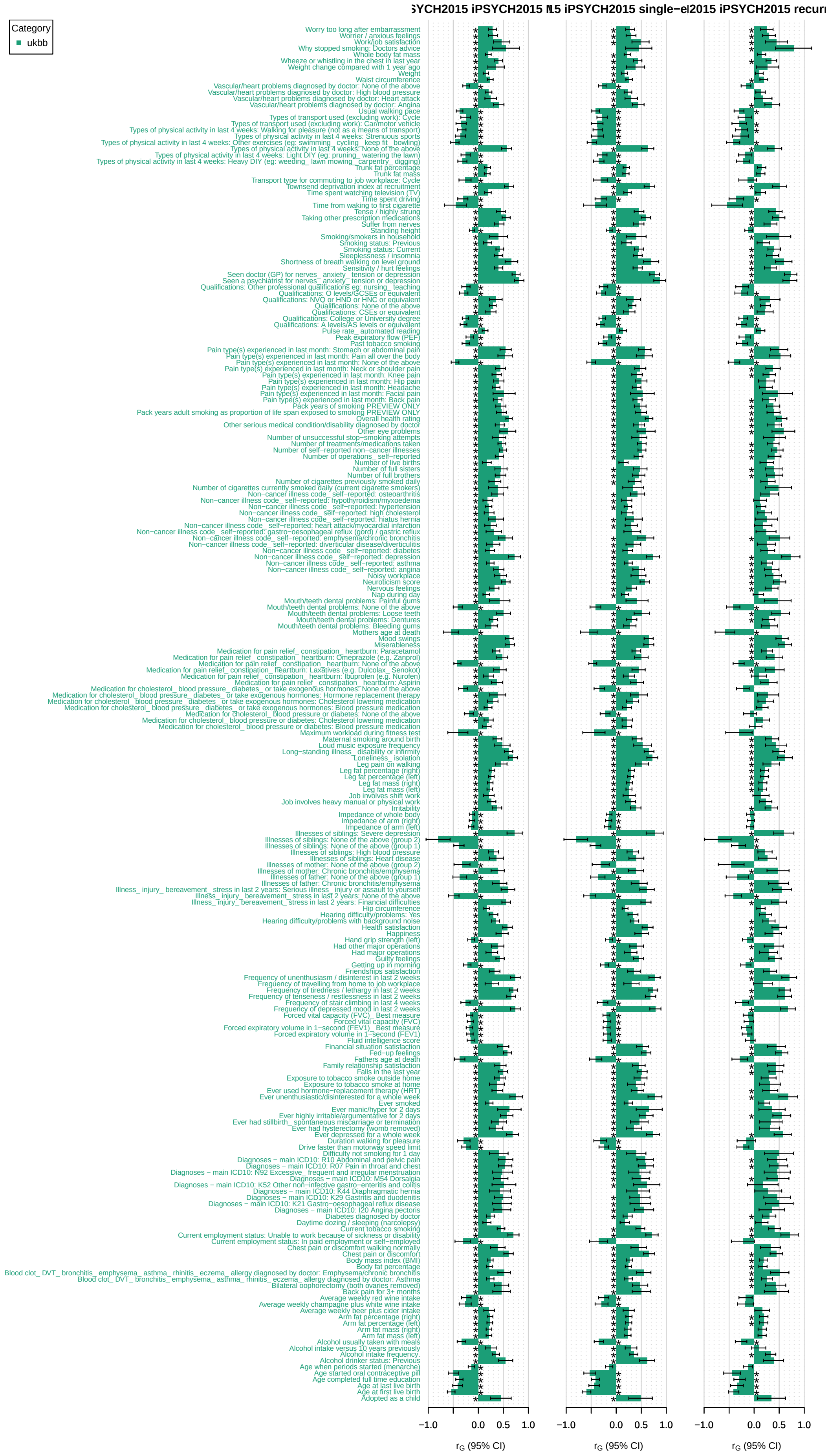

Figure S15B: $r_G$ of MD, single-episode and recurrent depresion with UKB phenotypes A total of 219 UKBB traits showed significant genetic correlation with either MD ($N_{cases}$ = 29158 and $N_{ctrls}$ = 38142), single-episode ($N_{cases}$ = 23140 and $N_{ctrls}$ = 38142) or recurrent depression ($N_{cases}$ = 6018 and $N_{ctrls}$ = 38142) within the iPSYCH2015 cohors

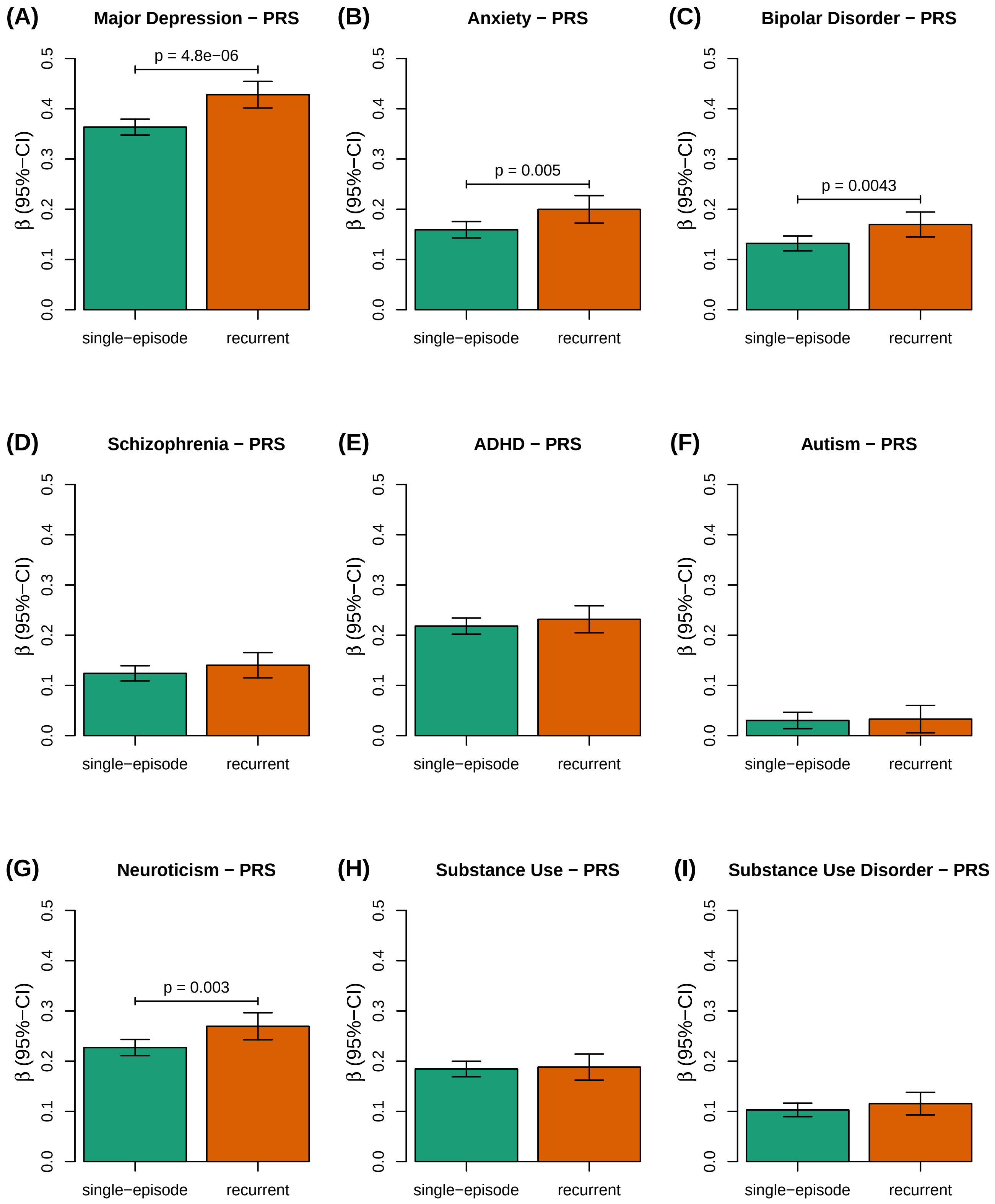

Figure S16: Multivariate PRS analyses of recurrent vs single-episode depression (overall p-value = 0.00075). A total of 23140 cases with single-episode, 6018 with recurrent depression and 38142 controls were analysed for their polygenic load of (A) Howard-FinnGen-MVP-MD. (B) ANX-FinnGenCore-MVP. (C) PGC3-BP-woDK. (D) PGC3-SCZ-woDK. (E) ADHD-iPSYCHwoPGC2. (F) ASD-PGC-AUT14. (G) Neurot2018-Sum. (H) SU. (I) CUDALD.

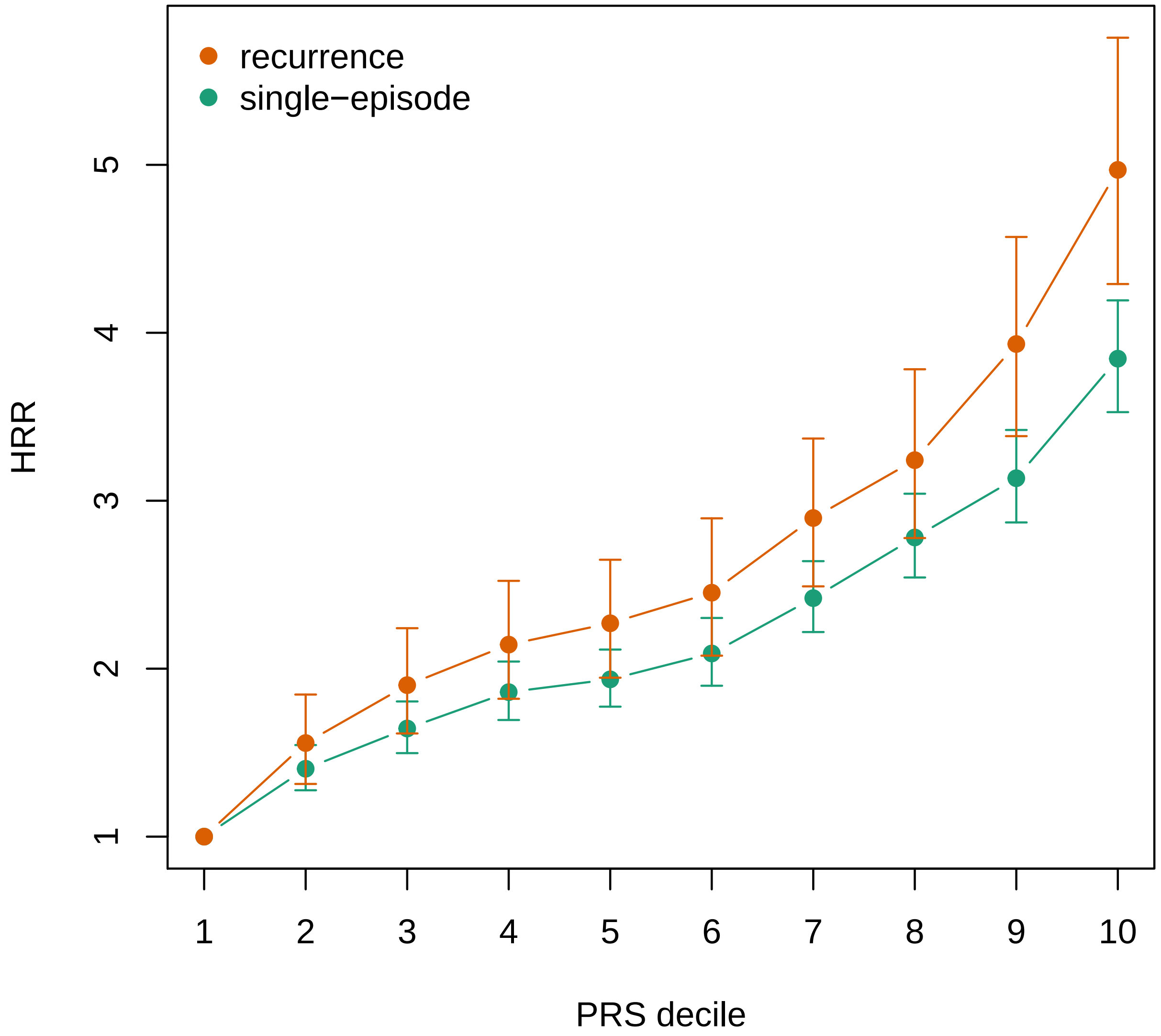

Figure S17: Hazard rate ratios (HRR) of single-episode and recurrent depression stratified by MD-PRS deciles . Results from two seperate cox-regression analyses were conducted: (A) A total of 23140 cases with single-episode depression and 38142 controls were stratified according to MD-PRS deciles and HRR were calculated for decile 2:10 compared to the first decile. (B) A total of 6018 cases with recurrent depression and 38142 controls were stratified according to MD-PRS deciles and HRR were calculated for decile 2:10 compared to the first decile.

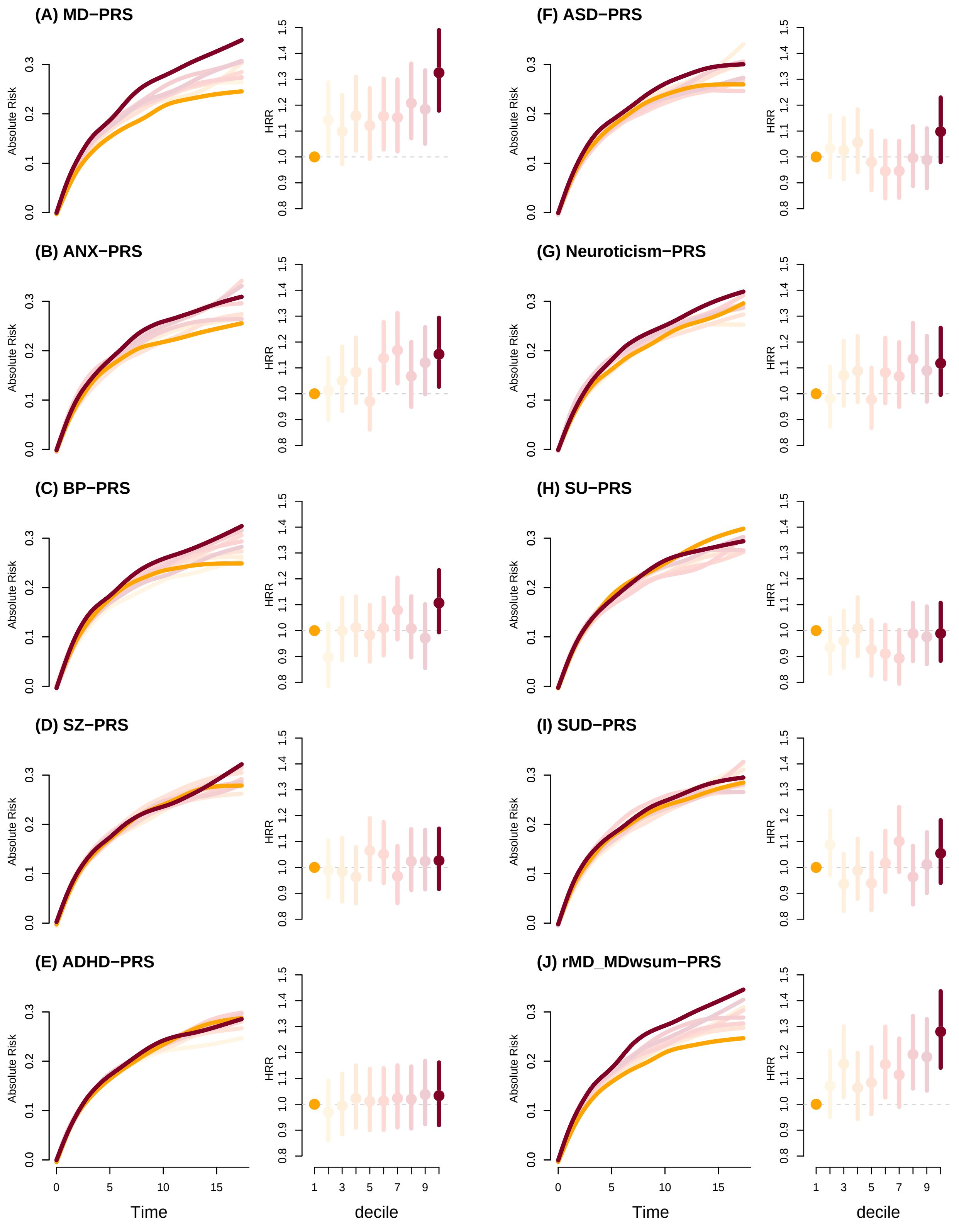

Figure S18: Absolute risk and Hazard rate ratios (HRR) of developing second-episode of depression for deciles of ten different PRSs estimated using cox's regression. HRRs (95%) were calculated for decile 2-10 compared to the first decile. The analyses were conducted for PRS deciles of MD, BP, SCZ, ADHD, ASD, Neuroticism, Substance Use, Substance Use Disorder and sum of PRSs weighted by $\log(OR_{\text{rMD vs sMD}})$. (A) MD-PRS, (B) ANX-PRS, (C) BP-PRS, (D) SZ-PRS, (E) ADHD-PRS, (F) ASD-PRS, (G) Neuroticism-PRS, (H) SU-PRS, (I) SUD-PRS, (J) rMD-MDwsum-PRS,

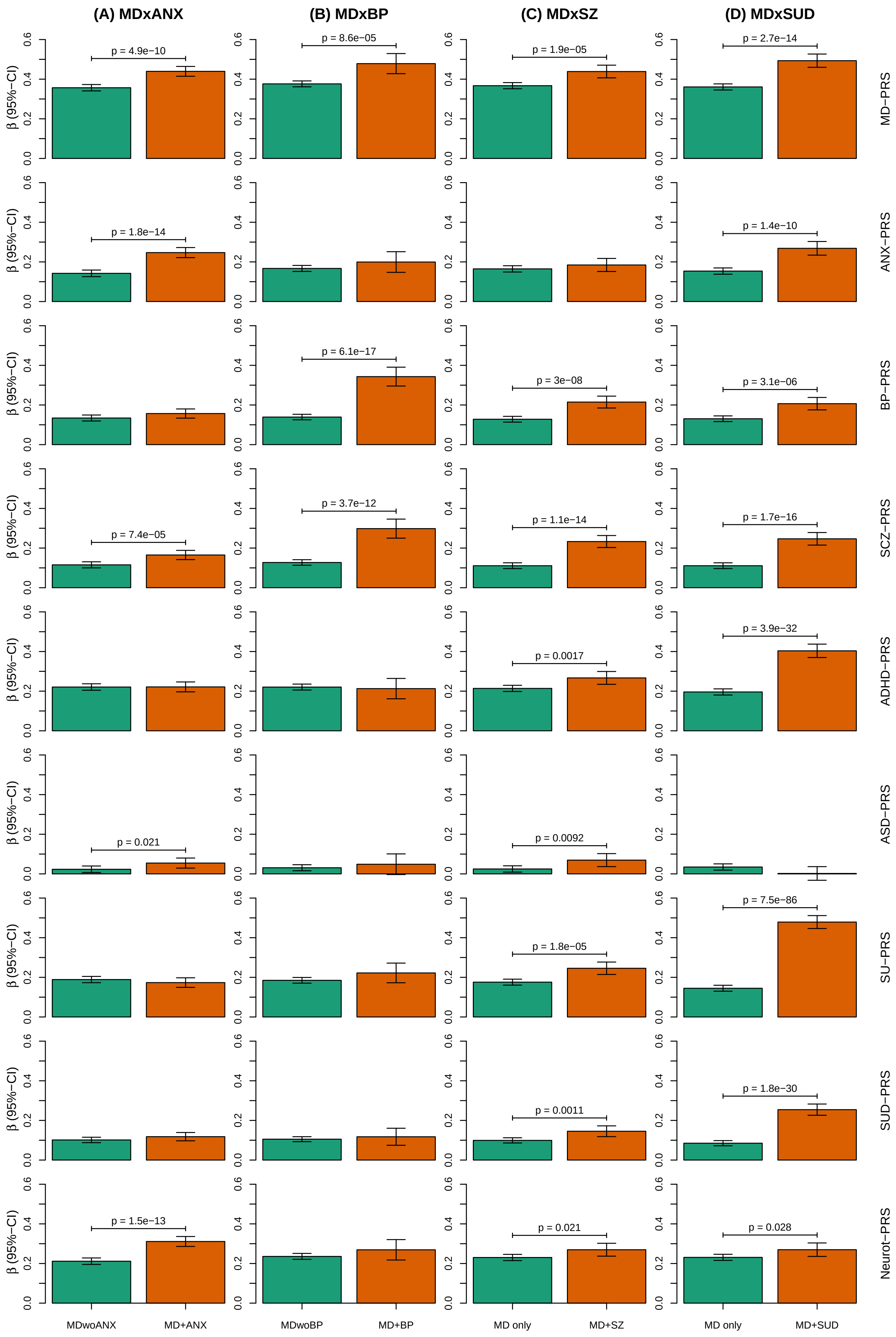

(A) MDxANX
(B) MDxBP
(C) MDxSZ
(D) MDxSUD
β (95%−CI)
MD−PRS
ANX−PRS
BP−PRS
SCZ−PRS
ADHD−PRS
ASD−PRS
SU−PRS
SUD−PRS
Neurot−PRS
p = 4.9e−10
p = 8.6e−05
p = 1.9e−05
p = 2.7e−14
p = 1.8e−14
p = 1.4e−10
p = 6.1e−17
p = 3e−08
p = 3.1e−06
p = 7.4e−05
p = 3.7e−12
p = 1.1e−14
p = 1.7e−16
p = 0.0017
p = 3.9e−32
p = 0.021
p = 0.0092
p = 1.8e−05
p = 7.5e−86
p = 0.0011
p = 1.8e−30
p = 1.5e−13
p = 0.021
p = 0.028
MDwoANX
MD+ANX
MDwoBP
MD+BP
MD only
MD+SZ
MD only
MD+SUD

Figure S19 *(previous page)*: Multivariate PRS analyses of MD with/without diagnoses of ANX, BP, SZ or SUD.. Each collumn represents one of four seperate mvPRS analyses of four different sets of MD-subphenotypes, one mvPRS analysis corresponds to one collumn, i.e. MD without vs (A) MD with ANX, (B) MD with BP, (C) MD with SZ and (D) MD with SUD respectively. On the x-axis: MD-subphenotype. On the y-axis: $\beta$ is the slope of the linear regression (95% CI). Significant difference between $\beta$ for MD without/with an additional diagnosis is indicated with horizontal line with nominal p-value above, i.e. the Wald test of equal group effect. (A) MDxANX ($N_{ctrl}$ = 38142, $N_{MDwoANX}$ = 22114, $N_{MDwANX}$ = 7044), p-values corresponds to $P_{MDwoANX}$ and $P_{MDwANX}$ for various PRSs in supplementary table 19A, overall p-value =1.2e-21. (B) MDxBP, ($N_{ctrl}$ = 38200, $N_{MDwoBP}$ = 29158, $N_{MDwBP}$ = 1460), p-values corresponds to $P_{MDwoBP}$ and $P_{MDwBP}$ for various PRSs in supplementary table 19B,overall p-value =1.5e-15. (C) MDxSZ, ($N_{ctrl}$ = 38142, $N_{MDwoSZ}$ = 25253, $N_{MDwSZ}$ = 3905), p-values corresponds to $P_{MDwoSZ}$ and $P_{MDwSZ}$ for various PRSs in supplementary table 19C, overall p-value =1.7e-15. (D) MDxSUD, ($N_{ctrl}$ = 38142, $N_{MDwoSUD}$ = 25620, $N_{MDwSUD}$ = 3538), p-values corresponds to $P_{MDwoSUD}$ and $P_{MDwSUD}$ for various PRSs in supplementary table 19D, overall p-value =6.4e-97. Cases with BP disorder were excluded from MDxANX, MDxSZ and MDxSUD analyses.

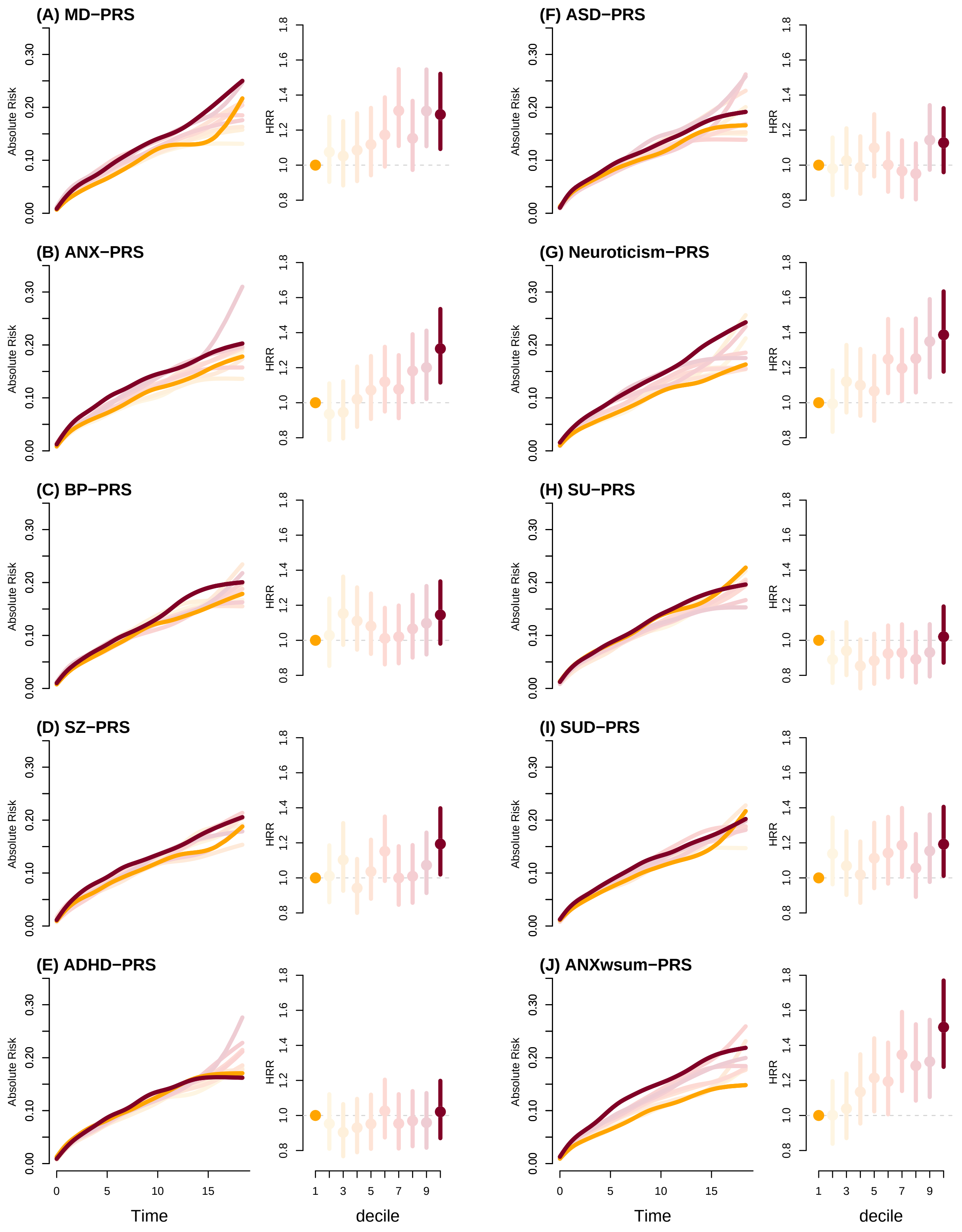

Figure S20: Absolute risk and Hazard rate ratios (HRR) of developing anxiety for deciles of ten different PRSs estimated using cox's regression. HRRs (95%) were calculated for decile 2-10 compared to the first decile. The analyses were conducted for PRS deciles of MD, BP, SCZ, ADHD, ASD, Neuroticism, Substance Use, Substance Use Disorder and sum of PRSs weighted by log($OR_{ANX}$). (A) MD-PRS, (B) ANX-PRS, (C) BP-PRS, (D) SZ-PRS, (E) ADHD-PRS, (F) ASD-PRS, (G) Neuroticism-PRS, (H) SU-PRS, (I) SUD-PRS, (J) ANXwsum-PRS,

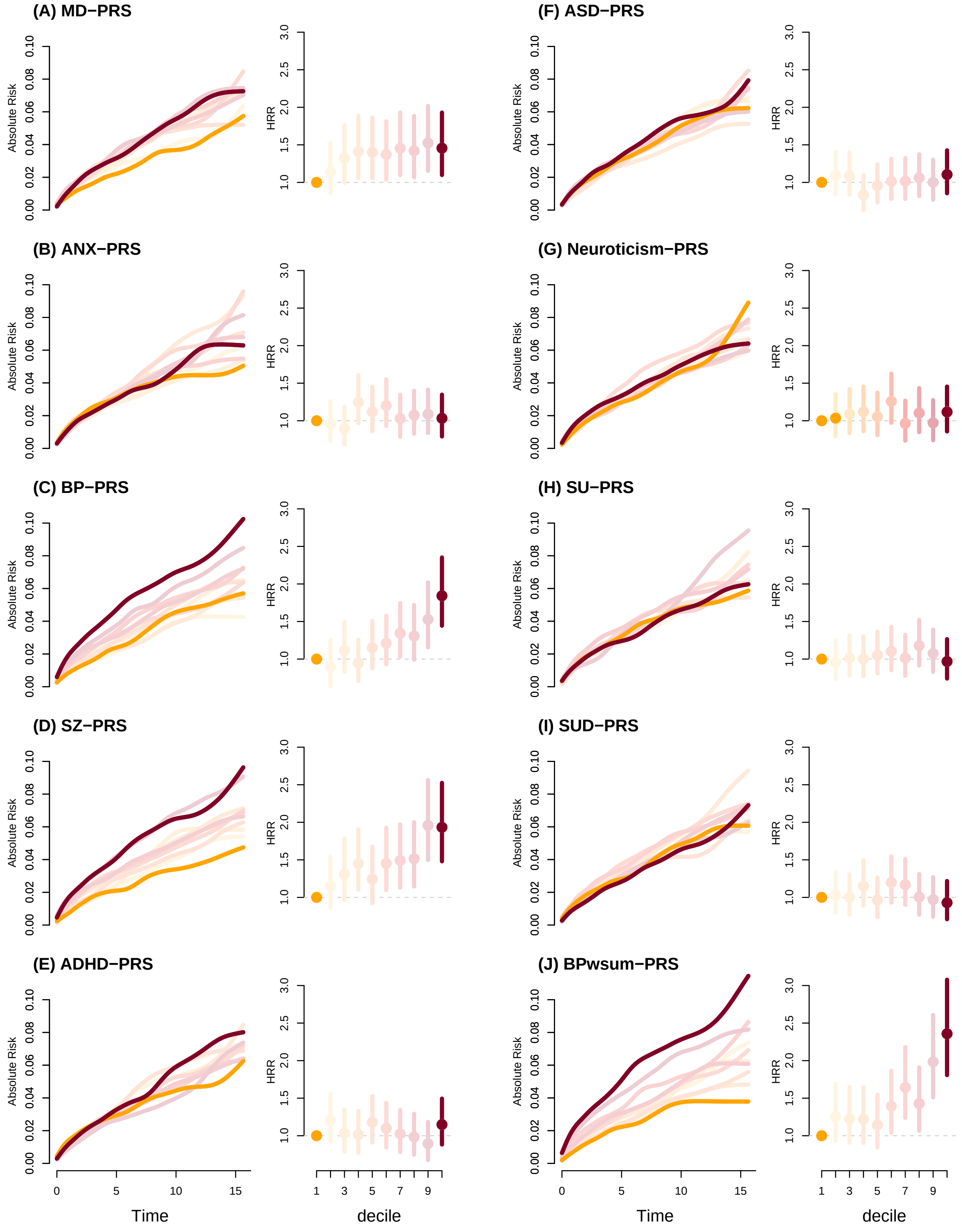

Figure S21: Absolute risk and Hazard rate ratios (HRR) transition into a bipolar disorder diagnosis for deciles of ten different PRSs estimated using cox's regression. HRRs (95%) were calculated for decile 2-10 compared to the first decile. The analyses were conducted for PRS deciles of MD, BP, SCZ, ADHD, ASD, Neuroticism, Substance Use, Substance Use Disorder and sum of PRSs weighted by log($OR_{BP}$). (A) MD-PRS, (B) ANX-PRS, (C) BP-PRS, (D) SZ-PRS, (E) ADHD-PRS, (F) ASD-PRS, (G) Neuroticism-PRS, (H) SU-PRS, (I) SUD-PRS, (J) BPwsum-PRS, but see Musliner, K. L. et al. Polygenic Risk and Progression to Bipolar or Psychotic Disorders Among Individuals Diagnosed With Unipolar Depression in Early Life. Am J Psychiatry 177, 936-943, doi:10.1176/appi.ajp.2020.19111195 (2020).

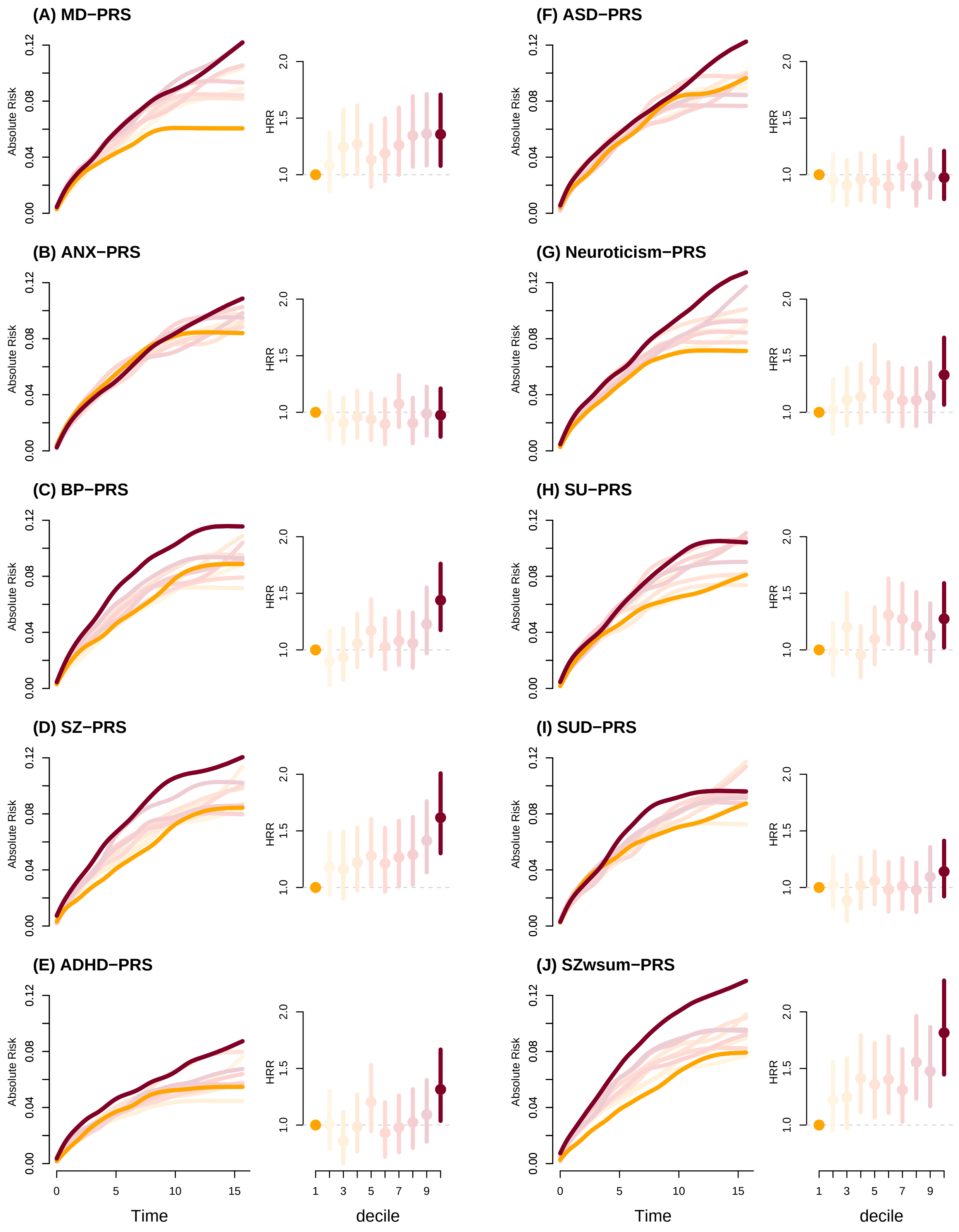

Figure S22: Absolute risk and Hazard rate ratios (HRR) transition into a schizophrenia diagnosis for deciles of ten different PRSs estimated using cox's regression. HRRs (95%) were calculated for decile 2-10 compared to the first decile. The analyses were conducted for PRS deciles of MD, BP, SCZ, ADHD, ASD, Neuroticism, Substance Use, Substance Use Disorder and sum of PRSs weighted by log($OR_{SZ}$). (A) MD-PRS, (B) ANX-PRS, (C) BP-PRS, (D) SZ-PRS, (E) ADHD-PRS, (F) ASD-PRS, (G) Neuroticism-PRS, (H) SU-PRS, (I) SUD-PRS, (J) SZwsum-PRS,

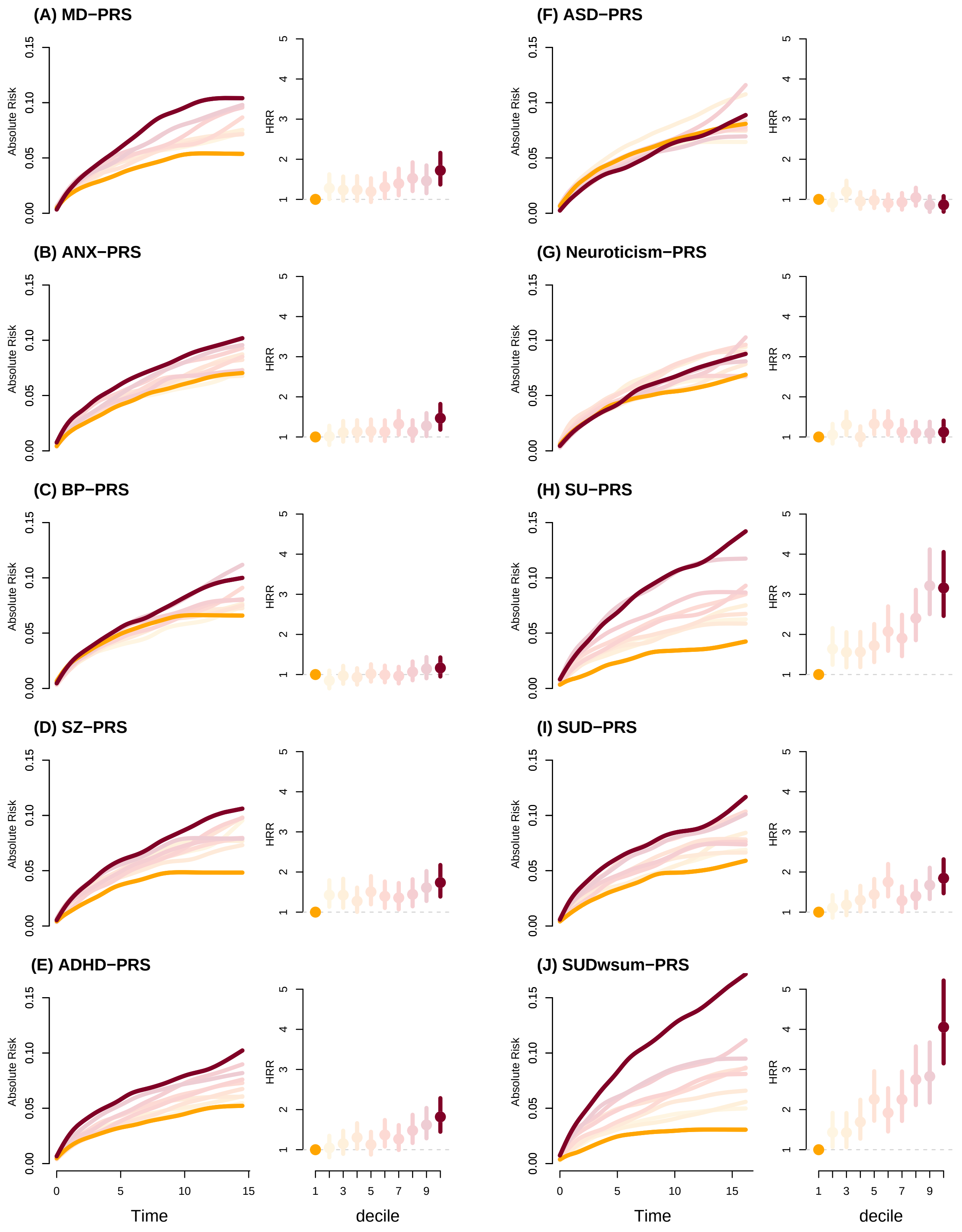

Figure S23: Absolute risk and Hazard rate ratios (HRR) of devloping substance use disorder for deciles of ten different PRSs estimated using cox's regression. HRRs (95%) were calculated for decile 2-10 compared to the first decile. The analyses were conducted for PRS deciles of MD, BP, SCZ, ADHD, ASD, Neuroticism, Substance Use, Substance Use Disorder and sum of PRSs weighted by log($OR_{\text{MDwSUD vs MDwoSUD}}$). (A) MD-PRS, (B) ANX-PRS, (C) BP-PRS, (D) SZ-PRS, (E) ADHD-PRS, (F) ASD-PRS, (G) Neuroticism-PRS, (H) SU-PRS, (I) SUD-PRS, (J) SUDwsum-PRS,
